# Implementation and results of active vaccine safety monitoring during the COVID-19 pandemic in the UK

**DOI:** 10.1101/2024.11.06.24316782

**Authors:** Jenny Wong, Katherine Donegan, Kendal Harrison, Tahira Jan, Alison Cave, Phil Tregunno

**Author notes:** Corresponding Author: Phil Tregunno.

## Abstract

**Introduction:** Yellow Card Vaccine Monitor (YCVM) was established by the UK Medicines and Healthcare products Regulatory Agency (MHRA) to facilitate active monitoring of adverse reactions following COVID-19 vaccination and further characterise safety in populations under-represented in clinical trials.

**Methods:** Randomly selected individuals were invited to register and actively contacted to seek further information on the vaccines received and adverse events they experienced. Demographics of patients recruited, and summaries of reported data, are presented alongside detailed analyses of the sub-cohort of pregnant and breast-feeding patients and analyses conducted to support regulatory assessment of two safety signals, menstrual disorders, and tinnitus.

**Results:** 36,604 individuals registered, with 30,281 reporting vaccination. Median [IQR] follow-up was 184 days [14-367]. Demographics of the recruited cohort reflected the vaccinated population and timing of invitations. 15,764 (52.1%) of those reporting vaccination, reported experiencing at least one adverse reaction. However, nearly all were expected acute reactions and only 4,134 (13.7%) reported an event considered medically serious. The data raised no safety concerns in pregnant and breast-feeding patients. Reporting of menstrual disorders appeared stimulated by media interest, as seen in spontaneous reporting systems. Data on the incidence of tinnitus were used to support regulatory action on this signal.

**Discussion:** Active surveillance provided a complimentary data source for monitoring the safety of COVID- 19 vaccines. However, further efforts are needed to recruit ethnic minorities. The technology developed has enhanced regulatory vigilance options and could be valuable in the future for actively monitoring the safety of innovative products used in small populations.

## 1 Introduction

Upon the introduction of any new vaccination programme, a large and diverse patient population receives the vaccine over a very short period. While clinical trials provide data on adverse events, particularly common and acute reactions in the trial population, there remains a clear need to continuously monitor safety and rapidly increase the evidence base on the benefits and risks [1]. This is in order to identify rare adverse events, which trials are underpowered to detect, as well as to further explore the overall safety profile of the vaccine in wider routine use including in groups excluded from clinical trials.

The Yellow Card Vaccine Monitor (YCVM) was set up by the UK Medicines and Healthcare products Regulatory Agency (MHRA) to actively collect suspected adverse vaccine reactions reported in association with a COVID-19 vaccine within targeted populations in a real-world setting [2]. The YCVM was implemented by the MHRA under their statutory responsibility to operate a national pharmacovigilance system and is classed as surveillance.

This platform was one of the four complimentary strands of the MHRA’s proactive vigilance strategy for COVID-19 vaccines [3]. It was designed to complement the existing Yellow Card scheme, through which healthcare providers, patients, and their carers can report incidences of suspected side effects of medicines including vaccines. Established spontaneous reporting systems are a critical in the detection of rare risks however they are limited in particular by underreporting meaning the data cannot be reliably used to estimate the frequency of an adverse event and, because of the challenges in obtaining follow up, spontaneously reported data generally capture a snapshot in time, with ongoing longitudinal information and patient outcomes not routinely available. This can limit the use of the data in monitoring vaccine safety [1.a.i.44].

The primary purpose of the YCVM was not to detect safety signals or identify rare risks, although, if signals of potentially more common adverse events arose from other sources, it could be used as supportive evidence. Rather it was designed to compare the frequency and severity of adverse reactions seen in routine use to those seen in clinical trials to allow further characterisation of the safety profile particularly in subgroups under-represented in clinical trials.

In response to potential signals identified by the MHRA through the data amassed through the different strands of the COVID-19 surveillance strategy or from elsewhere, safety reviews were conducted to evaluate evidence from multiple sources such as clinical trial data, spontaneous ADR data, and epidemiological data from literature.

YCVM was established prior to deployment of the first vaccine outside of a clinical trial which occurred on 8^th^ December. In England, the COVID-19 vaccination programme began by offering vaccinations to priority groups including those in residential care, frontline health and social care workers, older patients, and those considered clinically vulnerable before vaccinations were offered to younger patients and subsequently to those under 18 years and pregnant women [5,6,7,8]. Those with severe immunosuppression, who may not have mounted a full immune response, were offered a third dose after their primary course (two doses) with other patients offered a first booster [9]. Later doses were offered according to eligibility criteria recommended by the Joint Committee on Vaccination and Immunisation (JCVI).

This paper presents the data profile of the individuals registered to the YCVM along with the reports of suspected adverse reactions following a COVID-19 vaccination that they submitted. Three case studies are also discussed in further detail to demonstrate how the data from YCVM contributed to safety evaluations. This includes two safety signals which first arose from other data sources, and which were evaluated as part of the MHRAs continuous benefit risk assessment process during the pandemic, menstrual disorders and tinnitus, and the specific surveillance of pregnant women.

Learnings from the experience with implementing active surveillance using this technology in a regulatory setting are presented and proposals and opportunities for future development and use discussed.

## 2 Methods

### 2.1 Description of Yellow Card Vaccine Monitor platform

In England, a central NHS system identified individuals who were eligible for the COVID-19 vaccine and invited them by letter to book their COVID-19 vaccination. Through this call/recall system, randomly selected individuals were additionally invited to voluntarily register for follow-up via the YCVM digital platform. The groups of individuals sent invitations by the NHS to register with the YCVM aligned with the call-in priorities for vaccination during vaccination programme, taking into account the distribution of individuals registered by region and ethnicity. The timing of invitations was largely determined by the deployment of the vaccine programme to new groups of individuals. In addition to random invites, pregnant women were also encouraged to register with the YCVM in information provided to them when they were considering or having their vaccination [10].

Enrolment was sought prior to vaccination and individuals, or their carers, were required to read and agree to the User Sign-up Agreement and the privacy policy, in order to participate. To support accessibility, a telephone service and an easy read invitation letter were also offered.

At enrolment, patient information including demographics, NHS number, and pregnancy and breastfeeding information, were collected where relevant. Information on past medical history, receipt of a seasonal flu vaccine and COVID-19 infection were also sought.

Once registered, individuals were contacted via email or SMS at set intervals to ask for further information, including about the vaccinations received and whether they had experienced any suspected adverse drug reactions (ADR). Follow up was sought at one week, one month, and six months post vaccination. They were also provided the opportunity to update or add to their previously submitted information, including outcomes for any previously reported reactions. The seriousness of an ADR could be reported by selecting from the optional seriousness categories. When new information was submitted by the patient, a new submission was recorded. Where a suspected ADR was recorded, the data was entered onto the MHRA’s ADR reporting database and evaluated alongside all other data in safety surveillance activities as outlined in the COVID-19 vigilance strategy.

The YCVM data specification met ICH E2B guidelines on the electronic transmission of individual case reports [11].

### 2.2 Statistical analysis

This paper presents all data submitted up to 31^st^ December 2022, with data locked at this point. Only data reported directly through the YCVM are reported, with any additional follow up of individual cases undertaken separately excluded.

Descriptive analyses were conducted to evaluate the extent and timing of registrations with the YCVM and all data submissions and the duration of follow up. In these analyses, where there were multiple submissions by a patient on a single day, only one per patient was counted. The follow-up time contributed per patient was derived from the later of the individual’s registration date or first reported vaccination date and the date of their last submission of information on the YCVM.

Patient characteristics are summarised for all registered patients, patients who reported vaccination data (further stratified by vaccine brand and dose), and patients who also reported an ADR following a vaccination. To note, information on patient sex, i.e. male/female, was requested within the YCVM as opposed to gender. Patients were classified as having immunosuppression if they indicated that they had any of a prespecified list of medical conditions or were taking specific medications that might lower their immune response.

To explore those actively submitting ADR information to the YCVM over time, analyses were conducted to characterise the cohorts who had reported a 1^st^ dose, those who reported a 1^st^ and 2^nd^ dose, and also those who reported 1^st^, 2^nd^ and 3^rd^/booster doses. Amongst these cohorts, the proportion reporting an ADR or a MedDRA serious ADR was calculated and stratified by patient demographics. The dose sequence in the analyses was derived by the vaccination date reported by the patient or carer. The YCVM makes no distinction between first booster and third doses in the data collection stage, therefore 3rd dose in the analyses refers to the dose taken after their primary course (1st and 2^nd^ dose). The number of ADRs reported and the most common ADR events reported for each vaccine and each dose was extracted. These most commonly reported ADR events were explored using the corresponding MedDRA preferred term (PT) and system organ class (SOC) terms [12].

Reporters were able to indicate the seriousness of their ADRs against at least one of a number of non-mandatory options describing their impact. The most serious indicator was then assigned to the individual’s whole report regardless of which dose an ADR occurred in temporal association with. Note the categories for the level of seriousness (including added terms: mild or lightly uncomfortable; uncomfortable, a nuisance or irritation, but able to carry on with everyday activities; and had short term effect that was bad enough to affect every day activities) were adapted from CIOMS guidance to allow patients to better describe events that were non-serious [13]. The number of patients reporting ADRs identified as MedDRA serious events and the number of these ADRs were analysed. MedDRA seriousness is as defined by MedDRA through agreement of clinical experts.

A subgroup analysis was conducted on female patients who reported as being pregnant or breastfeeding at the time of vaccination or during their follow-up time in the YCVM. The proportion reporting an ADR after a COVID-19 vaccination dose was calculated and stratified by patient demographics. In the pregnancy analysis, the reported stage of their pregnancy at the time of vaccination entered by a pregnant individual is presented. These categories include first trimester (1- 12 weeks), second trimester (13-28 weeks) and third trimester (29-40 weeks). When an individual patient reports vaccination in more than one trimester, they are assigned into a category indicating more than one stage was reported. If no stage of pregnancy was reported, they were categorised as unknown.

For the two case studies, which are presented as examples of analyses conducted as part of ongoing safety monitoring, the numbers of patients reporting an ADR relating to menstrual disorders (female patients) or tinnitus were extracted. MedDRA search term lists for identifying cases of menstrual disorders and tinnitus are in Appendix A. The onset time for tinnitus was derived by subtracting the date of the ADR event from the date of the most recent vaccination. The outcomes of ADRs are based on the outcome as reported at the time of the individual’s last submission.

## 3 Results

### 3.1 Registered Patients

36,604 individuals registered with the YCVM, with the first individual registering in November 2020 and the last registration within the study period in December 2022. The number of registrations and submissions by calendar month and year are shown in Figure 1. The time period for the majority of registrations aligns with the period in which first doses were initially offered in the UK, while peaks in later submissions align with the offer of second and subsequent doses. A total of 190,034 submissions were submitted by the 36,604 registered individuals during the study period. The median number of submissions submitted was 3 (IQR 2-7).

**Figure 1.**
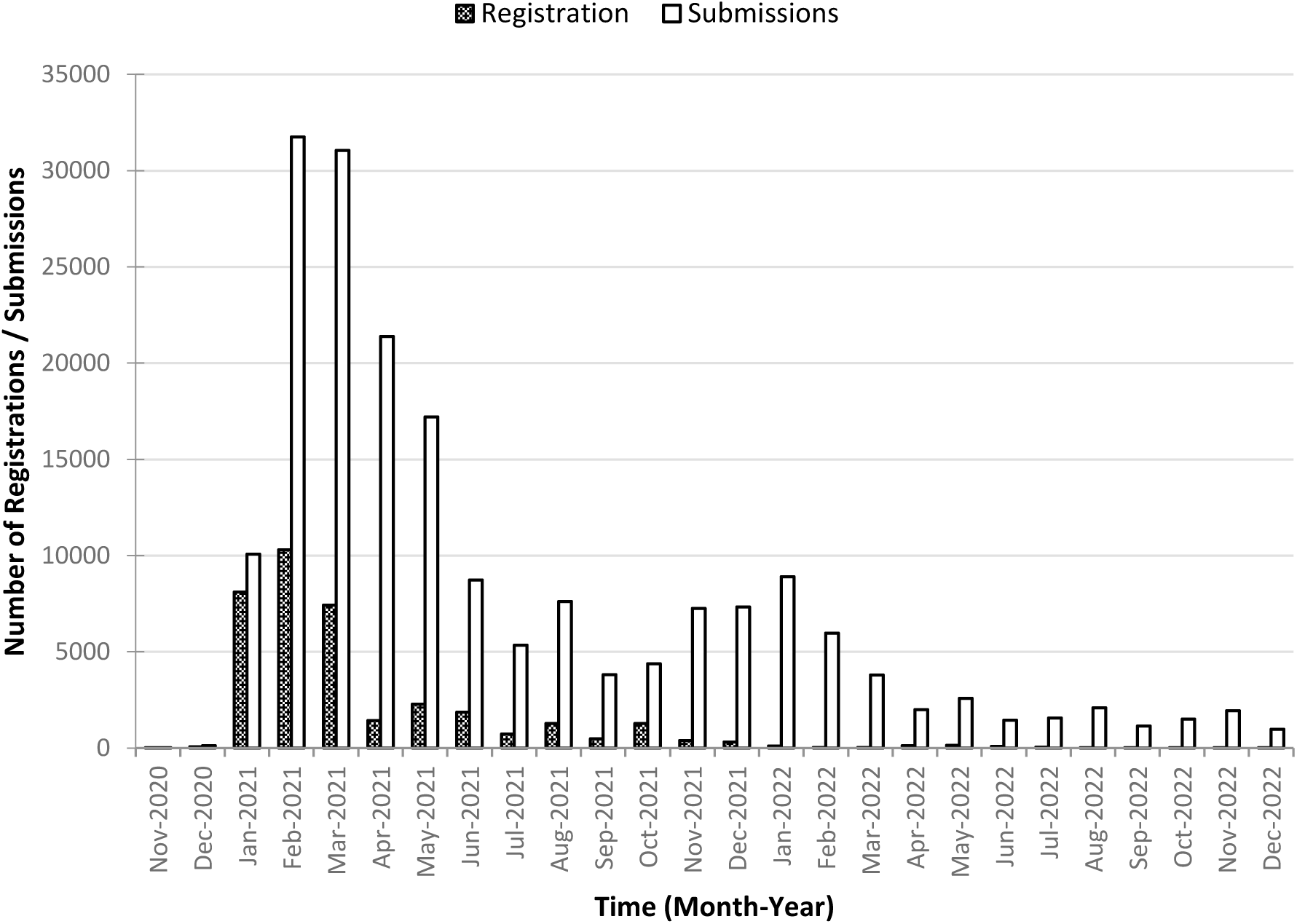
Number of registrations and submissions to the Yellow Card Vaccine Monitor up to 31^st^ December 2022

Table 1 presents the demographics of patients registered in the YCVM and the subset of those who reported a vaccination during their follow-up time in the YCVM. The median age of the cohort reported at registration was 62 years (interquartile range, IQR: 42-73). 10,791 (35.6%) of those reporting a vaccination had a registration date before their first vaccination, while 14,369 (47.5%) registered after the date of their first vaccination. Despite vaccination only being recommended in patients aged 5+ years in the UK within the study period, 63 patients reported an age < 5 years. The median age amongst those who had reported a vaccination was 63 years (IQR: 43-74). Other demographics of those reporting a vaccination were also broadly the same as those registering. The median follow-up in those reporting a vaccination was 184 patient days (IQR:14-367, range:1-697) and 82.1% of the total follow-up time was from those aged 50 years or older. Female individuals contributed slightly more to YCVM than males, approximately 55% of the total follow-up time. Females were predominant contributors amongst the younger age groups up until the 70-79 year age group with particularly greater levels of follow up in the 50-59 year and 30–39 year age groups compared to men (median 273 vs 264 patient days and 60 vs 52 patient days respectively). In the older age groups, men contributed greater levels of follow-up than women (median 181 days vs 93 patient day amongst the 80+ year age group, and 324 vs 286 patient days in the 70–79-year age group.) The follow-up time stratified by patient sex is presented in Appendix B to support contextualisation of the analyses on vaccinations in pregnancy.

**Table 1.** Number of individuals registered in the Yellow Card Vaccine Monitor stratified by vaccination reporting status and total follow up time accumulated in individuals reporting a vaccination.

|  | Patient count, n (%) |  |  |  | Follow-up time (days) |  |
| --- | --- | --- | --- | --- | --- | --- |
|  | All registered patients |  | Patients reporting vaccination data |  | Total | Median (IQR) |
| Number of registered patients | 36,604 |  | 30,281 |  | 6,808,705 | 184 (14-367) |
| Sex |  |  |  |  |  |  |
| Male | 15,223 | (41.6) | 12,424 | (41.0) | 3,040,806 | 249 (16-383) |
| Female | 20,868 | (57.0) | 17,463 | (57.7) | 3,701,682 | 179 (10-363) |
| Unknown | 513 | (1.4) | 394 | (1.3) | 66,217 | 74 (1-333) |
| Age-bands |  |  |  |  |  |  |
| Under 12 | 195 | (0.5) | 120 | (0.4) | 11,489 | 18 (1-183) |
| 12-17 | 2,109 | (5.8) | 1,238 | (4.1) | 107,662 | 17 (2-127) |
| 18-29 | 1,747 | (4.8) | 1,399 | (4.6) | 141,719 | 15 (1-184) |
| 30-39 | 4,133 | (11.3) | 3,613 | (11.9) | 489,190 | 59 (3-244) |
| 40-49 | 3,286 | (9.0) | 2,713 | (9.0) | 465,917 | 79 (8-285) |
| 50-59 | 4,926 | (13.5) | 3,936 | (13.0) | 941,784 | 270 (17-365) |
| 60-69 | 7,118 | (19.4) | 6,029 | (19.9) | 1,738,576 | 296 (57-450) |
| 70-79 | 10,464 | (28.6) | 8,899 | (29.4) | 2,435,668 | 304 (53-407) |
| 80+ | 2,596 | (7.1) | 2,309 | (7.6) | 471,414 | 123 (8-365) |
| Unknown | 30 | (0.1) | 25 | (0.1) | 5,286 | 91 (1-380) |
| Ethnicity |  |  |  |  |  |  |
| White British, White Irish, or any other white background | 31,331 | (85.6) | 26,281 | (86.8) | 6,208,814 | 213 (15-372) |
| Other | 2,392 | (6.5) | 1,759 | (5.8) | 262,287 | 57 (4-273) |
| Unknown | 2,881 | (7.9) | 2,241 | (7.4) | 337,604 | 72 (2-276) |
| BMI Category |  |  |  |  |  |  |
|  | All registered patients |  | Patients reporting vaccination data |  | Total | Median (IQR) |
| Underweight | 842 | (2.3) | 667 | (2.2) | 146,265 | 183 (8-369) |
| Normal | 10,471 | (28.6) | 8,928 | (29.5) | 2,189,172 | 245 (16-381) |
| Overweight | 9,727 | (26.6) | 8,184 | (27.0) | 2,008,210 | 248 (17-383) |
| Obese | 5,362 | (14.6) | 4,436 | (14.6) | 1,005,914 | 184 (15-367) |
| Unknown | 10,202 | (27.9) | 8,066 | (26.6) | 1,459,144 | 88 (8-333) |
| **Immunocompromised** |  |  |  |  |  |  |
| Yes | 4,162 | (11.4) | 3,483 | (11.5) | 811,593 | 195 (16-368) |
| No/Unknown | 32,442 | (88.6) | 26,798 | (88.5) | 5,997,112 | 183 (13-367) |
| **Reported as pregnant at the time of a vaccination** |  |  |  |  |  |  |
| Yes | 2,517 | (6.9) | 2,416 | (8.0) | 330,018 | 73 (5-244) |
| No/Unknown | 34,087 | (93.1) | 27,865 | (92.0) | 6,478,687 | 189 (15-370) |

### 3.2 COVID-19 vaccinations reported

The demographics of patients stratified by the vaccination brand(s) they reported receiving across all doses (N=56,371) are presented in Table 2. A patient could receive doses from different product brands during their COVID-19 vaccination regimen, therefore a patient could appear in more than one column. Data are presented for the Pfizer, AstraZeneca, and Moderna monovalent vaccines. Of those reporting receiving another vaccine brand, 2 patients reported receiving a Moderna bivalent vaccine, 10 a Novavax vaccine, and 2 a Janssen vaccine.

**Table 2.** Demographics of patients stratified by vaccine brand administered (% of doses reported of the vaccine brand).

|  | Pfizer | AstraZeneca | Moderna | Unknown or other brand |
| --- | --- | --- | --- | --- |
| Number of patients reporting a vaccine dose | 16,509 | 17,713 | 4,467 | 1,125 |
| Number of doses (% of total doses) | 24,405 (43.3) | 25,515 (45.3) | 5,228 (9.3) | 1,223 (2.2) |
| **Sex** |  |  |  |  |
| Male | 6,614 (40.1) | 7,738 (43.7) | 1,944 (43.5) | 504 (44.8) |
| Female | 9,687 (58.7) | 9,763 (55.1) | 2,483 (55.6) | 594 (52.8) |
| Unknown | 208 (1.3) | 212 (1.2) | 40 (0.9) | 27 (2.4) |
| **Age-bands** |  |  |  |  |
| Under 12 | 90 (0.5) | 22 (0.1) | 12 (0.3) | 11 (1.0) |
| 12-17 | 1,207 (7.3) | 4 (0.0) | 18 (0.4) | 40 (3.6) |
| 18-29 | 1,023 (6.2) | 286 (1.6) | 199 (4.5) | 22 (2.0) |
| 30-39 | 2,630 (15.9) | 801 (4.5) | 552 (12.4) | 25 (2.2) |
| 40-49 | 958 (5.8) | 1,960 (11.1) | 394 (8.8) | 37 (3.3) |
| 50-59 | 1,280 (7.8) | 3,476 (19.6) | 592 (13.3) | 103 (9.2) |
| 60-69 | 2,748 (16.6) | 4,732 (26.7) | 1,023 (22.9) | 214 (19.0) |
| 70-79 | 5,156 (31.2) | 5,372 (30.3) | 1,451 (32.5) | 512 (45.5) |

|  | Pfizer |  | AstraZeneca |  | Moderna |  | Unknown or other brand |  |
| --- | --- | --- | --- | --- | --- | --- | --- | --- |
| 80+ | 1,405 | (8.5) | 1,043 | (5.9) | 223 | (5.0) | 160 | (14.2) |
| Unknown | 12 | (0.1) | 17 | (0.1) | 3 | (0.1) | 1 | (0.1) |
| **Ethnicity** |  |  |  |  |  |  |  |  |
| White British, White Irish, or any other white background | 14,218 | (86.1) | 15,833 | (89.4) | 4,039 | (90.4) | 1,016 | (90.3) |
| Other | 1,111 | (6.7) | 720 | (4.1) | 183 | (4.1) | 28 | (2.5) |
| Unknown | 1,180 | (7.1) | 1,160 | (6.5) | 245 | (5.5) | 81 | (7.2) |
| **BMI Category** |  |  |  |  |  |  |  |  |
| Underweight | 423 | (2.6) | 323 | (1.8) | 84 | (1.9) | 33 | (2.9) |
| Normal | 5,041 | (30.5) | 5,229 | (29.5) | 1,556 | (34.8) | 289 | (25.7) |
| Overweight | 4,256 | (25.8) | 5,205 | (29.4) | 1,277 | (28.6) | 342 | (30.4) |
| Obese | 2,222 | (13.5) | 2,839 | (16.0) | 613 | (13.7) | 153 | (13.6) |
| Unknown | 4,567 | (27.7) | 4,117 | (23.2) | 937 | (21.0) | 308 | (27.4) |
| **Immunocompromised** |  |  |  |  |  |  |  |  |
| Yes | 1,916 | (11.6) | 2,096 | (11.8) | 519 | (11.6) | 166 | (14.8) |
| No/Unknown | 14,593 | (88.4) | 15,617 | (88.2) | 3,948 | (88.4) | 959 | (85.2) |

Table 3 presents a summary of the number and demographics of patients reporting a vaccination stratified by dose number and vaccine brand. In addition, three patients reported a 6^th^ dose. The median age of patients in the subgroup reporting for 4^th^ or 5^th^ doses was older compared to those reporting primary vaccination doses. This reflects the narrowing eligibility criteria for booster doses. The higher median age of patients reporting a first or second dose of the AstraZeneca vaccine reflects the advised preferential use of alternatives in patients aged <40 years following the signal of thrombosis with thrombocytopenia adverse events. Amongst those who reported receiving a 1^st^ dose, 11.5% (n=3,146) were identified as immunocompromised. The proportion of immunocompromised increased across the doses, with 11.5% (n=1,691) in 2^nd^ dose, 12.2% (n=1,217) in 3^rd^ dose, 15.5% (n=527) in 4^th^ dose and 25.1% (n=242) in 5^th^ dose recipients. Again this reflects the narrowing eligibility criteria for booster doses. The proportion of reported vaccines marked as unknown or other brand increased slightly with dose number.

**Table 3.** The demographics of vaccine recipients stratified by reported dose number and vaccine brand.

|  | Number of patients<br>(% of those receiving dose) |  | Median age<br>(IQR) |  | Number (%) female |  | Number (%) immuno-compromised |  |
| --- | --- | --- | --- | --- | --- | --- | --- | --- |
| Dose 1 | 27,403 | (100) | 63 | (43-74) | 15,895 | (100) | 3146 | (100) |
| Pfizer | 10,249 | (37.4) | 54 | (31-75) | 6300 | (39.6) | 1152 | (36.6) |
| AstraZeneca | 16,228 | (59.2) | 65 | (55-73) | 8988 | (56.5) | 1910 | (60.7) |
| Moderna | 550 | (2.0) | 34 | (30-40) | 402 | (2.5) | 24 | (0.8) |
| Unknown/other | 376 | (1.3) | 73 | (57-77) | 205 | (1.3) | 60 | (1.9) |
| Dose 2 | 14,657 | (100) | 65 | (50-74) | 8344 | (100) | 1691 | (100) |
| Pfizer | 5,023 | (34.4) | 66 | (34-75) | 3072 | (36.8) | 586 | (34.7) |
| AstraZeneca | 9,105 | (62.1) | 65 | (57-73) | 4923 | (59.0) | 1060 | (62.7) |
| Moderna | 302 | (2.1) | 35.5 | (31-42) | 226 | (2.7) | 13 | (0.8) |
| Unknown/other | 227 | (1.5) | 73 | (61-78) | 123 | (1.5) | 32 | (1.9) |
| Dose 3 | 9,935 | (100) | 67 | (56-74) | 5389 | (100) | 1217 | (100) |
| Pfizer | 7,505 | (75.5) | 69 | (58-75) | 4018 | (74.6) | 972 | (79.9) |
| AstraZeneca | 161 | (1.6) | 71 | (62-75) | 76 | (1.4) | 25 | (2.1) |
| Moderna | 1,992 | (20.1) | 58 | (46-65) | 1157 | (21.5) | 176 | (14.5) |
| Unknown/other | 277 | (2.8) | 74 | (66-77) | 138 | (2.6) | 44 | (3.6) |
| Dose 4 | 3,409 | (100) | 72 | (64-75) | 1691 | (100) | 527 | (100) |
| Pfizer | 1,386 | (40.7) | 68 | (59-75) | 728 | (43.1) | 245 | (46.5) |
| AstraZeneca | 18 | (0.5) | 73 | (62-77) | 8 | (0.5) | 3 | (0.6) |
| Moderna | 1,753 | (51.4) | 73 | (67-76) | 830 | (49.1) | 244 | (46.3) |
| Unknown/other | 252 | (7.4) | 72 | (63-75) | 125 | (7.4) | 35 | (6.6) |
| Dose 5 | 964 | (100) | 75 | (74-78) | 463 | (100) | 242 | (100) |
| Pfizer | 242 | (25.1) | 75 | (74-78) | 121 | (26.1) | 66 | (27.3) |
| AstraZeneca | 3 | (0.3) | 74 | (73-78) | 2 | (0.4) | 0 | (0.0) |
| Moderna | 628 | (65.1) | 75 | (74-78) | 296 | (63.9) | 160 | (66.1) |
| Unknown/other | 91 | (9.4) | 76 | (74-78) | 44 | (9.5) | 16 | (6.6) |

Of the 30,281 patients reporting a vaccine dose, 27,403 (90.5%) reported receiving a 1^st^ dose, 12,508 (41.3%) reported receiving a 1^st^ and 2^nd^ dose, and 7891 (26.1%) reported receiving a 1^st^, 2^nd^, and 3^rd^ dose. 2,878 (9.5%) reported receiving a second or subsequent vaccine dose but did not report details for their first dose. Within the UK COVID-19 vaccine programme, it was recommended an individual should receive the same vaccine product for their 1^st^ and 2^nd^ doses (primary course). Of those reporting both primary course doses, 99% reported receiving homologous dose regimens (n=12,070). Less than 1% of patients reported receiving a heterogenous course (n=148). Amongst the patients who had reported a 1^st^ dose and received a homogenous schedule, 63.4% (n=7,654) received the AstraZeneca vaccine whilst 34.8% (n=4,203) received the Pfizer vaccine and 1.8% (n=213) the Moderna vaccine.

### 3.3 Adverse events reported following vaccination

Patient demographics for those reporting an ADR are presented in Table 4. 15,764 patients (52.1%) of those reporting a vaccine dose, reported experiencing at least one ADR following vaccination. The median age of those who reported an ADR was 59 years (IQR: 42-71) and 68 years (IQR: 49-75) amongst those who did not report an ADR. Female patients more commonly reported ADRs and the rate of reporting of ADRs decreased with increasing dose number. Demographics of patients reporting an ADR for each dose stratified by vaccination brand are presented in Appendix C.

**Table 4.** Number of patients reporting an ADR following vaccination, stratified by patient demographics and vaccination dose reporting (Row %^1^).

|  | Patients reporting an ADR<br>(% of patient cohort reporting any vaccine dose) |  | Patients reporting an ADR after the 1<sup>st</sup> dose<br>(% of patient cohort reporting a 1<sup>st</sup> vaccine dose) |  | Patients reporting an ADR after the 2<sup>nd</sup> dose (% of patient cohort reporting 1<sup>st</sup> and 2<sup>nd</sup> vaccine doses) |  | Patients reporting an ADR after the 3<sup>rd</sup> dose<br>(% of patient cohort reporting 1<sup>st</sup>, 2<sup>nd</sup>, and 3<sup>rd</sup> vaccine doses) |  |
| --- | --- | --- | --- | --- | --- | --- | --- | --- |
| Sex |  |  |  |  |  |  |  |  |
| Male | 5,407 | (43.5) | 4173 | (37.4) | 839 | (16.0) | 592 | (17.1) |
| Female | 10,145 | (58.1) | 8009 | (50.4) | 1,764 | (24.7) | 1,134 | (26.1) |
| Unknown | 212 | (53.8) | 145 | (42.2) | 22 | (16.3) | 16 | (22.2) |
| Age bands |  |  |  |  |  |  |  |  |
| Under 12 | 45 | (37.5) | 36 | (34.0) | 4 | (10.5) | 1 | (9.1) |
| 12-17 | 592 | (47.8) | 519 | (42.5) | 81 | (30.7) | 15 | (19.5) |
| 18-29 | 829 | (59.3) | 669 | (51.0) | 141 | (30.8) | 64 | (29.8) |
| 30-39 | 1,983 | (54.9) | 1,567 | (47.6) | 359 | (28.0) | 133 | (20.7) |
| 40-49 | 1,937 | (71.4) | 1,668 | (66.6) | 275 | (27.0) | 201 | (32.4) |
| 50-59 | 2,505 | (63.6) | 2,041 | (58.2) | 379 | (22.9) | 367 | (31.7) |
| 60-69 | 3,430 | (56.9) | 2,639 | (49.3) | 618 | (21.0) | 523 | (25.1) |
| 70-79 | 3,715 | (41.7) | 2,666 | (33.4) | 666 | (16.6) | 391 | (14.7) |
| 80+ | 719 | (31.1) | 515 | (24.5) | 101 | (12.3) | 47 | (11.2) |
| Unknown | 9 | (36.0) | 7 | (31.8) | 1 | (20.0) | 0 | (0.0) |
| Ethnicity |  |  |  |  |  |  |  |  |
| White British, White Irish, or any other white background | 13,705 | (52.1) | 10,770 | (45.2) | 2,369 | (21.1) | 1,574 | (21.8) |
| Other | 966 | (54.9) | 789 | (48.3) | 149 | (26.5) | 97 | (30.8) |
| Unknown | 1,093 | (48.8) | 768 | (39.1) | 107 | (14.7) | 71 | (19.8) |
| BMI Category |  |  |  |  |  |  |  |  |
| Underweight | 386 | (57.9) | 291 | (47.3) | 79 | (28.5) | 50 | (31.1) |
| Normal | 4,904 | (54.9) | 3,840 | (47.5) | 906 | (22.5) | 594 | (22.4) |
| Overweight | 4,177 | (51.0) | 3,293 | (44.5) | 691 | (19.1) | 478 | (20.5) |
| Obese | 2,315 | (52.2) | 1,852 | (46.0) | 383 | (21.6) | 252 | (22.0) |
| Unknown | 3,982 | (49.4) | 3,051 | (41.9) | 566 | (20.1) | 368 | (23.0) |
| Immunocompromised category |  |  |  |  |  |  |  |  |
| Yes | 1,738 | (49.9) | 1,318 | (41.9) | 306 | (21.2) | 175 | (18.7) |
| No/Unknown | 14,026 | (52.3) | 11,009 | (45.4) | 2,319 | (21.0) | 1,567 | (22.5) |

<sup>1</sup> [TABLE FOOTNOTE] Proportion of patients who reported an ADR in association with a dose of a COVID-19 vaccine, stratified by patient demographics and by the patient cohort definition. Example: Amongst the male patients who had reported a 1<sup>st</sup> dose and a 2<sup>nd</sup> dose vaccination, 16.0% (n=839) had reported an ADR that occurred after the 2<sup>nd</sup> dose vaccination.

Reporters were able to select at least one level of seriousness for their report to describe the impact of an ADR on their lives although this was non-mandatory. The levels of seriousness indicated by reporters are presented in Table 5. The number of patients who had reported their ADRs with a level of seriousness was 9,670 (61.3%). These reports were associated with 22,920 events (60.1%). In the cohort of patients who had reported a 1^st^ dose in YCVM (n=14,587), 8,955 (61.4%) reported their ADRs with a level of seriousness. There were 21,484 (60.3%) reactions associated with these reports.

**Table 5.**
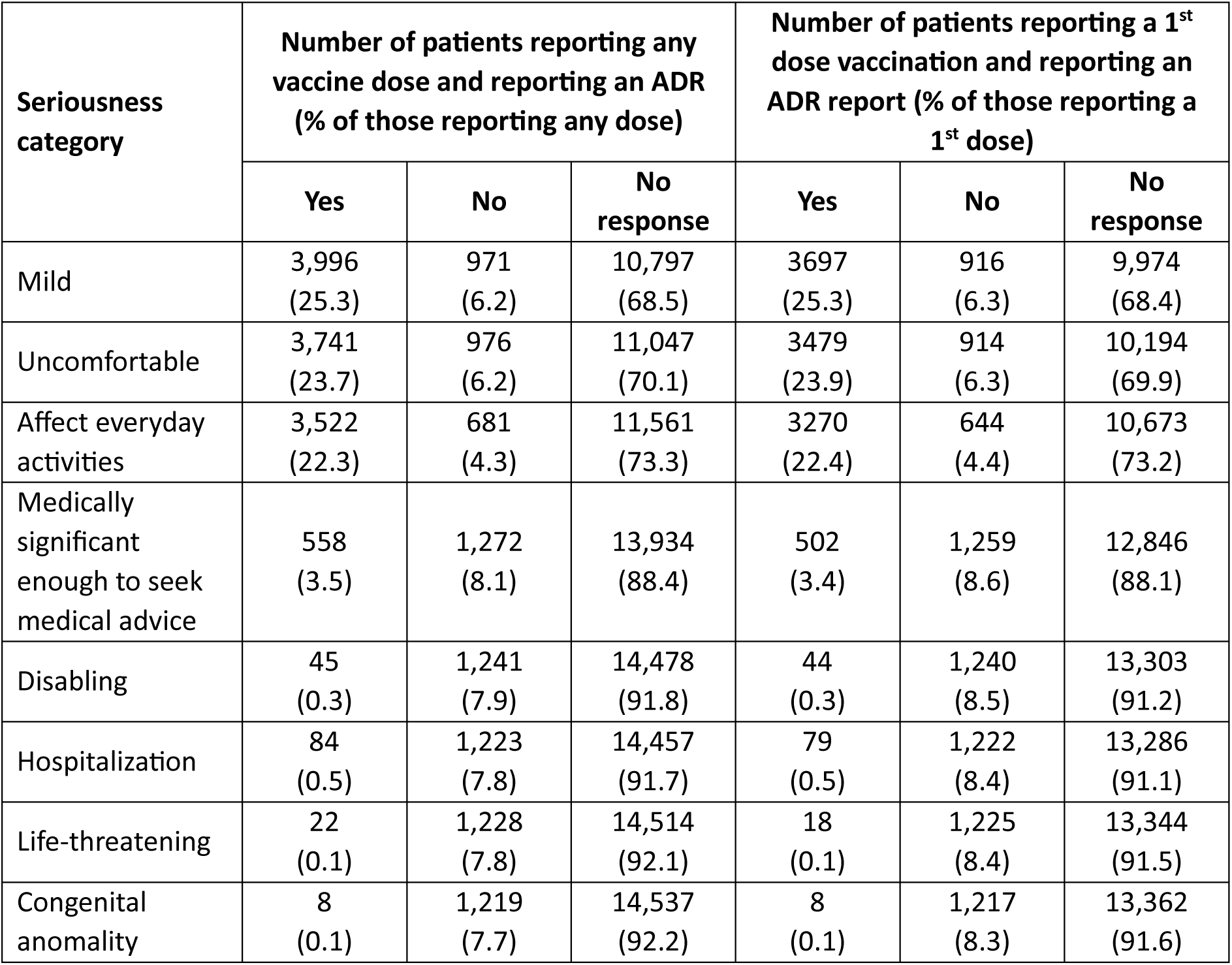
Impact of ADRs experienced using the self-reported seriousness categories, as reported by the patient.

When the reported medical events were categorised as medically serious according to the MedDRA classification, 4,134 (13.7% of those reporting any vaccination dose) patients reported an ADR considered medically serious. Table 6 presents a summary of the patient demographics for those reporting a MedDRA serious ADR.

**Table 6.** Number of patients reporting and not reporting a MedDRA serious ADR following vaccination, stratified by patient demographics and vaccination dose reporting (Row %^2^).

|  | Patients reporting a MedDRA serious ADR (% of patient cohort reporting any vaccine dose) |  | Patients reporting a MedDRA serious ADR after the 1<sup>st</sup> dose (% of patient cohort reporting a 1<sup>st</sup> vaccine dose) |  | Patients reporting a MedDRA serious ADR after the 2<sup>nd</sup> dose (% of patient cohort reporting 1<sup>st</sup> and 2<sup>nd</sup> vaccine doses) |  | Patients reporting a MedDRA serious ADR after the 3<sup>rd</sup> dose (% of patient cohort reporting 1<sup>st</sup>, 2<sup>nd</sup>, and 3<sup>rd</sup> vaccine doses) |  |
| --- | --- | --- | --- | --- | --- | --- | --- | --- |
| Sex |  |  |  |  |  |  |  |  |
| Male | 1,387 | (11.2) | 978 | (8.8) | 182 | (3.5) | 118 | (3.4) |
| Female | 2,696 | (15.4) | 1,890 | (11.9) | 342 | (4.8) | 208 | (4.8) |
| Unknown | 51 | (12.9) | 31 | (9.0) | 7 | (5.2) | 3 | (4.2) |
| Age-bands |  |  |  |  |  |  |  |  |
| Under 12 | 3 | (2.5) | 3 | (2.8) | 0 | (0.0) | 0 | (0.0) |
| 12-17 | 111 | (9.0) | 86 | (7.0) | 15 | (5.7) | 5 | (6.5) |
| 18-29 | 250 | (17.9) | 178 | (13.6) | 33 | (7.2) | 13 | (6.0) |
| 30-39 | 498 | (13.8) | 357 | (10.8) | 69 | (5.4) | 16 | (2.5) |
| 40-49 | 581 | (21.4) | 462 | (18.4) | 52 | (5.1) | 37 | (6.0) |
| 50-59 | 731 | (18.6) | 539 | (15.4) | 68 | (4.1) | 72 | (6.2) |
| 60-69 | 893 | (14.8) | 600 | (11.2) | 126 | (4.3) | 98 | (4.7) |
| 70-79 | 910 | (10.2) | 576 | (7.2) | 141 | (3.5) | 83 | (3.1) |
| 80+ | 153 | (6.6) | 97 | (4.6) | 26 | (3.2) | 5 | (1.2) |
| Unknown | 4 | (16.0) | 1 | (4.5) | 1 | (4.2) | 0 | (0.0) |
| Ethnicity |  |  |  |  |  |  |  |  |
| White British, White Irish, or any other white background | 3,592 | (13.7) | 2,522 | (10.6) | 477 | (4.3) | 298 | (4.1) |
| Other | 254 | (14.4) | 187 | (11.5) | 32 | (5.7) | 15 | (4.8) |
| Unknown | 288 | (12.9) | 190 | (9.7) | 22 | (3.0) | 16 | (4.5) |
| BMI Category |  |  |  |  |  |  |  |  |
| Underweight | 107 | (16.0) | 70 | (11.4) | 19 | (6.9) | 14 | (8.7) |
| Normal | 1,304 | (14.6) | 907 | (11.2) | 193 | (4.8) | 101 | (3.8) |
| Overweight | 1,088 | (13.3) | 773 | (10.4) | 138 | (3.8) | 94 | (4.0) |
| Obese | 637 | (14.4) | 453 | (11.3) | 77 | (4.3) | 52 | (4.5) |
| Unknown | 998 | (12.4) | 696 | (9.6) | 104 | (3.7) | 68 | (4.2) |
| Immunocompromised |  |  |  |  |  |  |  |  |
| Yes | 514 | (14.8) | 346 | (11.0) | 75 | (5.2) | 43 | (4.6) |

<sup>2</sup> [TABLE FOOTNOTE] Proportion of patients who reported a MedDRA serious ADR in association with a dose of a COVID-19 vaccine, stratified by patient demographics and by the patient cohort definition. Example: Amongst the male patients who had reported a 1<sup>st</sup> dose and a 2<sup>nd</sup> dose vaccination, 3.5% (n=182) had reported a MedDRA serious ADR occurring after the 2<sup>nd</sup> dose vaccination.

|  | Patients reporting a MedDRA serious ADR (% of patient cohort reporting any vaccine dose) |  | Patients reporting a MedDRA serious ADR after the 1<sup>st</sup> dose (% of patient cohort reporting a 1<sup>st</sup> vaccine dose) |  | Patients reporting a MedDRA serious ADR after the 2<sup>nd</sup> dose (% of patient cohort reporting 1<sup>st</sup> and 2<sup>nd</sup> vaccine doses) |  | Patients reporting a MedDRA serious ADR after the 3<sup>rd</sup> dose (% of patient cohort reporting 1<sup>st</sup>, 2<sup>nd</sup>, and 3<sup>rd</sup> vaccine doses) |  |
| --- | --- | --- | --- | --- | --- | --- | --- | --- |
| No/Unknown | 3,620 | (13.5) | 2553 | (10.5) | 456 | (4.1) | 286 | (4.1) |

Amongst the patients who had reported at least their 1^st^ dose of vaccination, 14,587 patients had reported a total of 35,647 ADRs. Figure 2 presents the ADR reporting rates by dose for all ADRs reported and for MedDRA serious ADRs. Patient records for each dose are included if they reported all previous doses. In addition, one patient reported a single ADR following a 6^th^ dose. 1,093 patients reported an ADR with no start date therefore it could not be determined which dose the event followed.

**Figure 2.**
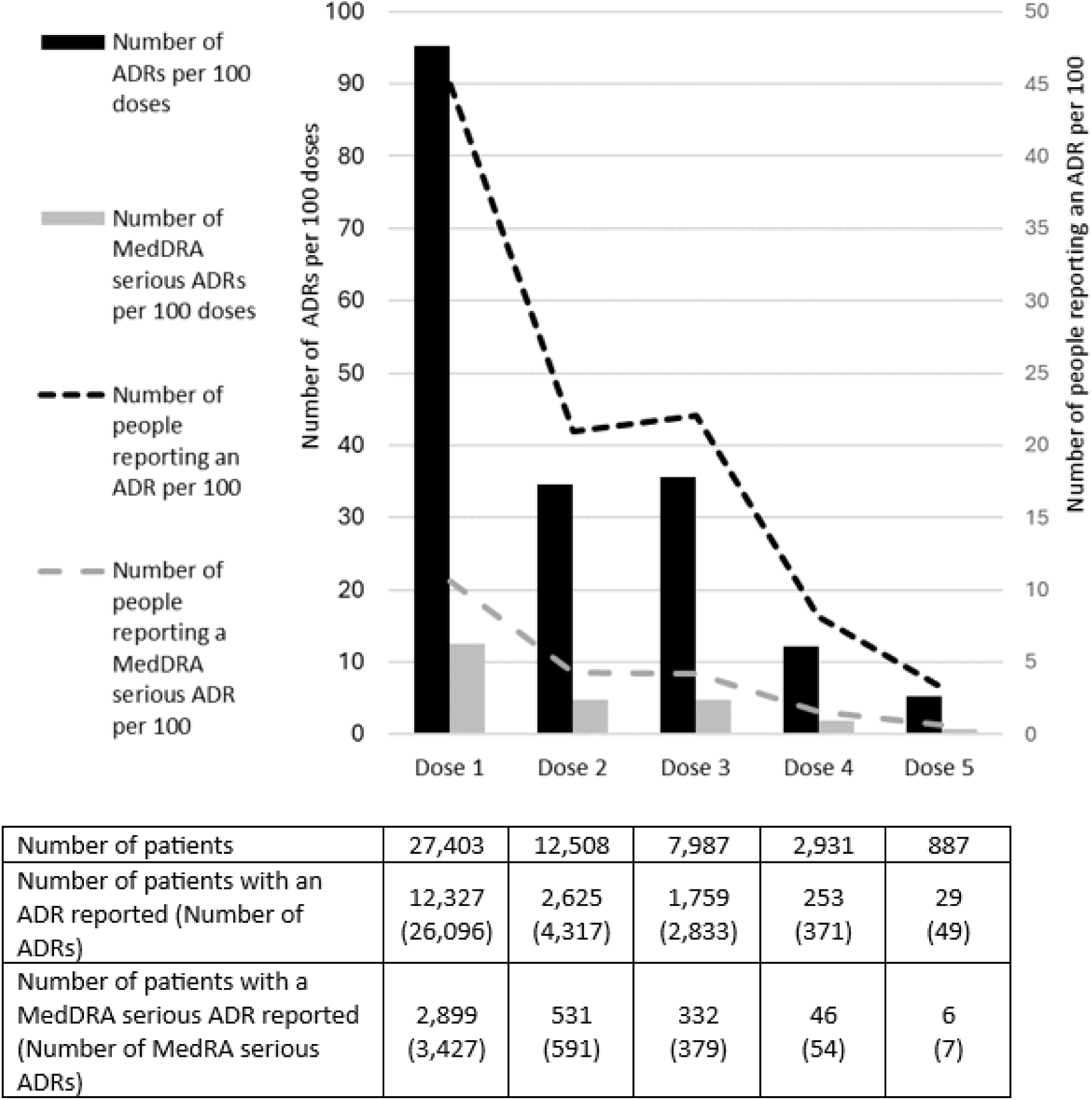
Average reporting rates of ADRs – Overall ADRs and MedDRA serious ADRs.

The 10 most prevalently reported ADRs for the first, second and third doses of a COVID-19 vaccination are presented for each vaccine brand in Table 7. Of these ADRs, only arthralgia and myalgia were considered medically serious events according to the MedDRA medical terminology system.

**Table 7:** Most commonly reported ADRs (MedDRA Preferred Term) by vaccine brand and dose (% of total ADRs reported for each dose/brand).

| **Vaccine Brand** | **Dose 1** | **Count (%)** | **Dose 2** | **Count (%)** | **Dose 3** | **Count (%)** |
| --- | --- | --- | --- | --- | --- | --- |
| **Pfizer** | Pain in extremity | 1,059 (18.6) | Fatigue | 230 (14.6) | Pain in extremity | 235 (13.5) |
|  | Fatigue | 654 (11.5) | Pain in extremity | 207 (13.1) | Headache | 223 (12.9) |
|  | Headache | 618 (10.8) | Headache | 166 (10.5) | Fatigue | 186 (10.7) |
|  | Myalgia | 271 (4.8) | Pyrexia | 74 (4.7) | Arthralgia | 72 (4.1) |
|  | Pain | 217(3.8) | Myalgia | 67 (4.3) | Myalgia | 72 (4.1) |
|  | Nausea | 174 (3.0) | Pain | 57 (3.6) | Pyrexia | 69 (4.0) |
|  | Arthralgia | 174 (3.0) | Nausea | 48 (3.0) | Chills | 66 (3.8) |
|  | Pyrexia | 156 (2.7) | Arthralgia | 47 (3.0) | Nausea | 56 (3.2) |
|  | Injection site pain | 155 (2.7) | Limb discomfort | 40 (2.5) | Pain | 54 (3.1) |
|  | Limb discomfort | 150 (2.6) | Dizziness | 33 (2.1) | Limb discomfort | 38 (2.2) |
|  | **TOTAL** | **5,705** | **TOTAL** | **1,576** | **TOTAL** | **1735** |
| **AstraZeneca** | Headache | 3,063 (15.7) | Fatigue | 359 (14.8) | Myalgia | 2 (16.7) |
|  | Fatigue | 2,205 (11.3) | Headache | 351 (14.4) | Pain in extremity | 2 (16.7) |
|  | Pyrexia | 1,470 (7.5) | Pain in extremity | 225 (9.2) | Fatigue | 1 (8.3) |
|  | Chills | 1,235 (6.3) | Myalgia | 123 (5.1) | Tenderness | 1 (8.3) |
|  | Pain in extremity | 1,200 (6.1) | Arthralgia | 91 (3.7) | Chills | 1 (8.3) |
|  | Myalgia | 992 (5.1) | Pyrexia | 90 (3.7) | Palpitations | 1 (8.3) |
|  | Nausea | 789 (4.0) | Chills | 74 (3.0) | Malaise | 1 (8.3) |
|  | Arthralgia | 760 (3.9) | Nausea | 71 (2.9) | Injection related reaction | 1 (8.3) |
|  | Pain | 485 (2.5) | Dizziness | 63 (2.6) | Body temp increased | 1 (8.3) |
|  | Influenza like illness | 410 (2.1) | Limb discomfort | 59 (2.4) | Arthralgia | 1 (8.3) |
|  | **TOTAL** | **19,559** | **TOTAL** | **2,433** | **TOTAL** | **12** |
| **Moderna** | Pain in extremity | 141 (22.6) | Pyrexia | 39 (13.9) | Pain in extremity | 155 (15.0) |
|  | Fatigue | 63 (10.1) | Fatigue | 37 (13.2) | Fatigue | 106 (10.3) |
|  | Headache | 57 (9.1) | Headache | 28 (10.0) | Headache | 106 (10.3) |
|  | Pyrexia | 33 (5.3) | Pain in extremity | 17 (6.0) | Pyrexia | 64 (6.2) |
|  | Nausea | 23 (3.7) | Chills | 17 (6.0) | Chills | 57 (5.5) |
|  | Myalgia | 21 (3.4) | Pain | 15 (5.3) | Nausea | 40 (3.9) |
|  | Pain | 20 (3.2) | Myalgia | 11 (3.9) | Myalgia | 40 (3.9) |
|  | Arthralgia | 16 (2.6) | Influenza | 8 (2.8) | Arthralgia | 37 (3.6) |
|  | Rash | 15 (2.4) | Arthralgia | 8 (2.8) | Influenza like illness | 25 (2.4) |
|  | Injection site pain | 15 (2.4) | Nausea | 6 (2.1) | Limb discomfort | 24 (2.2) |
|  | **TOTAL** | **623** | **TOTAL** | **281** | **TOTAL** | **1,034** |

Appendix D presents the number and proportion of events reported by MedDRA SOC level for each dose and by dose sequence for each vaccine brand. See ‘Supplementary Material S1’ for ADR count listings at MedDRA SOC, HLGT, HLT and PT levels.

Due to the approach taken to rollout of the national vaccination campaign to the UK population, analyses of ADR reporting across vaccine brands will potentially be confounded by age and other risk factors for severe outcomes following COVID-19 infection with the AstraZeneca vaccine being predominantly used in patients aged 40+ years and those with underlying health conditions and the Moderna vaccine being predominantly used as a 3^rd^ or subsequent dose again in an older population. Age-stratified analyses exploring the ADRs reported in those aged under 40 years and in those aged 40 years or older, presented in Appendix E, do not indicate any particular differences in the ADR reporting patterns between the age groups.

As part of the COVID-19 vaccination surveillance strategy, a list of specific medical events was identified of adverse events of special interest which were subject to enhanced monitoring. In the YCVM, there were 2 cases of Guillain-Barre Syndrome (GBS) reported, both were with a 1^st^ dose of the AstraZeneca vaccine. One case of GBS was medically confirmed, while the remaining case was unconfirmed. There were also 2 cases of transverse myelitis (including “myelitis” events) reported with a 1^st^ dose of the AstraZeneca vaccine. Information provided was too limited to medically confirm these cases. Finally, there were two cases of Bell’s Palsy reported. One occurred after receiving a 1^st^ dose of the AstraZeneca vaccine but was not medically confirmed. The other case was confirmed by a healthcare professional and occurred after a 2^nd^ dose vaccination using Pfizer vaccine.

### 3.4 COVID-19 Vaccination during Pregnancy and Breast-feeding

2,517 females registered on the YCVM platform reported they were pregnant at the time of a COVID-19 vaccination although only 2,416 provided details of that vaccination. 641 females reported breast-feeding at the time of a COVID-19 vaccination. Demographics of these cohorts and those who experienced an ADR after a vaccination are presented in Table 8. The demographics of females registered, including those with no vaccination reported are presented in Appendix F.

**Table 8:** Number of patients reporting an ADR reporting in vaccinated females, vaccinated pregnant females, and vaccinated breast-feeding females registered in the Yellow Card Vaccine Monitor, stratified by patient demographics (row %^3^).

|  | Female cohort |  |  | Pregnant females' cohort |  |  | Breast-feeding cohort |  |
| --- | --- | --- | --- | --- | --- | --- | --- | --- |
|  | Reported a vaccination<br>(n=17,463) | Reported an ADR after any vaccination<br>(n=10,145)<br>(% of vaccinated females cohort) |  | Reported a vaccination<br>(n=2,416) | Reported an ADR after any vaccination (n=995)<br>(% of vaccinated pregnant females cohort) |  | Reporting a vaccination<br>(n=641) | Reported an ADR after any vaccination<br>(n=343)<br>(% of vaccinated breastfeeding females cohort) |
| **Age-bands** |  |  |  |  |  |  |  |  |
| Under 12 | 61 | 27 (44.3) |  | 0 | 0 (0.0) |  | 0 | 0 (0) |
| 12-17 | 728 | 374 (51.4) |  | 2 | 0 (0.0) |  | 0 | 0 (0) |
| 18-29 | 1,074 | 636 (59.2) |  | 403 | 148 (36.7) |  | 76 | 43 (56.6) |
| 30-39 | 3,057 | 1,655 (54.1) |  | 1,857 | 781 (42.1) |  | 487 | 260 (53.4) |
| 40-49 | 1,738 | 1,302 (74.9) |  | 153 | 66 (43.1) |  | 77 | 40 (51.9) |
| 50-59 | 2,235 | 1,563 (69.9) |  | 1 | 0 (0.0) |  | 1 | 0 (0.0) |
| 60-69 | 3,214 | 2,051 (63.8) |  | 0 | 0 (0.0) |  | 0 | 0 (0.0) |

<sup>3</sup> [TABLE FOOTNOTE] Proportion of females who reported an ADR in association with a dose of a COVID-19 vaccine, stratified by patient demographics and by the patient cohort definition. Example: Amongst the pregnant females aged 30-39 years who had reported a vaccination, 42.1% (n=781) had reported an ADR occurring after vaccination.

|  | Female cohort |  |  | Pregnant females' cohort |  |  | Breast-feeding cohort |  |
| --- | --- | --- | --- | --- | --- | --- | --- | --- |
|  | Reported a vaccination<br>(n=17,463) | Reported an ADR after any vaccination<br>(n=10,145)<br>(% of vaccinated females cohort) |  | Reported a vaccination<br>(n=2,416) | Reported an ADR after any vaccination (n=995)<br>(% of vaccinated pregnant females cohort) |  | Reporting a vaccination<br>(n=641) | Reported an ADR after any vaccination<br>(n=343)<br>(% of vaccinated breastfeeding females cohort) |
| 70-79 | 4,267 | 2,110<br>(49.4) |  | 0 | 0<br>(0.0) |  | 0 | 0<br>(0.0) |
| 80+ | 1,077 | 422<br>(39.2) |  | 0 | 0<br>(0.0) |  | 0 | 0<br>(0.0) |
| Unknown | 12 | 5<br>(41.7) |  | 0 | 0<br>(0.0) |  | 0 | 0<br>(0.0) |
| **Ethnicity** |  |  |  |  |  |  |  |  |
| White British, White Irish, or any other white background | 15,049 | 8,815<br>(58.6) |  | 1,909 | 809<br>(42.4) |  | 538 | 295<br>(54.8) |
| Other | 1,101 | 642<br>(58.3) |  | 248 | 100<br>(40.3) |  | 65 | 36<br>(55.4) |
| Unknown | 1,313 | 688<br>(52.4) |  | 259 | 86<br>(33.2) |  | 38 | 12<br>(31.6) |
| **BMI Category** |  |  |  |  |  |  |  |  |
| Underweight | 433 | 278<br>(64.2) |  | 26 | 9<br>(34.6) |  | 5 | 3<br>(60.0) |
| Normal | 5,492 | 3,298<br>(60.1) |  | 690 | 325<br>(47.1) |  | 204 | 115<br>(56.4) |
| Overweight | 3,772 | 2,237<br>(59.3) |  | 489 | 209<br>(42.7) |  | 153 | 92<br>(60.1) |
| Obese | 2,405 | 1,461<br>(60.7) |  | 266 | 110<br>(41.4) |  | 77 | 40<br>(51.9) |
| Unknown | 5,361 | 2,871<br>(53.6) |  | 945 | 342<br>(36.2) |  | 202 | 93<br>(46.0) |
| **Immunocompromised** |  |  |  |  |  |  |  |  |
| Yes | 2,195 | 1,067<br>(58.2) |  | 111 | 50<br>(45.0) |  | 40 | 27<br>(67.5) |
| No | 18,673 | 9,078<br>(58.1) |  | 2,305 | 945<br>(41.0) |  | 601 | 316<br>(52.6) |

In pregnant females and breast-feeding females reporting a vaccination, the median age was 33 years (IQR 31-36) and 34 years (IQR 31-37) respectively. The cohort of vaccinated pregnant females contributed 903.5 patient years follow-up with a median follow-up of 72.5 days (IQR 5-244).

#### 3.4.1 Pregnant women

41.2% (n=995) of the vaccinated pregnant women reported a total of 1,934 ADRs in the YCVM. There were 1,255 ADRs reported in association with a 1^st^ dose, 476 with a 2^nd^ dose, 118 with a 3^rd^ dose and 3 with a 4^th^ dose. There were 82 ADRs with no onset date reported.

Due to the method in which pregnancy status data was collected and stored, it was not always possible to identify which dose or doses were received during pregnancy if they had reported more than one vaccination dose. As a result, all vaccination dose(s) and vaccine brand(s) reported in the YCVM by those who were pregnant are presented here. It does not confirm each reported dose was administered when the individual was pregnant. 1,654 (68.5%) had reported the stage of their pregnancy at the time of vaccination, with 310 reporting more than one stage of pregnancy in which they were vaccinated. The stage of pregnancy reported by these individuals and those who also reported at least one ADR are presented in Table 13. The proportion of patients experiencing an ADR after vaccination were similar across the different stages of vaccination during pregnancy.

**Table 13:** Stage of Pregnancy during exposure to vaccine amongst the cohort of pregnant females.

| Stage of Pregnancy | Number of vaccinated pregnant females (N=2,416) | Number of patients with an ADR reported (n=995), (% of vaccinated pregnant females) |
| --- | --- | --- |
| First trimester (1-12 weeks) | 281 | 132 (47.0) |
| Second trimester (13-28 weeks) | 663 | 305 (46.0) |
| Third trimester (29-40 weeks) | 400 | 174 (43.5) |
| More than one stage reported* | 310 | 144 (46.5) |
| Unknown | 762 | 240 (31.5) |

\* 310 patients reported exposure in more than one pregnancy stage and contributed the following counts: first trimester (110), second trimester (283) and third trimester (248).

Table 14 shows that the majority of doses reported by pregnant individuals were for the 1^st^ dose of COVID-19 vaccination and that the dominant vaccine brand was Pfizer BioNTech. Amongst those who had reported a vaccination with AstraZeneca, 116 had reported receiving only one COVID-19 vaccination in their YCVM data and 71 reported at least 1 ADR. Based on the guidelines that Pfizer or Moderna vaccines were the preferred vaccines for pregnant women it is more likely these AstraZeneca vaccinations were received prior to them becoming pregnant or when they were unaware, they were pregnant during their first trimester. The median number of doses reported in the YCVM by a patient who had indicated they were pregnant at the time of a vaccination was 1 dose (IQR: 1-2 doses). 499 patients (20.7%) reported receiving a third dose or booster dose as per guidance which recommends continued doses should be offered to pregnant women [14].

**Table 14:** Number of pregnant individuals registered by brand and dose and (number), [%], who reported an ADR.

|  | Pfizer | AstraZeneca | Moderna | Other or Unknown | Total |
| --- | --- | --- | --- | --- | --- |
| Dose 1 | 1729<br>(534), [30.9] | 241<br>(129), [53.5] | 183<br>(85), [46.4] | 3<br>(2), [66.7] | 2156<br>(750), [34.8] |
| Dose 2 | 901<br>(189), [21.0] | 155<br>(34), [21.9] | 104<br>(48), [46.2] | 5<br>(0), [0.0] | 1165<br>(271), [23.3] |
| Dose 3 | 321<br>(34), [10.6] | 1<br>(0), [0.0] | 169<br>(37), [21.9] | 8<br>(1), [12.5] | 499<br>(72), [14.4] |
| Dose 4 | 12<br>(1), [8.3] | 0<br>(0), [0.0] | 10<br>(1), [10.0] | 0<br>(0), [0.0] | 22<br>(2), [9.1] |
| Dose 5 | 0<br>(0), [0.0] | 0<br>(0), [0.0] | 3 (0), [0.0] | 0<br>(0), [0.0] | 3<br>(0), [0.0] |

The most commonly reported ADRs in this cohort for the 1^st^ dose, 2^nd^ dose, and 3^rd^ dose of COVID-19 vaccination are shown in Table 15. The events reported are common and recognised ADRs associated with vaccines. Pain in extremity, fatigue, headache, and pyrexia are the events that appear in the top 4 reported ADRs across the three doses.

**Table 15.** Most commonly reported ADRs by vaccine dose in the cohort of pregnant females.

| **Dose** | **Ranked Medical event reported (PT level)** | **Number of events (% of total ADR events per dose)** |  |
| --- | --- | --- | --- |
| *First dose* | Pain in extremity | 302 | (24.1) |
|  | Fatigue | 167 | (13.3) |
|  | Headache | 134 | (10.7) |
|  | Pyrexia | 74 | (5.9) |
|  | Myalgia | 50 | (4.0) |
|  | Nausea | 43 | (3.4) |
|  | Injection site pain | 36 | (2.9) |
|  | Chills | 35 | (2.8) |
|  | Pain | 35 | (2.8) |
|  | Limb discomfort | 34 | (2.7) |
|  | **TOTAL EVENTS** | **1255** |  |
| *Second dose* | Fatigue | 74 | (15.5) |
|  | Headache | 58 | (12.2) |
|  | Pain in extremity | 54 | (11.3) |
|  | Pyrexia | 33 | (6.9) |
|  | Nausea | 23 | (4.8) |
|  | Pain | 22 | (4.6) |
|  | Myalgia | 21 | (4.4) |
|  | Chills | 16 | (3.4) |
|  | Arthralgia | 12 | (2.5) |
|  | Dizziness | 11 | (2.3) |
|  | **TOTAL EVENTS** | **476** |  |
| *Third dose* | Fatigue | 17 | (14.4) |
|  | Pain in extremity | 16 | (13.6) |
|  | Headache | 14 | (11.9) |
|  | Pyrexia | 8 | (6.8) |
|  | Chills | 7 | (5.9) |
|  | Myalgia | 5 | (4.2) |
|  | Pain | 5 | (4.2) |
|  | Nausea | 4 | (3.4) |
|  | Diarrhoea | 3 | (2.5) |
|  | Dizziness | 3 | (2.5) |
|  | Influenza | 3 | (2.5) |
|  | Injection site pain | 3 | (2.5) |
|  | ***TOTAL EVENTS*** | ***118*** |  |

There were no reports of ADRs in the “Congenital, Familial and Genetic Disorders” SOC. The reports of ADRs belonging to the “Pregnancy, Puerperium and Perinatal Conditions” and “Reproductive System and Breast Disorders” SOCs are presented in Table 16.

**Table 16:** Individual MedDRA PT Terms reported in the “Pregnancy, Puerperium and Perinatal Conditions” and “Reproductive System and Breast Disorders” SOCs: Vaccine brand and dose stratification.

| **Number of PTs reported**<br>**(Frequency %)** | **Total** | **Vaccine brand** |  |  |  | **Dose** |  |  |  |
| --- | --- | --- | --- | --- | --- | --- | --- | --- | --- |
| **SOC & PT Term** |  | **Pfizer** | **AstraZeneca** | **Moderna** | **Other or**<br>**Unknown**<br>**brand** | **Dose 1** | **Dose 2** | **Dose 3** | **Missing**<br>**ADR date** |
| **Pregnancy, Puerperium**<br>**and Perinatal Conditions**<br>**SOC** | **28** | **17** | **7** | **1** | **1** | **17** | **8** | **1** | **2** |
| Abortion spontaneous | 19 | 13 | 5 | 1 | 0 | 11 | 8 | 0 | 0 |
| Foetal death | 1 | 0 | 1 | 0 | 0 | 1 | 0 | 0 | 0 |
| Foetal disorder | 1 | 0 | 0 | 0 | 1 | 1 | 0 | 0 | 0 |
| Foetal hypokinesia | 2 | 2 | 0 | 0 | 0 | 1 | 0 | 1 | 0 |
| Foetal macrosomia | 1 | 1 | 0 | 0 | 0 | 1 | 0 | 0 | 0 |
| Morning sickness | 1 | 0 | 0 | 0 | 0 | 0 | 0 | 0 | 1 |
| Placental infarction | 1 | 0 | 1 | 0 | 0 | 1 | 0 | 0 | 0 |
| Pregnancy | 1 | 0 | 0 | 0 | 0 | 0 | 0 | 0 | 1 |
| Uterine contractions<br>during pregnancy | 1 | 1 | 0 | 0 | 0 | 1 | 0 | 0 | 0 |
| **Reproductive System**<br>**and Breast Disorders**<br>**SOC** | **12** | **7** | **2** | **0** | **0** | **4** | **3** | **2** | **3** |
| Menstrual disorder | 1 | 1 | 0 | 0 | 0 | 0 | 0 | 1 | 0 |
| Menstruation delayed | 2 | 0 | 1 | 0 | 0 | 0 | 1 | 0 | 1 |
| Oligomenorrhoea | 1 | 0 | 0 | 0 | 0 | 0 | 0 | 0 | 1 |
| Heavy menstrual<br>bleeding | 2 | 1 | 0 | 0 | 0 | 0 | 0 | 1 | 1 |
| Vaginal haemorrhage | 6 | 5 | 1 | 0 | 0 | 4 | 2 | 0 | 0 |
| **TOTAL ADR EVENTS (All**<br>**SOCs)** | **1934** | **1161** | **377** | **309** | **5** | **1255** | **476** | **118** | **82** |

There were 26 individuals who reported an ADR from the “Pregnancy, Puerperium and Perinatal Conditions” SOC with a total of 28 ADRs reported. Of the total 28 events reported in the “Pregnancy, Puerperium and perinatal condition” SOC category, 13 events (35.7%) were associated with 13 pregnant females who reported receiving a vaccination during their first trimester.

There were 19 reports of spontaneous abortion. The majority of these reports were reported in association with a 1^st^ dose of COVID-19 vaccination (n=11), 7 with the Pfizer BioNTech vaccine, 3 with AstraZeneca and 1 with Moderna. The one case of foetal death was reported alongside a spontaneous abortion, and this was reported in association with a 1^st^ dose of AstraZeneca. In 8 (72.7%) of the reports occurring after a dose 1 vaccination, individuals had reported vaccination during the 1^st^ trimester, whilst 3 reports (27.3%) did not report the trimester in which they were vaccinated. The median time from the last reported vaccination to the reported date of spontaneous abortions occurring after receiving a 1^st^ dose vaccination (n=11) was 27 days (IQR 18-47 days), and in those occurring after a 2^nd^ dose vaccination (n=8) was 33.5 days (IQR 24-53).

In the Reproductive System and Breast Disorders SOC, there were 12 events associated with 12 females who had received a vaccination during their pregnancy. There were 4 events (33.3%) associated with 4 pregnant females who reported receiving a vaccination during their first trimester, 3 reporting second trimester, 2 with third trimester and 2 with unknown stage of pregnancy. There was 1 event reported by an individual with multiple stages of pregnancy reported in their YCVM data.

At the time of the individual’s last submission, there were 7 events resolved, 2 recovering, 9 ongoing, and 10 unknowns, in the “Pregnancy, Puerperium and Perinatal Conditions”. In the “Reproductive System and Breast Disorders” SOC, there were 7 resolved, 2 recovering, and 3 ongoing. The full list of ADRs reported in this cohort of pregnant individuals are tabulated in ‘Supplementary Material S2’.

For those who had reported an estimated pregnancy due date, a follow-up was sent 10 weeks after the due date requesting additional information on the outcome of the pregnancy such full-term births or pre-term births. This information was recorded in the free-text narrative field of the YCVM report. There had been no reports of pre-term births reported as an ADR, however a search of the free-text field identified 4 females amongst the vaccinated pregnant females who had reported a premature birth.

#### 3.4.2 Breast-feeding Individuals

Of the 694 females reported as breastfeeding at the time of a vaccination, 641 had entered details of at least one COVID-19 vaccination; 53.5% (n=343) of these vaccinated females who were breastfeeding reported a total of 900 ADRs in the YCVM. There were 564 ADRs reported in association with a 1^st^ dose, 199 with a 2^nd^ dose, 81 with a 3^rd^ dose and 56 with no onset date. Approximately 64.8% (n=450) of breast-feeding females and 61.5% (n=211) of those who went on to report an ADR, had reported being vaccinated during their pregnancy.

The information collected on breastfeeding in the YCVM platform was derived from a binary yes or no question. As a result, where a patient recorded more than one dose, it is not possible to determine at which dose the breastfeeding occurred. The median number of doses reported in the YCVM by a patient who had indicated they were breastfeeding at the time of a vaccination was 2 doses (IQR: 1-3 doses). At least 235 patients had reported receiving a third dose or booster dose.

Table 17 presents the number of doses received by patients by sequence and by brand, during the whole follow-up time in the YCVM. The majority of reported doses were for the 1^st^ dose vaccination, with the most frequently reported vaccine brand being the Pfizer vaccine (n=391).

**Table 17:**
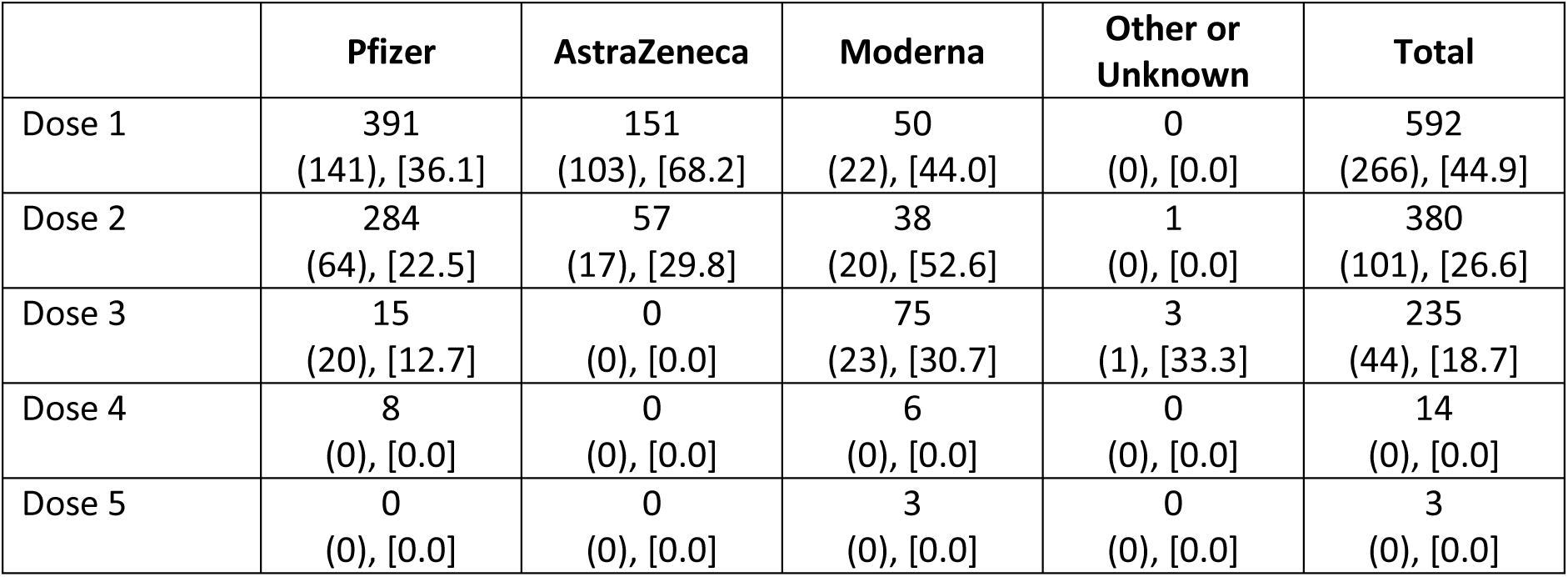
Number of breastfeeding individuals registered by brand and dose and (number), [%], who reported an ADR.

There were no reports of events relating to lactation disorders reported in this cohort. There was 1 report of mastitis occurring after a 3^rd^ dose vaccination with the Moderna vaccine. In the “Pregnancy, Puerperium and Perinatal Conditions” SOC there was one ADR reported where placental infarction occurred after a 1^st^ COVID-19 vaccination with the AstraZeneca vaccine. In the “Reproductive System and Breast Disorders” SOC, there were in total 6 events reported with 1 report of each of the following events: irregular menstruation, hypomenorrhea, delayed menstruation, oligomenorrhoea, heavy menstrual bleeding and vaginal haemorrhage. There were no reports of ADRs in the “Congenital, Familial and Genetic Disorders” SOC.

It is possible a female patient may not indicate they were breast-feeding using the optional breast-feeding question. Amongst the ADRs reported by any female patient, there were no reports of lactation related ADRs. However, there were 25 reports of breast disorders – 4 reports of breast mass, 1 report of nipple enlargement, 1 report of breast discharge, 15 reports of breast pain, 1 report of breast sweeling, 1 report of breast tenderness and 2 reports of nipple pain.

## 4 Case studies

### 4.1 Menstrual disorders

The potential safety signal was first identified in spontaneous Yellow Card data through routine signal detection activities in January 2021. Of the 20,868 females registered in the Yellow Card Vaccine Monitor cohort, 219 (1.05%) reported 258 ADRs relating to menstrual disorder. There were 152 related events reported following a 1^st^ dose, 45 events following a 2^nd^ dose and 23 events following a 3^rd^ dose. There were 38 events which did not have sufficient information to determine the preceding vaccination dose and brand.

The demographics of the patients reporting menstrual disorder ADRs are presented in Table 21, including vaccine brand-specific patient demographics.

**Table 21:** Patient characteristics of female patients who reported menstrual disorder ADRs.

|  | **Total Patients** | **Pfizer** | **AstraZeneca** | **Moderna** | **Other/unknown brand** |
| --- | --- | --- | --- | --- | --- |
| Number of female patients | **219** | **102** | **72** | **17** | **1** |
| **Age (years)** |  |  |  |  |  |
| <18 years | 12 | 11 | 0 | 0 | 1 |
| 18-49 years | 196 | 88 | 67 | 17 | 0 |
| 50+ years | 11 | 3 | 5 | 0 | 0 |
| **Ethnicity** |  |  |  |  |  |
| White | 179 | 79 | 63 | 14 | 1 |
| Non-white | 23 | 15 | 4 | 2 | 0 |
| Not stated | 17 | 8 | 5 | 1 | 0 |

The menstrual disorder-related PT events reported are presented in Table 22. More than half of the three most commonly reported events, heavy menstrual bleeding, irregular or delayed menstruation, and dysmenorrhoea, occurred after a 1^st^ dose vaccination.

**Table 22:** Individual MedDRA PTs reported relating to Menstrual disorders.

| **Menstrual disorder related PT Term** | **Number of PTs reported (Frequency %)** |  |
| --- | --- | --- |
| Amenorrhoea | 1 | (0.4) |
| Dysmenorrhoea | 29 | (11.2) |
| Heavy menstrual bleeding | 61 | (23.6) |
| Hypomenorrhoea | 9 | (3.5) |
| Intermenstrual bleeding | 13 | (5.0) |
| Menstrual disorder | 22 | (8.5) |
| Menstruation delayed | 47 | (18.2) |
| Menstruation irregular | 47 | (18.2) |
| Oligomenorrhoea | 3 | (1.2) |
| Polymenorrhoea | 9 | (3.5) |
| Postmenopausal haemorrhage | 1 | (0.4) |
| Premenstrual pain | 2 | (0.8) |
| Retrograde menstruation | 1 | (0.4) |
| Vaginal haemorrhage | 13 | (5.0) |
| **TOTAL ADR EVENTS** | **258** |  |

At the time of the last submission submitted prior to or on 31st December 2022, 105 events were reported as resolved with or without sequalae, 40 as resolving, and 91 as ongoing. There were 22 events with no outcome reported.

To explore time trends in the reporting of menstrual disorder related adverse events, the report submission dates of these events are shown in Figure 8. The number of reports grew with increasing social media attention and the publication of several news stories on the internet (A in Figure 8) with subsequent peaks coinciding with publication of a key major news article (B in Figure 8) and a mass booster vaccination campaign in where a large cohort of females aged 18-49 years of age were made eligible to receive their booster dose (C in Figure 8).

**Figure 8:**
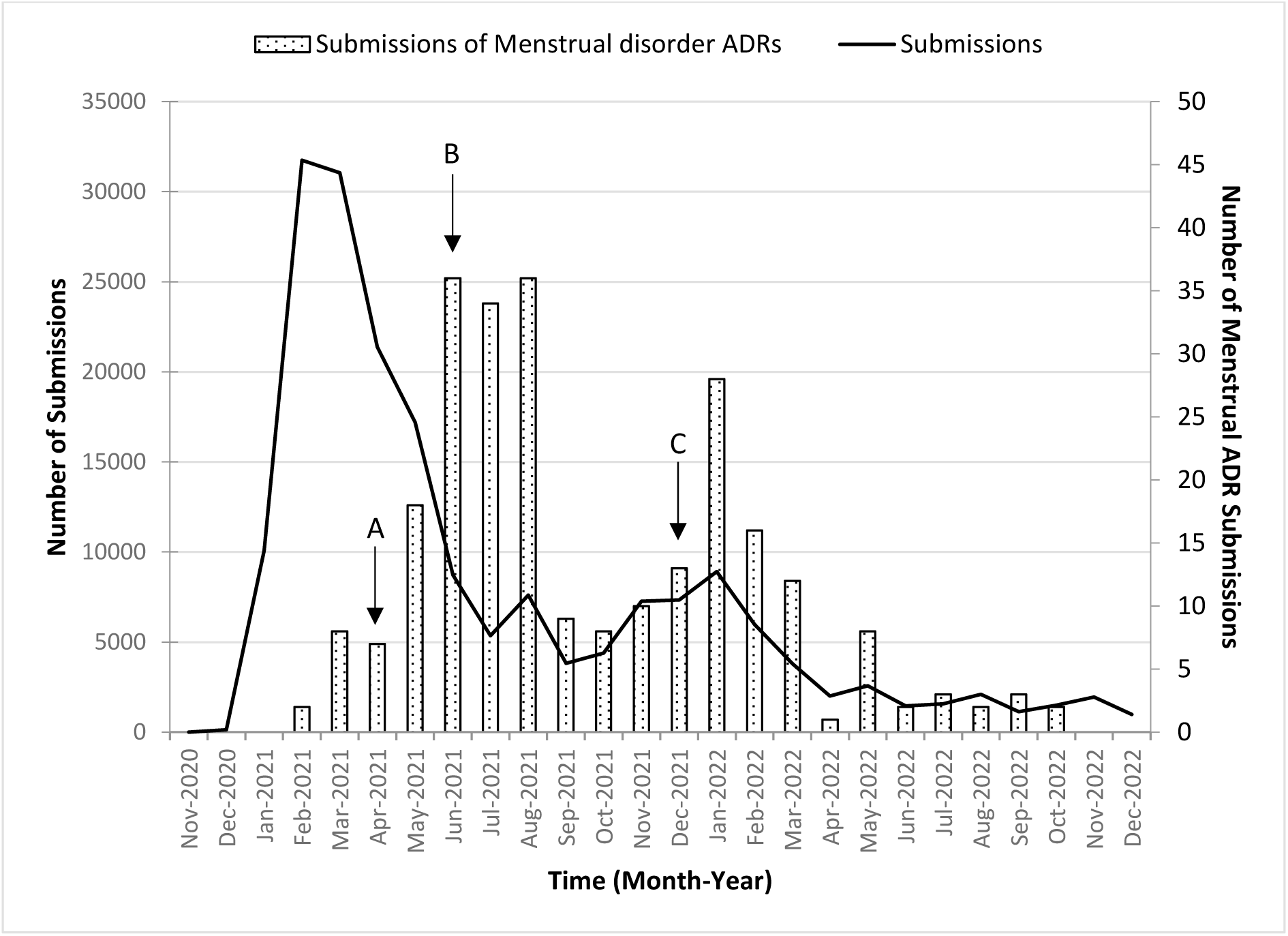
Number of Menstrual ADRs submissions in the Yellow Card Vaccine Monitor up to 31 December 2022. *(Arrows indicate key events: A^4^ for the first wave of published news stories, B^5^ for the key news article by Sunday Times, and C for rapid rollout of booster to under 50’s.)* ^4^ Wave A included articles published by the Huffington Post website (“How The Covid-19 Vaccine May Affect Your Period (And What To Do) | HuffPost UK Life”), the Euronews (“Experts cool claims about impact of vaccines on menstrual cycles | Euronews”), the BBC News (“Covid vaccine: Period changes could be a short-term side effect - BBC News”) and The Telegraph (“Post-menopausal women report periods coming back after having Covid vaccine (telegraph.co.uk)”). ^5^ Article published by the Sunday Times on 20 June 2021, titled “4,000 women report period problems after Covid jab”. Weblink: “https://www.thetimes.com/uk/healthcare/article/4-000-women-report-period-problems-after-jab-3sdgwgx8v”.

### 4.2 Tinnitus

Amongst the patients who had reported receiving a 1^st^ dose in the YCVM, 1.9% (n=73) reported 83 tinnitus or related PT events. 54 events (from 50 patients) were reported in association with the 1^st^ dose, 16 (from 13 patients) with the 2^nd^ dose, 6 (from 6 patients) with the 3^rd^ dose. There were 7 events (from 7 patients) which did not have sufficient information to determine which dose the event followed.

**Table 26:** Patient characteristics of patients who reported hearing loss and tinnitus ADRs:

|  | Patients (n=73) | Pfizer (n=21) | AstraZeneca (n=43) | Moderna (n=4) |
| --- | --- | --- | --- | --- |
| **Sex** |  |  |  |  |
| Male | 26 | 7 | 14 | 1 |
| Female | 46 | 14 | 28 | 3 |
| Unknown | 1 | 0 | 1 | 0 |
| **Age (years)** |  |  |  |  |
| <65 years | 58 | 17 | 32 | 4 |
| 65+ years | 15 | 4 | 11 | 0 |
| **Ethnicity** |  |  |  |  |
| White | 61 | 18 | 37 | 2 |
| Non-white | 6 | 2 | 3 | 1 |
| Not stated | 6 | 1 | 3 | 1 |

The reports of tinnitus were predominantly reported in association with an AstraZeneca vaccine, with “Tinnitus” as the most commonly reported PT. Table 27 presents PT Terms reported, the dose onset, vaccine brand and the median time to onset of the ADR after receiving the vaccination.

**Table 27:** Individual MedDRA PTs reported and onset time information by vaccination brand.

|  | TOTAL | Pfizer | AstraZeneca | Moderna |
| --- | --- | --- | --- | --- |
| **PT Term reported** |  |  |  |  |
| DEAFNESS | 4 | 3 | 0 | 1 |
| DEAFNESS NEUROSENSORY | 1 | 1 | 0 | 0 |
| DEAFNESS UNILATERAL | 2 | 0 | 2 | 0 |
| HYPERACUSIS | 1 | 0 | 1 | 0 |
| HYPOACUSIS | 3 | 1 | 2 | 0 |
| SUDDEN HEARING LOSS | 1 | 0 | 1 | 0 |
| TINNITUS | 71 | 17 | 44 | 3 |
| **Total Tinnitus and related terms:** | **83** | **22** | **50** | **4** |
| **ADR timing** |  |  |  |  |
| ADR after 1<sup>st</sup> dose | 54 | 13 | 40 | 1 |
| ADR after 2<sup>nd</sup> dose | 16 | 5 | 10 | 1 |
| ADR after 3<sup>rd</sup> dose | 6 | 4 | 0 | 2 |
| Timing unknown | 7 | - | - | - |
| **Time to onset\* (median days, IQR)** | 2 (1-7.5) | 4.5 (0-23) | 2 (1-5) | 0.5 (0-3.5) |
| 1<sup>st</sup> doses | 1 (0-10) | 1 (0-23) | 1 (0.5-5) | 1 (1-1) |
| 2<sup>nd</sup> doses | 4 (1-5) | 4 (0-4) | 4 (1-5) | 0 (0-0) |
| 3<sup>rd</sup> doses | 5 (1-7) | 6 (3-79.5) | - | 3 (0-6) |

\* Note: In relation to most recent dose prior to ADR onset.

Approximately two thirds of the reported tinnitus related events were ongoing at the most recent submission (n=51) whilst less than a third were reported as resolved (n=25), and the remaining events were resolving (n=6) or unknown (n=1).

## 5 Discussion

The scale of the COVID-19 pandemic necessitated the implementation of a safety surveillance strategy utilising multiple data sources to provide the timely and robust data needed for regulatory decision making. While spontaneous reporting of adverse events through the UK Yellow Card (YC) scheme has been these a central component of the MHRA’s approach to medicines and vaccines vigilance for the last 60 years, these data represent the first time that active surveillance has been directly conducted by the MHRA and the first time the technology and infrastructure used have been available. Active surveillance was also conducted for COVID-19 vaccines by other organisations, including by public health bodies and regulators [15,16]. However, these were not particularly designed to be integrated with spontaneous reporting directly into a regulatory adverse drug reaction database.

### 5.1 Recruitment and engagement

Up to 31^st^ December 2022, over 1 million individuals were invited to register with the YCVM. Female patients and those aged over 50 years were more likely to register and provide vaccination data although the characteristics of patients included will have been influenced by differences in vaccine uptake, timing of recruitment drives, and an individual’s propensity to report. Invitations to register could be targeted to new at-risk groups through the system established for inviting people in England to book their vaccination, while pregnant women were targeted through printed materials given to them at their vaccine appointment. However, as with any voluntary reporting scheme it is challenging to ensure a representative population, and we are unable to access data on those who were invited to register but did not. One of the aims of YCVM was to target enrolment of ethnic minorities underrepresented in clinical trials. In the YCVM, approximately 86% of registered patients belonged to the “White British, White Irish or Other White” ethnicity group. This proportion is larger than the estimate across the UK population [17], suggesting that representation of the other ethnic groups was limited despite targeted recruitment. Approximately 35,000 invites were sent during one call for registrations in May 2021, with 80% of these sent to non-white British/non-white Irish individuals. These targeted invites to under-represented ethnic groups had little to no impact on the numbers registering from this group, reflecting the lower levels of engagement with passive reporting systems seen in non-white populations. The need to increase the diversity of reporting on adverse reactions is well recognised even within spontaneous reporting systems and broader efforts to increase the visibility of, and engagement with, vigilance systems in different communities is needed.

Due to the system implemented during the pandemic period to directly invite people to book and receive a vaccination there was a particular opportunity for prospective recruitment, however nearly half of patients registered had a registration date after their earliest vaccination date. This limits interpretation of the totality of the data with regards to acute reactogenicity reports as the propensity to register after vaccination may be influenced by the occurrence of early reactions. Consideration also needs to be given as to how drop out from reporting of second and subsequent doses can be reduced.

### 5.2 Adverse drug reaction reporting and contribution to evidence base

Generally, the types of ADRs reported through the YCVM aligned with those reported through the Yellow Card passive reporting system [18]. Patterns of reporting also reflected those seen in pre-authorisation clinical trials with higher reporting of local and systemic events following administration of the Pfizer mRNA vaccine in younger patients and after a second dose [19] and in younger patients after a first dose following administration of the AstraZeneca adenoviral vectored vaccine [20]. However, the interpretation of these data, and those data for the Moderna mRNA vaccine, are complicated by the rollout of the vaccines in the UK with correlation between the vaccine brand used and the patient age and dose number.

Data from YCVM has supported signal assessment alongside routine signal detection activities using other data sources. It has not identified any additional safety signals in isolation, which is to be expected given the size of the cohort involved and the design. Information from ADR reports submitted via the YCVM has contributed to the evaluation of potential safety signals with a COVID-19 vaccine alongside that accrued through the other three pillars of the COVID-19 vaccine safety surveillance strategy.

Vaccination in pregnancy was not recommended in England until April 2021. Inclusion of YCVM data has contributed to the safety review for COVID-19 vaccines in pregnancy and breastfeeding, which concluded the YC data did not raise any safety concerns with COVID-19 vaccine exposure during pregnancy. However, as with passive spontaneous reporting, the completeness of reporting is unknown. The rate of early pregnancy loss is uncertain but is estimated to occur in around 15% of recognised pregnancies with a greater risk earlier in pregnancy and in older mothers; with certain long term health conditions, weight, and lifestyle factors also being risk factors [21]. The absolute rate of pregnancy loss reported into the YCVM was lower than the expected rate. However, these figures cannot be directly compared as the underlying risk at the point of vaccination for these pregnant women is unknown. Longer follow-up would be required from pregnant and breastfeeding patients to collect data on ADRs relating to their infants and developmental outcomes. Subsequent epidemiological studies demonstrate the safety of these vaccines in pregnancy, showing no evidence of an increased risk of adverse pregnancy outcomes [22,23]. These studies enable comparative analyses across substantially larger cohorts of vaccinated and unvaccinated pregnant women and allow for the adjustment of key confounding variables including gestational and maternal age meaning they present more robust evidence of vaccine safety in pregnancy.

In the menstrual disorders case study, YCVM data was used to support and facilitate discussions on the emerging safety signal. Whilst overall assessment at the time concluded there was no evidence of an increased risk of menstrual disorders, it was recognised the reporting of menstrual disorder events in the YCVM and the YC Scheme was heavily influenced by media interest. Data from the YCVM cannot be used to assess if there is an association between vaccination and menstrual events but improvements to follow up could support better assessment of patient experiences and outcomes. Heavy menstrual bleeding was included in the product information for the Pfizer and Moderna vaccines following review [24].

For tinnitus, the potential safety signal first arose from the YC data during routine signal detection activities. Data from YCVM was included in the MHRA reviews conducted on the available evidence, alongside spontaneous YC data, clinical trial data and published literature. The number of patients reporting a tinnitus-related ADR in the YCVM was small, which emphasised the use of YCVM data in a supportive role in signal assessment and not in isolation as well as providing reassurance on the potential incidence rate. The average time to onset for the reports related to the AstraZeneca vaccine was slightly shorter compared to the reports with Pfizer vaccine. Tinnitus was added as an adverse reaction to the product information for the AstraZeneca COVID-19 vaccine [25].

### 5.3 Technology and infrastructure

The platform benefits from an automated follow-up reminder system which aims to capture up-to-date information on a patient’s characteristics, vaccination records and any ADR(s) reported, and extend the active follow-up time for data collection. This supported a 70% successful follow up rate in June 2021, which only dropped to 62% in March 2022, which can be compared to less than a 10% success rate for follow up of spontaneous cases using traditional routes. It allows patients to have control of the information they share and how frequently they update their information. These include acute but short-lived conditions that may not be well captured by electronic health records because medical assistance was not sought from a clinician. Although follow up was substantially increased compared to that achieved for spontaneously reported data, improvement could be made to better capture final patient outcomes.

The YCVM is currently not automatically linked to other sources of health data, therefore it is not possible to medically confirm self-reported ADRs reported through the YCVM. This was of particular importance when adverse events of special interest such as GBS and Bell’s palsy were reported.

The data architecture of the YCVM was driven by the processing and integration of data in YCVM into the MHRA’s ADR reporting database. This has an impact on the data model and consequently the structure of the YCVM dataset used in conducting data analysis outside of the MHRA’s standard signal detection procedures. In addition, a number of variables are captured once in each submission report, which makes relationship assertions between variables unclear.

The capture of data from pregnant individuals could be improved further to enable use of the YCVM as an active surveillance tool for prospective monitoring of pregnant women. For example, at present, it is not possible differentiate between due dates and the date of the pregnancy outcome and is not possible to indicate if an individual experienced multiple pregnancy episodes during their time in the YCVM.

### 5.4 Future development and use

YCVM demonstrated capability to collect longitudinal data from vaccinees through a user-friendly interface and engagement model. Data were collected in standard formats for medicines regulators, meaning that any reports of side effects could be rapidly evaluated alongside spontaneous reports received via the Yellow Card Scheme. However, due to the nature of collecting reports in this format analysis of the totality of data (specifically where there were no side effects, or where there was a change between follow up cycles) was challenging.

Future developments will focus on improving data formats for this type of analysis and increasing levels of validation to reduce the level of data cleansing required. Updates to improve data quality and data collection and storage methods could facilitate the creation of a pregnancy outcome data set for the purpose of close monitoring of pregnancy outcomes and exposure to vaccine to support case assessment; such approaches, including smart forms and conditional questions based on the information provided by the reporter could deliver data akin to registry data, but at significantly lower cost.

Key to the successful collection of data through YCVM was that information was included within the system for vaccine invites. This was a unique opportunity during the pandemic, but even with this capability a substantial proportion registered post vaccination. For a genuine unbiased dataset stricter control would be required, either through exclusion criteria applied to the overall analysis or through a mandate for registration prior to receiving the product. This latter option would require careful coordination and management across the healthcare system.

More realistic may be an active follow-up approach where individuals who report spontaneously to the Yellow Card Scheme are subsequently consented for ongoing engagement to enable better understanding of the long-term nature of any events experienced, and their recovery. An alternative or additional approach would be linkage of Yellow Card data to that within the Electronic Healthcare record for assessment of specific signals.

The technology behind the Yellow Card Scheme now has the capability for all such types of engagement with reporter’s incidents related to medicines, vaccines, and medical devices, substantially increasing the MHRA’s vigilance options, depending on the specific products under consideration.

In addition to validation of the side effect profiles for vaccines in situations such as a pandemic, the MHRA considers that active surveillance may be particularly valuable in situations where there is extremely low or niche usage of a product or where long follow up is required. One example may be for products such as for Advanced Therapy Medicinal Products (ATMPs), where review of high quality data for small numbers of patients will be critical. Such situations are likely to have highly engaged patient populations and are less likely to require the volume of invites that were required to obtain a representative population in YCVM. Although ensuring representativeness of the enrolled population, for example across ethnicities, would be challenging, further efforts to increase engagement with vigilance systems through community leaders and through patient charities could be helpful.

## 6 Conclusions

Overall, adverse event reporting into the YCVM provided a complimentary data source to the other strands of the MHRA COVID-19 vaccine safety surveillance strategy, which utilised both spontaneous reports and large electronic healthcare record databases. However, the investment in infrastructure has advanced UK regulatory pharmacovigilance systems which can support targeted prospective monitoring of products in real world use in the future capturing data directly from patients on their experiences.

## Supporting information

Supplementary Material S1. ADR listing

Supplementary Material S2. ADR listing in Pregnant and Breastfeeding Cohort

## Data Availability

The Agency provides information to the public in line with requirements of the Freedom of Information Act. In addition, organisations and individuals can apply for Yellow Card data for research purposes. Applications include provision of a protocol and will be considered by the UK Commission of Human Medicine's (CHM) Pharmacovigilance Expert Advisory Group.

## 8 Appendices

## Appendix A. MedDRA Event Search Terms Lists for Case Studies

**Table A.1.** MedDRA PT terms considered for Menstrual Disorders Case Study. All events within the HLT Term “"MENSTRUATION AND UTERINE BLEEDING NEC","MENSTRUATION WITH DECREASED BLEEDING","MENSTRUATION WITH INCREASED BLEEDING","VULVOVAGINAL DISORDERS NEC" were included in the search criteria.

| PT Term |
| --- |
| POSTMENOPAUSAL HAEMORRHAGE |
| UTERINE HAEMORRHAGE |
| VAGINAL HAEMORRHAGE |
| ABNORMAL UTERINE BLEEDING |
| AMENORRHOEA |
| DYSMENORRHOEA |
| HEAVY MENSTRUAL BLEEDING |
| HYPOMENORRHOEA |
| INTERMENSTRUAL BLEEDING |
| MENOMETRORRHAGIA |
| MENSTRUAL DISCOMFORT |
| MENSTRUAL DISORDER |
| MENSTRUATION DELAYED |
| MENSTRUATION IRREGULAR |
| OLIGOMENORRHOEA |
| POLYMENORRHOEA |
| PREMENSTRUAL DYSPHORIC DISORDER |
| PREMENSTRUAL HEADACHE |
| PREMENSTRUAL PAIN |
| PREMENSTRUAL SYNDROME |
| WITHDRAWAL BLEED |
| RETROGRADE MENSTRUATION |

**Table A.2.**
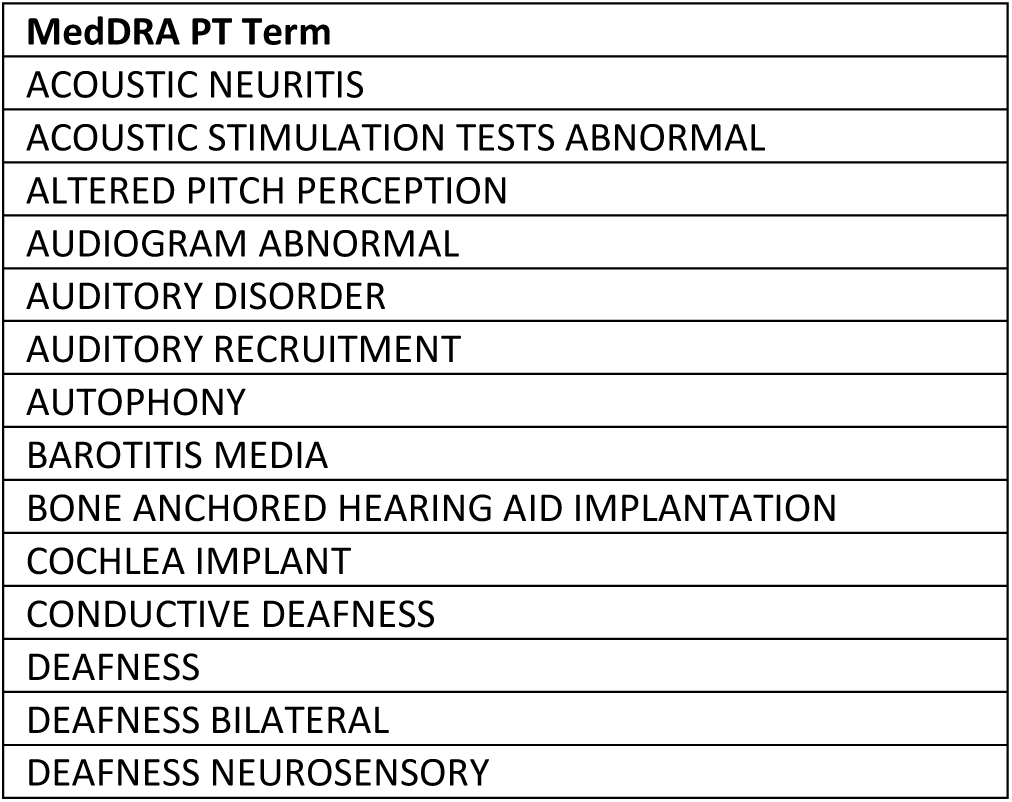
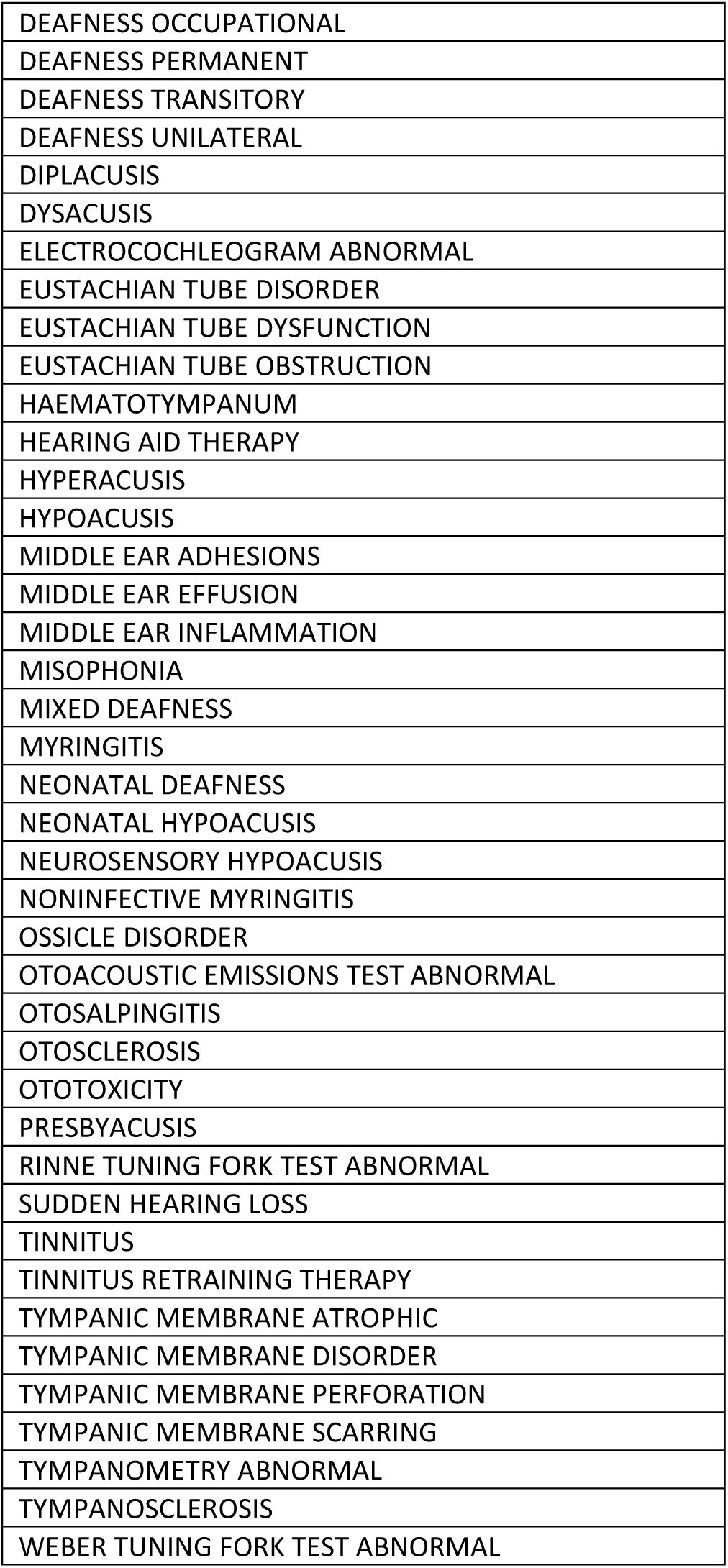
MedDRA PT terms considered for Tinnitus Case Study.

| MedDRA PT Term |
| --- |
| ACOUSTIC NEURITIS |
| ACOUSTIC STIMULATION TESTS ABNORMAL |
| ALTERED PITCH PERCEPTION |
| AUDIOGRAM ABNORMAL |
| AUDITORY DISORDER |
| AUDITORY RECRUITMENT |
| AUTOPHONY |
| BAROTITIS MEDIA |
| BONE ANCHORED HEARING AID IMPLANTATION |
| COCHLEA IMPLANT |
| CONDUCTIVE DEAFNESS |
| DEAFNESS |
| DEAFNESS BILATERAL |
| DEAFNESS NEUROSENSORY |
| DEAFNESS OCCUPATIONAL |
| DEAFNESS PERMANENT |
| DEAFNESS TRANSITORY |
| DEAFNESS UNILATERAL |
| DIPLACUSIS |
| DYSACUSIS |
| ELECTROCOCHLEOGRAM ABNORMAL |
| EUSTACHIAN TUBE DISORDER |
| EUSTACHIAN TUBE DYSFUNCTION |
| EUSTACHIAN TUBE OBSTRUCTION |
| HAEMATOTYMPANUM |
| HEARING AID THERAPY |
| HYPERACUSIS |
| HYPOACUSIS |
| MIDDLE EAR ADHESIONS |
| MIDDLE EAR EFFUSION |
| MIDDLE EAR INFLAMMATION |
| MISOPHONIA |
| MIXED DEAFNESS |
| MYRINGITIS |
| NEONATAL DEAFNESS |
| NEONATAL HYPOACUSIS |
| NEUROSENSORY HYPOACUSIS |
| NONINFECTIVE MYRINGITIS |
| OSSICLE DISORDER |
| OTOACOUSTIC EMISSIONS TEST ABNORMAL |
| OTOSALPINGITIS |
| OTOSCLEROSIS |
| OTOTOXICITY |
| PRESBYACUSIS |
| RINNE TUNING FORK TEST ABNORMAL |
| SUDDEN HEARING LOSS |
| TINNITUS |
| TINNITUS RETRAINING THERAPY |
| TYMPANIC MEMBRANE ATROPHIC |
| TYMPANIC MEMBRANE DISORDER |
| TYMPANIC MEMBRANE PERFORATION |
| TYMPANIC MEMBRANE SCARRING |
| TYMPANOMETRY ABNORMAL |
| TYMPANOSCLEROSIS |
| WEBER TUNING FORK TEST ABNORMAL |

## Appendix B. Follow-up Time contributed from Vaccinated Cohort

**Table B.1.** Follow-up time by patient sex.

| Follow-up time (days) | Male Patients (n=12,424) |  | Female Patients (n=17,463) |  | Unknown sex (n=394) |  |
| --- | --- | --- | --- | --- | --- | --- |
|  | Total | Median (IQR) | Total | Median (IQR) | Total | Median (IQR) |
| Number of registered patients | 3,040,806 | 248.5 (16-383) | 3,701,682 | 179 (10-363) | 66,217 | 73.5 (1-333) |
| **Age-bands** |  |  |  |  |  |  |
| Under 12 | 3,979 | 23 (1-182.5) | 6,749 | 18 (1-183) | 761 | 8 (1-135) |
| 12-17 | 43,521 | 16 (2-135) | 63,311 | 18 (2-127) | 830 | 12 (1.5-71) |
| 18-29 | 32,875 | 16 (1-204) | 107,257 | 15 (1-183) | 1,587 | 8 (1-183) |
| 30-39 | 71,421 | 52 (1-244) | 412,934 | 60 (4-244) | 4,835 | 16 (1-208) |
| 40-49 | 173,242 | 81 (8.5-287) | 289,443 | 79 (8-283) | 3,232 | 23 (5-244) |
| 50-59 | 391,706 | 264 (17-359) | 541,444 | 273 (17-366) | 8,634 | 141.5 (6.5-346) |
| 60-69 | 796,961 | 297 (58-450) | 925,917 | 296 (57-451) | 15,698 | 121 (1-367) |
| 70-79 | 1,262,335 | 324 (57-408) | 1,148,674 | 286 (39-405) | 24,659 | 183 (8-365) |
| 80+ | 262,237 | 181 (11-369) | 203,497 | 93 (6-360) | 5,680 | 22 (1-173) |
| Unknown | 2,529 | 54.5 (3-508) | 2,456 | 181.5 (1-352) | 301 | 301 (301-301) |
| **Ethnicity** |  |  |  |  |  |  |
| White British, White Irish, or any other white background | 2,811,518 | 273 (18-387) | 3,361,621 | 184 (14-366) | 35,675 | 220.5 (15.5-366) |
| Other | 106,435 | 69.5 (6-276) | 153,617 | 57 (3-253) | 2,235 | 82.5 (4-242) |
| Unknown | 122,853 | 89 (8-317) | 186,444 | 62 (1-259) | 28,307 | 19 (1-205) |
| **BMI Category** |  |  |  |  |  |  |
| Underweight | 45,870 | 133 (5-366) | 98,737 | 189 (9-375) | 1,658 | 259.5 (14-367) |
| Normal | 895,431 | 275 (23-404.5) | 1,282,993 | 204 (15-367.5) | 10,748 | 110 (7.5-366) |
| Overweight | 1,126,374 | 275 (30-390) | 871,081 | 204 (15-367) | 10,755 | 184 (65-312) |
| Obese | 490,424 | 269.5 (16-373.5) | 509,143 | 172 (13-362) | 6,347 | 312 (6-436) |
| Unknown | 482,707 | 99 (9-358) | 939,728 | 85 (7-317) | 36,709 | 22 (1-296) |
| **Immunocompromised** |  |  |  |  |  |  |
| Yes | 397,508 | 270 (25-378) | 405,225 | 183 (11-366) | 8,860 | 312 (47-367) |
| No/Unknown | 2,643,298 | 247 (16-384) | 3,296,457 | 178 (10-363) | 57,357 | 66 (1-312) |
| **Reported as pregnant at time of a vaccination** |  |  |  |  |  |  |
| Yes | - | - | 330,018 | 72.5 (5-244) | 0 | 0 (0-0) |
| No/Unknown | 3,040,806 | 248.5 (16-383) | 3,371,664 | 183 (12-367) | 66,217 | 73.5 (1-333) |

## Appendix C. Patient Demographics by Vaccine Brand

These tables display the proportion of patients who reported an ADR in association with a dose of the branded vaccine – any dose, 1^st^ dose, 2^nd^ dose, and 3^rd^ dose. The denominators are the total number of patients who received the branded dose (i) at any dose, or for their 1^st^ dose, or (ii) for their 2^nd^ dose and also received any branded 1^st^ dose, or for their third dose and had also received any branded 1^st^ and 2^nd^ doses.

**Table C.1.** Pfizer COVID-19 Vaccine.

|  | Patients reporting an ADR after a Pfizer dose, n=5,385 (% of those reporting any vaccine dose) |  | Patients reporting an ADR after Pfizer 1<sup>st</sup> dose, n=3,353 (% of those reporting a 1<sup>st</sup> vaccine dose) |  | Patients reporting an ADR after Pfizer 2<sup>nd</sup> dose, n=977 (% of those reporting 1<sup>st</sup> and 2<sup>nd</sup> vaccine doses) |  | Patients reporting an ADR after Pfizer 3<sup>rd</sup> dose, n=1,124(% of those reporting 1<sup>st</sup>, 2<sup>nd</sup>, and 3<sup>rd</sup> vaccine doses) |  |
| --- | --- | --- | --- | --- | --- | --- | --- | --- |
| Sex |  |  |  |  |  |  |  |  |
| Male | 1,719 | (26.0) | 1,034 | (27.1) | 304 | (18.4) | 377 | (14.1) |
| Female | 3,598 | (37.1) | 2,274 | (36.1) | 662 | (25.2) | 736 | (22.7) |
| Unknown | 68 | (32.7) | 45 | (32.4) | 11 | (22.4) | 11 | (19.3) |
| Age-bands |  |  |  |  |  |  |  |  |
| Under 12 | 30 | (33.3) | 28 | (34.6) | 4 | (12.5) | 1 | (16.7) |
| 12-17 | 559 | (46.3) | 510 | (42.8) | 76 | (30.4) | 11 | (17.5) |
| 18-29 | 488 | (47.7) | 396 | (43.2) | 99 | (29.3) | 30 | (23.1) |
| 30-39 | 1,135 | (43.2) | 896 | (39.7) | 245 | (27.7) | 66 | (15.9) |
| 40-49 | 396 | (41.3) | 229 | (43.6) | 61 | (28.5) | 114 | (28.3) |
| 50-59 | 406 | (31.7) | 132 | (40.0) | 59 | (34.7) | 189 | (25.7) |
| 60-69 | 842 | (30.6) | 316 | (31.6) | 129 | (22.9) | 357 | (23.3) |
| 70-79 | 1,237 | (24.0) | 652 | (22.5) | 250 | (16.9) | 311 | (13.4) |
| 80+ | 291 | (20.7) | 194 | (18.6) | 54 | (13.4) | 45 | (12.1) |
| Unknown | 1 | (8.3) | 0 | (0.0) | 0 | (0.0) | 0 | (0.0) |
| Ethnicity |  |  |  |  |  |  |  |  |
| White British, White Irish, or any other white background | 4,570 | (32.1) | 2,753 | (32.3) | 847 | (22.6) | 1,019 | (18.6) |
| Other | 469 | (42.2) | 358 | (40.4) | 81 | (28.7) | 60 | (26.9) |
| Unknown | 346 | (29.3) | 242 | (29.2) | 49 | (16.2) | 45 | (16.9) |
| BMI Category |  |  |  |  |  |  |  |  |
| Underweight | 170 | (40.2) | 114 | (38.5) | 31 | (27.9) | 34 | (27.9) |
| Normal | 1,737 | (34.5) | 1,008 | (33.5) | 330 | (23.7) | 399 | (19.7) |
| Overweight | 1,266 | (29.7) | 752 | (31.4) | 236 | (21.1) | 300 | (16.6) |
| Obese | 697 | (31.4) | 433 | (33.0) | 130 | (23.3) | 152 | (18.1) |
| Unknown | 1,515 | (33.2) | 1,046 | (32.3) | 250 | (21.8) | 239 | (20.3) |
| Immunocompromised |  |  |  |  |  |  |  |  |
| Yes | 608 | (31.7) | 347 | (30.1) | 109 | (21.4) | 127 | (17.0) |
| No/Unknown | 4,777 | (32.7) | 3,006 | (33.0) | 868 | (22.7) | 997 | (19.1) |
| Reported as pregnant at time of vaccination |  |  |  |  |  |  |  |  |
| Yes | 687 | (34.3) | 534 | (30.9) | 145 | (20.0) | 29 | (10.2) |
| No/Unknown | 4,698 | (32.4) | 2819 | (33.1) | 832 | (23.1) | 1095 | (19.3) |

**Table C.2.**
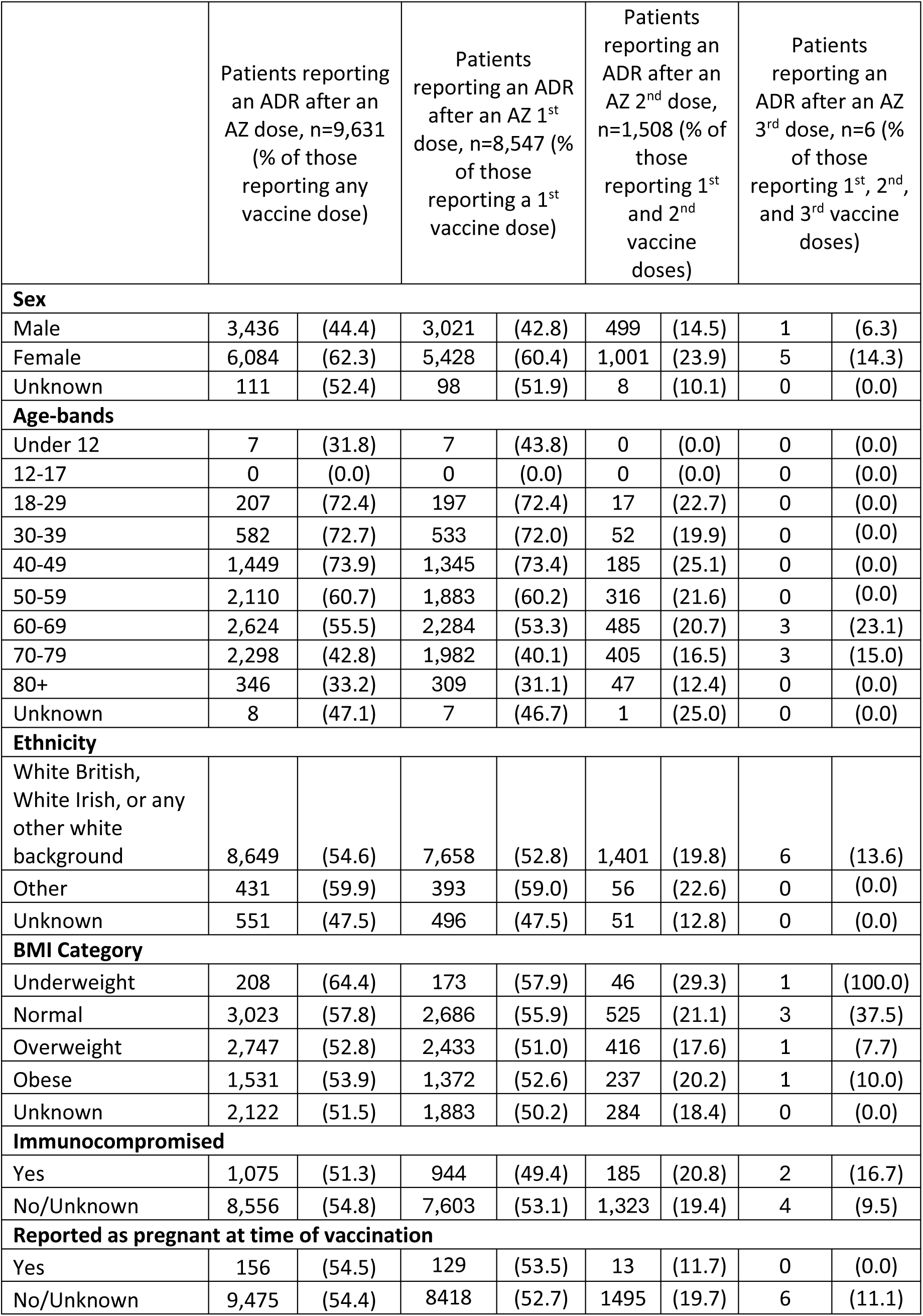
AstraZeneca COVID-19 Vaccine.

**Table C.3.** Moderna COVID-19 Vaccine.

|  | Patients reporting an ADR after a Moderna dose, n= 1,302 (% of those reporting any vaccine dose) |  | Patients reporting an ADR after Moderna 1<sup>st</sup> dose, n=300 (% of those reporting a 1<sup>st</sup> vaccine dose) |  | Patients reporting an ADR after Moderna 2<sup>nd</sup> dose, n=121 (% of those reporting 1<sup>st</sup> and 2<sup>nd</sup> vaccine doses) |  | Patients reporting an ADR after Moderna 3<sup>rd</sup> dose, n=576 (% of those reporting 1<sup>st</sup>, 2<sup>nd</sup>, and 3<sup>rd</sup> vaccine doses) |  |
| --- | --- | --- | --- | --- | --- | --- | --- | --- |
| Sex |  |  |  |  |  |  |  |  |
| Male | 428 | (22.0) | 73 | (51.0) | 29 | (47.5) | 199 | (29.7) |
| Female | 864 | (34.8) | 225 | (56.0) | 90 | (48.1) | 373 | (38.5) |
| Unknown | 10 | (25.0) | 2 | (40.0) | 2 | (66.7) | 4 | (44.4) |
| Age-bands |  |  |  |  |  |  |  |  |
| Under 12 | 3 | (25.0) | 1 | (50.0) | 0 | (0.0) | 0 | (0.0) |
| 12-17 | 6 | (33.3) | 1 | (20.0) | 1 | (100.0) | 4 | (44.4) |
| 18-29 | 118 | (59.3) | 72 | (63.7) | 25 | (58.1) | 32 | (41.6) |
| 30-39 | 254 | (46.0) | 133 | (46.8) | 62 | (48.4) | 67 | (30.0) |
| 40-49 | 202 | (51.3) | 89 | (65.9) | 29 | (46.0) | 85 | (40.9) |
| 50-59 | 228 | (38.5) | 0 | (0.0) | 2 | (33.3) | 176 | (43.2) |
| 60-69 | 228 | (22.3) | 3 | (100.0) | 1 | (20.0) | 160 | (31.3) |
| 70-79 | 238 | (16.4) | 1 | (20.0) | 1 | (33.3) | 51 | (26.6) |
| 80+ | 25 | (11.2) | 0 | (0.0) | 0 | (0.0) | 1 | (6.7) |
| Unknown | 0 | (0.0) | 0 | (0.0) | 0 | (0.0) | 0 | (0.0) |
| Ethnicity |  |  |  |  |  |  |  |  |
| White British, White Irish, or any other white background | 1,149 | (28.4) | 251 | (57.2) | 102 | (49.8) | 517 | (34.8) |
| Other | 78 | (42.6) | 30 | (46.9) | 12 | (42.9) | 36 | (41.9) |
| Unknown | 75 | (30.6) | 19 | (40.4) | 7 | (38.9) | 23 | (30.7) |
| BMI Category |  |  |  |  |  |  |  |  |
| Underweight | 28 | (33.3) | 1 | (20.0) | 1 | (50.0) | 14 | (42.4) |
| Normal | 450 | (28.9) | 115 | (58.4) | 48 | (52.7) | 189 | (33.9) |
| Overweight | 349 | (27.3) | 75 | (55.6) | 31 | (47.7) | 161 | (35.5) |
| Obese | 176 | (28.7) | 26 | (47.3) | 13 | (56.5) | 95 | (37.5) |
| Unknown | 299 | (31.9) | 83 | (52.5) | 28 | (40.0) | 117 | (33.5) |
| Immunocompromised |  |  |  |  |  |  |  |  |
| Yes | 103 | (19.8) | 8 | (33.3) | 6 | (54.5) | 43 | (30.9) |
| No/Unknown | 1,199 | (30.4) | 292 | (55.5) | 115 | (47.9) | 533 | (35.3) |
| Reported as pregnant at time of vaccination |  |  |  |  |  |  |  |  |
| Yes | 149 | (40.6) | 85 | (46.4) | 38 | (44.7) | 33 | (22.3) |
| No/Unknown | 1,153 | (28.1) | 215 | (58.6) | 83 | (50.0) | 543 | (36.2) |

**Table C.4.** Other remaining brands or unknown brands of a COVID-19 vaccine.

|  | Patients reporting an ADR after an Other/unknown vaccine dose, n=200 (% of those reporting any vaccine dose) |  | Patients reporting an ADR after Other/unknown 1<sup>st</sup> dose, n=127 (% of those reporting a 1<sup>st</sup> vaccine dose) |  | Patients reporting an ADR after Other/unknown 2<sup>nd</sup> dose, n=19 (% of those reporting 1<sup>st</sup> and 2<sup>nd</sup> vaccine doses) |  | Patients reporting an ADR after Other/unknown 3<sup>rd</sup> dose, n=36 (% of those reporting 1<sup>st</sup>, 2<sup>nd</sup>, and 3<sup>rd</sup> vaccine doses) |  |
| --- | --- | --- | --- | --- | --- | --- | --- | --- |
| Sex |  |  |  |  |  |  |  |  |
| Male | 75 | (14.9) | 45 | (28.1) | 7 | (7.8) | 15 | (14.4) |
| Female | 122 | (20.5) | 82 | (40.0) | 11 | (9.6) | 20 | (17.9) |
| Unknown | 3 | (11.1) | 0 | (0.0) | 1 | (25.0) | 1 | (33.3) |
| Age-bands |  |  |  |  |  |  |  |  |
| Under 12 | 0 | (0.0) | 0 | (0.0) | 0 | (0.0) | 0 | (0.0) |
| 12-17 | 13 | (32.5) | 8 | (36.4) | 4 | (30.8) | 0 | (0.0) |
| 18-29 | 7 | (31.8) | 4 | (40.0) | 0 | (0.0) | 2 | (28.6) |
| 30-39 | 5 | (20.0) | 5 | (38.5) | 0 | (0.0) | 0 | (0.0) |
| 40-49 | 7 | (18.9) | 5 | (38.5) | 0 | (0.0) | 2 | (25.0) |
| 50-59 | 34 | (33.0) | 26 | (59.1) | 2 | (12.5) | 2 | (20.0) |
| 60-69 | 47 | (22.0) | 36 | (56.3) | 3 | (10.3) | 3 | (10.0) |
| 70-79 | 73 | (14.3) | 31 | (22.5) | 10 | (10.4) | 26 | (20.0) |
| 80+ | 14 | (8.8) | 12 | (18.5) | 0 | (0.0) | 1 | (4.3) |
| Unknown | 0 | (0.0) | 0 | (0.0) | 0 | (0.0) | 0 | (0.0) |
| Ethnicity |  |  |  |  |  |  |  |  |
| White British, White Irish, or any other white background | 176 | (17.3) | 108 | (34.0) | 19 | (9.7) | 32 | (15.6) |
| Other | 9 | (32.1) | 8 | (50.0) | 0 | (0.0) | 1 | (25.0) |
| Unknown | 15 | (18.5) | 11 | (26.2) | 0 | (0.0) | 3 | (30.0) |
| BMI Category |  |  |  |  |  |  |  |  |
| Underweight | 6 | (18.2) | 3 | (20.0) | 1 | (14.3) | 2 | (40.0) |
| Normal | 42 | (14.5) | 31 | (40.3) | 3 | (5.1) | 5 | (9.3) |
| Overweight | 66 | (19.3) | 33 | (32.7) | 8 | (11.3) | 14 | (21.2) |
| Obese | 29 | (19.0) | 21 | (42.0) | 3 | (15.0) | 4 | (10.3) |
| Unknown | 57 | (18.5) | 39 | (29.3) | 4 | (7.8) | 11 | (20.0) |
| Immunocompromised |  |  |  |  |  |  |  |  |
| Yes | 30 | (18.1) | 19 | (31.7) | 6 | (20.0) | 3 | (8.1) |
| No | 170 | (17.7) | 108 | (34.2) | 13 | (7.3) | 33 | (18.1) |
| Reported as pregnant at time of vaccination |  |  |  |  |  |  |  |  |
| Yes | 3 | (18.8) | 2 | (66.7) | 0 | (0.0) | 1 | (14.3) |
| No/Unknown | 197 | (17.8) | 125 | (33.5) | 19 | (9.4) | 35 | (16.5) |

## Appendix D. Adverse Events Reported in association with a COVID-19 Vaccination for Any Dose and by Dose Sequence for each Vaccine Brand

### Proportion of ADRs events by MedDRA SOC level by vaccine brand and dose sequence

Figures in Appendix D presents an overview of the proportion of events by SOC level by dose sequence for each vaccine brand. Of all the adverse reactions reported, the majority of events belonged to three system organ class categories, the General Disorders and Administration Site Conditions SOC (35.50%, n=12,656), followed by Musculoskeletal and Connective Tissue Disorders (22.22%, n=7,921) and then Nervous System Disorders (20.61%, n=7,347) (Figure D.1 and Table D.1).

A similar trend in SOC event reporting was reflected in the dose specific analysis and the vaccine brand-specific analyses. In the dose-specific analyses, the top 3 most frequently reported SOCs remained the same. There were differences in the proportions of events in these SOC groups across the doses, however the use of branded vaccines differed over time and as a result impacted the dose patterns for each brand.

The most SOC categories with the most ADRs reported were similar to what was seen in the SOC level analysis for the overall cohort. The Nervous system SOC, Musculoskeletal and connective tissue conditions SOC and General disorders and Administration site conditions SOC remained the most reported SOC for each of the vaccine brands. For the Pfizer vaccine in Figure D.2 and Table D.2, the reporting of events in the Nervous system SOC remained consistent for each dose (17.4-17.8%). There was a higher proportion of reporting of musculoskeletal and connective tissue conditions with the 1^st^ dose (30.9%) compared to the 2^nd^ and 3^rd^ doses (25.4% and 26.9%), whereas there was a higher proportion of reporting with 2^nd^ dose for general disorders and administration site conditions (36.7%) compared to 1^st^ and 3^rd^ doses (29.5% vs 34.5%).

In the data presented for AstraZeneca (Figure D.3 and Table D.3), there was no events reported in the Nervous system SOC for the 3^rd^ dose, but the proportion of reporting remained consistent for 1^st^ and 2^nd^ doses (22.3% vs 22.4%). For general or administration site events, the 1^st^ dose saw a larger proportion of reporting compared to the 2^nd^ and 3^rd^ doses (37.4% vs 33.5% vs 33.3%). However, in contrast, the proportionate reporting of musculoskeletal events increased across the doses with an increase from 18.7% with 1^st^ dose to 41.7% with 3^rd^ dose. A similar increase in reporting was seen in the “Other SOC grouped” category (5.9% to 25.0%).

For the Moderna COVID-19 vaccine (Figure D.4 and Table D.4), the proportionate reporting in the Nervous system SOC were similar across the 3 doses (15.1% vs 16.7% vs 15.7%). In the Musculoskeletal and connective tissue SOC, the proportion of events reported with the 1^st^ dose was larger than that reported for the other two doses (32.7% vs 18.1% vs 26.9%). In contrast, in the General and administration site disorders SOC, the proportion of events reported was the highest with the 2^nd^ dose at 45.9%, followed by the 3^rd^ dose at 37.1% and then the lowest at 30.3% with the 1^st^ dose vaccination.

Figure D.5 and Table D.5 presents the distribution of reporting by SOC level for the other remaining vaccines or when the vaccine brand was unknown.

See ‘Supplementary Material S1’ for tables presenting the full listing of ADRs reported by MedDRA SOC, HLGT, HLT and PT level terms for each brand and dose.

**FIGURE D.1.**
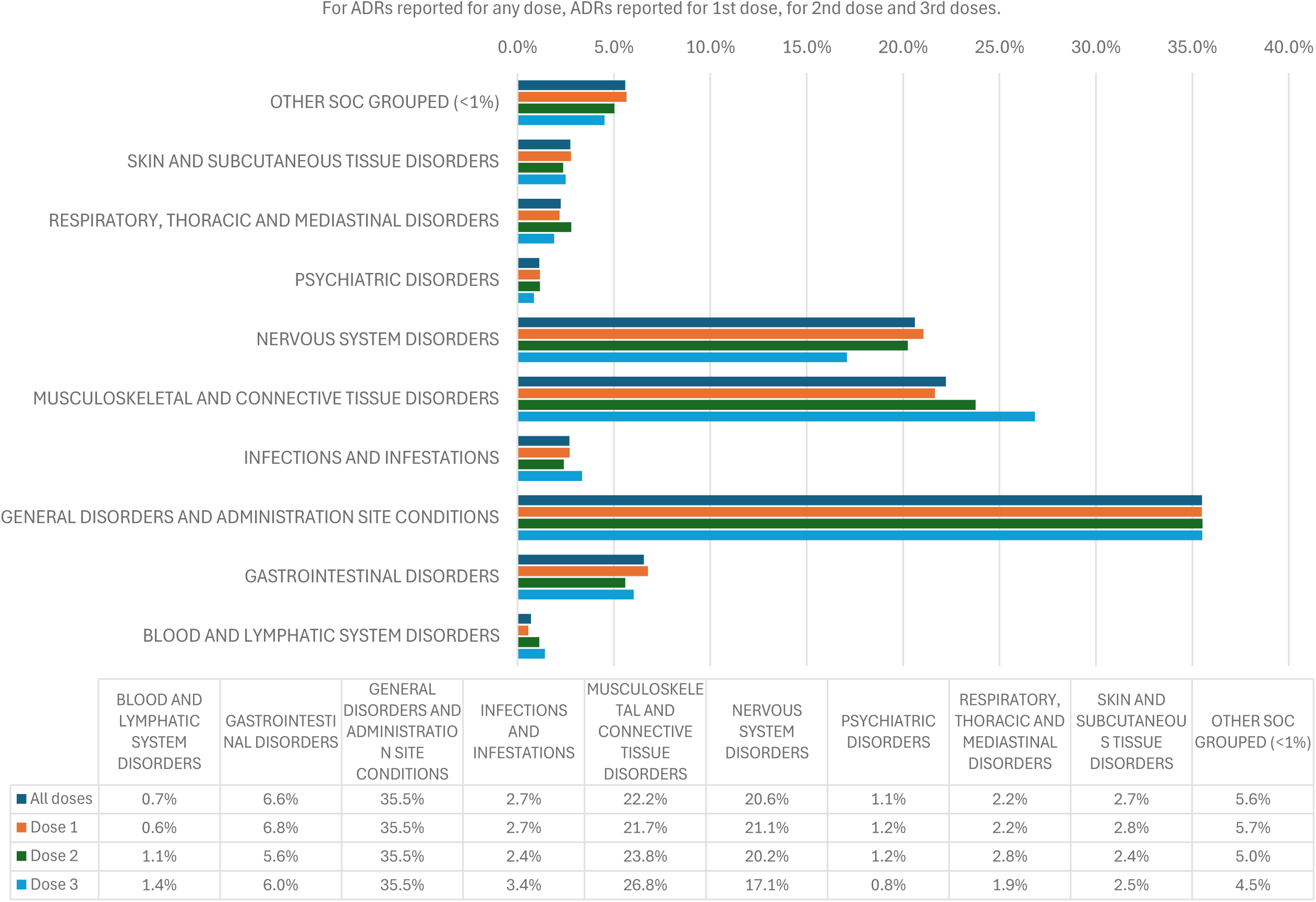
ALL COVID-19 VACCINES: TYPES OF ADR REPORTING BY SOC LEVEL

**Table D.1.** Overall COVID-19 vaccines Data table for Figure D.1.

|  | BLOOD & LYMPHATIC<br>SYSTEM DISORDERS | GASTRO-INTESTINAL<br>DISORDERS | GENERAL DISORDERS<br>& ADMINISTRATION<br>SITE CONDITIONS | INFECTIONS &<br>INFESTATIONS | MUSCULOSKELETAL<br>& CONNECTIVE<br>TISSUE DISORDERS | NERVOUS SYSTEM<br>DISORDERS | PSYCHIATRIC<br>DISORDERS | RESPIRATORY,<br>THORACIC &<br>MEDIASTINAL<br>DISORDERS | SKIN &<br>SUBCUTANEOUS<br>TISSUE DISORDERS | OTHER SOC<br>GROUPED (<1%) |
| --- | --- | --- | --- | --- | --- | --- | --- | --- | --- | --- |
| **ADR EVENTS**<br>**REPORTED** |  |  |  |  |  |  |  |  |  |  |
| ***All doses (n=35647)*** | 251 (0.7) | 2339 (6.6) | 12656 (35.5) | 961 (2.7) | 7921 (22.2) | 7347 (20.6) | 405 (1.1) | 799 (2.2) | 976 (2.7) | 1992 (5.6) |
| ***Dose 1 (n=26096)*** | 148 (0.6) | 1764 (6.8) | 9263 (35.5) | 707 (2.7) | 5651 (21.7) | 5496 (21.1) | 303 (1.2) | 566 (2.2) | 723 (2.8) | 1475 (5.7) |
| ***Dose 2 (4317)*** | 49 (1.1) | 241 (5.6) | 1534 (35.5) | 104 (2.4) | 1026 (23.8) | 874 (20.2) | 50 (1.2) | 120 (2.8) | 102 (2.4) | 217 (5.0) |
| ***Dose3 (2833)*** | 40 (1.4) | 171 (6.0) | 1006 (35.5) | 95 (3.4) | 760 (26.8) | 484 (17.1) | 24 (0.8) | 54 (1.9) | 71 (2.5) | 128 (4.5) |

Percentages reflect the proportion of ADRs by SOC level reported for each dose.

**FIGURE D.2.**
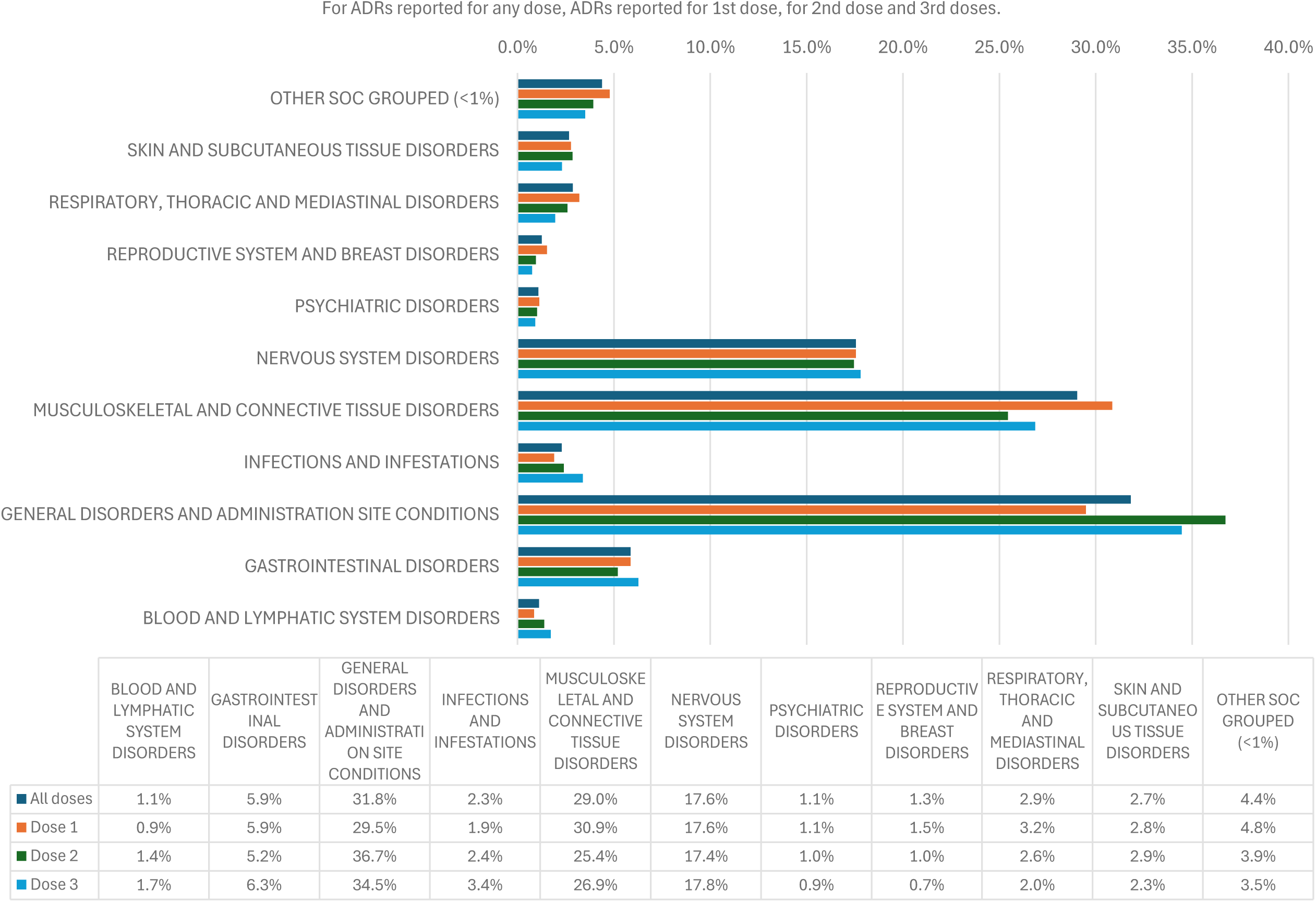
PFIZER BIONTECH VACCINE: TYPES OF ADR REPORTING BY SOC LEVEL

**Table D.2.** Pfizer BioNTech vaccine data table for Figure D.2.

|  | BLOOD AND<br>LYMPHATIC SYSTEM<br>DISORDERS | GASTRO-INTESTINAL<br>DISORDERS | GENERAL DISORDERS<br>& ADMINISTRATION<br>SITE CONDITIONS | INFECTIONS AND<br>INFESTATIONS | MUSCULOSKELETAL<br>& CONNECTIVE<br>TISSUE DISORDERS | NERVOUS SYSTEM<br>DISORDERS | PSYCHIATRIC<br>DISORDERS | REPRODUCTIVE<br>SYSTEM & BREAST<br>DISORDERS | RESPIRATORY,<br>THORACIC &<br>MEDIASTINAL<br>DISORDERS | SKIN &<br>SUBCUTANEOUS<br>TISSUE DISORDERS | OTHER SOC<br>GROUPED (<1%) |
| --- | --- | --- | --- | --- | --- | --- | --- | --- | --- | --- | --- |
| **ADR EVENTS**<br>**REPORTED** |  |  |  |  |  |  |  |  |  |  |  |
| ***All doses (n=9139)*** | 102 (1.1) | 537 (5.9) | 2909 (31.8) | 210 (2.3) | 2654 (29.0) | 1605 (17.6) | 99 (1.1) | 115 (1.3) | 262 (2.9) | 245 (2.7) | 401 (4.4) |
| ***Dose 1 (n=5705)*** | 49 (0.9) | 335 (5.9) | 1683 (29.5) | 109 (1.9) | 1761 (30.9) | 1002 (17.6) | 65 (1.1) | 87 (1.5) | 183 (3.2) | 158 (2.8) | 273 (4.8) |
| ***Dose 2 (n=1576)*** | 22 (1.4) | 82 (5.2) | 579 (36.7) | 38 (2.4) | 401 (25.4) | 275 (17.4) | 16 (1.0) | 15 (1.0) | 41 (2.6) | 45 (2.9) | 62 (3.9) |
| ***Dose3 (n=1735)*** | 30 (1.7) | 109 (5.2) | 598 (34.5) | 59 (3.4) | 466 (26.9) | 309 (17.8) | 16 (0.9) | 13 (0.7) | 34 (2.0) | 40 (2.3) | 61 (3.5) |

Percentages reflect the proportion of ADRs by SOC level reported for each dose.

**FIGURE D.3.**
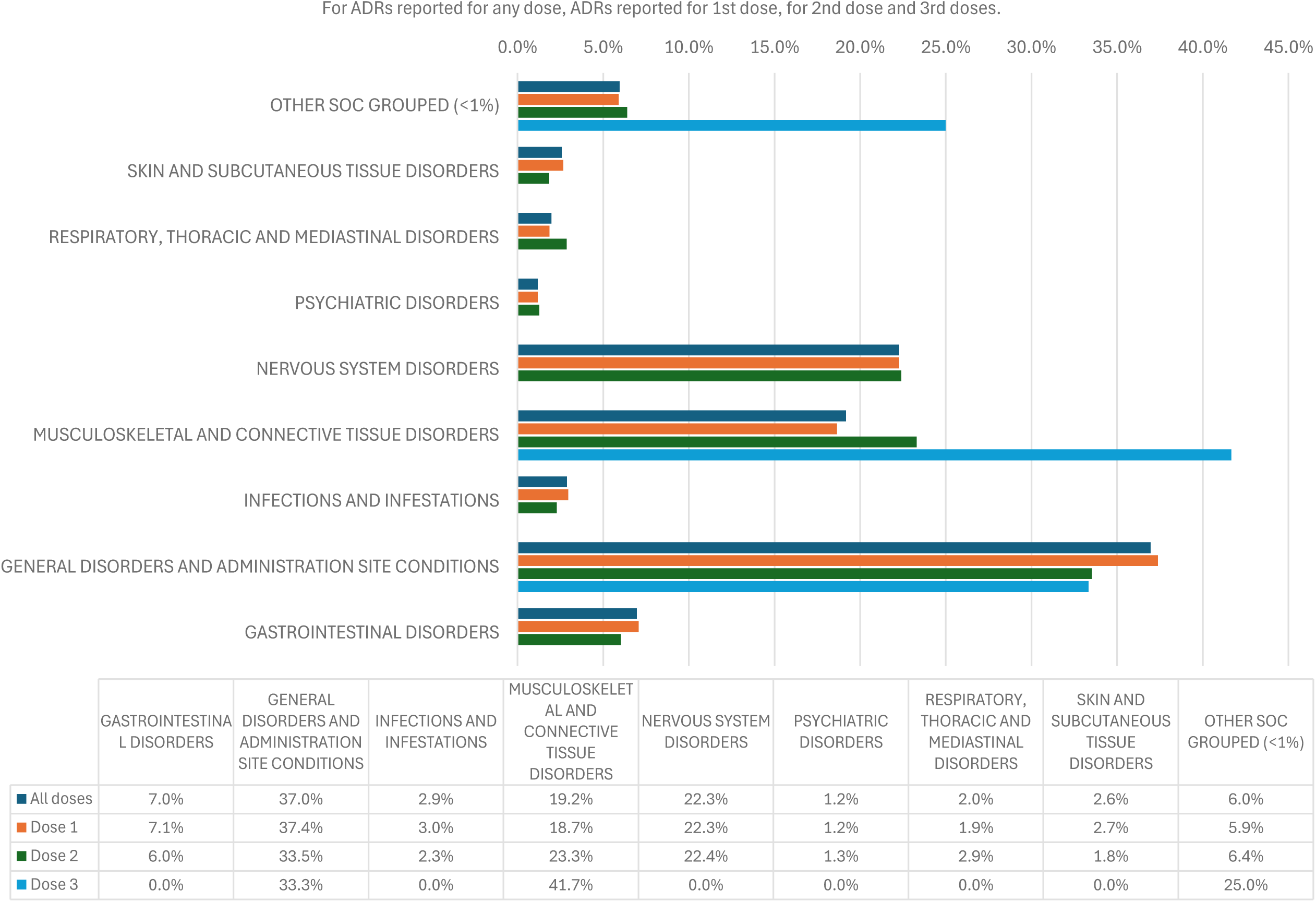
ASTRAZENECA VACCINE: TYPES OF ADR REPORTING BY SOC LEVEL

**Table D.3.** AstraZeneca vaccine data table for Figure D.3.

|  | GASTROINTESTINAL DISORDERS | GENERAL DISORDERS & ADMINISTRATION SITE CONDITIONS | INFECTIONS & INFESTATIONS | MUSCULOSKELETAL & CONNECTIVE TISSUE DISORDERS | NERVOUS SYSTEM DISORDERS | PSYCHIATRIC DISORDERS | RESPIRATORY, THORACIC & MEDIASTINAL DISORDERS | SKIN & SUBCUTANEOUS TISSUE DISORDERS | OTHER SOC GROUPED (<1%) |
| --- | --- | --- | --- | --- | --- | --- | --- | --- | --- |
| **ADR EVENTS REPORTED** |  |  |  |  |  |  |  |  |  |
| ***All doses (n=22004)*** | 1532 (7.0) | 8132 (37.0) | 635 (2.9) | 4220 (19.2) | 4904 (22.3) | 262 (1.2) | 435 (2.0) | 570 (2.6) | 1314 (6.0) |
| ***Dose 1 (n=19559)*** | 1385 (7.1) | 7312 (37.4) | 579 (3.0) | 3648 (18.7) | 4359 (22.3) | 231 (1.2) | 365 (1.9) | 525 (2.7) | 1155 (5.9) |
| ***Dose 2 (n=2433)*** | 147 (6.0) | 816 (33.5) | 56 (2.3) | 567 (23.3) | 545 (22.4) | 31 (1.3) | 70 (2.9) | 45 (1.8) | 156 (6.4) |
| ***Dose3 (n=12)*** | 0 (0.0) | 4 (33.3) | 0 (0.0) | 5 (41.7) | 0 (0.0) | 0 (0.0) | 0 (0.0) | 0 (0.0) | 3 (25.0) |

Percentages reflect the proportion of ADRs by SOC level reported for each dose.

**FIGURE D.4.**
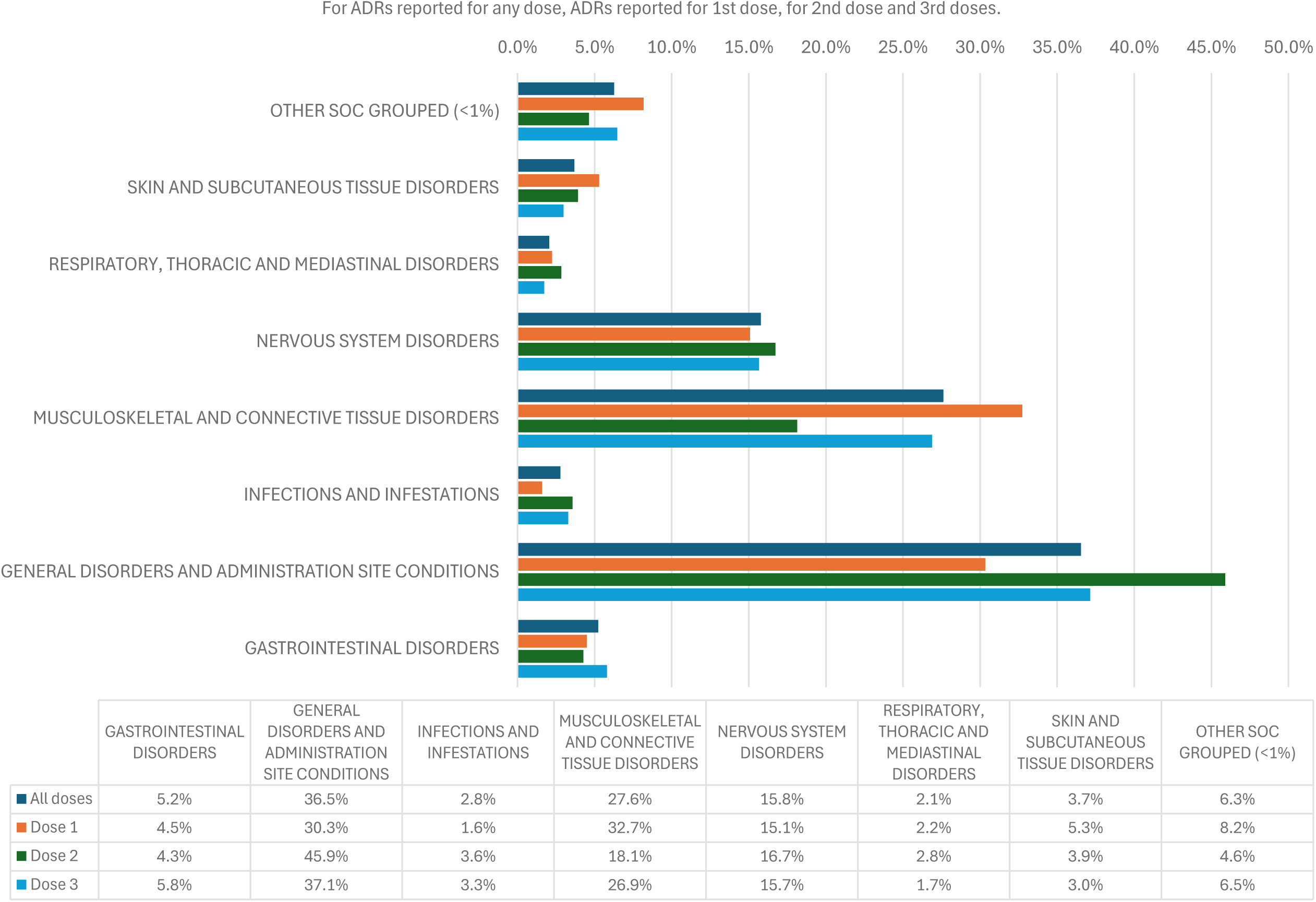
MODERNA VACCINE: TYPES OF ADR REPORTING BY SOC LEVEL

**Table D.4.** Moderna vaccine data table for Figure D.4.

|  | GASTROINTESTINAL DISORDERS | GENERAL DISORDERS & ADMINISTRATION SITE CONDITIONS | INFECTIONS & INFESTATIONS | MUSCULOSKELETAL & CONNECTIVE TISSUE DISORDERS | NERVOUS SYSTEM DISORDERS | RESPIRATORY, THORACIC & MEDIASTINAL DISORDERS | SKIN & SUBCUTANEOUS TISSUE DISORDERS | OTHER SOC GROUPED (<1%) |
| --- | --- | --- | --- | --- | --- | --- | --- | --- |
| **ADR EVENTS REPORTED** |  |  |  |  |  |  |  |  |
| ***All doses (n=2230)*** | 117 (5.2) | 815 (36.5) | 62 (2.8) | 616 (27.6) | 352 (15.8) | 46 (2.1) | 82 (3.7) | 140 (6.3) |
| ***Dose 1 (n=623)*** | 28 (4.5) | 189 (30.3) | 10 (1.6) | 204 (32.7) | 94 (15.1) | 14 (2.2) | 33 (5.3) | 51 (8.2) |
| ***Dose 2 (n=281)*** | 12 (4.3) | 129 (45.9) | 10 (3.6) | 51 (18.1) | 47 (16.7) | 8 (2.8) | 11 (3.9) | 13 (4.6) |
| ***Dose3 (n=1034)*** | 60 (5.8) | 384 (37.1) | 34 (3.3) | 278 (26.9) | 162 (15.7) | 18 (1.7) | 31 (3.0) | 67 (6.5) |

Percentages reflect the proportion of ADRs by SOC level reported for each dose.

**FIGURE D.5.**
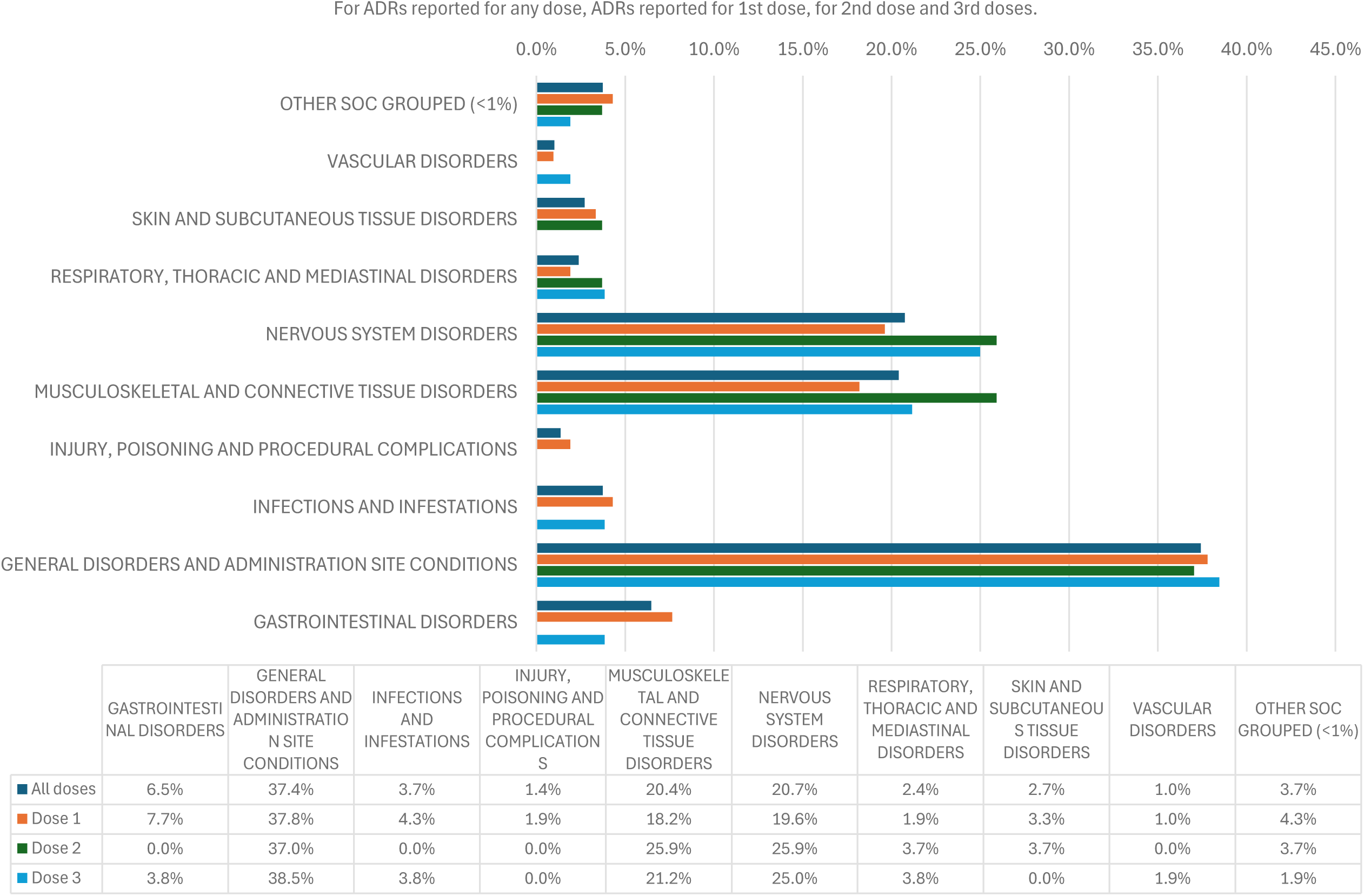
OTHER OR UNKNOWN BRANDED VACCINE: TYPES OF ADR REPORTING BY SOC LEVEL

**Table D.5.** Other or unknown branded vaccine data table for Figure D.5.

|  | GASTROINTESTINAL DISORDERS | GENERAL DISORDERS & ADMINISTRATION SITE CONDITIONS | INFECTIONS & INFESTATIONS | INJURY, POISONING AND PROCEDURAL COMPLICATIONS | MUSCULOSKELETAL & CONNECTIVE TISSUE DISORDERS | NERVOUS SYSTEM DISORDERS | RESPIRATORY, THORACIC & MEDIASTINAL DISORDERS | SKIN & SUBCUTANEOUS TISSUE DISORDERS | VASCULAR DISORDERS | OTHER SOC GROUPED (<1%) |
| --- | --- | --- | --- | --- | --- | --- | --- | --- | --- | --- |
| **ADR EVENTS REPORTED** |  |  |  |  |  |  |  |  |  |  |
| ***All doses (n=294)*** | 19 (6.5) | 110 (37.4) | 11 (3.7) | 4 (1.4) | 60 (20.4) | 61 (20.7) | 7 (2.4) | 8 (2.7) | 3 (1.0) | 11 (3.7) |
| ***Dose 1 (n=209)*** | 16 (7.7) | 79 (37.8) | 9 (4.3) | 4 (1.9) | 38 (18.2) | 41 (19.6) | 4 (1.9) | 7 (3.3) | 2 (1.0) | 9 (4.3) |
| ***Dose 2 (n=27)*** | 0 (0.0) | 10 (37.0) | 0 (0.0) | 0 (0.0) | 7 (25.9) | 7 (25.9) | 1 (3.7) | 1 (3.7) | 0 (0.0) | 1 (3.7) |
| ***Dose3 (n=52)*** | 2 (3.8) | 20 (38.5) | 2 (3.8) | 0 (0.0) | 11 (21.2) | 13 (25.0) | 2 (3.8) | 0 (0.0) | 1 (1.9) | 1 (1.9) |

Percentages reflect the proportion of ADRs by SOC level reported for each dose.

## Appendix E. Adverse Events Reported in Patients aged under 40 years versus Patients aged 40 years or over

Due to the approach taken to rollout the national vaccination campaign to the UK population, the brand and dose analysis is confounded by age. COVID-19 vaccination was prioritised by descending age groups, with the older population being the first to receive Pfizer and AstraZeneca before the younger population. Additionally, the supply availability of different vaccine brands over time would have impacted which vaccine an individual would receive. The brand of vaccine recommended would also be dependent on the age of the patient or if they were clinically vulnerable. The AstraZeneca vaccine was not recommended for use in those aged under 40 years after early May 2021 and therefore there could be differences in the ADR reporting profile between those aged under 40 years and those aged 40 years or older.

In the age-stratified analysis exploring the ADRs reported in those aged under 40 years and in those aged 40 years or older, the age-stratified analyses indicate that any differences in the ADR reporting patterns overall between the age groups remain small (Table E.1).

**Table E.1.** Top 10 ADRs reported for the individuals aged under 40 years compared against those aged 40 years and older.

| **Dose** | **Under 40 years** |  |  | **40 years and older** |  |  |
| --- | --- | --- | --- | --- | --- | --- |
|  | **Ranked Medical event (PT level) reported** | **Number of events** | **Percentage of ADRs reported for each dose** | **Ranked Medical event (PT level) reported** | **Number of events** | **Percentage of ADRs reported for each dose** |
| *First dose* | Pain in extremity | 945 | 15.3% | Headache | 3033 | 15.2% |
|  | Headache | 730 | 11.8% | Fatigue | 2323 | 11.7% |
|  | Fatigue | 621 | 10.1% | Pain in extremity | 1464 | 7.4% |
|  | Pyrexia | 439 | 7.1% | Pyrexia | 1232 | 6.2% |
|  | Myalgia | 294 | 4.8% | Chills | 1131 | 5.7% |
|  | Nausea | 240 | 3.9% | Myalgia | 1000 | 5.0% |
|  | Pain | 219 | 3.5% | Arthralgia | 805 | 4.0% |
|  | Chills | 209 | 3.4% | Nausea | 753 | 3.8% |
|  | Arthralgia | 151 | 2.4% | Pain | 507 | 2.5% |
|  | Injection site pain | 147 | 2.4% | Influenza | 413 | 2.1% |
|  | **TOTAL EVENTS** | **6179** |  | **TOTAL EVENTS** | **19905** |  |
| *Second dose* | Fatigue | 136 | 12.7% | Fatigue | 495 | 15.3% |
|  | Headache | 120 | 11.2% | Headache | 426 | 13.2% |
|  | Pain in extremity | 108 | 10.1% | Pain in extremity | 345 | 10.7% |
|  | Pyrexia | 91 | 8.5% | Myalgia | 167 | 5.2% |
|  | Pain | 52 | 4.8% | Arthralgia | 120 | 3.7% |
|  | Nausea | 37 | 3.4% | Pyrexia | 110 | 3.4% |
|  | Myalgia | 34 | 3.2% | Chills | 91 | 2.8% |
|  | Chills | 31 | 2.9% | Nausea | 87 | 2.7% |
|  | Arthralgia | 29 | 2.7% | Limb discomfort | 82 | 2.5% |
|  | Dizziness | 26 | 2.4% | Pain | 78 | 2.4% |
|  | **TOTAL EVENTS** | **1073** |  | **TOTAL EVENTS** | **3238** |  |
| *Third dose* | Headache | 48 | 12.8% | Pain in extremity | 360 | 14.6% |
|  | Fatigue | 47 | 12.6% | Headache | 289 | 11.8% |
|  | Pain in extremity | 40 | 10.7% | Fatigue | 255 | 10.4% |
|  | Pyrexia | 27 | 7.2% | Pyrexia | 107 | 4.4% |
|  | Chills | 22 | 5.9% | Arthralgia | 107 | 4.4% |
|  | Myalgia | 18 | 4.8% | Chills | 104 | 4.2% |
|  | Nausea | 16 | 4.3% | Myalgia | 96 | 3.9% |
|  | Pain | 13 | 3.5% | Nausea | 82 | 3.3% |
|  | Injection site pain | 8 | 2.1% | Pain | 65 | 2.6% |
|  | Influenza | 8 | 2.1% | Limb discomfort | 62 | 2.5% |
|  | **TOTAL EVENTS** | **374** |  | **TOTAL EVENTS** | **2459** |  |

## Appendix F. Demographics of Registered Females

**Table F.1.**
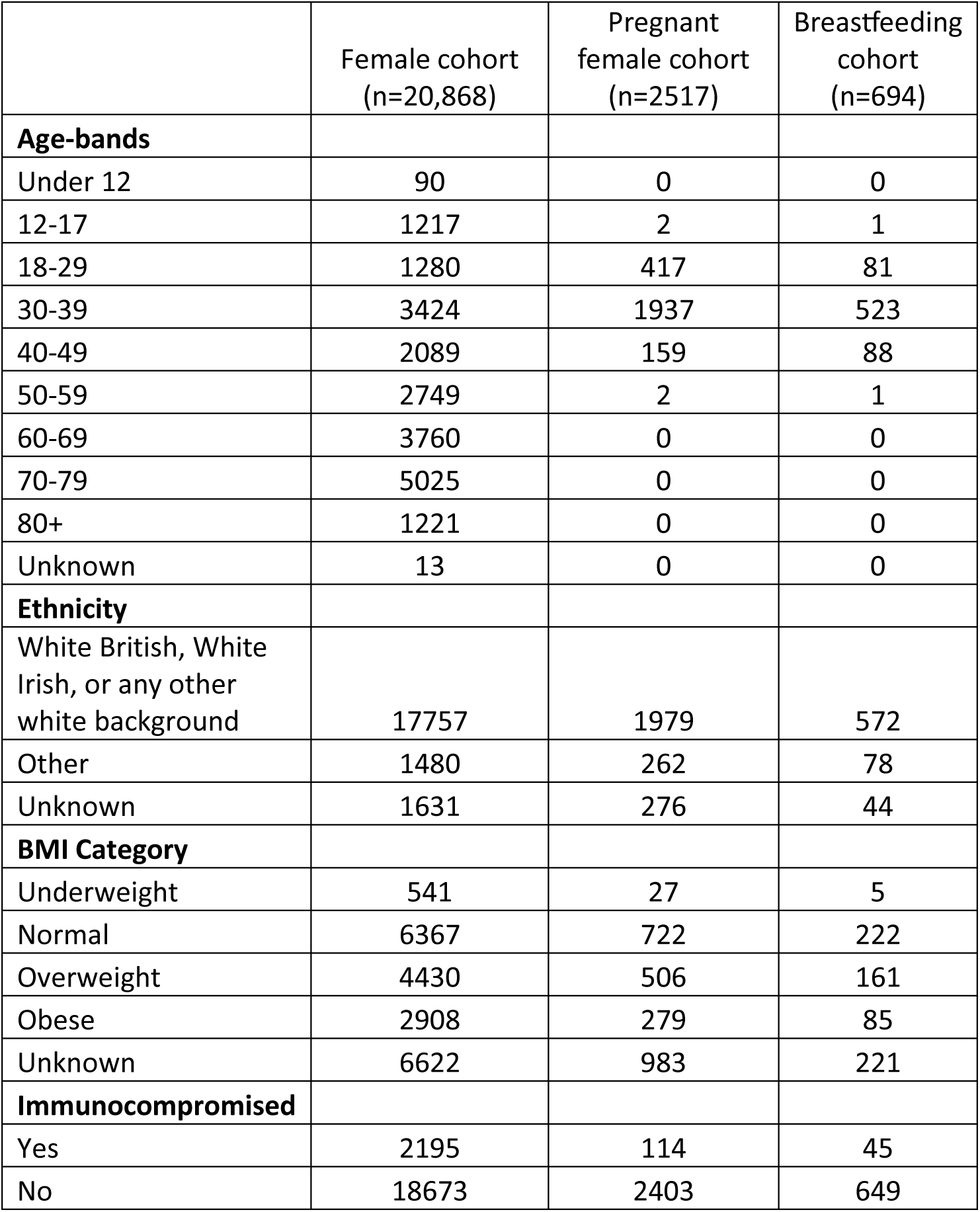
Demographics of registered females: female patients, those reporting being pregnant or breastfeeding at the time of a vaccination.

|  | Female cohort<br>(n=20,868) | Pregnant<br>female cohort<br>(n=2517) | Breastfeeding<br>cohort<br>(n=694) |
| --- | --- | --- | --- |
| **Age-bands** |  |  |  |
| Under 12 | 90 | 0 | 0 |
| 12-17 | 1217 | 2 | 1 |
| 18-29 | 1280 | 417 | 81 |
| 30-39 | 3424 | 1937 | 523 |
| 40-49 | 2089 | 159 | 88 |
| 50-59 | 2749 | 2 | 1 |
| 60-69 | 3760 | 0 | 0 |
| 70-79 | 5025 | 0 | 0 |
| 80+ | 1221 | 0 | 0 |
| Unknown | 13 | 0 | 0 |
| **Ethnicity** |  |  |  |
| White British, White Irish, or any other white background | 17757 | 1979 | 572 |
| Other | 1480 | 262 | 78 |
| Unknown | 1631 | 276 | 44 |
| **BMI Category** |  |  |  |
| Underweight | 541 | 27 | 5 |
| Normal | 6367 | 722 | 222 |
| Overweight | 4430 | 506 | 161 |
| Obese | 2908 | 279 | 85 |
| Unknown | 6622 | 983 | 221 |
| **Immunocompromised** |  |  |  |
| Yes | 2195 | 114 | 45 |
| No | 18673 | 2403 | 649 |

