## Supplementary Material S1. ADR listing for "Implementation and results of active vaccine safety monitoring during the COVID-19 pandemic in the UK"

### Supplementary Material S1. Adverse Events (ADRs) Reported in association with a COVID-19 Vaccination for Any Dose and by Dose Sequence for each Vaccine Brand.

<u>Total ADR events reported</u>

The tables listed below are presented in the file named "Supplementary_ S1-ADR_Listing". Each of these tables capture the full listing of ADRs reported by MedDRA SOC, HLGT, HLT and PT level terms for each brand and dose.

| Table Number | Description |
|---|---|
| **S1-1** | ADR listing for events reported in the YCVM in (a) those reporting any vaccination dose and (b) in those who had reported a 1st dose vaccination and any subsequent doses. |
| **S1-2** | Pfizer BioNTech COVID-19 vaccine: ADR listing for events reported in the YCVM in (a) those reporting any vaccination dose and (b) in those who had reported a 1st dose vaccination and any subsequent doses. |
| **S1-3** | AstraZeneca COVID-19 vaccine: ADR listing for events reported in the YCVM in (a) those reporting any vaccination dose and (b) in those who had reported a 1st dose vaccination and any subsequent doses. |
| **S1-4** | Moderna COVID-19 vaccine: ADR listing for events reported in the YCVM in (a) those reporting any vaccination dose and (b) in those who had reported a 1st dose vaccination and any subsequent doses. |
| **S1-5** | Others/Unknown COVID-19 vaccine: ADR listing for events reported in the YCVM in (a) those reporting any vaccination dose and (b) in those who had reported a 1st dose vaccination and any subsequent doses. |
| **S1-6** | COVID-19 vaccine ADR listing for events reported in the YCVM with no dose identification information in (a) those reporting any vaccination dose and (b) in those who had reported a 1st dose vaccination and any subsequent doses. |

**TABLE S1-1. ADR listing for events reported in the YCVM in (a) those reporting any vaccination dose and (b) in those who had reported a 1st dose vaccination and any subsequent doses.**

| | Patients with any vaccination dose: ADR Counts | | | | | | Patients reporting a 1st dose: ADR Counts | | | | | |
|---|---|---|---|---|---|---|---|---|---|---|---|---|
| **REACTION TERM (SOC, *HLGT*, *HLT*, PT)** | All doses | 1st dose | 2nd dose | 3rd dose | Other doses | Unknown | All doses | 1st dose | 2nd dose | 3rd dose | Other doses | Unknown |
| ***(freetext)*** | 70 | 65 | 2 | 0 | 0 | 3 | 69 | 65 | 1 | 0 | 0 | 3 |
| **BLOOD AND LYMPHATIC SYSTEM DISORDERS** | | | | | | | | | | | | |
| ***COAGULOPATHIES AND BLEEDING DIATHESES (EXCL THROMBOCYTOPENIC)*** | | | | | | | | | | | | |
| *BLEEDING TENDENCIES* | | | | | | | | | | | | |
| INCREASED TENDENCY TO BRUISE | 2 | 0 | 1 | 0 | 0 | 1 | 2 | 0 | 1 | 0 | 0 | 1 |
| ***HAEMOGLOBINOPATHIES*** | | | | | | | | | | | | |
| *SICKLE CELL TRAIT AND DISORDERS* | | | | | | | | | | | | |
| SICKLE CELL ANAEMIA WITH CRISIS | 1 | 1 | 0 | 0 | 0 | 0 | 1 | 1 | 0 | 0 | 0 | 0 |
| ***PLATELET DISORDERS*** | | | | | | | | | | | | |
| *THROMBOCYTOPENIAS* | | | | | | | | | | | | |
| IMMUNE THROMBOCYTOPENIA | 1 | 0 | 1 | 0 | 0 | 0 | 0 | 0 | 0 | 0 | 0 | 0 |
| ***SPLEEN, LYMPHATIC AND RETICULOENDOTHELIAL SYSTEM DISORDERS*** | | | | | | | | | | | | |
| *LYMPHATIC SYSTEM DISORDERS NEC* | | | | | | | | | | | | |
| LYMPH NODE PAIN | 39 | 15 | 12 | 8 | 0 | 4 | 34 | 15 | 10 | 5 | 0 | 4 |
| LYMPHADENITIS | 3 | 1 | 1 | 1 | 0 | 0 | 3 | 1 | 1 | 1 | 0 | 0 |
| LYMPHADENOPATHY | 234 | 131 | 52 | 41 | 1 | 9 | 211 | 131 | 37 | 34 | 1 | 8 |
| **CARDIAC DISORDERS** | | | | | | | | | | | | |
| ***CARDIAC ARRHYTHMIAS*** | | | | | | | | | | | | |
| *RATE AND RHYTHM DISORDERS NEC* | | | | | | | | | | | | |
| ARRHYTHMIA | 1 | 1 | 0 | 0 | 0 | 0 | 1 | 1 | 0 | 0 | 0 | 0 |
| CARDIAC FLUTTER | 12 | 4 | 5 | 3 | 0 | 0 | 9 | 4 | 3 | 2 | 0 | 0 |
| EXTRASYSTOLES | 2 | 0 | 1 | 1 | 0 | 0 | 2 | 0 | 1 | 1 | 0 | 0 |
| TACHYCARDIA | 16 | 14 | 2 | 0 | 0 | 0 | 14 | 14 | 0 | 0 | 0 | 0 |
| *SUPRAVENTRICULAR ARRHYTHMIAS* | | | | | | | | | | | | |
| ATRIAL FIBRILLATION | 4 | 2 | 1 | 1 | 0 | 0 | 2 | 2 | 0 | 0 | 0 | 0 |
| SUPRAVENTRICULAR TACHYCARDIA | 1 | 1 | 0 | 0 | 0 | 0 | 1 | 1 | 0 | 0 | 0 | 0 |
| ***CARDIAC DISORDERS, SIGNS AND SYMPTOMS NEC*** | | | | | | | | | | | | |

| | | | | | | | | | | | | |
|---|---|---|---|---|---|---|---|---|---|---|---|---|
| *CARDIAC DISORDERS NEC* | | | | | | | | | | | | |
| CARDIOVASCULAR DISORDER | 1 | 1 | 0 | 0 | 0 | 0 | 1 | 1 | 0 | 0 | 0 | 0 |
| *CARDIAC SIGNS AND SYMPTOMS NEC* | | | | | | | | | | | | |
| PALPITATIONS | 100 | 61 | 17 | 11 | 1 | 10 | 92 | 61 | 12 | 10 | 1 | 8 |
| ***CORONARY ARTERY DISORDERS*** | | | | | | | | | | | | |
| *ISCHAEMIC CORONARY ARTERY DISORDERS* | | | | | | | | | | | | |
| ANGINA PECTORIS | 2 | 2 | 0 | 0 | 0 | 0 | 2 | 2 | 0 | 0 | 0 | 0 |
| MYOCARDIAL INFARCTION | 2 | 2 | 0 | 0 | 0 | 0 | 2 | 2 | 0 | 0 | 0 | 0 |
| ***HEART FAILURES*** | | | | | | | | | | | | |
| *HEART FAILURES NEC* | | | | | | | | | | | | |
| CARDIAC FAILURE | 1 | 0 | 0 | 1 | 0 | 0 | 0 | 0 | 0 | 0 | 0 | 0 |
| ***MYOCARDIAL DISORDERS*** | | | | | | | | | | | | |
| *NONINFECTIOUS MYOCARDITIS* | | | | | | | | | | | | |
| MYOCARDITIS | 5 | 1 | 1 | 0 | 1 | 2 | 4 | 1 | 0 | 0 | 1 | 2 |
| ***PERICARDIAL DISORDERS*** | | | | | | | | | | | | |
| *NONINFECTIOUS PERICARDITIS* | | | | | | | | | | | | |
| PERICARDITIS | 1 | 1 | 0 | 0 | 0 | 0 | 1 | 1 | 0 | 0 | 0 | 0 |
| **CONGENITAL, FAMILIAL AND GENETIC DISORDERS** | | | | | | | | | | | | |
| ***CARDIAC AND VASCULAR DISORDERS CONGENITAL*** | | | | | | | | | | | | |
| *CARDIAC DISORDERS CONGENITAL NEC* | | | | | | | | | | | | |
| HEART DISEASE CONGENITAL | 1 | 0 | 0 | 1 | 0 | 0 | 1 | 0 | 0 | 1 | 0 | 0 |
| ***METABOLIC AND NUTRITIONAL DISORDERS CONGENITAL*** | | | | | | | | | | | | |
| *INBORN ERRORS OF AMINO ACID METABOLISM* | | | | | | | | | | | | |
| HYPERGLYCINAEMIA | 1 | 1 | 0 | 0 | 0 | 0 | 1 | 1 | 0 | 0 | 0 | 0 |
| *INBORN ERRORS OF STEROID SYNTHESIS* | | | | | | | | | | | | |
| 11-BETA-HYDROXYLASE DEFICIENCY | 1 | 1 | 0 | 0 | 0 | 0 | 1 | 1 | 0 | 0 | 0 | 0 |
| ***NEUROLOGICAL DISORDERS CONGENITAL*** | | | | | | | | | | | | |
| *PERIPHERAL NERVOUS SYSTEM DISORDERS CONGENITAL NEC* | | | | | | | | | | | | |
| PAROXYSMAL EXTREME PAIN DISORDER | 1 | 0 | 1 | 0 | 0 | 0 | 1 | 0 | 1 | 0 | 0 | 0 |
| **EAR AND LABYRINTH DISORDERS** | | | | | | | | | | | | |
| ***AURAL DISORDERS NEC*** | | | | | | | | | | | | |
| *EAR DISORDERS NEC* | | | | | | | | | | | | |
| EAR DISCOMFORT | 4 | 2 | 2 | 0 | 0 | 0 | 4 | 2 | 2 | 0 | 0 | 0 |

| | | | | | | | | | | | | |
|---|---|---|---|---|---|---|---|---|---|---|---|---|
| EAR DISORDER | 1 | 0 | 1 | 0 | 0 | 0 | 1 | 0 | 1 | 0 | 0 | 0 |
| EAR PAIN | 49 | 37 | 9 | 1 | 0 | 2 | 44 | 37 | 5 | 0 | 0 | 2 |
| EAR SWELLING | 1 | 1 | 0 | 0 | 0 | 0 | 1 | 1 | 0 | 0 | 0 | 0 |
| ***EXTERNAL EAR DISORDERS (EXCL CONGENITAL)*** | | | | | | | | | | | | |
| *EXTERNAL EAR DISORDERS NEC* | | | | | | | | | | | | |
| EXCESSIVE CERUMEN PRODUCTION | 2 | 2 | 0 | 0 | 0 | 0 | 2 | 2 | 0 | 0 | 0 | 0 |
| ***HEARING DISORDERS*** | | | | | | | | | | | | |
| *HEARING LOSSES* | | | | | | | | | | | | |
| DEAFNESS | 5 | 3 | 2 | 0 | 0 | 0 | 4 | 3 | 1 | 0 | 0 | 0 |
| DEAFNESS NEUROSENSORY | 1 | 1 | 0 | 0 | 0 | 0 | 1 | 1 | 0 | 0 | 0 | 0 |
| DEAFNESS PERMANENT | 1 | 0 | 1 | 0 | 0 | 0 | 0 | 0 | 0 | 0 | 0 | 0 |
| DEAFNESS UNILATERAL | 2 | 1 | 1 | 0 | 0 | 0 | 2 | 1 | 1 | 0 | 0 | 0 |
| HYPOACUSIS | 3 | 2 | 1 | 0 | 0 | 0 | 3 | 2 | 1 | 0 | 0 | 0 |
| SUDDEN HEARING LOSS | 1 | 1 | 0 | 0 | 0 | 0 | 1 | 1 | 0 | 0 | 0 | 0 |
| *HYPERACUSIA* | | | | | | | | | | | | |
| HYPERACUSIS | 1 | 0 | 1 | 0 | 0 | 0 | 1 | 0 | 1 | 0 | 0 | 0 |
| ***INNER EAR AND VIIITH CRANIAL NERVE DISORDERS*** | | | | | | | | | | | | |
| *INNER EAR SIGNS AND SYMPTOMS* | | | | | | | | | | | | |
| MOTION SICKNESS | 2 | 2 | 0 | 0 | 0 | 0 | 2 | 2 | 0 | 0 | 0 | 0 |
| TINNITUS | 75 | 46 | 15 | 7 | 0 | 7 | 71 | 46 | 12 | 6 | 0 | 7 |
| VERTIGO | 47 | 30 | 10 | 3 | 1 | 3 | 45 | 30 | 9 | 2 | 1 | 3 |
| VERTIGO LABYRINTHINE | 2 | 2 | 0 | 0 | 0 | 0 | 2 | 2 | 0 | 0 | 0 | 0 |
| VERTIGO POSITIONAL | 5 | 4 | 1 | 0 | 0 | 0 | 5 | 4 | 1 | 0 | 0 | 0 |
| **ENDOCRINE DISORDERS** | | | | | | | | | | | | |
| ***THYROID GLAND DISORDERS*** | | | | | | | | | | | | |
| *THYROID HYPERFUNCTION DISORDERS* | | | | | | | | | | | | |
| HYPERTHYROIDISM | 1 | 1 | 0 | 0 | 0 | 0 | 1 | 1 | 0 | 0 | 0 | 0 |
| **EYE DISORDERS** | | | | | | | | | | | | |
| ***EYE DISORDERS NEC*** | | | | | | | | | | | | |
| *LACRIMATION DISORDERS* | | | | | | | | | | | | |
| DRY EYE | 8 | 3 | 1 | 4 | 0 | 0 | 7 | 3 | 1 | 3 | 0 | 0 |
| LACRIMATION INCREASED | 4 | 2 | 0 | 1 | 0 | 1 | 3 | 2 | 0 | 0 | 0 | 1 |
| *OCULAR DISORDERS NEC* | | | | | | | | | | | | |
| EYE OEDEMA | 1 | 1 | 0 | 0 | 0 | 0 | 1 | 1 | 0 | 0 | 0 | 0 |
| EYE PAIN | 95 | 72 | 8 | 6 | 0 | 9 | 94 | 72 | 7 | 6 | 0 | 9 |
| EYE SWELLING | 5 | 3 | 0 | 2 | 0 | 0 | 3 | 3 | 0 | 0 | 0 | 0 |
| EYE ULCER | 1 | 1 | 0 | 0 | 0 | 0 | 1 | 1 | 0 | 0 | 0 | 0 |
| EYELID PAIN | 1 | 1 | 0 | 0 | 0 | 0 | 1 | 1 | 0 | 0 | 0 | 0 |

| | | | | | | | | | | | | |
|---|---|---|---|---|---|---|---|---|---|---|---|---|
| OCULAR DISCOMFORT | 2 | 2 | 0 | 0 | 0 | 0 | 2 | 2 | 0 | 0 | 0 | 0 |
| PERIORBITAL DISCOMFORT | 1 | 0 | 1 | 0 | 0 | 0 | 0 | 0 | 0 | 0 | 0 | 0 |
| PERIORBITAL SWELLING | 2 | 0 | 1 | 1 | 0 | 0 | 2 | 0 | 1 | 1 | 0 | 0 |
| ***OCULAR HAEMORRHAGES AND VASCULAR DISORDERS NEC*** | | | | | | | | | | | | |
| *CONJUNCTIVAL AND CORNEAL BLEEDING AND VASCULAR DISORDERS* | | | | | | | | | | | | |
| CONJUNCTIVAL HAEMORRHAGE | 2 | 1 | 0 | 0 | 0 | 1 | 2 | 1 | 0 | 0 | 0 | 1 |
| *LID BLEEDING AND VASCULAR DISORDERS* | | | | | | | | | | | | |
| EYELID BLEEDING | 1 | 1 | 0 | 0 | 0 | 0 | 1 | 1 | 0 | 0 | 0 | 0 |
| ***OCULAR INFECTIONS, IRRITATIONS AND INFLAMMATIONS*** | | | | | | | | | | | | |
| *LID, LASH AND LACRIMAL INFECTIONS, IRRITATIONS AND INFLAMMATIONS* | | | | | | | | | | | | |
| BLEPHARITIS | 1 | 1 | 0 | 0 | 0 | 0 | 1 | 1 | 0 | 0 | 0 | 0 |
| ERYTHEMA OF EYELID | 1 | 1 | 0 | 0 | 0 | 0 | 1 | 1 | 0 | 0 | 0 | 0 |
| EYELID IRRITATION | 1 | 1 | 0 | 0 | 0 | 0 | 1 | 1 | 0 | 0 | 0 | 0 |
| *OCULAR INFECTIONS, INFLAMMATIONS AND ASSOCIATED MANIFESTATIONS* | | | | | | | | | | | | |
| EYE DISCHARGE | 1 | 1 | 0 | 0 | 0 | 0 | 1 | 1 | 0 | 0 | 0 | 0 |
| EYE IRRITATION | 1 | 1 | 0 | 0 | 0 | 0 | 1 | 1 | 0 | 0 | 0 | 0 |
| EYE PRURITUS | 9 | 8 | 0 | 1 | 0 | 0 | 8 | 8 | 0 | 0 | 0 | 0 |
| LIMBAL SWELLING | 1 | 0 | 0 | 1 | 0 | 0 | 1 | 0 | 0 | 1 | 0 | 0 |
| OCULAR HYPERAEMIA | 10 | 6 | 1 | 3 | 0 | 0 | 10 | 6 | 1 | 3 | 0 | 0 |
| ***OCULAR NEUROMUSCULAR DISORDERS*** | | | | | | | | | | | | |
| *EYELID MOVEMENT DISORDERS* | | | | | | | | | | | | |
| BLEPHAROSPASM | 2 | 2 | 0 | 0 | 0 | 0 | 2 | 2 | 0 | 0 | 0 | 0 |
| ***OCULAR SENSORY SYMPTOMS NEC*** | | | | | | | | | | | | |
| *OCULAR SENSATION DISORDERS* | | | | | | | | | | | | |
| ABNORMAL SENSATION IN EYE | 2 | 1 | 1 | 0 | 0 | 0 | 2 | 1 | 1 | 0 | 0 | 0 |
| ASTHENOPIA | 26 | 18 | 3 | 3 | 0 | 2 | 23 | 18 | 1 | 2 | 0 | 2 |
| FOREIGN BODY SENSATION IN EYES | 1 | 1 | 0 | 0 | 0 | 0 | 1 | 1 | 0 | 0 | 0 | 0 |
| PHOTOPHOBIA | 19 | 14 | 3 | 0 | 0 | 2 | 18 | 14 | 2 | 0 | 0 | 2 |
| ***OCULAR STRUCTURAL CHANGE, DEPOSIT AND DEGENERATION NEC*** | | | | | | | | | | | | |
| *CHOROID AND VITREOUS STRUCTURAL CHANGE, DEPOSIT AND DEGENERATION* | | | | | | | | | | | | |
| VITREOUS DETACHMENT | 1 | 1 | 0 | 0 | 0 | 0 | 1 | 1 | 0 | 0 | 0 | 0 |
| VITREOUS FLOATERS | 2 | 2 | 0 | 0 | 0 | 0 | 2 | 2 | 0 | 0 | 0 | 0 |

| | | | | | | | | | | | | |
|---|---|---|---|---|---|---|---|---|---|---|---|---|
| ***RETINA, CHOROID AND VITREOUS HAEMORRHAGES AND VASCULAR DISORDERS*** | | | | | | | | | | | | |
| *RETINAL BLEEDING AND VASCULAR DISORDERS (EXCL RETINOPATHY)* | | | | | | | | | | | | |
| RETINAL VEIN OCCLUSION | 2 | 0 | 2 | 0 | 0 | 0 | 2 | 0 | 2 | 0 | 0 | 0 |
| *RETINOPATHIES NEC* | | | | | | | | | | | | |
| RETINAL EXUDATES | 1 | 1 | 0 | 0 | 0 | 0 | 1 | 1 | 0 | 0 | 0 | 0 |
| ***VISION DISORDERS*** | | | | | | | | | | | | |
| *VISUAL DISORDERS NEC* | | | | | | | | | | | | |
| DIPLOPIA | 2 | 1 | 0 | 0 | 0 | 1 | 2 | 1 | 0 | 0 | 0 | 1 |
| HALO VISION | 2 | 1 | 1 | 0 | 0 | 0 | 2 | 1 | 1 | 0 | 0 | 0 |
| METAMORPHOPSIA | 1 | 1 | 0 | 0 | 0 | 0 | 1 | 1 | 0 | 0 | 0 | 0 |
| PHOTOPSIA | 4 | 2 | 0 | 0 | 0 | 2 | 4 | 2 | 0 | 0 | 0 | 2 |
| VISION BLURRED | 38 | 27 | 9 | 1 | 0 | 1 | 34 | 27 | 7 | 0 | 0 | 0 |
| *VISUAL IMPAIRMENT AND BLINDNESS (EXCL COLOUR BLINDNESS)* | | | | | | | | | | | | |
| BLINDNESS | 1 | 0 | 1 | 0 | 0 | 0 | 1 | 0 | 1 | 0 | 0 | 0 |
| BLINDNESS TRANSIENT | 1 | 1 | 0 | 0 | 0 | 0 | 1 | 1 | 0 | 0 | 0 | 0 |
| SUDDEN VISUAL LOSS | 1 | 1 | 0 | 0 | 0 | 0 | 1 | 1 | 0 | 0 | 0 | 0 |
| VISUAL IMPAIRMENT | 16 | 16 | 0 | 0 | 0 | 0 | 16 | 16 | 0 | 0 | 0 | 0 |
| **GASTROINTESTINAL DISORDERS** | | | | | | | | | | | | |
| ***ANAL AND RECTAL CONDITIONS NEC*** | | | | | | | | | | | | |
| *ANAL AND RECTAL SIGNS AND SYMPTOMS* | | | | | | | | | | | | |
| ANAL PARAESTHESIA | 1 | 1 | 0 | 0 | 0 | 0 | 1 | 1 | 0 | 0 | 0 | 0 |
| ***BENIGN NEOPLASMS GASTROINTESTINAL*** | | | | | | | | | | | | |
| *BENIGN ORAL CAVITY NEOPLASMS* | | | | | | | | | | | | |
| MOUTH CYST | 1 | 1 | 0 | 0 | 0 | 0 | 1 | 1 | 0 | 0 | 0 | 0 |
| ***DENTAL AND GINGIVAL CONDITIONS*** | | | | | | | | | | | | |
| *DENTAL DISORDERS NEC* | 1 | 1 | 0 | 0 | 0 | 0 | 1 | 1 | 0 | 0 | 0 | 0 |
| TEETHING | | | | | | | | | | | | |
| *DENTAL PAIN AND SENSATION DISORDERS* | | | | | | | | | | | | |
| DENTAL PARAESTHESIA | 1 | 1 | 0 | 0 | 0 | 0 | 1 | 1 | 0 | 0 | 0 | 0 |
| TOOTHACHE | 11 | 4 | 5 | 2 | 0 | 0 | 9 | 4 | 3 | 2 | 0 | 0 |
| *GINGIVAL DISORDERS, SIGNS AND SYMPTOMS NEC* | | | | | | | | | | | | |
| GINGIVAL BLISTER | 1 | 1 | 0 | 0 | 0 | 0 | 1 | 1 | 0 | 0 | 0 | 0 |
| GINGIVAL PAIN | 7 | 5 | 2 | 0 | 0 | 0 | 7 | 5 | 2 | 0 | 0 | 0 |
| GINGIVAL SWELLING | 1 | 1 | 0 | 0 | 0 | 0 | 1 | 1 | 0 | 0 | 0 | 0 |
| ***GASTROINTESTINAL CONDITIONS NEC*** | | | | | | | | | | | | |

| | | | | | | | | | | | | |
|---|---|---|---|---|---|---|---|---|---|---|---|---|
| *GASTROINTESTINAL MUCOSAL DYSTROPHIES AND SECRETION DISORDERS* | | | | | | | | | | | | |
| BARRETT'S OESOPHAGUS | 1 | 1 | 0 | 0 | 0 | 0 | 1 | 1 | 0 | 0 | 0 | 0 |
| ***GASTROINTESTINAL HAEMORRHAGES NEC*** | | | | | | | | | | | | |
| *INTESTINAL HAEMORRHAGES* | | | | | | | | | | | | |
| RECTAL HAEMORRHAGE | 1 | 1 | 0 | 0 | 0 | 0 | 1 | 1 | 0 | 0 | 0 | 0 |
| *NON-SITE SPECIFIC GASTROINTESTINAL HAEMORRHAGES* | | | | | | | | | | | | |
| HAEMATEMESIS | 1 | 1 | 0 | 0 | 0 | 0 | 1 | 1 | 0 | 0 | 0 | 0 |
| ***GASTROINTESTINAL INFLAMMATORY CONDITIONS*** | | | | | | | | | | | | |
| *COLITIS (EXCL INFECTIVE)* | | | | | | | | | | | | |
| COLITIS | 1 | 0 | 1 | 0 | 0 | 0 | 1 | 0 | 1 | 0 | 0 | 0 |
| COLITIS MICROSCOPIC | 1 | 0 | 1 | 0 | 0 | 0 | 1 | 0 | 1 | 0 | 0 | 0 |
| COLITIS ULCERATIVE | 2 | 1 | 1 | 0 | 0 | 0 | 1 | 1 | 0 | 0 | 0 | 0 |
| *GASTRITIS (EXCL INFECTIVE)* | | | | | | | | | | | | |
| GASTRITIS | 2 | 2 | 0 | 0 | 0 | 0 | 2 | 2 | 0 | 0 | 0 | 0 |
| REFLUX GASTRITIS | 1 | 1 | 0 | 0 | 0 | 0 | 1 | 1 | 0 | 0 | 0 | 0 |
| *GASTROINTESTINAL INFLAMMATORY DISORDERS NEC* | | | | | | | | | | | | |
| GASTROINTESTINAL TRACT IRRITATION | 1 | 0 | 0 | 1 | 0 | 0 | 1 | 0 | 0 | 1 | 0 | 0 |
| ***GASTROINTESTINAL MOTILITY AND DEFAECATION CONDITIONS*** | | | | | | | | | | | | |
| *DIARRHOEA (EXCL INFECTIVE)* | | | | | | | | | | | | |
| DIARRHOEA | 331 | 234 | 46 | 32 | 6 | 13 | 309 | 234 | 32 | 26 | 5 | 12 |
| *GASTROINTESTINAL ATONIC AND HYPOMOTILITY DISORDERS NEC* | | | | | | | | | | | | |
| CONSTIPATION | 10 | 6 | 2 | 2 | 0 | 0 | 8 | 6 | 1 | 1 | 0 | 0 |
| GASTROOESOPHAGEAL REFLUX DISEASE | 3 | 1 | 1 | 0 | 0 | 1 | 3 | 1 | 1 | 0 | 0 | 1 |
| *GASTROINTESTINAL DYSKINETIC DISORDERS* | | | | | | | | | | | | |
| CHANGE OF BOWEL HABIT | 2 | 1 | 1 | 0 | 0 | 0 | 2 | 1 | 1 | 0 | 0 | 0 |
| *GASTROINTESTINAL SPASTIC AND HYPERMOTILITY DISORDERS* | | | | | | | | | | | | |
| IRRITABLE BOWEL SYNDROME | 3 | 1 | 1 | 1 | 0 | 0 | 3 | 1 | 1 | 1 | 0 | 0 |
| ***GASTROINTESTINAL SIGNS AND SYMPTOMS*** | | | | | | | | | | | | |
| *DYSPEPTIC SIGNS AND SYMPTOMS* | | | | | | | | | | | | |
| DYSPEPSIA | 36 | 24 | 5 | 4 | 1 | 2 | 33 | 24 | 3 | 4 | 1 | 1 |
| ERUCTATION | 3 | 3 | 0 | 0 | 0 | 0 | 3 | 3 | 0 | 0 | 0 | 0 |
| *FAECAL ABNORMALITIES NEC* | | | | | | | | | | | | |
| FAECALOMA | 1 | 0 | 1 | 0 | 0 | 0 | 1 | 0 | 1 | 0 | 0 | 0 |
| FAECES DISCOLOURED | 1 | 1 | 0 | 0 | 0 | 0 | 1 | 1 | 0 | 0 | 0 | 0 |
| FAECES SOFT | 3 | 1 | 1 | 1 | 0 | 0 | 3 | 1 | 1 | 1 | 0 | 0 |
| *FLATULENCE, BLOATING AND DISTENSION* | | | | | | | | | | | | |
| ABDOMINAL DISTENSION | 11 | 9 | 0 | 1 | 1 | 0 | 11 | 9 | 0 | 1 | 1 | 0 |
| FLATULENCE | 18 | 16 | 1 | 1 | 0 | 0 | 17 | 16 | 1 | 0 | 0 | 0 |

| | | | | | | | | | | | | |
|---|---|---|---|---|---|---|---|---|---|---|---|---|
| *GASTROINTESTINAL AND ABDOMINAL PAINS (EXCL ORAL AND THROAT)* | | | | | | | | | | | | |
| ABDOMINAL PAIN | 72 | 54 | 12 | 2 | 1 | 3 | 67 | 54 | 8 | 2 | 0 | 3 |
| ABDOMINAL PAIN LOWER | 4 | 3 | 1 | 0 | 0 | 0 | 3 | 3 | 0 | 0 | 0 | 0 |
| ABDOMINAL PAIN UPPER | 137 | 98 | 21 | 7 | 1 | 10 | 128 | 98 | 14 | 6 | 0 | 10 |
| GASTROINTESTINAL PAIN | 11 | 9 | 0 | 2 | 0 | 0 | 10 | 9 | 0 | 1 | 0 | 0 |
| *GASTROINTESTINAL SIGNS AND SYMPTOMS NEC* | | | | | | | | | | | | |
| ABDOMINAL DISCOMFORT | 78 | 53 | 16 | 5 | 0 | 4 | 73 | 53 | 12 | 5 | 0 | 3 |
| ABDOMINAL SYMPTOM | 1 | 1 | 0 | 0 | 0 | 0 | 1 | 1 | 0 | 0 | 0 | 0 |
| ACUTE ABDOMEN | 1 | 1 | 0 | 0 | 0 | 0 | 1 | 1 | 0 | 0 | 0 | 0 |
| ANAL INCONTINENCE | 1 | 1 | 0 | 0 | 0 | 0 | 1 | 1 | 0 | 0 | 0 | 0 |
| BREATH ODOUR | 1 | 1 | 0 | 0 | 0 | 0 | 1 | 1 | 0 | 0 | 0 | 0 |
| DYSPHAGIA | 2 | 1 | 1 | 0 | 0 | 0 | 1 | 1 | 0 | 0 | 0 | 0 |
| ODYNOPHAGIA | 2 | 1 | 1 | 0 | 0 | 0 | 1 | 1 | 0 | 0 | 0 | 0 |
| *NAUSEA AND VOMITING SYMPTOMS* | | | | | | | | | | | | |
| NAUSEA | 1376 | 993 | 175 | 112 | 17 | 79 | 1307 | 993 | 125 | 98 | 16 | 75 |
| RETCHING | 3 | 3 | 0 | 0 | 0 | 0 | 3 | 3 | 0 | 0 | 0 | 0 |
| VOMITING | 180 | 122 | 20 | 17 | 4 | 17 | 173 | 122 | 18 | 13 | 3 | 17 |
| VOMITING PROJECTILE | 2 | 2 | 0 | 0 | 0 | 0 | 2 | 2 | 0 | 0 | 0 | 0 |
| ***MALABSORPTION CONDITIONS*** | | | | | | | | | | | | |
| *MALABSORPTION SYNDROMES* | | | | | | | | | | | | |
| COELIAC DISEASE | 1 | 0 | 1 | 0 | 0 | 0 | 1 | 0 | 1 | 0 | 0 | 0 |
| ***ORAL SOFT TISSUE CONDITIONS*** | | | | | | | | | | | | |
| *ORAL SOFT TISSUE DISORDERS NEC* | | | | | | | | | | | | |
| CHEILITIS | 1 | 1 | 0 | 0 | 0 | 0 | 1 | 1 | 0 | 0 | 0 | 0 |
| LIP BLISTER | 2 | 2 | 0 | 0 | 0 | 0 | 2 | 2 | 0 | 0 | 0 | 0 |
| ORAL LICHEN PLANUS | 1 | 1 | 0 | 0 | 0 | 0 | 1 | 1 | 0 | 0 | 0 | 0 |
| *ORAL SOFT TISSUE INFECTIONS* | | | | | | | | | | | | |
| ANGULAR CHEILITIS | 1 | 1 | 0 | 0 | 0 | 0 | 1 | 1 | 0 | 0 | 0 | 0 |
| *ORAL SOFT TISSUE SIGNS AND SYMPTOMS* | | | | | | | | | | | | |
| HYPOAESTHESIA ORAL | 8 | 7 | 1 | 0 | 0 | 0 | 7 | 7 | 0 | 0 | 0 | 0 |
| LIP PAIN | 3 | 2 | 1 | 0 | 0 | 0 | 3 | 2 | 1 | 0 | 0 | 0 |
| ORAL DISCOMFORT | 3 | 2 | 1 | 0 | 0 | 0 | 2 | 2 | 0 | 0 | 0 | 0 |
| ORAL MUCOSAL ROUGHENING | 1 | 1 | 0 | 0 | 0 | 0 | 1 | 1 | 0 | 0 | 0 | 0 |
| ORAL PAIN | 10 | 7 | 2 | 1 | 0 | 0 | 9 | 7 | 1 | 1 | 0 | 0 |
| PARAESTHESIA ORAL | 27 | 23 | 3 | 0 | 0 | 1 | 26 | 23 | 2 | 0 | 0 | 1 |
| LIP SWELLING | 11 | 7 | 1 | 1 | 0 | 2 | 11 | 7 | 1 | 1 | 0 | 2 |
| MOUTH SWELLING | 1 | 1 | 0 | 0 | 0 | 0 | 1 | 1 | 0 | 0 | 0 | 0 |
| *STOMATITIS AND ULCERATION* | | | | | | | | | | | | |

| | | | | | | | | | | | | |
|---|---|---|---|---|---|---|---|---|---|---|---|---|
| APHTHOUS ULCER | 1 | 0 | 1 | 0 | 0 | 0 | 0 | 0 | 0 | 0 | 0 | 0 |
| LIP ULCERATION | 1 | 0 | 0 | 0 | 1 | 0 | 0 | 0 | 0 | 0 | 0 | 0 |
| MOUTH ULCERATION | 14 | 8 | 3 | 2 | 1 | 0 | 14 | 8 | 3 | 2 | 1 | 0 |
| STOMATITIS | 2 | 1 | 1 | 0 | 0 | 0 | 2 | 1 | 1 | 0 | 0 | 0 |
| ***SALIVARY GLAND CONDITIONS*** | | | | | | | | | | | | |
| *ORAL DRYNESS AND SALIVA ALTERED* | | | | | | | | | | | | |
| DRY MOUTH | 35 | 20 | 4 | 0 | 4 | 7 | 32 | 20 | 3 | 0 | 2 | 7 |
| LIP DRY | 6 | 3 | 1 | 1 | 0 | 1 | 6 | 3 | 1 | 1 | 0 | 1 |
| SALIVARY HYPOSECRETION | 1 | 1 | 0 | 0 | 0 | 0 | 1 | 1 | 0 | 0 | 0 | 0 |
| ***TONGUE CONDITIONS*** | | | | | | | | | | | | |
| *TONGUE SIGNS AND SYMPTOMS* | | | | | | | | | | | | |
| GLOSSODYNIA | 9 | 6 | 1 | 2 | 0 | 0 | 8 | 6 | 1 | 1 | 0 | 0 |
| SWOLLEN TONGUE | 6 | 4 | 0 | 1 | 0 | 1 | 6 | 4 | 0 | 1 | 0 | 1 |
| TONGUE COATED | 3 | 3 | 0 | 0 | 0 | 0 | 3 | 3 | 0 | 0 | 0 | 0 |
| TONGUE DISCOMFORT | 2 | 1 | 1 | 0 | 0 | 0 | 1 | 1 | 0 | 0 | 0 | 0 |
| TONGUE OEDEMA | 1 | 0 | 0 | 1 | 0 | 0 | 1 | 0 | 0 | 1 | 0 | 0 |
| TONGUE SPASM | 1 | 0 | 0 | 1 | 0 | 0 | 1 | 0 | 0 | 1 | 0 | 0 |
| **GENERAL DISORDERS AND ADMINISTRATION SITE CONDITIONS** | | | | | | | | | | | | |
| ***ADMINISTRATION SITE REACTIONS*** | | | | | | | | | | | | |
| *ADMINISTRATION SITE REACTIONS NEC* | | | | | | | | | | | | |
| ADMINISTRATION SITE BRUISE | 2 | 2 | 0 | 0 | 0 | 0 | 2 | 2 | 0 | 0 | 0 | 0 |
| ADMINISTRATION SITE PAIN | 1 | 1 | 0 | 0 | 0 | 0 | 1 | 1 | 0 | 0 | 0 | 0 |
| PUNCTURE SITE BRUISE | 17 | 9 | 3 | 2 | 0 | 3 | 17 | 9 | 3 | 2 | 0 | 3 |
| PUNCTURE SITE PAIN | 3 | 2 | 0 | 0 | 0 | 1 | 3 | 2 | 0 | 0 | 0 | 1 |
| *APPLICATION SITE REACTIONS* | | | | | | | | | | | | |
| APPLICATION SITE BRUISE | 9 | 5 | 2 | 1 | 1 | 0 | 8 | 5 | 1 | 1 | 1 | 0 |
| APPLICATION SITE ERYTHEMA | 2 | 1 | 1 | 0 | 0 | 0 | 2 | 1 | 1 | 0 | 0 | 0 |
| APPLICATION SITE PAIN | 6 | 2 | 1 | 3 | 0 | 0 | 5 | 2 | 1 | 2 | 0 | 0 |
| *IMPLANT AND CATHETER SITE REACTIONS* | | | | | | | | | | | | |
| IMPLANT SITE PAIN | 2 | 0 | 2 | 0 | 0 | 0 | 1 | 0 | 1 | 0 | 0 | 0 |
| IMPLANT SITE WARMTH | 2 | 2 | 0 | 0 | 0 | 0 | 2 | 2 | 0 | 0 | 0 | 0 |
| *INFUSION SITE REACTIONS* | | | | | | | | | | | | |
| INFUSION SITE PAIN | 2 | 1 | 0 | 1 | 0 | 0 | 2 | 1 | 0 | 1 | 0 | 0 |
| INFUSION SITE SCAB | 1 | 1 | 0 | 0 | 0 | 0 | 1 | 1 | 0 | 0 | 0 | 0 |
| INFUSION SITE WARMTH | 1 | 1 | 0 | 0 | 0 | 0 | 1 | 1 | 0 | 0 | 0 | 0 |
| *INJECTION SITE REACTIONS* | | | | | | | | | | | | |
| INJECTION SITE BRUISING | 8 | 4 | 2 | 2 | 0 | 0 | 8 | 4 | 2 | 2 | 0 | 0 |
| INJECTION SITE DISCOMFORT | 3 | 2 | 1 | 0 | 0 | 0 | 2 | 2 | 0 | 0 | 0 | 0 |

| | | | | | | | | | | | | |
|---|---|---|---|---|---|---|---|---|---|---|---|---|
| INJECTION SITE ERYTHEMA | 32 | 21 | 5 | 3 | 2 | 1 | 30 | 21 | 4 | 3 | 1 | 1 |
| INJECTION SITE HYPERSENSITIVITY | 1 | 0 | 1 | 0 | 0 | 0 | 1 | 0 | 1 | 0 | 0 | 0 |
| INJECTION SITE INFLAMMATION | 2 | 1 | 0 | 0 | 0 | 1 | 2 | 1 | 0 | 0 | 0 | 1 |
| INJECTION SITE INJURY | 1 | 1 | 0 | 0 | 0 | 0 | 1 | 1 | 0 | 0 | 0 | 0 |
| INJECTION SITE IRRITATION | 1 | 0 | 1 | 0 | 0 | 0 | 1 | 0 | 1 | 0 | 0 | 0 |
| INJECTION SITE JOINT PAIN | 3 | 1 | 1 | 0 | 1 | 0 | 1 | 1 | 0 | 0 | 0 | 0 |
| INJECTION SITE MASS | 46 | 29 | 9 | 4 | 2 | 2 | 44 | 29 | 8 | 3 | 2 | 2 |
| INJECTION SITE NODULE | 1 | 0 | 0 | 1 | 0 | 0 | 1 | 0 | 0 | 1 | 0 | 0 |
| INJECTION SITE OEDEMA | 1 | 1 | 0 | 0 | 0 | 0 | 1 | 1 | 0 | 0 | 0 | 0 |
| INJECTION SITE PAIN | 539 | 353 | 92 | 64 | 9 | 21 | 505 | 353 | 72 | 50 | 9 | 21 |
| INJECTION SITE PAPULE | 1 | 1 | 0 | 0 | 0 | 0 | 1 | 1 | 0 | 0 | 0 | 0 |
| INJECTION SITE PARAESTHESIA | 1 | 1 | 0 | 0 | 0 | 0 | 1 | 1 | 0 | 0 | 0 | 0 |
| INJECTION SITE PRURITUS | 16 | 13 | 1 | 2 | 0 | 0 | 15 | 13 | 0 | 2 | 0 | 0 |
| INJECTION SITE RASH | 9 | 7 | 1 | 1 | 0 | 0 | 9 | 7 | 1 | 1 | 0 | 0 |
| INJECTION SITE REACTION | 4 | 2 | 2 | 0 | 0 | 0 | 4 | 2 | 2 | 0 | 0 | 0 |
| INJECTION SITE SCAB | 2 | 1 | 1 | 0 | 0 | 0 | 2 | 1 | 1 | 0 | 0 | 0 |
| INJECTION SITE SWELLING | 18 | 10 | 1 | 5 | 0 | 2 | 18 | 10 | 1 | 5 | 0 | 2 |
| INJECTION SITE URTICARIA | 5 | 0 | 2 | 0 | 3 | 0 | 5 | 0 | 2 | 0 | 3 | 0 |
| INJECTION SITE WARMTH | 16 | 12 | 2 | 1 | 1 | 0 | 16 | 12 | 2 | 1 | 1 | 0 |
| *INSTILLATION SITE REACTIONS* | | | | | | | | | | | | |
| INSTILLATION SITE PRURITUS | 1 | 0 | 1 | 0 | 0 | 0 | 1 | 0 | 1 | 0 | 0 | 0 |
| INSTILLATION SITE WARMTH | 3 | 2 | 0 | 0 | 0 | 1 | 3 | 2 | 0 | 0 | 0 | 1 |
| *VACCINATION SITE REACTIONS* | | | | | | | | | | | | |
| SHOULDER INJURY RELATED TO VACCINE ADMINIST | 1 | 1 | 0 | 0 | 0 | 0 | 1 | 1 | 0 | 0 | 0 | 0 |
| VACCINATION SITE BRUISING | 4 | 1 | 3 | 0 | 0 | 0 | 3 | 1 | 2 | 0 | 0 | 0 |
| VACCINATION SITE DISCOMFORT | 1 | 1 | 0 | 0 | 0 | 0 | 1 | 1 | 0 | 0 | 0 | 0 |
| VACCINATION SITE ERYTHEMA | 2 | 1 | 0 | 0 | 0 | 1 | 2 | 1 | 0 | 0 | 0 | 1 |
| VACCINATION SITE JOINT ERYTHEMA | 1 | 1 | 0 | 0 | 0 | 0 | 1 | 1 | 0 | 0 | 0 | 0 |
| VACCINATION SITE JOINT PAIN | 6 | 0 | 0 | 1 | 0 | 5 | 6 | 0 | 0 | 1 | 0 | 5 |
| VACCINATION SITE MASS | 16 | 11 | 3 | 1 | 0 | 1 | 13 | 11 | 1 | 0 | 0 | 1 |
| VACCINATION SITE PAIN | 87 | 49 | 11 | 12 | 1 | 14 | 83 | 49 | 8 | 12 | 1 | 13 |
| VACCINATION SITE RASH | 2 | 1 | 0 | 0 | 0 | 1 | 2 | 1 | 0 | 0 | 0 | 1 |
| VACCINATION SITE SWELLING | 4 | 1 | 1 | 0 | 0 | 2 | 4 | 1 | 1 | 0 | 0 | 2 |
| VACCINATION SITE WARMTH | 6 | 4 | 1 | 0 | 0 | 1 | 5 | 4 | 0 | 0 | 0 | 1 |
| ***BODY TEMPERATURE CONDITIONS*** | | | | | | | | | | | | |
| *BODY TEMPERATURE ALTERED* | | | | | | | | | | | | |
| HYPERTHERMIA | 1 | 1 | 0 | 0 | 0 | 0 | 1 | 1 | 0 | 0 | 0 | 0 |
| HYPOTHERMIA | 1 | 1 | 0 | 0 | 0 | 0 | 1 | 1 | 0 | 0 | 0 | 0 |

| | | | | | | | | | | | | |
|---|---|---|---|---|---|---|---|---|---|---|---|---|
| *FEBRILE DISORDERS* | | | | | | | | | | | | |
| PYREXIA | 2274 | 1671 | 313 | 160 | 30 | 100 | 2134 | 1671 | 203 | 134 | 27 | 99 |
| ***FATAL OUTCOMES*** | | | | | | | | | | | | |
| *DEATH AND SUDDEN DEATH* | | | | | | | | | | | | |
| DEATH | 3 | 3 | 0 | 0 | 0 | 0 | 3 | 3 | 0 | 0 | 0 | 0 |
| ***GENERAL SYSTEM DISORDERS NEC*** | | | | | | | | | | | | |
| *ADVERSE EFFECT ABSENT* | | | | | | | | | | | | |
| NO ADVERSE EVENT | 1 | 0 | 0 | 0 | 0 | 1 | 1 | 0 | 0 | 0 | 0 | 1 |
| *ASTHENIC CONDITIONS* | | | | | | | | | | | | |
| ASTHENIA | 130 | 89 | 21 | 9 | 2 | 9 | 118 | 89 | 15 | 5 | 1 | 8 |
| CHRONIC FATIGUE SYNDROME | 6 | 2 | 2 | 2 | 0 | 0 | 5 | 2 | 2 | 1 | 0 | 0 |
| DECREASED ACTIVITY | 1 | 1 | 0 | 0 | 0 | 0 | 1 | 1 | 0 | 0 | 0 | 0 |
| FATIGUE | 4468 | 2946 | 833 | 362 | 70 | 257 | 4182 | 2946 | 631 | 302 | 58 | 245 |
| MALAISE | 438 | 286 | 71 | 46 | 11 | 24 | 403 | 286 | 50 | 36 | 8 | 23 |
| SLUGGISHNESS | 2 | 0 | 1 | 0 | 1 | 0 | 2 | 0 | 1 | 0 | 1 | 0 |
| *FEELINGS AND SENSATIONS NEC* | | | | | | | | | | | | |
| CHILLS | 1770 | 1340 | 190 | 144 | 13 | 83 | 1677 | 1340 | 122 | 126 | 10 | 79 |
| FEELING ABNORMAL | 123 | 75 | 22 | 12 | 5 | 9 | 112 | 75 | 14 | 9 | 5 | 9 |
| FEELING COLD | 353 | 258 | 51 | 25 | 7 | 12 | 316 | 258 | 25 | 17 | 5 | 11 |
| FEELING HOT | 89 | 61 | 15 | 8 | 1 | 4 | 85 | 61 | 12 | 8 | 0 | 4 |
| FEELING JITTERY | 1 | 0 | 1 | 0 | 0 | 0 | 1 | 0 | 1 | 0 | 0 | 0 |
| FEELING OF BODY TEMPERATURE CHANGE | 61 | 45 | 9 | 4 | 0 | 3 | 58 | 45 | 6 | 4 | 0 | 3 |
| FEELING OF RELAXATION | 1 | 1 | 0 | 0 | 0 | 0 | 1 | 1 | 0 | 0 | 0 | 0 |
| HANGOVER | 10 | 7 | 0 | 1 | 0 | 2 | 10 | 7 | 0 | 1 | 0 | 2 |
| HUNGER | 7 | 3 | 1 | 1 | 0 | 2 | 6 | 3 | 1 | 0 | 0 | 2 |
| SENSATION OF BLOOD FLOW | 1 | 1 | 0 | 0 | 0 | 0 | 1 | 1 | 0 | 0 | 0 | 0 |
| THIRST | 39 | 32 | 4 | 2 | 0 | 1 | 36 | 32 | 1 | 2 | 0 | 1 |
| *GAIT DISTURBANCES* | | | | | | | | | | | | |
| GAIT DISTURBANCE | 9 | 3 | 3 | 0 | 0 | 3 | 9 | 3 | 3 | 0 | 0 | 3 |
| GAIT INABILITY | 3 | 3 | 0 | 0 | 0 | 0 | 3 | 3 | 0 | 0 | 0 | 0 |
| *GENERAL SIGNS AND SYMPTOMS NEC* | | | | | | | | | | | | |
| CRYING | 1 | 1 | 0 | 0 | 0 | 0 | 1 | 1 | 0 | 0 | 0 | 0 |
| ENERGY INCREASED | 1 | 0 | 1 | 0 | 0 | 0 | 1 | 0 | 1 | 0 | 0 | 0 |
| EXERCISE TOLERANCE DECREASED | 2 | 1 | 0 | 0 | 0 | 1 | 2 | 1 | 0 | 0 | 0 | 1 |
| GENERAL SYMPTOM | 2 | 2 | 0 | 0 | 0 | 0 | 2 | 2 | 0 | 0 | 0 | 0 |
| ILLNESS | 146 | 93 | 22 | 20 | 0 | 11 | 134 | 93 | 14 | 16 | 0 | 11 |
| INFLUENZA LIKE ILLNESS | 665 | 442 | 100 | 77 | 9 | 37 | 599 | 442 | 54 | 60 | 8 | 35 |
| LOCAL REACTION | 5 | 4 | 1 | 0 | 0 | 0 | 5 | 4 | 1 | 0 | 0 | 0 |

| | | | | | | | | | | | | |
|---|---|---|---|---|---|---|---|---|---|---|---|---|
| PERIPHERAL SWELLING | 236 | 139 | 41 | 39 | 5 | 12 | 221 | 139 | 36 | 30 | 4 | 12 |
| SWELLING | 80 | 50 | 13 | 13 | 0 | 4 | 73 | 50 | 10 | 10 | 0 | 3 |
| SWELLING FACE | 14 | 11 | 1 | 0 | 0 | 2 | 14 | 11 | 1 | 0 | 0 | 2 |
| TISSUE IRRITATION | 1 | 0 | 1 | 0 | 0 | 0 | 1 | 0 | 1 | 0 | 0 | 0 |
| *INFLAMMATIONS* | | | | | | | | | | | | |
| INFLAMMATION | 13 | 7 | 6 | 0 | 0 | 0 | 10 | 7 | 3 | 0 | 0 | 0 |
| SYSTEMIC INFLAMMATORY RESPONSE SYNDROME | 1 | 0 | 1 | 0 | 0 | 0 | 0 | 0 | 0 | 0 | 0 | 0 |
| *OEDEMA NEC* | | | | | | | | | | | | |
| OEDEMA | 1 | 1 | 0 | 0 | 0 | 0 | 1 | 1 | 0 | 0 | 0 | 0 |
| OEDEMA PERIPHERAL | 2 | 1 | 1 | 0 | 0 | 0 | 2 | 1 | 1 | 0 | 0 | 0 |
| *PAIN AND DISCOMFORT NEC* | | | | | | | | | | | | |
| AXILLARY PAIN | 90 | 39 | 22 | 27 | 1 | 1 | 86 | 39 | 19 | 26 | 1 | 1 |
| CHEST DISCOMFORT | 39 | 28 | 2 | 7 | 0 | 2 | 38 | 28 | 1 | 7 | 0 | 2 |
| CHEST PAIN | 95 | 64 | 14 | 9 | 2 | 6 | 87 | 64 | 10 | 7 | 1 | 5 |
| DISCOMFORT | 22 | 8 | 5 | 5 | 2 | 2 | 17 | 8 | 3 | 4 | 0 | 2 |
| FACIAL PAIN | 3 | 2 | 0 | 1 | 0 | 0 | 2 | 2 | 0 | 0 | 0 | 0 |
| HERNIA PAIN | 1 | 1 | 0 | 0 | 0 | 0 | 1 | 1 | 0 | 0 | 0 | 0 |
| INFLAMMATORY PAIN | 3 | 2 | 0 | 0 | 1 | 0 | 2 | 2 | 0 | 0 | 0 | 0 |
| NON-CARDIAC CHEST PAIN | 1 | 0 | 0 | 1 | 0 | 0 | 1 | 0 | 0 | 1 | 0 | 0 |
| PAIN | 1075 | 727 | 187 | 97 | 15 | 49 | 994 | 727 | 130 | 78 | 12 | 47 |
| TENDERNESS | 364 | 247 | 54 | 39 | 4 | 20 | 349 | 247 | 44 | 35 | 4 | 19 |
| ***THERAPEUTIC AND NONTHERAPEUTIC EFFECTS (EXCL TOXICITY)*** | | | | | | | | | | | | |
| *THERAPEUTIC AND NONTHERAPEUTIC RESPONSES* | | | | | | | | | | | | |
| ADVERSE DRUG REACTION | 3 | 3 | 0 | 0 | 0 | 0 | 3 | 3 | 0 | 0 | 0 | 0 |
| ADVERSE EVENT | 5 | 0 | 0 | 0 | 0 | 5 | 4 | 0 | 0 | 0 | 0 | 4 |
| ADVERSE REACTION | 1 | 1 | 0 | 0 | 0 | 0 | 1 | 1 | 0 | 0 | 0 | 0 |
| IMMEDIATE POST-INJECTION REACTION | 1 | 0 | 1 | 0 | 0 | 0 | 1 | 0 | 1 | 0 | 0 | 0 |
| **HEPATOBILIARY DISORDERS** | | | | | | | | | | | | |
| ***HEPATIC AND HEPATOBILIARY DISORDERS*** | | | | | | | | | | | | |
| *HEPATOBILIARY SIGNS AND SYMPTOMS* | | | | | | | | | | | | |
| HEPATIC PAIN | 2 | 2 | 0 | 0 | 0 | 0 | 2 | 2 | 0 | 0 | 0 | 0 |
| *HEPATOCELLULAR DAMAGE AND HEPATITIS NEC* | | | | | | | | | | | | |
| LIVER INJURY | 1 | 1 | 0 | 0 | 0 | 0 | 1 | 1 | 0 | 0 | 0 | 0 |
| **IMMUNE SYSTEM DISORDERS** | | | | | | | | | | | | |
| ***ALLERGIC CONDITIONS*** | | | | | | | | | | | | |
| *ALLERGIC CONDITIONS NEC* | | | | | | | | | | | | |
| HYPERSENSITIVITY | 8 | 4 | 3 | 1 | 0 | 0 | 6 | 4 | 1 | 1 | 0 | 0 |
| MULTIPLE ALLERGIES | 1 | 1 | 0 | 0 | 0 | 0 | 1 | 1 | 0 | 0 | 0 | 0 |

| | | | | | | | | | | | | |
|---|---|---|---|---|---|---|---|---|---|---|---|---|
| *ALLERGIES TO FOODS, FOOD ADDITIVES, DRUGS AND OTHER CHEMICALS* | | | | | | | | | | | | |
| ALLERGY TO CHEMICALS | 1 | 0 | 1 | 0 | 0 | 0 | 1 | 0 | 1 | 0 | 0 | 0 |
| *ANAPHYLACTIC AND ANAPHYLACTOID RESPONSES* | | | | | | | | | | | | |
| ANAPHYLACTIC REACTION | 3 | 3 | 0 | 0 | 0 | 0 | 3 | 3 | 0 | 0 | 0 | 0 |
| *ATOPIC DISORDERS* | | | | | | | | | | | | |
| SEASONAL ALLERGY | 1 | 0 | 1 | 0 | 0 | 0 | 1 | 0 | 1 | 0 | 0 | 0 |
| ***IMMUNE DISORDERS NEC*** | | | | | | | | | | | | |
| *IMMUNE AND ASSOCIATED CONDITIONS NEC* | | | | | | | | | | | | |
| BACILLE CALMETTE-GUERIN SCAR REACTIVATION | 1 | 1 | 0 | 0 | 0 | 0 | 1 | 1 | 0 | 0 | 0 | 0 |
| SENSITISATION | 1 | 1 | 0 | 0 | 0 | 0 | 1 | 1 | 0 | 0 | 0 | 0 |
| **INFECTIONS AND INFESTATIONS** | | | | | | | | | | | | |
| ***BACTERIAL INFECTIOUS DISORDERS*** | | | | | | | | | | | | |
| *BACTERIAL INFECTIONS NEC* | | | | | | | | | | | | |
| CELLULITIS | 4 | 3 | 1 | 0 | 0 | 0 | 3 | 3 | 0 | 0 | 0 | 0 |
| PERIORBITAL CELLULITIS | 1 | 0 | 1 | 0 | 0 | 0 | 1 | 0 | 1 | 0 | 0 | 0 |
| *HELICOBACTER INFECTIONS* | | | | | | | | | | | | |
| HELICOBACTER GASTRITIS | 1 | 0 | 0 | 1 | 0 | 0 | 1 | 0 | 0 | 1 | 0 | 0 |
| *STAPHYLOCOCCAL INFECTIONS* | | | | | | | | | | | | |
| FURUNCLE | 4 | 2 | 1 | 0 | 0 | 1 | 3 | 2 | 0 | 0 | 0 | 1 |
| ***FUNGAL INFECTIOUS DISORDERS*** | | | | | | | | | | | | |
| *CANDIDA INFECTIONS* | | | | | | | | | | | | |
| CANDIDA INFECTION | 1 | 1 | 0 | 0 | 0 | 0 | 1 | 1 | 0 | 0 | 0 | 0 |
| VULVOVAGINAL CANDIDIASIS | 2 | 2 | 0 | 0 | 0 | 0 | 2 | 2 | 0 | 0 | 0 | 0 |
| ***INFECTIONS - PATHOGEN UNSPECIFIED*** | | | | | | | | | | | | |
| *ABDOMINAL AND GASTROINTESTINAL INFECTIONS* | | | | | | | | | | | | |
| APPENDICITIS | 1 | 1 | 0 | 0 | 0 | 0 | 1 | 1 | 0 | 0 | 0 | 0 |
| DIARRHOEA INFECTIOUS | 1 | 0 | 0 | 1 | 0 | 0 | 1 | 0 | 0 | 1 | 0 | 0 |
| GASTROINTESTINAL INFECTION | 1 | 1 | 0 | 0 | 0 | 0 | 1 | 1 | 0 | 0 | 0 | 0 |
| *BREAST INFECTIONS* | | | | | | | | | | | | |
| MASTITIS | 3 | 0 | 1 | 2 | 0 | 0 | 3 | 0 | 1 | 2 | 0 | 0 |
| *CENTRAL NERVOUS SYSTEM AND SPINAL INFECTIONS* | | | | | | | | | | | | |
| MYELITIS | 1 | 1 | 0 | 0 | 0 | 0 | 1 | 1 | 0 | 0 | 0 | 0 |
| *DENTAL AND ORAL SOFT TISSUE INFECTIONS* | | | | | | | | | | | | |
| ABSCESS ORAL | 1 | 1 | 0 | 0 | 0 | 0 | 1 | 1 | 0 | 0 | 0 | 0 |
| ORAL PUSTULE | 1 | 1 | 0 | 0 | 0 | 0 | 1 | 1 | 0 | 0 | 0 | 0 |
| PERICORONITIS | 1 | 1 | 0 | 0 | 0 | 0 | 1 | 1 | 0 | 0 | 0 | 0 |
| TOOTH ABSCESS | 1 | 1 | 0 | 0 | 0 | 0 | 1 | 1 | 0 | 0 | 0 | 0 |

| | | | | | | | | | | | | |
|---|---|---|---|---|---|---|---|---|---|---|---|---|
| *EAR INFECTIONS* | | | | | | | | | | | | |
| EAR INFECTION | 2 | 1 | 0 | 0 | 0 | 1 | 2 | 1 | 0 | 0 | 0 | 1 |
| LABYRINTHITIS | 3 | 1 | 2 | 0 | 0 | 0 | 3 | 1 | 2 | 0 | 0 | 0 |
| OTITIS EXTERNA | 1 | 1 | 0 | 0 | 0 | 0 | 1 | 1 | 0 | 0 | 0 | 0 |
| *EYE AND EYELID INFECTIONS* | | | | | | | | | | | | |
| CONJUNCTIVITIS | 2 | 0 | 2 | 0 | 0 | 0 | 1 | 0 | 1 | 0 | 0 | 0 |
| HORDEOLUM | 1 | 1 | 0 | 0 | 0 | 0 | 1 | 1 | 0 | 0 | 0 | 0 |
| *INFECTIONS NEC* | | | | | | | | | | | | |
| ABSCESS | 1 | 1 | 0 | 0 | 0 | 0 | 1 | 1 | 0 | 0 | 0 | 0 |
| INFECTION | 2 | 1 | 0 | 0 | 0 | 1 | 2 | 1 | 0 | 0 | 0 | 1 |
| INJECTION SITE INFECTION | 1 | 1 | 0 | 0 | 0 | 0 | 1 | 1 | 0 | 0 | 0 | 0 |
| LOCALISED INFECTION | 6 | 5 | 0 | 0 | 0 | 1 | 5 | 5 | 0 | 0 | 0 | 0 |
| WOUND INFECTION | 1 | 0 | 1 | 0 | 0 | 0 | 0 | 0 | 0 | 0 | 0 | 0 |
| *LOWER RESPIRATORY TRACT AND LUNG INFECTIONS* | | | | | | | | | | | | |
| LOWER RESPIRATORY TRACT INFECTION | 8 | 4 | 2 | 1 | 0 | 1 | 8 | 4 | 2 | 1 | 0 | 1 |
| PNEUMONIA | 5 | 2 | 1 | 0 | 0 | 2 | 4 | 2 | 0 | 0 | 0 | 2 |
| *MALE REPRODUCTIVE TRACT INFECTIONS* | | | | | | | | | | | | |
| ORCHITIS | 1 | 1 | 0 | 0 | 0 | 0 | 1 | 1 | 0 | 0 | 0 | 0 |
| *SEPSIS, BACTERAEMIA, VIRAEMIA AND FUNGAEMIA NEC* | | | | | | | | | | | | |
| SEPTIC RASH | 1 | 0 | 1 | 0 | 0 | 0 | 0 | 0 | 0 | 0 | 0 | 0 |
| *SKIN STRUCTURES AND SOFT TISSUE INFECTIONS* | | | | | | | | | | | | |
| INFECTED DERMAL CYST | 1 | 1 | 0 | 0 | 0 | 0 | 1 | 1 | 0 | 0 | 0 | 0 |
| INJECTION SITE PUSTULE | 1 | 0 | 1 | 0 | 0 | 0 | 0 | 0 | 0 | 0 | 0 | 0 |
| SKIN INFECTION | 1 | 1 | 0 | 0 | 0 | 0 | 1 | 1 | 0 | 0 | 0 | 0 |
| *UPPER RESPIRATORY TRACT INFECTIONS* | | | | | | | | | | | | |
| LARYNGITIS | 2 | 1 | 1 | 0 | 0 | 0 | 2 | 1 | 1 | 0 | 0 | 0 |
| NASOPHARYNGITIS | 208 | 129 | 37 | 32 | 1 | 9 | 191 | 129 | 26 | 27 | 1 | 8 |
| PHARYNGITIS | 1 | 1 | 0 | 0 | 0 | 0 | 1 | 1 | 0 | 0 | 0 | 0 |
| RHINITIS | 2 | 1 | 0 | 0 | 0 | 1 | 2 | 1 | 0 | 0 | 0 | 1 |
| SINUSITIS | 13 | 9 | 2 | 1 | 0 | 1 | 11 | 9 | 0 | 1 | 0 | 1 |
| TONSILLITIS | 1 | 0 | 0 | 0 | 0 | 1 | 1 | 0 | 0 | 0 | 0 | 1 |
| *URINARY TRACT INFECTIONS* | | | | | | | | | | | | |
| CYSTITIS | 2 | 1 | 1 | 0 | 0 | 0 | 2 | 1 | 1 | 0 | 0 | 0 |
| URINARY TRACT INFECTION | 3 | 2 | 1 | 0 | 0 | 0 | 2 | 2 | 0 | 0 | 0 | 0 |
| ***PROTOZOAL INFECTIOUS DISORDERS*** | | | | | | | | | | | | |
| *TRYPANOSOMAL INFECTIONS* | | | | | | | | | | | | |
| AFRICAN TRYPANOSOMIASIS | 1 | 1 | 0 | 0 | 0 | 0 | 1 | 1 | 0 | 0 | 0 | 0 |
| ***RICKETTSIAL INFECTIOUS DISORDERS*** | | | | | | | | | | | | |

| | | | | | | | | | | | | |
|---|---|---|---|---|---|---|---|---|---|---|---|---|
| *COXIELLA INFECTIONS* | | | | | | | | | | | | |
| Q FEVER | 5 | 5 | 0 | 0 | 0 | 0 | 5 | 5 | 0 | 0 | 0 | 0 |
| ***VIRAL INFECTIOUS DISORDERS*** | | | | | | | | | | | | |
| *CORONAVIRUS INFECTIONS* | | | | | | | | | | | | |
| COVID-19 | 7 | 1 | 1 | 1 | 3 | 1 | 4 | 1 | 0 | 1 | 2 | 0 |
| *HERPES VIRAL INFECTIONS* | | | | | | | | | | | | |
| GENITAL HERPES | 1 | 1 | 0 | 0 | 0 | 0 | 1 | 1 | 0 | 0 | 0 | 0 |
| HERPES OPHTHALMIC | 1 | 1 | 0 | 0 | 0 | 0 | 1 | 1 | 0 | 0 | 0 | 0 |
| HERPES SIMPLEX | 2 | 1 | 1 | 0 | 0 | 0 | 1 | 1 | 0 | 0 | 0 | 0 |
| HERPES ZOSTER | 15 | 8 | 4 | 1 | 0 | 2 | 14 | 8 | 4 | 0 | 0 | 2 |
| NASAL HERPES | 1 | 1 | 0 | 0 | 0 | 0 | 1 | 1 | 0 | 0 | 0 | 0 |
| OPHTHALMIC HERPES ZOSTER | 1 | 1 | 0 | 0 | 0 | 0 | 1 | 1 | 0 | 0 | 0 | 0 |
| ORAL HERPES | 33 | 26 | 5 | 1 | 0 | 1 | 32 | 26 | 4 | 1 | 0 | 1 |
| *INFLUENZA VIRAL INFECTIONS* | | | | | | | | | | | | |
| H2N2 INFLUENZA | 1 | 1 | 0 | 0 | 0 | 0 | 1 | 1 | 0 | 0 | 0 | 0 |
| INFLUENZA | 668 | 464 | 91 | 82 | 9 | 22 | 614 | 464 | 61 | 60 | 8 | 21 |
| *RETROVIRAL INFECTIONS* | | | | | | | | | | | | |
| AIDS RELATED COMPLEX | 1 | 1 | 0 | 0 | 0 | 0 | 1 | 1 | 0 | 0 | 0 | 0 |
| *VIRAL INFECTIONS NEC* | | | | | | | | | | | | |
| GASTROENTERITIS VIRAL | 4 | 4 | 0 | 0 | 0 | 0 | 4 | 4 | 0 | 0 | 0 | 0 |
| SWEATING FEVER | 10 | 8 | 0 | 0 | 0 | 2 | 10 | 8 | 0 | 0 | 0 | 2 |
| VESTIBULAR NEURONITIS | 1 | 0 | 0 | 0 | 1 | 0 | 0 | 0 | 0 | 0 | 0 | 0 |
| VIRAL DIARRHOEA | 1 | 1 | 0 | 0 | 0 | 0 | 1 | 1 | 0 | 0 | 0 | 0 |
| VIRAL RASH | 2 | 1 | 0 | 0 | 1 | 0 | 2 | 1 | 0 | 0 | 1 | 0 |
| VIRAL UPPER RESPIRATORY TRACT INFECTION | 1 | 1 | 0 | 0 | 0 | 0 | 1 | 1 | 0 | 0 | 0 | 0 |
| **INJURY, POISONING AND PROCEDURAL COMPLICATIONS** | | | | | | | | | | | | |
| ***BONE AND JOINT INJURIES*** | | | | | | | | | | | | |
| *FRACTURES AND DISLOCATIONS NEC* | | | | | | | | | | | | |
| JOINT DISLOCATION | 1 | 1 | 0 | 0 | 0 | 0 | 1 | 1 | 0 | 0 | 0 | 0 |
| ***EXPOSURES, CHEMICAL INJURIES AND POISONING*** | | | | | | | | | | | | |
| *POISONING AND TOXICITY* | | | | | | | | | | | | |
| SYSTEMIC TOXICITY | 1 | 1 | 0 | 0 | 0 | 0 | 1 | 1 | 0 | 0 | 0 | 0 |
| TOXICITY TO VARIOUS AGENTS | 2 | 2 | 0 | 0 | 0 | 0 | 2 | 2 | 0 | 0 | 0 | 0 |
| ***INJURIES BY PHYSICAL AGENTS*** | | | | | | | | | | | | |
| *CONDITIONS CAUSED BY COLD* | | | | | | | | | | | | |
| CHILLBLAINS | 3 | 2 | 0 | 0 | 0 | 1 | 3 | 2 | 0 | 0 | 0 | 1 |
| *HEAT INJURIES (EXCL THERMAL BURNS)* | | | | | | | | | | | | |
| HEAT EXHAUSTION | 1 | 1 | 0 | 0 | 0 | 0 | 1 | 1 | 0 | 0 | 0 | 0 |

| | | | | | | | | | | | | |
|---|---|---|---|---|---|---|---|---|---|---|---|---|
| HEAT OEDEMA | 1 | 1 | 0 | 0 | 0 | 0 | 1 | 1 | 0 | 0 | 0 | 0 |
| *RADIATION INJURIES* | | | | | | | | | | | | |
| SUNBURN | 1 | 0 | 1 | 0 | 0 | 0 | 0 | 0 | 0 | 0 | 0 | 0 |
| *THERMAL BURNS* | | | | | | | | | | | | |
| THERMAL BURN | 1 | 1 | 0 | 0 | 0 | 0 | 1 | 1 | 0 | 0 | 0 | 0 |
| THERMAL BURNS OF EYE | 1 | 1 | 0 | 0 | 0 | 0 | 1 | 1 | 0 | 0 | 0 | 0 |
| ***INJURIES NEC*** | | | | | | | | | | | | |
| *CHEST AND RESPIRATORY TRACT INJURIES NEC* | | | | | | | | | | | | |
| CHEST CRUSHING | 1 | 1 | 0 | 0 | 0 | 0 | 1 | 1 | 0 | 0 | 0 | 0 |
| *EYE INJURIES NEC* | | | | | | | | | | | | |
| EYE CONTUSION | 1 | 1 | 0 | 0 | 0 | 0 | 1 | 1 | 0 | 0 | 0 | 0 |
| *MUSCLE, TENDON AND LIGAMENT INJURIES* | | | | | | | | | | | | |
| LIGAMENT SPRAIN | 1 | 1 | 0 | 0 | 0 | 0 | 1 | 1 | 0 | 0 | 0 | 0 |
| MUSCLE INJURY | 2 | 1 | 0 | 1 | 0 | 0 | 1 | 1 | 0 | 0 | 0 | 0 |
| MUSCLE STRAIN | 2 | 1 | 1 | 0 | 0 | 0 | 2 | 1 | 1 | 0 | 0 | 0 |
| *NERVE INJURIES NEC* | | | | | | | | | | | | |
| NERVE INJURY | 1 | 1 | 0 | 0 | 0 | 0 | 1 | 1 | 0 | 0 | 0 | 0 |
| *NON-SITE SPECIFIC INJURIES NEC* | | | | | | | | | | | | |
| ARTHROPOD STING | 2 | 2 | 0 | 0 | 0 | 0 | 2 | 2 | 0 | 0 | 0 | 0 |
| BITE | 1 | 0 | 1 | 0 | 0 | 0 | 1 | 0 | 1 | 0 | 0 | 0 |
| FALL | 5 | 4 | 0 | 1 | 0 | 0 | 5 | 4 | 0 | 1 | 0 | 0 |
| INFLAMMATION OF WOUND | 2 | 2 | 0 | 0 | 0 | 0 | 2 | 2 | 0 | 0 | 0 | 0 |
| TISSUE INJURY | 1 | 1 | 0 | 0 | 0 | 0 | 1 | 1 | 0 | 0 | 0 | 0 |
| WOUND COMPLICATION | 1 | 1 | 0 | 0 | 0 | 0 | 1 | 1 | 0 | 0 | 0 | 0 |
| WOUND SECRETION | 1 | 1 | 0 | 0 | 0 | 0 | 1 | 1 | 0 | 0 | 0 | 0 |
| *SITE SPECIFIC INJURIES NEC* | | | | | | | | | | | | |
| LIMB INJURY | 6 | 6 | 0 | 0 | 0 | 0 | 6 | 6 | 0 | 0 | 0 | 0 |
| *SKIN INJURIES NEC* | | | | | | | | | | | | |
| CONTUSION | 145 | 85 | 27 | 18 | 3 | 12 | 132 | 85 | 18 | 15 | 3 | 11 |
| SCAR | 2 | 1 | 1 | 0 | 0 | 0 | 2 | 1 | 1 | 0 | 0 | 0 |
| SKIN WOUND | 1 | 0 | 1 | 0 | 0 | 0 | 0 | 0 | 0 | 0 | 0 | 0 |
| ***PROCEDURAL RELATED INJURIES AND COMPLICATIONS NEC*** | | | | | | | | | | | | |
| *CARDIAC AND VASCULAR PROCEDURAL COMPLICATIONS* | | | | | | | | | | | | |
| CARDIAC PROCEDURE COMPLICATION | 1 | 0 | 1 | 0 | 0 | 0 | 0 | 0 | 0 | 0 | 0 | 0 |
| *GASTROINTESTINAL AND HEPATOBILIARY PROCEDURAL COMPLICATIONS* | | | | | | | | | | | | |
| PROCEDURAL NAUSEA | 6 | 6 | 0 | 0 | 0 | 0 | 6 | 6 | 0 | 0 | 0 | 0 |
| *NEUROLOGICAL AND PSYCHIATRIC PROCEDURAL COMPLICATIONS* | | | | | | | | | | | | |
| PROCEDURAL DIZZINESS | 2 | 2 | 0 | 0 | 0 | 0 | 2 | 2 | 0 | 0 | 0 | 0 |

| | | | | | | | | | | | | |
|---|---|---|---|---|---|---|---|---|---|---|---|---|
| *NON-SITE SPECIFIC PROCEDURAL COMPLICATIONS* | | | | | | | | | | | | |
| INCISION SITE PAIN | 1 | 1 | 0 | 0 | 0 | 0 | 1 | 1 | 0 | 0 | 0 | 0 |
| INCISION SITE SWELLING | 1 | 0 | 1 | 0 | 0 | 0 | 1 | 0 | 1 | 0 | 0 | 0 |
| INJECTION RELATED REACTION | 49 | 41 | 3 | 2 | 3 | 0 | 49 | 41 | 3 | 2 | 3 | 0 |
| POST PROCEDURAL COMPLICATION | 10 | 6 | 2 | 1 | 1 | 0 | 6 | 6 | 0 | 0 | 0 | 0 |
| PROCEDURAL PAIN | 1 | 0 | 0 | 0 | 0 | 1 | 1 | 0 | 0 | 0 | 0 | 1 |
| *VACCINATION RELATED COMPLICATIONS* | | | | | | | | | | | | |
| IMMUNISATION REACTION | 8 | 3 | 4 | 1 | 0 | 0 | 7 | 3 | 3 | 1 | 0 | 0 |
| **INVESTIGATIONS** | | | | | | | | | | | | |
| ***CARDIAC AND VASCULAR INVESTIGATIONS (EXCL ENZYME TESTS)*** | | | | | | | | | | | | |
| *HEART RATE AND PULSE INVESTIGATIONS* | | | | | | | | | | | | |
| HEART RATE | 29 | 21 | 3 | 2 | 0 | 3 | 29 | 21 | 3 | 2 | 0 | 3 |
| HEART RATE DECREASED | 3 | 2 | 0 | 0 | 0 | 1 | 3 | 2 | 0 | 0 | 0 | 1 |
| HEART RATE INCREASED | 42 | 34 | 3 | 2 | 0 | 3 | 40 | 34 | 1 | 2 | 0 | 3 |
| HEART RATE IRREGULAR | 2 | 0 | 1 | 1 | 0 | 0 | 2 | 0 | 1 | 1 | 0 | 0 |
| *VASCULAR TESTS NEC (INCL BLOOD PRESSURE)* | | | | | | | | | | | | |
| BLOOD PRESSURE DECREASED | 1 | 0 | 1 | 0 | 0 | 0 | 1 | 0 | 1 | 0 | 0 | 0 |
| BLOOD PRESSURE INCREASED | 7 | 5 | 1 | 1 | 0 | 0 | 7 | 5 | 1 | 1 | 0 | 0 |
| BLOOD PRESSURE MEASUREMENT | 3 | 1 | 0 | 1 | 0 | 1 | 2 | 1 | 0 | 0 | 0 | 1 |
| ***ENDOCRINE INVESTIGATIONS (INCL SEX HORMONES)*** | | | | | | | | | | | | |
| *ENDOCRINE ANALYSES AND IMAGING NEC* | | | | | | | | | | | | |
| HORMONE LEVEL ABNORMAL | 1 | 1 | 0 | 0 | 0 | 0 | 1 | 1 | 0 | 0 | 0 | 0 |
| *PITUITARY ANALYSES ANTERIOR* | | | | | | | | | | | | |
| BLOOD FOLLICLE STIMULATING HORMONE INCREA | 1 | 0 | 0 | 1 | 0 | 0 | 1 | 0 | 0 | 1 | 0 | 0 |
| BLOOD LUTEINISING HORMONE | 1 | 1 | 0 | 0 | 0 | 0 | 1 | 1 | 0 | 0 | 0 | 0 |
| *THYROID ANALYSES* | | | | | | | | | | | | |
| TRI-IODOTHYRONINE | 1 | 1 | 0 | 0 | 0 | 0 | 1 | 1 | 0 | 0 | 0 | 0 |
| ***HAEMATOLOGY INVESTIGATIONS (INCL BLOOD GROUPS)*** | | | | | | | | | | | | |
| *COAGULATION AND BLEEDING ANALYSES* | | | | | | | | | | | | |
| BLEEDING TIME | 1 | 0 | 0 | 0 | 0 | 1 | 1 | 0 | 0 | 0 | 0 | 1 |
| INTERNATIONAL NORMALISED RATIO DECREASED | 1 | 0 | 1 | 0 | 0 | 0 | 0 | 0 | 0 | 0 | 0 | 0 |
| *PLATELET ANALYSES* | | | | | | | | | | | | |
| PLATELET COUNT INCREASED | 2 | 0 | 1 | 0 | 0 | 1 | 2 | 0 | 1 | 0 | 0 | 1 |
| *RED BLOOD CELL ANALYSES* | | | | | | | | | | | | |
| HAEMOGLOBIN | 1 | 1 | 0 | 0 | 0 | 0 | 1 | 1 | 0 | 0 | 0 | 0 |
| ***METABOLIC, NUTRITIONAL AND BLOOD GAS INVESTIGATIONS*** | | | | | | | | | | | | |
| *BLOOD GAS AND ACID BASE ANALYSES* | | | | | | | | | | | | |
| OXYGEN SATURATION DECREASED | 2 | 2 | 0 | 0 | 0 | 0 | 2 | 2 | 0 | 0 | 0 | 0 |

| | | | | | | | | | | | | |
|---|---|---|---|---|---|---|---|---|---|---|---|---|
| *CARBOHYDRATE TOLERANCE ANALYSES (INCL DIABETES)* | | | | | | | | | | | | |
| BLOOD GLUCOSE | 3 | 3 | 0 | 0 | 0 | 0 | 3 | 3 | 0 | 0 | 0 | 0 |
| BLOOD GLUCOSE ABNORMAL | 1 | 1 | 0 | 0 | 0 | 0 | 1 | 1 | 0 | 0 | 0 | 0 |
| BLOOD GLUCOSE DECREASED | 1 | 1 | 0 | 0 | 0 | 0 | 1 | 1 | 0 | 0 | 0 | 0 |
| BLOOD GLUCOSE INCREASED | 6 | 6 | 0 | 0 | 0 | 0 | 6 | 6 | 0 | 0 | 0 | 0 |
| ***MICROBIOLOGY AND SEROLOGY INVESTIGATIONS*** | | | | | | | | | | | | |
| *VIRUS IDENTIFICATION AND SEROLOGY* | | | | | | | | | | | | |
| SARS-COV-2 TEST | 1 | 1 | 0 | 0 | 0 | 0 | 1 | 1 | 0 | 0 | 0 | 0 |
| SARS-COV-2 TEST POSITIVE | 1 | 0 | 0 | 1 | 0 | 0 | 1 | 0 | 0 | 1 | 0 | 0 |
| ***MUSCULOSKELETAL AND SOFT TISSUE INVESTIGATIONS (EXCL ENZYME TESTS)*** | | | | | | | | | | | | |
| *MUSCULOSKELETAL AND SOFT TISSUE IMAGING PROCEDURES* | | | | | | | | | | | | |
| BONE SCAN | 1 | 1 | 0 | 0 | 0 | 0 | 1 | 1 | 0 | 0 | 0 | 0 |
| ***NEUROLOGICAL, SPECIAL SENSES AND PSYCHIATRIC INVESTIGATIONS*** | | | | | | | | | | | | |
| *CENTRAL NERVOUS SYSTEM IMAGING PROCEDURES* | | | | | | | | | | | | |
| MAGNETIC RESONANCE IMAGING HEAD | 8 | 8 | 0 | 0 | 0 | 0 | 8 | 8 | 0 | 0 | 0 | 0 |
| SCAN BRAIN | 1 | 0 | 0 | 1 | 0 | 0 | 1 | 0 | 0 | 1 | 0 | 0 |
| ***PHYSICAL EXAMINATION AND ORGAN SYSTEM STATUS TOPICS*** | | | | | | | | | | | | |
| *PHYSICAL EXAMINATION PROCEDURES AND ORGAN SYSTEM STATUS* | | | | | | | | | | | | |
| BODY TEMPERATURE | 64 | 43 | 10 | 2 | 0 | 9 | 62 | 43 | 8 | 2 | 0 | 9 |
| BODY TEMPERATURE ABNORMAL | 3 | 2 | 0 | 1 | 0 | 0 | 3 | 2 | 0 | 1 | 0 | 0 |
| BODY TEMPERATURE DECREASED | 2 | 2 | 0 | 0 | 0 | 0 | 2 | 2 | 0 | 0 | 0 | 0 |
| BODY TEMPERATURE FLUCTUATION | 10 | 7 | 2 | 1 | 0 | 0 | 10 | 7 | 2 | 1 | 0 | 0 |
| BODY TEMPERATURE INCREASED | 64 | 43 | 9 | 10 | 1 | 1 | 59 | 43 | 6 | 9 | 0 | 1 |
| GRIP STRENGTH DECREASED | 1 | 1 | 0 | 0 | 0 | 0 | 1 | 1 | 0 | 0 | 0 | 0 |
| HEAD LAG | 2 | 2 | 0 | 0 | 0 | 0 | 2 | 2 | 0 | 0 | 0 | 0 |
| LEFT-HANDEDNESS | 1 | 1 | 0 | 0 | 0 | 0 | 1 | 1 | 0 | 0 | 0 | 0 |
| LYMPH NODE PALPABLE | 1 | 0 | 0 | 1 | 0 | 0 | 1 | 0 | 0 | 1 | 0 | 0 |
| RESPIRATORY RATE DECREASED | 1 | 0 | 0 | 0 | 0 | 1 | 1 | 0 | 0 | 0 | 0 | 1 |
| SKIN TEMPERATURE | 5 | 3 | 1 | 1 | 0 | 0 | 5 | 3 | 1 | 1 | 0 | 0 |
| WEIGHT DECREASED | 3 | 3 | 0 | 0 | 0 | 0 | 3 | 3 | 0 | 0 | 0 | 0 |
| WEIGHT INCREASED | 3 | 2 | 0 | 0 | 0 | 1 | 3 | 2 | 0 | 0 | 0 | 1 |
| ***RENAL AND URINARY TRACT INVESTIGATIONS AND URINALYSES*** | | | | | | | | | | | | |
| *URINALYSIS NEC* | | | | | | | | | | | | |
| BLOOD URINE | 2 | 2 | 0 | 0 | 0 | 0 | 2 | 2 | 0 | 0 | 0 | 0 |
| NITRITE URINE PRESENT | 1 | 1 | 0 | 0 | 0 | 0 | 1 | 1 | 0 | 0 | 0 | 0 |
| PH URINE | 2 | 1 | 0 | 0 | 0 | 1 | 2 | 1 | 0 | 0 | 0 | 1 |
| *URINARY TRACT FUNCTION ANALYSES NEC* | | | | | | | | | | | | |
| URINE OUTPUT | 3 | 2 | 0 | 1 | 0 | 0 | 3 | 2 | 0 | 1 | 0 | 0 |

| | | | | | | | | | | | | |
|---|---|---|---|---|---|---|---|---|---|---|---|---|
| URINE OUTPUT INCREASED | 1 | 1 | 0 | 0 | 0 | 0 | 1 | 1 | 0 | 0 | 0 | 0 |
| ***REPRODUCTIVE ORGAN AND BREAST INVESTIGATIONS (EXCL HORMONE ANALYSES)*** | | | | | | | | | | | | |
| *REPRODUCTIVE ORGAN AND BREAST IMAGING PROCEDURES* | | | | | | | | | | | | |
| BREAST SCAN | 1 | 0 | 1 | 0 | 0 | 0 | 1 | 0 | 1 | 0 | 0 | 0 |
| ***RESPIRATORY AND PULMONARY INVESTIGATIONS (EXCL BLOOD GASES)*** | | | | | | | | | | | | |
| *RESPIRATORY AND PULMONARY FUNCTION DIAGNOSTIC PROCEDURES* | | | | | | | | | | | | |
| FORCED EXPIRATORY VOLUME DECREASED | 1 | 1 | 0 | 0 | 0 | 0 | 1 | 1 | 0 | 0 | 0 | 0 |
| FORCED EXPIRATORY VOLUME INCREASED | 1 | 0 | 1 | 0 | 0 | 0 | 1 | 0 | 1 | 0 | 0 | 0 |
| ***WATER, ELECTROLYTE AND MINERAL INVESTIGATIONS*** | | | | | | | | | | | | |
| *WATER AND ELECTROLYTE ANALYSES NEC* | | | | | | | | | | | | |
| VOLUME BLOOD | 1 | 0 | 1 | 0 | 0 | 0 | 1 | 0 | 1 | 0 | 0 | 0 |
| **METABOLISM AND NUTRITION DISORDERS** | | | | | | | | | | | | |
| ***APPETITE AND GENERAL NUTRITIONAL DISORDERS*** | | | | | | | | | | | | |
| *APPETITE DISORDERS* | | | | | | | | | | | | |
| APPETITE DISORDER | 1 | 0 | 0 | 0 | 0 | 1 | 1 | 0 | 0 | 0 | 0 | 1 |
| DECREASED APPETITE | 154 | 113 | 14 | 9 | 3 | 15 | 146 | 113 | 8 | 9 | 2 | 14 |
| FOOD CRAVING | 1 | 1 | 0 | 0 | 0 | 0 | 1 | 1 | 0 | 0 | 0 | 0 |
| FOOD REFUSAL | 2 | 1 | 0 | 1 | 0 | 0 | 1 | 1 | 0 | 0 | 0 | 0 |
| INCREASED APPETITE | 2 | 1 | 0 | 0 | 0 | 1 | 2 | 1 | 0 | 0 | 0 | 1 |
| *GENERAL NUTRITIONAL DISORDERS NEC* | | | | | | | | | | | | |
| FOOD AVERSION | 2 | 2 | 0 | 0 | 0 | 0 | 2 | 2 | 0 | 0 | 0 | 0 |
| ***ELECTROLYTE AND FLUID BALANCE CONDITIONS*** | | | | | | | | | | | | |
| *TOTAL FLUID VOLUME DECREASED* | | | | | | | | | | | | |
| DEHYDRATION | 14 | 13 | 1 | 0 | 0 | 0 | 13 | 13 | 0 | 0 | 0 | 0 |
| *TOTAL FLUID VOLUME INCREASED* | | | | | | | | | | | | |
| FLUID RETENTION | 1 | 0 | 0 | 1 | 0 | 0 | 0 | 0 | 0 | 0 | 0 | 0 |
| ***FOOD INTOLERANCE SYNDROMES*** | | | | | | | | | | | | |
| *FOOD MALABSORPTION AND INTOLERANCE SYNDROMES (EXCL SUGAR INTOLERANCE)* | | | | | | | | | | | | |
| ALCOHOL INTOLERANCE | 1 | 1 | 0 | 0 | 0 | 0 | 1 | 1 | 0 | 0 | 0 | 0 |
| ***GLUCOSE METABOLISM DISORDERS (INCL DIABETES MELLITUS)*** | | | | | | | | | | | | |
| *DIABETES MELLITUS (INCL SUBTYPES)* | | | | | | | | | | | | |
| DIABETES MELLITUS | 1 | 1 | 0 | 0 | 0 | 0 | 1 | 1 | 0 | 0 | 0 | 0 |
| DIABETES MELLITUS INADEQUATE CONTROL | 3 | 2 | 0 | 0 | 0 | 1 | 3 | 2 | 0 | 0 | 0 | 1 |
| *HYPERGLYCAEMIC CONDITIONS NEC* | | | | | | | | | | | | |
| HYPERGLYCAEMIA | 1 | 1 | 0 | 0 | 0 | 0 | 1 | 1 | 0 | 0 | 0 | 0 |
| *HYPOGLYCAEMIC CONDITIONS NEC* | | | | | | | | | | | | |
| HYPOGLYCAEMIA | 3 | 3 | 0 | 0 | 0 | 0 | 3 | 3 | 0 | 0 | 0 | 0 |
| ***PURINE AND PYRIMIDINE METABOLISM DISORDERS*** | | | | | | | | | | | | |

| | | | | | | | | | | | | |
|---|---|---|---|---|---|---|---|---|---|---|---|---|
| *DISORDERS OF PURINE METABOLISM* | | | | | | | | | | | | |
| GOUT | 1 | 1 | 0 | 0 | 0 | 0 | 1 | 1 | 0 | 0 | 0 | 0 |
| **MUSCULOSKELETAL AND CONNECTIVE TISSUE DISORDERS** | | | | | | | | | | | | |
| ***BONE DISORDERS (EXCL CONGENITAL AND FRACTURES)*** | | | | | | | | | | | | |
| *BONE DISORDERS NEC* | | | | | | | | | | | | |
| JAW DISORDER | 1 | 1 | 0 | 0 | 0 | 0 | 1 | 1 | 0 | 0 | 0 | 0 |
| OSTEITIS | 1 | 1 | 0 | 0 | 0 | 0 | 1 | 1 | 0 | 0 | 0 | 0 |
| *BONE RELATED SIGNS AND SYMPTOMS* | | | | | | | | | | | | |
| BONE PAIN | 16 | 12 | 1 | 1 | 1 | 1 | 14 | 12 | 0 | 1 | 0 | 1 |
| BONE SWELLING | 1 | 0 | 0 | 0 | 0 | 1 | 1 | 0 | 0 | 0 | 0 | 1 |
| PAIN IN JAW | 15 | 10 | 4 | 1 | 0 | 0 | 15 | 10 | 4 | 1 | 0 | 0 |
| PUBIC PAIN | 1 | 1 | 0 | 0 | 0 | 0 | 1 | 1 | 0 | 0 | 0 | 0 |
| SPINAL PAIN | 3 | 2 | 1 | 0 | 0 | 0 | 3 | 2 | 1 | 0 | 0 | 0 |
| ***CONNECTIVE TISSUE DISORDERS (EXCL CONGENITAL)*** | | | | | | | | | | | | |
| *CONNECTIVE TISSUE DISORDERS NEC* | | | | | | | | | | | | |
| POLYMYALGIA RHEUMATICA | 4 | 1 | 1 | 1 | 1 | 0 | 4 | 1 | 1 | 1 | 1 | 0 |
| *LUPUS ERYTHEMATOSUS (INCL SUBTYPES)* | | | | | | | | | | | | |
| SYSTEMIC LUPUS ERYTHEMATOSUS | 1 | 1 | 0 | 0 | 0 | 0 | 1 | 1 | 0 | 0 | 0 | 0 |
| ***JOINT DISORDERS*** | | | | | | | | | | | | |
| *ARTHROPATHIES NEC* | | | | | | | | | | | | |
| ARTHRITIS | 13 | 7 | 3 | 2 | 0 | 1 | 12 | 7 | 3 | 2 | 0 | 0 |
| ARTHROPATHY | 2 | 1 | 1 | 0 | 0 | 0 | 2 | 1 | 1 | 0 | 0 | 0 |
| POLYARTHRITIS | 1 | 0 | 1 | 0 | 0 | 0 | 1 | 0 | 1 | 0 | 0 | 0 |
| RHEUMATIC FEVER | 1 | 1 | 0 | 0 | 0 | 0 | 1 | 1 | 0 | 0 | 0 | 0 |
| *JOINT RELATED DISORDERS NEC* | | | | | | | | | | | | |
| JOINT LOCK | 2 | 1 | 0 | 0 | 0 | 1 | 2 | 1 | 0 | 0 | 0 | 1 |
| PERIARTHRITIS | 6 | 2 | 2 | 2 | 0 | 0 | 6 | 2 | 2 | 2 | 0 | 0 |
| TEMPOROMANDIBULAR PAIN AND DYSFUNCTION S | 1 | 1 | 0 | 0 | 0 | 0 | 1 | 1 | 0 | 0 | 0 | 0 |
| *JOINT RELATED SIGNS AND SYMPTOMS* | | | | | | | | | | | | |
| ARTHRALGIA | 1380 | 956 | 208 | 136 | 20 | 60 | 1285 | 956 | 149 | 111 | 15 | 54 |
| JAW CLICKING | 1 | 1 | 0 | 0 | 0 | 0 | 1 | 1 | 0 | 0 | 0 | 0 |
| JOINT STIFFNESS | 39 | 29 | 4 | 3 | 1 | 2 | 35 | 29 | 2 | 2 | 1 | 1 |
| JOINT SWELLING | 12 | 7 | 2 | 3 | 0 | 0 | 10 | 7 | 2 | 1 | 0 | 0 |
| *OSTEOARTHROPATHIES* | | | | | | | | | | | | |
| OSTEOARTHRITIS | 2 | 1 | 1 | 0 | 0 | 0 | 1 | 1 | 0 | 0 | 0 | 0 |
| *PSORIATIC ARTHROPATHIES* | | | | | | | | | | | | |
| PSORIATIC ARTHROPATHY | 1 | 1 | 0 | 0 | 0 | 0 | 1 | 1 | 0 | 0 | 0 | 0 |
| *RHEUMATOID ARTHROPATHIES* | | | | | | | | | | | | |

| | | | | | | | | | | | | |
|---|---|---|---|---|---|---|---|---|---|---|---|---|
| RHEUMATOID ARTHRITIS | 3 | 3 | 0 | 0 | 0 | 0 | 3 | 3 | 0 | 0 | 0 | 0 |
| ***MUSCLE DISORDERS*** | | | | | | | | | | | | |
| *MUSCLE INFECTIONS AND INFLAMMATIONS* | | | | | | | | | | | | |
| MYOSITIS | 1 | 0 | 1 | 0 | 0 | 0 | 1 | 0 | 1 | 0 | 0 | 0 |
| *MUSCLE PAINS* | | | | | | | | | | | | |
| FIBROMYALGIA | 4 | 2 | 0 | 2 | 0 | 0 | 4 | 2 | 0 | 2 | 0 | 0 |
| MYALGIA | 1820 | 1294 | 280 | 150 | 31 | 65 | 1697 | 1294 | 201 | 114 | 27 | 61 |
| *MUSCLE RELATED SIGNS AND SYMPTOMS NEC* | | | | | | | | | | | | |
| MUSCLE ATROPHY | 2 | 1 | 0 | 0 | 0 | 1 | 2 | 1 | 0 | 0 | 0 | 1 |
| MUSCLE DISCOMFORT | 1 | 1 | 0 | 0 | 0 | 0 | 1 | 1 | 0 | 0 | 0 | 0 |
| MUSCLE FATIGUE | 52 | 39 | 7 | 4 | 0 | 2 | 49 | 39 | 5 | 3 | 0 | 2 |
| MUSCLE FIBROSIS | 1 | 0 | 0 | 0 | 0 | 1 | 1 | 0 | 0 | 0 | 0 | 1 |
| MUSCLE MASS | 3 | 3 | 0 | 0 | 0 | 0 | 3 | 3 | 0 | 0 | 0 | 0 |
| MUSCLE SPASMS | 59 | 34 | 12 | 4 | 1 | 8 | 54 | 34 | 8 | 3 | 1 | 8 |
| MUSCLE TIGHTNESS | 3 | 2 | 1 | 0 | 0 | 0 | 3 | 2 | 1 | 0 | 0 | 0 |
| MUSCLE TWITCHING | 9 | 8 | 0 | 0 | 0 | 1 | 9 | 8 | 0 | 0 | 0 | 1 |
| *MUSCLE TONE ABNORMALITIES* | | | | | | | | | | | | |
| MUSCLE RIGIDITY | 1 | 1 | 0 | 0 | 0 | 0 | 1 | 1 | 0 | 0 | 0 | 0 |
| TRISMUS | 2 | 1 | 1 | 0 | 0 | 0 | 2 | 1 | 1 | 0 | 0 | 0 |
| *MUSCLE WEAKNESS CONDITIONS* | | | | | | | | | | | | |
| MUSCULAR WEAKNESS | 57 | 41 | 10 | 3 | 0 | 3 | 55 | 41 | 9 | 2 | 0 | 3 |
| ***MUSCULOSKELETAL AND CONNECTIVE TISSUE DISORDERS NEC*** | | | | | | | | | | | | |
| *MUSCULOSKELETAL AND CONNECTIVE TISSUE CONDITIONS NEC* | | | | | | | | | | | | |
| MOBILITY DECREASED | 3 | 1 | 1 | 0 | 1 | 0 | 2 | 1 | 0 | 0 | 1 | 0 |
| MUSCULOSKELETAL STIFFNESS | 222 | 149 | 34 | 30 | 6 | 3 | 207 | 149 | 28 | 22 | 5 | 3 |
| BACK PAIN | 194 | 133 | 29 | 19 | 0 | 13 | 181 | 133 | 22 | 14 | 0 | 12 |
| FLANK PAIN | 1 | 1 | 0 | 0 | 0 | 0 | 1 | 1 | 0 | 0 | 0 | 0 |
| LIMB DISCOMFORT | 623 | 399 | 121 | 73 | 2 | 28 | 595 | 399 | 102 | 64 | 2 | 28 |
| MUSCULOSKELETAL CHEST PAIN | 10 | 4 | 2 | 0 | 0 | 4 | 10 | 4 | 2 | 0 | 0 | 4 |
| MUSCULOSKELETAL DISCOMFORT | 17 | 10 | 3 | 4 | 0 | 0 | 16 | 10 | 2 | 4 | 0 | 0 |
| MUSCULOSKELETAL PAIN | 5 | 1 | 3 | 1 | 0 | 0 | 5 | 1 | 3 | 1 | 0 | 0 |
| NECK PAIN | 109 | 71 | 20 | 10 | 2 | 6 | 107 | 71 | 19 | 9 | 2 | 6 |
| PAIN IN EXTREMITY | 3757 | 2409 | 599 | 476 | 75 | 198 | 3502 | 2409 | 453 | 400 | 58 | 182 |
| *SOFT TISSUE DISORDERS NEC* | | | | | | | | | | | | |
| AXILLARY MASS | 1 | 0 | 0 | 1 | 0 | 0 | 0 | 0 | 0 | 0 | 0 | 0 |
| GROIN PAIN | 5 | 3 | 1 | 0 | 0 | 1 | 5 | 3 | 1 | 0 | 0 | 1 |
| ***SYNOVIAL AND BURSAL DISORDERS*** | | | | | | | | | | | | |
| *SYNOVIAL DISORDERS* | | | | | | | | | | | | |

| | | | | | | | | | | | | |
|---|---|---|---|---|---|---|---|---|---|---|---|---|
| SYNOVITIS | 1 | 0 | 1 | 0 | 0 | 0 | 1 | 0 | 1 | 0 | 0 | 0 |
| ***TENDON, LIGAMENT AND CARTILAGE DISORDERS*** | | | | | | | | | | | | |
| *CARTILAGE DISORDERS* | | | | | | | | | | | | |
| COSTOCHONDRITIS | 2 | 2 | 0 | 0 | 0 | 0 | 2 | 2 | 0 | 0 | 0 | 0 |
| *TENDON DISORDERS* | | | | | | | | | | | | |
| TENDONITIS | 1 | 1 | 0 | 0 | 0 | 0 | 1 | 1 | 0 | 0 | 0 | 0 |
| TENOSYNOVITIS | 1 | 0 | 1 | 0 | 0 | 0 | 1 | 0 | 1 | 0 | 0 | 0 |
| TRIGGER FINGER | 1 | 0 | 0 | 1 | 0 | 0 | 1 | 0 | 0 | 1 | 0 | 0 |
| **NEOPLASMS BENIGN, MALIGNANT AND UNSPECIFIED (INCL CYSTS AND POLYPS)** | | | | | | | | | | | | |
| ***CUTANEOUS NEOPLASMS BENIGN*** | | | | | | | | | | | | |
| *SKIN NEOPLASMS BENIGN* | | | | | | | | | | | | |
| MELANOCYTIC NAEVUS | 1 | 0 | 0 | 0 | 0 | 1 | 1 | 0 | 0 | 0 | 0 | 1 |
| SEBORRHOEIC KERATOSIS | 1 | 0 | 0 | 1 | 0 | 0 | 1 | 0 | 0 | 1 | 0 | 0 |
| SKIN PAPILLOMA | 1 | 1 | 0 | 0 | 0 | 0 | 1 | 1 | 0 | 0 | 0 | 0 |
| **NERVOUS SYSTEM DISORDERS** | | | | | | | | | | | | |
| ***CENTRAL NERVOUS SYSTEM INFECTIONS AND INFLAMMATIONS*** | | | | | | | | | | | | |
| *MYELITIS (INCL INFECTIVE)* | | | | | | | | | | | | |
| MYELITIS TRANSVERSE | 1 | 1 | 0 | 0 | 0 | 0 | 1 | 1 | 0 | 0 | 0 | 0 |
| ***CENTRAL NERVOUS SYSTEM VASCULAR DISORDERS*** | | | | | | | | | | | | |
| *CENTRAL NERVOUS SYSTEM HAEMORRHAGES AND CEREBROVASCULAR ACCIDENTS* | | | | | | | | | | | | |
| CEREBRAL HAEMORRHAGE | 1 | 0 | 1 | 0 | 0 | 0 | 1 | 0 | 1 | 0 | 0 | 0 |
| CEREBROVASCULAR ACCIDENT | 6 | 4 | 1 | 1 | 0 | 0 | 6 | 4 | 1 | 1 | 0 | 0 |
| *TRANSIENT CEREBROVASCULAR EVENTS* | | | | | | | | | | | | |
| TRANSIENT ISCHAEMIC ATTACK | 5 | 4 | 1 | 0 | 0 | 0 | 4 | 4 | 0 | 0 | 0 | 0 |
| ***CRANIAL NERVE DISORDERS (EXCL NEOPLASMS)*** | | | | | | | | | | | | |
| *FACIAL CRANIAL NERVE DISORDERS* | | | | | | | | | | | | |
| BELL'S PALSY | 2 | 1 | 1 | 0 | 0 | 0 | 2 | 1 | 1 | 0 | 0 | 0 |
| FACIAL PARALYSIS | 3 | 3 | 0 | 0 | 0 | 0 | 3 | 3 | 0 | 0 | 0 | 0 |
| FACIAL PARESIS | 1 | 0 | 0 | 1 | 0 | 0 | 1 | 0 | 0 | 1 | 0 | 0 |
| *OLFACTORY NERVE DISORDERS* | | | | | | | | | | | | |
| ANOSMIA | 14 | 10 | 1 | 1 | 0 | 2 | 14 | 10 | 1 | 1 | 0 | 2 |
| HYPOSMIA | 1 | 1 | 0 | 0 | 0 | 0 | 1 | 1 | 0 | 0 | 0 | 0 |
| PAROSMIA | 16 | 9 | 6 | 1 | 0 | 0 | 15 | 9 | 5 | 1 | 0 | 0 |
| *TRIGEMINAL DISORDERS* | | | | | | | | | | | | |
| TRIGEMINAL NEURALGIA | 3 | 3 | 0 | 0 | 0 | 0 | 3 | 3 | 0 | 0 | 0 | 0 |
| TRIGEMINAL NEURITIS | 1 | 0 | 0 | 1 | 0 | 0 | 1 | 0 | 0 | 1 | 0 | 0 |
| ***DEMYELINATING DISORDERS*** | | | | | | | | | | | | |
| *MULTIPLE SCLEROSIS ACUTE AND PROGRESSIVE* | | | | | | | | | | | | |

| | | | | | | | | | | | | |
|---|---|---|---|---|---|---|---|---|---|---|---|---|
| MULTIPLE SCLEROSIS RELAPSE | 1 | 1 | 0 | 0 | 0 | 0 | 1 | 1 | 0 | 0 | 0 | 0 |
| ***HEADACHES*** | | | | | | | | | | | | |
| *HEADACHES NEC* | | | | | | | | | | | | |
| CLUSTER HEADACHE | 29 | 19 | 3 | 2 | 1 | 4 | 27 | 19 | 1 | 2 | 1 | 4 |
| COLD-STIMULUS HEADACHE | 1 | 1 | 0 | 0 | 0 | 0 | 1 | 1 | 0 | 0 | 0 | 0 |
| DRUG WITHDRAWAL HEADACHE | 1 | 0 | 1 | 0 | 0 | 0 | 1 | 0 | 1 | 0 | 0 | 0 |
| HEADACHE | 5317 | 3764 | 792 | 413 | 55 | 293 | 4969 | 3764 | 547 | 337 | 47 | 274 |
| PRIMARY STABBING HEADACHE | 3 | 3 | 0 | 0 | 0 | 0 | 3 | 3 | 0 | 0 | 0 | 0 |
| SINUS HEADACHE | 68 | 50 | 11 | 4 | 2 | 1 | 62 | 50 | 8 | 3 | 0 | 1 |
| TENSION HEADACHE | 126 | 87 | 22 | 11 | 4 | 2 | 119 | 87 | 17 | 9 | 4 | 2 |
| THUNDERCLAP HEADACHE | 1 | 1 | 0 | 0 | 0 | 0 | 1 | 1 | 0 | 0 | 0 | 0 |
| VASCULAR HEADACHE | 2 | 2 | 0 | 0 | 0 | 0 | 2 | 2 | 0 | 0 | 0 | 0 |
| *MIGRAINE HEADACHES* | | | | | | | | | | | | |
| MIGRAINE | 154 | 113 | 22 | 13 | 0 | 6 | 144 | 113 | 15 | 10 | 0 | 6 |
| MIGRAINE WITH AURA | 8 | 5 | 3 | 0 | 0 | 0 | 7 | 5 | 2 | 0 | 0 | 0 |
| RETINAL MIGRAINE | 3 | 3 | 0 | 0 | 0 | 0 | 3 | 3 | 0 | 0 | 0 | 0 |
| TYPICAL AURA WITHOUT HEADACHE | 1 | 0 | 1 | 0 | 0 | 0 | 1 | 0 | 1 | 0 | 0 | 0 |
| ***MENTAL IMPAIRMENT DISORDERS*** | | | | | | | | | | | | |
| *INTELLECTUAL DISABILITIES* | | | | | | | | | | | | |
| INTELLECTUAL DISABILITY | 2 | 0 | 0 | 2 | 0 | 0 | 2 | 0 | 0 | 2 | 0 | 0 |
| *MEMORY LOSS (EXCL DEMENTIA)* | | | | | | | | | | | | |
| AMNESIA | 4 | 2 | 1 | 0 | 0 | 1 | 4 | 2 | 1 | 0 | 0 | 1 |
| MEMORY IMPAIRMENT | 10 | 7 | 2 | 1 | 0 | 0 | 9 | 7 | 1 | 1 | 0 | 0 |
| *MENTAL IMPAIRMENT (EXCL DEMENTIA AND MEMORY LOSS)* | | | | | | | | | | | | |
| COGNITIVE DISORDER | 3 | 2 | 0 | 1 | 0 | 0 | 3 | 2 | 0 | 1 | 0 | 0 |
| DISTURBANCE IN ATTENTION | 16 | 12 | 3 | 0 | 0 | 1 | 16 | 12 | 3 | 0 | 0 | 1 |
| MENTAL IMPAIRMENT | 4 | 3 | 0 | 1 | 0 | 0 | 3 | 3 | 0 | 0 | 0 | 0 |
| ***MOVEMENT DISORDERS (INCL PARKINSONISM)*** | | | | | | | | | | | | |
| *DYSKINESIAS AND MOVEMENT DISORDERS NEC* | | | | | | | | | | | | |
| BRADYKINESIA | 2 | 1 | 0 | 1 | 0 | 0 | 2 | 1 | 0 | 1 | 0 | 0 |
| CLUMSINESS | 1 | 1 | 0 | 0 | 0 | 0 | 1 | 1 | 0 | 0 | 0 | 0 |
| DYSKINESIA | 3 | 3 | 0 | 0 | 0 | 0 | 3 | 3 | 0 | 0 | 0 | 0 |
| EXTRAPYRAMIDAL DISORDER | 1 | 1 | 0 | 0 | 0 | 0 | 1 | 1 | 0 | 0 | 0 | 0 |
| PSYCHOMOTOR HYPERACTIVITY | 1 | 0 | 1 | 0 | 0 | 0 | 1 | 0 | 1 | 0 | 0 | 0 |
| *PARALYSIS AND PARESIS (EXCL CRANIAL NERVE)* | | | | | | | | | | | | |
| HEMIPLEGIA | 2 | 0 | 0 | 1 | 0 | 1 | 2 | 0 | 0 | 1 | 0 | 1 |
| MONOPARESIS | 4 | 2 | 0 | 1 | 0 | 1 | 2 | 2 | 0 | 0 | 0 | 0 |
| MONOPLEGIA | 1 | 1 | 0 | 0 | 0 | 0 | 1 | 1 | 0 | 0 | 0 | 0 |

| | | | | | | | | | | | | |
|---|---|---|---|---|---|---|---|---|---|---|---|---|
| PARALYSIS | 4 | 2 | 0 | 1 | 0 | 1 | 4 | 2 | 0 | 1 | 0 | 1 |
| *PARKINSON'S DISEASE AND PARKINSONISM* | | | | | | | | | | | | |
| FREEZING PHENOMENON | 3 | 2 | 0 | 1 | 0 | 0 | 3 | 2 | 0 | 1 | 0 | 0 |
| PARKINSON'S DISEASE | 1 | 1 | 0 | 0 | 0 | 0 | 1 | 1 | 0 | 0 | 0 | 0 |
| *TREMOR (EXCL CONGENITAL)* | | | | | | | | | | | | |
| TREMOR | 162 | 113 | 16 | 10 | 1 | 22 | 151 | 113 | 9 | 7 | 1 | 21 |
| ***NEUROLOGICAL DISORDERS NEC*** | | | | | | | | | | | | |
| *COMA STATES* | | | | | | | | | | | | |
| DIABETIC HYPERGLYCAEMIC COMA | 1 | 0 | 0 | 0 | 0 | 1 | 0 | 0 | 0 | 0 | 0 | 0 |
| *COORDINATION AND BALANCE DISTURBANCES* | | | | | | | | | | | | |
| BALANCE DISORDER | 26 | 17 | 3 | 2 | 1 | 3 | 26 | 17 | 3 | 2 | 1 | 3 |
| COORDINATION ABNORMAL | 2 | 1 | 1 | 0 | 0 | 0 | 2 | 1 | 1 | 0 | 0 | 0 |
| DYSSTASIA | 1 | 1 | 0 | 0 | 0 | 0 | 1 | 1 | 0 | 0 | 0 | 0 |
| VESTIBULAR NYSTAGMUS | 1 | 1 | 0 | 0 | 0 | 0 | 1 | 1 | 0 | 0 | 0 | 0 |
| *DISTURBANCES IN CONSCIOUSNESS NEC* | | | | | | | | | | | | |
| DEPRESSED LEVEL OF CONSCIOUSNESS | 1 | 0 | 0 | 0 | 0 | 1 | 1 | 0 | 0 | 0 | 0 | 1 |
| LETHARGY | 322 | 204 | 62 | 31 | 6 | 19 | 296 | 204 | 47 | 26 | 2 | 17 |
| LOSS OF CONSCIOUSNESS | 11 | 7 | 2 | 1 | 0 | 1 | 11 | 7 | 2 | 1 | 0 | 1 |
| SEDATION | 1 | 0 | 0 | 0 | 1 | 0 | 1 | 0 | 0 | 0 | 1 | 0 |
| SOMNOLENCE | 146 | 95 | 30 | 9 | 0 | 12 | 138 | 95 | 26 | 7 | 0 | 10 |
| SYNCOPE | 41 | 24 | 10 | 3 | 0 | 4 | 34 | 24 | 4 | 2 | 0 | 4 |
| *NERVOUS SYSTEM DISORDERS NEC* | | | | | | | | | | | | |
| NERVOUS SYSTEM DISORDER | 1 | 0 | 0 | 0 | 0 | 1 | 1 | 0 | 0 | 0 | 0 | 1 |
| *NEUROLOGICAL SIGNS AND SYMPTOMS NEC* | | | | | | | | | | | | |
| AGITATION NEONATAL | 1 | 0 | 0 | 0 | 0 | 1 | 0 | 0 | 0 | 0 | 0 | 0 |
| BRAIN FOG | 33 | 21 | 6 | 1 | 0 | 5 | 32 | 21 | 5 | 1 | 0 | 5 |
| DIZZINESS | 655 | 443 | 125 | 38 | 9 | 40 | 617 | 443 | 98 | 31 | 6 | 39 |
| DIZZINESS EXERTIONAL | 7 | 5 | 2 | 0 | 0 | 0 | 6 | 5 | 1 | 0 | 0 | 0 |
| DIZZINESS POSTURAL | 93 | 65 | 18 | 4 | 1 | 5 | 87 | 65 | 13 | 4 | 1 | 4 |
| HEAD DISCOMFORT | 17 | 11 | 4 | 1 | 0 | 1 | 16 | 11 | 4 | 1 | 0 | 0 |
| INFANT IRRITABILITY | 1 | 0 | 1 | 0 | 0 | 0 | 1 | 0 | 1 | 0 | 0 | 0 |
| MENINGISM | 1 | 1 | 0 | 0 | 0 | 0 | 1 | 1 | 0 | 0 | 0 | 0 |
| MYOCLONUS | 1 | 1 | 0 | 0 | 0 | 0 | 1 | 1 | 0 | 0 | 0 | 0 |
| NEUROLOGICAL SYMPTOM | 1 | 1 | 0 | 0 | 0 | 0 | 1 | 1 | 0 | 0 | 0 | 0 |
| PERSISTENT POSTURAL-PERCEPTUAL DIZZINESS | 1 | 0 | 1 | 0 | 0 | 0 | 0 | 0 | 0 | 0 | 0 | 0 |
| PRESYNCOPE | 17 | 14 | 0 | 3 | 0 | 0 | 17 | 14 | 0 | 3 | 0 | 0 |
| *PARAESTHESIAS AND DYSAESTHESIAS* | | | | | | | | | | | | |
| BURNING FEET SYNDROME | 1 | 1 | 0 | 0 | 0 | 0 | 1 | 1 | 0 | 0 | 0 | 0 |

| | | | | | | | | | | | | |
|---|---|---|---|---|---|---|---|---|---|---|---|---|
| BURNING SENSATION | 7 | 3 | 0 | 0 | 0 | 4 | 7 | 3 | 0 | 0 | 0 | 4 |
| FORMICATION | 1 | 0 | 1 | 0 | 0 | 0 | 1 | 0 | 1 | 0 | 0 | 0 |
| HYPERAESTHESIA | 1 | 1 | 0 | 0 | 0 | 0 | 1 | 1 | 0 | 0 | 0 | 0 |
| HYPOAESTHESIA | 112 | 96 | 8 | 5 | 0 | 3 | 107 | 96 | 4 | 5 | 0 | 2 |
| PARAESTHESIA | 157 | 114 | 26 | 4 | 2 | 11 | 152 | 114 | 22 | 3 | 2 | 11 |
| REVERSED HOT-COLD SENSATION | 1 | 1 | 0 | 0 | 0 | 0 | 1 | 1 | 0 | 0 | 0 | 0 |
| *SENSORY ABNORMALITIES NEC* | | | | | | | | | | | | |
| AGEUSIA | 32 | 21 | 5 | 1 | 1 | 4 | 31 | 21 | 4 | 1 | 1 | 4 |
| ALLODYNIA | 2 | 1 | 1 | 0 | 0 | 0 | 1 | 1 | 0 | 0 | 0 | 0 |
| DYSGEUSIA | 70 | 49 | 14 | 5 | 1 | 1 | 67 | 49 | 11 | 5 | 1 | 1 |
| HYPOGEUSIA | 1 | 1 | 0 | 0 | 0 | 0 | 1 | 1 | 0 | 0 | 0 | 0 |
| NEURALGIA | 25 | 18 | 5 | 1 | 0 | 1 | 22 | 18 | 3 | 0 | 0 | 1 |
| POST HERPETIC NEURALGIA | 1 | 0 | 0 | 1 | 0 | 0 | 1 | 0 | 0 | 1 | 0 | 0 |
| RESTLESS ARM SYNDROME | 1 | 1 | 0 | 0 | 0 | 0 | 1 | 1 | 0 | 0 | 0 | 0 |
| RESTLESS LEGS SYNDROME | 5 | 4 | 0 | 0 | 0 | 1 | 5 | 4 | 0 | 0 | 0 | 1 |
| SENSORY LOSS | 2 | 2 | 0 | 0 | 0 | 0 | 2 | 2 | 0 | 0 | 0 | 0 |
| TASTE DISORDER | 17 | 10 | 5 | 1 | 0 | 1 | 16 | 10 | 4 | 1 | 0 | 1 |
| *SPEECH AND LANGUAGE ABNORMALITIES* | | | | | | | | | | | | |
| DYSARTHRIA | 1 | 1 | 0 | 0 | 0 | 0 | 1 | 1 | 0 | 0 | 0 | 0 |
| SPEECH DISORDER DEVELOPMENTAL | 1 | 0 | 0 | 1 | 0 | 0 | 1 | 0 | 0 | 1 | 0 | 0 |
| ***NEUROLOGICAL DISORDERS OF THE EYE*** | | | | | | | | | | | | |
| *NEUROLOGIC VISUAL PROBLEMS NEC* | | | | | | | | | | | | |
| TUNNEL VISION | 2 | 1 | 1 | 0 | 0 | 0 | 2 | 1 | 1 | 0 | 0 | 0 |
| ***NEUROMUSCULAR DISORDERS*** | | | | | | | | | | | | |
| *MUSCLE TONE ABNORMAL* | | | | | | | | | | | | |
| HYPOTONIA | 1 | 1 | 0 | 0 | 0 | 0 | 1 | 1 | 0 | 0 | 0 | 0 |
| STIFF LEG SYNDROME | 1 | 0 | 1 | 0 | 0 | 0 | 1 | 0 | 1 | 0 | 0 | 0 |
| *NEUROMUSCULAR DISORDERS NEC* | | | | | | | | | | | | |
| MUSCLE SPASTICITY | 1 | 1 | 0 | 0 | 0 | 0 | 1 | 1 | 0 | 0 | 0 | 0 |
| ***PERIPHERAL NEUROPATHIES*** | | | | | | | | | | | | |
| *ACUTE POLYNEUROPATHIES* | | | | | | | | | | | | |
| GUILLAIN-BARRE SYNDROME | 2 | 2 | 0 | 0 | 0 | 0 | 2 | 2 | 0 | 0 | 0 | 0 |
| *PERIPHERAL NEUROPATHIES NEC* | | | | | | | | | | | | |
| AXONAL NEUROPATHY | 1 | 1 | 0 | 0 | 0 | 0 | 1 | 1 | 0 | 0 | 0 | 0 |
| NEUROPATHY PERIPHERAL | 3 | 1 | 0 | 2 | 0 | 0 | 3 | 1 | 0 | 2 | 0 | 0 |
| ***SEIZURES (INCL SUBTYPES)*** | | | | | | | | | | | | |
| *SEIZURES AND SEIZURE DISORDERS NEC* | | | | | | | | | | | | |
| EPILEPSY | 1 | 0 | 0 | 1 | 0 | 0 | 1 | 0 | 0 | 1 | 0 | 0 |

| | | | | | | | | | | | | |
|---|---|---|---|---|---|---|---|---|---|---|---|---|
| SEIZURE | 10 | 8 | 1 | 1 | 0 | 0 | 10 | 8 | 1 | 1 | 0 | 0 |
| ***SLEEP DISTURBANCES (INCL SUBTYPES)*** | | | | | | | | | | | | |
| *DISTURBANCES IN SLEEP PHASE RHYTHM* | | | | | | | | | | | | |
| CIRCADIAN RHYTHM SLEEP DISORDER | 1 | 1 | 0 | 0 | 0 | 0 | 1 | 1 | 0 | 0 | 0 | 0 |
| *SLEEP DISTURBANCES NEC* | | | | | | | | | | | | |
| SLEEP DEFICIT | 2 | 2 | 0 | 0 | 0 | 0 | 2 | 2 | 0 | 0 | 0 | 0 |
| ***SPINAL CORD AND NERVE ROOT DISORDERS*** | | | | | | | | | | | | |
| *CERVICAL SPINAL CORD AND NERVE ROOT DISORDERS* | | | | | | | | | | | | |
| CERVICOBRACHIAL SYNDROME | 1 | 0 | 0 | 1 | 0 | 0 | 1 | 0 | 0 | 1 | 0 | 0 |
| *LUMBAR SPINAL CORD AND NERVE ROOT DISORDERS* | | | | | | | | | | | | |
| CAUDA EQUINA SYNDROME | 1 | 0 | 0 | 1 | 0 | 0 | 1 | 0 | 0 | 1 | 0 | 0 |
| SCIATICA | 8 | 4 | 1 | 2 | 0 | 1 | 8 | 4 | 1 | 2 | 0 | 1 |
| **PREGNANCY, PUERPERIUM AND PERINATAL CONDITIONS** | | | | | | | | | | | | |
| ***ABORTIONS AND STILLBIRTH*** | | | | | | | | | | | | |
| *ABORTIONS SPONTANEOUS* | | | | | | | | | | | | |
| ABORTION SPONTANEOUS | 23 | 13 | 10 | 0 | 0 | 0 | 20 | 13 | 7 | 0 | 0 | 0 |
| *STILLBIRTH AND FOETAL DEATH* | | | | | | | | | | | | |
| FOETAL DEATH | 1 | 1 | 0 | 0 | 0 | 0 | 1 | 1 | 0 | 0 | 0 | 0 |
| ***FOETAL COMPLICATIONS*** | | | | | | | | | | | | |
| *FOETAL COMPLICATIONS NEC* | | | | | | | | | | | | |
| FOETAL DISORDER | 1 | 1 | 0 | 0 | 0 | 0 | 1 | 1 | 0 | 0 | 0 | 0 |
| FOETAL HYPOKINESIA | 2 | 1 | 0 | 1 | 0 | 0 | 1 | 1 | 0 | 0 | 0 | 0 |
| *FOETAL GROWTH COMPLICATIONS* | | | | | | | | | | | | |
| FOETAL MACROSOMIA | 1 | 1 | 0 | 0 | 0 | 0 | 1 | 1 | 0 | 0 | 0 | 0 |
| ***MATERNAL COMPLICATIONS OF PREGNANCY*** | | | | | | | | | | | | |
| *MATERNAL COMPLICATIONS OF PREGNANCY NEC* | | | | | | | | | | | | |
| MORNING SICKNESS | 3 | 2 | 0 | 0 | 0 | 1 | 2 | 2 | 0 | 0 | 0 | 0 |
| ***PLACENTAL, AMNIOTIC AND CAVITY DISORDERS (EXCL HAEMORRHAGES)*** | | | | | | | | | | | | |
| *PLACENTAL ABNORMALITIES (EXCL NEOPLASMS)* | | | | | | | | | | | | |
| PLACENTAL INFARCTION | 1 | 1 | 0 | 0 | 0 | 0 | 1 | 1 | 0 | 0 | 0 | 0 |
| ***PREGNANCY, LABOUR, DELIVERY AND POSTPARTUM CONDITIONS*** | | | | | | | | | | | | |
| *NORMAL PREGNANCY, LABOUR AND DELIVERY* | | | | | | | | | | | | |
| PREGNANCY | 1 | 0 | 0 | 0 | 0 | 1 | 0 | 0 | 0 | 0 | 0 | 0 |
| UTERINE CONTRACTIONS DURING PREGNANCY | 1 | 1 | 0 | 0 | 0 | 0 | 1 | 1 | 0 | 0 | 0 | 0 |
| **PRODUCT ISSUES** | | | | | | | | | | | | |
| ***DEVICE ISSUES*** | | | | | | | | | | | | |
| *DEVICE MALFUNCTION EVENTS NEC* | | | | | | | | | | | | |
| OVERSENSING | 1 | 1 | 0 | 0 | 0 | 0 | 1 | 1 | 0 | 0 | 0 | 0 |

| | | | | | | | | | | | | |
|---|---|---|---|---|---|---|---|---|---|---|---|---|
| **PSYCHIATRIC DISORDERS** | | | | | | | | | | | | |
| ***ANXIETY DISORDERS AND SYMPTOMS*** | | | | | | | | | | | | |
| *ANXIETY SYMPTOMS* | | | | | | | | | | | | |
| AGITATION | 3 | 1 | 2 | 0 | 0 | 0 | 3 | 1 | 2 | 0 | 0 | 0 |
| ANXIETY | 23 | 13 | 4 | 2 | 0 | 4 | 22 | 13 | 3 | 2 | 0 | 4 |
| NERVOUSNESS | 14 | 11 | 1 | 0 | 0 | 2 | 14 | 11 | 1 | 0 | 0 | 2 |
| TENSION | 6 | 5 | 1 | 0 | 0 | 0 | 5 | 5 | 0 | 0 | 0 | 0 |
| *PANIC ATTACKS AND DISORDERS* | | | | | | | | | | | | |
| PANIC ATTACK | 4 | 3 | 0 | 0 | 0 | 1 | 4 | 3 | 0 | 0 | 0 | 1 |
| ***CHANGES IN PHYSICAL ACTIVITY*** | | | | | | | | | | | | |
| *INCREASED PHYSICAL ACTIVITY LEVELS* | | | | | | | | | | | | |
| RESTLESSNESS | 9 | 6 | 1 | 0 | 1 | 1 | 9 | 6 | 1 | 0 | 1 | 1 |
| *STEREOTYPIES AND AUTOMATISMS* | | | | | | | | | | | | |
| BRUXISM | 1 | 1 | 0 | 0 | 0 | 0 | 1 | 1 | 0 | 0 | 0 | 0 |
| ***COGNITIVE AND ATTENTION DISORDERS AND DISTURBANCES*** | | | | | | | | | | | | |
| *COGNITIVE AND ATTENTION DISORDERS AND DISTURBANCES NEC* | | | | | | | | | | | | |
| DAYDREAMING | 1 | 1 | 0 | 0 | 0 | 0 | 1 | 1 | 0 | 0 | 0 | 0 |
| MENTAL FATIGUE | 30 | 19 | 6 | 2 | 0 | 3 | 26 | 19 | 3 | 1 | 0 | 3 |
| ***COMMUNICATION DISORDERS AND DISTURBANCES*** | | | | | | | | | | | | |
| *SPEECH ARTICULATION AND RHYTHM DISTURBANCES* | | | | | | | | | | | | |
| DYSPHEMIA | 1 | 0 | 1 | 0 | 0 | 0 | 0 | 0 | 0 | 0 | 0 | 0 |
| ***DELIRIA (INCL CONFUSION)*** | | | | | | | | | | | | |
| *CONFUSION AND DISORIENTATION* | | | | | | | | | | | | |
| CONFUSIONAL STATE | 34 | 27 | 4 | 2 | 1 | 0 | 31 | 27 | 2 | 1 | 1 | 0 |
| DISORIENTATION | 7 | 5 | 1 | 0 | 0 | 1 | 7 | 5 | 1 | 0 | 0 | 1 |
| *DELIRIA* | | | | | | | | | | | | |
| DELIRIUM | 2 | 1 | 0 | 0 | 0 | 1 | 2 | 1 | 0 | 0 | 0 | 1 |
| ***DEPRESSED MOOD DISORDERS AND DISTURBANCES*** | | | | | | | | | | | | |
| *DEPRESSIVE DISORDERS* | | | | | | | | | | | | |
| DEPRESSION | 12 | 10 | 2 | 0 | 0 | 0 | 11 | 10 | 1 | 0 | 0 | 0 |
| *MOOD ALTERATIONS WITH DEPRESSIVE SYMPTOMS* | | | | | | | | | | | | |
| DEPRESSED MOOD | 16 | 10 | 5 | 1 | 0 | 0 | 16 | 10 | 5 | 1 | 0 | 0 |
| TEARFULNESS | 1 | 1 | 0 | 0 | 0 | 0 | 1 | 1 | 0 | 0 | 0 | 0 |
| ***DISSOCIATIVE DISORDERS*** | | | | | | | | | | | | |
| *DISSOCIATIVE STATES* | | | | | | | | | | | | |
| DISSOCIATION | 3 | 1 | 0 | 2 | 0 | 0 | 3 | 1 | 0 | 2 | 0 | 0 |
| ***DISTURBANCES IN THINKING AND PERCEPTION*** | | | | | | | | | | | | |
| *DELUSIONAL SYMPTOMS* | | | | | | | | | | | | |

| | | | | | | | | | | | | |
|---|---|---|---|---|---|---|---|---|---|---|---|---|
| DELUSION | 1 | 0 | 0 | 0 | 0 | 1 | 1 | 0 | 0 | 0 | 0 | 1 |
| *HALLUCINATIONS (EXCL SLEEP-RELATED)* | | | | | | | | | | | | |
| HALLUCINATION | 5 | 4 | 0 | 0 | 0 | 1 | 5 | 4 | 0 | 0 | 0 | 1 |
| *PERCEPTION DISTURBANCES NEC* | | | | | | | | | | | | |
| DEREALISATION | 2 | 2 | 0 | 0 | 0 | 0 | 2 | 2 | 0 | 0 | 0 | 0 |
| *THINKING DISTURBANCES* | | | | | | | | | | | | |
| BRADYPHRENIA | 4 | 1 | 2 | 1 | 0 | 0 | 4 | 1 | 2 | 1 | 0 | 0 |
| THOUGHT BLOCKING | 1 | 1 | 0 | 0 | 0 | 0 | 1 | 1 | 0 | 0 | 0 | 0 |
| ***MOOD DISORDERS AND DISTURBANCES NEC*** | | | | | | | | | | | | |
| *AFFECT ALTERATIONS NEC* | | | | | | | | | | | | |
| AFFECT LABILITY | 1 | 0 | 1 | 0 | 0 | 0 | 1 | 0 | 1 | 0 | 0 | 0 |
| INAPPROPRIATE AFFECT | 2 | 2 | 0 | 0 | 0 | 0 | 2 | 2 | 0 | 0 | 0 | 0 |
| *EMOTIONAL AND MOOD DISTURBANCES NEC* | | | | | | | | | | | | |
| ANGER | 1 | 1 | 0 | 0 | 0 | 0 | 1 | 1 | 0 | 0 | 0 | 0 |
| EMOTIONAL DISORDER | 1 | 0 | 1 | 0 | 0 | 0 | 1 | 0 | 1 | 0 | 0 | 0 |
| EMOTIONAL DISTRESS | 1 | 1 | 0 | 0 | 0 | 0 | 1 | 1 | 0 | 0 | 0 | 0 |
| EUPHORIC MOOD | 2 | 2 | 0 | 0 | 0 | 0 | 2 | 2 | 0 | 0 | 0 | 0 |
| IRRITABILITY | 14 | 13 | 0 | 0 | 0 | 1 | 14 | 13 | 0 | 0 | 0 | 1 |
| MOOD ALTERED | 3 | 0 | 2 | 1 | 0 | 0 | 2 | 0 | 1 | 1 | 0 | 0 |
| *FLUCTUATING MOOD SYMPTOMS* | | | | | | | | | | | | |
| MOOD SWINGS | 3 | 1 | 2 | 0 | 0 | 0 | 2 | 1 | 1 | 0 | 0 | 0 |
| *MOOD DISORDERS NEC* | | | | | | | | | | | | |
| APATHY | 3 | 1 | 1 | 1 | 0 | 0 | 3 | 1 | 1 | 1 | 0 | 0 |
| LISTLESS | 3 | 1 | 1 | 0 | 1 | 0 | 3 | 1 | 1 | 0 | 1 | 0 |
| ***PERSONALITY DISORDERS AND DISTURBANCES IN BEHAVIOUR*** | | | | | | | | | | | | |
| *BEHAVIOUR AND SOCIALISATION DISTURBANCES* | | | | | | | | | | | | |
| AGGRESSION | 1 | 1 | 0 | 0 | 0 | 0 | 1 | 1 | 0 | 0 | 0 | 0 |
| ***PSYCHIATRIC AND BEHAVIOURAL SYMPTOMS NEC*** | | | | | | | | | | | | |
| *PSYCHIATRIC SYMPTOMS NEC* | | | | | | | | | | | | |
| PSYCHIATRIC SYMPTOM | 1 | 0 | 0 | 0 | 0 | 1 | 1 | 0 | 0 | 0 | 0 | 1 |
| ***PSYCHIATRIC DISORDERS NEC*** | | | | | | | | | | | | |
| *MENTAL DISORDERS NEC* | | | | | | | | | | | | |
| MENTAL DISORDER | 1 | 0 | 1 | 0 | 0 | 0 | 1 | 0 | 1 | 0 | 0 | 0 |
| ***SCHIZOPHRENIA AND OTHER PSYCHOTIC DISORDERS*** | | | | | | | | | | | | |
| *PSYCHOTIC DISORDER NEC* | | | | | | | | | | | | |
| PSYCHOTIC DISORDER | 1 | 1 | 0 | 0 | 0 | 0 | 1 | 1 | 0 | 0 | 0 | 0 |
| ***SEXUAL DYSFUNCTIONS, DISTURBANCES AND GENDER IDENTITY DISORDERS*** | | | | | | | | | | | | |
| *SEXUAL DESIRE DISORDERS* | | | | | | | | | | | | |

| | | | | | | | | | | | | |
|---|---|---|---|---|---|---|---|---|---|---|---|---|
| LOSS OF LIBIDO | 3 | 3 | 0 | 0 | 0 | 0 | 3 | 3 | 0 | 0 | 0 | 0 |
| ***SLEEP DISORDERS AND DISTURBANCES*** | | | | | | | | | | | | |
| *DISTURBANCES IN INITIATING AND MAINTAINING SLEEP* | | | | | | | | | | | | |
| INITIAL INSOMNIA | 1 | 1 | 0 | 0 | 0 | 0 | 1 | 1 | 0 | 0 | 0 | 0 |
| INSOMNIA | 118 | 88 | 16 | 9 | 1 | 4 | 110 | 88 | 11 | 7 | 0 | 4 |
| *DYSSOMNIAS* | | | | | | | | | | | | |
| BREATHING-RELATED SLEEP DISORDER | 1 | 1 | 0 | 0 | 0 | 0 | 1 | 1 | 0 | 0 | 0 | 0 |
| POOR QUALITY SLEEP | 37 | 30 | 4 | 3 | 0 | 0 | 36 | 30 | 3 | 3 | 0 | 0 |
| *PARASOMNIAS* | | | | | | | | | | | | |
| ABNORMAL DREAMS | 18 | 11 | 2 | 1 | 1 | 3 | 17 | 11 | 2 | 1 | 0 | 3 |
| EXPLODING HEAD SYNDROME | 1 | 0 | 1 | 0 | 0 | 0 | 1 | 0 | 1 | 0 | 0 | 0 |
| NIGHTMARE | 13 | 7 | 3 | 2 | 0 | 1 | 13 | 7 | 3 | 2 | 0 | 1 |
| *SLEEP DISORDERS NEC* | | | | | | | | | | | | |
| SLEEP DISORDER | 13 | 11 | 2 | 0 | 0 | 0 | 13 | 11 | 2 | 0 | 0 | 0 |
| ***SOMATIC SYMPTOM AND RELATED DISORDERS*** | | | | | | | | | | | | |
| *SOMATIC SYMPTOM DISORDERS* | | | | | | | | | | | | |
| HABIT COUGH | 4 | 3 | 0 | 1 | 0 | 0 | 4 | 3 | 0 | 1 | 0 | 0 |
| ***SUICIDAL AND SELF-INJURIOUS BEHAVIOURS NEC*** | | | | | | | | | | | | |
| *SUICIDAL AND SELF-INJURIOUS BEHAVIOUR* | | | | | | | | | | | | |
| SUICIDAL IDEATION | 1 | 1 | 0 | 0 | 0 | 0 | 1 | 1 | 0 | 0 | 0 | 0 |
| **RENAL AND URINARY DISORDERS** | | | | | | | | | | | | |
| ***RENAL DISORDERS (EXCL NEPHROPATHIES)*** | | | | | | | | | | | | |
| *RENAL FAILURE AND IMPAIRMENT* | | | | | | | | | | | | |
| RENAL FAILURE | 1 | 1 | 0 | 0 | 0 | 0 | 1 | 1 | 0 | 0 | 0 | 0 |
| ***URINARY TRACT SIGNS AND SYMPTOMS*** | | | | | | | | | | | | |
| *BLADDER AND URETHRAL SYMPTOMS* | | | | | | | | | | | | |
| BLADDER PAIN | 2 | 1 | 0 | 0 | 0 | 1 | 2 | 1 | 0 | 0 | 0 | 1 |
| MICTURITION URGENCY | 3 | 1 | 0 | 2 | 0 | 0 | 3 | 1 | 0 | 2 | 0 | 0 |
| POLLAKIURIA | 12 | 10 | 1 | 0 | 0 | 1 | 12 | 10 | 1 | 0 | 0 | 1 |
| URINARY INCONTINENCE | 1 | 0 | 1 | 0 | 0 | 0 | 0 | 0 | 0 | 0 | 0 | 0 |
| *URINARY ABNORMALITIES* | | | | | | | | | | | | |
| HAEMATURIA | 1 | 1 | 0 | 0 | 0 | 0 | 1 | 1 | 0 | 0 | 0 | 0 |
| URINE ABNORMALITY | 1 | 0 | 0 | 1 | 0 | 0 | 1 | 0 | 0 | 1 | 0 | 0 |
| URINE ODOUR ABNORMAL | 2 | 2 | 0 | 0 | 0 | 0 | 2 | 2 | 0 | 0 | 0 | 0 |
| *URINARY TRACT SIGNS AND SYMPTOMS NEC* | | | | | | | | | | | | |
| POLYURIA | 3 | 2 | 0 | 0 | 1 | 0 | 3 | 2 | 0 | 0 | 1 | 0 |
| RENAL PAIN | 8 | 5 | 2 | 0 | 0 | 1 | 8 | 5 | 2 | 0 | 0 | 1 |
| **REPRODUCTIVE SYSTEM AND BREAST DISORDERS** | | | | | | | | | | | | |

| | | | | | | | | | | | | |
|---|---|---|---|---|---|---|---|---|---|---|---|---|
| ***BREAST DISORDERS*** | | | | | | | | | | | | |
| *BREAST DISORDERS NEC* | | | | | | | | | | | | |
| BREAST MASS | 4 | 3 | 0 | 0 | 0 | 1 | 4 | 3 | 0 | 0 | 0 | 1 |
| NIPPLE ENLARGEMENT | 1 | 1 | 0 | 0 | 0 | 0 | 1 | 1 | 0 | 0 | 0 | 0 |
| *BREAST SIGNS AND SYMPTOMS* | | | | | | | | | | | | |
| BREAST DISCHARGE | 1 | 0 | 0 | 0 | 0 | 1 | 1 | 0 | 0 | 0 | 0 | 1 |
| BREAST PAIN | 16 | 7 | 4 | 3 | 0 | 2 | 14 | 7 | 3 | 2 | 0 | 2 |
| BREAST SWELLING | 1 | 1 | 0 | 0 | 0 | 0 | 1 | 1 | 0 | 0 | 0 | 0 |
| BREAST TENDERNESS | 1 | 1 | 0 | 0 | 0 | 0 | 1 | 1 | 0 | 0 | 0 | 0 |
| NIPPLE PAIN | 2 | 2 | 0 | 0 | 0 | 0 | 2 | 2 | 0 | 0 | 0 | 0 |
| ***MENOPAUSE RELATED CONDITIONS*** | | | | | | | | | | | | |
| *MENOPAUSAL EFFECTS ON THE GENITOURINARY TRACT* | | | | | | | | | | | | |
| POSTMENOPAUSAL HAEMORRHAGE | 1 | 1 | 0 | 0 | 0 | 0 | 1 | 1 | 0 | 0 | 0 | 0 |
| ***MENSTRUAL CYCLE AND UTERINE BLEEDING DISORDERS*** | | | | | | | | | | | | |
| *MENSTRUATION AND UTERINE BLEEDING NEC* | | | | | | | | | | | | |
| DYSMENORRHOEA | 29 | 20 | 4 | 2 | 0 | 3 | 25 | 20 | 2 | 0 | 0 | 3 |
| INTERMENSTRUAL BLEEDING | 13 | 9 | 1 | 2 | 0 | 1 | 12 | 9 | 1 | 1 | 0 | 1 |
| MENSTRUAL DISORDER | 22 | 12 | 2 | 7 | 0 | 1 | 22 | 12 | 2 | 7 | 0 | 1 |
| MENSTRUATION IRREGULAR | 47 | 27 | 11 | 2 | 0 | 7 | 43 | 27 | 8 | 2 | 0 | 6 |
| PREMENSTRUAL PAIN | 2 | 1 | 0 | 0 | 0 | 1 | 2 | 1 | 0 | 0 | 0 | 1 |
| RETROGRADE MENSTRUATION | 1 | 1 | 0 | 0 | 0 | 0 | 1 | 1 | 0 | 0 | 0 | 0 |
| *MENSTRUATION WITH DECREASED BLEEDING* | | | | | | | | | | | | |
| AMENORRHOEA | 1 | 1 | 0 | 0 | 0 | 0 | 1 | 1 | 0 | 0 | 0 | 0 |
| HYPOMENORRHOEA | 9 | 4 | 2 | 0 | 0 | 3 | 8 | 4 | 1 | 0 | 0 | 3 |
| MENSTRUATION DELAYED | 47 | 23 | 11 | 1 | 0 | 12 | 43 | 23 | 8 | 1 | 0 | 11 |
| OLIGOMENORRHOEA | 3 | 1 | 1 | 0 | 0 | 1 | 3 | 1 | 1 | 0 | 0 | 1 |
| *MENSTRUATION WITH INCREASED BLEEDING* | | | | | | | | | | | | |
| HEAVY MENSTRUAL BLEEDING | 61 | 37 | 8 | 7 | 0 | 9 | 57 | 37 | 7 | 6 | 0 | 7 |
| POLYMENORRHOEA | 9 | 8 | 0 | 1 | 0 | 0 | 9 | 8 | 0 | 1 | 0 | 0 |
| ***PENILE AND SCROTAL DISORDERS (EXCL INFECTIONS AND INFLAMMATIONS)*** | | | | | | | | | | | | |
| *SCROTAL DISORDERS NEC* | | | | | | | | | | | | |
| SCROTAL SWELLING | 1 | 0 | 1 | 0 | 0 | 0 | 1 | 0 | 1 | 0 | 0 | 0 |
| ***REPRODUCTIVE TRACT DISORDERS NEC*** | | | | | | | | | | | | |
| *REPRODUCTIVE TRACT DISORDERS NEC (EXCL NEOPLASMS)* | | | | | | | | | | | | |
| GENITAL LESION | 1 | 1 | 0 | 0 | 0 | 0 | 1 | 1 | 0 | 0 | 0 | 0 |
| *REPRODUCTIVE TRACT SIGNS AND SYMPTOMS NEC* | | | | | | | | | | | | |
| GENITAL DISCOMFORT | 1 | 1 | 0 | 0 | 0 | 0 | 1 | 1 | 0 | 0 | 0 | 0 |
| GENITAL PAIN | 1 | 1 | 0 | 0 | 0 | 0 | 1 | 1 | 0 | 0 | 0 | 0 |

| | | | | | | | | | | | | |
|---|---|---|---|---|---|---|---|---|---|---|---|---|
| ***UTERINE, PELVIC AND BROAD LIGAMENT DISORDERS*** | | | | | | | | | | | | |
| *PELVIS AND BROAD LIGAMENT DISORDERS NEC* | | | | | | | | | | | | |
| ADNEXA UTERI PAIN | 1 | 1 | 0 | 0 | 0 | 0 | 1 | 1 | 0 | 0 | 0 | 0 |
| ***VULVOVAGINAL DISORDERS (EXCL INFECTIONS AND INFLAMMATIONS)*** | | | | | | | | | | | | |
| *VULVOVAGINAL DISORDERS NEC* | | | | | | | | | | | | |
| VAGINAL HAEMORRHAGE | 13 | 7 | 5 | 1 | 0 | 0 | 9 | 7 | 2 | 0 | 0 | 0 |
| *VULVOVAGINAL SIGNS AND SYMPTOMS* | | | | | | | | | | | | |
| VAGINAL DISCHARGE | 1 | 1 | 0 | 0 | 0 | 0 | 1 | 1 | 0 | 0 | 0 | 0 |
| **RESPIRATORY, THORACIC AND MEDIASTINAL DISORDERS** | | | | | | | | | | | | |
| ***BRONCHIAL DISORDERS (EXCL NEOPLASMS)*** | | | | | | | | | | | | |
| *BRONCHOSPASM AND OBSTRUCTION* | | | | | | | | | | | | |
| ASTHMA | 6 | 4 | 2 | 0 | 0 | 0 | 5 | 4 | 1 | 0 | 0 | 0 |
| WHEEZING | 12 | 8 | 2 | 1 | 0 | 1 | 10 | 8 | 1 | 0 | 0 | 1 |
| ***LOWER RESPIRATORY TRACT DISORDERS (EXCL OBSTRUCTION AND INFECTION)*** | | | | | | | | | | | | |
| *PULMONARY OEDEMAS* | | | | | | | | | | | | |
| PULMONARY CONGESTION | 1 | 1 | 0 | 0 | 0 | 0 | 1 | 1 | 0 | 0 | 0 | 0 |
| ***PULMONARY VASCULAR DISORDERS*** | | | | | | | | | | | | |
| *PULMONARY THROMBOTIC AND EMBOLIC CONDITIONS* | | | | | | | | | | | | |
| PULMONARY EMBOLISM | 2 | 1 | 0 | 0 | 0 | 1 | 2 | 1 | 0 | 0 | 0 | 1 |
| ***RESPIRATORY DISORDERS NEC*** | | | | | | | | | | | | |
| *BREATHING ABNORMALITIES* | | | | | | | | | | | | |
| DYSPNOEA | 129 | 79 | 20 | 11 | 5 | 14 | 124 | 79 | 17 | 10 | 5 | 13 |
| HYPERVENTILATION | 2 | 2 | 0 | 0 | 0 | 0 | 2 | 2 | 0 | 0 | 0 | 0 |
| HYPOPNOEA | 7 | 4 | 1 | 1 | 0 | 1 | 7 | 4 | 1 | 1 | 0 | 1 |
| IRREGULAR BREATHING | 1 | 1 | 0 | 0 | 0 | 0 | 1 | 1 | 0 | 0 | 0 | 0 |
| MOUTH BREATHING | 1 | 1 | 0 | 0 | 0 | 0 | 1 | 1 | 0 | 0 | 0 | 0 |
| RESPIRATION ABNORMAL | 1 | 1 | 0 | 0 | 0 | 0 | 1 | 1 | 0 | 0 | 0 | 0 |
| RESPIRATORY ARREST | 1 | 1 | 0 | 0 | 0 | 0 | 1 | 1 | 0 | 0 | 0 | 0 |
| RESPIRATORY FATIGUE | 2 | 2 | 0 | 0 | 0 | 0 | 2 | 2 | 0 | 0 | 0 | 0 |
| SLEEP APNOEA SYNDROME | 1 | 0 | 1 | 0 | 0 | 0 | 1 | 0 | 1 | 0 | 0 | 0 |
| *COUGHING AND ASSOCIATED SYMPTOMS* | | | | | | | | | | | | |
| COUGH | 132 | 87 | 20 | 16 | 3 | 6 | 126 | 87 | 18 | 14 | 1 | 6 |
| PRODUCTIVE COUGH | 9 | 3 | 4 | 0 | 0 | 2 | 8 | 3 | 4 | 0 | 0 | 1 |
| *RESPIRATORY TRACT DISORDERS NEC* | | | | | | | | | | | | |
| RESPIRATORY TRACT IRRITATION | 1 | 1 | 0 | 0 | 0 | 0 | 1 | 1 | 0 | 0 | 0 | 0 |
| ***RESPIRATORY TRACT SIGNS AND SYMPTOMS*** | | | | | | | | | | | | |
| *LOWER RESPIRATORY TRACT SIGNS AND SYMPTOMS* | | | | | | | | | | | | |
| PULMONARY PAIN | 3 | 3 | 0 | 0 | 0 | 0 | 3 | 3 | 0 | 0 | 0 | 0 |

| | | | | | | | | | | | | |
|---|---|---|---|---|---|---|---|---|---|---|---|---|
| *RESPIRATORY SIGNS AND SYMPTOMS NEC* | | | | | | | | | | | | |
| RESPIRATORY SYMPTOM | 1 | 1 | 0 | 0 | 0 | 0 | 1 | 1 | 0 | 0 | 0 | 0 |
| *UPPER RESPIRATORY TRACT SIGNS AND SYMPTOMS* | | | | | | | | | | | | |
| APHONIA | 3 | 2 | 0 | 1 | 0 | 0 | 3 | 2 | 0 | 1 | 0 | 0 |
| CATARRH | 7 | 3 | 1 | 2 | 0 | 1 | 6 | 3 | 1 | 1 | 0 | 1 |
| DRY THROAT | 11 | 7 | 3 | 0 | 0 | 1 | 9 | 7 | 1 | 0 | 0 | 1 |
| DYSPHONIA | 7 | 7 | 0 | 0 | 0 | 0 | 7 | 7 | 0 | 0 | 0 | 0 |
| INCREASED UPPER AIRWAY SECRETION | 1 | 1 | 0 | 0 | 0 | 0 | 1 | 1 | 0 | 0 | 0 | 0 |
| INCREASED VISCOSITY OF UPPER RESPIRATORY SE( | 1 | 0 | 0 | 1 | 0 | 0 | 1 | 0 | 0 | 1 | 0 | 0 |
| NASAL DISCOMFORT | 1 | 1 | 0 | 0 | 0 | 0 | 1 | 1 | 0 | 0 | 0 | 0 |
| OROPHARYNGEAL DISCOMFORT | 2 | 2 | 0 | 0 | 0 | 0 | 2 | 2 | 0 | 0 | 0 | 0 |
| OROPHARYNGEAL PAIN | 238 | 174 | 40 | 11 | 2 | 11 | 230 | 174 | 33 | 10 | 2 | 11 |
| PARANASAL SINUS DISCOMFORT | 3 | 3 | 0 | 0 | 0 | 0 | 3 | 3 | 0 | 0 | 0 | 0 |
| RHINALGIA | 2 | 1 | 1 | 0 | 0 | 0 | 2 | 1 | 1 | 0 | 0 | 0 |
| RHINORRHOEA | 126 | 76 | 25 | 15 | 2 | 8 | 112 | 76 | 15 | 12 | 1 | 8 |
| SINUS PAIN | 19 | 13 | 5 | 1 | 0 | 0 | 19 | 13 | 5 | 1 | 0 | 0 |
| SNEEZING | 31 | 22 | 5 | 1 | 1 | 2 | 30 | 22 | 4 | 1 | 1 | 2 |
| THROAT CLEARING | 1 | 1 | 0 | 0 | 0 | 0 | 1 | 1 | 0 | 0 | 0 | 0 |
| THROAT IRRITATION | 4 | 3 | 0 | 0 | 0 | 1 | 4 | 3 | 0 | 0 | 0 | 1 |
| THROAT TIGHTNESS | 5 | 4 | 1 | 0 | 0 | 0 | 5 | 4 | 1 | 0 | 0 | 0 |
| UPPER-AIRWAY COUGH SYNDROME | 1 | 0 | 0 | 1 | 0 | 0 | 1 | 0 | 0 | 1 | 0 | 0 |
| YAWNING | 3 | 2 | 0 | 0 | 0 | 1 | 3 | 2 | 0 | 0 | 0 | 1 |
| ***UPPER RESPIRATORY TRACT DISORDERS (EXCL INFECTIONS)*** | | | | | | | | | | | | |
| *NASAL CONGESTION AND INFLAMMATIONS* | | | | | | | | | | | | |
| NASAL CONGESTION | 18 | 11 | 6 | 1 | 0 | 0 | 16 | 11 | 4 | 1 | 0 | 0 |
| RHINITIS ALLERGIC | 2 | 2 | 0 | 0 | 0 | 0 | 2 | 2 | 0 | 0 | 0 | 0 |
| RHINITIS ATROPHIC | 1 | 0 | 1 | 0 | 0 | 0 | 1 | 0 | 1 | 0 | 0 | 0 |
| *NASAL DISORDERS NEC* | | | | | | | | | | | | |
| EPISTAXIS | 32 | 21 | 9 | 2 | 0 | 0 | 29 | 21 | 8 | 0 | 0 | 0 |
| NASAL DRYNESS | 4 | 3 | 1 | 0 | 0 | 0 | 3 | 3 | 0 | 0 | 0 | 0 |
| *PARANASAL SINUS DISORDERS (EXCL INFECTIONS AND NEOPLASMS)* | | | | | | | | | | | | |
| SINUS CONGESTION | 5 | 3 | 2 | 0 | 0 | 0 | 5 | 3 | 2 | 0 | 0 | 0 |
| *PHARYNGEAL DISORDERS (EXCL INFECTIONS AND NEOPLASMS)* | | | | | | | | | | | | |
| PHARYNGEAL SWELLING | 3 | 2 | 1 | 0 | 0 | 0 | 3 | 2 | 1 | 0 | 0 | 0 |
| PHARYNGEAL ULCERATION | 1 | 1 | 0 | 0 | 0 | 0 | 1 | 1 | 0 | 0 | 0 | 0 |
| TONSILLAR ERYTHEMA | 1 | 0 | 0 | 0 | 0 | 1 | 1 | 0 | 0 | 0 | 0 | 1 |
| *TRACHEAL DISORDERS (EXCL INFECTIONS AND NEOPLASMS)* | | | | | | | | | | | | |
| TRACHEAL PAIN | 1 | 1 | 0 | 0 | 0 | 0 | 1 | 1 | 0 | 0 | 0 | 0 |

| | | | | | | | | | | | | |
|---|---|---|---|---|---|---|---|---|---|---|---|---|
| **SKIN AND SUBCUTANEOUS TISSUE DISORDERS** | | | | | | | | | | | | |
| ***ANGIOEDEMA AND URTICARIA*** | | | | | | | | | | | | |
| *ANGIOEDEMAS* | | | | | | | | | | | | |
| ANGIOEDEMA | 3 | 3 | 0 | 0 | 0 | 0 | 3 | 3 | 0 | 0 | 0 | 0 |
| *URTICARIAS* | | | | | | | | | | | | |
| COLD URTICARIA | 1 | 0 | 1 | 0 | 0 | 0 | 1 | 0 | 1 | 0 | 0 | 0 |
| SOLAR URTICARIA | 1 | 1 | 0 | 0 | 0 | 0 | 1 | 1 | 0 | 0 | 0 | 0 |
| URTICARIA | 21 | 12 | 2 | 7 | 0 | 0 | 21 | 12 | 2 | 7 | 0 | 0 |
| URTICARIA CHRONIC | 1 | 0 | 0 | 0 | 0 | 1 | 1 | 0 | 0 | 0 | 0 | 1 |
| ***CORNIFICATION AND DYSTROPHIC SKIN DISORDERS*** | | | | | | | | | | | | |
| *SKIN DYSTROPHIES* | | | | | | | | | | | | |
| HYPERTROPHIC SCAR | 1 | 1 | 0 | 0 | 0 | 0 | 1 | 1 | 0 | 0 | 0 | 0 |
| ***CUTANEOUS NEOPLASMS BENIGN*** | | | | | | | | | | | | |
| *SKIN CYSTS AND POLYPS* | | | | | | | | | | | | |
| DERMAL CYST | 1 | 1 | 0 | 0 | 0 | 0 | 1 | 1 | 0 | 0 | 0 | 0 |
| ***EPIDERMAL AND DERMAL CONDITIONS*** | | | | | | | | | | | | |
| *BULLOUS CONDITIONS* | | | | | | | | | | | | |
| BLISTER | 5 | 3 | 1 | 0 | 0 | 1 | 5 | 3 | 1 | 0 | 0 | 1 |
| BLOOD BLISTER | 1 | 0 | 0 | 0 | 0 | 1 | 1 | 0 | 0 | 0 | 0 | 1 |
| PEMPHIGOID | 1 | 0 | 0 | 0 | 1 | 0 | 1 | 0 | 0 | 0 | 1 | 0 |
| TOXIC EPIDERMAL NECROLYSIS | 2 | 2 | 0 | 0 | 0 | 0 | 2 | 2 | 0 | 0 | 0 | 0 |
| *DERMAL AND EPIDERMAL CONDITIONS NEC* | | | | | | | | | | | | |
| DRY SKIN | 9 | 4 | 0 | 1 | 0 | 4 | 8 | 4 | 0 | 1 | 0 | 3 |
| PAIN OF SKIN | 37 | 26 | 3 | 4 | 1 | 3 | 35 | 26 | 2 | 4 | 0 | 3 |
| SENSITIVE SKIN | 28 | 22 | 2 | 1 | 1 | 2 | 26 | 22 | 1 | 1 | 0 | 2 |
| SKIN BURNING SENSATION | 13 | 9 | 1 | 1 | 0 | 2 | 13 | 9 | 1 | 1 | 0 | 2 |
| SKIN DISCOLOURATION | 1 | 1 | 0 | 0 | 0 | 0 | 1 | 1 | 0 | 0 | 0 | 0 |
| SKIN FRAGILITY | 1 | 1 | 0 | 0 | 0 | 0 | 1 | 1 | 0 | 0 | 0 | 0 |
| SKIN LESION | 1 | 0 | 0 | 0 | 0 | 1 | 0 | 0 | 0 | 0 | 0 | 0 |
| SKIN ODOUR ABNORMAL | 2 | 2 | 0 | 0 | 0 | 0 | 2 | 2 | 0 | 0 | 0 | 0 |
| SKIN REACTION | 3 | 2 | 0 | 1 | 0 | 0 | 3 | 2 | 0 | 1 | 0 | 0 |
| SKIN SENSITISATION | 1 | 1 | 0 | 0 | 0 | 0 | 1 | 1 | 0 | 0 | 0 | 0 |
| SKIN WARM | 33 | 23 | 4 | 5 | 0 | 1 | 31 | 23 | 2 | 5 | 0 | 1 |
| *DERMATITIS AND ECZEMA* | | | | | | | | | | | | |
| DERMATITIS | 4 | 4 | 0 | 0 | 0 | 0 | 4 | 4 | 0 | 0 | 0 | 0 |
| DERMATITIS ALLERGIC | 4 | 4 | 0 | 0 | 0 | 0 | 4 | 4 | 0 | 0 | 0 | 0 |
| DERMATITIS ATOPIC | 2 | 2 | 0 | 0 | 0 | 0 | 2 | 2 | 0 | 0 | 0 | 0 |
| DERMATITIS CONTACT | 1 | 1 | 0 | 0 | 0 | 0 | 1 | 1 | 0 | 0 | 0 | 0 |

| | | | | | | | | | | | | |
|---|---|---|---|---|---|---|---|---|---|---|---|---|
| ECZEMA | 6 | 3 | 1 | 0 | 0 | 2 | 5 | 3 | 1 | 0 | 0 | 1 |
| ECZEMA ASTEATOTIC | 1 | 0 | 0 | 1 | 0 | 0 | 1 | 0 | 0 | 1 | 0 | 0 |
| NEURODERMATITIS | 1 | 1 | 0 | 0 | 0 | 0 | 1 | 1 | 0 | 0 | 0 | 0 |
| SEBORRHOEIC DERMATITIS | 2 | 2 | 0 | 0 | 0 | 0 | 2 | 2 | 0 | 0 | 0 | 0 |
| SKIN IRRITATION | 9 | 6 | 2 | 1 | 0 | 0 | 8 | 6 | 1 | 1 | 0 | 0 |
| *ERYTHEMAS* | | | | | | | | | | | | |
| ERYTHEMA | 59 | 39 | 11 | 3 | 1 | 5 | 54 | 39 | 9 | 2 | 0 | 4 |
| *EXFOLIATIVE CONDITIONS* | | | | | | | | | | | | |
| SKIN EXFOLIATION | 2 | 1 | 0 | 0 | 1 | 0 | 1 | 1 | 0 | 0 | 0 | 0 |
| *PAPULOSQUAMOUS CONDITIONS* | | | | | | | | | | | | |
| LICHEN PLANUS | 1 | 0 | 1 | 0 | 0 | 0 | 1 | 0 | 1 | 0 | 0 | 0 |
| PITYRIASIS ROSEA | 1 | 1 | 0 | 0 | 0 | 0 | 1 | 1 | 0 | 0 | 0 | 0 |
| *PHOTOSENSITIVITY AND PHOTODERMATOSIS CONDITIONS* | | | | | | | | | | | | |
| PHOTOSENSITIVITY REACTION | 6 | 4 | 2 | 0 | 0 | 0 | 5 | 4 | 1 | 0 | 0 | 0 |
| *PRURITUS NEC* | | | | | | | | | | | | |
| PRURITUS | 176 | 122 | 29 | 10 | 2 | 13 | 165 | 122 | 21 | 8 | 2 | 12 |
| *PSORIATIC CONDITIONS* | | | | | | | | | | | | |
| PSORIASIS | 10 | 5 | 3 | 1 | 0 | 1 | 10 | 5 | 3 | 1 | 0 | 1 |
| *RASHES, ERUPTIONS AND EXANTHEMS NEC* | | | | | | | | | | | | |
| RASH | 178 | 125 | 25 | 22 | 2 | 4 | 162 | 125 | 17 | 14 | 2 | 4 |
| RASH ERYTHEMATOUS | 39 | 26 | 3 | 6 | 0 | 4 | 37 | 26 | 2 | 6 | 0 | 3 |
| RASH MACULAR | 5 | 3 | 2 | 0 | 0 | 0 | 5 | 3 | 2 | 0 | 0 | 0 |
| RASH MORBILLIFORM | 1 | 0 | 0 | 1 | 0 | 0 | 0 | 0 | 0 | 0 | 0 | 0 |
| RASH PAPULAR | 13 | 10 | 2 | 0 | 0 | 1 | 13 | 10 | 2 | 0 | 0 | 1 |
| RASH PRURITIC | 36 | 22 | 4 | 5 | 2 | 3 | 35 | 22 | 4 | 5 | 1 | 3 |
| *SKIN INJURIES AND MECHANICAL DERMATOSES* | | | | | | | | | | | | |
| DECUBITUS ULCER | 2 | 1 | 0 | 1 | 0 | 0 | 1 | 1 | 0 | 0 | 0 | 0 |
| NEEDLE TRACK MARKS | 1 | 1 | 0 | 0 | 0 | 0 | 1 | 1 | 0 | 0 | 0 | 0 |
| ***SKIN AND SUBCUTANEOUS TISSUE DISORDERS NEC*** | | | | | | | | | | | | |
| *SKIN AND SUBCUTANEOUS TISSUE ULCERATIONS* | | | | | | | | | | | | |
| SKIN EROSION | 2 | 1 | 1 | 0 | 0 | 0 | 1 | 1 | 0 | 0 | 0 | 0 |
| ***SKIN APPENDAGE CONDITIONS*** | | | | | | | | | | | | |
| *ACNES* | | | | | | | | | | | | |
| ACNE | 1 | 1 | 0 | 0 | 0 | 0 | 1 | 1 | 0 | 0 | 0 | 0 |
| ACNE CYSTIC | 1 | 0 | 1 | 0 | 0 | 0 | 1 | 0 | 1 | 0 | 0 | 0 |
| *ALOPECIAS* | | | | | | | | | | | | |
| ALOPECIA | 14 | 6 | 2 | 1 | 0 | 5 | 12 | 6 | 1 | 1 | 0 | 4 |
| *APOCRINE AND ECCRINE GLAND DISORDERS* | | | | | | | | | | | | |

| | | | | | | | | | | | | |
|---|---|---|---|---|---|---|---|---|---|---|---|---|
| COLD SWEAT | 35 | 28 | 5 | 1 | 1 | 0 | 33 | 28 | 3 | 1 | 1 | 0 |
| HYPERHIDROSIS | 169 | 122 | 19 | 8 | 1 | 19 | 163 | 122 | 15 | 6 | 1 | 19 |
| MILIARIA | 6 | 3 | 2 | 0 | 0 | 1 | 5 | 3 | 1 | 0 | 0 | 1 |
| NIGHT SWEATS | 81 | 59 | 12 | 6 | 1 | 3 | 74 | 59 | 7 | 5 | 0 | 3 |
| *HYPERTRICHOSES* | | | | | | | | | | | | |
| HIRSUTISM | 1 | 1 | 0 | 0 | 0 | 0 | 1 | 1 | 0 | 0 | 0 | 0 |
| *NAIL AND NAIL BED CONDITIONS (EXCL INFECTIONS AND INFESTATIONS)* | | | | | | | | | | | | |
| NAIL DISCOLOURATION | 1 | 0 | 0 | 0 | 0 | 1 | 1 | 0 | 0 | 0 | 0 | 1 |
| ONYCHOCLASIS | 1 | 0 | 0 | 0 | 0 | 1 | 0 | 0 | 0 | 0 | 0 | 0 |
| *PILAR DISORDERS NEC* | | | | | | | | | | | | |
| PILOERECTION | 3 | 2 | 1 | 0 | 0 | 0 | 2 | 2 | 0 | 0 | 0 | 0 |
| ***SKIN VASCULAR ABNORMALITIES*** | | | | | | | | | | | | |
| *PURPURA AND RELATED CONDITIONS* | | | | | | | | | | | | |
| PETECHIAE | 2 | 2 | 0 | 0 | 0 | 0 | 2 | 2 | 0 | 0 | 0 | 0 |
| PURPURA | 1 | 0 | 0 | 0 | 1 | 0 | 1 | 0 | 0 | 0 | 1 | 0 |
| *SKIN HAEMORRHAGES* | | | | | | | | | | | | |
| SKIN HAEMORRHAGE | 1 | 1 | 0 | 0 | 0 | 0 | 1 | 1 | 0 | 0 | 0 | 0 |
| **SOCIAL CIRCUMSTANCES** | | | | | | | | | | | | |
| ***ECONOMIC AND HOUSING ISSUES*** | | | | | | | | | | | | |
| *EMPLOYMENT ISSUES* | | | | | | | | | | | | |
| RETIREMENT | 2 | 2 | 0 | 0 | 0 | 0 | 2 | 2 | 0 | 0 | 0 | 0 |
| ***ENVIRONMENTAL ISSUES*** | | | | | | | | | | | | |
| *NON-OCCUPATIONAL AND UNSPECIFIED ENVIRONMENTAL PROBLEMS* | | | | | | | | | | | | |
| POLLUTION | 1 | 1 | 0 | 0 | 0 | 0 | 1 | 1 | 0 | 0 | 0 | 0 |
| **SURGICAL AND MEDICAL PROCEDURES** | | | | | | | | | | | | |
| ***BONE AND JOINT THERAPEUTIC PROCEDURES*** | | | | | | | | | | | | |
| *JOINT THERAPEUTIC PROCEDURES* | | | | | | | | | | | | |
| KNEE OPERATION | 1 | 1 | 0 | 0 | 0 | 0 | 1 | 1 | 0 | 0 | 0 | 0 |
| ***BREAST THERAPEUTIC PROCEDURES*** | | | | | | | | | | | | |
| *MASTECTOMIES* | | | | | | | | | | | | |
| BREAST CONSERVING SURGERY | 1 | 0 | 1 | 0 | 0 | 0 | 1 | 0 | 1 | 0 | 0 | 0 |
| ***GASTROINTESTINAL THERAPEUTIC PROCEDURES*** | | | | | | | | | | | | |
| *GASTROINTESTINAL THERAPEUTIC PROCEDURES NEC* | | | | | | | | | | | | |
| PROPHYLAXIS OF NAUSEA AND VOMITING | 1 | 1 | 0 | 0 | 0 | 0 | 1 | 1 | 0 | 0 | 0 | 0 |
| ***HEAD AND NECK THERAPEUTIC PROCEDURES*** | | | | | | | | | | | | |
| *DENTAL AND GINGIVAL THERAPEUTIC PROCEDURES* | | | | | | | | | | | | |
| DENTAL CARE | 2 | 2 | 0 | 0 | 0 | 0 | 2 | 2 | 0 | 0 | 0 | 0 |
| ***NERVOUS SYSTEM, SKULL AND SPINE THERAPEUTIC PROCEDURES*** | | | | | | | | | | | | |

| | | | | | | | | | | | | |
|---|---|---|---|---|---|---|---|---|---|---|---|---|
| *SKULL AND BRAIN THERAPEUTIC PROCEDURES* | | | | | | | | | | | | |
| BRAIN TUMOUR OPERATION | 1 | 1 | 0 | 0 | 0 | 0 | 1 | 1 | 0 | 0 | 0 | 0 |
| ***OBSTETRIC AND GYNAECOLOGICAL THERAPEUTIC PROCEDURES*** | | | | | | | | | | | | |
| *FERTILITY AND FERTILISATION INTERVENTIONS FEMALE* | | | | | | | | | | | | |
| ENDOMETRIAL SCRATCHING | 1 | 1 | 0 | 0 | 0 | 0 | 1 | 1 | 0 | 0 | 0 | 0 |
| ***RESPIRATORY TRACT THERAPEUTIC PROCEDURES*** | | | | | | | | | | | | |
| *RESPIRATORY TRACT THERAPEUTIC PROCEDURES NEC* | | | | | | | | | | | | |
| OXYGEN THERAPY | 1 | 0 | 0 | 0 | 0 | 1 | 1 | 0 | 0 | 0 | 0 | 1 |
| ***THERAPEUTIC PROCEDURES AND SUPPORTIVE CARE NEC*** | | | | | | | | | | | | |
| *ANAESTHESIA AND ALLIED PROCEDURES* | | | | | | | | | | | | |
| NERVE BLOCK | 2 | 2 | 0 | 0 | 0 | 0 | 2 | 2 | 0 | 0 | 0 | 0 |
| *DIETARY AND NUTRITIONAL THERAPIES* | | | | | | | | | | | | |
| NOTHING BY MOUTH ORDER | 1 | 0 | 0 | 0 | 0 | 1 | 1 | 0 | 0 | 0 | 0 | 1 |
| *IMMUNISATIONS* | | | | | | | | | | | | |
| COVID-19 IMMUNISATION | 1 | 0 | 1 | 0 | 0 | 0 | 0 | 0 | 0 | 0 | 0 | 0 |
| *THERAPEUTIC PROCEDURES NEC* | | | | | | | | | | | | |
| INJECTION | 3 | 3 | 0 | 0 | 0 | 0 | 3 | 3 | 0 | 0 | 0 | 0 |
| LOCALISED ALTERNATING HOT AND COLD THERAPY | 2 | 2 | 0 | 0 | 0 | 0 | 2 | 2 | 0 | 0 | 0 | 0 |
| MASS EXCISION | 1 | 0 | 1 | 0 | 0 | 0 | 1 | 0 | 1 | 0 | 0 | 0 |
| REINFUSION | 1 | 0 | 0 | 1 | 0 | 0 | 1 | 0 | 0 | 1 | 0 | 0 |
| **VASCULAR DISORDERS** | | | | | | | | | | | | |
| ***ARTERIOSCLEROSIS, STENOSIS, VASCULAR INSUFFICIENCY AND NECROSIS*** | | | | | | | | | | | | |
| *NON-SITE SPECIFIC NECROSIS AND VASCULAR INSUFFICIENCY NEC* | | | | | | | | | | | | |
| VASOSPASM | 1 | 1 | 0 | 0 | 0 | 0 | 1 | 1 | 0 | 0 | 0 | 0 |
| *PERIPHERAL VASOCONSTRICTION, NECROSIS AND VASCULAR INSUFFICIENCY* | | | | | | | | | | | | |
| PERIPHERAL COLDNESS | 55 | 48 | 3 | 2 | 0 | 2 | 54 | 48 | 2 | 2 | 0 | 2 |
| RAYNAUD'S PHENOMENON | 1 | 1 | 0 | 0 | 0 | 0 | 1 | 1 | 0 | 0 | 0 | 0 |
| ***DECREASED AND NONSPECIFIC BLOOD PRESSURE DISORDERS AND SHOCK*** | | | | | | | | | | | | |
| *BLOOD PRESSURE DISORDERS NEC* | | | | | | | | | | | | |
| BLOOD PRESSURE FLUCTUATION | 1 | 1 | 0 | 0 | 0 | 0 | 1 | 1 | 0 | 0 | 0 | 0 |
| *VASCULAR HYPOTENSIVE DISORDERS* | | | | | | | | | | | | |
| CAPILLARY LEAK SYNDROME | 1 | 1 | 0 | 0 | 0 | 0 | 1 | 1 | 0 | 0 | 0 | 0 |
| HYPOTENSION | 8 | 7 | 0 | 1 | 0 | 0 | 7 | 7 | 0 | 0 | 0 | 0 |
| ORTHOSTATIC HYPOTENSION | 2 | 1 | 1 | 0 | 0 | 0 | 2 | 1 | 1 | 0 | 0 | 0 |
| ***EMBOLISM AND THROMBOSIS*** | | | | | | | | | | | | |
| *NON-SITE SPECIFIC EMBOLISM AND THROMBOSIS* | | | | | | | | | | | | |
| EMBOLISM | 1 | 1 | 0 | 0 | 0 | 0 | 1 | 1 | 0 | 0 | 0 | 0 |
| THROMBOSIS | 11 | 9 | 1 | 0 | 0 | 1 | 11 | 9 | 1 | 0 | 0 | 1 |

| | | | | | | | | | | | | |
|---|---|---|---|---|---|---|---|---|---|---|---|---|
| VENOUS THROMBOSIS | 1 | 0 | 1 | 0 | 0 | 0 | 0 | 0 | 0 | 0 | 0 | 0 |
| *PERIPHERAL EMBOLISM AND THROMBOSIS* | | | | | | | | | | | | |
| BLUE TOE SYNDROME | 2 | 2 | 0 | 0 | 0 | 0 | 2 | 2 | 0 | 0 | 0 | 0 |
| DEEP VEIN THROMBOSIS | 5 | 3 | 1 | 0 | 0 | 1 | 4 | 3 | 0 | 0 | 0 | 1 |
| SUPERFICIAL VEIN THROMBOSIS | 1 | 0 | 0 | 0 | 0 | 1 | 1 | 0 | 0 | 0 | 0 | 1 |
| ***LYMPHATIC VESSEL DISORDERS*** | | | | | | | | | | | | |
| *LYMPHOEDEMAS* | | | | | | | | | | | | |
| LYMPHOEDEMA | 4 | 2 | 0 | 2 | 0 | 0 | 3 | 2 | 0 | 1 | 0 | 0 |
| ***VASCULAR DISORDERS NEC*** | | | | | | | | | | | | |
| *NON-SITE SPECIFIC VASCULAR DISORDERS NEC* | | | | | | | | | | | | |
| VASCULAR PAIN | 1 | 0 | 1 | 0 | 0 | 0 | 1 | 0 | 1 | 0 | 0 | 0 |
| VEIN DISCOLOURATION | 1 | 0 | 0 | 0 | 0 | 1 | 1 | 0 | 0 | 0 | 0 | 1 |
| VEIN RUPTURE | 1 | 0 | 1 | 0 | 0 | 0 | 1 | 0 | 1 | 0 | 0 | 0 |
| *PERIPHERAL VASCULAR DISORDERS NEC* | | | | | | | | | | | | |
| FLUSHING | 30 | 24 | 4 | 1 | 0 | 1 | 28 | 24 | 2 | 1 | 0 | 1 |
| HOT FLUSH | 119 | 88 | 17 | 6 | 0 | 8 | 113 | 88 | 13 | 4 | 0 | 8 |
| *SITE SPECIFIC VASCULAR DISORDERS NEC* | | | | | | | | | | | | |
| PALLOR | 9 | 5 | 2 | 0 | 0 | 2 | 7 | 5 | 0 | 0 | 0 | 2 |
| ***VASCULAR HAEMORRHAGIC DISORDERS*** | | | | | | | | | | | | |
| *HAEMORRHAGES NEC* | | | | | | | | | | | | |
| HAEMATOMA | 1 | 1 | 0 | 0 | 0 | 0 | 1 | 1 | 0 | 0 | 0 | 0 |
| HAEMORRHAGE | 7 | 3 | 1 | 2 | 0 | 1 | 6 | 3 | 0 | 2 | 0 | 1 |
| ***VASCULAR HYPERTENSIVE DISORDERS*** | | | | | | | | | | | | |
| *ACCELERATED AND MALIGNANT HYPERTENSION* | | | | | | | | | | | | |
| HYPERTENSIVE URGENCY | 1 | 1 | 0 | 0 | 0 | 0 | 1 | 1 | 0 | 0 | 0 | 0 |
| *VASCULAR HYPERTENSIVE DISORDERS NEC* | | | | | | | | | | | | |
| HYPERTENSION | 15 | 9 | 4 | 1 | 0 | 1 | 14 | 9 | 3 | 1 | 0 | 1 |
| SYSTOLIC HYPERTENSION | 1 | 1 | 0 | 0 | 0 | 0 | 1 | 1 | 0 | 0 | 0 | 0 |
| ***VASCULAR INFECTIONS AND INFLAMMATIONS*** | | | | | | | | | | | | |
| *ARTERIAL INFECTIONS AND INFLAMMATIONS* | | | | | | | | | | | | |
| GIANT CELL ARTERITIS | 1 | 1 | 0 | 0 | 0 | 0 | 1 | 1 | 0 | 0 | 0 | 0 |
| *PHLEBITIS NEC* | | | | | | | | | | | | |
| PHLEBITIS | 3 | 1 | 1 | 1 | 0 | 0 | 2 | 1 | 1 | 0 | 0 | 0 |
| *VASCULITIDES NEC* | | | | | | | | | | | | |
| VASCULITIS | 1 | 0 | 0 | 0 | 1 | 0 | 1 | 0 | 0 | 0 | 1 | 0 |
| ***VENOUS VARICES*** | | | | | | | | | | | | |
| *VARICOSE VEINS NEC* | | | | | | | | | | | | |
| SPIDER VEIN | 1 | 1 | 0 | 0 | 0 | 0 | 1 | 1 | 0 | 0 | 0 | 0 |

| | | | | | | | | | | | | |
|---|---|---|---|---|---|---|---|---|---|---|---|---|
| VARICOSE VEIN | 3 | 3 | 0 | 0 | 0 | 0 | 3 | 3 | 0 | 0 | 0 | 0 |
| **TOTAL ADR EVENTS** | ***38144*** | ***26096*** | ***5973*** | ***3446*** | ***527*** | ***2102*** | ***35647*** | ***26096*** | ***4317*** | ***2833*** | ***421*** | ***1980*** |

**TABLE S1-2. Pfizer BioNTech COVID-19 vaccine: ADR listing for events reported in the YCVM in (a) those reporting any vaccination dose and (b) in those who had reported a 1st dose vaccination and any subsequent doses.**

| | Patients with any vaccination dose: ADR Counts | | | | | Patients reporting a 1st dose: ADR Counts | | | | |
|---|---|---|---|---|---|---|---|---|---|---|
| **REACTION TERM (SOC, *HLGT*, *HLT*, PT)** | All doses | 1st dose | 2nd dose | 3rd dose | Other doses | All doses | 1st dose | 2nd dose | 3rd dose | Other doses |
| ***(freetext)*** | 30 | 30 | 0 | 0 | 0 | 30 | 30 | 0 | 0 | 0 |
| **BLOOD AND LYMPHATIC SYSTEM DISORDERS** | | | | | | | | | | |
| ***COAGULOPATHIES AND BLEEDING DIATHESES (EXCL THROMBOCYTOPENIC)*** | | | | | | | | | | |
| *BLEEDING TENDENCIES* | | | | | | | | | | |
| INCREASED TENDENCY TO BRUISE | 0 | 0 | 0 | 0 | 0 | 0 | 0 | 0 | 0 | 0 |
| ***HAEMOGLOBINOPATHIES*** | | | | | | | | | | |
| *SICKLE CELL TRAIT AND DISORDERS* | | | | | | | | | | |
| SICKLE CELL ANAEMIA WITH CRISIS | 0 | 0 | 0 | 0 | 0 | 0 | 0 | 0 | 0 | 0 |
| ***PLATELET DISORDERS*** | | | | | | | | | | |
| *THROMBOCYTOPENIAS* | | | | | | | | | | |
| IMMUNE THROMBOCYTOPENIA | 0 | 0 | 0 | 0 | 0 | 0 | 0 | 0 | 0 | 0 |
| ***SPLEEN, LYMPHATIC AND RETICULOENDOTHELIAL SYSTEM DISORDERS*** | | | | | | | | | | |
| *LYMPHATIC SYSTEM DISORDERS NEC* | | | | | | | | | | |
| LYMPH NODE PAIN | 18 | 4 | 7 | 7 | 0 | 13 | 4 | 5 | 4 | 0 |
| LYMPHADENITIS | 1 | 0 | 1 | 0 | 0 | 1 | 0 | 1 | 0 | 0 |
| LYMPHADENOPATHY | 98 | 45 | 21 | 31 | 1 | 88 | 45 | 16 | 26 | 1 |
| **CARDIAC DISORDERS** | | | | | | | | | | |
| ***CARDIAC ARRHYTHMIAS*** | | | | | | | | | | |
| *RATE AND RHYTHM DISORDERS NEC* | | | | | | | | | | |
| ARRHYTHMIA | 0 | 0 | 0 | 0 | 0 | 0 | 0 | 0 | 0 | 0 |
| CARDIAC FLUTTER | 7 | 2 | 3 | 2 | 0 | 4 | 2 | 1 | 1 | 0 |
| EXTRASYSTOLES | 2 | 0 | 1 | 1 | 0 | 2 | 0 | 1 | 1 | 0 |
| TACHYCARDIA | 1 | 1 | 0 | 0 | 0 | 1 | 1 | 0 | 0 | 0 |
| *SUPRAVENTRICULAR ARRHYTHMIAS* | | | | | | | | | | |
| ATRIAL FIBRILLATION | 0 | 0 | 0 | 0 | 0 | 0 | 0 | 0 | 0 | 0 |
| SUPRAVENTRICULAR TACHYCARDIA | 0 | 0 | 0 | 0 | 0 | 0 | 0 | 0 | 0 | 0 |
| ***CARDIAC DISORDERS, SIGNS AND SYMPTOMS NEC*** | | | | | | | | | | |

| | | | | | | | | | | |
|---|---|---|---|---|---|---|---|---|---|---|
| *CARDIAC DISORDERS NEC* | | | | | | | | | | |
| CARDIOVASCULAR DISORDER | 0 | 0 | 0 | 0 | 0 | 0 | 0 | 0 | 0 | 0 |
| *CARDIAC SIGNS AND SYMPTOMS NEC* | | | | | | | | | | |
| PALPITATIONS | 21 | 7 | 9 | 4 | 1 | 17 | 7 | 5 | 4 | 1 |
| ***CORONARY ARTERY DISORDERS*** | | | | | | | | | | |
| *ISCHAEMIC CORONARY ARTERY DISORDERS* | | | | | | | | | | |
| ANGINA PECTORIS | 1 | 1 | 0 | 0 | 0 | 1 | 1 | 0 | 0 | 0 |
| MYOCARDIAL INFARCTION | 1 | 1 | 0 | 0 | 0 | 1 | 1 | 0 | 0 | 0 |
| ***HEART FAILURES*** | | | | | | | | | | |
| *HEART FAILURES NEC* | | | | | | | | | | |
| CARDIAC FAILURE | 1 | 0 | 0 | 1 | 0 | 0 | 0 | 0 | 0 | 0 |
| ***MYOCARDIAL DISORDERS*** | | | | | | | | | | |
| *NONINFECTIOUS MYOCARDITIS* | | | | | | | | | | |
| MYOCARDITIS | 2 | 1 | 1 | 0 | 0 | 1 | 1 | 0 | 0 | 0 |
| ***PERICARDIAL DISORDERS*** | | | | | | | | | | |
| *NONINFECTIOUS PERICARDITIS* | | | | | | | | | | |
| PERICARDITIS | 1 | 1 | 0 | 0 | 0 | 1 | 1 | 0 | 0 | 0 |
| **CONGENITAL, FAMILIAL AND GENETIC DISORDERS** | | | | | | | | | | |
| ***CARDIAC AND VASCULAR DISORDERS CONGENITAL*** | | | | | | | | | | |
| *CARDIAC DISORDERS CONGENITAL NEC* | | | | | | | | | | |
| HEART DISEASE CONGENITAL | 1 | 0 | 0 | 1 | 0 | 1 | 0 | 0 | 1 | 0 |
| ***METABOLIC AND NUTRITIONAL DISORDERS CONGENITAL*** | | | | | | | | | | |
| *INBORN ERRORS OF AMINO ACID METABOLISM* | | | | | | | | | | |
| HYPERGLYCINAEMIA | 0 | 0 | 0 | 0 | 0 | 0 | 0 | 0 | 0 | 0 |
| *INBORN ERRORS OF STEROID SYNTHESIS* | | | | | | | | | | |
| 11-BETA-HYDROXYLASE DEFICIENCY | 0 | 0 | 0 | 0 | 0 | 0 | 0 | 0 | 0 | 0 |
| ***NEUROLOGICAL DISORDERS CONGENITAL*** | | | | | | | | | | |
| *PERIPHERAL NERVOUS SYSTEM DISORDERS CONGENITAL NEC* | | | | | | | | | | |
| PAROXYSMAL EXTREME PAIN DISORDER | 0 | 0 | 0 | 0 | 0 | 0 | 0 | 0 | 0 | 0 |
| **EAR AND LABYRINTH DISORDERS** | | | | | | | | | | |
| ***AURAL DISORDERS NEC*** | | | | | | | | | | |
| *EAR DISORDERS NEC* | | | | | | | | | | |
| EAR DISCOMFORT | 1 | 0 | 1 | 0 | 0 | 1 | 0 | 1 | 0 | 0 |

| | | | | | | | | | | |
|---|---|---|---|---|---|---|---|---|---|---|
| EAR DISORDER | 0 | 0 | 0 | 0 | 0 | 0 | 0 | 0 | 0 | 0 |
| EAR PAIN | 9 | 7 | 1 | 1 | 0 | 8 | 7 | 1 | 0 | 0 |
| EAR SWELLING | 0 | 0 | 0 | 0 | 0 | 0 | 0 | 0 | 0 | 0 |
| ***EXTERNAL EAR DISORDERS (EXCL CONGENITAL)*** | | | | | | | | | | |
| *EXTERNAL EAR DISORDERS NEC* | | | | | | | | | | |
| EXCESSIVE CERUMEN PRODUCTION | 0 | 0 | 0 | 0 | 0 | 0 | 0 | 0 | 0 | 0 |
| ***HEARING DISORDERS*** | | | | | | | | | | |
| *HEARING LOSSES* | | | | | | | | | | |
| DEAFNESS | 3 | 2 | 1 | 0 | 0 | 3 | 2 | 1 | 0 | 0 |
| DEAFNESS NEUROSENSORY | 1 | 1 | 0 | 0 | 0 | 1 | 1 | 0 | 0 | 0 |
| DEAFNESS PERMANENT | 1 | 0 | 1 | 0 | 0 | 0 | 0 | 0 | 0 | 0 |
| DEAFNESS UNILATERAL | 0 | 0 | 0 | 0 | 0 | 0 | 0 | 0 | 0 | 0 |
| HYPOACUSIS | 1 | 1 | 0 | 0 | 0 | 1 | 1 | 0 | 0 | 0 |
| SUDDEN HEARING LOSS | 0 | 0 | 0 | 0 | 0 | 0 | 0 | 0 | 0 | 0 |
| *HYPERACUSIA* | | | | | | | | | | |
| HYPERACUSIS | 0 | 0 | 0 | 0 | 0 | 0 | 0 | 0 | 0 | 0 |
| ***INNER EAR AND VIIITH CRANIAL NERVE DISORDERS*** | | | | | | | | | | |
| *INNER EAR SIGNS AND SYMPTOMS* | | | | | | | | | | |
| MOTION SICKNESS | 1 | 1 | 0 | 0 | 0 | 1 | 1 | 0 | 0 | 0 |
| TINNITUS | 18 | 9 | 4 | 5 | 0 | 17 | 9 | 4 | 4 | 0 |
| VERTIGO | 10 | 6 | 3 | 1 | 0 | 10 | 6 | 3 | 1 | 0 |
| VERTIGO LABYRINTHINE | 0 | 0 | 0 | 0 | 0 | 0 | 0 | 0 | 0 | 0 |
| VERTIGO POSITIONAL | 0 | 0 | 0 | 0 | 0 | 0 | 0 | 0 | 0 | 0 |
| **ENDOCRINE DISORDERS** | | | | | | | | | | |
| ***THYROID GLAND DISORDERS*** | | | | | | | | | | |
| *THYROID HYPERFUNCTION DISORDERS* | | | | | | | | | | |
| HYPERTHYROIDISM | 0 | 0 | 0 | 0 | 0 | 0 | 0 | 0 | 0 | 0 |
| **EYE DISORDERS** | | | | | | | | | | |
| ***EYE DISORDERS NEC*** | | | | | | | | | | |
| *LACRIMATION DISORDERS* | | | | | | | | | | |
| DRY EYE | 5 | 0 | 1 | 4 | 0 | 4 | 0 | 1 | 3 | 0 |
| LACRIMATION INCREASED | 3 | 2 | 0 | 1 | 0 | 2 | 2 | 0 | 0 | 0 |
| *OCULAR DISORDERS NEC* | | | | | | | | | | |
| EYE OEDEMA | 0 | 0 | 0 | 0 | 0 | 0 | 0 | 0 | 0 | 0 |
| EYE PAIN | 22 | 14 | 3 | 5 | 0 | 22 | 14 | 3 | 5 | 0 |
| EYE SWELLING | 2 | 2 | 0 | 0 | 0 | 2 | 2 | 0 | 0 | 0 |
| EYE ULCER | 0 | 0 | 0 | 0 | 0 | 0 | 0 | 0 | 0 | 0 |
| EYELID PAIN | 0 | 0 | 0 | 0 | 0 | 0 | 0 | 0 | 0 | 0 |

| | | | | | | | | | | |
|---|---|---|---|---|---|---|---|---|---|---|
| OCULAR DISCOMFORT | 0 | 0 | 0 | 0 | 0 | 0 | 0 | 0 | 0 | 0 |
| PERIORBITAL DISCOMFORT | 0 | 0 | 0 | 0 | 0 | 0 | 0 | 0 | 0 | 0 |
| PERIORBITAL SWELLING | 0 | 0 | 0 | 0 | 0 | 0 | 0 | 0 | 0 | 0 |
| ***OCULAR HAEMORRHAGES AND VASCULAR DISORDERS NEC*** | | | | | | | | | | |
| *CONJUNCTIVAL AND CORNEAL BLEEDING AND VASCULAR DISORDERS* | | | | | | | | | | |
| CONJUNCTIVAL HAEMORRHAGE | 1 | 1 | 0 | 0 | 0 | 1 | 1 | 0 | 0 | 0 |
| *LID BLEEDING AND VASCULAR DISORDERS* | | | | | | | | | | |
| EYELID BLEEDING | 0 | 0 | 0 | 0 | 0 | 0 | 0 | 0 | 0 | 0 |
| ***OCULAR INFECTIONS, IRRITATIONS AND INFLAMMATIONS*** | | | | | | | | | | |
| *LID, LASH AND LACRIMAL INFECTIONS, IRRITATIONS AND INFLAMMATIONS* | | | | | | | | | | |
| BLEPHARITIS | 0 | 0 | 0 | 0 | 0 | 0 | 0 | 0 | 0 | 0 |
| ERYTHEMA OF EYELID | 0 | 0 | 0 | 0 | 0 | 0 | 0 | 0 | 0 | 0 |
| EYELID IRRITATION | 0 | 0 | 0 | 0 | 0 | 0 | 0 | 0 | 0 | 0 |
| *OCULAR INFECTIONS, INFLAMMATIONS AND ASSOCIATED MANIFESTATIONS* | | | | | | | | | | |
| EYE DISCHARGE | 0 | 0 | 0 | 0 | 0 | 0 | 0 | 0 | 0 | 0 |
| EYE IRRITATION | 0 | 0 | 0 | 0 | 0 | 0 | 0 | 0 | 0 | 0 |
| EYE PRURITUS | 5 | 4 | 0 | 1 | 0 | 4 | 4 | 0 | 0 | 0 |
| LIMBAL SWELLING | 0 | 0 | 0 | 0 | 0 | 0 | 0 | 0 | 0 | 0 |
| OCULAR HYPERAEMIA | 2 | 1 | 1 | 0 | 0 | 2 | 1 | 1 | 0 | 0 |
| ***OCULAR NEUROMUSCULAR DISORDERS*** | | | | | | | | | | |
| *EYELID MOVEMENT DISORDERS* | | | | | | | | | | |
| BLEPHAROSPASM | 1 | 1 | 0 | 0 | 0 | 1 | 1 | 0 | 0 | 0 |
| ***OCULAR SENSORY SYMPTOMS NEC*** | | | | | | | | | | |
| *OCULAR SENSATION DISORDERS* | | | | | | | | | | |
| ABNORMAL SENSATION IN EYE | 2 | 1 | 1 | 0 | 0 | 2 | 1 | 1 | 0 | 0 |
| ASTHENOPIA | 11 | 6 | 2 | 3 | 0 | 8 | 6 | 0 | 2 | 0 |
| FOREIGN BODY SENSATION IN EYES | 0 | 0 | 0 | 0 | 0 | 0 | 0 | 0 | 0 | 0 |
| PHOTOPHOBIA | 1 | 1 | 0 | 0 | 0 | 1 | 1 | 0 | 0 | 0 |
| ***OCULAR STRUCTURAL CHANGE, DEPOSIT AND DEGENERATION NEC*** | | | | | | | | | | |
| *CHOROID AND VITREOUS STRUCTURAL CHANGE, DEPOSIT AND DEGENERATION* | | | | | | | | | | |
| VITREOUS DETACHMENT | 0 | 0 | 0 | 0 | 0 | 0 | 0 | 0 | 0 | 0 |
| VITREOUS FLOATERS | 0 | 0 | 0 | 0 | 0 | 0 | 0 | 0 | 0 | 0 |

| | | | | | | | | | | |
|---|---|---|---|---|---|---|---|---|---|---|
| ***RETINA, CHOROID AND VITREOUS HAEMORRHAGES AND VASCULAR DISORDERS*** | | | | | | | | | | |
| *RETINAL BLEEDING AND VASCULAR DISORDERS (EXCL RETINOPATHY)* | | | | | | | | | | |
| RETINAL VEIN OCCLUSION | 1 | 0 | 1 | 0 | 0 | 1 | 0 | 1 | 0 | 0 |
| *RETINOPATHIES NEC* | | | | | | | | | | |
| RETINAL EXUDATES | 0 | 0 | 0 | 0 | 0 | 0 | 0 | 0 | 0 | 0 |
| ***VISION DISORDERS*** | | | | | | | | | | |
| *VISUAL DISORDERS NEC* | | | | | | | | | | |
| DIPLOPIA | 0 | 0 | 0 | 0 | 0 | 0 | 0 | 0 | 0 | 0 |
| HALO VISION | 0 | 0 | 0 | 0 | 0 | 0 | 0 | 0 | 0 | 0 |
| METAMORPHOPSIA | 0 | 0 | 0 | 0 | 0 | 0 | 0 | 0 | 0 | 0 |
| PHOTOPSIA | 1 | 1 | 0 | 0 | 0 | 1 | 1 | 0 | 0 | 0 |
| VISION BLURRED | 10 | 6 | 4 | 0 | 0 | 9 | 6 | 3 | 0 | 0 |
| *VISUAL IMPAIRMENT AND BLINDNESS (EXCL COLOUR BLINDNESS)* | | | | | | | | | | |
| BLINDNESS | 0 | 0 | 0 | 0 | 0 | 0 | 0 | 0 | 0 | 0 |
| BLINDNESS TRANSIENT | 0 | 0 | 0 | 0 | 0 | 0 | 0 | 0 | 0 | 0 |
| SUDDEN VISUAL LOSS | 0 | 0 | 0 | 0 | 0 | 0 | 0 | 0 | 0 | 0 |
| VISUAL IMPAIRMENT | 5 | 5 | 0 | 0 | 0 | 5 | 5 | 0 | 0 | 0 |
| **GASTROINTESTINAL DISORDERS** | | | | | | | | | | |
| ***ANAL AND RECTAL CONDITIONS NEC*** | | | | | | | | | | |
| *ANAL AND RECTAL SIGNS AND SYMPTOMS* | | | | | | | | | | |
| ANAL PARAESTHESIA | 0 | 0 | 0 | 0 | 0 | 0 | 0 | 0 | 0 | 0 |
| ***BENIGN NEOPLASMS GASTROINTESTINAL*** | | | | | | | | | | |
| *BENIGN ORAL CAVITY NEOPLASMS* | | | | | | | | | | |
| MOUTH CYST | 0 | 0 | 0 | 0 | 0 | 0 | 0 | 0 | 0 | 0 |
| ***DENTAL AND GINGIVAL CONDITIONS*** | | | | | | | | | | |
| *DENTAL DISORDERS NEC* | | | | | | | | | | |
| TEETHING | 1 | 1 | 0 | 0 | 0 | 1 | 1 | 0 | 0 | 0 |
| *DENTAL PAIN AND SENSATION DISORDERS* | | | | | | | | | | |
| DENTAL PARAESTHESIA | 0 | 0 | 0 | 0 | 0 | 0 | 0 | 0 | 0 | 0 |
| TOOTHACHE | 4 | 0 | 2 | 2 | 0 | 3 | 0 | 1 | 2 | 0 |
| *GINGIVAL DISORDERS, SIGNS AND SYMPTOMS NEC* | | | | | | | | | | |
| GINGIVAL BLISTER | 0 | 0 | 0 | 0 | 0 | 0 | 0 | 0 | 0 | 0 |
| GINGIVAL PAIN | 0 | 0 | 0 | 0 | 0 | 0 | 0 | 0 | 0 | 0 |
| GINGIVAL SWELLING | 0 | 0 | 0 | 0 | 0 | 0 | 0 | 0 | 0 | 0 |
| ***GASTROINTESTINAL CONDITIONS NEC*** | | | | | | | | | | |

| | | | | | | | | | | |
|---|---|---|---|---|---|---|---|---|---|---|
| *GASTROINTESTINAL MUCOSAL DYSTROPHIES AND SECRETION DISORDERS* | | | | | | | | | | |
| BARRETT'S OESOPHAGUS | 0 | 0 | 0 | 0 | 0 | 0 | 0 | 0 | 0 | 0 |
| ***GASTROINTESTINAL HAEMORRHAGES NEC*** | | | | | | | | | | |
| *INTESTINAL HAEMORRHAGES* | 0 | 0 | 0 | 0 | 0 | 0 | 0 | 0 | 0 | 0 |
| RECTAL HAEMORRHAGE | 0 | 0 | 0 | 0 | 0 | 0 | 0 | 0 | 0 | 0 |
| *NON-SITE SPECIFIC GASTROINTESTINAL HAEMORRHAGES* | | | | | | | | | | |
| HAEMATEMESIS | 0 | 0 | 0 | 0 | 0 | 0 | 0 | 0 | 0 | 0 |
| ***GASTROINTESTINAL INFLAMMATORY CONDITIONS*** | | | | | | | | | | |
| *COLITIS (EXCL INFECTIVE)* | | | | | | | | | | |
| COLITIS | 1 | 0 | 1 | 0 | 0 | 1 | 0 | 1 | 0 | 0 |
| COLITIS MICROSCOPIC | 0 | 0 | 0 | 0 | 0 | 0 | 0 | 0 | 0 | 0 |
| COLITIS ULCERATIVE | 1 | 0 | 1 | 0 | 0 | 0 | 0 | 0 | 0 | 0 |
| *GASTRITIS (EXCL INFECTIVE)* | | | | | | | | | | |
| GASTRITIS | 0 | 0 | 0 | 0 | 0 | 0 | 0 | 0 | 0 | 0 |
| REFLUX GASTRITIS | 0 | 0 | 0 | 0 | 0 | 0 | 0 | 0 | 0 | 0 |
| *GASTROINTESTINAL INFLAMMATORY DISORDERS NEC* | | | | | | | | | | |
| GASTROINTESTINAL TRACT IRRITATION | 1 | 0 | 0 | 1 | 0 | 1 | 0 | 0 | 1 | 0 |
| ***GASTROINTESTINAL MOTILITY AND DEFAECATION CONDITIONS*** | | | | | | | | | | |
| *DIARRHOEA (EXCL INFECTIVE)* | | | | | | | | | | |
| DIARRHOEA | 104 | 57 | 18 | 28 | 1 | 95 | 57 | 14 | 23 | 1 |
| *GASTROINTESTINAL ATONIC AND HYPOMOTILITY DISORDERS NEC* | | | | | | | | | | |
| CONSTIPATION | 5 | 3 | 0 | 2 | 0 | 4 | 3 | 0 | 1 | 0 |
| GASTROOESOPHAGEAL REFLUX DISEASE | 0 | 0 | 0 | 0 | 0 | 0 | 0 | 0 | 0 | 0 |
| *GASTROINTESTINAL DYSKINETIC DISORDERS* | | | | | | | | | | |
| CHANGE OF BOWEL HABIT | 2 | 1 | 1 | 0 | 0 | 2 | 1 | 1 | 0 | 0 |
| *GASTROINTESTINAL SPASTIC AND HYPERMOTILITY DISORDERS* | | | | | | | | | | |
| IRRITABLE BOWEL SYNDROME | 1 | 0 | 0 | 1 | 0 | 1 | 0 | 0 | 1 | 0 |
| ***GASTROINTESTINAL SIGNS AND SYMPTOMS*** | | | | | | | | | | |
| *DYSPEPTIC SIGNS AND SYMPTOMS* | | | | | | | | | | |
| DYSPEPSIA | 5 | 2 | 0 | 3 | 0 | 5 | 2 | 0 | 3 | 0 |
| ERUCTATION | 2 | 2 | 0 | 0 | 0 | 2 | 2 | 0 | 0 | 0 |
| *FAECAL ABNORMALITIES NEC* | | | | | | | | | | |
| FAECALOMA | 1 | 0 | 1 | 0 | 0 | 1 | 0 | 1 | 0 | 0 |
| FAECES DISCOLOURED | 1 | 1 | 0 | 0 | 0 | 1 | 1 | 0 | 0 | 0 |
| FAECES SOFT | 0 | 0 | 0 | 0 | 0 | 0 | 0 | 0 | 0 | 0 |
| *FLATULENCE, BLOATING AND DISTENSION* | | | | | | | | | | |
| ABDOMINAL DISTENSION | 1 | 0 | 0 | 0 | 1 | 1 | 0 | 0 | 0 | 1 |

| | | | | | | | | | | |
|---|---|---|---|---|---|---|---|---|---|---|
| FLATULENCE | 3 | 2 | 0 | 1 | 0 | 2 | 2 | 0 | 0 | 0 |
| *GASTROINTESTINAL AND ABDOMINAL PAINS (EXCL ORAL AND THROAT)* | | | | | | | | | | |
| ABDOMINAL PAIN | 3 | 2 | 1 | 0 | 0 | 3 | 2 | 1 | 0 | 0 |
| ABDOMINAL PAIN LOWER | 0 | 0 | 0 | 0 | 0 | 0 | 0 | 0 | 0 | 0 |
| ABDOMINAL PAIN UPPER | 31 | 21 | 4 | 5 | 1 | 29 | 21 | 3 | 5 | 0 |
| GASTROINTESTINAL PAIN | 3 | 2 | 0 | 1 | 0 | 2 | 2 | 0 | 0 | 0 |
| *GASTROINTESTINAL SIGNS AND SYMPTOMS NEC* | | | | | | | | | | |
| ABDOMINAL DISCOMFORT | 17 | 8 | 4 | 5 | 0 | 16 | 8 | 3 | 5 | 0 |
| ABDOMINAL SYMPTOM | 0 | 0 | 0 | 0 | 0 | 0 | 0 | 0 | 0 | 0 |
| ACUTE ABDOMEN | 0 | 0 | 0 | 0 | 0 | 0 | 0 | 0 | 0 | 0 |
| ANAL INCONTINENCE | 1 | 1 | 0 | 0 | 0 | 1 | 1 | 0 | 0 | 0 |
| BREATH ODOUR | 1 | 1 | 0 | 0 | 0 | 1 | 1 | 0 | 0 | 0 |
| DYSPHAGIA | 2 | 1 | 1 | 0 | 0 | 1 | 1 | 0 | 0 | 0 |
| ODYNOPHAGIA | 1 | 0 | 1 | 0 | 0 | 0 | 0 | 0 | 0 | 0 |
| *NAUSEA AND VOMITING SYMPTOMS* | | | | | | | | | | |
| NAUSEA | 304 | 174 | 58 | 65 | 7 | 285 | 174 | 48 | 56 | 7 |
| RETCHING | 0 | 0 | 0 | 0 | 0 | 0 | 0 | 0 | 0 | 0 |
| VOMITING | 43 | 27 | 6 | 8 | 2 | 41 | 27 | 6 | 7 | 1 |
| VOMITING PROJECTILE | 0 | 0 | 0 | 0 | 0 | 0 | 0 | 0 | 0 | 0 |
| ***MALABSORPTION CONDITIONS*** | | | | | | | | | | |
| *MALABSORPTION SYNDROMES* | | | | | | | | | | |
| COELIAC DISEASE | 0 | 0 | 0 | 0 | 0 | 0 | 0 | 0 | 0 | 0 |
| ***ORAL SOFT TISSUE CONDITIONS*** | | | | | | | | | | |
| *ORAL SOFT TISSUE DISORDERS NEC* | | | | | | | | | | |
| CHEILITIS | 0 | 0 | 0 | 0 | 0 | 0 | 0 | 0 | 0 | 0 |
| LIP BLISTER | 0 | 0 | 0 | 0 | 0 | 0 | 0 | 0 | 0 | 0 |
| ORAL LICHEN PLANUS | 0 | 0 | 0 | 0 | 0 | 0 | 0 | 0 | 0 | 0 |
| *ORAL SOFT TISSUE INFECTIONS* | | | | | | | | | | |
| ANGULAR CHEILITIS | 0 | 0 | 0 | 0 | 0 | 0 | 0 | 0 | 0 | 0 |
| *ORAL SOFT TISSUE SIGNS AND SYMPTOMS* | | | | | | | | | | |
| HYPOAESTHESIA ORAL | 5 | 4 | 1 | 0 | 0 | 4 | 4 | 0 | 0 | 0 |
| LIP PAIN | 0 | 0 | 0 | 0 | 0 | 0 | 0 | 0 | 0 | 0 |
| ORAL DISCOMFORT | 1 | 0 | 1 | 0 | 0 | 0 | 0 | 0 | 0 | 0 |
| ORAL MUCOSAL ROUGHENING | 1 | 1 | 0 | 0 | 0 | 1 | 1 | 0 | 0 | 0 |
| ORAL PAIN | 2 | 0 | 1 | 1 | 0 | 2 | 0 | 1 | 1 | 0 |

| | | | | | | | | | | |
|---|---|---|---|---|---|---|---|---|---|---|
| PARAESTHESIA ORAL | 8 | 7 | 1 | 0 | 0 | 8 | 7 | 1 | 0 | 0 |
| LIP SWELLING | 2 | 1 | 0 | 1 | 0 | 2 | 1 | 0 | 1 | 0 |
| MOUTH SWELLING | 0 | 0 | 0 | 0 | 0 | 0 | 0 | 0 | 0 | 0 |
| *STOMATITIS AND ULCERATION* | | | | | | | | | | |
| APHTHOUS ULCER | 0 | 0 | 0 | 0 | 0 | 0 | 0 | 0 | 0 | 0 |
| LIP ULCERATION | 1 | 0 | 0 | 0 | 1 | 0 | 0 | 0 | 0 | 0 |
| MOUTH ULCERATION | 7 | 4 | 1 | 1 | 1 | 7 | 4 | 1 | 1 | 1 |
| STOMATITIS | 1 | 1 | 0 | 0 | 0 | 1 | 1 | 0 | 0 | 0 |
| ***SALIVARY GLAND CONDITIONS*** | | | | | | | | | | |
| *ORAL DRYNESS AND SALIVA ALTERED* | | | | | | | | | | |
| DRY MOUTH | 6 | 5 | 0 | 0 | 1 | 5 | 5 | 0 | 0 | 0 |
| LIP DRY | 0 | 0 | 0 | 0 | 0 | 0 | 0 | 0 | 0 | 0 |
| SALIVARY HYPOSECRETION | 0 | 0 | 0 | 0 | 0 | 0 | 0 | 0 | 0 | 0 |
| ***TONGUE CONDITIONS*** | | | | | | | | | | |
| *TONGUE SIGNS AND SYMPTOMS* | | | | | | | | | | |
| GLOSSODYNIA | 5 | 4 | 0 | 1 | 0 | 5 | 4 | 0 | 1 | 0 |
| SWOLLEN TONGUE | 0 | 0 | 0 | 0 | 0 | 0 | 0 | 0 | 0 | 0 |
| TONGUE COATED | 2 | 2 | 0 | 0 | 0 | 2 | 2 | 0 | 0 | 0 |
| TONGUE DISCOMFORT | 1 | 0 | 1 | 0 | 0 | 0 | 0 | 0 | 0 | 0 |
| TONGUE OEDEMA | 1 | 0 | 0 | 1 | 0 | 1 | 0 | 0 | 1 | 0 |
| TONGUE SPASM | 0 | 0 | 0 | 0 | 0 | 0 | 0 | 0 | 0 | 0 |
| **GENERAL DISORDERS AND ADMINISTRATION SITE CONDITIONS** | | | | | | | | | | |
| ***ADMINISTRATION SITE REACTIONS*** | | | | | | | | | | |
| *ADMINISTRATION SITE REACTIONS NEC* | | | | | | | | | | |
| ADMINISTRATION SITE BRUISE | 0 | 0 | 0 | 0 | 0 | 0 | 0 | 0 | 0 | 0 |
| ADMINISTRATION SITE PAIN | 1 | 1 | 0 | 0 | 0 | 1 | 1 | 0 | 0 | 0 |
| PUNCTURE SITE BRUISE | 6 | 4 | 1 | 1 | 0 | 6 | 4 | 1 | 1 | 0 |
| PUNCTURE SITE PAIN | 1 | 1 | 0 | 0 | 0 | 1 | 1 | 0 | 0 | 0 |
| *APPLICATION SITE REACTIONS* | | | | | | | | | | |
| APPLICATION SITE BRUISE | 3 | 1 | 1 | 1 | 0 | 2 | 1 | 0 | 1 | 0 |
| APPLICATION SITE ERYTHEMA | 0 | 0 | 0 | 0 | 0 | 0 | 0 | 0 | 0 | 0 |
| APPLICATION SITE PAIN | 1 | 0 | 0 | 1 | 0 | 0 | 0 | 0 | 0 | 0 |
| *IMPLANT AND CATHETER SITE REACTIONS* | | | | | | | | | | |
| IMPLANT SITE PAIN | 1 | 0 | 1 | 0 | 0 | 0 | 0 | 0 | 0 | 0 |
| IMPLANT SITE WARMTH | 0 | 0 | 0 | 0 | 0 | 0 | 0 | 0 | 0 | 0 |

| | | | | | | | | | | |
|---|---|---|---|---|---|---|---|---|---|---|
| *INFUSION SITE REACTIONS* | | | | | | | | | | |
| INFUSION SITE PAIN | 1 | 0 | 0 | 1 | 0 | 1 | 0 | 0 | 1 | 0 |
| INFUSION SITE SCAB | 0 | 0 | 0 | 0 | 0 | 0 | 0 | 0 | 0 | 0 |
| INFUSION SITE WARMTH | 0 | 0 | 0 | 0 | 0 | 0 | 0 | 0 | 0 | 0 |
| *INJECTION SITE REACTIONS* | | | | | | | | | | |
| INJECTION SITE BRUISING | 3 | 2 | 1 | 0 | 0 | 3 | 2 | 1 | 0 | 0 |
| INJECTION SITE DISCOMFORT | 2 | 1 | 1 | 0 | 0 | 1 | 1 | 0 | 0 | 0 |
| INJECTION SITE ERYTHEMA | 11 | 6 | 2 | 2 | 1 | 11 | 6 | 2 | 2 | 1 |
| INJECTION SITE HYPERSENSITIVITY | 1 | 0 | 1 | 0 | 0 | 1 | 0 | 1 | 0 | 0 |
| INJECTION SITE INFLAMMATION | 0 | 0 | 0 | 0 | 0 | 0 | 0 | 0 | 0 | 0 |
| INJECTION SITE INJURY | 1 | 1 | 0 | 0 | 0 | 1 | 1 | 0 | 0 | 0 |
| INJECTION SITE IRRITATION | 1 | 0 | 1 | 0 | 0 | 1 | 0 | 1 | 0 | 0 |
| INJECTION SITE JOINT PAIN | 0 | 0 | 0 | 0 | 0 | 0 | 0 | 0 | 0 | 0 |
| INJECTION SITE MASS | 12 | 2 | 5 | 4 | 1 | 11 | 2 | 5 | 3 | 1 |
| INJECTION SITE NODULE | 1 | 0 | 0 | 1 | 0 | 1 | 0 | 0 | 1 | 0 |
| INJECTION SITE OEDEMA | 0 | 0 | 0 | 0 | 0 | 0 | 0 | 0 | 0 | 0 |
| INJECTION SITE PAIN | 237 | 155 | 39 | 41 | 2 | 216 | 155 | 31 | 28 | 2 |
| INJECTION SITE PAPULE | 0 | 0 | 0 | 0 | 0 | 0 | 0 | 0 | 0 | 0 |
| INJECTION SITE PARAESTHESIA | 1 | 1 | 0 | 0 | 0 | 1 | 1 | 0 | 0 | 0 |
| INJECTION SITE PRURITUS | 4 | 3 | 0 | 1 | 0 | 4 | 3 | 0 | 1 | 0 |
| INJECTION SITE RASH | 2 | 1 | 0 | 1 | 0 | 2 | 1 | 0 | 1 | 0 |
| INJECTION SITE REACTION | 2 | 0 | 2 | 0 | 0 | 2 | 0 | 2 | 0 | 0 |
| INJECTION SITE SCAB | 0 | 0 | 0 | 0 | 0 | 0 | 0 | 0 | 0 | 0 |
| INJECTION SITE SWELLING | 6 | 3 | 0 | 3 | 0 | 6 | 3 | 0 | 3 | 0 |
| INJECTION SITE URTICARIA | 2 | 0 | 2 | 0 | 0 | 2 | 0 | 2 | 0 | 0 |
| INJECTION SITE WARMTH | 2 | 1 | 1 | 0 | 0 | 2 | 1 | 1 | 0 | 0 |
| *INSTILLATION SITE REACTIONS* | | | | | | | | | | |
| INSTILLATION SITE PRURITUS | 1 | 0 | 1 | 0 | 0 | 1 | 0 | 1 | 0 | 0 |
| INSTILLATION SITE WARMTH | 1 | 1 | 0 | 0 | 0 | 1 | 1 | 0 | 0 | 0 |
| *VACCINATION SITE REACTIONS* | | | | | | | | | | |
| SHOULDER INJURY RELATED TO VACCINE ADMINISTRATION | 0 | 0 | 0 | 0 | 0 | 0 | 0 | 0 | 0 | 0 |
| VACCINATION SITE BRUISING | 0 | 0 | 0 | 0 | 0 | 0 | 0 | 0 | 0 | 0 |
| VACCINATION SITE DISCOMFORT | 1 | 1 | 0 | 0 | 0 | 1 | 1 | 0 | 0 | 0 |
| VACCINATION SITE ERYTHEMA | 1 | 1 | 0 | 0 | 0 | 1 | 1 | 0 | 0 | 0 |

| | | | | | | | | | | |
|---|---|---|---|---|---|---|---|---|---|---|
| VACCINATION SITE JOINT ERYTHEMA | 0 | 0 | 0 | 0 | 0 | 0 | 0 | 0 | 0 | 0 |
| VACCINATION SITE JOINT PAIN | 0 | 0 | 0 | 0 | 0 | 0 | 0 | 0 | 0 | 0 |
| VACCINATION SITE MASS | 2 | 1 | 0 | 1 | 0 | 1 | 1 | 0 | 0 | 0 |
| VACCINATION SITE PAIN | 27 | 17 | 2 | 8 | 0 | 27 | 17 | 2 | 8 | 0 |
| VACCINATION SITE RASH | 0 | 0 | 0 | 0 | 0 | 0 | 0 | 0 | 0 | 0 |
| VACCINATION SITE SWELLING | 2 | 1 | 1 | 0 | 0 | 2 | 1 | 1 | 0 | 0 |
| VACCINATION SITE WARMTH | 1 | 1 | 0 | 0 | 0 | 1 | 1 | 0 | 0 | 0 |
| ***BODY TEMPERATURE CONDITIONS*** | | | | | | | | | | |
| *BODY TEMPERATURE ALTERED* | | | | | | | | | | |
| HYPERTHERMIA | 0 | 0 | 0 | 0 | 0 | 0 | 0 | 0 | 0 | 0 |
| HYPOTHERMIA | 1 | 1 | 0 | 0 | 0 | 1 | 1 | 0 | 0 | 0 |
| *FEBRILE DISORDERS* | | | | | | | | | | |
| PYREXIA | 338 | 156 | 88 | 84 | 10 | 307 | 156 | 74 | 69 | 8 |
| ***FATAL OUTCOMES*** | | | | | | | | | | |
| *DEATH AND SUDDEN DEATH* | | | | | | | | | | |
| DEATH | 1 | 1 | 0 | 0 | 0 | 1 | 1 | 0 | 0 | 0 |
| ***GENERAL SYSTEM DISORDERS NEC*** | | | | | | | | | | |
| *ADVERSE EFFECT ABSENT* | | | | | | | | | | |
| NO ADVERSE EVENT | 0 | 0 | 0 | 0 | 0 | 0 | 0 | 0 | 0 | 0 |
| *ASTHENIC CONDITIONS* | | | | | | | | | | |
| ASTHENIA | 34 | 21 | 6 | 7 | 0 | 29 | 21 | 4 | 4 | 0 |
| CHRONIC FATIGUE SYNDROME | 5 | 1 | 2 | 2 | 0 | 4 | 1 | 2 | 1 | 0 |
| DECREASED ACTIVITY | 1 | 1 | 0 | 0 | 0 | 1 | 1 | 0 | 0 | 0 |
| FATIGUE | 1180 | 654 | 277 | 229 | 20 | 1089 | 654 | 230 | 186 | 19 |
| MALAISE | 107 | 49 | 28 | 27 | 3 | 93 | 49 | 21 | 20 | 3 |
| SLUGGISHNESS | 0 | 0 | 0 | 0 | 0 | 0 | 0 | 0 | 0 | 0 |
| *FEELINGS AND SENSATIONS NEC* | | | | | | | | | | |
| CHILLS | 204 | 87 | 36 | 78 | 3 | 186 | 87 | 31 | 66 | 2 |
| FEELING ABNORMAL | 29 | 10 | 6 | 11 | 2 | 24 | 10 | 4 | 8 | 2 |
| FEELING COLD | 54 | 30 | 10 | 12 | 2 | 47 | 30 | 8 | 8 | 1 |
| FEELING HOT | 13 | 5 | 3 | 5 | 0 | 13 | 5 | 3 | 5 | 0 |
| FEELING JITTERY | 0 | 0 | 0 | 0 | 0 | 0 | 0 | 0 | 0 | 0 |
| FEELING OF BODY TEMPERATURE CHANGE | 12 | 8 | 2 | 2 | 0 | 12 | 8 | 2 | 2 | 0 |
| FEELING OF RELAXATION | 0 | 0 | 0 | 0 | 0 | 0 | 0 | 0 | 0 | 0 |
| HANGOVER | 2 | 1 | 0 | 1 | 0 | 2 | 1 | 0 | 1 | 0 |

| | | | | | | | | | |
|---|---|---|---|---|---|---|---|---|---|
| HUNGER | 1 | 1 | 0 | 0 | 0 | 1 | 1 | 0 | 0 | 0 |
| SENSATION OF BLOOD FLOW | 0 | 0 | 0 | 0 | 0 | 0 | 0 | 0 | 0 | 0 |
| THIRST | 6 | 3 | 1 | 2 | 0 | 5 | 3 | 0 | 2 | 0 |
| *GAIT DISTURBANCES* | | | | | | | | | | |
| GAIT DISTURBANCE | 2 | 1 | 1 | 0 | 0 | 2 | 1 | 1 | 0 | 0 |
| GAIT INABILITY | 1 | 1 | 0 | 0 | 0 | 1 | 1 | 0 | 0 | 0 |
| *GENERAL SIGNS AND SYMPTOMS NEC* | | | | | | | | | | |
| CRYING | 0 | 0 | 0 | 0 | 0 | 0 | 0 | 0 | 0 | 0 |
| ENERGY INCREASED | 0 | 0 | 0 | 0 | 0 | 0 | 0 | 0 | 0 | 0 |
| EXERCISE TOLERANCE DECREASED | 0 | 0 | 0 | 0 | 0 | 0 | 0 | 0 | 0 | 0 |
| GENERAL SYMPTOM | 1 | 1 | 0 | 0 | 0 | 1 | 1 | 0 | 0 | 0 |
| ILLNESS | 34 | 12 | 8 | 14 | 0 | 30 | 12 | 7 | 11 | 0 |
| INFLUENZA LIKE ILLNESS | 87 | 25 | 21 | 39 | 2 | 76 | 25 | 16 | 33 | 2 |
| LOCAL REACTION | 2 | 1 | 1 | 0 | 0 | 2 | 1 | 1 | 0 | 0 |
| PERIPHERAL SWELLING | 90 | 49 | 19 | 21 | 1 | 82 | 49 | 17 | 15 | 1 |
| SWELLING | 34 | 20 | 5 | 9 | 0 | 31 | 20 | 4 | 7 | 0 |
| SWELLING FACE | 4 | 4 | 0 | 0 | 0 | 4 | 4 | 0 | 0 | 0 |
| TISSUE IRRITATION | 0 | 0 | 0 | 0 | 0 | 0 | 0 | 0 | 0 | 0 |
| *INFLAMMATIONS* | | | | | | | | | | |
| INFLAMMATION | 5 | 3 | 2 | 0 | 0 | 4 | 3 | 1 | 0 | 0 |
| SYSTEMIC INFLAMMATORY RESPONSE SYNDROME | 0 | 0 | 0 | 0 | 0 | 0 | 0 | 0 | 0 | 0 |
| *OEDEMA NEC* | | | | | | | | | | |
| OEDEMA | 0 | 0 | 0 | 0 | 0 | 0 | 0 | 0 | 0 | 0 |
| OEDEMA PERIPHERAL | 1 | 0 | 1 | 0 | 0 | 1 | 0 | 1 | 0 | 0 |
| *PAIN AND DISCOMFORT NEC* | | | | | | | | | | |
| AXILLARY PAIN | 50 | 12 | 16 | 21 | 1 | 47 | 12 | 14 | 20 | 1 |
| CHEST DISCOMFORT | 9 | 6 | 0 | 3 | 0 | 9 | 6 | 0 | 3 | 0 |
| CHEST PAIN | 34 | 23 | 6 | 5 | 0 | 32 | 23 | 5 | 4 | 0 |
| DISCOMFORT | 10 | 3 | 3 | 3 | 1 | 8 | 3 | 2 | 3 | 0 |
| FACIAL PAIN | 1 | 0 | 0 | 1 | 0 | 0 | 0 | 0 | 0 | 0 |
| HERNIA PAIN | 1 | 1 | 0 | 0 | 0 | 1 | 1 | 0 | 0 | 0 |
| INFLAMMATORY PAIN | 2 | 1 | 0 | 0 | 1 | 1 | 1 | 0 | 0 | 0 |
| NON-CARDIAC CHEST PAIN | 1 | 0 | 0 | 1 | 0 | 1 | 0 | 0 | 1 | 0 |
| PAIN | 363 | 217 | 74 | 66 | 6 | 333 | 217 | 57 | 54 | 5 |
| TENDERNESS | 121 | 67 | 26 | 27 | 1 | 115 | 67 | 22 | 25 | 1 |

| | | | | | | | | | | |
|---|---|---|---|---|---|---|---|---|---|---|
| ***THERAPEUTIC AND NONTHERAPEUTIC EFFECTS (EXCL TOXICITY)*** | | | | | | | | | | |
| *THERAPEUTIC AND NONTHERAPEUTIC RESPONSES* | | | | | | | | | | |
| ADVERSE DRUG REACTION | 0 | 0 | 0 | 0 | 0 | 0 | 0 | 0 | 0 | 0 |
| ADVERSE EVENT | 0 | 0 | 0 | 0 | 0 | 0 | 0 | 0 | 0 | 0 |
| ADVERSE REACTION | 1 | 1 | 0 | 0 | 0 | 1 | 1 | 0 | 0 | 0 |
| IMMEDIATE POST-INJECTION REACTION | 1 | 0 | 1 | 0 | 0 | 1 | 0 | 1 | 0 | 0 |
| **HEPATOBILIARY DISORDERS** | | | | | | | | | | |
| ***HEPATIC AND HEPATOBILIARY DISORDERS*** | | | | | | | | | | |
| *HEPATOBILIARY SIGNS AND SYMPTOMS* | | | | | | | | | | |
| HEPATIC PAIN | 0 | 0 | 0 | 0 | 0 | 0 | 0 | 0 | 0 | 0 |
| *HEPATOCELLULAR DAMAGE AND HEPATITIS NEC* | | | | | | | | | | |
| LIVER INJURY | 0 | 0 | 0 | 0 | 0 | 0 | 0 | 0 | 0 | 0 |
| **IMMUNE SYSTEM DISORDERS** | | | | | | | | | | |
| ***ALLERGIC CONDITIONS*** | | | | | | | | | | |
| *ALLERGIC CONDITIONS NEC* | | | | | | | | | | |
| HYPERSENSITIVITY | 1 | 0 | 0 | 1 | 0 | 1 | 0 | 0 | 1 | 0 |
| MULTIPLE ALLERGIES | 0 | 0 | 0 | 0 | 0 | 0 | 0 | 0 | 0 | 0 |
| *ALLERGIES TO FOODS, FOOD ADDITIVES, DRUGS AND OTHER CHEMICALS* | | | | | | | | | | |
| ALLERGY TO CHEMICALS | 0 | 0 | 0 | 0 | 0 | 0 | 0 | 0 | 0 | 0 |
| *ANAPHYLACTIC AND ANAPHYLACTOID RESPONSES* | | | | | | | | | | |
| ANAPHYLACTIC REACTION | 0 | 0 | 0 | 0 | 0 | 0 | 0 | 0 | 0 | 0 |
| *ATOPIC DISORDERS* | | | | | | | | | | |
| SEASONAL ALLERGY | 0 | 0 | 0 | 0 | 0 | 0 | 0 | 0 | 0 | 0 |
| ***IMMUNE DISORDERS NEC*** | | | | | | | | | | |
| *IMMUNE AND ASSOCIATED CONDITIONS NEC* | | | | | | | | | | |
| BACILLE CALMETTE-GUERIN SCAR REACTIVATION | 0 | 0 | 0 | 0 | 0 | 0 | 0 | 0 | 0 | 0 |
| SENSITISATION | 0 | 0 | 0 | 0 | 0 | 0 | 0 | 0 | 0 | 0 |
| **INFECTIONS AND INFESTATIONS** | | | | | | | | | | |
| ***BACTERIAL INFECTIOUS DISORDERS*** | | | | | | | | | | |
| *BACTERIAL INFECTIONS NEC* | | | | | | | | | | |
| CELLULITIS | 0 | 0 | 0 | 0 | 0 | 0 | 0 | 0 | 0 | 0 |
| PERIORBITAL CELLULITIS | 1 | 0 | 1 | 0 | 0 | 1 | 0 | 1 | 0 | 0 |
| *HELICOBACTER INFECTIONS* | | | | | | | | | | |
| HELICOBACTER GASTRITIS | 1 | 0 | 0 | 1 | 0 | 1 | 0 | 0 | 1 | 0 |
| *STAPHYLOCOCCAL INFECTIONS* | | | | | | | | | | |

| | | | | | | | | | | |
|---|---|---|---|---|---|---|---|---|---|---|
| FURUNCLE | 1 | 1 | 0 | 0 | 0 | 1 | 1 | 0 | 0 | 0 |
| ***FUNGAL INFECTIOUS DISORDERS*** | | | | | | | | | | |
| *CANDIDA INFECTIONS* | | | | | | | | | | |
| CANDIDA INFECTION | 1 | 1 | 0 | 0 | 0 | 1 | 1 | 0 | 0 | 0 |
| VULVOVAGINAL CANDIDIASIS | 0 | 0 | 0 | 0 | 0 | 0 | 0 | 0 | 0 | 0 |
| ***INFECTIONS - PATHOGEN UNSPECIFIED*** | | | | | | | | | | |
| *ABDOMINAL AND GASTROINTESTINAL INFECTIONS* | | | | | | | | | | |
| APPENDICITIS | 0 | 0 | 0 | 0 | 0 | 0 | 0 | 0 | 0 | 0 |
| DIARRHOEA INFECTIOUS | 0 | 0 | 0 | 0 | 0 | 0 | 0 | 0 | 0 | 0 |
| GASTROINTESTINAL INFECTION | 0 | 0 | 0 | 0 | 0 | 0 | 0 | 0 | 0 | 0 |
| *BREAST INFECTIONS* | | | | | | | | | | |
| MASTITIS | 1 | 0 | 0 | 1 | 0 | 1 | 0 | 0 | 1 | 0 |
| *CENTRAL NERVOUS SYSTEM AND SPINAL INFECTIONS* | | | | | | | | | | |
| MYELITIS | 0 | 0 | 0 | 0 | 0 | 0 | 0 | 0 | 0 | 0 |
| *DENTAL AND ORAL SOFT TISSUE INFECTIONS* | | | | | | | | | | |
| ABSCESS ORAL | 1 | 1 | 0 | 0 | 0 | 1 | 1 | 0 | 0 | 0 |
| ORAL PUSTULE | 0 | 0 | 0 | 0 | 0 | 0 | 0 | 0 | 0 | 0 |
| PERICORONITIS | 0 | 0 | 0 | 0 | 0 | 0 | 0 | 0 | 0 | 0 |
| TOOTH ABSCESS | 0 | 0 | 0 | 0 | 0 | 0 | 0 | 0 | 0 | 0 |
| *EAR INFECTIONS* | | | | | | | | | | |
| EAR INFECTION | 0 | 0 | 0 | 0 | 0 | 0 | 0 | 0 | 0 | 0 |
| LABYRINTHITIS | 3 | 1 | 2 | 0 | 0 | 3 | 1 | 2 | 0 | 0 |
| OTITIS EXTERNA | 0 | 0 | 0 | 0 | 0 | 0 | 0 | 0 | 0 | 0 |
| *EYE AND EYELID INFECTIONS* | | | | | | | | | | |
| CONJUNCTIVITIS | 0 | 0 | 0 | 0 | 0 | 0 | 0 | 0 | 0 | 0 |
| HORDEOLUM | 0 | 0 | 0 | 0 | 0 | 0 | 0 | 0 | 0 | 0 |
| *INFECTIONS NEC* | | | | | | | | | | |
| ABSCESS | 0 | 0 | 0 | 0 | 0 | 0 | 0 | 0 | 0 | 0 |
| INFECTION | 0 | 0 | 0 | 0 | 0 | 0 | 0 | 0 | 0 | 0 |
| INJECTION SITE INFECTION | 0 | 0 | 0 | 0 | 0 | 0 | 0 | 0 | 0 | 0 |
| LOCALISED INFECTION | 1 | 1 | 0 | 0 | 0 | 1 | 1 | 0 | 0 | 0 |
| WOUND INFECTION | 0 | 0 | 0 | 0 | 0 | 0 | 0 | 0 | 0 | 0 |
| *LOWER RESPIRATORY TRACT AND LUNG INFECTIONS* | | | | | | | | | | |
| LOWER RESPIRATORY TRACT INFECTION | 3 | 1 | 1 | 1 | 0 | 3 | 1 | 1 | 1 | 0 |
| PNEUMONIA | 0 | 0 | 0 | 0 | 0 | 0 | 0 | 0 | 0 | 0 |

| | | | | | | | | | | |
|---|---|---|---|---|---|---|---|---|---|---|
| *MALE REPRODUCTIVE TRACT INFECTIONS* | | | | | | | | | | |
| ORCHITIS | 0 | 0 | 0 | 0 | 0 | 0 | 0 | 0 | 0 | 0 |
| *SEPSIS, BACTERAEMIA, VIRAEMIA AND FUNGAEMIA NEC* | | | | | | | | | | |
| SEPTIC RASH | 0 | 0 | 0 | 0 | 0 | 0 | 0 | 0 | 0 | 0 |
| *SKIN STRUCTURES AND SOFT TISSUE INFECTIONS* | | | | | | | | | | |
| INFECTED DERMAL CYST | 1 | 1 | 0 | 0 | 0 | 1 | 1 | 0 | 0 | 0 |
| INJECTION SITE PUSTULE | 0 | 0 | 0 | 0 | 0 | 0 | 0 | 0 | 0 | 0 |
| SKIN INFECTION | 1 | 1 | 0 | 0 | 0 | 1 | 1 | 0 | 0 | 0 |
| *UPPER RESPIRATORY TRACT INFECTIONS* | | | | | | | | | | |
| LARYNGITIS | 1 | 0 | 1 | 0 | 0 | 1 | 0 | 1 | 0 | 0 |
| NASOPHARYNGITIS | 58 | 30 | 9 | 19 | 0 | 52 | 30 | 6 | 16 | 0 |
| PHARYNGITIS | 0 | 0 | 0 | 0 | 0 | 0 | 0 | 0 | 0 | 0 |
| RHINITIS | 0 | 0 | 0 | 0 | 0 | 0 | 0 | 0 | 0 | 0 |
| SINUSITIS | 3 | 1 | 1 | 1 | 0 | 2 | 1 | 0 | 1 | 0 |
| TONSILLITIS | 0 | 0 | 0 | 0 | 0 | 0 | 0 | 0 | 0 | 0 |
| *URINARY TRACT INFECTIONS* | | | | | | | | | | |
| CYSTITIS | 2 | 1 | 1 | 0 | 0 | 2 | 1 | 1 | 0 | 0 |
| URINARY TRACT INFECTION | 0 | 0 | 0 | 0 | 0 | 0 | 0 | 0 | 0 | 0 |
| ***PROTOZOAL INFECTIOUS DISORDERS*** | | | | | | | | | | |
| *TRYPANOSOMAL INFECTIONS* | | | | | | | | | | |
| AFRICAN TRYPANOSOMIASIS | 1 | 1 | 0 | 0 | 0 | 1 | 1 | 0 | 0 | 0 |
| ***RICKETTSIAL INFECTIOUS DISORDERS*** | | | | | | | | | | |
| *COXIELLA INFECTIONS* | | | | | | | | | | |
| Q FEVER | 1 | 1 | 0 | 0 | 0 | 1 | 1 | 0 | 0 | 0 |
| ***VIRAL INFECTIOUS DISORDERS*** | | | | | | | | | | |
| *CORONAVIRUS INFECTIONS* | | | | | | | | | | |
| COVID-19 | 2 | 1 | 0 | 1 | 0 | 2 | 1 | 0 | 1 | 0 |
| *HERPES VIRAL INFECTIONS* | | | | | | | | | | |
| GENITAL HERPES | 1 | 1 | 0 | 0 | 0 | 1 | 1 | 0 | 0 | 0 |
| HERPES OPHTHALMIC | 0 | 0 | 0 | 0 | 0 | 0 | 0 | 0 | 0 | 0 |
| HERPES SIMPLEX | 1 | 1 | 0 | 0 | 0 | 1 | 1 | 0 | 0 | 0 |
| HERPES ZOSTER | 7 | 4 | 2 | 1 | 0 | 6 | 4 | 2 | 0 | 0 |
| NASAL HERPES | 0 | 0 | 0 | 0 | 0 | 0 | 0 | 0 | 0 | 0 |
| OPHTHALMIC HERPES ZOSTER | 0 | 0 | 0 | 0 | 0 | 0 | 0 | 0 | 0 | 0 |
| ORAL HERPES | 13 | 10 | 2 | 1 | 0 | 13 | 10 | 2 | 1 | 0 |
| *INFLUENZA VIRAL INFECTIONS* | | | | | | | | | | |

| | | | | | | | | | | |
|---|---|---|---|---|---|---|---|---|---|---|
| H2N2 INFLUENZA | 0 | 0 | 0 | 0 | 0 | 0 | 0 | 0 | 0 | 0 |
| INFLUENZA | 129 | 48 | 27 | 51 | 3 | 110 | 48 | 22 | 37 | 3 |
| *RETROVIRAL INFECTIONS* | | | | | | | | | | |
| AIDS RELATED COMPLEX | 0 | 0 | 0 | 0 | 0 | 0 | 0 | 0 | 0 | 0 |
| *VIRAL INFECTIONS NEC* | | | | | | | | | | |
| GASTROENTERITIS VIRAL | 0 | 0 | 0 | 0 | 0 | 0 | 0 | 0 | 0 | 0 |
| SWEATING FEVER | 1 | 1 | 0 | 0 | 0 | 1 | 1 | 0 | 0 | 0 |
| VESTIBULAR NEURONITIS | 0 | 0 | 0 | 0 | 0 | 0 | 0 | 0 | 0 | 0 |
| VIRAL DIARRHOEA | 1 | 1 | 0 | 0 | 0 | 1 | 1 | 0 | 0 | 0 |
| VIRAL RASH | 1 | 0 | 0 | 0 | 1 | 1 | 0 | 0 | 0 | 1 |
| VIRAL UPPER RESPIRATORY TRACT INFECTION | 0 | 0 | 0 | 0 | 0 | 0 | 0 | 0 | 0 | 0 |
| **INJURY, POISONING AND PROCEDURAL COMPLICATIONS** | | | | | | | | | | |
| ***BONE AND JOINT INJURIES*** | | | | | | | | | | |
| *FRACTURES AND DISLOCATIONS NEC* | | | | | | | | | | |
| JOINT DISLOCATION | 0 | 0 | 0 | 0 | 0 | 0 | 0 | 0 | 0 | 0 |
| ***EXPOSURES, CHEMICAL INJURIES AND POISONING*** | | | | | | | | | | |
| *POISONING AND TOXICITY* | | | | | | | | | | |
| SYSTEMIC TOXICITY | 0 | 0 | 0 | 0 | 0 | 0 | 0 | 0 | 0 | 0 |
| TOXICITY TO VARIOUS AGENTS | 1 | 1 | 0 | 0 | 0 | 1 | 1 | 0 | 0 | 0 |
| ***INJURIES BY PHYSICAL AGENTS*** | | | | | | | | | | |
| *CONDITIONS CAUSED BY COLD* | | | | | | | | | | |
| CHILLBLAINS | 1 | 1 | 0 | 0 | 0 | 1 | 1 | 0 | 0 | 0 |
| *HEAT INJURIES (EXCL THERMAL BURNS)* | | | | | | | | | | |
| HEAT EXHAUSTION | 0 | 0 | 0 | 0 | 0 | 0 | 0 | 0 | 0 | 0 |
| HEAT OEDEMA | 0 | 0 | 0 | 0 | 0 | 0 | 0 | 0 | 0 | 0 |
| *RADIATION INJURIES* | | | | | | | | | | |
| SUNBURN | 0 | 0 | 0 | 0 | 0 | 0 | 0 | 0 | 0 | 0 |
| *THERMAL BURNS* | | | | | | | | | | |
| THERMAL BURN | 0 | 0 | 0 | 0 | 0 | 0 | 0 | 0 | 0 | 0 |
| THERMAL BURNS OF EYE | 0 | 0 | 0 | 0 | 0 | 0 | 0 | 0 | 0 | 0 |
| ***INJURIES NEC*** | | | | | | | | | | |
| *CHEST AND RESPIRATORY TRACT INJURIES NEC* | | | | | | | | | | |
| CHEST CRUSHING | 0 | 0 | 0 | 0 | 0 | 0 | 0 | 0 | 0 | 0 |
| *EYE INJURIES NEC* | | | | | | | | | | |
| EYE CONTUSION | 1 | 1 | 0 | 0 | 0 | 1 | 1 | 0 | 0 | 0 |
| *MUSCLE, TENDON AND LIGAMENT INJURIES* | | | | | | | | | | |

| | | | | | | | | | | |
|---|---|---|---|---|---|---|---|---|---|---|
| LIGAMENT SPRAIN | 0 | 0 | 0 | 0 | 0 | 0 | 0 | 0 | 0 | 0 |
| MUSCLE INJURY | 1 | 0 | 0 | 1 | 0 | 0 | 0 | 0 | 0 | 0 |
| MUSCLE STRAIN | 0 | 0 | 0 | 0 | 0 | 0 | 0 | 0 | 0 | 0 |
| *NERVE INJURIES NEC* | | | | | | | | | | |
| NERVE INJURY | 0 | 0 | 0 | 0 | 0 | 0 | 0 | 0 | 0 | 0 |
| *NON-SITE SPECIFIC INJURIES NEC* | | | | | | | | | | |
| ARTHROPOD STING | 2 | 2 | 0 | 0 | 0 | 2 | 2 | 0 | 0 | 0 |
| BITE | 0 | 0 | 0 | 0 | 0 | 0 | 0 | 0 | 0 | 0 |
| FALL | 1 | 1 | 0 | 0 | 0 | 1 | 1 | 0 | 0 | 0 |
| INFLAMMATION OF WOUND | 1 | 1 | 0 | 0 | 0 | 1 | 1 | 0 | 0 | 0 |
| TISSUE INJURY | 0 | 0 | 0 | 0 | 0 | 0 | 0 | 0 | 0 | 0 |
| WOUND COMPLICATION | 0 | 0 | 0 | 0 | 0 | 0 | 0 | 0 | 0 | 0 |
| WOUND SECRETION | 0 | 0 | 0 | 0 | 0 | 0 | 0 | 0 | 0 | 0 |
| *SITE SPECIFIC INJURIES NEC* | | | | | | | | | | |
| LIMB INJURY | 0 | 0 | 0 | 0 | 0 | 0 | 0 | 0 | 0 | 0 |
| *SKIN INJURIES NEC* | | | | | | | | | | |
| CONTUSION | 56 | 29 | 10 | 14 | 3 | 52 | 29 | 8 | 12 | 3 |
| SCAR | 1 | 1 | 0 | 0 | 0 | 1 | 1 | 0 | 0 | 0 |
| SKIN WOUND | 0 | 0 | 0 | 0 | 0 | 0 | 0 | 0 | 0 | 0 |
| ***PROCEDURAL RELATED INJURIES AND COMPLICATIONS NEC*** | | | | | | | | | | |
| *CARDIAC AND VASCULAR PROCEDURAL COMPLICATIONS* | | | | | | | | | | |
| CARDIAC PROCEDURE COMPLICATION | 0 | 0 | 0 | 0 | 0 | 0 | 0 | 0 | 0 | 0 |
| *GASTROINTESTINAL AND HEPATOBILIARY PROCEDURAL COMPLICATIONS* | | | | | | | | | | |
| PROCEDURAL NAUSEA | 0 | 0 | 0 | 0 | 0 | 0 | 0 | 0 | 0 | 0 |
| *NEUROLOGICAL AND PSYCHIATRIC PROCEDURAL COMPLICATIONS* | | | | | | | | | | |
| PROCEDURAL DIZZINESS | 0 | 0 | 0 | 0 | 0 | 0 | 0 | 0 | 0 | 0 |
| *NON-SITE SPECIFIC PROCEDURAL COMPLICATIONS* | | | | | | | | | | |
| INCISION SITE PAIN | 1 | 1 | 0 | 0 | 0 | 1 | 1 | 0 | 0 | 0 |
| INCISION SITE SWELLING | 1 | 0 | 1 | 0 | 0 | 1 | 0 | 1 | 0 | 0 |
| INJECTION RELATED REACTION | 1 | 1 | 0 | 0 | 0 | 1 | 1 | 0 | 0 | 0 |
| POST PROCEDURAL COMPLICATION | 0 | 0 | 0 | 0 | 0 | 0 | 0 | 0 | 0 | 0 |
| PROCEDURAL PAIN | 0 | 0 | 0 | 0 | 0 | 0 | 0 | 0 | 0 | 0 |
| *VACCINATION RELATED COMPLICATIONS* | | | | | | | | | | |
| IMMUNISATION REACTION | 3 | 1 | 1 | 1 | 0 | 3 | 1 | 1 | 1 | 0 |
| **INVESTIGATIONS** | | | | | | | | | | |

| | | | | | | | | | | |
|---|---|---|---|---|---|---|---|---|---|---|
| ***CARDIAC AND VASCULAR INVESTIGATIONS (EXCL ENZYME TESTS)*** | | | | | | | | | | |
| *HEART RATE AND PULSE INVESTIGATIONS* | | | | | | | | | | |
| HEART RATE | 7 | 5 | 1 | 1 | 0 | 7 | 5 | 1 | 1 | 0 |
| HEART RATE DECREASED | 1 | 1 | 0 | 0 | 0 | 1 | 1 | 0 | 0 | 0 |
| HEART RATE INCREASED | 7 | 6 | 0 | 1 | 0 | 7 | 6 | 0 | 1 | 0 |
| HEART RATE IRREGULAR | 2 | 0 | 1 | 1 | 0 | 2 | 0 | 1 | 1 | 0 |
| *VASCULAR TESTS NEC (INCL BLOOD PRESSURE)* | | | | | | | | | | |
| BLOOD PRESSURE DECREASED | 0 | 0 | 0 | 0 | 0 | 0 | 0 | 0 | 0 | 0 |
| BLOOD PRESSURE INCREASED | 3 | 2 | 0 | 1 | 0 | 3 | 2 | 0 | 1 | 0 |
| BLOOD PRESSURE MEASUREMENT | 0 | 0 | 0 | 0 | 0 | 0 | 0 | 0 | 0 | 0 |
| ***ENDOCRINE INVESTIGATIONS (INCL SEX HORMONES)*** | | | | | | | | | | |
| *ENDOCRINE ANALYSES AND IMAGING NEC* | | | | | | | | | | |
| HORMONE LEVEL ABNORMAL | 1 | 1 | 0 | 0 | 0 | 1 | 1 | 0 | 0 | 0 |
| *PITUITARY ANALYSES ANTERIOR* | | | | | | | | | | |
| BLOOD FOLLICLE STIMULATING HORMONE INCREASED | 0 | 0 | 0 | 0 | 0 | 0 | 0 | 0 | 0 | 0 |
| BLOOD LUTEINISING HORMONE | 1 | 1 | 0 | 0 | 0 | 1 | 1 | 0 | 0 | 0 |
| *THYROID ANALYSES* | | | | | | | | | | |
| TRI-IODOTHYRONINE | 0 | 0 | 0 | 0 | 0 | 0 | 0 | 0 | 0 | 0 |
| ***HAEMATOLOGY INVESTIGATIONS (INCL BLOOD GROUPS)*** | | | | | | | | | | |
| *COAGULATION AND BLEEDING ANALYSES* | | | | | | | | | | |
| BLEEDING TIME | 0 | 0 | 0 | 0 | 0 | 0 | 0 | 0 | 0 | 0 |
| INTERNATIONAL NORMALISED RATIO DECREASED | 0 | 0 | 0 | 0 | 0 | 0 | 0 | 0 | 0 | 0 |
| *PLATELET ANALYSES* | | | | | | | | | | |
| PLATELET COUNT INCREASED | 0 | 0 | 0 | 0 | 0 | 0 | 0 | 0 | 0 | 0 |
| *RED BLOOD CELL ANALYSES* | | | | | | | | | | |
| HAEMOGLOBIN | 0 | 0 | 0 | 0 | 0 | 0 | 0 | 0 | 0 | 0 |
| ***METABOLIC, NUTRITIONAL AND BLOOD GAS INVESTIGATIONS*** | | | | | | | | | | |
| *BLOOD GAS AND ACID BASE ANALYSES* | | | | | | | | | | |
| OXYGEN SATURATION DECREASED | 0 | 0 | 0 | 0 | 0 | 0 | 0 | 0 | 0 | 0 |
| *CARBOHYDRATE TOLERANCE ANALYSES (INCL DIABETES)* | | | | | | | | | | |
| BLOOD GLUCOSE | 1 | 1 | 0 | 0 | 0 | 1 | 1 | 0 | 0 | 0 |
| BLOOD GLUCOSE ABNORMAL | 0 | 0 | 0 | 0 | 0 | 0 | 0 | 0 | 0 | 0 |
| BLOOD GLUCOSE DECREASED | 0 | 0 | 0 | 0 | 0 | 0 | 0 | 0 | 0 | 0 |
| BLOOD GLUCOSE INCREASED | 2 | 2 | 0 | 0 | 0 | 2 | 2 | 0 | 0 | 0 |
| ***MICROBIOLOGY AND SEROLOGY INVESTIGATIONS*** | | | | | | | | | | |

| | | | | | | | | | | |
|---|---|---|---|---|---|---|---|---|---|---|
| *VIRUS IDENTIFICATION AND SEROLOGY* | | | | | | | | | | |
| SARS-COV-2 TEST | 1 | 1 | 0 | 0 | 0 | 1 | 1 | 0 | 0 | 0 |
| SARS-COV-2 TEST POSITIVE | 1 | 0 | 0 | 1 | 0 | 1 | 0 | 0 | 1 | 0 |
| ***MUSCULOSKELETAL AND SOFT TISSUE INVESTIGATIONS (EXCL ENZYME TESTS)*** | | | | | | | | | | |
| *MUSCULOSKELETAL AND SOFT TISSUE IMAGING PROCEDURES* | | | | | | | | | | |
| BONE SCAN | 0 | 0 | 0 | 0 | 0 | 0 | 0 | 0 | 0 | 0 |
| ***NEUROLOGICAL, SPECIAL SENSES AND PSYCHIATRIC INVESTIGATIONS*** | | | | | | | | | | |
| *CENTRAL NERVOUS SYSTEM IMAGING PROCEDURES* | | | | | | | | | | |
| MAGNETIC RESONANCE IMAGING HEAD | 2 | 2 | 0 | 0 | 0 | 2 | 2 | 0 | 0 | 0 |
| SCAN BRAIN | 1 | 0 | 0 | 1 | 0 | 1 | 0 | 0 | 1 | 0 |
| ***PHYSICAL EXAMINATION AND ORGAN SYSTEM STATUS TOPICS*** | | | | | | | | | | |
| *PHYSICAL EXAMINATION PROCEDURES AND ORGAN SYSTEM STATUS* | | | | | | | | | | |
| BODY TEMPERATURE | 4 | 0 | 3 | 1 | 0 | 4 | 0 | 3 | 1 | 0 |
| BODY TEMPERATURE ABNORMAL | 1 | 1 | 0 | 0 | 0 | 1 | 1 | 0 | 0 | 0 |
| BODY TEMPERATURE DECREASED | 0 | 0 | 0 | 0 | 0 | 0 | 0 | 0 | 0 | 0 |
| BODY TEMPERATURE FLUCTUATION | 0 | 0 | 0 | 0 | 0 | 0 | 0 | 0 | 0 | 0 |
| BODY TEMPERATURE INCREASED | 10 | 4 | 2 | 4 | 0 | 8 | 4 | 1 | 3 | 0 |
| GRIP STRENGTH DECREASED | 0 | 0 | 0 | 0 | 0 | 0 | 0 | 0 | 0 | 0 |
| HEAD LAG | 0 | 0 | 0 | 0 | 0 | 0 | 0 | 0 | 0 | 0 |
| LEFT-HANDEDNESS | 1 | 1 | 0 | 0 | 0 | 1 | 1 | 0 | 0 | 0 |
| LYMPH NODE PALPABLE | 0 | 0 | 0 | 0 | 0 | 0 | 0 | 0 | 0 | 0 |
| RESPIRATORY RATE DECREASED | 0 | 0 | 0 | 0 | 0 | 0 | 0 | 0 | 0 | 0 |
| SKIN TEMPERATURE | 1 | 0 | 0 | 1 | 0 | 1 | 0 | 0 | 1 | 0 |
| WEIGHT DECREASED | 2 | 2 | 0 | 0 | 0 | 2 | 2 | 0 | 0 | 0 |
| WEIGHT INCREASED | 0 | 0 | 0 | 0 | 0 | 0 | 0 | 0 | 0 | 0 |
| ***RENAL AND URINARY TRACT INVESTIGATIONS AND URINALYSES*** | | | | | | | | | | |
| *URINALYSIS NEC* | | | | | | | | | | |
| BLOOD URINE | 0 | 0 | 0 | 0 | 0 | 0 | 0 | 0 | 0 | 0 |
| NITRITE URINE PRESENT | 0 | 0 | 0 | 0 | 0 | 0 | 0 | 0 | 0 | 0 |
| PH URINE | 1 | 1 | 0 | 0 | 0 | 1 | 1 | 0 | 0 | 0 |
| *URINARY TRACT FUNCTION ANALYSES NEC* | | | | | | | | | | |
| URINE OUTPUT | 1 | 0 | 0 | 1 | 0 | 1 | 0 | 0 | 1 | 0 |
| URINE OUTPUT INCREASED | 0 | 0 | 0 | 0 | 0 | 0 | 0 | 0 | 0 | 0 |
| ***REPRODUCTIVE ORGAN AND BREAST INVESTIGATIONS (EXCL HORMONE ANALYSES)*** | | | | | | | | | | |
| *REPRODUCTIVE ORGAN AND BREAST IMAGING PROCEDURES* | | | | | | | | | | |

| | | | | | | | | | | |
|---|---|---|---|---|---|---|---|---|---|---|
| BREAST SCAN | 1 | 0 | 1 | 0 | 0 | 1 | 0 | 1 | 0 | 0 |
| ***RESPIRATORY AND PULMONARY INVESTIGATIONS (EXCL BLOOD GASES)*** | | | | | | | | | | |
| *RESPIRATORY AND PULMONARY FUNCTION DIAGNOSTIC PROCEDURES* | | | | | | | | | | |
| FORCED EXPIRATORY VOLUME DECREASED | 0 | 0 | 0 | 0 | 0 | 0 | 0 | 0 | 0 | 0 |
| FORCED EXPIRATORY VOLUME INCREASED | 1 | 0 | 1 | 0 | 0 | 1 | 0 | 1 | 0 | 0 |
| ***WATER, ELECTROLYTE AND MINERAL INVESTIGATIONS*** | | | | | | | | | | |
| *WATER AND ELECTROLYTE ANALYSES NEC* | | | | | | | | | | |
| VOLUME BLOOD | 1 | 0 | 1 | 0 | 0 | 1 | 0 | 1 | 0 | 0 |
| **METABOLISM AND NUTRITION DISORDERS** | | | | | | | | | | |
| ***APPETITE AND GENERAL NUTRITIONAL DISORDERS*** | | | | | | | | | | |
| *APPETITE DISORDERS* | | | | | | | | | | |
| APPETITE DISORDER | 0 | 0 | 0 | 0 | 0 | 0 | 0 | 0 | 0 | 0 |
| DECREASED APPETITE | 23 | 14 | 2 | 5 | 2 | 22 | 14 | 2 | 5 | 1 |
| FOOD CRAVING | 1 | 1 | 0 | 0 | 0 | 1 | 1 | 0 | 0 | 0 |
| FOOD REFUSAL | 0 | 0 | 0 | 0 | 0 | 0 | 0 | 0 | 0 | 0 |
| INCREASED APPETITE | 0 | 0 | 0 | 0 | 0 | 0 | 0 | 0 | 0 | 0 |
| *GENERAL NUTRITIONAL DISORDERS NEC* | | | | | | | | | | |
| FOOD AVERSION | 0 | 0 | 0 | 0 | 0 | 0 | 0 | 0 | 0 | 0 |
| ***ELECTROLYTE AND FLUID BALANCE CONDITIONS*** | | | | | | | | | | |
| *TOTAL FLUID VOLUME DECREASED* | | | | | | | | | | |
| DEHYDRATION | 0 | 0 | 0 | 0 | 0 | 0 | 0 | 0 | 0 | 0 |
| *TOTAL FLUID VOLUME INCREASED* | | | | | | | | | | |
| FLUID RETENTION | 0 | 0 | 0 | 0 | 0 | 0 | 0 | 0 | 0 | 0 |
| ***FOOD INTOLERANCE SYNDROMES*** | | | | | | | | | | |
| *FOOD MALABSORPTION AND INTOLERANCE SYNDROMES (EXCL SUGAR INTOLERANCE)* | | | | | | | | | | |
| ALCOHOL INTOLERANCE | 0 | 0 | 0 | 0 | 0 | 0 | 0 | 0 | 0 | 0 |
| ***GLUCOSE METABOLISM DISORDERS (INCL DIABETES MELLITUS)*** | | | | | | | | | | |
| *DIABETES MELLITUS (INCL SUBTYPES)* | | | | | | | | | | |
| DIABETES MELLITUS | 1 | 1 | 0 | 0 | 0 | 1 | 1 | 0 | 0 | 0 |
| DIABETES MELLITUS INADEQUATE CONTROL | 1 | 1 | 0 | 0 | 0 | 1 | 1 | 0 | 0 | 0 |
| *HYPERGLYCAEMIC CONDITIONS NEC* | | | | | | | | | | |
| HYPERGLYCAEMIA | 1 | 1 | 0 | 0 | 0 | 1 | 1 | 0 | 0 | 0 |
| *HYPOGLYCAEMIC CONDITIONS NEC* | | | | | | | | | | |
| HYPOGLYCAEMIA | 1 | 1 | 0 | 0 | 0 | 1 | 1 | 0 | 0 | 0 |
| ***PURINE AND PYRIMIDINE METABOLISM DISORDERS*** | | | | | | | | | | |
| *DISORDERS OF PURINE METABOLISM* | | | | | | | | | | |

| | | | | | | | | | | |
|---|---|---|---|---|---|---|---|---|---|---|
| GOUT | 0 | 0 | 0 | 0 | 0 | 0 | 0 | 0 | 0 | 0 |
| **MUSCULOSKELETAL AND CONNECTIVE TISSUE DISORDERS** | | | | | | | | | | |
| ***BONE DISORDERS (EXCL CONGENITAL AND FRACTURES)*** | | | | | | | | | | |
| *BONE DISORDERS NEC* | | | | | | | | | | |
| JAW DISORDER | 0 | 0 | 0 | 0 | 0 | 0 | 0 | 0 | 0 | 0 |
| OSTEITIS | 0 | 0 | 0 | 0 | 0 | 0 | 0 | 0 | 0 | 0 |
| *BONE RELATED SIGNS AND SYMPTOMS* | | | | | | | | | | |
| BONE PAIN | 2 | 1 | 0 | 1 | 0 | 2 | 1 | 0 | 1 | 0 |
| BONE SWELLING | 0 | 0 | 0 | 0 | 0 | 0 | 0 | 0 | 0 | 0 |
| PAIN IN JAW | 4 | 3 | 0 | 1 | 0 | 4 | 3 | 0 | 1 | 0 |
| PUBIC PAIN | 0 | 0 | 0 | 0 | 0 | 0 | 0 | 0 | 0 | 0 |
| SPINAL PAIN | 1 | 0 | 1 | 0 | 0 | 1 | 0 | 1 | 0 | 0 |
| ***CONNECTIVE TISSUE DISORDERS (EXCL CONGENITAL)*** | | | | | | | | | | |
| *CONNECTIVE TISSUE DISORDERS NEC* | | | | | | | | | | |
| POLYMYALGIA RHEUMATICA | 1 | 0 | 0 | 1 | 0 | 1 | 0 | 0 | 1 | 0 |
| *LUPUS ERYTHEMATOSUS (INCL SUBTYPES)* | | | | | | | | | | |
| SYSTEMIC LUPUS ERYTHEMATOSUS | 0 | 0 | 0 | 0 | 0 | 0 | 0 | 0 | 0 | 0 |
| ***JOINT DISORDERS*** | | | | | | | | | | |
| *ARTHROPATHIES NEC* | | | | | | | | | | |
| ARTHRITIS | 5 | 1 | 2 | 2 | 0 | 5 | 1 | 2 | 2 | 0 |
| ARTHROPATHY | 1 | 0 | 1 | 0 | 0 | 1 | 0 | 1 | 0 | 0 |
| POLYARTHRITIS | 1 | 0 | 1 | 0 | 0 | 1 | 0 | 1 | 0 | 0 |
| RHEUMATIC FEVER | 0 | 0 | 0 | 0 | 0 | 0 | 0 | 0 | 0 | 0 |
| *JOINT RELATED DISORDERS NEC* | | | | | | | | | | |
| JOINT LOCK | 0 | 0 | 0 | 0 | 0 | 0 | 0 | 0 | 0 | 0 |
| PERIARTHRITIS | 2 | 1 | 0 | 1 | 0 | 2 | 1 | 0 | 1 | 0 |
| TEMPOROMANDIBULAR PAIN AND DYSFUNCTION SYNDROME | 1 | 1 | 0 | 0 | 0 | 1 | 1 | 0 | 0 | 0 |
| *JOINT RELATED SIGNS AND SYMPTOMS* | | | | | | | | | | |
| ARTHRALGIA | 332 | 174 | 64 | 87 | 7 | 298 | 174 | 47 | 72 | 5 |
| JAW CLICKING | 0 | 0 | 0 | 0 | 0 | 0 | 0 | 0 | 0 | 0 |
| JOINT STIFFNESS | 9 | 7 | 0 | 2 | 0 | 8 | 7 | 0 | 1 | 0 |
| JOINT SWELLING | 7 | 3 | 1 | 3 | 0 | 5 | 3 | 1 | 1 | 0 |
| *OSTEOARTHROPATHIES* | | | | | | | | | | |
| OSTEOARTHRITIS | 1 | 1 | 0 | 0 | 0 | 1 | 1 | 0 | 0 | 0 |
| *PSORIATIC ARTHROPATHIES* | | | | | | | | | | |

| | | | | | | | | | | |
|---|---|---|---|---|---|---|---|---|---|---|
| PSORIATIC ARTHROPATHY | 0 | 0 | 0 | 0 | 0 | 0 | 0 | 0 | 0 | 0 |
| *RHEUMATOID ARTHROPATHIES* | | | | | | | | | | |
| RHEUMATOID ARTHRITIS | 1 | 1 | 0 | 0 | 0 | 1 | 1 | 0 | 0 | 0 |
| ***MUSCLE DISORDERS*** | | | | | | | | | | |
| *MUSCLE INFECTIONS AND INFLAMMATIONS* | | | | | | | | | | |
| MYOSITIS | 1 | 0 | 1 | 0 | 0 | 1 | 0 | 1 | 0 | 0 |
| *MUSCLE PAINS* | | | | | | | | | | |
| FIBROMYALGIA | 2 | 0 | 0 | 2 | 0 | 2 | 0 | 0 | 2 | 0 |
| MYALGIA | 457 | 271 | 82 | 96 | 8 | 417 | 271 | 67 | 72 | 7 |
| *MUSCLE RELATED SIGNS AND SYMPTOMS NEC* | | | | | | | | | | |
| MUSCLE ATROPHY | 0 | 0 | 0 | 0 | 0 | 0 | 0 | 0 | 0 | 0 |
| MUSCLE DISCOMFORT | 1 | 1 | 0 | 0 | 0 | 1 | 1 | 0 | 0 | 0 |
| MUSCLE FATIGUE | 9 | 5 | 2 | 2 | 0 | 9 | 5 | 2 | 2 | 0 |
| MUSCLE FIBROSIS | 0 | 0 | 0 | 0 | 0 | 0 | 0 | 0 | 0 | 0 |
| MUSCLE MASS | 0 | 0 | 0 | 0 | 0 | 0 | 0 | 0 | 0 | 0 |
| MUSCLE SPASMS | 12 | 6 | 4 | 2 | 0 | 11 | 6 | 3 | 2 | 0 |
| MUSCLE TIGHTNESS | 0 | 0 | 0 | 0 | 0 | 0 | 0 | 0 | 0 | 0 |
| MUSCLE TWITCHING | 0 | 0 | 0 | 0 | 0 | 0 | 0 | 0 | 0 | 0 |
| *MUSCLE TONE ABNORMALITIES* | | | | | | | | | | |
| MUSCLE RIGIDITY | 1 | 1 | 0 | 0 | 0 | 1 | 1 | 0 | 0 | 0 |
| TRISMUS | 0 | 0 | 0 | 0 | 0 | 0 | 0 | 0 | 0 | 0 |
| *MUSCLE WEAKNESS CONDITIONS* | | | | | | | | | | |
| MUSCULAR WEAKNESS | 13 | 6 | 5 | 2 | 0 | 11 | 6 | 4 | 1 | 0 |
| ***MUSCULOSKELETAL AND CONNECTIVE TISSUE DISORDERS NEC*** | | | | | | | | | | |
| *MUSCULOSKELETAL AND CONNECTIVE TISSUE CONDITIONS NEC* | | | | | | | | | | |
| MOBILITY DECREASED | 1 | 0 | 1 | 0 | 0 | 0 | 0 | 0 | 0 | 0 |
| MUSCULOSKELETAL STIFFNESS | 78 | 44 | 9 | 25 | 0 | 71 | 44 | 9 | 18 | 0 |
| BACK PAIN | 34 | 16 | 8 | 10 | 0 | 30 | 16 | 7 | 7 | 0 |
| FLANK PAIN | 0 | 0 | 0 | 0 | 0 | 0 | 0 | 0 | 0 | 0 |
| LIMB DISCOMFORT | 242 | 150 | 47 | 45 | 0 | 228 | 150 | 40 | 38 | 0 |
| MUSCULOSKELETAL CHEST PAIN | 0 | 0 | 0 | 0 | 0 | 0 | 0 | 0 | 0 | 0 |
| MUSCULOSKELETAL DISCOMFORT | 2 | 0 | 1 | 1 | 0 | 1 | 0 | 0 | 1 | 0 |
| MUSCULOSKELETAL PAIN | 1 | 0 | 0 | 1 | 0 | 1 | 0 | 0 | 1 | 0 |
| NECK PAIN | 22 | 9 | 5 | 8 | 0 | 21 | 9 | 5 | 7 | 0 |
| PAIN IN EXTREMITY | 1622 | 1059 | 262 | 286 | 15 | 1515 | 1059 | 207 | 235 | 14 |

| | | | | | | | | | | |
|---|---|---|---|---|---|---|---|---|---|---|
| *SOFT TISSUE DISORDERS NEC* | | | | | | | | | | |
| AXILLARY MASS | 1 | 0 | 0 | 1 | 0 | 0 | 0 | 0 | 0 | 0 |
| GROIN PAIN | 1 | 0 | 1 | 0 | 0 | 1 | 0 | 1 | 0 | 0 |
| ***SYNOVIAL AND BURSAL DISORDERS*** | | | | | | | | | | |
| *SYNOVIAL DISORDERS* | | | | | | | | | | |
| SYNOVITIS | 1 | 0 | 1 | 0 | 0 | 1 | 0 | 1 | 0 | 0 |
| ***TENDON, LIGAMENT AND CARTILAGE DISORDERS*** | | | | | | | | | | |
| *CARTILAGE DISORDERS* | | | | | | | | | | |
| COSTOCHONDRITIS | 0 | 0 | 0 | 0 | 0 | 0 | 0 | 0 | 0 | 0 |
| *TENDON DISORDERS* | | | | | | | | | | |
| TENDONITIS | 0 | 0 | 0 | 0 | 0 | 0 | 0 | 0 | 0 | 0 |
| TENOSYNOVITIS | 1 | 0 | 1 | 0 | 0 | 1 | 0 | 1 | 0 | 0 |
| TRIGGER FINGER | 0 | 0 | 0 | 0 | 0 | 0 | 0 | 0 | 0 | 0 |
| **NEOPLASMS BENIGN, MALIGNANT AND UNSPECIFIED (INCL CYSTS AND POLYPS)** | | | | | | | | | | |
| ***CUTANEOUS NEOPLASMS BENIGN*** | | | | | | | | | | |
| *SKIN NEOPLASMS BENIGN* | | | | | | | | | | |
| MELANOCYTIC NAEVUS | 0 | 0 | 0 | 0 | 0 | 0 | 0 | 0 | 0 | 0 |
| SEBORRHOEIC KERATOSIS | 0 | 0 | 0 | 0 | 0 | 0 | 0 | 0 | 0 | 0 |
| SKIN PAPILLOMA | 1 | 1 | 0 | 0 | 0 | 1 | 1 | 0 | 0 | 0 |
| **NERVOUS SYSTEM DISORDERS** | | | | | | | | | | |
| ***CENTRAL NERVOUS SYSTEM INFECTIONS AND INFLAMMATIONS*** | | | | | | | | | | |
| *MYELITIS (INCL INFECTIVE)* | | | | | | | | | | |
| MYELITIS TRANSVERSE | 0 | 0 | 0 | 0 | 0 | 0 | 0 | 0 | 0 | 0 |
| ***CENTRAL NERVOUS SYSTEM VASCULAR DISORDERS*** | | | | | | | | | | |
| *CENTRAL NERVOUS SYSTEM HAEMORRHAGES AND CEREBROVASCULAR ACCIDENTS* | | | | | | | | | | |
| CEREBRAL HAEMORRHAGE | 0 | 0 | 0 | 0 | 0 | 0 | 0 | 0 | 0 | 0 |
| CEREBROVASCULAR ACCIDENT | 1 | 0 | 0 | 1 | 0 | 1 | 0 | 0 | 1 | 0 |
| *TRANSIENT CEREBROVASCULAR EVENTS* | | | | | | | | | | |
| TRANSIENT ISCHAEMIC ATTACK | 0 | 0 | 0 | 0 | 0 | 0 | 0 | 0 | 0 | 0 |
| ***CRANIAL NERVE DISORDERS (EXCL NEOPLASMS)*** | | | | | | | | | | |
| *FACIAL CRANIAL NERVE DISORDERS* | | | | | | | | | | |
| BELL'S PALSY | 1 | 0 | 1 | 0 | 0 | 1 | 0 | 1 | 0 | 0 |
| FACIAL PARALYSIS | 3 | 3 | 0 | 0 | 0 | 3 | 3 | 0 | 0 | 0 |
| FACIAL PARESIS | 0 | 0 | 0 | 0 | 0 | 0 | 0 | 0 | 0 | 0 |
| *OLFACTORY NERVE DISORDERS* | | | | | | | | | | |
| ANOSMIA | 5 | 4 | 1 | 0 | 0 | 5 | 4 | 1 | 0 | 0 |

| | | | | | | | | | | |
|---|---|---|---|---|---|---|---|---|---|---|
| HYPOSMIA | 0 | 0 | 0 | 0 | 0 | 0 | 0 | 0 | 0 | 0 |
| PAROSMIA | 6 | 4 | 1 | 1 | 0 | 6 | 4 | 1 | 1 | 0 |
| *TRIGEMINAL DISORDERS* | | | | | | | | | | |
| TRIGEMINAL NEURALGIA | 1 | 1 | 0 | 0 | 0 | 1 | 1 | 0 | 0 | 0 |
| TRIGEMINAL NEURITIS | 1 | 0 | 0 | 1 | 0 | 1 | 0 | 0 | 1 | 0 |
| ***DEMYELINATING DISORDERS*** | | | | | | | | | | |
| *MULTIPLE SCLEROSIS ACUTE AND PROGRESSIVE* | | | | | | | | | | |
| MULTIPLE SCLEROSIS RELAPSE | 0 | 0 | 0 | 0 | 0 | 0 | 0 | 0 | 0 | 0 |
| ***HEADACHES*** | | | | | | | | | | |
| *HEADACHES NEC* | | | | | | | | | | |
| CLUSTER HEADACHE | 5 | 0 | 3 | 2 | 0 | 3 | 0 | 1 | 2 | 0 |
| COLD-STIMULUS HEADACHE | 0 | 0 | 0 | 0 | 0 | 0 | 0 | 0 | 0 | 0 |
| DRUG WITHDRAWAL HEADACHE | 0 | 0 | 0 | 0 | 0 | 0 | 0 | 0 | 0 | 0 |
| HEADACHE | 1119 | 618 | 207 | 274 | 20 | 1024 | 618 | 166 | 223 | 17 |
| PRIMARY STABBING HEADACHE | 1 | 1 | 0 | 0 | 0 | 1 | 1 | 0 | 0 | 0 |
| SINUS HEADACHE | 18 | 11 | 4 | 3 | 0 | 16 | 11 | 3 | 2 | 0 |
| TENSION HEADACHE | 27 | 16 | 4 | 7 | 0 | 24 | 16 | 3 | 5 | 0 |
| THUNDERCLAP HEADACHE | 0 | 0 | 0 | 0 | 0 | 0 | 0 | 0 | 0 | 0 |
| VASCULAR HEADACHE | 1 | 1 | 0 | 0 | 0 | 1 | 1 | 0 | 0 | 0 |
| *MIGRAINE HEADACHES* | | | | | | | | | | |
| MIGRAINE | 45 | 26 | 9 | 10 | 0 | 41 | 26 | 8 | 7 | 0 |
| MIGRAINE WITH AURA | 4 | 4 | 0 | 0 | 0 | 4 | 4 | 0 | 0 | 0 |
| RETINAL MIGRAINE | 1 | 1 | 0 | 0 | 0 | 1 | 1 | 0 | 0 | 0 |
| TYPICAL AURA WITHOUT HEADACHE | 1 | 0 | 1 | 0 | 0 | 1 | 0 | 1 | 0 | 0 |
| ***MENTAL IMPAIRMENT DISORDERS*** | | | | | | | | | | |
| *INTELLECTUAL DISABILITIES* | | | | | | | | | | |
| INTELLECTUAL DISABILITY | 1 | 0 | 0 | 1 | 0 | 1 | 0 | 0 | 1 | 0 |
| *MEMORY LOSS (EXCL DEMENTIA)* | | | | | | | | | | |
| AMNESIA | 0 | 0 | 0 | 0 | 0 | 0 | 0 | 0 | 0 | 0 |
| MEMORY IMPAIRMENT | 3 | 2 | 1 | 0 | 0 | 2 | 2 | 0 | 0 | 0 |
| *MENTAL IMPAIRMENT (EXCL DEMENTIA AND MEMORY LOSS)* | | | | | | | | | | |
| COGNITIVE DISORDER | 2 | 1 | 0 | 1 | 0 | 2 | 1 | 0 | 1 | 0 |
| DISTURBANCE IN ATTENTION | 3 | 1 | 2 | 0 | 0 | 3 | 1 | 2 | 0 | 0 |
| MENTAL IMPAIRMENT | 3 | 2 | 0 | 1 | 0 | 2 | 2 | 0 | 0 | 0 |
| ***MOVEMENT DISORDERS (INCL PARKINSONISM)*** | | | | | | | | | | |

| | | | | | | | | | | |
|---|---|---|---|---|---|---|---|---|---|---|
| *DYSKINESIAS AND MOVEMENT DISORDERS NEC* | | | | | | | | | | |
| BRADYKINESIA | 0 | 0 | 0 | 0 | 0 | 0 | 0 | 0 | 0 | 0 |
| CLUMSINESS | 0 | 0 | 0 | 0 | 0 | 0 | 0 | 0 | 0 | 0 |
| DYSKINESIA | 0 | 0 | 0 | 0 | 0 | 0 | 0 | 0 | 0 | 0 |
| EXTRAPYRAMIDAL DISORDER | 0 | 0 | 0 | 0 | 0 | 0 | 0 | 0 | 0 | 0 |
| PSYCHOMOTOR HYPERACTIVITY | 0 | 0 | 0 | 0 | 0 | 0 | 0 | 0 | 0 | 0 |
| *PARALYSIS AND PARESIS (EXCL CRANIAL NERVE)* | | | | | | | | | | |
| HEMIPLEGIA | 0 | 0 | 0 | 0 | 0 | 0 | 0 | 0 | 0 | 0 |
| MONOPARESIS | 2 | 2 | 0 | 0 | 0 | 2 | 2 | 0 | 0 | 0 |
| MONOPLEGIA | 0 | 0 | 0 | 0 | 0 | 0 | 0 | 0 | 0 | 0 |
| PARALYSIS | 1 | 1 | 0 | 0 | 0 | 1 | 1 | 0 | 0 | 0 |
| *PARKINSON'S DISEASE AND PARKINSONISM* | | | | | | | | | | |
| FREEZING PHENOMENON | 0 | 0 | 0 | 0 | 0 | 0 | 0 | 0 | 0 | 0 |
| PARKINSON'S DISEASE | 1 | 1 | 0 | 0 | 0 | 1 | 1 | 0 | 0 | 0 |
| *TREMOR (EXCL CONGENITAL)* | | | | | | | | | | |
| TREMOR | 19 | 10 | 2 | 7 | 0 | 16 | 10 | 1 | 5 | 0 |
| ***NEUROLOGICAL DISORDERS NEC*** | | | | | | | | | | |
| *COMA STATES* | | | | | | | | | | |
| DIABETIC HYPERGLYCAEMIC COMA | 0 | 0 | 0 | 0 | 0 | 0 | 0 | 0 | 0 | 0 |
| *COORDINATION AND BALANCE DISTURBANCES* | | | | | | | | | | |
| BALANCE DISORDER | 6 | 3 | 1 | 2 | 0 | 6 | 3 | 1 | 2 | 0 |
| COORDINATION ABNORMAL | 1 | 0 | 1 | 0 | 0 | 1 | 0 | 1 | 0 | 0 |
| DYSSTASIA | 1 | 1 | 0 | 0 | 0 | 1 | 1 | 0 | 0 | 0 |
| VESTIBULAR NYSTAGMUS | 0 | 0 | 0 | 0 | 0 | 0 | 0 | 0 | 0 | 0 |
| *DISTURBANCES IN CONSCIOUSNESS NEC* | | | | | | | | | | |
| DEPRESSED LEVEL OF CONSCIOUSNESS | 0 | 0 | 0 | 0 | 0 | 0 | 0 | 0 | 0 | 0 |
| LETHARGY | 75 | 35 | 20 | 20 | 0 | 65 | 35 | 15 | 15 | 0 |
| LOSS OF CONSCIOUSNESS | 2 | 1 | 1 | 0 | 0 | 2 | 1 | 1 | 0 | 0 |
| SEDATION | 0 | 0 | 0 | 0 | 0 | 0 | 0 | 0 | 0 | 0 |
| SOMNOLENCE | 32 | 19 | 9 | 4 | 0 | 30 | 19 | 7 | 4 | 0 |
| SYNCOPE | 12 | 8 | 4 | 0 | 0 | 10 | 8 | 2 | 0 | 0 |
| *NERVOUS SYSTEM DISORDERS NEC* | | | | | | | | | | |
| NERVOUS SYSTEM DISORDER | 0 | 0 | 0 | 0 | 0 | 0 | 0 | 0 | 0 | 0 |
| *NEUROLOGICAL SIGNS AND SYMPTOMS NEC* | | | | | | | | | | |
| AGITATION NEONATAL | 0 | 0 | 0 | 0 | 0 | 0 | 0 | 0 | 0 | 0 |

| | | | | | | | | | | |
|---|---|---|---|---|---|---|---|---|---|---|
| BRAIN FOG | 7 | 5 | 1 | 1 | 0 | 7 | 5 | 1 | 1 | 0 |
| DIZZINESS | 172 | 112 | 38 | 21 | 1 | 163 | 112 | 33 | 18 | 0 |
| DIZZINESS EXERTIONAL | 1 | 0 | 1 | 0 | 0 | 0 | 0 | 0 | 0 | 0 |
| DIZZINESS POSTURAL | 21 | 13 | 5 | 3 | 0 | 19 | 13 | 3 | 3 | 0 |
| HEAD DISCOMFORT | 4 | 3 | 0 | 1 | 0 | 4 | 3 | 0 | 1 | 0 |
| INFANT IRRITABILITY | 0 | 0 | 0 | 0 | 0 | 0 | 0 | 0 | 0 | 0 |
| MENINGISM | 0 | 0 | 0 | 0 | 0 | 0 | 0 | 0 | 0 | 0 |
| MYOCLONUS | 1 | 1 | 0 | 0 | 0 | 1 | 1 | 0 | 0 | 0 |
| NEUROLOGICAL SYMPTOM | 0 | 0 | 0 | 0 | 0 | 0 | 0 | 0 | 0 | 0 |
| PERSISTENT POSTURAL-PERCEPTUAL DIZZINESS | 0 | 0 | 0 | 0 | 0 | 0 | 0 | 0 | 0 | 0 |
| PRESYNCOPE | 8 | 7 | 0 | 1 | 0 | 8 | 7 | 0 | 1 | 0 |
| *PARAESTHESIAS AND DYSAESTHESIAS* | | | | | | | | | | |
| BURNING FEET SYNDROME | 1 | 1 | 0 | 0 | 0 | 1 | 1 | 0 | 0 | 0 |
| BURNING SENSATION | 0 | 0 | 0 | 0 | 0 | 0 | 0 | 0 | 0 | 0 |
| FORMICATION | 1 | 0 | 1 | 0 | 0 | 1 | 0 | 1 | 0 | 0 |
| HYPERAESTHESIA | 0 | 0 | 0 | 0 | 0 | 0 | 0 | 0 | 0 | 0 |
| HYPOAESTHESIA | 45 | 37 | 3 | 5 | 0 | 44 | 37 | 2 | 5 | 0 |
| PARAESTHESIA | 35 | 22 | 9 | 3 | 1 | 34 | 22 | 9 | 2 | 1 |
| REVERSED HOT-COLD SENSATION | 0 | 0 | 0 | 0 | 0 | 0 | 0 | 0 | 0 | 0 |
| *SENSORY ABNORMALITIES NEC* | | | | | | | | | | |
| AGEUSIA | 8 | 5 | 2 | 0 | 1 | 8 | 5 | 2 | 0 | 1 |
| ALLODYNIA | 0 | 0 | 0 | 0 | 0 | 0 | 0 | 0 | 0 | 0 |
| DYSGEUSIA | 17 | 9 | 5 | 3 | 0 | 16 | 9 | 4 | 3 | 0 |
| HYPOGEUSIA | 0 | 0 | 0 | 0 | 0 | 0 | 0 | 0 | 0 | 0 |
| NEURALGIA | 7 | 4 | 2 | 1 | 0 | 6 | 4 | 2 | 0 | 0 |
| POST HERPETIC NEURALGIA | 1 | 0 | 0 | 1 | 0 | 1 | 0 | 0 | 1 | 0 |
| RESTLESS ARM SYNDROME | 0 | 0 | 0 | 0 | 0 | 0 | 0 | 0 | 0 | 0 |
| RESTLESS LEGS SYNDROME | 0 | 0 | 0 | 0 | 0 | 0 | 0 | 0 | 0 | 0 |
| SENSORY LOSS | 1 | 1 | 0 | 0 | 0 | 1 | 1 | 0 | 0 | 0 |
| TASTE DISORDER | 7 | 2 | 4 | 1 | 0 | 6 | 2 | 3 | 1 | 0 |
| *SPEECH AND LANGUAGE ABNORMALITIES* | | | | | | | | | | |
| DYSARTHRIA | 0 | 0 | 0 | 0 | 0 | 0 | 0 | 0 | 0 | 0 |
| SPEECH DISORDER DEVELOPMENTAL | 0 | 0 | 0 | 0 | 0 | 0 | 0 | 0 | 0 | 0 |
| ***NEUROLOGICAL DISORDERS OF THE EYE*** | | | | | | | | | | |
| *NEUROLOGIC VISUAL PROBLEMS NEC* | | | | | | | | | | |

| | | | | | | | | | | |
|---|---|---|---|---|---|---|---|---|---|---|
| TUNNEL VISION | 0 | 0 | 0 | 0 | 0 | 0 | 0 | 0 | 0 | 0 |
| ***NEUROMUSCULAR DISORDERS*** | | | | | | | | | | |
| *MUSCLE TONE ABNORMAL* | | | | | | | | | | |
| HYPOTONIA | 0 | 0 | 0 | 0 | 0 | 0 | 0 | 0 | 0 | 0 |
| STIFF LEG SYNDROME | 0 | 0 | 0 | 0 | 0 | 0 | 0 | 0 | 0 | 0 |
| *NEUROMUSCULAR DISORDERS NEC* | | | | | | | | | | |
| MUSCLE SPASTICITY | 0 | 0 | 0 | 0 | 0 | 0 | 0 | 0 | 0 | 0 |
| ***PERIPHERAL NEUROPATHIES*** | | | | | | | | | | |
| *ACUTE POLYNEUROPATHIES* | | | | | | | | | | |
| GUILLAIN-BARRE SYNDROME | 0 | 0 | 0 | 0 | 0 | 0 | 0 | 0 | 0 | 0 |
| *PERIPHERAL NEUROPATHIES NEC* | | | | | | | | | | |
| AXONAL NEUROPATHY | 0 | 0 | 0 | 0 | 0 | 0 | 0 | 0 | 0 | 0 |
| NEUROPATHY PERIPHERAL | 1 | 0 | 0 | 1 | 0 | 1 | 0 | 0 | 1 | 0 |
| ***SEIZURES (INCL SUBTYPES)*** | | | | | | | | | | |
| *SEIZURES AND SEIZURE DISORDERS NEC* | | | | | | | | | | |
| EPILEPSY | 0 | 0 | 0 | 0 | 0 | 0 | 0 | 0 | 0 | 0 |
| SEIZURE | 2 | 2 | 0 | 0 | 0 | 2 | 2 | 0 | 0 | 0 |
| ***SLEEP DISTURBANCES (INCL SUBTYPES)*** | | | | | | | | | | |
| *DISTURBANCES IN SLEEP PHASE RHYTHM* | | | | | | | | | | |
| CIRCADIAN RHYTHM SLEEP DISORDER | 0 | 0 | 0 | 0 | 0 | 0 | 0 | 0 | 0 | 0 |
| *SLEEP DISTURBANCES NEC* | | | | | | | | | | |
| SLEEP DEFICIT | 1 | 1 | 0 | 0 | 0 | 1 | 1 | 0 | 0 | 0 |
| ***SPINAL CORD AND NERVE ROOT DISORDERS*** | | | | | | | | | | |
| *CERVICAL SPINAL CORD AND NERVE ROOT DISORDERS* | | | | | | | | | | |
| CERVICOBRACHIAL SYNDROME | 0 | 0 | 0 | 0 | 0 | 0 | 0 | 0 | 0 | 0 |
| *LUMBAR SPINAL CORD AND NERVE ROOT DISORDERS* | | | | | | | | | | |
| CAUDA EQUINA SYNDROME | 0 | 0 | 0 | 0 | 0 | 0 | 0 | 0 | 0 | 0 |
| SCIATICA | 2 | 0 | 0 | 2 | 0 | 2 | 0 | 0 | 2 | 0 |
| **PREGNANCY, PUERPERIUM AND PERINATAL CONDITIONS** | | | | | | | | | | |
| ***ABORTIONS AND STILLBIRTH*** | | | | | | | | | | |
| *ABORTIONS SPONTANEOUS* | | | | | | | | | | |
| ABORTION SPONTANEOUS | 15 | 8 | 7 | 0 | 0 | 14 | 8 | 6 | 0 | 0 |
| *STILLBIRTH AND FOETAL DEATH* | | | | | | | | | | |
| FOETAL DEATH | 0 | 0 | 0 | 0 | 0 | 0 | 0 | 0 | 0 | 0 |
| ***FOETAL COMPLICATIONS*** | | | | | | | | | | |
| *FOETAL COMPLICATIONS NEC* | | | | | | | | | | |

| | | | | | | | | | | |
|---|---|---|---|---|---|---|---|---|---|---|
| FOETAL DISORDER | 0 | 0 | 0 | 0 | 0 | 0 | 0 | 0 | 0 | 0 |
| FOETAL HYPOKINESIA | 2 | 1 | 0 | 1 | 0 | 1 | 1 | 0 | 0 | 0 |
| *FOETAL GROWTH COMPLICATIONS* | | | | | | | | | | |
| FOETAL MACROSOMIA | 1 | 1 | 0 | 0 | 0 | 1 | 1 | 0 | 0 | 0 |
| ***MATERNAL COMPLICATIONS OF PREGNANCY*** | | | | | | | | | | |
| *MATERNAL COMPLICATIONS OF PREGNANCY NEC* | | | | | | | | | | |
| MORNING SICKNESS | 1 | 1 | 0 | 0 | 0 | 1 | 1 | 0 | 0 | 0 |
| ***PLACENTAL, AMNIOTIC AND CAVITY DISORDERS (EXCL HAEMORRHAGES)*** | | | | | | | | | | |
| *PLACENTAL ABNORMALITIES (EXCL NEOPLASMS)* | | | | | | | | | | |
| PLACENTAL INFARCTION | 0 | 0 | 0 | 0 | 0 | 0 | 0 | 0 | 0 | 0 |
| ***PREGNANCY, LABOUR, DELIVERY AND POSTPARTUM CONDITIONS*** | | | | | | | | | | |
| *NORMAL PREGNANCY, LABOUR AND DELIVERY* | | | | | | | | | | |
| PREGNANCY | 0 | 0 | 0 | 0 | 0 | 0 | 0 | 0 | 0 | 0 |
| UTERINE CONTRACTIONS DURING PREGNANCY | 1 | 1 | 0 | 0 | 0 | 1 | 1 | 0 | 0 | 0 |
| **PRODUCT ISSUES** | | | | | | | | | | |
| ***DEVICE ISSUES*** | | | | | | | | | | |
| *DEVICE MALFUNCTION EVENTS NEC* | | | | | | | | | | |
| OVERSENSING | 0 | 0 | 0 | 0 | 0 | 0 | 0 | 0 | 0 | 0 |
| **PSYCHIATRIC DISORDERS** | | | | | | | | | | |
| ***ANXIETY DISORDERS AND SYMPTOMS*** | | | | | | | | | | |
| *ANXIETY SYMPTOMS* | | | | | | | | | | |
| AGITATION | 2 | 0 | 2 | 0 | 0 | 2 | 0 | 2 | 0 | 0 |
| ANXIETY | 6 | 5 | 1 | 0 | 0 | 5 | 5 | 0 | 0 | 0 |
| NERVOUSNESS | 1 | 1 | 0 | 0 | 0 | 1 | 1 | 0 | 0 | 0 |
| TENSION | 1 | 1 | 0 | 0 | 0 | 1 | 1 | 0 | 0 | 0 |
| *PANIC ATTACKS AND DISORDERS* | | | | | | | | | | |
| PANIC ATTACK | 2 | 2 | 0 | 0 | 0 | 2 | 2 | 0 | 0 | 0 |
| ***CHANGES IN PHYSICAL ACTIVITY*** | | | | | | | | | | |
| *INCREASED PHYSICAL ACTIVITY LEVELS* | | | | | | | | | | |
| RESTLESSNESS | 2 | 2 | 0 | 0 | 0 | 2 | 2 | 0 | 0 | 0 |
| *STEREOTYPIES AND AUTOMATISMS* | | | | | | | | | | |
| BRUXISM | 0 | 0 | 0 | 0 | 0 | 0 | 0 | 0 | 0 | 0 |
| ***COGNITIVE AND ATTENTION DISORDERS AND DISTURBANCES*** | | | | | | | | | | |
| *COGNITIVE AND ATTENTION DISORDERS AND DISTURBANCES NEC* | | | | | | | | | | |
| DAYDREAMING | 0 | 0 | 0 | 0 | 0 | 0 | 0 | 0 | 0 | 0 |
| MENTAL FATIGUE | 10 | 7 | 1 | 2 | 0 | 9 | 7 | 1 | 1 | 0 |

| | | | | | | | | | | |
|---|---|---|---|---|---|---|---|---|---|---|
| ***COMMUNICATION DISORDERS AND DISTURBANCES*** | | | | | | | | | | |
| *SPEECH ARTICULATION AND RHYTHM DISTURBANCES* | | | | | | | | | | |
| DYSPHEMIA | 0 | 0 | 0 | 0 | 0 | 0 | 0 | 0 | 0 | 0 |
| ***DELIRIA (INCL CONFUSION)*** | | | | | | | | | | |
| *CONFUSION AND DISORIENTATION* | | | | | | | | | | |
| CONFUSIONAL STATE | 10 | 7 | 1 | 1 | 1 | 9 | 7 | 0 | 1 | 1 |
| DISORIENTATION | 0 | 0 | 0 | 0 | 0 | 0 | 0 | 0 | 0 | 0 |
| *DELIRIA* | | | | | | | | | | |
| DELIRIUM | 0 | 0 | 0 | 0 | 0 | 0 | 0 | 0 | 0 | 0 |
| ***DEPRESSED MOOD DISORDERS AND DISTURBANCES*** | | | | | | | | | | |
| *DEPRESSIVE DISORDERS* | | | | | | | | | | |
| DEPRESSION | 5 | 4 | 1 | 0 | 0 | 5 | 4 | 1 | 0 | 0 |
| *MOOD ALTERATIONS WITH DEPRESSIVE SYMPTOMS* | | | | | | | | | | |
| DEPRESSED MOOD | 3 | 1 | 1 | 1 | 0 | 3 | 1 | 1 | 1 | 0 |
| TEARFULNESS | 0 | 0 | 0 | 0 | 0 | 0 | 0 | 0 | 0 | 0 |
| ***DISSOCIATIVE DISORDERS*** | | | | | | | | | | |
| *DISSOCIATIVE STATES* | | | | | | | | | | |
| DISSOCIATION | 1 | 0 | 0 | 1 | 0 | 1 | 0 | 0 | 1 | 0 |
| ***DISTURBANCES IN THINKING AND PERCEPTION*** | | | | | | | | | | |
| *DELUSIONAL SYMPTOMS* | | | | | | | | | | |
| DELUSION | 0 | 0 | 0 | 0 | 0 | 0 | 0 | 0 | 0 | 0 |
| *HALLUCINATIONS (EXCL SLEEP-RELATED)* | | | | | | | | | | |
| HALLUCINATION | 2 | 2 | 0 | 0 | 0 | 2 | 2 | 0 | 0 | 0 |
| *PERCEPTION DISTURBANCES NEC* | | | | | | | | | | |
| DEREALISATION | 1 | 1 | 0 | 0 | 0 | 1 | 1 | 0 | 0 | 0 |
| *THINKING DISTURBANCES* | | | | | | | | | | |
| BRADYPHRENIA | 0 | 0 | 0 | 0 | 0 | 0 | 0 | 0 | 0 | 0 |
| THOUGHT BLOCKING | 0 | 0 | 0 | 0 | 0 | 0 | 0 | 0 | 0 | 0 |
| ***MOOD DISORDERS AND DISTURBANCES NEC*** | | | | | | | | | | |
| *AFFECT ALTERATIONS NEC* | | | | | | | | | | |
| AFFECT LABILITY | 0 | 0 | 0 | 0 | 0 | 0 | 0 | 0 | 0 | 0 |
| INAPPROPRIATE AFFECT | 1 | 1 | 0 | 0 | 0 | 1 | 1 | 0 | 0 | 0 |
| *EMOTIONAL AND MOOD DISTURBANCES NEC* | | | | | | | | | | |
| ANGER | 0 | 0 | 0 | 0 | 0 | 0 | 0 | 0 | 0 | 0 |
| EMOTIONAL DISORDER | 1 | 0 | 1 | 0 | 0 | 1 | 0 | 1 | 0 | 0 |
| EMOTIONAL DISTRESS | 0 | 0 | 0 | 0 | 0 | 0 | 0 | 0 | 0 | 0 |

| | | | | | | | | | | |
|---|---|---|---|---|---|---|---|---|---|---|
| EUPHORIC MOOD | 0 | 0 | 0 | 0 | 0 | 0 | 0 | 0 | 0 | 0 |
| IRRITABILITY | 5 | 5 | 0 | 0 | 0 | 5 | 5 | 0 | 0 | 0 |
| MOOD ALTERED | 1 | 0 | 1 | 0 | 0 | 0 | 0 | 0 | 0 | 0 |
| *FLUCTUATING MOOD SYMPTOMS* | | | | | | | | | | |
| MOOD SWINGS | 1 | 0 | 1 | 0 | 0 | 1 | 0 | 1 | 0 | 0 |
| *MOOD DISORDERS NEC* | | | | | | | | | | |
| APATHY | 2 | 1 | 0 | 1 | 0 | 2 | 1 | 0 | 1 | 0 |
| LISTLESS | 1 | 0 | 0 | 0 | 1 | 1 | 0 | 0 | 0 | 1 |
| ***PERSONALITY DISORDERS AND DISTURBANCES IN BEHAVIOUR*** | | | | | | | | | | |
| *BEHAVIOUR AND SOCIALISATION DISTURBANCES* | | | | | | | | | | |
| AGGRESSION | 0 | 0 | 0 | 0 | 0 | 0 | 0 | 0 | 0 | 0 |
| ***PSYCHIATRIC AND BEHAVIOURAL SYMPTOMS NEC*** | | | | | | | | | | |
| *PSYCHIATRIC SYMPTOMS NEC* | | | | | | | | | | |
| PSYCHIATRIC SYMPTOM | 0 | 0 | 0 | 0 | 0 | 0 | 0 | 0 | 0 | 0 |
| ***PSYCHIATRIC DISORDERS NEC*** | | | | | | | | | | |
| *MENTAL DISORDERS NEC* | | | | | | | | | | |
| MENTAL DISORDER | 0 | 0 | 0 | 0 | 0 | 0 | 0 | 0 | 0 | 0 |
| ***SCHIZOPHRENIA AND OTHER PSYCHOTIC DISORDERS*** | | | | | | | | | | |
| *PSYCHOTIC DISORDER NEC* | | | | | | | | | | |
| PSYCHOTIC DISORDER | 0 | 0 | 0 | 0 | 0 | 0 | 0 | 0 | 0 | 0 |
| ***SEXUAL DYSFUNCTIONS, DISTURBANCES AND GENDER IDENTITY DISORDERS*** | | | | | | | | | | |
| *SEXUAL DESIRE DISORDERS* | | | | | | | | | | |
| LOSS OF LIBIDO | 1 | 1 | 0 | 0 | 0 | 1 | 1 | 0 | 0 | 0 |
| ***SLEEP DISORDERS AND DISTURBANCES*** | | | | | | | | | | |
| *DISTURBANCES IN INITIATING AND MAINTAINING SLEEP* | | | | | | | | | | |
| INITIAL INSOMNIA | 1 | 1 | 0 | 0 | 0 | 1 | 1 | 0 | 0 | 0 |
| INSOMNIA | 22 | 11 | 4 | 6 | 1 | 19 | 11 | 3 | 5 | 0 |
| *DYSSOMNIAS* | | | | | | | | | | |
| BREATHING-RELATED SLEEP DISORDER | 0 | 0 | 0 | 0 | 0 | 0 | 0 | 0 | 0 | 0 |
| POOR QUALITY SLEEP | 9 | 5 | 1 | 3 | 0 | 9 | 5 | 1 | 3 | 0 |
| *PARASOMNIAS* | | | | | | | | | | |
| ABNORMAL DREAMS | 6 | 4 | 1 | 0 | 1 | 5 | 4 | 1 | 0 | 0 |
| EXPLODING HEAD SYNDROME | 1 | 0 | 1 | 0 | 0 | 1 | 0 | 1 | 0 | 0 |
| NIGHTMARE | 5 | 1 | 2 | 2 | 0 | 5 | 1 | 2 | 2 | 0 |
| *SLEEP DISORDERS NEC* | | | | | | | | | | |
| SLEEP DISORDER | 2 | 1 | 1 | 0 | 0 | 2 | 1 | 1 | 0 | 0 |

| | | | | | | | | | | |
|---|---|---|---|---|---|---|---|---|---|---|
| ***SOMATIC SYMPTOM AND RELATED DISORDERS*** | | | | | | | | | | |
| *SOMATIC SYMPTOM DISORDERS* | | | | | | | | | | |
| HABIT COUGH | 1 | 0 | 0 | 1 | 0 | 1 | 0 | 0 | 1 | 0 |
| ***SUICIDAL AND SELF-INJURIOUS BEHAVIOURS NEC*** | | | | | | | | | | |
| *SUICIDAL AND SELF-INJURIOUS BEHAVIOUR* | | | | | | | | | | |
| SUICIDAL IDEATION | 1 | 1 | 0 | 0 | 0 | 1 | 1 | 0 | 0 | 0 |
| **RENAL AND URINARY DISORDERS** | | | | | | | | | | |
| ***RENAL DISORDERS (EXCL NEPHROPATHIES)*** | | | | | | | | | | |
| *RENAL FAILURE AND IMPAIRMENT* | | | | | | | | | | |
| RENAL FAILURE | 1 | 1 | 0 | 0 | 0 | 1 | 1 | 0 | 0 | 0 |
| ***URINARY TRACT SIGNS AND SYMPTOMS*** | | | | | | | | | | |
| *BLADDER AND URETHRAL SYMPTOMS* | | | | | | | | | | |
| BLADDER PAIN | 0 | 0 | 0 | 0 | 0 | 0 | 0 | 0 | 0 | 0 |
| MICTURITION URGENCY | 1 | 0 | 0 | 1 | 0 | 1 | 0 | 0 | 1 | 0 |
| POLLAKIURIA | 1 | 1 | 0 | 0 | 0 | 1 | 1 | 0 | 0 | 0 |
| URINARY INCONTINENCE | 0 | 0 | 0 | 0 | 0 | 0 | 0 | 0 | 0 | 0 |
| *URINARY ABNORMALITIES* | | | | | | | | | | |
| HAEMATURIA | 1 | 1 | 0 | 0 | 0 | 1 | 1 | 0 | 0 | 0 |
| URINE ABNORMALITY | 0 | 0 | 0 | 0 | 0 | 0 | 0 | 0 | 0 | 0 |
| URINE ODOUR ABNORMAL | 0 | 0 | 0 | 0 | 0 | 0 | 0 | 0 | 0 | 0 |
| *URINARY TRACT SIGNS AND SYMPTOMS NEC* | | | | | | | | | | |
| POLYURIA | 0 | 0 | 0 | 0 | 0 | 0 | 0 | 0 | 0 | 0 |
| RENAL PAIN | 0 | 0 | 0 | 0 | 0 | 0 | 0 | 0 | 0 | 0 |
| **REPRODUCTIVE SYSTEM AND BREAST DISORDERS** | | | | | | | | | | |
| ***BREAST DISORDERS*** | | | | | | | | | | |
| *BREAST DISORDERS NEC* | | | | | | | | | | |
| BREAST MASS | 1 | 1 | 0 | 0 | 0 | 1 | 1 | 0 | 0 | 0 |
| NIPPLE ENLARGEMENT | 0 | 0 | 0 | 0 | 0 | 0 | 0 | 0 | 0 | 0 |
| *BREAST SIGNS AND SYMPTOMS* | | | | | | | | | | |
| BREAST DISCHARGE | 0 | 0 | 0 | 0 | 0 | 0 | 0 | 0 | 0 | 0 |
| BREAST PAIN | 4 | 1 | 1 | 2 | 0 | 2 | 1 | 0 | 1 | 0 |
| BREAST SWELLING | 0 | 0 | 0 | 0 | 0 | 0 | 0 | 0 | 0 | 0 |
| BREAST TENDERNESS | 0 | 0 | 0 | 0 | 0 | 0 | 0 | 0 | 0 | 0 |
| NIPPLE PAIN | 0 | 0 | 0 | 0 | 0 | 0 | 0 | 0 | 0 | 0 |
| ***MENOPAUSE RELATED CONDITIONS*** | | | | | | | | | | |
| *MENOPAUSAL EFFECTS ON THE GENITOURINARY TRACT* | | | | | | | | | | |

| | | | | | | | | | | |
|---|---|---|---|---|---|---|---|---|---|---|
| POSTMENOPAUSAL HAEMORRHAGE | 0 | 0 | 0 | 0 | 0 | 0 | 0 | 0 | 0 | 0 |
| ***MENSTRUAL CYCLE AND UTERINE BLEEDING DISORDERS*** | | | | | | | | | | |
| *MENSTRUATION AND UTERINE BLEEDING NEC* | | | | | | | | | | |
| DYSMENORRHOEA | 19 | 15 | 2 | 2 | 0 | 16 | 15 | 1 | 0 | 0 |
| INTERMENSTRUAL BLEEDING | 5 | 5 | 0 | 0 | 0 | 5 | 5 | 0 | 0 | 0 |
| MENSTRUAL DISORDER | 12 | 7 | 0 | 5 | 0 | 12 | 7 | 0 | 5 | 0 |
| MENSTRUATION IRREGULAR | 23 | 16 | 6 | 1 | 0 | 21 | 16 | 4 | 1 | 0 |
| PREMENSTRUAL PAIN | 0 | 0 | 0 | 0 | 0 | 0 | 0 | 0 | 0 | 0 |
| RETROGRADE MENSTRUATION | 0 | 0 | 0 | 0 | 0 | 0 | 0 | 0 | 0 | 0 |
| *MENSTRUATION WITH DECREASED BLEEDING* | | | | | | | | | | |
| AMENORRHOEA | 1 | 1 | 0 | 0 | 0 | 1 | 1 | 0 | 0 | 0 |
| HYPOMENORRHOEA | 2 | 1 | 1 | 0 | 0 | 2 | 1 | 1 | 0 | 0 |
| MENSTRUATION DELAYED | 20 | 14 | 5 | 1 | 0 | 19 | 14 | 4 | 1 | 0 |
| OLIGOMENORRHOEA | 2 | 1 | 1 | 0 | 0 | 2 | 1 | 1 | 0 | 0 |
| *MENSTRUATION WITH INCREASED BLEEDING* | | | | | | | | | | |
| HEAVY MENSTRUAL BLEEDING | 24 | 17 | 2 | 5 | 0 | 23 | 17 | 2 | 4 | 0 |
| POLYMENORRHOEA | 5 | 4 | 0 | 1 | 0 | 5 | 4 | 0 | 1 | 0 |
| ***PENILE AND SCROTAL DISORDERS (EXCL INFECTIONS AND INFLAMMATIONS)*** | | | | | | | | | | |
| *SCROTAL DISORDERS NEC* | | | | | | | | | | |
| SCROTAL SWELLING | 0 | 0 | 0 | 0 | 0 | 0 | 0 | 0 | 0 | 0 |
| ***REPRODUCTIVE TRACT DISORDERS NEC*** | | | | | | | | | | |
| *REPRODUCTIVE TRACT DISORDERS NEC (EXCL NEOPLASMS)* | | | | | | | | | | |
| GENITAL LESION | 0 | 0 | 0 | 0 | 0 | 0 | 0 | 0 | 0 | 0 |
| *REPRODUCTIVE TRACT SIGNS AND SYMPTOMS NEC* | | | | | | | | | | |
| GENITAL DISCOMFORT | 0 | 0 | 0 | 0 | 0 | 0 | 0 | 0 | 0 | 0 |
| GENITAL PAIN | 0 | 0 | 0 | 0 | 0 | 0 | 0 | 0 | 0 | 0 |
| ***UTERINE, PELVIC AND BROAD LIGAMENT DISORDERS*** | | | | | | | | | | |
| *PELVIS AND BROAD LIGAMENT DISORDERS NEC* | | | | | | | | | | |
| ADNEXA UTERI PAIN | 0 | 0 | 0 | 0 | 0 | 0 | 0 | 0 | 0 | 0 |
| ***VULVOVAGINAL DISORDERS (EXCL INFECTIONS AND INFLAMMATIONS)*** | | | | | | | | | | |
| *VULVOVAGINAL DISORDERS NEC* | | | | | | | | | | |
| VAGINAL HAEMORRHAGE | 9 | 4 | 4 | 1 | 0 | 6 | 4 | 2 | 0 | 0 |
| *VULVOVAGINAL SIGNS AND SYMPTOMS* | | | | | | | | | | |
| VAGINAL DISCHARGE | 0 | 0 | 0 | 0 | 0 | 0 | 0 | 0 | 0 | 0 |
| **RESPIRATORY, THORACIC AND MEDIASTINAL DISORDERS** | | | | | | | | | | |
| ***BRONCHIAL DISORDERS (EXCL NEOPLASMS)*** | | | | | | | | | | |

| | | | | | | | | | | |
|---|---|---|---|---|---|---|---|---|---|---|
| *BRONCHOSPASM AND OBSTRUCTION* | | | | | | | | | | |
| ASTHMA | 1 | 1 | 0 | 0 | 0 | 1 | 1 | 0 | 0 | 0 |
| WHEEZING | 1 | 0 | 0 | 1 | 0 | 0 | 0 | 0 | 0 | 0 |
| ***LOWER RESPIRATORY TRACT DISORDERS (EXCL OBSTRUCTION AND INFECTION)*** | | | | | | | | | | |
| *PULMONARY OEDEMAS* | | | | | | | | | | |
| PULMONARY CONGESTION | 1 | 1 | 0 | 0 | 0 | 1 | 1 | 0 | 0 | 0 |
| ***PULMONARY VASCULAR DISORDERS*** | | | | | | | | | | |
| *PULMONARY THROMBOTIC AND EMBOLIC CONDITIONS* | | | | | | | | | | |
| PULMONARY EMBOLISM | 0 | 0 | 0 | 0 | 0 | 0 | 0 | 0 | 0 | 0 |
| ***RESPIRATORY DISORDERS NEC*** | | | | | | | | | | |
| *BREATHING ABNORMALITIES* | | | | | | | | | | |
| DYSPNOEA | 36 | 20 | 6 | 8 | 2 | 35 | 20 | 5 | 8 | 2 |
| HYPERVENTILATION | 1 | 1 | 0 | 0 | 0 | 1 | 1 | 0 | 0 | 0 |
| HYPOPNOEA | 1 | 0 | 0 | 1 | 0 | 1 | 0 | 0 | 1 | 0 |
| IRREGULAR BREATHING | 0 | 0 | 0 | 0 | 0 | 0 | 0 | 0 | 0 | 0 |
| MOUTH BREATHING | 0 | 0 | 0 | 0 | 0 | 0 | 0 | 0 | 0 | 0 |
| RESPIRATION ABNORMAL | 1 | 1 | 0 | 0 | 0 | 1 | 1 | 0 | 0 | 0 |
| RESPIRATORY ARREST | 1 | 1 | 0 | 0 | 0 | 1 | 1 | 0 | 0 | 0 |
| RESPIRATORY FATIGUE | 0 | 0 | 0 | 0 | 0 | 0 | 0 | 0 | 0 | 0 |
| SLEEP APNOEA SYNDROME | 0 | 0 | 0 | 0 | 0 | 0 | 0 | 0 | 0 | 0 |
| *COUGHING AND ASSOCIATED SYMPTOMS* | | | | | | | | | | |
| COUGH | 49 | 32 | 5 | 11 | 1 | 47 | 32 | 5 | 10 | 0 |
| PRODUCTIVE COUGH | 5 | 2 | 3 | 0 | 0 | 5 | 2 | 3 | 0 | 0 |
| *RESPIRATORY TRACT DISORDERS NEC* | | | | | | | | | | |
| RESPIRATORY TRACT IRRITATION | 0 | 0 | 0 | 0 | 0 | 0 | 0 | 0 | 0 | 0 |
| ***RESPIRATORY TRACT SIGNS AND SYMPTOMS*** | | | | | | | | | | |
| *LOWER RESPIRATORY TRACT SIGNS AND SYMPTOMS* | | | | | | | | | | |
| PULMONARY PAIN | 1 | 1 | 0 | 0 | 0 | 1 | 1 | 0 | 0 | 0 |
| *RESPIRATORY SIGNS AND SYMPTOMS NEC* | | | | | | | | | | |
| RESPIRATORY SYMPTOM | 1 | 1 | 0 | 0 | 0 | 1 | 1 | 0 | 0 | 0 |
| *UPPER RESPIRATORY TRACT SIGNS AND SYMPTOMS* | | | | | | | | | | |
| APHONIA | 1 | 0 | 0 | 1 | 0 | 1 | 0 | 0 | 1 | 0 |
| CATARRH | 2 | 2 | 0 | 0 | 0 | 2 | 2 | 0 | 0 | 0 |
| DRY THROAT | 4 | 2 | 2 | 0 | 0 | 3 | 2 | 1 | 0 | 0 |
| DYSPHONIA | 2 | 2 | 0 | 0 | 0 | 2 | 2 | 0 | 0 | 0 |
| INCREASED UPPER AIRWAY SECRETION | 1 | 1 | 0 | 0 | 0 | 1 | 1 | 0 | 0 | 0 |

| | | | | | | | | | | |
|---|---|---|---|---|---|---|---|---|---|---|
| INCREASED VISCOSITY OF UPPER RESPIRATORY SE( | 1 | 0 | 0 | 1 | 0 | 1 | 0 | 0 | 1 | 0 |
| NASAL DISCOMFORT | 0 | 0 | 0 | 0 | 0 | 0 | 0 | 0 | 0 | 0 |
| OROPHARYNGEAL DISCOMFORT | 0 | 0 | 0 | 0 | 0 | 0 | 0 | 0 | 0 | 0 |
| OROPHARYNGEAL PAIN | 82 | 57 | 18 | 7 | 0 | 79 | 57 | 16 | 6 | 0 |
| PARANASAL SINUS DISCOMFORT | 1 | 1 | 0 | 0 | 0 | 1 | 1 | 0 | 0 | 0 |
| RHINALGIA | 0 | 0 | 0 | 0 | 0 | 0 | 0 | 0 | 0 | 0 |
| RHINORRHOEA | 48 | 30 | 8 | 9 | 1 | 41 | 30 | 4 | 6 | 1 |
| SINUS PAIN | 5 | 3 | 2 | 0 | 0 | 5 | 3 | 2 | 0 | 0 |
| SNEEZING | 13 | 10 | 2 | 0 | 1 | 13 | 10 | 2 | 0 | 1 |
| THROAT CLEARING | 1 | 1 | 0 | 0 | 0 | 1 | 1 | 0 | 0 | 0 |
| THROAT IRRITATION | 1 | 1 | 0 | 0 | 0 | 1 | 1 | 0 | 0 | 0 |
| THROAT TIGHTNESS | 1 | 1 | 0 | 0 | 0 | 1 | 1 | 0 | 0 | 0 |
| UPPER-AIRWAY COUGH SYNDROME | 1 | 0 | 0 | 1 | 0 | 1 | 0 | 0 | 1 | 0 |
| YAWNING | 0 | 0 | 0 | 0 | 0 | 0 | 0 | 0 | 0 | 0 |
| ***UPPER RESPIRATORY TRACT DISORDERS (EXCL INFECTIONS)*** | | | | | | | | | | |
| *NASAL CONGESTION AND INFLAMMATIONS* | | | | | | | | | | |
| NASAL CONGESTION | 5 | 2 | 3 | 0 | 0 | 4 | 2 | 2 | 0 | 0 |
| RHINITIS ALLERGIC | 1 | 1 | 0 | 0 | 0 | 1 | 1 | 0 | 0 | 0 |
| RHINITIS ATROPHIC | 0 | 0 | 0 | 0 | 0 | 0 | 0 | 0 | 0 | 0 |
| *NASAL DISORDERS NEC* | | | | | | | | | | |
| EPISTAXIS | 8 | 5 | 1 | 2 | 0 | 6 | 5 | 1 | 0 | 0 |
| NASAL DRYNESS | 2 | 2 | 0 | 0 | 0 | 2 | 2 | 0 | 0 | 0 |
| *PARANASAL SINUS DISORDERS (EXCL INFECTIONS AND NEOPLASMS)* | | | | | | | | | | |
| SINUS CONGESTION | 1 | 1 | 0 | 0 | 0 | 1 | 1 | 0 | 0 | 0 |
| *PHARYNGEAL DISORDERS (EXCL INFECTIONS AND NEOPLASMS)* | | | | | | | | | | |
| PHARYNGEAL SWELLING | 0 | 0 | 0 | 0 | 0 | 0 | 0 | 0 | 0 | 0 |
| PHARYNGEAL ULCERATION | 0 | 0 | 0 | 0 | 0 | 0 | 0 | 0 | 0 | 0 |
| TONSILLAR ERYTHEMA | 0 | 0 | 0 | 0 | 0 | 0 | 0 | 0 | 0 | 0 |
| *TRACHEAL DISORDERS (EXCL INFECTIONS AND NEOPLASMS)* | | | | | | | | | | |
| TRACHEAL PAIN | 0 | 0 | 0 | 0 | 0 | 0 | 0 | 0 | 0 | 0 |
| **SKIN AND SUBCUTANEOUS TISSUE DISORDERS** | | | | | | | | | | |
| ***ANGIOEDEMA AND URTICARIA*** | | | | | | | | | | |
| *ANGIOEDEMAS* | | | | | | | | | | |
| ANGIOEDEMA | 0 | 0 | 0 | 0 | 0 | 0 | 0 | 0 | 0 | 0 |
| *URTICARIAS* | | | | | | | | | | |

| | | | | | | | | | | |
|---|---|---|---|---|---|---|---|---|---|---|
| COLD URTICARIA | 0 | 0 | 0 | 0 | 0 | 0 | 0 | 0 | 0 | 0 |
| SOLAR URTICARIA | 0 | 0 | 0 | 0 | 0 | 0 | 0 | 0 | 0 | 0 |
| URTICARIA | 10 | 4 | 1 | 5 | 0 | 10 | 4 | 1 | 5 | 0 |
| URTICARIA CHRONIC | 0 | 0 | 0 | 0 | 0 | 0 | 0 | 0 | 0 | 0 |
| ***CORNIFICATION AND DYSTROPHIC SKIN DISORDERS*** | | | | | | | | | | |
| *SKIN DYSTROPHIES* | | | | | | | | | | |
| HYPERTROPHIC SCAR | 1 | 1 | 0 | 0 | 0 | 1 | 1 | 0 | 0 | 0 |
| ***CUTANEOUS NEOPLASMS BENIGN*** | | | | | | | | | | |
| *SKIN CYSTS AND POLYPS* | | | | | | | | | | |
| DERMAL CYST | 1 | 1 | 0 | 0 | 0 | 1 | 1 | 0 | 0 | 0 |
| ***EPIDERMAL AND DERMAL CONDITIONS*** | | | | | | | | | | |
| *BULLOUS CONDITIONS* | | | | | | | | | | |
| BLISTER | 1 | 1 | 0 | 0 | 0 | 1 | 1 | 0 | 0 | 0 |
| BLOOD BLISTER | 0 | 0 | 0 | 0 | 0 | 0 | 0 | 0 | 0 | 0 |
| PEMPHIGOID | 1 | 0 | 0 | 0 | 1 | 1 | 0 | 0 | 0 | 1 |
| TOXIC EPIDERMAL NECROLYSIS | 1 | 1 | 0 | 0 | 0 | 1 | 1 | 0 | 0 | 0 |
| *DERMAL AND EPIDERMAL CONDITIONS NEC* | | | | | | | | | | |
| DRY SKIN | 2 | 2 | 0 | 0 | 0 | 2 | 2 | 0 | 0 | 0 |
| PAIN OF SKIN | 12 | 8 | 1 | 3 | 0 | 12 | 8 | 1 | 3 | 0 |
| SENSITIVE SKIN | 7 | 3 | 2 | 1 | 1 | 5 | 3 | 1 | 1 | 0 |
| SKIN BURNING SENSATION | 3 | 2 | 0 | 1 | 0 | 3 | 2 | 0 | 1 | 0 |
| SKIN DISCOLOURATION | 1 | 1 | 0 | 0 | 0 | 1 | 1 | 0 | 0 | 0 |
| SKIN FRAGILITY | 1 | 1 | 0 | 0 | 0 | 1 | 1 | 0 | 0 | 0 |
| SKIN LESION | 0 | 0 | 0 | 0 | 0 | 0 | 0 | 0 | 0 | 0 |
| SKIN ODOUR ABNORMAL | 0 | 0 | 0 | 0 | 0 | 0 | 0 | 0 | 0 | 0 |
| SKIN REACTION | 1 | 0 | 0 | 1 | 0 | 1 | 0 | 0 | 1 | 0 |
| SKIN SENSITISATION | 0 | 0 | 0 | 0 | 0 | 0 | 0 | 0 | 0 | 0 |
| SKIN WARM | 14 | 9 | 1 | 4 | 0 | 14 | 9 | 1 | 4 | 0 |
| *DERMATITIS AND ECZEMA* | | | | | | | | | | |
| DERMATITIS | 2 | 2 | 0 | 0 | 0 | 2 | 2 | 0 | 0 | 0 |
| DERMATITIS ALLERGIC | 0 | 0 | 0 | 0 | 0 | 0 | 0 | 0 | 0 | 0 |
| DERMATITIS ATOPIC | 0 | 0 | 0 | 0 | 0 | 0 | 0 | 0 | 0 | 0 |
| DERMATITIS CONTACT | 0 | 0 | 0 | 0 | 0 | 0 | 0 | 0 | 0 | 0 |
| ECZEMA | 4 | 3 | 1 | 0 | 0 | 4 | 3 | 1 | 0 | 0 |
| ECZEMA ASTEATOTIC | 1 | 0 | 0 | 1 | 0 | 1 | 0 | 0 | 1 | 0 |

| | | | | | | | | | | |
|---|---|---|---|---|---|---|---|---|---|---|
| NEURODERMATITIS | 0 | 0 | 0 | 0 | 0 | 0 | 0 | 0 | 0 | 0 |
| SEBORRHOEIC DERMATITIS | 0 | 0 | 0 | 0 | 0 | 0 | 0 | 0 | 0 | 0 |
| SKIN IRRITATION | 2 | 0 | 1 | 1 | 0 | 1 | 0 | 0 | 1 | 0 |
| *ERYTHEMAS* | | | | | | | | | | |
| ERYTHEMA | 21 | 12 | 8 | 1 | 0 | 20 | 12 | 8 | 0 | 0 |
| *EXFOLIATIVE CONDITIONS* | | | | | | | | | | |
| SKIN EXFOLIATION | 0 | 0 | 0 | 0 | 0 | 0 | 0 | 0 | 0 | 0 |
| *PAPULOSQUAMOUS CONDITIONS* | | | | | | | | | | |
| LICHEN PLANUS | 0 | 0 | 0 | 0 | 0 | 0 | 0 | 0 | 0 | 0 |
| PITYRIASIS ROSEA | 0 | 0 | 0 | 0 | 0 | 0 | 0 | 0 | 0 | 0 |
| *PHOTOSENSITIVITY AND PHOTODERMATOSIS CONDITIONS* | | | | | | | | | | |
| PHOTOSENSITIVITY REACTION | 1 | 1 | 0 | 0 | 0 | 1 | 1 | 0 | 0 | 0 |
| *PRURITUS NEC* | | | | | | | | | | |
| PRURITUS | 51 | 30 | 13 | 8 | 0 | 45 | 30 | 9 | 6 | 0 |
| *PSORIATIC CONDITIONS* | | | | | | | | | | |
| PSORIASIS | 2 | 0 | 1 | 1 | 0 | 2 | 0 | 1 | 1 | 0 |
| *RASHES, ERUPTIONS AND EXANTHEMS NEC* | | | | | | | | | | |
| RASH | 46 | 31 | 5 | 10 | 0 | 42 | 31 | 5 | 6 | 0 |
| RASH ERYTHEMATOUS | 9 | 4 | 1 | 4 | 0 | 9 | 4 | 1 | 4 | 0 |
| RASH MACULAR | 2 | 2 | 0 | 0 | 0 | 2 | 2 | 0 | 0 | 0 |
| RASH MORBILLIFORM | 1 | 0 | 0 | 1 | 0 | 0 | 0 | 0 | 0 | 0 |
| RASH PAPULAR | 2 | 1 | 1 | 0 | 0 | 2 | 1 | 1 | 0 | 0 |
| RASH PRURITIC | 6 | 3 | 2 | 1 | 0 | 6 | 3 | 2 | 1 | 0 |
| *SKIN INJURIES AND MECHANICAL DERMATOSES* | | | | | | | | | | |
| DECUBITUS ULCER | 1 | 0 | 0 | 1 | 0 | 0 | 0 | 0 | 0 | 0 |
| NEEDLE TRACK MARKS | 0 | 0 | 0 | 0 | 0 | 0 | 0 | 0 | 0 | 0 |
| ***SKIN AND SUBCUTANEOUS TISSUE DISORDERS NEC*** | | | | | | | | | | |
| *SKIN AND SUBCUTANEOUS TISSUE ULCERATIONS* | | | | | | | | | | |
| SKIN EROSION | 0 | 0 | 0 | 0 | 0 | 0 | 0 | 0 | 0 | 0 |
| ***SKIN APPENDAGE CONDITIONS*** | | | | | | | | | | |
| *ACNES* | | | | | | | | | | |
| ACNE | 0 | 0 | 0 | 0 | 0 | 0 | 0 | 0 | 0 | 0 |
| ACNE CYSTIC | 0 | 0 | 0 | 0 | 0 | 0 | 0 | 0 | 0 | 0 |
| *ALOPECIAS* | | | | | | | | | | |
| ALOPECIA | 3 | 3 | 0 | 0 | 0 | 3 | 3 | 0 | 0 | 0 |
| *APOCRINE AND ECCRINE GLAND DISORDERS* | | | | | | | | | | |

| | | | | | | | | | | |
|---|---|---|---|---|---|---|---|---|---|---|
| COLD SWEAT | 6 | 2 | 3 | 0 | 1 | 5 | 2 | 2 | 0 | 1 |
| HYPERHIDROSIS | 27 | 19 | 5 | 3 | 0 | 26 | 19 | 5 | 2 | 0 |
| MILIARIA | 3 | 1 | 2 | 0 | 0 | 2 | 1 | 1 | 0 | 0 |
| NIGHT SWEATS | 20 | 9 | 6 | 4 | 1 | 17 | 9 | 5 | 3 | 0 |
| *HYPERTRICHOSES* | | | | | | | | | | |
| HIRSUTISM | 1 | 1 | 0 | 0 | 0 | 1 | 1 | 0 | 0 | 0 |
| *NAIL AND NAIL BED CONDITIONS (EXCL INFECTIONS AND INFESTATIONS)* | | | | | | | | | | |
| NAIL DISCOLOURATION | 0 | 0 | 0 | 0 | 0 | 0 | 0 | 0 | 0 | 0 |
| ONYCHOCLASIS | 0 | 0 | 0 | 0 | 0 | 0 | 0 | 0 | 0 | 0 |
| *PILAR DISORDERS NEC* | | | | | | | | | | |
| PILOERECTION | 0 | 0 | 0 | 0 | 0 | 0 | 0 | 0 | 0 | 0 |
| ***SKIN VASCULAR ABNORMALITIES*** | | | | | | | | | | |
| *PURPURA AND RELATED CONDITIONS* | | | | | | | | | | |
| PETECHIAE | 0 | 0 | 0 | 0 | 0 | 0 | 0 | 0 | 0 | 0 |
| PURPURA | 0 | 0 | 0 | 0 | 0 | 0 | 0 | 0 | 0 | 0 |
| *SKIN HAEMORRHAGES* | | | | | | | | | | |
| SKIN HAEMORRHAGE | 0 | 0 | 0 | 0 | 0 | 0 | 0 | 0 | 0 | 0 |
| **SOCIAL CIRCUMSTANCES** | | | | | | | | | | |
| ***ECONOMIC AND HOUSING ISSUES*** | | | | | | | | | | |
| *EMPLOYMENT ISSUES* | | | | | | | | | | |
| RETIREMENT | 1 | 1 | 0 | 0 | 0 | 1 | 1 | 0 | 0 | 0 |
| ***ENVIRONMENTAL ISSUES*** | | | | | | | | | | |
| *NON-OCCUPATIONAL AND UNSPECIFIED ENVIRONMENTAL PROBLEMS* | | | | | | | | | | |
| POLLUTION | 0 | 0 | 0 | 0 | 0 | 0 | 0 | 0 | 0 | 0 |
| **SURGICAL AND MEDICAL PROCEDURES** | | | | | | | | | | |
| ***BONE AND JOINT THERAPEUTIC PROCEDURES*** | | | | | | | | | | |
| *JOINT THERAPEUTIC PROCEDURES* | | | | | | | | | | |
| KNEE OPERATION | 0 | 0 | 0 | 0 | 0 | 0 | 0 | 0 | 0 | 0 |
| ***BREAST THERAPEUTIC PROCEDURES*** | | | | | | | | | | |
| *MASTECTOMIES* | | | | | | | | | | |
| BREAST CONSERVING SURGERY | 0 | 0 | 0 | 0 | 0 | 0 | 0 | 0 | 0 | 0 |
| ***GASTROINTESTINAL THERAPEUTIC PROCEDURES*** | | | | | | | | | | |
| *GASTROINTESTINAL THERAPEUTIC PROCEDURES NEC* | | | | | | | | | | |
| PROPHYLAXIS OF NAUSEA AND VOMITING | 0 | 0 | 0 | 0 | 0 | 0 | 0 | 0 | 0 | 0 |
| ***HEAD AND NECK THERAPEUTIC PROCEDURES*** | | | | | | | | | | |
| *DENTAL AND GINGIVAL THERAPEUTIC PROCEDURES* | | | | | | | | | | |

| | | | | | | | | | | |
|---|---|---|---|---|---|---|---|---|---|---|
| DENTAL CARE | 0 | 0 | 0 | 0 | 0 | 0 | 0 | 0 | 0 | 0 |
| ***NERVOUS SYSTEM, SKULL AND SPINE THERAPEUTIC PROCEDURES*** | | | | | | | | | | |
| *SKULL AND BRAIN THERAPEUTIC PROCEDURES* | | | | | | | | | | |
| BRAIN TUMOUR OPERATION | 0 | 0 | 0 | 0 | 0 | 0 | 0 | 0 | 0 | 0 |
| ***OBSTETRIC AND GYNAECOLOGICAL THERAPEUTIC PROCEDURES*** | | | | | | | | | | |
| *FERTILITY AND FERTILISATION INTERVENTIONS FEMALE* | | | | | | | | | | |
| ENDOMETRIAL SCRATCHING | 1 | 1 | 0 | 0 | 0 | 1 | 1 | 0 | 0 | 0 |
| ***RESPIRATORY TRACT THERAPEUTIC PROCEDURES*** | | | | | | | | | | |
| *RESPIRATORY TRACT THERAPEUTIC PROCEDURES NEC* | | | | | | | | | | |
| OXYGEN THERAPY | 0 | 0 | 0 | 0 | 0 | 0 | 0 | 0 | 0 | 0 |
| ***THERAPEUTIC PROCEDURES AND SUPPORTIVE CARE NEC*** | | | | | | | | | | |
| *ANAESTHESIA AND ALLIED PROCEDURES* | | | | | | | | | | |
| NERVE BLOCK | 0 | 0 | 0 | 0 | 0 | 0 | 0 | 0 | 0 | 0 |
| *DIETARY AND NUTRITIONAL THERAPIES* | | | | | | | | | | |
| NOTHING BY MOUTH ORDER | 0 | 0 | 0 | 0 | 0 | 0 | 0 | 0 | 0 | 0 |
| *IMMUNISATIONS* | | | | | | | | | | |
| COVID-19 IMMUNISATION | 0 | 0 | 0 | 0 | 0 | 0 | 0 | 0 | 0 | 0 |
| *THERAPEUTIC PROCEDURES NEC* | | | | | | | | | | |
| INJECTION | 1 | 1 | 0 | 0 | 0 | 1 | 1 | 0 | 0 | 0 |
| LOCALISED ALTERNATING HOT AND COLD THERAPY | 0 | 0 | 0 | 0 | 0 | 0 | 0 | 0 | 0 | 0 |
| MASS EXCISION | 0 | 0 | 0 | 0 | 0 | 0 | 0 | 0 | 0 | 0 |
| REINFUSION | 0 | 0 | 0 | 0 | 0 | 0 | 0 | 0 | 0 | 0 |
| **VASCULAR DISORDERS** | | | | | | | | | | |
| ***ARTERIOSCLEROSIS, STENOSIS, VASCULAR INSUFFICIENCY AND NECROSIS*** | | | | | | | | | | |
| *NON-SITE SPECIFIC NECROSIS AND VASCULAR INSUFFICIENCY NEC* | | | | | | | | | | |
| VASOSPASM | 0 | 0 | 0 | 0 | 0 | 0 | 0 | 0 | 0 | 0 |
| *PERIPHERAL VASOCONSTRICTION, NECROSIS AND VASCULAR INSUFFICIENCY* | | | | | | | | | | |
| PERIPHERAL COLDNESS | 4 | 3 | 1 | 0 | 0 | 4 | 3 | 1 | 0 | 0 |
| RAYNAUD'S PHENOMENON | 0 | 0 | 0 | 0 | 0 | 0 | 0 | 0 | 0 | 0 |
| ***DECREASED AND NONSPECIFIC BLOOD PRESSURE DISORDERS AND SHOCK*** | | | | | | | | | | |
| *BLOOD PRESSURE DISORDERS NEC* | | | | | | | | | | |
| BLOOD PRESSURE FLUCTUATION | 0 | 0 | 0 | 0 | 0 | 0 | 0 | 0 | 0 | 0 |
| *VASCULAR HYPOTENSIVE DISORDERS* | | | | | | | | | | |
| CAPILLARY LEAK SYNDROME | 0 | 0 | 0 | 0 | 0 | 0 | 0 | 0 | 0 | 0 |
| HYPOTENSION | 4 | 4 | 0 | 0 | 0 | 4 | 4 | 0 | 0 | 0 |

| | | | | | | | | | | |
|---|---|---|---|---|---|---|---|---|---|---|
| ORTHOSTATIC HYPOTENSION | 1 | 0 | 1 | 0 | 0 | 1 | 0 | 1 | 0 | 0 |
| ***EMBOLISM AND THROMBOSIS*** | | | | | | | | | | |
| *NON-SITE SPECIFIC EMBOLISM AND THROMBOSIS* | | | | | | | | | | |
| EMBOLISM | 0 | 0 | 0 | 0 | 0 | 0 | 0 | 0 | 0 | 0 |
| THROMBOSIS | 3 | 3 | 0 | 0 | 0 | 3 | 3 | 0 | 0 | 0 |
| VENOUS THROMBOSIS | 0 | 0 | 0 | 0 | 0 | 0 | 0 | 0 | 0 | 0 |
| *PERIPHERAL EMBOLISM AND THROMBOSIS* | | | | | | | | | | |
| BLUE TOE SYNDROME | 0 | 0 | 0 | 0 | 0 | 0 | 0 | 0 | 0 | 0 |
| DEEP VEIN THROMBOSIS | 0 | 0 | 0 | 0 | 0 | 0 | 0 | 0 | 0 | 0 |
| SUPERFICIAL VEIN THROMBOSIS | 0 | 0 | 0 | 0 | 0 | 0 | 0 | 0 | 0 | 0 |
| ***LYMPHATIC VESSEL DISORDERS*** | | | | | | | | | | |
| *LYMPHOEDEMAS* | | | | | | | | | | |
| LYMPHOEDEMA | 4 | 2 | 0 | 2 | 0 | 3 | 2 | 0 | 1 | 0 |
| ***VASCULAR DISORDERS NEC*** | | | | | | | | | | |
| *NON-SITE SPECIFIC VASCULAR DISORDERS NEC* | | | | | | | | | | |
| VASCULAR PAIN | 1 | 0 | 1 | 0 | 0 | 1 | 0 | 1 | 0 | 0 |
| VEIN DISCOLOURATION | 0 | 0 | 0 | 0 | 0 | 0 | 0 | 0 | 0 | 0 |
| VEIN RUPTURE | 0 | 0 | 0 | 0 | 0 | 0 | 0 | 0 | 0 | 0 |
| *PERIPHERAL VASCULAR DISORDERS NEC* | | | | | | | | | | |
| FLUSHING | 7 | 5 | 1 | 1 | 0 | 6 | 5 | 0 | 1 | 0 |
| HOT FLUSH | 27 | 19 | 6 | 2 | 0 | 26 | 19 | 5 | 2 | 0 |
| *SITE SPECIFIC VASCULAR DISORDERS NEC* | | | | | | | | | | |
| PALLOR | 2 | 2 | 0 | 0 | 0 | 2 | 2 | 0 | 0 | 0 |
| ***VASCULAR HAEMORRHAGIC DISORDERS*** | | | | | | | | | | |
| *HAEMORRHAGES NEC* | | | | | | | | | | |
| HAEMATOMA | 0 | 0 | 0 | 0 | 0 | 0 | 0 | 0 | 0 | 0 |
| HAEMORRHAGE | 4 | 2 | 0 | 2 | 0 | 4 | 2 | 0 | 2 | 0 |
| ***VASCULAR HYPERTENSIVE DISORDERS*** | | | | | | | | | | |
| *ACCELERATED AND MALIGNANT HYPERTENSION* | | | | | | | | | | |
| HYPERTENSIVE URGENCY | 1 | 1 | 0 | 0 | 0 | 1 | 1 | 0 | 0 | 0 |
| *VASCULAR HYPERTENSIVE DISORDERS NEC* | | | | | | | | | | |
| HYPERTENSION | 6 | 5 | 0 | 1 | 0 | 6 | 5 | 0 | 1 | 0 |
| SYSTOLIC HYPERTENSION | 0 | 0 | 0 | 0 | 0 | 0 | 0 | 0 | 0 | 0 |
| ***VASCULAR INFECTIONS AND INFLAMMATIONS*** | | | | | | | | | | |
| *ARTERIAL INFECTIONS AND INFLAMMATIONS* | | | | | | | | | | |
| GIANT CELL ARTERITIS | 0 | 0 | 0 | 0 | 0 | 0 | 0 | 0 | 0 | 0 |

| | | | | | | | | | | |
|---|---|---|---|---|---|---|---|---|---|---|
| *PHLEBITIS NEC* | | | | | | | | | | |
| PHLEBITIS | 1 | 0 | 0 | 1 | 0 | 0 | 0 | 0 | 0 | 0 |
| *VASCULITIDES NEC* | | | | | | | | | | |
| VASCULITIS | 0 | 0 | 0 | 0 | 0 | 0 | 0 | 0 | 0 | 0 |
| ***VENOUS VARICES*** | | | | | | | | | | |
| *VARICOSE VEINS NEC* | | | | | | | | | | |
| SPIDER VEIN | 1 | 1 | 0 | 0 | 0 | 1 | 1 | 0 | 0 | 0 |
| VARICOSE VEIN | 1 | 1 | 0 | 0 | 0 | 1 | 1 | 0 | 0 | 0 |
| **TOTAL ADR EVENTS** | ***9949*** | ***5705*** | ***1954*** | ***2141*** | ***149*** | ***9139*** | ***5705*** | ***1576*** | ***1735*** | ***123*** |

**TABLE S1-3. AstraZeneca COVID-19 vaccine: ADR listing for events reported in the YCVM in (a) those reporting any vaccination dose and (b) in those who had reported a 1st dose vaccination and any subsequent doses.**

| | Patients with any vaccination dose: ADR Counts | | | | | Patients reporting a 1st dose: ADR Counts | | | | |
|---|---|---|---|---|---|---|---|---|---|---|
| **REACTION TERM (SOC, *HLGT*, *HLT*, PT)** | All doses | 1st dose | 2nd dose | 3rd dose | Other doses | All doses | 1st dose | 2nd dose | 3rd dose | Other doses |
| ***(freetext)*** | 35 | 33 | 2 | 0 | 0 | 34 | 33 | 1 | 0 | 0 |
| **BLOOD AND LYMPHATIC SYSTEM DISORDERS** | | | | | | | | | | |
| ***COAGULOPATHIES AND BLEEDING DIATHESES (EXCL THROMBOCYTOPENIC)*** | | | | | | | | | | |
| *BLEEDING TENDENCIES* | | | | | | | | | | |
| INCREASED TENDENCY TO BRUISE | 1 | 0 | 1 | 0 | 0 | 1 | 0 | 1 | 0 | 0 |
| ***HAEMOGLOBINOPATHIES*** | | | | | | | | | | |
| *SICKLE CELL TRAIT AND DISORDERS* | | | | | | | | | | |
| SICKLE CELL ANAEMIA WITH CRISIS | 1 | 1 | 0 | 0 | 0 | 1 | 1 | 0 | 0 | 0 |
| ***PLATELET DISORDERS*** | | | | | | | | | | |
| *THROMBOCYTOPENIAS* | | | | | | | | | | |
| IMMUNE THROMBOCYTOPENIA | 1 | 0 | 1 | 0 | 0 | 0 | 0 | 0 | 0 | 0 |
| ***SPLEEN, LYMPHATIC AND RETICULOENDOTHELIAL SYSTEM DISORDERS*** | | | | | | | | | | |
| *LYMPHATIC SYSTEM DISORDERS NEC* | | | | | | | | | | |
| LYMPH NODE PAIN | 14 | 9 | 5 | 0 | 0 | 14 | 9 | 5 | 0 | 0 |
| LYMPHADENITIS | 0 | 0 | 0 | 0 | 0 | 0 | 0 | 0 | 0 | 0 |
| LYMPHADENOPATHY | 108 | 79 | 29 | 0 | 0 | 98 | 79 | 19 | 0 | 0 |
| **CARDIAC DISORDERS** | | | | | | | | | | |
| ***CARDIAC ARRHYTHMIAS*** | | | | | | | | | | |
| *RATE AND RHYTHM DISORDERS NEC* | | | | | | | | | | |
| ARRHYTHMIA | 1 | 1 | 0 | 0 | 0 | 1 | 1 | 0 | 0 | 0 |
| CARDIAC FLUTTER | 4 | 2 | 2 | 0 | 0 | 4 | 2 | 2 | 0 | 0 |
| EXTRASYSTOLES | 0 | 0 | 0 | 0 | 0 | 0 | 0 | 0 | 0 | 0 |
| TACHYCARDIA | 15 | 13 | 2 | 0 | 0 | 13 | 13 | 0 | 0 | 0 |
| *SUPRAVENTRICULAR ARRHYTHMIAS* | | | | | | | | | | |
| ATRIAL FIBRILLATION | 3 | 2 | 1 | 0 | 0 | 2 | 2 | 0 | 0 | 0 |
| SUPRAVENTRICULAR TACHYCARDIA | 1 | 1 | 0 | 0 | 0 | 1 | 1 | 0 | 0 | 0 |

| | | | | | | | | | | |
|---|---|---|---|---|---|---|---|---|---|---|
| ***CARDIAC DISORDERS, SIGNS AND SYMPTOMS NEC*** | | | | | | | | | | |
| *CARDIAC DISORDERS NEC* | | | | | | | | | | |
| CARDIOVASCULAR DISORDER | 1 | 1 | 0 | 0 | 0 | 1 | 1 | 0 | 0 | 0 |
| *CARDIAC SIGNS AND SYMPTOMS NEC* | | | | | | | | | | |
| PALPITATIONS | 58 | 50 | 7 | 1 | 0 | 57 | 50 | 6 | 1 | 0 |
| ***CORONARY ARTERY DISORDERS*** | | | | | | | | | | |
| *ISCHAEMIC CORONARY ARTERY DISORDERS* | | | | | | | | | | |
| ANGINA PECTORIS | 1 | 1 | 0 | 0 | 0 | 1 | 1 | 0 | 0 | 0 |
| MYOCARDIAL INFARCTION | 1 | 1 | 0 | 0 | 0 | 1 | 1 | 0 | 0 | 0 |
| ***HEART FAILURES*** | | | | | | | | | | |
| *HEART FAILURES NEC* | | | | | | | | | | |
| CARDIAC FAILURE | 0 | 0 | 0 | 0 | 0 | 0 | 0 | 0 | 0 | 0 |
| ***MYOCARDIAL DISORDERS*** | | | | | | | | | | |
| *NONINFECTIOUS MYOCARDITIS* | | | | | | | | | | |
| MYOCARDITIS | 0 | 0 | 0 | 0 | 0 | 0 | 0 | 0 | 0 | 0 |
| ***PERICARDIAL DISORDERS*** | | | | | | | | | | |
| *NONINFECTIOUS PERICARDITIS* | | | | | | | | | | |
| PERICARDITIS | 0 | 0 | 0 | 0 | 0 | 0 | 0 | 0 | 0 | 0 |
| **CONGENITAL, FAMILIAL AND GENETIC DISORDERS** | | | | | | | | | | |
| ***CARDIAC AND VASCULAR DISORDERS CONGENITAL*** | | | | | | | | | | |
| *CARDIAC DISORDERS CONGENITAL NEC* | | | | | | | | | | |
| HEART DISEASE CONGENITAL | 0 | 0 | 0 | 0 | 0 | 0 | 0 | 0 | 0 | 0 |
| ***METABOLIC AND NUTRITIONAL DISORDERS CONGENITAL*** | | | | | | | | | | |
| *INBORN ERRORS OF AMINO ACID METABOLISM* | | | | | | | | | | |
| HYPERGLYCINAEMIA | 1 | 1 | 0 | 0 | 0 | 1 | 1 | 0 | 0 | 0 |
| *INBORN ERRORS OF STEROID SYNTHESIS* | | | | | | | | | | |
| 11-BETA-HYDROXYLASE DEFICIENCY | 0 | 0 | 0 | 0 | 0 | 0 | 0 | 0 | 0 | 0 |
| ***NEUROLOGICAL DISORDERS CONGENITAL*** | | | | | | | | | | |
| *PERIPHERAL NERVOUS SYSTEM DISORDERS CONGENITAL NEC* | | | | | | | | | | |
| PAROXYSMAL EXTREME PAIN DISORDER | 1 | 0 | 1 | 0 | 0 | 1 | 0 | 1 | 0 | 0 |
| **EAR AND LABYRINTH DISORDERS** | | | | | | | | | | |

| | | | | | | | | | | |
|---|---|---|---|---|---|---|---|---|---|---|
| ***AURAL DISORDERS NEC*** | | | | | | | | | | |
| *EAR DISORDERS NEC* | | | | | | | | | | |
| EAR DISCOMFORT | 3 | 2 | 1 | 0 | 0 | 3 | 2 | 1 | 0 | 0 |
| EAR DISORDER | 1 | 0 | 1 | 0 | 0 | 1 | 0 | 1 | 0 | 0 |
| EAR PAIN | 36 | 28 | 8 | 0 | 0 | 32 | 28 | 4 | 0 | 0 |
| EAR SWELLING | 1 | 1 | 0 | 0 | 0 | 1 | 1 | 0 | 0 | 0 |
| ***EXTERNAL EAR DISORDERS (EXCL CONGENITAL)*** | | | | | | | | | | |
| *EXTERNAL EAR DISORDERS NEC* | | | | | | | | | | |
| EXCESSIVE CERUMEN PRODUCTION | 2 | 2 | 0 | 0 | 0 | 2 | 2 | 0 | 0 | 0 |
| ***HEARING DISORDERS*** | | | | | | | | | | |
| *HEARING LOSSES* | | | | | | | | | | |
| DEAFNESS | 1 | 0 | 1 | 0 | 0 | 0 | 0 | 0 | 0 | 0 |
| DEAFNESS NEUROSENSORY | 0 | 0 | 0 | 0 | 0 | 0 | 0 | 0 | 0 | 0 |
| DEAFNESS PERMANENT | 0 | 0 | 0 | 0 | 0 | 0 | 0 | 0 | 0 | 0 |
| DEAFNESS UNILATERAL | 2 | 1 | 1 | 0 | 0 | 2 | 1 | 1 | 0 | 0 |
| HYPOACUSIS | 2 | 1 | 1 | 0 | 0 | 2 | 1 | 1 | 0 | 0 |
| SUDDEN HEARING LOSS | 1 | 1 | 0 | 0 | 0 | 1 | 1 | 0 | 0 | 0 |
| *HYPERACUSIA* | | | | | | | | | | |
| HYPERACUSIS | 1 | 0 | 1 | 0 | 0 | 1 | 0 | 1 | 0 | 0 |
| ***INNER EAR AND VIIITH CRANIAL NERVE DISORDERS*** | | | | | | | | | | |
| *INNER EAR SIGNS AND SYMPTOMS* | | | | | | | | | | |
| MOTION SICKNESS | 1 | 1 | 0 | 0 | 0 | 1 | 1 | 0 | 0 | 0 |
| TINNITUS | 47 | 37 | 10 | 0 | 0 | 44 | 37 | 7 | 0 | 0 |
| VERTIGO | 31 | 24 | 7 | 0 | 0 | 30 | 24 | 6 | 0 | 0 |
| VERTIGO LABYRINTHINE | 2 | 2 | 0 | 0 | 0 | 2 | 2 | 0 | 0 | 0 |
| VERTIGO POSITIONAL | 4 | 3 | 1 | 0 | 0 | 4 | 3 | 1 | 0 | 0 |
| **ENDOCRINE DISORDERS** | | | | | | | | | | |
| ***THYROID GLAND DISORDERS*** | | | | | | | | | | |
| *THYROID HYPERFUNCTION DISORDERS* | | | | | | | | | | |
| HYPERTHYROIDISM | 1 | 1 | 0 | 0 | 0 | 1 | 1 | 0 | 0 | 0 |
| **EYE DISORDERS** | | | | | | | | | | |
| ***EYE DISORDERS NEC*** | | | | | | | | | | |
| *LACRIMATION DISORDERS* | | | | | | | | | | |
| DRY EYE | 3 | 3 | 0 | 0 | 0 | 3 | 3 | 0 | 0 | 0 |
| LACRIMATION INCREASED | 0 | 0 | 0 | 0 | 0 | 0 | 0 | 0 | 0 | 0 |
| *OCULAR DISORDERS NEC* | | | | | | | | | | |

| | | | | | | | | | | |
|---|---|---|---|---|---|---|---|---|---|---|
| EYE OEDEMA | 1 | 1 | 0 | 0 | 0 | 1 | 1 | 0 | 0 | 0 |
| EYE PAIN | 61 | 57 | 4 | 0 | 0 | 60 | 57 | 3 | 0 | 0 |
| EYE SWELLING | 1 | 1 | 0 | 0 | 0 | 1 | 1 | 0 | 0 | 0 |
| EYE ULCER | 1 | 1 | 0 | 0 | 0 | 1 | 1 | 0 | 0 | 0 |
| EYELID PAIN | 1 | 1 | 0 | 0 | 0 | 1 | 1 | 0 | 0 | 0 |
| OCULAR DISCOMFORT | 2 | 2 | 0 | 0 | 0 | 2 | 2 | 0 | 0 | 0 |
| PERIORBITAL DISCOMFORT | 1 | 0 | 1 | 0 | 0 | 0 | 0 | 0 | 0 | 0 |
| PERIORBITAL SWELLING | 1 | 0 | 1 | 0 | 0 | 1 | 0 | 1 | 0 | 0 |
| ***OCULAR HAEMORRHAGES AND VASCULAR DISORDERS NEC*** | | | | | | | | | | |
| *CONJUNCTIVAL AND CORNEAL BLEEDING AND VASCULAR DISORDERS* | | | | | | | | | | |
| CONJUNCTIVAL HAEMORRHAGE | 0 | 0 | 0 | 0 | 0 | 0 | 0 | 0 | 0 | 0 |
| *LID BLEEDING AND VASCULAR DISORDERS* | | | | | | | | | | |
| EYELID BLEEDING | 1 | 1 | 0 | 0 | 0 | 1 | 1 | 0 | 0 | 0 |
| ***OCULAR INFECTIONS, IRRITATIONS AND INFLAMMATIONS*** | | | | | | | | | | |
| *LID, LASH AND LACRIMAL INFECTIONS, IRRITATIONS AND INFLAMMATIONS* | | | | | | | | | | |
| BLEPHARITIS | 1 | 1 | 0 | 0 | 0 | 1 | 1 | 0 | 0 | 0 |
| ERYTHEMA OF EYELID | 1 | 1 | 0 | 0 | 0 | 1 | 1 | 0 | 0 | 0 |
| EYELID IRRITATION | 1 | 1 | 0 | 0 | 0 | 1 | 1 | 0 | 0 | 0 |
| *OCULAR INFECTIONS, INFLAMMATIONS AND ASSOCIATED MANIFESTATIONS* | | | | | | | | | | |
| EYE DISCHARGE | 1 | 1 | 0 | 0 | 0 | 1 | 1 | 0 | 0 | 0 |
| EYE IRRITATION | 0 | 0 | 0 | 0 | 0 | 0 | 0 | 0 | 0 | 0 |
| EYE PRURITUS | 4 | 4 | 0 | 0 | 0 | 4 | 4 | 0 | 0 | 0 |
| LIMBAL SWELLING | 0 | 0 | 0 | 0 | 0 | 0 | 0 | 0 | 0 | 0 |
| OCULAR HYPERAEMIA | 5 | 5 | 0 | 0 | 0 | 5 | 5 | 0 | 0 | 0 |
| ***OCULAR NEUROMUSCULAR DISORDERS*** | | | | | | | | | | |
| *EYELID MOVEMENT DISORDERS* | | | | | | | | | | |
| BLEPHAROSPASM | 1 | 1 | 0 | 0 | 0 | 1 | 1 | 0 | 0 | 0 |
| ***OCULAR SENSORY SYMPTOMS NEC*** | | | | | | | | | | |
| *OCULAR SENSATION DISORDERS* | | | | | | | | | | |
| ABNORMAL SENSATION IN EYE | 0 | 0 | 0 | 0 | 0 | 0 | 0 | 0 | 0 | 0 |
| ASTHENOPIA | 11 | 10 | 1 | 0 | 0 | 11 | 10 | 1 | 0 | 0 |
| FOREIGN BODY SENSATION IN EYES | 1 | 1 | 0 | 0 | 0 | 1 | 1 | 0 | 0 | 0 |
| PHOTOPHOBIA | 16 | 13 | 3 | 0 | 0 | 15 | 13 | 2 | 0 | 0 |

| | | | | | | | | | | |
|---|---|---|---|---|---|---|---|---|---|---|
| ***OCULAR STRUCTURAL CHANGE, DEPOSIT AND DEGENERATION NEC*** | | | | | | | | | | |
| *CHOROID AND VITREOUS STRUCTURAL CHANGE, DEPOSIT AND DEGENERATION* | | | | | | | | | | |
| VITREOUS DETACHMENT | 1 | 1 | 0 | 0 | 0 | 1 | 1 | 0 | 0 | 0 |
| VITREOUS FLOATERS | 2 | 2 | 0 | 0 | 0 | 2 | 2 | 0 | 0 | 0 |
| ***RETINA, CHOROID AND VITREOUS HAEMORRHAGES AND VASCULAR DISORDERS*** | | | | | | | | | | |
| *RETINAL BLEEDING AND VASCULAR DISORDERS (EXCL RETINOPATHY)* | | | | | | | | | | |
| RETINAL VEIN OCCLUSION | 1 | 0 | 1 | 0 | 0 | 1 | 0 | 1 | 0 | 0 |
| *RETINOPATHIES NEC* | | | | | | | | | | |
| RETINAL EXUDATES | 1 | 1 | 0 | 0 | 0 | 1 | 1 | 0 | 0 | 0 |
| ***VISION DISORDERS*** | | | | | | | | | | |
| *VISUAL DISORDERS NEC* | | | | | | | | | | |
| DIPLOPIA | 1 | 1 | 0 | 0 | 0 | 1 | 1 | 0 | 0 | 0 |
| HALO VISION | 2 | 1 | 1 | 0 | 0 | 2 | 1 | 1 | 0 | 0 |
| METAMORPHOPSIA | 1 | 1 | 0 | 0 | 0 | 1 | 1 | 0 | 0 | 0 |
| PHOTOPSIA | 1 | 1 | 0 | 0 | 0 | 1 | 1 | 0 | 0 | 0 |
| VISION BLURRED | 25 | 20 | 5 | 0 | 0 | 24 | 20 | 4 | 0 | 0 |
| *VISUAL IMPAIRMENT AND BLINDNESS (EXCL COLOUR BLINDNESS)* | | | | | | | | | | |
| BLINDNESS | 1 | 0 | 1 | 0 | 0 | 1 | 0 | 1 | 0 | 0 |
| BLINDNESS TRANSIENT | 1 | 1 | 0 | 0 | 0 | 1 | 1 | 0 | 0 | 0 |
| SUDDEN VISUAL LOSS | 1 | 1 | 0 | 0 | 0 | 1 | 1 | 0 | 0 | 0 |
| VISUAL IMPAIRMENT | 10 | 10 | 0 | 0 | 0 | 10 | 10 | 0 | 0 | 0 |
| **GASTROINTESTINAL DISORDERS** | | | | | | | | | | |
| ***ANAL AND RECTAL CONDITIONS NEC*** | | | | | | | | | | |
| *ANAL AND RECTAL SIGNS AND SYMPTOMS* | | | | | | | | | | |
| ANAL PARAESTHESIA | 1 | 1 | 0 | 0 | 0 | 1 | 1 | 0 | 0 | 0 |
| ***BENIGN NEOPLASMS GASTROINTESTINAL*** | | | | | | | | | | |
| *BENIGN ORAL CAVITY NEOPLASMS* | | | | | | | | | | |
| MOUTH CYST | 1 | 1 | 0 | 0 | 0 | 1 | 1 | 0 | 0 | 0 |
| ***DENTAL AND GINGIVAL CONDITIONS*** | | | | | | | | | | |
| *DENTAL DISORDERS NEC* | | | | | | | | | | |
| TEETHING | 0 | 0 | 0 | 0 | 0 | 0 | 0 | 0 | 0 | 0 |

| | | | | | | | | | | |
|---|---|---|---|---|---|---|---|---|---|---|
| *DENTAL PAIN AND SENSATION DISORDERS* | | | | | | | | | | |
| DENTAL PARAESTHESIA | 1 | 1 | 0 | 0 | 0 | 1 | 1 | 0 | 0 | 0 |
| TOOTHACHE | 6 | 4 | 2 | 0 | 0 | 6 | 4 | 2 | 0 | 0 |
| *GINGIVAL DISORDERS, SIGNS AND SYMPTOMS NEC* | | | | | | | | | | |
| GINGIVAL BLISTER | 1 | 1 | 0 | 0 | 0 | 1 | 1 | 0 | 0 | 0 |
| GINGIVAL PAIN | 6 | 4 | 2 | 0 | 0 | 6 | 4 | 2 | 0 | 0 |
| GINGIVAL SWELLING | 1 | 1 | 0 | 0 | 0 | 1 | 1 | 0 | 0 | 0 |
| ***GASTROINTESTINAL CONDITIONS NEC*** | | | | | | | | | | |
| *GASTROINTESTINAL MUCOSAL DYSTROPHIES AND SECRETION DISORDERS* | | | | | | | | | | |
| BARRETT'S OESOPHAGUS | 1 | 1 | 0 | 0 | 0 | 1 | 1 | 0 | 0 | 0 |
| ***GASTROINTESTINAL HAEMORRHAGES NEC*** | | | | | | | | | | |
| *INTESTINAL HAEMORRHAGES* | | | | | | | | | | |
| RECTAL HAEMORRHAGE | 1 | 1 | 0 | 0 | 0 | 1 | 1 | 0 | 0 | 0 |
| *NON-SITE SPECIFIC GASTROINTESTINAL HAEMORRHAGES* | | | | | | | | | | |
| HAEMATEMESIS | 1 | 1 | 0 | 0 | 0 | 1 | 1 | 0 | 0 | 0 |
| ***GASTROINTESTINAL INFLAMMATORY CONDITIONS*** | | | | | | | | | | |
| *COLITIS (EXCL INFECTIVE)* | | | | | | | | | | |
| COLITIS | 0 | 0 | 0 | 0 | 0 | 0 | 0 | 0 | 0 | 0 |
| COLITIS MICROSCOPIC | 1 | 0 | 1 | 0 | 0 | 1 | 0 | 1 | 0 | 0 |
| COLITIS ULCERATIVE | 1 | 1 | 0 | 0 | 0 | 1 | 1 | 0 | 0 | 0 |
| *GASTRITIS (EXCL INFECTIVE)* | | | | | | | | | | |
| GASTRITIS | 2 | 2 | 0 | 0 | 0 | 2 | 2 | 0 | 0 | 0 |
| REFLUX GASTRITIS | 1 | 1 | 0 | 0 | 0 | 1 | 1 | 0 | 0 | 0 |
| *GASTROINTESTINAL INFLAMMATORY DISORDERS NEC* | | | | | | | | | | |
| GASTROINTESTINAL TRACT IRRITATION | 0 | 0 | 0 | 0 | 0 | 0 | 0 | 0 | 0 | 0 |
| ***GASTROINTESTINAL MOTILITY AND DEFAECATION CONDITIONS*** | | | | | | | | | | |
| *DIARRHOEA (EXCL INFECTIVE)* | | | | | | | | | | |
| DIARRHOEA | 201 | 173 | 28 | 0 | 0 | 191 | 173 | 18 | 0 | 0 |
| *GASTROINTESTINAL ATONIC AND HYPOMOTILITY DISORDERS NEC* | | | | | | | | | | |
| CONSTIPATION | 4 | 2 | 2 | 0 | 0 | 3 | 2 | 1 | 0 | 0 |
| GASTROOESOPHAGEAL REFLUX DISEASE | 2 | 1 | 1 | 0 | 0 | 2 | 1 | 1 | 0 | 0 |
| *GASTROINTESTINAL DYSKINETIC DISORDERS* | | | | | | | | | | |
| CHANGE OF BOWEL HABIT | 0 | 0 | 0 | 0 | 0 | 0 | 0 | 0 | 0 | 0 |
| *GASTROINTESTINAL SPASTIC AND HYPERMOTILITY DISORDERS* | | | | | | | | | | |
| IRRITABLE BOWEL SYNDROME | 2 | 1 | 1 | 0 | 0 | 2 | 1 | 1 | 0 | 0 |
| ***GASTROINTESTINAL SIGNS AND SYMPTOMS*** | | | | | | | | | | |

| | | | | | | | | | | |
|---|---|---|---|---|---|---|---|---|---|---|
| *DYSPEPTIC SIGNS AND SYMPTOMS* | | | | | | | | | | |
| DYSPEPSIA | 26 | 21 | 5 | 0 | 0 | 24 | 21 | 3 | 0 | 0 |
| ERUCTATION | 1 | 1 | 0 | 0 | 0 | 1 | 1 | 0 | 0 | 0 |
| *FAECAL ABNORMALITIES NEC* | | | | | | | | | | |
| FAECALOMA | 0 | 0 | 0 | 0 | 0 | 0 | 0 | 0 | 0 | 0 |
| FAECES DISCOLOURED | 0 | 0 | 0 | 0 | 0 | 0 | 0 | 0 | 0 | 0 |
| FAECES SOFT | 2 | 1 | 1 | 0 | 0 | 2 | 1 | 1 | 0 | 0 |
| *FLATULENCE, BLOATING AND DISTENSION* | | | | | | | | | | |
| ABDOMINAL DISTENSION | 9 | 9 | 0 | 0 | 0 | 9 | 9 | 0 | 0 | 0 |
| FLATULENCE | 14 | 13 | 1 | 0 | 0 | 14 | 13 | 1 | 0 | 0 |
| *GASTROINTESTINAL AND ABDOMINAL PAINS (EXCL ORAL AND THROAT)* | | | | | | | | | | |
| ABDOMINAL PAIN | 61 | 50 | 11 | 0 | 0 | 57 | 50 | 7 | 0 | 0 |
| ABDOMINAL PAIN LOWER | 4 | 3 | 1 | 0 | 0 | 3 | 3 | 0 | 0 | 0 |
| ABDOMINAL PAIN UPPER | 91 | 76 | 15 | 0 | 0 | 86 | 76 | 10 | 0 | 0 |
| GASTROINTESTINAL PAIN | 7 | 7 | 0 | 0 | 0 | 7 | 7 | 0 | 0 | 0 |
| *GASTROINTESTINAL SIGNS AND SYMPTOMS NEC* | | | | | | | | | | |
| ABDOMINAL DISCOMFORT | 57 | 45 | 12 | 0 | 0 | 54 | 45 | 9 | 0 | 0 |
| ABDOMINAL SYMPTOM | 1 | 1 | 0 | 0 | 0 | 1 | 1 | 0 | 0 | 0 |
| ACUTE ABDOMEN | 1 | 1 | 0 | 0 | 0 | 1 | 1 | 0 | 0 | 0 |
| ANAL INCONTINENCE | 0 | 0 | 0 | 0 | 0 | 0 | 0 | 0 | 0 | 0 |
| BREATH ODOUR | 0 | 0 | 0 | 0 | 0 | 0 | 0 | 0 | 0 | 0 |
| DYSPHAGIA | 0 | 0 | 0 | 0 | 0 | 0 | 0 | 0 | 0 | 0 |
| ODYNOPHAGIA | 1 | 1 | 0 | 0 | 0 | 1 | 1 | 0 | 0 | 0 |
| *NAUSEA AND VOMITING SYMPTOMS* | | | | | | | | | | |
| NAUSEA | 897 | 789 | 108 | 0 | 0 | 860 | 789 | 71 | 0 | 0 |
| RETCHING | 3 | 3 | 0 | 0 | 0 | 3 | 3 | 0 | 0 | 0 |
| VOMITING | 102 | 93 | 9 | 0 | 0 | 101 | 93 | 8 | 0 | 0 |
| VOMITING PROJECTILE | 2 | 2 | 0 | 0 | 0 | 2 | 2 | 0 | 0 | 0 |
| ***MALABSORPTION CONDITIONS*** | | | | | | | | | | |
| *MALABSORPTION SYNDROMES* | | | | | | | | | | |
| COELIAC DISEASE | 1 | 0 | 1 | 0 | 0 | 1 | 0 | 1 | 0 | 0 |
| ***ORAL SOFT TISSUE CONDITIONS*** | | | | | | | | | | |
| *ORAL SOFT TISSUE DISORDERS NEC* | | | | | | | | | | |
| CHEILITIS | 1 | 1 | 0 | 0 | 0 | 1 | 1 | 0 | 0 | 0 |
| LIP BLISTER | 2 | 2 | 0 | 0 | 0 | 2 | 2 | 0 | 0 | 0 |

| | | | | | | | | | | |
|---|---|---|---|---|---|---|---|---|---|---|
| ORAL LICHEN PLANUS | 1 | 1 | 0 | 0 | 0 | 1 | 1 | 0 | 0 | 0 |
| *ORAL SOFT TISSUE INFECTIONS* | | | | | | | | | | |
| ANGULAR CHEILITIS | 1 | 1 | 0 | 0 | 0 | 1 | 1 | 0 | 0 | 0 |
| *ORAL SOFT TISSUE SIGNS AND SYMPTOMS* | | | | | | | | | | |
| HYPOAESTHESIA ORAL | 2 | 2 | 0 | 0 | 0 | 2 | 2 | 0 | 0 | 0 |
| LIP PAIN | 3 | 2 | 1 | 0 | 0 | 3 | 2 | 1 | 0 | 0 |
| ORAL DISCOMFORT | 2 | 2 | 0 | 0 | 0 | 2 | 2 | 0 | 0 | 0 |
| ORAL MUCOSAL ROUGHENING | 0 | 0 | 0 | 0 | 0 | 0 | 0 | 0 | 0 | 0 |
| ORAL PAIN | 8 | 7 | 1 | 0 | 0 | 7 | 7 | 0 | 0 | 0 |
| PARAESTHESIA ORAL | 18 | 16 | 2 | 0 | 0 | 17 | 16 | 1 | 0 | 0 |
| LIP SWELLING | 6 | 6 | 0 | 0 | 0 | 6 | 6 | 0 | 0 | 0 |
| MOUTH SWELLING | 1 | 1 | 0 | 0 | 0 | 1 | 1 | 0 | 0 | 0 |
| *STOMATITIS AND ULCERATION* | | | | | | | | | | |
| APHTHOUS ULCER | 1 | 0 | 1 | 0 | 0 | 0 | 0 | 0 | 0 | 0 |
| LIP ULCERATION | 0 | 0 | 0 | 0 | 0 | 0 | 0 | 0 | 0 | 0 |
| MOUTH ULCERATION | 6 | 4 | 2 | 0 | 0 | 6 | 4 | 2 | 0 | 0 |
| STOMATITIS | 1 | 0 | 1 | 0 | 0 | 1 | 0 | 1 | 0 | 0 |
| ***SALIVARY GLAND CONDITIONS*** | | | | | | | | | | |
| *ORAL DRYNESS AND SALIVA ALTERED* | | | | | | | | | | |
| DRY MOUTH | 19 | 15 | 4 | 0 | 0 | 18 | 15 | 3 | 0 | 0 |
| LIP DRY | 4 | 3 | 1 | 0 | 0 | 4 | 3 | 1 | 0 | 0 |
| SALIVARY HYPOSECRETION | 1 | 1 | 0 | 0 | 0 | 1 | 1 | 0 | 0 | 0 |
| ***TONGUE CONDITIONS*** | | | | | | | | | | |
| *TONGUE SIGNS AND SYMPTOMS* | | | | | | | | | | |
| GLOSSODYNIA | 3 | 2 | 1 | 0 | 0 | 3 | 2 | 1 | 0 | 0 |
| SWOLLEN TONGUE | 4 | 4 | 0 | 0 | 0 | 4 | 4 | 0 | 0 | 0 |
| TONGUE COATED | 1 | 1 | 0 | 0 | 0 | 1 | 1 | 0 | 0 | 0 |
| TONGUE DISCOMFORT | 1 | 1 | 0 | 0 | 0 | 1 | 1 | 0 | 0 | 0 |
| TONGUE OEDEMA | 0 | 0 | 0 | 0 | 0 | 0 | 0 | 0 | 0 | 0 |
| TONGUE SPASM | 0 | 0 | 0 | 0 | 0 | 0 | 0 | 0 | 0 | 0 |
| **GENERAL DISORDERS AND ADMINISTRATION SITE CONDITIONS** | | | | | | | | | | |
| ***ADMINISTRATION SITE REACTIONS*** | | | | | | | | | | |
| *ADMINISTRATION SITE REACTIONS NEC* | | | | | | | | | | |
| ADMINISTRATION SITE BRUISE | 2 | 2 | 0 | 0 | 0 | 2 | 2 | 0 | 0 | 0 |
| ADMINISTRATION SITE PAIN | 0 | 0 | 0 | 0 | 0 | 0 | 0 | 0 | 0 | 0 |

| | | | | | | | | | | |
|---|---|---|---|---|---|---|---|---|---|---|
| PUNCTURE SITE BRUISE | 7 | 5 | 2 | 0 | 0 | 7 | 5 | 2 | 0 | 0 |
| PUNCTURE SITE PAIN | 1 | 1 | 0 | 0 | 0 | 1 | 1 | 0 | 0 | 0 |
| *APPLICATION SITE REACTIONS* | | | | | | | | | | |
| APPLICATION SITE BRUISE | 5 | 4 | 1 | 0 | 0 | 5 | 4 | 1 | 0 | 0 |
| APPLICATION SITE ERYTHEMA | 2 | 1 | 1 | 0 | 0 | 2 | 1 | 1 | 0 | 0 |
| APPLICATION SITE PAIN | 3 | 2 | 1 | 0 | 0 | 3 | 2 | 1 | 0 | 0 |
| *IMPLANT AND CATHETER SITE REACTIONS* | | | | | | | | | | |
| IMPLANT SITE PAIN | 1 | 0 | 1 | 0 | 0 | 1 | 0 | 1 | 0 | 0 |
| IMPLANT SITE WARMTH | 2 | 2 | 0 | 0 | 0 | 2 | 2 | 0 | 0 | 0 |
| *INFUSION SITE REACTIONS* | | | | | | | | | | |
| INFUSION SITE PAIN | 1 | 1 | 0 | 0 | 0 | 1 | 1 | 0 | 0 | 0 |
| INFUSION SITE SCAB | 1 | 1 | 0 | 0 | 0 | 1 | 1 | 0 | 0 | 0 |
| INFUSION SITE WARMTH | 1 | 1 | 0 | 0 | 0 | 1 | 1 | 0 | 0 | 0 |
| *INJECTION SITE REACTIONS* | | | | | | | | | | |
| INJECTION SITE BRUISING | 2 | 1 | 1 | 0 | 0 | 2 | 1 | 1 | 0 | 0 |
| INJECTION SITE DISCOMFORT | 1 | 1 | 0 | 0 | 0 | 1 | 1 | 0 | 0 | 0 |
| INJECTION SITE ERYTHEMA | 15 | 13 | 2 | 0 | 0 | 14 | 13 | 1 | 0 | 0 |
| INJECTION SITE HYPERSENSITIVITY | 0 | 0 | 0 | 0 | 0 | 0 | 0 | 0 | 0 | 0 |
| INJECTION SITE INFLAMMATION | 1 | 1 | 0 | 0 | 0 | 1 | 1 | 0 | 0 | 0 |
| INJECTION SITE INJURY | 0 | 0 | 0 | 0 | 0 | 0 | 0 | 0 | 0 | 0 |
| INJECTION SITE IRRITATION | 0 | 0 | 0 | 0 | 0 | 0 | 0 | 0 | 0 | 0 |
| INJECTION SITE JOINT PAIN | 2 | 1 | 1 | 0 | 0 | 1 | 1 | 0 | 0 | 0 |
| INJECTION SITE MASS | 28 | 25 | 3 | 0 | 0 | 27 | 25 | 2 | 0 | 0 |
| INJECTION SITE NODULE | 0 | 0 | 0 | 0 | 0 | 0 | 0 | 0 | 0 | 0 |
| INJECTION SITE OEDEMA | 1 | 1 | 0 | 0 | 0 | 1 | 1 | 0 | 0 | 0 |
| INJECTION SITE PAIN | 229 | 180 | 49 | 0 | 0 | 219 | 180 | 39 | 0 | 0 |
| INJECTION SITE PAPULE | 1 | 1 | 0 | 0 | 0 | 1 | 1 | 0 | 0 | 0 |
| INJECTION SITE PARAESTHESIA | 0 | 0 | 0 | 0 | 0 | 0 | 0 | 0 | 0 | 0 |
| INJECTION SITE PRURITUS | 8 | 7 | 1 | 0 | 0 | 7 | 7 | 0 | 0 | 0 |
| INJECTION SITE RASH | 4 | 4 | 0 | 0 | 0 | 4 | 4 | 0 | 0 | 0 |
| INJECTION SITE REACTION | 2 | 2 | 0 | 0 | 0 | 2 | 2 | 0 | 0 | 0 |
| INJECTION SITE SCAB | 2 | 1 | 1 | 0 | 0 | 2 | 1 | 1 | 0 | 0 |
| INJECTION SITE SWELLING | 8 | 7 | 1 | 0 | 0 | 8 | 7 | 1 | 0 | 0 |
| INJECTION SITE URTICARIA | 0 | 0 | 0 | 0 | 0 | 0 | 0 | 0 | 0 | 0 |
| INJECTION SITE WARMTH | 10 | 9 | 1 | 0 | 0 | 10 | 9 | 1 | 0 | 0 |

| | | | | | | | | | | |
|---|---|---|---|---|---|---|---|---|---|---|
| *INSTILLATION SITE REACTIONS* | | | | | | | | | | |
| INSTILLATION SITE PRURITUS | 0 | 0 | 0 | 0 | 0 | 0 | 0 | 0 | 0 | 0 |
| INSTILLATION SITE WARMTH | 1 | 1 | 0 | 0 | 0 | 1 | 1 | 0 | 0 | 0 |
| *VACCINATION SITE REACTIONS* | | | | | | | | | | |
| SHOULDER INJURY RELATED TO VACCINE ADMINISTRATION | 1 | 1 | 0 | 0 | 0 | 1 | 1 | 0 | 0 | 0 |
| VACCINATION SITE BRUISING | 4 | 1 | 3 | 0 | 0 | 3 | 1 | 2 | 0 | 0 |
| VACCINATION SITE DISCOMFORT | 0 | 0 | 0 | 0 | 0 | 0 | 0 | 0 | 0 | 0 |
| VACCINATION SITE ERYTHEMA | 0 | 0 | 0 | 0 | 0 | 0 | 0 | 0 | 0 | 0 |
| VACCINATION SITE JOINT ERYTHEMA | 1 | 1 | 0 | 0 | 0 | 1 | 1 | 0 | 0 | 0 |
| VACCINATION SITE JOINT PAIN | 0 | 0 | 0 | 0 | 0 | 0 | 0 | 0 | 0 | 0 |
| VACCINATION SITE MASS | 12 | 9 | 3 | 0 | 0 | 10 | 9 | 1 | 0 | 0 |
| VACCINATION SITE PAIN | 41 | 32 | 9 | 0 | 0 | 38 | 32 | 6 | 0 | 0 |
| VACCINATION SITE RASH | 1 | 1 | 0 | 0 | 0 | 1 | 1 | 0 | 0 | 0 |
| VACCINATION SITE SWELLING | 0 | 0 | 0 | 0 | 0 | 0 | 0 | 0 | 0 | 0 |
| VACCINATION SITE WARMTH | 4 | 3 | 1 | 0 | 0 | 3 | 3 | 0 | 0 | 0 |
| ***BODY TEMPERATURE CONDITIONS*** | | | | | | | | | | |
| *BODY TEMPERATURE ALTERED* | | | | | | | | | | |
| HYPERTHERMIA | 1 | 1 | 0 | 0 | 0 | 1 | 1 | 0 | 0 | 0 |
| HYPOTHERMIA | 0 | 0 | 0 | 0 | 0 | 0 | 0 | 0 | 0 | 0 |
| *FEBRILE DISORDERS* | | | | | | | | | | |
| PYREXIA | 1644 | 1470 | 174 | 0 | 0 | 1560 | 1470 | 90 | 0 | 0 |
| ***FATAL OUTCOMES*** | | | | | | | | | | |
| *DEATH AND SUDDEN DEATH* | | | | | | | | | | |
| DEATH | 2 | 2 | 0 | 0 | 0 | 2 | 2 | 0 | 0 | 0 |
| ***GENERAL SYSTEM DISORDERS NEC*** | | | | | | | | | | |
| *ADVERSE EFFECT ABSENT* | | | | | | | | | | |
| NO ADVERSE EVENT | 0 | 0 | 0 | 0 | 0 | 0 | 0 | 0 | 0 | 0 |
| *ASTHENIC CONDITIONS* | | | | | | | | | | |
| ASTHENIA | 74 | 60 | 14 | 0 | 0 | 71 | 60 | 11 | 0 | 0 |
| CHRONIC FATIGUE SYNDROME | 1 | 1 | 0 | 0 | 0 | 1 | 1 | 0 | 0 | 0 |
| DECREASED ACTIVITY | 0 | 0 | 0 | 0 | 0 | 0 | 0 | 0 | 0 | 0 |
| FATIGUE | 2711 | 2205 | 503 | 3 | 0 | 2565 | 2205 | 359 | 1 | 0 |
| MALAISE | 270 | 229 | 40 | 1 | 0 | 256 | 229 | 26 | 1 | 0 |
| SLUGGISHNESS | 1 | 0 | 1 | 0 | 0 | 1 | 0 | 1 | 0 | 0 |
| *FEELINGS AND SENSATIONS NEC* | | | | | | | | | | |

| | | | | | | | | | | |
|---|---|---|---|---|---|---|---|---|---|---|
| CHILLS | 1369 | 1235 | 132 | 2 | 0 | 1310 | 1235 | 74 | 1 | 0 |
| FEELING ABNORMAL | 78 | 63 | 15 | 0 | 0 | 72 | 63 | 9 | 0 | 0 |
| FEELING COLD | 264 | 226 | 37 | 1 | 0 | 242 | 226 | 16 | 0 | 0 |
| FEELING HOT | 65 | 54 | 11 | 0 | 0 | 62 | 54 | 8 | 0 | 0 |
| FEELING JITTERY | 1 | 0 | 1 | 0 | 0 | 1 | 0 | 1 | 0 | 0 |
| FEELING OF BODY TEMPERATURE CHANGE | 42 | 37 | 5 | 0 | 0 | 39 | 37 | 2 | 0 | 0 |
| FEELING OF RELAXATION | 1 | 1 | 0 | 0 | 0 | 1 | 1 | 0 | 0 | 0 |
| HANGOVER | 6 | 6 | 0 | 0 | 0 | 6 | 6 | 0 | 0 | 0 |
| HUNGER | 3 | 2 | 0 | 1 | 0 | 2 | 2 | 0 | 0 | 0 |
| SENSATION OF BLOOD FLOW | 1 | 1 | 0 | 0 | 0 | 1 | 1 | 0 | 0 | 0 |
| THIRST | 32 | 29 | 3 | 0 | 0 | 30 | 29 | 1 | 0 | 0 |
| *GAIT DISTURBANCES* | | | | | | | | | | |
| GAIT DISTURBANCE | 3 | 2 | 1 | 0 | 0 | 3 | 2 | 1 | 0 | 0 |
| GAIT INABILITY | 2 | 2 | 0 | 0 | 0 | 2 | 2 | 0 | 0 | 0 |
| *GENERAL SIGNS AND SYMPTOMS NEC* | | | | | | | | | | |
| CRYING | 1 | 1 | 0 | 0 | 0 | 1 | 1 | 0 | 0 | 0 |
| ENERGY INCREASED | 1 | 0 | 1 | 0 | 0 | 1 | 0 | 1 | 0 | 0 |
| EXERCISE TOLERANCE DECREASED | 1 | 1 | 0 | 0 | 0 | 1 | 1 | 0 | 0 | 0 |
| GENERAL SYMPTOM | 1 | 1 | 0 | 0 | 0 | 1 | 1 | 0 | 0 | 0 |
| ILLNESS | 85 | 75 | 10 | 0 | 0 | 79 | 75 | 4 | 0 | 0 |
| INFLUENZA LIKE ILLNESS | 488 | 410 | 77 | 1 | 0 | 447 | 410 | 37 | 0 | 0 |
| LOCAL REACTION | 3 | 3 | 0 | 0 | 0 | 3 | 3 | 0 | 0 | 0 |
| PERIPHERAL SWELLING | 103 | 84 | 19 | 0 | 0 | 100 | 84 | 16 | 0 | 0 |
| SWELLING | 34 | 26 | 8 | 0 | 0 | 32 | 26 | 6 | 0 | 0 |
| SWELLING FACE | 8 | 7 | 1 | 0 | 0 | 8 | 7 | 1 | 0 | 0 |
| TISSUE IRRITATION | 1 | 0 | 1 | 0 | 0 | 1 | 0 | 1 | 0 | 0 |
| *INFLAMMATIONS* | | | | | | | | | | |
| INFLAMMATION | 7 | 4 | 3 | 0 | 0 | 5 | 4 | 1 | 0 | 0 |
| SYSTEMIC INFLAMMATORY RESPONSE SYNDROME | 1 | 0 | 1 | 0 | 0 | 0 | 0 | 0 | 0 | 0 |
| *OEDEMA NEC* | | | | | | | | | | |
| OEDEMA | 1 | 1 | 0 | 0 | 0 | 1 | 1 | 0 | 0 | 0 |
| OEDEMA PERIPHERAL | 1 | 1 | 0 | 0 | 0 | 1 | 1 | 0 | 0 | 0 |
| *PAIN AND DISCOMFORT NEC* | | | | | | | | | | |
| AXILLARY PAIN | 25 | 20 | 5 | 0 | 0 | 24 | 20 | 4 | 0 | 0 |

| | | | | | | | | | | |
|---|---|---|---|---|---|---|---|---|---|---|
| CHEST DISCOMFORT | 24 | 22 | 2 | 0 | 0 | 23 | 22 | 1 | 0 | 0 |
| CHEST PAIN | 47 | 40 | 7 | 0 | 0 | 44 | 40 | 4 | 0 | 0 |
| DISCOMFORT | 7 | 5 | 2 | 0 | 0 | 6 | 5 | 1 | 0 | 0 |
| FACIAL PAIN | 2 | 2 | 0 | 0 | 0 | 2 | 2 | 0 | 0 | 0 |
| HERNIA PAIN | 0 | 0 | 0 | 0 | 0 | 0 | 0 | 0 | 0 | 0 |
| INFLAMMATORY PAIN | 1 | 1 | 0 | 0 | 0 | 1 | 1 | 0 | 0 | 0 |
| NON-CARDIAC CHEST PAIN | 0 | 0 | 0 | 0 | 0 | 0 | 0 | 0 | 0 | 0 |
| PAIN | 578 | 485 | 93 | 0 | 0 | 542 | 485 | 57 | 0 | 0 |
| TENDERNESS | 202 | 173 | 28 | 1 | 0 | 196 | 173 | 22 | 1 | 0 |
| ***THERAPEUTIC AND NONTHERAPEUTIC EFFECTS (EXCL TOXICITY)*** | | | | | | | | | | |
| *THERAPEUTIC AND NONTHERAPEUTIC RESPONSES* | | | | | | | | | | |
| ADVERSE DRUG REACTION | 1 | 1 | 0 | 0 | 0 | 1 | 1 | 0 | 0 | 0 |
| ADVERSE EVENT | 0 | 0 | 0 | 0 | 0 | 0 | 0 | 0 | 0 | 0 |
| ADVERSE REACTION | 0 | 0 | 0 | 0 | 0 | 0 | 0 | 0 | 0 | 0 |
| IMMEDIATE POST-INJECTION REACTION | 0 | 0 | 0 | 0 | 0 | 0 | 0 | 0 | 0 | 0 |
| **HEPATOBILIARY DISORDERS** | | | | | | | | | | |
| ***HEPATIC AND HEPATOBILIARY DISORDERS*** | | | | | | | | | | |
| *HEPATOBILIARY SIGNS AND SYMPTOMS* | | | | | | | | | | |
| HEPATIC PAIN | 2 | 2 | 0 | 0 | 0 | 2 | 2 | 0 | 0 | 0 |
| *HEPATOCELLULAR DAMAGE AND HEPATITIS NEC* | | | | | | | | | | |
| LIVER INJURY | 1 | 1 | 0 | 0 | 0 | 1 | 1 | 0 | 0 | 0 |
| **IMMUNE SYSTEM DISORDERS** | | | | | | | | | | |
| ***ALLERGIC CONDITIONS*** | | | | | | | | | | |
| *ALLERGIC CONDITIONS NEC* | | | | | | | | | | |
| HYPERSENSITIVITY | 7 | 4 | 3 | 0 | 0 | 5 | 4 | 1 | 0 | 0 |
| MULTIPLE ALLERGIES | 1 | 1 | 0 | 0 | 0 | 1 | 1 | 0 | 0 | 0 |
| *ALLERGIES TO FOODS, FOOD ADDITIVES, DRUGS AND OTHER CHEMICALS* | | | | | | | | | | |
| ALLERGY TO CHEMICALS | 1 | 0 | 1 | 0 | 0 | 1 | 0 | 1 | 0 | 0 |
| *ANAPHYLACTIC AND ANAPHYLACTOID RESPONSES* | | | | | | | | | | |
| ANAPHYLACTIC REACTION | 3 | 3 | 0 | 0 | 0 | 3 | 3 | 0 | 0 | 0 |
| *ATOPIC DISORDERS* | | | | | | | | | | |
| SEASONAL ALLERGY | 1 | 0 | 1 | 0 | 0 | 1 | 0 | 1 | 0 | 0 |
| ***IMMUNE DISORDERS NEC*** | | | | | | | | | | |
| *IMMUNE AND ASSOCIATED CONDITIONS NEC* | | | | | | | | | | |
| BACILLE CALMETTE-GUERIN SCAR REACTIVATION | 1 | 1 | 0 | 0 | 0 | 1 | 1 | 0 | 0 | 0 |

| | | | | | | | | | | |
|---|---|---|---|---|---|---|---|---|---|---|
| SENSITISATION | 1 | 1 | 0 | 0 | 0 | 1 | 1 | 0 | 0 | 0 |
| **INFECTIONS AND INFESTATIONS** | | | | | | | | | | |
| ***BACTERIAL INFECTIOUS DISORDERS*** | | | | | | | | | | |
| *BACTERIAL INFECTIONS NEC* | | | | | | | | | | |
| CELLULITIS | 2 | 2 | 0 | 0 | 0 | 2 | 2 | 0 | 0 | 0 |
| PERIORBITAL CELLULITIS | 0 | 0 | 0 | 0 | 0 | 0 | 0 | 0 | 0 | 0 |
| *HELICOBACTER INFECTIONS* | | | | | | | | | | |
| HELICOBACTER GASTRITIS | 0 | 0 | 0 | 0 | 0 | 0 | 0 | 0 | 0 | 0 |
| *STAPHYLOCOCCAL INFECTIONS* | | | | | | | | | | |
| FURUNCLE | 2 | 1 | 1 | 0 | 0 | 1 | 1 | 0 | 0 | 0 |
| ***FUNGAL INFECTIOUS DISORDERS*** | | | | | | | | | | |
| *CANDIDA INFECTIONS* | | | | | | | | | | |
| CANDIDA INFECTION | 0 | 0 | 0 | 0 | 0 | 0 | 0 | 0 | 0 | 0 |
| VULVOVAGINAL CANDIDIASIS | 2 | 2 | 0 | 0 | 0 | 2 | 2 | 0 | 0 | 0 |
| ***INFECTIONS - PATHOGEN UNSPECIFIED*** | | | | | | | | | | |
| *ABDOMINAL AND GASTROINTESTINAL INFECTIONS* | | | | | | | | | | |
| APPENDICITIS | 1 | 1 | 0 | 0 | 0 | 1 | 1 | 0 | 0 | 0 |
| DIARRHOEA INFECTIOUS | 0 | 0 | 0 | 0 | 0 | 0 | 0 | 0 | 0 | 0 |
| GASTROINTESTINAL INFECTION | 0 | 0 | 0 | 0 | 0 | 0 | 0 | 0 | 0 | 0 |
| *BREAST INFECTIONS* | | | | | | | | | | |
| MASTITIS | 1 | 0 | 1 | 0 | 0 | 1 | 0 | 1 | 0 | 0 |
| *CENTRAL NERVOUS SYSTEM AND SPINAL INFECTIONS* | | | | | | | | | | |
| MYELITIS | 1 | 1 | 0 | 0 | 0 | 1 | 1 | 0 | 0 | 0 |
| *DENTAL AND ORAL SOFT TISSUE INFECTIONS* | | | | | | | | | | |
| ABSCESS ORAL | 0 | 0 | 0 | 0 | 0 | 0 | 0 | 0 | 0 | 0 |
| ORAL PUSTULE | 1 | 1 | 0 | 0 | 0 | 1 | 1 | 0 | 0 | 0 |
| PERICORONITIS | 1 | 1 | 0 | 0 | 0 | 1 | 1 | 0 | 0 | 0 |
| TOOTH ABSCESS | 1 | 1 | 0 | 0 | 0 | 1 | 1 | 0 | 0 | 0 |
| *EAR INFECTIONS* | | | | | | | | | | |
| EAR INFECTION | 1 | 1 | 0 | 0 | 0 | 1 | 1 | 0 | 0 | 0 |
| LABYRINTHITIS | 0 | 0 | 0 | 0 | 0 | 0 | 0 | 0 | 0 | 0 |
| OTITIS EXTERNA | 1 | 1 | 0 | 0 | 0 | 1 | 1 | 0 | 0 | 0 |
| *EYE AND EYELID INFECTIONS* | | | | | | | | | | |
| CONJUNCTIVITIS | 1 | 0 | 1 | 0 | 0 | 1 | 0 | 1 | 0 | 0 |
| HORDEOLUM | 1 | 1 | 0 | 0 | 0 | 1 | 1 | 0 | 0 | 0 |
| *INFECTIONS NEC* | | | | | | | | | | |

| | | | | | | | | | | |
|---|---|---|---|---|---|---|---|---|---|---|
| ABSCESS | 1 | 1 | 0 | 0 | 0 | 1 | 1 | 0 | 0 | 0 |
| INFECTION | 0 | 0 | 0 | 0 | 0 | 0 | 0 | 0 | 0 | 0 |
| INJECTION SITE INFECTION | 1 | 1 | 0 | 0 | 0 | 1 | 1 | 0 | 0 | 0 |
| LOCALISED INFECTION | 3 | 3 | 0 | 0 | 0 | 3 | 3 | 0 | 0 | 0 |
| WOUND INFECTION | 1 | 0 | 1 | 0 | 0 | 0 | 0 | 0 | 0 | 0 |
| *LOWER RESPIRATORY TRACT AND LUNG INFECTIONS* | | | | | | | | | | |
| LOWER RESPIRATORY TRACT INFECTION | 3 | 2 | 1 | 0 | 0 | 3 | 2 | 1 | 0 | 0 |
| PNEUMONIA | 3 | 2 | 1 | 0 | 0 | 2 | 2 | 0 | 0 | 0 |
| *MALE REPRODUCTIVE TRACT INFECTIONS* | | | | | | | | | | |
| ORCHITIS | 1 | 1 | 0 | 0 | 0 | 1 | 1 | 0 | 0 | 0 |
| *SEPSIS, BACTERAEMIA, VIRAEMIA AND FUNGAEMIA NEC* | | | | | | | | | | |
| SEPTIC RASH | 1 | 0 | 1 | 0 | 0 | 0 | 0 | 0 | 0 | 0 |
| *SKIN STRUCTURES AND SOFT TISSUE INFECTIONS* | | | | | | | | | | |
| INFECTED DERMAL CYST | 0 | 0 | 0 | 0 | 0 | 0 | 0 | 0 | 0 | 0 |
| INJECTION SITE PUSTULE | 1 | 0 | 1 | 0 | 0 | 0 | 0 | 0 | 0 | 0 |
| SKIN INFECTION | 0 | 0 | 0 | 0 | 0 | 0 | 0 | 0 | 0 | 0 |
| *UPPER RESPIRATORY TRACT INFECTIONS* | | | | | | | | | | |
| LARYNGITIS | 0 | 0 | 0 | 0 | 0 | 0 | 0 | 0 | 0 | 0 |
| NASOPHARYNGITIS | 120 | 94 | 26 | 0 | 0 | 112 | 94 | 18 | 0 | 0 |
| PHARYNGITIS | 1 | 1 | 0 | 0 | 0 | 1 | 1 | 0 | 0 | 0 |
| RHINITIS | 1 | 1 | 0 | 0 | 0 | 1 | 1 | 0 | 0 | 0 |
| SINUSITIS | 9 | 8 | 1 | 0 | 0 | 8 | 8 | 0 | 0 | 0 |
| TONSILLITIS | 0 | 0 | 0 | 0 | 0 | 0 | 0 | 0 | 0 | 0 |
| *URINARY TRACT INFECTIONS* | | | | | | | | | | |
| CYSTITIS | 0 | 0 | 0 | 0 | 0 | 0 | 0 | 0 | 0 | 0 |
| URINARY TRACT INFECTION | 3 | 2 | 1 | 0 | 0 | 2 | 2 | 0 | 0 | 0 |
| ***PROTOZOAL INFECTIOUS DISORDERS*** | | | | | | | | | | |
| *TRYPANOSOMAL INFECTIONS* | | | | | | | | | | |
| AFRICAN TRYPANOSOMIASIS | 0 | 0 | 0 | 0 | 0 | 0 | 0 | 0 | 0 | 0 |
| ***RICKETTSIAL INFECTIOUS DISORDERS*** | | | | | | | | | | |
| *COXIELLA INFECTIONS* | | | | | | | | | | |
| Q FEVER | 4 | 4 | 0 | 0 | 0 | 4 | 4 | 0 | 0 | 0 |
| ***VIRAL INFECTIOUS DISORDERS*** | | | | | | | | | | |
| *CORONAVIRUS INFECTIONS* | | | | | | | | | | |
| COVID-19 | 1 | 0 | 1 | 0 | 0 | 0 | 0 | 0 | 0 | 0 |
| *HERPES VIRAL INFECTIONS* | | | | | | | | | | |

| | | | | | | | | | | |
|---|---|---|---|---|---|---|---|---|---|---|
| GENITAL HERPES | 0 | 0 | 0 | 0 | 0 | 0 | 0 | 0 | 0 | 0 |
| HERPES OPHTHALMIC | 1 | 1 | 0 | 0 | 0 | 1 | 1 | 0 | 0 | 0 |
| HERPES SIMPLEX | 1 | 0 | 1 | 0 | 0 | 0 | 0 | 0 | 0 | 0 |
| HERPES ZOSTER | 6 | 4 | 2 | 0 | 0 | 6 | 4 | 2 | 0 | 0 |
| NASAL HERPES | 1 | 1 | 0 | 0 | 0 | 1 | 1 | 0 | 0 | 0 |
| OPHTHALMIC HERPES ZOSTER | 1 | 1 | 0 | 0 | 0 | 1 | 1 | 0 | 0 | 0 |
| ORAL HERPES | 18 | 15 | 3 | 0 | 0 | 17 | 15 | 2 | 0 | 0 |
| *INFLUENZA VIRAL INFECTIONS* | | | | | | | | | | |
| H2N2 INFLUENZA | 1 | 1 | 0 | 0 | 0 | 1 | 1 | 0 | 0 | 0 |
| INFLUENZA | 463 | 409 | 54 | 0 | 0 | 440 | 409 | 31 | 0 | 0 |
| *RETROVIRAL INFECTIONS* | | | | | | | | | | |
| AIDS RELATED COMPLEX | 1 | 1 | 0 | 0 | 0 | 1 | 1 | 0 | 0 | 0 |
| *VIRAL INFECTIONS NEC* | | | | | | | | | | |
| GASTROENTERITIS VIRAL | 4 | 4 | 0 | 0 | 0 | 4 | 4 | 0 | 0 | 0 |
| SWEATING FEVER | 7 | 7 | 0 | 0 | 0 | 7 | 7 | 0 | 0 | 0 |
| VESTIBULAR NEURONITIS | 0 | 0 | 0 | 0 | 0 | 0 | 0 | 0 | 0 | 0 |
| VIRAL DIARRHOEA | 0 | 0 | 0 | 0 | 0 | 0 | 0 | 0 | 0 | 0 |
| VIRAL RASH | 1 | 1 | 0 | 0 | 0 | 1 | 1 | 0 | 0 | 0 |
| VIRAL UPPER RESPIRATORY TRACT INFECTION | 1 | 1 | 0 | 0 | 0 | 1 | 1 | 0 | 0 | 0 |
| **INJURY, POISONING AND PROCEDURAL COMPLICATIONS** | | | | | | | | | | |
| ***BONE AND JOINT INJURIES*** | | | | | | | | | | |
| *FRACTURES AND DISLOCATIONS NEC* | | | | | | | | | | |
| JOINT DISLOCATION | 1 | 1 | 0 | 0 | 0 | 1 | 1 | 0 | 0 | 0 |
| ***EXPOSURES, CHEMICAL INJURIES AND POISONING*** | | | | | | | | | | |
| *POISONING AND TOXICITY* | | | | | | | | | | |
| SYSTEMIC TOXICITY | 1 | 1 | 0 | 0 | 0 | 1 | 1 | 0 | 0 | 0 |
| TOXICITY TO VARIOUS AGENTS | 1 | 1 | 0 | 0 | 0 | 1 | 1 | 0 | 0 | 0 |
| ***INJURIES BY PHYSICAL AGENTS*** | | | | | | | | | | |
| *CONDITIONS CAUSED BY COLD* | | | | | | | | | | |
| CHILLBLAINS | 1 | 1 | 0 | 0 | 0 | 1 | 1 | 0 | 0 | 0 |
| *HEAT INJURIES (EXCL THERMAL BURNS)* | | | | | | | | | | |
| HEAT EXHAUSTION | 1 | 1 | 0 | 0 | 0 | 1 | 1 | 0 | 0 | 0 |
| HEAT OEDEMA | 1 | 1 | 0 | 0 | 0 | 1 | 1 | 0 | 0 | 0 |
| *RADIATION INJURIES* | | | | | | | | | | |
| SUNBURN | 1 | 0 | 1 | 0 | 0 | 0 | 0 | 0 | 0 | 0 |
| *THERMAL BURNS* | | | | | | | | | | |

| | | | | | | | | | | |
|---|---|---|---|---|---|---|---|---|---|---|
| THERMAL BURN | 1 | 1 | 0 | 0 | 0 | 1 | 1 | 0 | 0 | 0 |
| THERMAL BURNS OF EYE | 1 | 1 | 0 | 0 | 0 | 1 | 1 | 0 | 0 | 0 |
| ***INJURIES NEC*** | | | | | | | | | | |
| *CHEST AND RESPIRATORY TRACT INJURIES NEC* | | | | | | | | | | |
| CHEST CRUSHING | 1 | 1 | 0 | 0 | 0 | 1 | 1 | 0 | 0 | 0 |
| *EYE INJURIES NEC* | | | | | | | | | | |
| EYE CONTUSION | 0 | 0 | 0 | 0 | 0 | 0 | 0 | 0 | 0 | 0 |
| *MUSCLE, TENDON AND LIGAMENT INJURIES* | | | | | | | | | | |
| LIGAMENT SPRAIN | 1 | 1 | 0 | 0 | 0 | 1 | 1 | 0 | 0 | 0 |
| MUSCLE INJURY | 1 | 1 | 0 | 0 | 0 | 1 | 1 | 0 | 0 | 0 |
| MUSCLE STRAIN | 2 | 1 | 1 | 0 | 0 | 2 | 1 | 1 | 0 | 0 |
| *NERVE INJURIES NEC* | | | | | | | | | | |
| NERVE INJURY | 1 | 1 | 0 | 0 | 0 | 1 | 1 | 0 | 0 | 0 |
| *NON-SITE SPECIFIC INJURIES NEC* | | | | | | | | | | |
| ARTHROPOD STING | 0 | 0 | 0 | 0 | 0 | 0 | 0 | 0 | 0 | 0 |
| BITE | 1 | 0 | 1 | 0 | 0 | 1 | 0 | 1 | 0 | 0 |
| FALL | 3 | 3 | 0 | 0 | 0 | 3 | 3 | 0 | 0 | 0 |
| INFLAMMATION OF WOUND | 1 | 1 | 0 | 0 | 0 | 1 | 1 | 0 | 0 | 0 |
| TISSUE INJURY | 1 | 1 | 0 | 0 | 0 | 1 | 1 | 0 | 0 | 0 |
| WOUND COMPLICATION | 1 | 1 | 0 | 0 | 0 | 1 | 1 | 0 | 0 | 0 |
| WOUND SECRETION | 1 | 1 | 0 | 0 | 0 | 1 | 1 | 0 | 0 | 0 |
| *SITE SPECIFIC INJURIES NEC* | | | | | | | | | | |
| LIMB INJURY | 6 | 6 | 0 | 0 | 0 | 6 | 6 | 0 | 0 | 0 |
| *SKIN INJURIES NEC* | | | | | | | | | | |
| CONTUSION | 70 | 53 | 17 | 0 | 0 | 63 | 53 | 10 | 0 | 0 |
| SCAR | 1 | 0 | 1 | 0 | 0 | 1 | 0 | 1 | 0 | 0 |
| SKIN WOUND | 1 | 0 | 1 | 0 | 0 | 0 | 0 | 0 | 0 | 0 |
| ***PROCEDURAL RELATED INJURIES AND COMPLICATIONS NEC*** | | | | | | | | | | |
| *CARDIAC AND VASCULAR PROCEDURAL COMPLICATIONS* | | | | | | | | | | |
| CARDIAC PROCEDURE COMPLICATION | 1 | 0 | 1 | 0 | 0 | 0 | 0 | 0 | 0 | 0 |
| *GASTROINTESTINAL AND HEPATOBILIARY PROCEDURAL COMPLICATIONS* | | | | | | | | | | |
| PROCEDURAL NAUSEA | 6 | 6 | 0 | 0 | 0 | 6 | 6 | 0 | 0 | 0 |
| *NEUROLOGICAL AND PSYCHIATRIC PROCEDURAL COMPLICATIONS* | | | | | | | | | | |
| PROCEDURAL DIZZINESS | 2 | 2 | 0 | 0 | 0 | 2 | 2 | 0 | 0 | 0 |
| *NON-SITE SPECIFIC PROCEDURAL COMPLICATIONS* | | | | | | | | | | |
| INCISION SITE PAIN | 0 | 0 | 0 | 0 | 0 | 0 | 0 | 0 | 0 | 0 |

| | | | | | | | | | | |
|---|---|---|---|---|---|---|---|---|---|---|
| INCISION SITE SWELLING | 0 | 0 | 0 | 0 | 0 | 0 | 0 | 0 | 0 | 0 |
| INJECTION RELATED REACTION | 43 | 39 | 3 | 1 | 0 | 43 | 39 | 3 | 1 | 0 |
| POST PROCEDURAL COMPLICATION | 5 | 3 | 2 | 0 | 0 | 3 | 3 | 0 | 0 | 0 |
| PROCEDURAL PAIN | 0 | 0 | 0 | 0 | 0 | 0 | 0 | 0 | 0 | 0 |
| *VACCINATION RELATED COMPLICATIONS* | | | | | | | | | | |
| IMMUNISATION REACTION | 5 | 2 | 3 | 0 | 0 | 4 | 2 | 2 | 0 | 0 |
| **INVESTIGATIONS** | | | | | | | | | | |
| ***CARDIAC AND VASCULAR INVESTIGATIONS (EXCL ENZYME TESTS)*** | | | | | | | | | | |
| *HEART RATE AND PULSE INVESTIGATIONS* | | | | | | | | | | |
| HEART RATE | 18 | 16 | 2 | 0 | 0 | 18 | 16 | 2 | 0 | 0 |
| HEART RATE DECREASED | 1 | 1 | 0 | 0 | 0 | 1 | 1 | 0 | 0 | 0 |
| HEART RATE INCREASED | 28 | 25 | 3 | 0 | 0 | 26 | 25 | 1 | 0 | 0 |
| HEART RATE IRREGULAR | 0 | 0 | 0 | 0 | 0 | 0 | 0 | 0 | 0 | 0 |
| *VASCULAR TESTS NEC (INCL BLOOD PRESSURE)* | | | | | | | | | | |
| BLOOD PRESSURE DECREASED | 1 | 0 | 1 | 0 | 0 | 1 | 0 | 1 | 0 | 0 |
| BLOOD PRESSURE INCREASED | 4 | 3 | 1 | 0 | 0 | 4 | 3 | 1 | 0 | 0 |
| BLOOD PRESSURE MEASUREMENT | 1 | 1 | 0 | 0 | 0 | 1 | 1 | 0 | 0 | 0 |
| ***ENDOCRINE INVESTIGATIONS (INCL SEX HORMONES)*** | | | | | | | | | | |
| *ENDOCRINE ANALYSES AND IMAGING NEC* | | | | | | | | | | |
| HORMONE LEVEL ABNORMAL | 0 | 0 | 0 | 0 | 0 | 0 | 0 | 0 | 0 | 0 |
| *PITUITARY ANALYSES ANTERIOR* | | | | | | | | | | |
| BLOOD FOLLICLE STIMULATING HORMONE INCREASED | 0 | 0 | 0 | 0 | 0 | 0 | 0 | 0 | 0 | 0 |
| BLOOD LUTEINISING HORMONE | 0 | 0 | 0 | 0 | 0 | 0 | 0 | 0 | 0 | 0 |
| *THYROID ANALYSES* | | | | | | | | | | |
| TRI-IODOTHYRONINE | 1 | 1 | 0 | 0 | 0 | 1 | 1 | 0 | 0 | 0 |
| ***HAEMATOLOGY INVESTIGATIONS (INCL BLOOD GROUPS)*** | | | | | | | | | | |
| *COAGULATION AND BLEEDING ANALYSES* | | | | | | | | | | |
| BLEEDING TIME | 0 | 0 | 0 | 0 | 0 | 0 | 0 | 0 | 0 | 0 |
| INTERNATIONAL NORMALISED RATIO DECREASED | 1 | 0 | 1 | 0 | 0 | 0 | 0 | 0 | 0 | 0 |
| *PLATELET ANALYSES* | | | | | | | | | | |
| PLATELET COUNT INCREASED | 1 | 0 | 1 | 0 | 0 | 1 | 0 | 1 | 0 | 0 |
| *RED BLOOD CELL ANALYSES* | | | | | | | | | | |
| HAEMOGLOBIN | 1 | 1 | 0 | 0 | 0 | 1 | 1 | 0 | 0 | 0 |
| ***METABOLIC, NUTRITIONAL AND BLOOD GAS INVESTIGATIONS*** | | | | | | | | | | |
| *BLOOD GAS AND ACID BASE ANALYSES* | | | | | | | | | | |

| | | | | | | | | | | |
|---|---|---|---|---|---|---|---|---|---|---|
| OXYGEN SATURATION DECREASED | 2 | 2 | 0 | 0 | 0 | 2 | 2 | 0 | 0 | 0 |
| *CARBOHYDRATE TOLERANCE ANALYSES (INCL DIABETES)* | | | | | | | | | | |
| BLOOD GLUCOSE | 2 | 2 | 0 | 0 | 0 | 2 | 2 | 0 | 0 | 0 |
| BLOOD GLUCOSE ABNORMAL | 1 | 1 | 0 | 0 | 0 | 1 | 1 | 0 | 0 | 0 |
| BLOOD GLUCOSE DECREASED | 1 | 1 | 0 | 0 | 0 | 1 | 1 | 0 | 0 | 0 |
| BLOOD GLUCOSE INCREASED | 4 | 4 | 0 | 0 | 0 | 4 | 4 | 0 | 0 | 0 |
| ***MICROBIOLOGY AND SEROLOGY INVESTIGATIONS*** | | | | | | | | | | |
| *VIRUS IDENTIFICATION AND SEROLOGY* | | | | | | | | | | |
| SARS-COV-2 TEST | 0 | 0 | 0 | 0 | 0 | 0 | 0 | 0 | 0 | 0 |
| SARS-COV-2 TEST POSITIVE | 0 | 0 | 0 | 0 | 0 | 0 | 0 | 0 | 0 | 0 |
| ***MUSCULOSKELETAL AND SOFT TISSUE INVESTIGATIONS (EXCL ENZYME TESTS)*** | | | | | | | | | | |
| *MUSCULOSKELETAL AND SOFT TISSUE IMAGING PROCEDURES* | | | | | | | | | | |
| BONE SCAN | 1 | 1 | 0 | 0 | 0 | 1 | 1 | 0 | 0 | 0 |
| ***NEUROLOGICAL, SPECIAL SENSES AND PSYCHIATRIC INVESTIGATIONS*** | | | | | | | | | | |
| *CENTRAL NERVOUS SYSTEM IMAGING PROCEDURES* | | | | | | | | | | |
| MAGNETIC RESONANCE IMAGING HEAD | 6 | 6 | 0 | 0 | 0 | 6 | 6 | 0 | 0 | 0 |
| SCAN BRAIN | 0 | 0 | 0 | 0 | 0 | 0 | 0 | 0 | 0 | 0 |
| ***PHYSICAL EXAMINATION AND ORGAN SYSTEM STATUS TOPICS*** | | | | | | | | | | |
| *PHYSICAL EXAMINATION PROCEDURES AND ORGAN SYSTEM STATUS* | | | | | | | | | | |
| BODY TEMPERATURE | 49 | 42 | 7 | 0 | 0 | 47 | 42 | 5 | 0 | 0 |
| BODY TEMPERATURE ABNORMAL | 1 | 1 | 0 | 0 | 0 | 1 | 1 | 0 | 0 | 0 |
| BODY TEMPERATURE DECREASED | 2 | 2 | 0 | 0 | 0 | 2 | 2 | 0 | 0 | 0 |
| BODY TEMPERATURE FLUCTUATION | 8 | 7 | 1 | 0 | 0 | 8 | 7 | 1 | 0 | 0 |
| BODY TEMPERATURE INCREASED | 45 | 36 | 7 | 1 | 1 | 42 | 36 | 5 | 1 | 0 |
| GRIP STRENGTH DECREASED | 1 | 1 | 0 | 0 | 0 | 1 | 1 | 0 | 0 | 0 |
| HEAD LAG | 1 | 1 | 0 | 0 | 0 | 1 | 1 | 0 | 0 | 0 |
| LEFT-HANDEDNESS | 0 | 0 | 0 | 0 | 0 | 0 | 0 | 0 | 0 | 0 |
| LYMPH NODE PALPABLE | 0 | 0 | 0 | 0 | 0 | 0 | 0 | 0 | 0 | 0 |
| RESPIRATORY RATE DECREASED | 0 | 0 | 0 | 0 | 0 | 0 | 0 | 0 | 0 | 0 |
| SKIN TEMPERATURE | 4 | 3 | 1 | 0 | 0 | 4 | 3 | 1 | 0 | 0 |
| WEIGHT DECREASED | 1 | 1 | 0 | 0 | 0 | 1 | 1 | 0 | 0 | 0 |
| WEIGHT INCREASED | 2 | 2 | 0 | 0 | 0 | 2 | 2 | 0 | 0 | 0 |
| ***RENAL AND URINARY TRACT INVESTIGATIONS AND URINALYSES*** | | | | | | | | | | |
| *URINALYSIS NEC* | | | | | | | | | | |
| BLOOD URINE | 2 | 2 | 0 | 0 | 0 | 2 | 2 | 0 | 0 | 0 |

| | | | | | | | | | | |
|---|---|---|---|---|---|---|---|---|---|---|
| NITRITE URINE PRESENT | 1 | 1 | 0 | 0 | 0 | 1 | 1 | 0 | 0 | 0 |
| PH URINE | 0 | 0 | 0 | 0 | 0 | 0 | 0 | 0 | 0 | 0 |
| *URINARY TRACT FUNCTION ANALYSES NEC* | | | | | | | | | | |
| URINE OUTPUT | 2 | 2 | 0 | 0 | 0 | 2 | 2 | 0 | 0 | 0 |
| URINE OUTPUT INCREASED | 1 | 1 | 0 | 0 | 0 | 1 | 1 | 0 | 0 | 0 |
| ***REPRODUCTIVE ORGAN AND BREAST INVESTIGATIONS (EXCL HORMONE ANALYSES)*** | | | | | | | | | | |
| *REPRODUCTIVE ORGAN AND BREAST IMAGING PROCEDURES* | | | | | | | | | | |
| BREAST SCAN | 0 | 0 | 0 | 0 | 0 | 0 | 0 | 0 | 0 | 0 |
| ***RESPIRATORY AND PULMONARY INVESTIGATIONS (EXCL BLOOD GASES)*** | | | | | | | | | | |
| *RESPIRATORY AND PULMONARY FUNCTION DIAGNOSTIC PROCEDURES* | | | | | | | | | | |
| FORCED EXPIRATORY VOLUME DECREASED | 1 | 1 | 0 | 0 | 0 | 1 | 1 | 0 | 0 | 0 |
| FORCED EXPIRATORY VOLUME INCREASED | 0 | 0 | 0 | 0 | 0 | 0 | 0 | 0 | 0 | 0 |
| ***WATER, ELECTROLYTE AND MINERAL INVESTIGATIONS*** | | | | | | | | | | |
| *WATER AND ELECTROLYTE ANALYSES NEC* | | | | | | | | | | |
| VOLUME BLOOD | 0 | 0 | 0 | 0 | 0 | 0 | 0 | 0 | 0 | 0 |
| **METABOLISM AND NUTRITION DISORDERS** | | | | | | | | | | |
| ***APPETITE AND GENERAL NUTRITIONAL DISORDERS*** | | | | | | | | | | |
| *APPETITE DISORDERS* | | | | | | | | | | |
| APPETITE DISORDER | 0 | 0 | 0 | 0 | 0 | 0 | 0 | 0 | 0 | 0 |
| DECREASED APPETITE | 108 | 97 | 11 | 0 | 0 | 102 | 97 | 5 | 0 | 0 |
| FOOD CRAVING | 0 | 0 | 0 | 0 | 0 | 0 | 0 | 0 | 0 | 0 |
| FOOD REFUSAL | 1 | 1 | 0 | 0 | 0 | 1 | 1 | 0 | 0 | 0 |
| INCREASED APPETITE | 1 | 1 | 0 | 0 | 0 | 1 | 1 | 0 | 0 | 0 |
| *GENERAL NUTRITIONAL DISORDERS NEC* | | | | | | | | | | |
| FOOD AVERSION | 2 | 2 | 0 | 0 | 0 | 2 | 2 | 0 | 0 | 0 |
| ***ELECTROLYTE AND FLUID BALANCE CONDITIONS*** | | | | | | | | | | |
| *TOTAL FLUID VOLUME DECREASED* | | | | | | | | | | |
| DEHYDRATION | 13 | 12 | 1 | 0 | 0 | 12 | 12 | 0 | 0 | 0 |
| *TOTAL FLUID VOLUME INCREASED* | | | | | | | | | | |
| FLUID RETENTION | 1 | 0 | 0 | 1 | 0 | 0 | 0 | 0 | 0 | 0 |
| ***FOOD INTOLERANCE SYNDROMES*** | | | | | | | | | | |
| *FOOD MALABSORPTION AND INTOLERANCE SYNDROMES (EXCL SUGAR INTOLERANCE)* | | | | | | | | | | |
| ALCOHOL INTOLERANCE | 1 | 1 | 0 | 0 | 0 | 1 | 1 | 0 | 0 | 0 |
| ***GLUCOSE METABOLISM DISORDERS (INCL DIABETES MELLITUS)*** | | | | | | | | | | |
| *DIABETES MELLITUS (INCL SUBTYPES)* | | | | | | | | | | |
| DIABETES MELLITUS | 0 | 0 | 0 | 0 | 0 | 0 | 0 | 0 | 0 | 0 |

| | | | | | | | | | | |
|---|---|---|---|---|---|---|---|---|---|---|
| DIABETES MELLITUS INADEQUATE CONTROL | 1 | 1 | 0 | 0 | 0 | 1 | 1 | 0 | 0 | 0 |
| *HYPERGLYCAEMIC CONDITIONS NEC* | | | | | | | | | | |
| HYPERGLYCAEMIA | 0 | 0 | 0 | 0 | 0 | 0 | 0 | 0 | 0 | 0 |
| *HYPOGLYCAEMIC CONDITIONS NEC* | | | | | | | | | | |
| HYPOGLYCAEMIA | 2 | 2 | 0 | 0 | 0 | 2 | 2 | 0 | 0 | 0 |
| ***PURINE AND PYRIMIDINE METABOLISM DISORDERS*** | | | | | | | | | | |
| *DISORDERS OF PURINE METABOLISM* | | | | | | | | | | |
| GOUT | 1 | 1 | 0 | 0 | 0 | 1 | 1 | 0 | 0 | 0 |
| **MUSCULOSKELETAL AND CONNECTIVE TISSUE DISORDERS** | | | | | | | | | | |
| ***BONE DISORDERS (EXCL CONGENITAL AND FRACTURES)*** | | | | | | | | | | |
| *BONE DISORDERS NEC* | | | | | | | | | | |
| JAW DISORDER | 1 | 1 | 0 | 0 | 0 | 1 | 1 | 0 | 0 | 0 |
| OSTEITIS | 1 | 1 | 0 | 0 | 0 | 1 | 1 | 0 | 0 | 0 |
| *BONE RELATED SIGNS AND SYMPTOMS* | | | | | | | | | | |
| BONE PAIN | 12 | 11 | 1 | 0 | 0 | 11 | 11 | 0 | 0 | 0 |
| BONE SWELLING | 0 | 0 | 0 | 0 | 0 | 0 | 0 | 0 | 0 | 0 |
| PAIN IN JAW | 10 | 7 | 3 | 0 | 0 | 10 | 7 | 3 | 0 | 0 |
| PUBIC PAIN | 1 | 1 | 0 | 0 | 0 | 1 | 1 | 0 | 0 | 0 |
| SPINAL PAIN | 2 | 2 | 0 | 0 | 0 | 2 | 2 | 0 | 0 | 0 |
| ***CONNECTIVE TISSUE DISORDERS (EXCL CONGENITAL)*** | | | | | | | | | | |
| *CONNECTIVE TISSUE DISORDERS NEC* | | | | | | | | | | |
| POLYMYALGIA RHEUMATICA | 2 | 1 | 1 | 0 | 0 | 2 | 1 | 1 | 0 | 0 |
| *LUPUS ERYTHEMATOSUS (INCL SUBTYPES)* | | | | | | | | | | |
| SYSTEMIC LUPUS ERYTHEMATOSUS | 1 | 1 | 0 | 0 | 0 | 1 | 1 | 0 | 0 | 0 |
| ***JOINT DISORDERS*** | | | | | | | | | | |
| *ARTHROPATHIES NEC* | | | | | | | | | | |
| ARTHRITIS | 7 | 6 | 1 | 0 | 0 | 7 | 6 | 1 | 0 | 0 |
| ARTHROPATHY | 1 | 1 | 0 | 0 | 0 | 1 | 1 | 0 | 0 | 0 |
| POLYARTHRITIS | 0 | 0 | 0 | 0 | 0 | 0 | 0 | 0 | 0 | 0 |
| RHEUMATIC FEVER | 1 | 1 | 0 | 0 | 0 | 1 | 1 | 0 | 0 | 0 |
| *JOINT RELATED DISORDERS NEC* | | | | | | | | | | |
| JOINT LOCK | 1 | 1 | 0 | 0 | 0 | 1 | 1 | 0 | 0 | 0 |
| PERIARTHRITIS | 3 | 1 | 2 | 0 | 0 | 3 | 1 | 2 | 0 | 0 |
| TEMPOROMANDIBULAR PAIN AND DYSFUNCTION SYNDROME | 0 | 0 | 0 | 0 | 0 | 0 | 0 | 0 | 0 | 0 |
| *JOINT RELATED SIGNS AND SYMPTOMS* | | | | | | | | | | |

| | | | | | | | | | | |
|---|---|---|---|---|---|---|---|---|---|---|
| ARTHRALGIA | 892 | 760 | 131 | 1 | 0 | 852 | 760 | 91 | 1 | 0 |
| JAW CLICKING | 1 | 1 | 0 | 0 | 0 | 1 | 1 | 0 | 0 | 0 |
| JOINT STIFFNESS | 25 | 21 | 4 | 0 | 0 | 23 | 21 | 2 | 0 | 0 |
| JOINT SWELLING | 5 | 4 | 1 | 0 | 0 | 5 | 4 | 1 | 0 | 0 |
| *OSTEOARTHROPATHIES* | | | | | | | | | | |
| OSTEOARTHRITIS | 1 | 0 | 1 | 0 | 0 | 0 | 0 | 0 | 0 | 0 |
| *PSORIATIC ARTHROPATHIES* | | | | | | | | | | |
| PSORIATIC ARTHROPATHY | 1 | 1 | 0 | 0 | 0 | 1 | 1 | 0 | 0 | 0 |
| *RHEUMATOID ARTHROPATHIES* | | | | | | | | | | |
| RHEUMATOID ARTHRITIS | 2 | 2 | 0 | 0 | 0 | 2 | 2 | 0 | 0 | 0 |
| ***MUSCLE DISORDERS*** | | | | | | | | | | |
| *MUSCLE INFECTIONS AND INFLAMMATIONS* | | | | | | | | | | |
| MYOSITIS | 0 | 0 | 0 | 0 | 0 | 0 | 0 | 0 | 0 | 0 |
| *MUSCLE PAINS* | | | | | | | | | | |
| FIBROMYALGIA | 2 | 2 | 0 | 0 | 0 | 2 | 2 | 0 | 0 | 0 |
| MYALGIA | 1174 | 992 | 179 | 3 | 0 | 1117 | 992 | 123 | 2 | 0 |
| *MUSCLE RELATED SIGNS AND SYMPTOMS NEC* | | | | | | | | | | |
| MUSCLE ATROPHY | 1 | 1 | 0 | 0 | 0 | 1 | 1 | 0 | 0 | 0 |
| MUSCLE DISCOMFORT | 0 | 0 | 0 | 0 | 0 | 0 | 0 | 0 | 0 | 0 |
| MUSCLE FATIGUE | 31 | 27 | 4 | 0 | 0 | 30 | 27 | 3 | 0 | 0 |
| MUSCLE FIBROSIS | 0 | 0 | 0 | 0 | 0 | 0 | 0 | 0 | 0 | 0 |
| MUSCLE MASS | 3 | 3 | 0 | 0 | 0 | 3 | 3 | 0 | 0 | 0 |
| MUSCLE SPASMS | 36 | 27 | 8 | 1 | 0 | 32 | 27 | 5 | 0 | 0 |
| MUSCLE TIGHTNESS | 3 | 2 | 1 | 0 | 0 | 3 | 2 | 1 | 0 | 0 |
| MUSCLE TWITCHING | 8 | 8 | 0 | 0 | 0 | 8 | 8 | 0 | 0 | 0 |
| *MUSCLE TONE ABNORMALITIES* | | | | | | | | | | |
| MUSCLE RIGIDITY | 0 | 0 | 0 | 0 | 0 | 0 | 0 | 0 | 0 | 0 |
| TRISMUS | 1 | 1 | 0 | 0 | 0 | 1 | 1 | 0 | 0 | 0 |
| *MUSCLE WEAKNESS CONDITIONS* | | | | | | | | | | |
| MUSCULAR WEAKNESS | 36 | 32 | 4 | 0 | 0 | 36 | 32 | 4 | 0 | 0 |
| ***MUSCULOSKELETAL AND CONNECTIVE TISSUE DISORDERS NEC*** | | | | | | | | | | |
| *MUSCULOSKELETAL AND CONNECTIVE TISSUE CONDITIONS NEC* | | | | | | | | | | |
| MOBILITY DECREASED | 1 | 1 | 0 | 0 | 0 | 1 | 1 | 0 | 0 | 0 |
| MUSCULOSKELETAL STIFFNESS | 123 | 101 | 22 | 0 | 0 | 117 | 101 | 16 | 0 | 0 |
| BACK PAIN | 128 | 112 | 16 | 0 | 0 | 122 | 112 | 10 | 0 | 0 |

| | | | | | | | | | | |
|---|---|---|---|---|---|---|---|---|---|---|
| FLANK PAIN | 1 | 1 | 0 | 0 | 0 | 1 | 1 | 0 | 0 | 0 |
| LIMB DISCOMFORT | 307 | 235 | 71 | 1 | 0 | 294 | 235 | 59 | 0 | 0 |
| MUSCULOSKELETAL CHEST PAIN | 6 | 4 | 2 | 0 | 0 | 6 | 4 | 2 | 0 | 0 |
| MUSCULOSKELETAL DISCOMFORT | 12 | 10 | 2 | 0 | 0 | 12 | 10 | 2 | 0 | 0 |
| MUSCULOSKELETAL PAIN | 4 | 1 | 3 | 0 | 0 | 4 | 1 | 3 | 0 | 0 |
| NECK PAIN | 72 | 58 | 14 | 0 | 0 | 71 | 58 | 13 | 0 | 0 |
| PAIN IN EXTREMITY | 1509 | 1200 | 306 | 3 | 0 | 1427 | 1200 | 225 | 2 | 0 |
| *SOFT TISSUE DISORDERS NEC* | | | | | | | | | | |
| AXILLARY MASS | 0 | 0 | 0 | 0 | 0 | 0 | 0 | 0 | 0 | 0 |
| GROIN PAIN | 3 | 3 | 0 | 0 | 0 | 3 | 3 | 0 | 0 | 0 |
| ***SYNOVIAL AND BURSAL DISORDERS*** | | | | | | | | | | |
| *SYNOVIAL DISORDERS* | | | | | | | | | | |
| SYNOVITIS | 0 | 0 | 0 | 0 | 0 | 0 | 0 | 0 | 0 | 0 |
| ***TENDON, LIGAMENT AND CARTILAGE DISORDERS*** | | | | | | | | | | |
| *CARTILAGE DISORDERS* | | | | | | | | | | |
| COSTOCHONDRITIS | 2 | 2 | 0 | 0 | 0 | 2 | 2 | 0 | 0 | 0 |
| *TENDON DISORDERS* | | | | | | | | | | |
| TENDONITIS | 1 | 1 | 0 | 0 | 0 | 1 | 1 | 0 | 0 | 0 |
| TENOSYNOVITIS | 0 | 0 | 0 | 0 | 0 | 0 | 0 | 0 | 0 | 0 |
| TRIGGER FINGER | 0 | 0 | 0 | 0 | 0 | 0 | 0 | 0 | 0 | 0 |
| **NEOPLASMS BENIGN, MALIGNANT AND UNSPECIFIED (INCL CYSTS AND POLYPS)** | | | | | | | | | | |
| ***CUTANEOUS NEOPLASMS BENIGN*** | | | | | | | | | | |
| *SKIN NEOPLASMS BENIGN* | | | | | | | | | | |
| MELANOCYTIC NAEVUS | 0 | 0 | 0 | 0 | 0 | 0 | 0 | 0 | 0 | 0 |
| SEBORRHOEIC KERATOSIS | 0 | 0 | 0 | 0 | 0 | 0 | 0 | 0 | 0 | 0 |
| SKIN PAPILLOMA | 0 | 0 | 0 | 0 | 0 | 0 | 0 | 0 | 0 | 0 |
| **NERVOUS SYSTEM DISORDERS** | | | | | | | | | | |
| ***CENTRAL NERVOUS SYSTEM INFECTIONS AND INFLAMMATIONS*** | | | | | | | | | | |
| *MYELITIS (INCL INFECTIVE)* | | | | | | | | | | |
| MYELITIS TRANSVERSE | 1 | 1 | 0 | 0 | 0 | 1 | 1 | 0 | 0 | 0 |
| ***CENTRAL NERVOUS SYSTEM VASCULAR DISORDERS*** | | | | | | | | | | |
| *CENTRAL NERVOUS SYSTEM HAEMORRHAGES AND CEREBROVASCULAR ACCIDENTS* | | | | | | | | | | |
| CEREBRAL HAEMORRHAGE | 1 | 0 | 1 | 0 | 0 | 1 | 0 | 1 | 0 | 0 |
| CEREBROVASCULAR ACCIDENT | 5 | 4 | 1 | 0 | 0 | 5 | 4 | 1 | 0 | 0 |
| *TRANSIENT CEREBROVASCULAR EVENTS* | | | | | | | | | | |
| TRANSIENT ISCHAEMIC ATTACK | 5 | 4 | 1 | 0 | 0 | 4 | 4 | 0 | 0 | 0 |

| | | | | | | | | | | |
|---|---|---|---|---|---|---|---|---|---|---|
| ***CRANIAL NERVE DISORDERS (EXCL NEOPLASMS)*** | | | | | | | | | | |
| *FACIAL CRANIAL NERVE DISORDERS* | | | | | | | | | | |
| BELL'S PALSY | 1 | 1 | 0 | 0 | 0 | 1 | 1 | 0 | 0 | 0 |
| FACIAL PARALYSIS | 0 | 0 | 0 | 0 | 0 | 0 | 0 | 0 | 0 | 0 |
| FACIAL PARESIS | 0 | 0 | 0 | 0 | 0 | 0 | 0 | 0 | 0 | 0 |
| *OLFACTORY NERVE DISORDERS* | | | | | | | | | | |
| ANOSMIA | 6 | 6 | 0 | 0 | 0 | 6 | 6 | 0 | 0 | 0 |
| HYPOSMIA | 1 | 1 | 0 | 0 | 0 | 1 | 1 | 0 | 0 | 0 |
| PAROSMIA | 10 | 5 | 5 | 0 | 0 | 9 | 5 | 4 | 0 | 0 |
| *TRIGEMINAL DISORDERS* | | | | | | | | | | |
| TRIGEMINAL NEURALGIA | 2 | 2 | 0 | 0 | 0 | 2 | 2 | 0 | 0 | 0 |
| TRIGEMINAL NEURITIS | 0 | 0 | 0 | 0 | 0 | 0 | 0 | 0 | 0 | 0 |
| ***DEMYELINATING DISORDERS*** | | | | | | | | | | |
| *MULTIPLE SCLEROSIS ACUTE AND PROGRESSIVE* | | | | | | | | | | |
| MULTIPLE SCLEROSIS RELAPSE | 1 | 1 | 0 | 0 | 0 | 1 | 1 | 0 | 0 | 0 |
| ***HEADACHES*** | | | | | | | | | | |
| *HEADACHES NEC* | | | | | | | | | | |
| CLUSTER HEADACHE | 19 | 19 | 0 | 0 | 0 | 19 | 19 | 0 | 0 | 0 |
| COLD-STIMULUS HEADACHE | 1 | 1 | 0 | 0 | 0 | 1 | 1 | 0 | 0 | 0 |
| DRUG WITHDRAWAL HEADACHE | 1 | 0 | 1 | 0 | 0 | 1 | 0 | 1 | 0 | 0 |
| HEADACHE | 3605 | 3063 | 541 | 1 | 0 | 3414 | 3063 | 351 | 0 | 0 |
| PRIMARY STABBING HEADACHE | 2 | 2 | 0 | 0 | 0 | 2 | 2 | 0 | 0 | 0 |
| SINUS HEADACHE | 42 | 37 | 5 | 0 | 0 | 40 | 37 | 3 | 0 | 0 |
| TENSION HEADACHE | 82 | 66 | 16 | 0 | 0 | 78 | 66 | 12 | 0 | 0 |
| THUNDERCLAP HEADACHE | 1 | 1 | 0 | 0 | 0 | 1 | 1 | 0 | 0 | 0 |
| VASCULAR HEADACHE | 1 | 1 | 0 | 0 | 0 | 1 | 1 | 0 | 0 | 0 |
| *MIGRAINE HEADACHES* | | | | | | | | | | |
| MIGRAINE | 94 | 81 | 13 | 0 | 0 | 88 | 81 | 7 | 0 | 0 |
| MIGRAINE WITH AURA | 3 | 1 | 2 | 0 | 0 | 2 | 1 | 1 | 0 | 0 |
| RETINAL MIGRAINE | 2 | 2 | 0 | 0 | 0 | 2 | 2 | 0 | 0 | 0 |
| TYPICAL AURA WITHOUT HEADACHE | 0 | 0 | 0 | 0 | 0 | 0 | 0 | 0 | 0 | 0 |
| ***MENTAL IMPAIRMENT DISORDERS*** | | | | | | | | | | |
| *INTELLECTUAL DISABILITIES* | | | | | | | | | | |
| INTELLECTUAL DISABILITY | 0 | 0 | 0 | 0 | 0 | 0 | 0 | 0 | 0 | 0 |
| *MEMORY LOSS (EXCL DEMENTIA)* | | | | | | | | | | |

| | | | | | | | | | | |
|---|---|---|---|---|---|---|---|---|---|---|
| AMNESIA | 3 | 2 | 1 | 0 | 0 | 3 | 2 | 1 | 0 | 0 |
| MEMORY IMPAIRMENT | 6 | 5 | 1 | 0 | 0 | 6 | 5 | 1 | 0 | 0 |
| *MENTAL IMPAIRMENT (EXCL DEMENTIA AND MEMORY LOSS)* | | | | | | | | | | |
| COGNITIVE DISORDER | 1 | 1 | 0 | 0 | 0 | 1 | 1 | 0 | 0 | 0 |
| DISTURBANCE IN ATTENTION | 12 | 11 | 1 | 0 | 0 | 12 | 11 | 1 | 0 | 0 |
| MENTAL IMPAIRMENT | 1 | 1 | 0 | 0 | 0 | 1 | 1 | 0 | 0 | 0 |
| ***MOVEMENT DISORDERS (INCL PARKINSONISM)*** | | | | | | | | | | |
| *DYSKINESIAS AND MOVEMENT DISORDERS NEC* | | | | | | | | | | |
| BRADYKINESIA | 0 | 0 | 0 | 0 | 0 | 0 | 0 | 0 | 0 | 0 |
| CLUMSINESS | 1 | 1 | 0 | 0 | 0 | 1 | 1 | 0 | 0 | 0 |
| DYSKINESIA | 3 | 3 | 0 | 0 | 0 | 3 | 3 | 0 | 0 | 0 |
| EXTRAPYRAMIDAL DISORDER | 1 | 1 | 0 | 0 | 0 | 1 | 1 | 0 | 0 | 0 |
| PSYCHOMOTOR HYPERACTIVITY | 1 | 0 | 1 | 0 | 0 | 1 | 0 | 1 | 0 | 0 |
| *PARALYSIS AND PARESIS (EXCL CRANIAL NERVE)* | | | | | | | | | | |
| HEMIPLEGIA | 0 | 0 | 0 | 0 | 0 | 0 | 0 | 0 | 0 | 0 |
| MONOPARESIS | 0 | 0 | 0 | 0 | 0 | 0 | 0 | 0 | 0 | 0 |
| MONOPLEGIA | 1 | 1 | 0 | 0 | 0 | 1 | 1 | 0 | 0 | 0 |
| PARALYSIS | 1 | 1 | 0 | 0 | 0 | 1 | 1 | 0 | 0 | 0 |
| *PARKINSON'S DISEASE AND PARKINSONISM* | | | | | | | | | | |
| FREEZING PHENOMENON | 2 | 2 | 0 | 0 | 0 | 2 | 2 | 0 | 0 | 0 |
| PARKINSON'S DISEASE | 0 | 0 | 0 | 0 | 0 | 0 | 0 | 0 | 0 | 0 |
| *TREMOR (EXCL CONGENITAL)* | | | | | | | | | | |
| TREMOR | 111 | 100 | 11 | 0 | 0 | 105 | 100 | 5 | 0 | 0 |
| ***NEUROLOGICAL DISORDERS NEC*** | | | | | | | | | | |
| *COMA STATES* | | | | | | | | | | |
| DIABETIC HYPERGLYCAEMIC COMA | 0 | 0 | 0 | 0 | 0 | 0 | 0 | 0 | 0 | 0 |
| *COORDINATION AND BALANCE DISTURBANCES* | | | | | | | | | | |
| BALANCE DISORDER | 16 | 14 | 2 | 0 | 0 | 16 | 14 | 2 | 0 | 0 |
| COORDINATION ABNORMAL | 1 | 1 | 0 | 0 | 0 | 1 | 1 | 0 | 0 | 0 |
| DYSSTASIA | 0 | 0 | 0 | 0 | 0 | 0 | 0 | 0 | 0 | 0 |
| VESTIBULAR NYSTAGMUS | 1 | 1 | 0 | 0 | 0 | 1 | 1 | 0 | 0 | 0 |
| *DISTURBANCES IN CONSCIOUSNESS NEC* | | | | | | | | | | |
| DEPRESSED LEVEL OF CONSCIOUSNESS | 0 | 0 | 0 | 0 | 0 | 0 | 0 | 0 | 0 | 0 |
| LETHARGY | 204 | 165 | 39 | 0 | 0 | 194 | 165 | 29 | 0 | 0 |
| LOSS OF CONSCIOUSNESS | 7 | 6 | 1 | 0 | 0 | 7 | 6 | 1 | 0 | 0 |

| | | | | | | | | | | |
|---|---|---|---|---|---|---|---|---|---|---|
| SEDATION | 0 | 0 | 0 | 0 | 0 | 0 | 0 | 0 | 0 | 0 |
| SOMNOLENCE | 86 | 72 | 14 | 0 | 0 | 84 | 72 | 12 | 0 | 0 |
| SYNCOPE | 20 | 14 | 6 | 0 | 0 | 16 | 14 | 2 | 0 | 0 |
| *NERVOUS SYSTEM DISORDERS NEC* | | | | | | | | | | |
| NERVOUS SYSTEM DISORDER | 0 | 0 | 0 | 0 | 0 | 0 | 0 | 0 | 0 | 0 |
| *NEUROLOGICAL SIGNS AND SYMPTOMS NEC* | | | | | | | | | | |
| AGITATION NEONATAL | 0 | 0 | 0 | 0 | 0 | 0 | 0 | 0 | 0 | 0 |
| BRAIN FOG | 20 | 15 | 5 | 0 | 0 | 19 | 15 | 4 | 0 | 0 |
| DIZZINESS | 401 | 319 | 82 | 0 | 0 | 382 | 319 | 63 | 0 | 0 |
| DIZZINESS EXERTIONAL | 5 | 4 | 1 | 0 | 0 | 5 | 4 | 1 | 0 | 0 |
| DIZZINESS POSTURAL | 61 | 49 | 12 | 0 | 0 | 58 | 49 | 9 | 0 | 0 |
| HEAD DISCOMFORT | 10 | 8 | 2 | 0 | 0 | 10 | 8 | 2 | 0 | 0 |
| INFANT IRRITABILITY | 1 | 0 | 1 | 0 | 0 | 1 | 0 | 1 | 0 | 0 |
| MENINGISM | 1 | 1 | 0 | 0 | 0 | 1 | 1 | 0 | 0 | 0 |
| MYOCLONUS | 0 | 0 | 0 | 0 | 0 | 0 | 0 | 0 | 0 | 0 |
| NEUROLOGICAL SYMPTOM | 1 | 1 | 0 | 0 | 0 | 1 | 1 | 0 | 0 | 0 |
| PERSISTENT POSTURAL-PERCEPTUAL DIZZINESS | 1 | 0 | 1 | 0 | 0 | 0 | 0 | 0 | 0 | 0 |
| PRESYNCOPE | 7 | 7 | 0 | 0 | 0 | 7 | 7 | 0 | 0 | 0 |
| *PARAESTHESIAS AND DYSAESTHESIAS* | | | | | | | | | | |
| BURNING FEET SYNDROME | 0 | 0 | 0 | 0 | 0 | 0 | 0 | 0 | 0 | 0 |
| BURNING SENSATION | 3 | 3 | 0 | 0 | 0 | 3 | 3 | 0 | 0 | 0 |
| FORMICATION | 0 | 0 | 0 | 0 | 0 | 0 | 0 | 0 | 0 | 0 |
| HYPERAESTHESIA | 1 | 1 | 0 | 0 | 0 | 1 | 1 | 0 | 0 | 0 |
| HYPOAESTHESIA | 62 | 57 | 5 | 0 | 0 | 59 | 57 | 2 | 0 | 0 |
| PARAESTHESIA | 105 | 89 | 16 | 0 | 0 | 101 | 89 | 12 | 0 | 0 |
| REVERSED HOT-COLD SENSATION | 1 | 1 | 0 | 0 | 0 | 1 | 1 | 0 | 0 | 0 |
| *SENSORY ABNORMALITIES NEC* | | | | | | | | | | |
| AGEUSIA | 19 | 16 | 3 | 0 | 0 | 18 | 16 | 2 | 0 | 0 |
| ALLODYNIA | 1 | 1 | 0 | 0 | 0 | 1 | 1 | 0 | 0 | 0 |
| DYSGEUSIA | 49 | 40 | 9 | 0 | 0 | 47 | 40 | 7 | 0 | 0 |
| HYPOGEUSIA | 1 | 1 | 0 | 0 | 0 | 1 | 1 | 0 | 0 | 0 |
| NEURALGIA | 17 | 14 | 3 | 0 | 0 | 15 | 14 | 1 | 0 | 0 |
| POST HERPETIC NEURALGIA | 0 | 0 | 0 | 0 | 0 | 0 | 0 | 0 | 0 | 0 |
| RESTLESS ARM SYNDROME | 1 | 1 | 0 | 0 | 0 | 1 | 1 | 0 | 0 | 0 |
| RESTLESS LEGS SYNDROME | 4 | 4 | 0 | 0 | 0 | 4 | 4 | 0 | 0 | 0 |

| | | | | | | | | | | |
|---|---|---|---|---|---|---|---|---|---|---|
| SENSORY LOSS | 1 | 1 | 0 | 0 | 0 | 1 | 1 | 0 | 0 | 0 |
| TASTE DISORDER | 8 | 7 | 1 | 0 | 0 | 8 | 7 | 1 | 0 | 0 |
| *SPEECH AND LANGUAGE ABNORMALITIES* | | | | | | | | | | |
| DYSARTHRIA | 0 | 0 | 0 | 0 | 0 | 0 | 0 | 0 | 0 | 0 |
| SPEECH DISORDER DEVELOPMENTAL | 0 | 0 | 0 | 0 | 0 | 0 | 0 | 0 | 0 | 0 |
| ***NEUROLOGICAL DISORDERS OF THE EYE*** | | | | | | | | | | |
| *NEUROLOGIC VISUAL PROBLEMS NEC* | | | | | | | | | | |
| TUNNEL VISION | 2 | 1 | 1 | 0 | 0 | 2 | 1 | 1 | 0 | 0 |
| ***NEUROMUSCULAR DISORDERS*** | | | | | | | | | | |
| *MUSCLE TONE ABNORMAL* | | | | | | | | | | |
| HYPOTONIA | 1 | 1 | 0 | 0 | 0 | 1 | 1 | 0 | 0 | 0 |
| STIFF LEG SYNDROME | 1 | 0 | 1 | 0 | 0 | 1 | 0 | 1 | 0 | 0 |
| *NEUROMUSCULAR DISORDERS NEC* | | | | | | | | | | |
| MUSCLE SPASTICITY | 1 | 1 | 0 | 0 | 0 | 1 | 1 | 0 | 0 | 0 |
| ***PERIPHERAL NEUROPATHIES*** | | | | | | | | | | |
| *ACUTE POLYNEUROPATHIES* | | | | | | | | | | |
| GUILLAIN-BARRE SYNDROME | 2 | 2 | 0 | 0 | 0 | 2 | 2 | 0 | 0 | 0 |
| *PERIPHERAL NEUROPATHIES NEC* | | | | | | | | | | |
| AXONAL NEUROPATHY | 0 | 0 | 0 | 0 | 0 | 0 | 0 | 0 | 0 | 0 |
| NEUROPATHY PERIPHERAL | 1 | 1 | 0 | 0 | 0 | 1 | 1 | 0 | 0 | 0 |
| ***SEIZURES (INCL SUBTYPES)*** | | | | | | | | | | |
| *SEIZURES AND SEIZURE DISORDERS NEC* | | | | | | | | | | |
| EPILEPSY | 0 | 0 | 0 | 0 | 0 | 0 | 0 | 0 | 0 | 0 |
| SEIZURE | 7 | 6 | 1 | 0 | 0 | 7 | 6 | 1 | 0 | 0 |
| ***SLEEP DISTURBANCES (INCL SUBTYPES)*** | | | | | | | | | | |
| *DISTURBANCES IN SLEEP PHASE RHYTHM* | | | | | | | | | | |
| CIRCADIAN RHYTHM SLEEP DISORDER | 1 | 1 | 0 | 0 | 0 | 1 | 1 | 0 | 0 | 0 |
| *SLEEP DISTURBANCES NEC* | | | | | | | | | | |
| SLEEP DEFICIT | 1 | 1 | 0 | 0 | 0 | 1 | 1 | 0 | 0 | 0 |
| ***SPINAL CORD AND NERVE ROOT DISORDERS*** | | | | | | | | | | |
| *CERVICAL SPINAL CORD AND NERVE ROOT DISORDERS* | | | | | | | | | | |
| CERVICOBRACHIAL SYNDROME | 0 | 0 | 0 | 0 | 0 | 0 | 0 | 0 | 0 | 0 |
| *LUMBAR SPINAL CORD AND NERVE ROOT DISORDERS* | | | | | | | | | | |
| CAUDA EQUINA SYNDROME | 0 | 0 | 0 | 0 | 0 | 0 | 0 | 0 | 0 | 0 |
| SCIATICA | 5 | 4 | 1 | 0 | 0 | 5 | 4 | 1 | 0 | 0 |
| **PREGNANCY, PUERPERIUM AND PERINATAL CONDITIONS** | | | | | | | | | | |

| | | | | | | | | | | |
|---|---|---|---|---|---|---|---|---|---|---|
| ***ABORTIONS AND STILLBIRTH*** | | | | | | | | | | |
| *ABORTIONS SPONTANEOUS* | | | | | | | | | | |
| ABORTION SPONTANEOUS | 7 | 4 | 3 | 0 | 0 | 5 | 4 | 1 | 0 | 0 |
| *STILLBIRTH AND FOETAL DEATH* | | | | | | | | | | |
| FOETAL DEATH | 1 | 1 | 0 | 0 | 0 | 1 | 1 | 0 | 0 | 0 |
| ***FOETAL COMPLICATIONS*** | | | | | | | | | | |
| *FOETAL COMPLICATIONS NEC* | | | | | | | | | | |
| FOETAL DISORDER | 0 | 0 | 0 | 0 | 0 | 0 | 0 | 0 | 0 | 0 |
| FOETAL HYPOKINESIA | 0 | 0 | 0 | 0 | 0 | 0 | 0 | 0 | 0 | 0 |
| *FOETAL GROWTH COMPLICATIONS* | | | | | | | | | | |
| FOETAL MACROSOMIA | 0 | 0 | 0 | 0 | 0 | 0 | 0 | 0 | 0 | 0 |
| ***MATERNAL COMPLICATIONS OF PREGNANCY*** | | | | | | | | | | |
| *MATERNAL COMPLICATIONS OF PREGNANCY NEC* | | | | | | | | | | |
| MORNING SICKNESS | 1 | 1 | 0 | 0 | 0 | 1 | 1 | 0 | 0 | 0 |
| ***PLACENTAL, AMNIOTIC AND CAVITY DISORDERS (EXCL HAEMORRHAGES)*** | | | | | | | | | | |
| *PLACENTAL ABNORMALITIES (EXCL NEOPLASMS)* | | | | | | | | | | |
| PLACENTAL INFARCTION | 1 | 1 | 0 | 0 | 0 | 1 | 1 | 0 | 0 | 0 |
| ***PREGNANCY, LABOUR, DELIVERY AND POSTPARTUM CONDITIONS*** | | | | | | | | | | |
| *NORMAL PREGNANCY, LABOUR AND DELIVERY* | | | | | | | | | | |
| PREGNANCY | 0 | 0 | 0 | 0 | 0 | 0 | 0 | 0 | 0 | 0 |
| UTERINE CONTRACTIONS DURING PREGNANCY | 0 | 0 | 0 | 0 | 0 | 0 | 0 | 0 | 0 | 0 |
| **PRODUCT ISSUES** | | | | | | | | | | |
| ***DEVICE ISSUES*** | | | | | | | | | | |
| *DEVICE MALFUNCTION EVENTS NEC* | | | | | | | | | | |
| OVERSENSING | 1 | 1 | 0 | 0 | 0 | 1 | 1 | 0 | 0 | 0 |
| **PSYCHIATRIC DISORDERS** | | | | | | | | | | |
| ***ANXIETY DISORDERS AND SYMPTOMS*** | | | | | | | | | | |
| *ANXIETY SYMPTOMS* | | | | | | | | | | |
| AGITATION | 1 | 1 | 0 | 0 | 0 | 1 | 1 | 0 | 0 | 0 |
| ANXIETY | 11 | 8 | 3 | 0 | 0 | 11 | 8 | 3 | 0 | 0 |
| NERVOUSNESS | 11 | 10 | 1 | 0 | 0 | 11 | 10 | 1 | 0 | 0 |
| TENSION | 5 | 4 | 1 | 0 | 0 | 4 | 4 | 0 | 0 | 0 |
| *PANIC ATTACKS AND DISORDERS* | | | | | | | | | | |
| PANIC ATTACK | 1 | 1 | 0 | 0 | 0 | 1 | 1 | 0 | 0 | 0 |
| ***CHANGES IN PHYSICAL ACTIVITY*** | | | | | | | | | | |
| *INCREASED PHYSICAL ACTIVITY LEVELS* | | | | | | | | | | |

| | | | | | | | | | | |
|---|---|---|---|---|---|---|---|---|---|---|
| RESTLESSNESS | 5 | 4 | 1 | 0 | 0 | 5 | 4 | 1 | 0 | 0 |
| *STEREOTYPIES AND AUTOMATISMS* | | | | | | | | | | |
| BRUXISM | 1 | 1 | 0 | 0 | 0 | 1 | 1 | 0 | 0 | 0 |
| ***COGNITIVE AND ATTENTION DISORDERS AND DISTURBANCES*** | | | | | | | | | | |
| *COGNITIVE AND ATTENTION DISORDERS AND DISTURBANCES NEC* | | | | | | | | | | |
| DAYDREAMING | 1 | 1 | 0 | 0 | 0 | 1 | 1 | 0 | 0 | 0 |
| MENTAL FATIGUE | 15 | 10 | 5 | 0 | 0 | 12 | 10 | 2 | 0 | 0 |
| ***COMMUNICATION DISORDERS AND DISTURBANCES*** | | | | | | | | | | |
| *SPEECH ARTICULATION AND RHYTHM DISTURBANCES* | | | | | | | | | | |
| DYSPHEMIA | 1 | 0 | 1 | 0 | 0 | 0 | 0 | 0 | 0 | 0 |
| ***DELIRIA (INCL CONFUSION)*** | | | | | | | | | | |
| *CONFUSION AND DISORIENTATION* | | | | | | | | | | |
| CONFUSIONAL STATE | 21 | 19 | 2 | 0 | 0 | 20 | 19 | 1 | 0 | 0 |
| DISORIENTATION | 6 | 5 | 1 | 0 | 0 | 6 | 5 | 1 | 0 | 0 |
| *DELIRIA* | | | | | | | | | | |
| DELIRIUM | 1 | 1 | 0 | 0 | 0 | 1 | 1 | 0 | 0 | 0 |
| ***DEPRESSED MOOD DISORDERS AND DISTURBANCES*** | | | | | | | | | | |
| *DEPRESSIVE DISORDERS* | | | | | | | | | | |
| DEPRESSION | 7 | 6 | 1 | 0 | 0 | 6 | 6 | 0 | 0 | 0 |
| *MOOD ALTERATIONS WITH DEPRESSIVE SYMPTOMS* | | | | | | | | | | |
| DEPRESSED MOOD | 12 | 8 | 4 | 0 | 0 | 12 | 8 | 4 | 0 | 0 |
| TEARFULNESS | 1 | 1 | 0 | 0 | 0 | 1 | 1 | 0 | 0 | 0 |
| ***DISSOCIATIVE DISORDERS*** | | | | | | | | | | |
| *DISSOCIATIVE STATES* | | | | | | | | | | |
| DISSOCIATION | 1 | 1 | 0 | 0 | 0 | 1 | 1 | 0 | 0 | 0 |
| ***DISTURBANCES IN THINKING AND PERCEPTION*** | | | | | | | | | | |
| *DELUSIONAL SYMPTOMS* | | | | | | | | | | |
| DELUSION | 0 | 0 | 0 | 0 | 0 | 0 | 0 | 0 | 0 | 0 |
| *HALLUCINATIONS (EXCL SLEEP-RELATED)* | | | | | | | | | | |
| HALLUCINATION | 2 | 2 | 0 | 0 | 0 | 2 | 2 | 0 | 0 | 0 |
| *PERCEPTION DISTURBANCES NEC* | | | | | | | | | | |
| DEREALISATION | 1 | 1 | 0 | 0 | 0 | 1 | 1 | 0 | 0 | 0 |
| *THINKING DISTURBANCES* | | | | | | | | | | |
| BRADYPHRENIA | 2 | 0 | 2 | 0 | 0 | 2 | 0 | 2 | 0 | 0 |
| THOUGHT BLOCKING | 1 | 1 | 0 | 0 | 0 | 1 | 1 | 0 | 0 | 0 |
| ***MOOD DISORDERS AND DISTURBANCES NEC*** | | | | | | | | | | |

| | | | | | | | | | | |
|---|---|---|---|---|---|---|---|---|---|---|
| *AFFECT ALTERATIONS NEC* | | | | | | | | | | |
| AFFECT LABILITY | 0 | 0 | 0 | 0 | 0 | 0 | 0 | 0 | 0 | 0 |
| INAPPROPRIATE AFFECT | 1 | 1 | 0 | 0 | 0 | 1 | 1 | 0 | 0 | 0 |
| *EMOTIONAL AND MOOD DISTURBANCES NEC* | | | | | | | | | | |
| ANGER | 1 | 1 | 0 | 0 | 0 | 1 | 1 | 0 | 0 | 0 |
| EMOTIONAL DISORDER | 0 | 0 | 0 | 0 | 0 | 0 | 0 | 0 | 0 | 0 |
| EMOTIONAL DISTRESS | 1 | 1 | 0 | 0 | 0 | 1 | 1 | 0 | 0 | 0 |
| EUPHORIC MOOD | 2 | 2 | 0 | 0 | 0 | 2 | 2 | 0 | 0 | 0 |
| IRRITABILITY | 8 | 8 | 0 | 0 | 0 | 8 | 8 | 0 | 0 | 0 |
| MOOD ALTERED | 1 | 0 | 1 | 0 | 0 | 1 | 0 | 1 | 0 | 0 |
| *FLUCTUATING MOOD SYMPTOMS* | | | | | | | | | | |
| MOOD SWINGS | 2 | 1 | 1 | 0 | 0 | 1 | 1 | 0 | 0 | 0 |
| *MOOD DISORDERS NEC* | | | | | | | | | | |
| APATHY | 1 | 0 | 1 | 0 | 0 | 1 | 0 | 1 | 0 | 0 |
| LISTLESS | 2 | 1 | 1 | 0 | 0 | 2 | 1 | 1 | 0 | 0 |
| ***PERSONALITY DISORDERS AND DISTURBANCES IN BEHAVIOUR*** | | | | | | | | | | |
| *BEHAVIOUR AND SOCIALISATION DISTURBANCES* | | | | | | | | | | |
| AGGRESSION | 1 | 1 | 0 | 0 | 0 | 1 | 1 | 0 | 0 | 0 |
| ***PSYCHIATRIC AND BEHAVIOURAL SYMPTOMS NEC*** | | | | | | | | | | |
| *PSYCHIATRIC SYMPTOMS NEC* | | | | | | | | | | |
| PSYCHIATRIC SYMPTOM | 0 | 0 | 0 | 0 | 0 | 0 | 0 | 0 | 0 | 0 |
| ***PSYCHIATRIC DISORDERS NEC*** | | | | | | | | | | |
| *MENTAL DISORDERS NEC* | | | | | | | | | | |
| MENTAL DISORDER | 1 | 0 | 1 | 0 | 0 | 1 | 0 | 1 | 0 | 0 |
| ***SCHIZOPHRENIA AND OTHER PSYCHOTIC DISORDERS*** | | | | | | | | | | |
| *PSYCHOTIC DISORDER NEC* | | | | | | | | | | |
| PSYCHOTIC DISORDER | 1 | 1 | 0 | 0 | 0 | 1 | 1 | 0 | 0 | 0 |
| ***SEXUAL DYSFUNCTIONS, DISTURBANCES AND GENDER IDENTITY DISORDERS*** | | | | | | | | | | |
| *SEXUAL DESIRE DISORDERS* | | | | | | | | | | |
| LOSS OF LIBIDO | 2 | 2 | 0 | 0 | 0 | 2 | 2 | 0 | 0 | 0 |
| ***SLEEP DISORDERS AND DISTURBANCES*** | | | | | | | | | | |
| *DISTURBANCES IN INITIATING AND MAINTAINING SLEEP* | | | | | | | | | | |
| INITIAL INSOMNIA | 0 | 0 | 0 | 0 | 0 | 0 | 0 | 0 | 0 | 0 |
| INSOMNIA | 87 | 76 | 11 | 0 | 0 | 83 | 76 | 7 | 0 | 0 |
| *DYSSOMNIAS* | | | | | | | | | | |
| BREATHING-RELATED SLEEP DISORDER | 1 | 1 | 0 | 0 | 0 | 1 | 1 | 0 | 0 | 0 |

| | | | | | | | | | | |
|---|---|---|---|---|---|---|---|---|---|---|
| POOR QUALITY SLEEP | 28 | 25 | 3 | 0 | 0 | 27 | 25 | 2 | 0 | 0 |
| *PARASOMNIAS* | | | | | | | | | | |
| ABNORMAL DREAMS | 8 | 7 | 1 | 0 | 0 | 8 | 7 | 1 | 0 | 0 |
| EXPLODING HEAD SYNDROME | 0 | 0 | 0 | 0 | 0 | 0 | 0 | 0 | 0 | 0 |
| NIGHTMARE | 6 | 5 | 1 | 0 | 0 | 6 | 5 | 1 | 0 | 0 |
| *SLEEP DISORDERS NEC* | | | | | | | | | | |
| SLEEP DISORDER | 11 | 10 | 1 | 0 | 0 | 11 | 10 | 1 | 0 | 0 |
| ***SOMATIC SYMPTOM AND RELATED DISORDERS*** | | | | | | | | | | |
| *SOMATIC SYMPTOM DISORDERS* | | | | | | | | | | |
| HABIT COUGH | 3 | 3 | 0 | 0 | 0 | 3 | 3 | 0 | 0 | 0 |
| ***SUICIDAL AND SELF-INJURIOUS BEHAVIOURS NEC*** | | | | | | | | | | |
| *SUICIDAL AND SELF-INJURIOUS BEHAVIOUR* | | | | | | | | | | |
| SUICIDAL IDEATION | 0 | 0 | 0 | 0 | 0 | 0 | 0 | 0 | 0 | 0 |
| **RENAL AND URINARY DISORDERS** | | | | | | | | | | |
| ***RENAL DISORDERS (EXCL NEPHROPATHIES)*** | | | | | | | | | | |
| *RENAL FAILURE AND IMPAIRMENT* | | | | | | | | | | |
| RENAL FAILURE | 0 | 0 | 0 | 0 | 0 | 0 | 0 | 0 | 0 | 0 |
| ***URINARY TRACT SIGNS AND SYMPTOMS*** | | | | | | | | | | |
| *BLADDER AND URETHRAL SYMPTOMS* | | | | | | | | | | |
| BLADDER PAIN | 1 | 1 | 0 | 0 | 0 | 1 | 1 | 0 | 0 | 0 |
| MICTURITION URGENCY | 1 | 1 | 0 | 0 | 0 | 1 | 1 | 0 | 0 | 0 |
| POLLAKIURIA | 10 | 9 | 1 | 0 | 0 | 10 | 9 | 1 | 0 | 0 |
| URINARY INCONTINENCE | 1 | 0 | 1 | 0 | 0 | 0 | 0 | 0 | 0 | 0 |
| *URINARY ABNORMALITIES* | | | | | | | | | | |
| HAEMATURIA | 0 | 0 | 0 | 0 | 0 | 0 | 0 | 0 | 0 | 0 |
| URINE ABNORMALITY | 0 | 0 | 0 | 0 | 0 | 0 | 0 | 0 | 0 | 0 |
| URINE ODOUR ABNORMAL | 2 | 2 | 0 | 0 | 0 | 2 | 2 | 0 | 0 | 0 |
| *URINARY TRACT SIGNS AND SYMPTOMS NEC* | | | | | | | | | | |
| POLYURIA | 2 | 2 | 0 | 0 | 0 | 2 | 2 | 0 | 0 | 0 |
| RENAL PAIN | 7 | 5 | 2 | 0 | 0 | 7 | 5 | 2 | 0 | 0 |
| **REPRODUCTIVE SYSTEM AND BREAST DISORDERS** | | | | | | | | | | |
| ***BREAST DISORDERS*** | | | | | | | | | | |
| *BREAST DISORDERS NEC* | | | | | | | | | | |
| BREAST MASS | 2 | 2 | 0 | 0 | 0 | 2 | 2 | 0 | 0 | 0 |
| NIPPLE ENLARGEMENT | 1 | 1 | 0 | 0 | 0 | 1 | 1 | 0 | 0 | 0 |
| *BREAST SIGNS AND SYMPTOMS* | | | | | | | | | | |

| | | | | | | | | | | |
|---|---|---|---|---|---|---|---|---|---|---|
| BREAST DISCHARGE | 0 | 0 | 0 | 0 | 0 | 0 | 0 | 0 | 0 | 0 |
| BREAST PAIN | 8 | 6 | 2 | 0 | 0 | 8 | 6 | 2 | 0 | 0 |
| BREAST SWELLING | 1 | 1 | 0 | 0 | 0 | 1 | 1 | 0 | 0 | 0 |
| BREAST TENDERNESS | 1 | 1 | 0 | 0 | 0 | 1 | 1 | 0 | 0 | 0 |
| NIPPLE PAIN | 2 | 2 | 0 | 0 | 0 | 2 | 2 | 0 | 0 | 0 |
| ***MENOPAUSE RELATED CONDITIONS*** | | | | | | | | | | |
| *MENOPAUSAL EFFECTS ON THE GENITOURINARY TRACT* | | | | | | | | | | |
| POSTMENOPAUSAL HAEMORRHAGE | 1 | 1 | 0 | 0 | 0 | 1 | 1 | 0 | 0 | 0 |
| ***MENSTRUAL CYCLE AND UTERINE BLEEDING DISORDERS*** | | | | | | | | | | |
| *MENSTRUATION AND UTERINE BLEEDING NEC* | | | | | | | | | | |
| DYSMENORRHOEA | 5 | 4 | 1 | 0 | 0 | 5 | 4 | 1 | 0 | 0 |
| INTERMENSTRUAL BLEEDING | 5 | 4 | 1 | 0 | 0 | 5 | 4 | 1 | 0 | 0 |
| MENSTRUAL DISORDER | 7 | 5 | 2 | 0 | 0 | 7 | 5 | 2 | 0 | 0 |
| MENSTRUATION IRREGULAR | 12 | 9 | 3 | 0 | 0 | 12 | 9 | 3 | 0 | 0 |
| PREMENSTRUAL PAIN | 0 | 0 | 0 | 0 | 0 | 0 | 0 | 0 | 0 | 0 |
| RETROGRADE MENSTRUATION | 1 | 1 | 0 | 0 | 0 | 1 | 1 | 0 | 0 | 0 |
| *MENSTRUATION WITH DECREASED BLEEDING* | | | | | | | | | | |
| AMENORRHOEA | 0 | 0 | 0 | 0 | 0 | 0 | 0 | 0 | 0 | 0 |
| HYPOMENORRHOEA | 4 | 3 | 1 | 0 | 0 | 3 | 3 | 0 | 0 | 0 |
| MENSTRUATION DELAYED | 14 | 9 | 5 | 0 | 0 | 12 | 9 | 3 | 0 | 0 |
| OLIGOMENORRHOEA | 0 | 0 | 0 | 0 | 0 | 0 | 0 | 0 | 0 | 0 |
| *MENSTRUATION WITH INCREASED BLEEDING* | | | | | | | | | | |
| HEAVY MENSTRUAL BLEEDING | 20 | 16 | 4 | 0 | 0 | 20 | 16 | 4 | 0 | 0 |
| POLYMENORRHOEA | 4 | 4 | 0 | 0 | 0 | 4 | 4 | 0 | 0 | 0 |
| ***PENILE AND SCROTAL DISORDERS (EXCL INFECTIONS AND INFLAMMATIONS)*** | | | | | | | | | | |
| *SCROTAL DISORDERS NEC* | | | | | | | | | | |
| SCROTAL SWELLING | 1 | 0 | 1 | 0 | 0 | 1 | 0 | 1 | 0 | 0 |
| ***REPRODUCTIVE TRACT DISORDERS NEC*** | | | | | | | | | | |
| *REPRODUCTIVE TRACT DISORDERS NEC (EXCL NEOPLASMS)* | | | | | | | | | | |
| GENITAL LESION | 1 | 1 | 0 | 0 | 0 | 1 | 1 | 0 | 0 | 0 |
| *REPRODUCTIVE TRACT SIGNS AND SYMPTOMS NEC* | | | | | | | | | | |
| GENITAL DISCOMFORT | 1 | 1 | 0 | 0 | 0 | 1 | 1 | 0 | 0 | 0 |
| GENITAL PAIN | 1 | 1 | 0 | 0 | 0 | 1 | 1 | 0 | 0 | 0 |
| ***UTERINE, PELVIC AND BROAD LIGAMENT DISORDERS*** | | | | | | | | | | |
| *PELVIS AND BROAD LIGAMENT DISORDERS NEC* | | | | | | | | | | |
| ADNEXA UTERI PAIN | 1 | 1 | 0 | 0 | 0 | 1 | 1 | 0 | 0 | 0 |

| | | | | | | | | | | |
|---|---|---|---|---|---|---|---|---|---|---|
| ***VULVOVAGINAL DISORDERS (EXCL INFECTIONS AND INFLAMMATIONS)*** | | | | | | | | | | |
| *VULVOVAGINAL DISORDERS NEC* | | | | | | | | | | |
| VAGINAL HAEMORRHAGE | 4 | 3 | 1 | 0 | 0 | 3 | 3 | 0 | 0 | 0 |
| *VULVOVAGINAL SIGNS AND SYMPTOMS* | | | | | | | | | | |
| VAGINAL DISCHARGE | 1 | 1 | 0 | 0 | 0 | 1 | 1 | 0 | 0 | 0 |
| **RESPIRATORY, THORACIC AND MEDIASTINAL DISORDERS** | | | | | | | | | | |
| ***BRONCHIAL DISORDERS (EXCL NEOPLASMS)*** | | | | | | | | | | |
| *BRONCHOSPASM AND OBSTRUCTION* | | | | | | | | | | |
| ASTHMA | 5 | 3 | 2 | 0 | 0 | 4 | 3 | 1 | 0 | 0 |
| WHEEZING | 9 | 8 | 1 | 0 | 0 | 8 | 8 | 0 | 0 | 0 |
| ***LOWER RESPIRATORY TRACT DISORDERS (EXCL OBSTRUCTION AND INFECTION)*** | | | | | | | | | | |
| *PULMONARY OEDEMAS* | | | | | | | | | | |
| PULMONARY CONGESTION | 0 | 0 | 0 | 0 | 0 | 0 | 0 | 0 | 0 | 0 |
| ***PULMONARY VASCULAR DISORDERS*** | | | | | | | | | | |
| *PULMONARY THROMBOTIC AND EMBOLIC CONDITIONS* | | | | | | | | | | |
| PULMONARY EMBOLISM | 1 | 1 | 0 | 0 | 0 | 1 | 1 | 0 | 0 | 0 |
| ***RESPIRATORY DISORDERS NEC*** | | | | | | | | | | |
| *BREATHING ABNORMALITIES* | | | | | | | | | | |
| DYSPNOEA | 68 | 56 | 12 | 0 | 0 | 66 | 56 | 10 | 0 | 0 |
| HYPERVENTILATION | 1 | 1 | 0 | 0 | 0 | 1 | 1 | 0 | 0 | 0 |
| HYPOPNOEA | 4 | 4 | 0 | 0 | 0 | 4 | 4 | 0 | 0 | 0 |
| IRREGULAR BREATHING | 1 | 1 | 0 | 0 | 0 | 1 | 1 | 0 | 0 | 0 |
| MOUTH BREATHING | 1 | 1 | 0 | 0 | 0 | 1 | 1 | 0 | 0 | 0 |
| RESPIRATION ABNORMAL | 0 | 0 | 0 | 0 | 0 | 0 | 0 | 0 | 0 | 0 |
| RESPIRATORY ARREST | 0 | 0 | 0 | 0 | 0 | 0 | 0 | 0 | 0 | 0 |
| RESPIRATORY FATIGUE | 2 | 2 | 0 | 0 | 0 | 2 | 2 | 0 | 0 | 0 |
| SLEEP APNOEA SYNDROME | 1 | 0 | 1 | 0 | 0 | 1 | 0 | 1 | 0 | 0 |
| *COUGHING AND ASSOCIATED SYMPTOMS* | | | | | | | | | | |
| COUGH | 66 | 51 | 15 | 0 | 0 | 64 | 51 | 13 | 0 | 0 |
| PRODUCTIVE COUGH | 2 | 1 | 1 | 0 | 0 | 2 | 1 | 1 | 0 | 0 |
| *RESPIRATORY TRACT DISORDERS NEC* | | | | | | | | | | |
| RESPIRATORY TRACT IRRITATION | 1 | 1 | 0 | 0 | 0 | 1 | 1 | 0 | 0 | 0 |
| ***RESPIRATORY TRACT SIGNS AND SYMPTOMS*** | | | | | | | | | | |
| *LOWER RESPIRATORY TRACT SIGNS AND SYMPTOMS* | | | | | | | | | | |
| PULMONARY PAIN | 2 | 2 | 0 | 0 | 0 | 2 | 2 | 0 | 0 | 0 |
| *RESPIRATORY SIGNS AND SYMPTOMS NEC* | | | | | | | | | | |

| | | | | | | | | | | |
|---|---|---|---|---|---|---|---|---|---|---|
| RESPIRATORY SYMPTOM | 0 | 0 | 0 | 0 | 0 | 0 | 0 | 0 | 0 | 0 |
| *UPPER RESPIRATORY TRACT SIGNS AND SYMPTOMS* | | | | | | | | | | |
| APHONIA | 2 | 2 | 0 | 0 | 0 | 2 | 2 | 0 | 0 | 0 |
| CATARRH | 2 | 1 | 1 | 0 | 0 | 2 | 1 | 1 | 0 | 0 |
| DRY THROAT | 6 | 5 | 1 | 0 | 0 | 5 | 5 | 0 | 0 | 0 |
| DYSPHONIA | 5 | 5 | 0 | 0 | 0 | 5 | 5 | 0 | 0 | 0 |
| INCREASED UPPER AIRWAY SECRETION | 0 | 0 | 0 | 0 | 0 | 0 | 0 | 0 | 0 | 0 |
| INCREASED VISCOSITY OF UPPER RESPIRATORY SE | 0 | 0 | 0 | 0 | 0 | 0 | 0 | 0 | 0 | 0 |
| NASAL DISCOMFORT | 1 | 1 | 0 | 0 | 0 | 1 | 1 | 0 | 0 | 0 |
| OROPHARYNGEAL DISCOMFORT | 2 | 2 | 0 | 0 | 0 | 2 | 2 | 0 | 0 | 0 |
| OROPHARYNGEAL PAIN | 131 | 110 | 21 | 0 | 0 | 126 | 110 | 16 | 0 | 0 |
| PARANASAL SINUS DISCOMFORT | 2 | 2 | 0 | 0 | 0 | 2 | 2 | 0 | 0 | 0 |
| RHINALGIA | 1 | 0 | 1 | 0 | 0 | 1 | 0 | 1 | 0 | 0 |
| RHINORRHOEA | 63 | 46 | 17 | 0 | 0 | 57 | 46 | 11 | 0 | 0 |
| SINUS PAIN | 11 | 9 | 2 | 0 | 0 | 11 | 9 | 2 | 0 | 0 |
| SNEEZING | 15 | 12 | 3 | 0 | 0 | 14 | 12 | 2 | 0 | 0 |
| THROAT CLEARING | 0 | 0 | 0 | 0 | 0 | 0 | 0 | 0 | 0 | 0 |
| THROAT IRRITATION | 1 | 1 | 0 | 0 | 0 | 1 | 1 | 0 | 0 | 0 |
| THROAT TIGHTNESS | 4 | 3 | 1 | 0 | 0 | 4 | 3 | 1 | 0 | 0 |
| UPPER-AIRWAY COUGH SYNDROME | 0 | 0 | 0 | 0 | 0 | 0 | 0 | 0 | 0 | 0 |
| YAWNING | 2 | 2 | 0 | 0 | 0 | 2 | 2 | 0 | 0 | 0 |
| ***UPPER RESPIRATORY TRACT DISORDERS (EXCL INFECTIONS)*** | | | | | | | | | | |
| *NASAL CONGESTION AND INFLAMMATIONS* | | | | | | | | | | |
| NASAL CONGESTION | 11 | 9 | 2 | 0 | 0 | 10 | 9 | 1 | 0 | 0 |
| RHINITIS ALLERGIC | 1 | 1 | 0 | 0 | 0 | 1 | 1 | 0 | 0 | 0 |
| RHINITIS ATROPHIC | 1 | 0 | 1 | 0 | 0 | 1 | 0 | 1 | 0 | 0 |
| *NASAL DISORDERS NEC* | | | | | | | | | | |
| EPISTAXIS | 22 | 15 | 7 | 0 | 0 | 21 | 15 | 6 | 0 | 0 |
| NASAL DRYNESS | 2 | 1 | 1 | 0 | 0 | 1 | 1 | 0 | 0 | 0 |
| *PARANASAL SINUS DISORDERS (EXCL INFECTIONS AND NEOPLASMS)* | | | | | | | | | | |
| SINUS CONGESTION | 4 | 2 | 2 | 0 | 0 | 4 | 2 | 2 | 0 | 0 |
| *PHARYNGEAL DISORDERS (EXCL INFECTIONS AND NEOPLASMS)* | | | | | | | | | | |
| PHARYNGEAL SWELLING | 2 | 2 | 0 | 0 | 0 | 2 | 2 | 0 | 0 | 0 |
| PHARYNGEAL ULCERATION | 1 | 1 | 0 | 0 | 0 | 1 | 1 | 0 | 0 | 0 |
| TONSILLAR ERYTHEMA | 0 | 0 | 0 | 0 | 0 | 0 | 0 | 0 | 0 | 0 |

| | | | | | | | | | | |
|---|---|---|---|---|---|---|---|---|---|---|
| *TRACHEAL DISORDERS (EXCL INFECTIONS AND NEOPLASMS)* | | | | | | | | | | |
| TRACHEAL PAIN | 1 | 1 | 0 | 0 | 0 | 1 | 1 | 0 | 0 | 0 |
| **SKIN AND SUBCUTANEOUS TISSUE DISORDERS** | | | | | | | | | | |
| ***ANGIOEDEMA AND URTICARIA*** | | | | | | | | | | |
| *ANGIOEDEMAS* | | | | | | | | | | |
| ANGIOEDEMA | 3 | 3 | 0 | 0 | 0 | 3 | 3 | 0 | 0 | 0 |
| *URTICARIAS* | | | | | | | | | | |
| COLD URTICARIA | 1 | 0 | 1 | 0 | 0 | 1 | 0 | 1 | 0 | 0 |
| SOLAR URTICARIA | 1 | 1 | 0 | 0 | 0 | 1 | 1 | 0 | 0 | 0 |
| URTICARIA | 9 | 8 | 1 | 0 | 0 | 9 | 8 | 1 | 0 | 0 |
| URTICARIA CHRONIC | 0 | 0 | 0 | 0 | 0 | 0 | 0 | 0 | 0 | 0 |
| ***CORNIFICATION AND DYSTROPHIC SKIN DISORDERS*** | | | | | | | | | | |
| *SKIN DYSTROPHIES* | | | | | | | | | | |
| HYPERTROPHIC SCAR | 0 | 0 | 0 | 0 | 0 | 0 | 0 | 0 | 0 | 0 |
| ***CUTANEOUS NEOPLASMS BENIGN*** | | | | | | | | | | |
| *SKIN CYSTS AND POLYPS* | | | | | | | | | | |
| DERMAL CYST | 0 | 0 | 0 | 0 | 0 | 0 | 0 | 0 | 0 | 0 |
| ***EPIDERMAL AND DERMAL CONDITIONS*** | | | | | | | | | | |
| *BULLOUS CONDITIONS* | | | | | | | | | | |
| BLISTER | 3 | 2 | 1 | 0 | 0 | 3 | 2 | 1 | 0 | 0 |
| BLOOD BLISTER | 0 | 0 | 0 | 0 | 0 | 0 | 0 | 0 | 0 | 0 |
| PEMPHIGOID | 0 | 0 | 0 | 0 | 0 | 0 | 0 | 0 | 0 | 0 |
| TOXIC EPIDERMAL NECROLYSIS | 1 | 1 | 0 | 0 | 0 | 1 | 1 | 0 | 0 | 0 |
| *DERMAL AND EPIDERMAL CONDITIONS NEC* | | | | | | | | | | |
| DRY SKIN | 2 | 2 | 0 | 0 | 0 | 2 | 2 | 0 | 0 | 0 |
| PAIN OF SKIN | 20 | 18 | 2 | 0 | 0 | 19 | 18 | 1 | 0 | 0 |
| SENSITIVE SKIN | 18 | 18 | 0 | 0 | 0 | 18 | 18 | 0 | 0 | 0 |
| SKIN BURNING SENSATION | 8 | 7 | 1 | 0 | 0 | 8 | 7 | 1 | 0 | 0 |
| SKIN DISCOLOURATION | 0 | 0 | 0 | 0 | 0 | 0 | 0 | 0 | 0 | 0 |
| SKIN FRAGILITY | 0 | 0 | 0 | 0 | 0 | 0 | 0 | 0 | 0 | 0 |
| SKIN LESION | 0 | 0 | 0 | 0 | 0 | 0 | 0 | 0 | 0 | 0 |
| SKIN ODOUR ABNORMAL | 2 | 2 | 0 | 0 | 0 | 2 | 2 | 0 | 0 | 0 |
| SKIN REACTION | 1 | 1 | 0 | 0 | 0 | 1 | 1 | 0 | 0 | 0 |
| SKIN SENSITISATION | 1 | 1 | 0 | 0 | 0 | 1 | 1 | 0 | 0 | 0 |
| SKIN WARM | 16 | 14 | 2 | 0 | 0 | 15 | 14 | 1 | 0 | 0 |
| *DERMATITIS AND ECZEMA* | | | | | | | | | | |

| | | | | | | | | | | |
|---|---|---|---|---|---|---|---|---|---|---|
| DERMATITIS | 2 | 2 | 0 | 0 | 0 | 2 | 2 | 0 | 0 | 0 |
| DERMATITIS ALLERGIC | 3 | 3 | 0 | 0 | 0 | 3 | 3 | 0 | 0 | 0 |
| DERMATITIS ATOPIC | 2 | 2 | 0 | 0 | 0 | 2 | 2 | 0 | 0 | 0 |
| DERMATITIS CONTACT | 1 | 1 | 0 | 0 | 0 | 1 | 1 | 0 | 0 | 0 |
| ECZEMA | 0 | 0 | 0 | 0 | 0 | 0 | 0 | 0 | 0 | 0 |
| ECZEMA ASTEATOTIC | 0 | 0 | 0 | 0 | 0 | 0 | 0 | 0 | 0 | 0 |
| NEURODERMATITIS | 1 | 1 | 0 | 0 | 0 | 1 | 1 | 0 | 0 | 0 |
| SEBORRHOEIC DERMATITIS | 2 | 2 | 0 | 0 | 0 | 2 | 2 | 0 | 0 | 0 |
| SKIN IRRITATION | 6 | 5 | 1 | 0 | 0 | 6 | 5 | 1 | 0 | 0 |
| *ERYTHEMAS* | | | | | | | | | | |
| ERYTHEMA | 29 | 26 | 3 | 0 | 0 | 27 | 26 | 1 | 0 | 0 |
| *EXFOLIATIVE CONDITIONS* | | | | | | | | | | |
| SKIN EXFOLIATION | 1 | 1 | 0 | 0 | 0 | 1 | 1 | 0 | 0 | 0 |
| *PAPULOSQUAMOUS CONDITIONS* | | | | | | | | | | |
| LICHEN PLANUS | 1 | 0 | 1 | 0 | 0 | 1 | 0 | 1 | 0 | 0 |
| PITYRIASIS ROSEA | 1 | 1 | 0 | 0 | 0 | 1 | 1 | 0 | 0 | 0 |
| *PHOTOSENSITIVITY AND PHOTODERMATOSIS CONDITIONS* | | | | | | | | | | |
| PHOTOSENSITIVITY REACTION | 5 | 3 | 2 | 0 | 0 | 4 | 3 | 1 | 0 | 0 |
| *PRURITUS NEC* | | | | | | | | | | |
| PRURITUS | 101 | 88 | 13 | 0 | 0 | 97 | 88 | 9 | 0 | 0 |
| *PSORIATIC CONDITIONS* | | | | | | | | | | |
| PSORIASIS | 6 | 5 | 1 | 0 | 0 | 6 | 5 | 1 | 0 | 0 |
| *RASHES, ERUPTIONS AND EXANTHEMS NEC* | | | | | | | | | | |
| RASH | 93 | 78 | 15 | 0 | 0 | 88 | 78 | 10 | 0 | 0 |
| RASH ERYTHEMATOUS | 21 | 19 | 2 | 0 | 0 | 20 | 19 | 1 | 0 | 0 |
| RASH MACULAR | 2 | 0 | 2 | 0 | 0 | 2 | 0 | 2 | 0 | 0 |
| RASH MORBILLIFORM | 0 | 0 | 0 | 0 | 0 | 0 | 0 | 0 | 0 | 0 |
| RASH PAPULAR | 9 | 8 | 1 | 0 | 0 | 9 | 8 | 1 | 0 | 0 |
| RASH PRURITIC | 19 | 17 | 2 | 0 | 0 | 19 | 17 | 2 | 0 | 0 |
| *SKIN INJURIES AND MECHANICAL DERMATOSES* | | | | | | | | | | |
| DECUBITUS ULCER | 1 | 1 | 0 | 0 | 0 | 1 | 1 | 0 | 0 | 0 |
| NEEDLE TRACK MARKS | 1 | 1 | 0 | 0 | 0 | 1 | 1 | 0 | 0 | 0 |
| ***SKIN AND SUBCUTANEOUS TISSUE DISORDERS NEC*** | | | | | | | | | | |
| *SKIN AND SUBCUTANEOUS TISSUE ULCERATIONS* | | | | | | | | | | |
| SKIN EROSION | 2 | 1 | 1 | 0 | 0 | 1 | 1 | 0 | 0 | 0 |

| | | | | | | | | | | |
|---|---|---|---|---|---|---|---|---|---|---|
| ***SKIN APPENDAGE CONDITIONS*** | | | | | | | | | | |
| *ACNES* | | | | | | | | | | |
| ACNE | 1 | 1 | 0 | 0 | 0 | 1 | 1 | 0 | 0 | 0 |
| ACNE CYSTIC | 1 | 0 | 1 | 0 | 0 | 1 | 0 | 1 | 0 | 0 |
| *ALOPECIAS* | | | | | | | | | | |
| ALOPECIA | 5 | 3 | 2 | 0 | 0 | 4 | 3 | 1 | 0 | 0 |
| *APOCRINE AND ECCRINE GLAND DISORDERS* | | | | | | | | | | |
| COLD SWEAT | 23 | 22 | 1 | 0 | 0 | 22 | 22 | 0 | 0 | 0 |
| HYPERHIDROSIS | 111 | 100 | 10 | 1 | 0 | 107 | 100 | 7 | 0 | 0 |
| MILIARIA | 2 | 2 | 0 | 0 | 0 | 2 | 2 | 0 | 0 | 0 |
| NIGHT SWEATS | 53 | 49 | 4 | 0 | 0 | 49 | 49 | 0 | 0 | 0 |
| *HYPERTRICHOSES* | | | | | | | | | | |
| HIRSUTISM | 0 | 0 | 0 | 0 | 0 | 0 | 0 | 0 | 0 | 0 |
| *NAIL AND NAIL BED CONDITIONS (EXCL INFECTIONS AND INFESTATIONS)* | | | | | | | | | | |
| NAIL DISCOLOURATION | 0 | 0 | 0 | 0 | 0 | 0 | 0 | 0 | 0 | 0 |
| ONYCHOCLASIS | 0 | 0 | 0 | 0 | 0 | 0 | 0 | 0 | 0 | 0 |
| *PILAR DISORDERS NEC* | | | | | | | | | | |
| PILOERECTION | 3 | 2 | 1 | 0 | 0 | 2 | 2 | 0 | 0 | 0 |
| ***SKIN VASCULAR ABNORMALITIES*** | | | | | | | | | | |
| *PURPURA AND RELATED CONDITIONS* | | | | | | | | | | |
| PETECHIAE | 2 | 2 | 0 | 0 | 0 | 2 | 2 | 0 | 0 | 0 |
| PURPURA | 0 | 0 | 0 | 0 | 0 | 0 | 0 | 0 | 0 | 0 |
| *SKIN HAEMORRHAGES* | | | | | | | | | | |
| SKIN HAEMORRHAGE | 1 | 1 | 0 | 0 | 0 | 1 | 1 | 0 | 0 | 0 |
| **SOCIAL CIRCUMSTANCES** | | | | | | | | | | |
| ***ECONOMIC AND HOUSING ISSUES*** | | | | | | | | | | |
| *EMPLOYMENT ISSUES* | | | | | | | | | | |
| RETIREMENT | 1 | 1 | 0 | 0 | 0 | 1 | 1 | 0 | 0 | 0 |
| ***ENVIRONMENTAL ISSUES*** | | | | | | | | | | |
| *NON-OCCUPATIONAL AND UNSPECIFIED ENVIRONMENTAL PROBLEMS* | | | | | | | | | | |
| POLLUTION | 1 | 1 | 0 | 0 | 0 | 1 | 1 | 0 | 0 | 0 |
| **SURGICAL AND MEDICAL PROCEDURES** | | | | | | | | | | |
| ***BONE AND JOINT THERAPEUTIC PROCEDURES*** | | | | | | | | | | |
| *JOINT THERAPEUTIC PROCEDURES* | | | | | | | | | | |
| KNEE OPERATION | 1 | 1 | 0 | 0 | 0 | 1 | 1 | 0 | 0 | 0 |
| ***BREAST THERAPEUTIC PROCEDURES*** | | | | | | | | | | |

| | | | | | | | | | | |
|---|---|---|---|---|---|---|---|---|---|---|
| *MASTECTOMIES* | | | | | | | | | | |
| BREAST CONSERVING SURGERY | 1 | 0 | 1 | 0 | 0 | 1 | 0 | 1 | 0 | 0 |
| ***GASTROINTESTINAL THERAPEUTIC PROCEDURES*** | | | | | | | | | | |
| *GASTROINTESTINAL THERAPEUTIC PROCEDURES NEC* | | | | | | | | | | |
| PROPHYLAXIS OF NAUSEA AND VOMITING | 1 | 1 | 0 | 0 | 0 | 1 | 1 | 0 | 0 | 0 |
| ***HEAD AND NECK THERAPEUTIC PROCEDURES*** | | | | | | | | | | |
| *DENTAL AND GINGIVAL THERAPEUTIC PROCEDURES* | | | | | | | | | | |
| DENTAL CARE | 2 | 2 | 0 | 0 | 0 | 2 | 2 | 0 | 0 | 0 |
| ***NERVOUS SYSTEM, SKULL AND SPINE THERAPEUTIC PROCEDURES*** | | | | | | | | | | |
| *SKULL AND BRAIN THERAPEUTIC PROCEDURES* | | | | | | | | | | |
| BRAIN TUMOUR OPERATION | 1 | 1 | 0 | 0 | 0 | 1 | 1 | 0 | 0 | 0 |
| ***OBSTETRIC AND GYNAECOLOGICAL THERAPEUTIC PROCEDURES*** | | | | | | | | | | |
| *FERTILITY AND FERTILISATION INTERVENTIONS FEMALE* | | | | | | | | | | |
| ENDOMETRIAL SCRATCHING | 0 | 0 | 0 | 0 | 0 | 0 | 0 | 0 | 0 | 0 |
| ***RESPIRATORY TRACT THERAPEUTIC PROCEDURES*** | | | | | | | | | | |
| *RESPIRATORY TRACT THERAPEUTIC PROCEDURES NEC* | | | | | | | | | | |
| OXYGEN THERAPY | 0 | 0 | 0 | 0 | 0 | 0 | 0 | 0 | 0 | 0 |
| ***THERAPEUTIC PROCEDURES AND SUPPORTIVE CARE NEC*** | | | | | | | | | | |
| *ANAESTHESIA AND ALLIED PROCEDURES* | | | | | | | | | | |
| NERVE BLOCK | 2 | 2 | 0 | 0 | 0 | 2 | 2 | 0 | 0 | 0 |
| *DIETARY AND NUTRITIONAL THERAPIES* | | | | | | | | | | |
| NOTHING BY MOUTH ORDER | 0 | 0 | 0 | 0 | 0 | 0 | 0 | 0 | 0 | 0 |
| *IMMUNISATIONS* | | | | | | | | | | |
| COVID-19 IMMUNISATION | 1 | 0 | 1 | 0 | 0 | 0 | 0 | 0 | 0 | 0 |
| *THERAPEUTIC PROCEDURES NEC* | | | | | | | | | | |
| INJECTION | 2 | 2 | 0 | 0 | 0 | 2 | 2 | 0 | 0 | 0 |
| LOCALISED ALTERNATING HOT AND COLD THERAPY | 2 | 2 | 0 | 0 | 0 | 2 | 2 | 0 | 0 | 0 |
| MASS EXCISION | 1 | 0 | 1 | 0 | 0 | 1 | 0 | 1 | 0 | 0 |
| REINFUSION | 0 | 0 | 0 | 0 | 0 | 0 | 0 | 0 | 0 | 0 |
| **VASCULAR DISORDERS** | | | | | | | | | | |
| ***ARTERIOSCLEROSIS, STENOSIS, VASCULAR INSUFFICIENCY AND NECROSIS*** | | | | | | | | | | |
| *NON-SITE SPECIFIC NECROSIS AND VASCULAR INSUFFICIENCY NEC* | | | | | | | | | | |
| VASOSPASM | 1 | 1 | 0 | 0 | 0 | 1 | 1 | 0 | 0 | 0 |
| *PERIPHERAL VASOCONSTRICTION, NECROSIS AND VASCULAR INSUFFICIENCY* | | | | | | | | | | |
| PERIPHERAL COLDNESS | 47 | 45 | 2 | 0 | 0 | 46 | 45 | 1 | 0 | 0 |

| | | | | | | | | | | |
|---|---|---|---|---|---|---|---|---|---|---|
| RAYNAUD'S PHENOMENON | 1 | 1 | 0 | 0 | 0 | 1 | 1 | 0 | 0 | 0 |
| ***DECREASED AND NONSPECIFIC BLOOD PRESSURE DISORDERS AND SHOCK*** | | | | | | | | | | |
| *BLOOD PRESSURE DISORDERS NEC* | | | | | | | | | | |
| BLOOD PRESSURE FLUCTUATION | 1 | 1 | 0 | 0 | 0 | 1 | 1 | 0 | 0 | 0 |
| *VASCULAR HYPOTENSIVE DISORDERS* | | | | | | | | | | |
| CAPILLARY LEAK SYNDROME | 1 | 1 | 0 | 0 | 0 | 1 | 1 | 0 | 0 | 0 |
| HYPOTENSION | 3 | 3 | 0 | 0 | 0 | 3 | 3 | 0 | 0 | 0 |
| ORTHOSTATIC HYPOTENSION | 1 | 1 | 0 | 0 | 0 | 1 | 1 | 0 | 0 | 0 |
| ***EMBOLISM AND THROMBOSIS*** | | | | | | | | | | |
| *NON-SITE SPECIFIC EMBOLISM AND THROMBOSIS* | | | | | | | | | | |
| EMBOLISM | 0 | 0 | 0 | 0 | 0 | 0 | 0 | 0 | 0 | 0 |
| THROMBOSIS | 7 | 6 | 1 | 0 | 0 | 7 | 6 | 1 | 0 | 0 |
| VENOUS THROMBOSIS | 1 | 0 | 1 | 0 | 0 | 0 | 0 | 0 | 0 | 0 |
| *PERIPHERAL EMBOLISM AND THROMBOSIS* | | | | | | | | | | |
| BLUE TOE SYNDROME | 2 | 2 | 0 | 0 | 0 | 2 | 2 | 0 | 0 | 0 |
| DEEP VEIN THROMBOSIS | 4 | 3 | 1 | 0 | 0 | 3 | 3 | 0 | 0 | 0 |
| SUPERFICIAL VEIN THROMBOSIS | 0 | 0 | 0 | 0 | 0 | 0 | 0 | 0 | 0 | 0 |
| ***LYMPHATIC VESSEL DISORDERS*** | | | | | | | | | | |
| *LYMPHOEDEMAS* | | | | | | | | | | |
| LYMPHOEDEMA | 0 | 0 | 0 | 0 | 0 | 0 | 0 | 0 | 0 | 0 |
| ***VASCULAR DISORDERS NEC*** | | | | | | | | | | |
| *NON-SITE SPECIFIC VASCULAR DISORDERS NEC* | | | | | | | | | | |
| VASCULAR PAIN | 0 | 0 | 0 | 0 | 0 | 0 | 0 | 0 | 0 | 0 |
| VEIN DISCOLOURATION | 0 | 0 | 0 | 0 | 0 | 0 | 0 | 0 | 0 | 0 |
| VEIN RUPTURE | 1 | 0 | 1 | 0 | 0 | 1 | 0 | 1 | 0 | 0 |
| *PERIPHERAL VASCULAR DISORDERS NEC* | | | | | | | | | | |
| FLUSHING | 22 | 19 | 3 | 0 | 0 | 21 | 19 | 2 | 0 | 0 |
| HOT FLUSH | 78 | 67 | 11 | 0 | 0 | 75 | 67 | 8 | 0 | 0 |
| *SITE SPECIFIC VASCULAR DISORDERS NEC* | | | | | | | | | | |
| PALLOR | 4 | 3 | 1 | 0 | 0 | 3 | 3 | 0 | 0 | 0 |
| ***VASCULAR HAEMORRHAGIC DISORDERS*** | | | | | | | | | | |
| *HAEMORRHAGES NEC* | | | | | | | | | | |
| HAEMATOMA | 1 | 1 | 0 | 0 | 0 | 1 | 1 | 0 | 0 | 0 |
| HAEMORRHAGE | 2 | 1 | 1 | 0 | 0 | 1 | 1 | 0 | 0 | 0 |
| ***VASCULAR HYPERTENSIVE DISORDERS*** | | | | | | | | | | |
| *ACCELERATED AND MALIGNANT HYPERTENSION* | | | | | | | | | | |

| | | | | | | | | | | |
|---|---|---|---|---|---|---|---|---|---|---|
| HYPERTENSIVE URGENCY | 0 | 0 | 0 | 0 | 0 | 0 | 0 | 0 | 0 | 0 |
| *VASCULAR HYPERTENSIVE DISORDERS NEC* | | | | | | | | | | |
| HYPERTENSION | 7 | 3 | 4 | 0 | 0 | 6 | 3 | 3 | 0 | 0 |
| SYSTOLIC HYPERTENSION | 1 | 1 | 0 | 0 | 0 | 1 | 1 | 0 | 0 | 0 |
| ***VASCULAR INFECTIONS AND INFLAMMATIONS*** | | | | | | | | | | |
| *ARTERIAL INFECTIONS AND INFLAMMATIONS* | | | | | | | | | | |
| GIANT CELL ARTERITIS | 1 | 1 | 0 | 0 | 0 | 1 | 1 | 0 | 0 | 0 |
| *PHLEBITIS NEC* | | | | | | | | | | |
| PHLEBITIS | 2 | 1 | 1 | 0 | 0 | 2 | 1 | 1 | 0 | 0 |
| *VASCULITIDES NEC* | | | | | | | | | | |
| VASCULITIS | 0 | 0 | 0 | 0 | 0 | 0 | 0 | 0 | 0 | 0 |
| ***VENOUS VARICES*** | | | | | | | | | | |
| *VARICOSE VEINS NEC* | | | | | | | | | | |
| SPIDER VEIN | 0 | 0 | 0 | 0 | 0 | 0 | 0 | 0 | 0 | 0 |
| VARICOSE VEIN | 2 | 2 | 0 | 0 | 0 | 2 | 2 | 0 | 0 | 0 |
| **TOTAL ADR EVENTS** | ***23198*** | ***19559*** | ***3613*** | ***25*** | ***1*** | ***22004*** | ***19559*** | ***2433*** | ***12*** | ***0*** |

**TABLE S1-4. Moderna COVID-19 vaccine: ADR listing for events reported in the YCVM in (a) those reporting any vaccination dose and (b) in those who had reported a 1st dose vaccination and any subsequent doses.**

| | Patients with any vaccination dose: ADR Counts | | | | | Patients reporting a 1st dose: ADR Counts | | | | |
|---|---|---|---|---|---|---|---|---|---|---|
| **REACTION TERM (SOC, *HLGT*, *HLT*, PT)** | All doses | 1st dose | 2nd dose | 3rd dose | Other doses | All doses | 1st dose | 2nd dose | 3rd dose | Other doses |
| ***(freetext)*** | 0 | 0 | 0 | 0 | 0 | 0 | 0 | 0 | 0 | 0 |
| **BLOOD AND LYMPHATIC SYSTEM DISORDERS** | | | | | | | | | | |
| ***COAGULOPATHIES AND BLEEDING DIATHESES (EXCL THROMBOCYTOPENIC)*** | | | | | | | | | | |
| *BLEEDING TENDENCIES* | | | | | | | | | | |
| INCREASED TENDENCY TO BRUISE | 0 | 0 | 0 | 0 | 0 | 0 | 0 | 0 | 0 | 0 |
| ***HAEMOGLOBINOPATHIES*** | | | | | | | | | | |
| *SICKLE CELL TRAIT AND DISORDERS* | | | | | | | | | | |
| SICKLE CELL ANAEMIA WITH CRISIS | 0 | 0 | 0 | 0 | 0 | 0 | 0 | 0 | 0 | 0 |
| ***PLATELET DISORDERS*** | | | | | | | | | | |
| *THROMBOCYTOPENIAS* | | | | | | | | | | |
| IMMUNE THROMBOCYTOPENIA | 0 | 0 | 0 | 0 | 0 | 0 | 0 | 0 | 0 | 0 |
| ***SPLEEN, LYMPHATIC AND RETICULOENDOTHELIAL SYSTEM DISORDERS*** | | | | | | | | | | |
| *LYMPHATIC SYSTEM DISORDERS NEC* | | | | | | | | | | |
| LYMPH NODE PAIN | 3 | 2 | 0 | 1 | 0 | 3 | 2 | 0 | 1 | 0 |
| LYMPHADENITIS | 2 | 1 | 0 | 1 | 0 | 2 | 1 | 0 | 1 | 0 |
| LYMPHADENOPATHY | 19 | 7 | 2 | 10 | 0 | 17 | 7 | 2 | 8 | 0 |
| **CARDIAC DISORDERS** | | | | | | | | | | |
| ***CARDIAC ARRHYTHMIAS*** | | | | | | | | | | |
| *RATE AND RHYTHM DISORDERS NEC* | | | | | | | | | | |
| ARRHYTHMIA | 0 | 0 | 0 | 0 | 0 | 0 | 0 | 0 | 0 | 0 |
| CARDIAC FLUTTER | 1 | 0 | 0 | 1 | 0 | 1 | 0 | 0 | 1 | 0 |
| EXTRASYSTOLES | 0 | 0 | 0 | 0 | 0 | 0 | 0 | 0 | 0 | 0 |
| TACHYCARDIA | 0 | 0 | 0 | 0 | 0 | 0 | 0 | 0 | 0 | 0 |
| *SUPRAVENTRICULAR ARRHYTHMIAS* | | | | | | | | | | |
| ATRIAL FIBRILLATION | 1 | 0 | 0 | 1 | 0 | 0 | 0 | 0 | 0 | 0 |
| SUPRAVENTRICULAR TACHYCARDIA | 0 | 0 | 0 | 0 | 0 | 0 | 0 | 0 | 0 | 0 |

| | | | | | | | | | | |
|---|---|---|---|---|---|---|---|---|---|---|
| ***CARDIAC DISORDERS, SIGNS AND SYMPTOMS NEC*** | | | | | | | | | | |
| *CARDIAC DISORDERS NEC* | | | | | | | | | | |
| CARDIOVASCULAR DISORDER | 0 | 0 | 0 | 0 | 0 | 0 | 0 | 0 | 0 | 0 |
| *CARDIAC SIGNS AND SYMPTOMS NEC* | | | | | | | | | | |
| PALPITATIONS | 10 | 3 | 1 | 6 | 0 | 9 | 3 | 1 | 5 | 0 |
| ***CORONARY ARTERY DISORDERS*** | | | | | | | | | | |
| *ISCHAEMIC CORONARY ARTERY DISORDERS* | | | | | | | | | | |
| ANGINA PECTORIS | 0 | 0 | 0 | 0 | 0 | 0 | 0 | 0 | 0 | 0 |
| MYOCARDIAL INFARCTION | 0 | 0 | 0 | 0 | 0 | 0 | 0 | 0 | 0 | 0 |
| ***HEART FAILURES*** | | | | | | | | | | |
| *HEART FAILURES NEC* | | | | | | | | | | |
| CARDIAC FAILURE | 0 | 0 | 0 | 0 | 0 | 0 | 0 | 0 | 0 | 0 |
| ***MYOCARDIAL DISORDERS*** | | | | | | | | | | |
| *NONINFECTIOUS MYOCARDITIS* | | | | | | | | | | |
| MYOCARDITIS | 1 | 0 | 0 | 0 | 1 | 1 | 0 | 0 | 0 | 1 |
| ***PERICARDIAL DISORDERS*** | | | | | | | | | | |
| *NONINFECTIOUS PERICARDITIS* | | | | | | | | | | |
| PERICARDITIS | 0 | 0 | 0 | 0 | 0 | 0 | 0 | 0 | 0 | 0 |
| **CONGENITAL, FAMILIAL AND GENETIC DISORDERS** | | | | | | | | | | |
| ***CARDIAC AND VASCULAR DISORDERS CONGENITAL*** | | | | | | | | | | |
| *CARDIAC DISORDERS CONGENITAL NEC* | | | | | | | | | | |
| HEART DISEASE CONGENITAL | 0 | 0 | 0 | 0 | 0 | 0 | 0 | 0 | 0 | 0 |
| ***METABOLIC AND NUTRITIONAL DISORDERS CONGENITAL*** | | | | | | | | | | |
| *INBORN ERRORS OF AMINO ACID METABOLISM* | | | | | | | | | | |
| HYPERGLYCINAEMIA | 0 | 0 | 0 | 0 | 0 | 0 | 0 | 0 | 0 | 0 |
| *INBORN ERRORS OF STEROID SYNTHESIS* | | | | | | | | | | |
| 11-BETA-HYDROXYLASE DEFICIENCY | 0 | 0 | 0 | 0 | 0 | 0 | 0 | 0 | 0 | 0 |
| ***NEUROLOGICAL DISORDERS CONGENITAL*** | | | | | | | | | | |
| *PERIPHERAL NERVOUS SYSTEM DISORDERS CONGENITAL NEC* | | | | | | | | | | |
| PAROXYSMAL EXTREME PAIN DISORDER | 0 | 0 | 0 | 0 | 0 | 0 | 0 | 0 | 0 | 0 |
| **EAR AND LABYRINTH DISORDERS** | | | | | | | | | | |

| | | | | | | | | | | |
|---|---|---|---|---|---|---|---|---|---|---|
| ***AURAL DISORDERS NEC*** | | | | | | | | | | |
| *EAR DISORDERS NEC* | | | | | | | | | | |
| EAR DISCOMFORT | 0 | 0 | 0 | 0 | 0 | 0 | 0 | 0 | 0 | 0 |
| EAR DISORDER | 0 | 0 | 0 | 0 | 0 | 0 | 0 | 0 | 0 | 0 |
| EAR PAIN | 2 | 2 | 0 | 0 | 0 | 2 | 2 | 0 | 0 | 0 |
| EAR SWELLING | 0 | 0 | 0 | 0 | 0 | 0 | 0 | 0 | 0 | 0 |
| ***EXTERNAL EAR DISORDERS (EXCL CONGENITAL)*** | | | | | | | | | | |
| *EXTERNAL EAR DISORDERS NEC* | | | | | | | | | | |
| EXCESSIVE CERUMEN PRODUCTION | 0 | 0 | 0 | 0 | 0 | 0 | 0 | 0 | 0 | 0 |
| ***HEARING DISORDERS*** | | | | | | | | | | |
| *HEARING LOSSES* | | | | | | | | | | |
| DEAFNESS | 1 | 1 | 0 | 0 | 0 | 1 | 1 | 0 | 0 | 0 |
| DEAFNESS NEUROSENSORY | 0 | 0 | 0 | 0 | 0 | 0 | 0 | 0 | 0 | 0 |
| DEAFNESS PERMANENT | 0 | 0 | 0 | 0 | 0 | 0 | 0 | 0 | 0 | 0 |
| DEAFNESS UNILATERAL | 0 | 0 | 0 | 0 | 0 | 0 | 0 | 0 | 0 | 0 |
| HYPOACUSIS | 0 | 0 | 0 | 0 | 0 | 0 | 0 | 0 | 0 | 0 |
| SUDDEN HEARING LOSS | 0 | 0 | 0 | 0 | 0 | 0 | 0 | 0 | 0 | 0 |
| *HYPERACUSIA* | | | | | | | | | | |
| HYPERACUSIS | 0 | 0 | 0 | 0 | 0 | 0 | 0 | 0 | 0 | 0 |
| ***INNER EAR AND VIIITH CRANIAL NERVE DISORDERS*** | | | | | | | | | | |
| *INNER EAR SIGNS AND SYMPTOMS* | | | | | | | | | | |
| MOTION SICKNESS | 0 | 0 | 0 | 0 | 0 | 0 | 0 | 0 | 0 | 0 |
| TINNITUS | 3 | 0 | 1 | 2 | 0 | 3 | 0 | 1 | 2 | 0 |
| VERTIGO | 2 | 0 | 0 | 1 | 1 | 1 | 0 | 0 | 0 | 1 |
| VERTIGO LABYRINTHINE | 0 | 0 | 0 | 0 | 0 | 0 | 0 | 0 | 0 | 0 |
| VERTIGO POSITIONAL | 1 | 1 | 0 | 0 | 0 | 1 | 1 | 0 | 0 | 0 |
| **ENDOCRINE DISORDERS** | | | | | | | | | | |
| ***THYROID GLAND DISORDERS*** | | | | | | | | | | |
| *THYROID HYPERFUNCTION DISORDERS* | | | | | | | | | | |
| HYPERTHYROIDISM | 0 | 0 | 0 | 0 | 0 | 0 | 0 | 0 | 0 | 0 |
| **EYE DISORDERS** | | | | | | | | | | |
| ***EYE DISORDERS NEC*** | | | | | | | | | | |
| *LACRIMATION DISORDERS* | | | | | | | | | | |
| DRY EYE | 0 | 0 | 0 | 0 | 0 | 0 | 0 | 0 | 0 | 0 |
| LACRIMATION INCREASED | 0 | 0 | 0 | 0 | 0 | 0 | 0 | 0 | 0 | 0 |
| *OCULAR DISORDERS NEC* | | | | | | | | | | |

| | | | | | | | | | | |
|---|---|---|---|---|---|---|---|---|---|---|
| EYE OEDEMA | 0 | 0 | 0 | 0 | 0 | 0 | 0 | 0 | 0 | 0 |
| EYE PAIN | 3 | 1 | 1 | 1 | 0 | 3 | 1 | 1 | 1 | 0 |
| EYE SWELLING | 2 | 0 | 0 | 2 | 0 | 0 | 0 | 0 | 0 | 0 |
| EYE ULCER | 0 | 0 | 0 | 0 | 0 | 0 | 0 | 0 | 0 | 0 |
| EYELID PAIN | 0 | 0 | 0 | 0 | 0 | 0 | 0 | 0 | 0 | 0 |
| OCULAR DISCOMFORT | 0 | 0 | 0 | 0 | 0 | 0 | 0 | 0 | 0 | 0 |
| PERIORBITAL DISCOMFORT | 0 | 0 | 0 | 0 | 0 | 0 | 0 | 0 | 0 | 0 |
| PERIORBITAL SWELLING | 1 | 0 | 0 | 1 | 0 | 1 | 0 | 0 | 1 | 0 |
| ***OCULAR HAEMORRHAGES AND VASCULAR DISORDERS NEC*** | | | | | | | | | | |
| *CONJUNCTIVAL AND CORNEAL BLEEDING AND VASCULAR DISORDERS* | | | | | | | | | | |
| CONJUNCTIVAL HAEMORRHAGE | 0 | 0 | 0 | 0 | 0 | 0 | 0 | 0 | 0 | 0 |
| *LID BLEEDING AND VASCULAR DISORDERS* | | | | | | | | | | |
| EYELID BLEEDING | 0 | 0 | 0 | 0 | 0 | 0 | 0 | 0 | 0 | 0 |
| ***OCULAR INFECTIONS, IRRITATIONS AND INFLAMMATIONS*** | | | | | | | | | | |
| *LID, LASH AND LACRIMAL INFECTIONS, IRRITATIONS AND INFLAMMATIONS* | | | | | | | | | | |
| BLEPHARITIS | 0 | 0 | 0 | 0 | 0 | 0 | 0 | 0 | 0 | 0 |
| ERYTHEMA OF EYELID | 0 | 0 | 0 | 0 | 0 | 0 | 0 | 0 | 0 | 0 |
| EYELID IRRITATION | 0 | 0 | 0 | 0 | 0 | 0 | 0 | 0 | 0 | 0 |
| *OCULAR INFECTIONS, INFLAMMATIONS AND ASSOCIATED MANIFESTATIONS* | | | | | | | | | | |
| EYE DISCHARGE | 0 | 0 | 0 | 0 | 0 | 0 | 0 | 0 | 0 | 0 |
| EYE IRRITATION | 1 | 1 | 0 | 0 | 0 | 1 | 1 | 0 | 0 | 0 |
| EYE PRURITUS | 0 | 0 | 0 | 0 | 0 | 0 | 0 | 0 | 0 | 0 |
| LIMBAL SWELLING | 1 | 0 | 0 | 1 | 0 | 1 | 0 | 0 | 1 | 0 |
| OCULAR HYPERAEMIA | 3 | 0 | 0 | 3 | 0 | 3 | 0 | 0 | 3 | 0 |
| ***OCULAR NEUROMUSCULAR DISORDERS*** | | | | | | | | | | |
| *EYELID MOVEMENT DISORDERS* | | | | | | | | | | |
| BLEPHAROSPASM | 0 | 0 | 0 | 0 | 0 | 0 | 0 | 0 | 0 | 0 |
| ***OCULAR SENSORY SYMPTOMS NEC*** | | | | | | | | | | |
| *OCULAR SENSATION DISORDERS* | | | | | | | | | | |
| ABNORMAL SENSATION IN EYE | 0 | 0 | 0 | 0 | 0 | 0 | 0 | 0 | 0 | 0 |
| ASTHENOPIA | 2 | 2 | 0 | 0 | 0 | 2 | 2 | 0 | 0 | 0 |
| FOREIGN BODY SENSATION IN EYES | 0 | 0 | 0 | 0 | 0 | 0 | 0 | 0 | 0 | 0 |
| PHOTOPHOBIA | 0 | 0 | 0 | 0 | 0 | 0 | 0 | 0 | 0 | 0 |

| | | | | | | | | | | |
|---|---|---|---|---|---|---|---|---|---|---|
| ***OCULAR STRUCTURAL CHANGE, DEPOSIT AND DEGENERATION NEC*** | | | | | | | | | | |
| *CHOROID AND VITREOUS STRUCTURAL CHANGE, DEPOSIT AND DEGENERATION* | | | | | | | | | | |
| VITREOUS DETACHMENT | 0 | 0 | 0 | 0 | 0 | 0 | 0 | 0 | 0 | 0 |
| VITREOUS FLOATERS | 0 | 0 | 0 | 0 | 0 | 0 | 0 | 0 | 0 | 0 |
| ***RETINA, CHOROID AND VITREOUS HAEMORRHAGES AND VASCULAR DISORDERS*** | | | | | | | | | | |
| *RETINAL BLEEDING AND VASCULAR DISORDERS (EXCL RETINOPATHY)* | | | | | | | | | | |
| RETINAL VEIN OCCLUSION | 0 | 0 | 0 | 0 | 0 | 0 | 0 | 0 | 0 | 0 |
| *RETINOPATHIES NEC* | | | | | | | | | | |
| RETINAL EXUDATES | 0 | 0 | 0 | 0 | 0 | 0 | 0 | 0 | 0 | 0 |
| ***VISION DISORDERS*** | | | | | | | | | | |
| *VISUAL DISORDERS NEC* | | | | | | | | | | |
| DIPLOPIA | 0 | 0 | 0 | 0 | 0 | 0 | 0 | 0 | 0 | 0 |
| HALO VISION | 0 | 0 | 0 | 0 | 0 | 0 | 0 | 0 | 0 | 0 |
| METAMORPHOPSIA | 0 | 0 | 0 | 0 | 0 | 0 | 0 | 0 | 0 | 0 |
| PHOTOPSIA | 0 | 0 | 0 | 0 | 0 | 0 | 0 | 0 | 0 | 0 |
| VISION BLURRED | 2 | 1 | 0 | 1 | 0 | 1 | 1 | 0 | 0 | 0 |
| *VISUAL IMPAIRMENT AND BLINDNESS (EXCL COLOUR BLINDNESS)* | | | | | | | | | | |
| BLINDNESS | 0 | 0 | 0 | 0 | 0 | 0 | 0 | 0 | 0 | 0 |
| BLINDNESS TRANSIENT | 0 | 0 | 0 | 0 | 0 | 0 | 0 | 0 | 0 | 0 |
| SUDDEN VISUAL LOSS | 0 | 0 | 0 | 0 | 0 | 0 | 0 | 0 | 0 | 0 |
| VISUAL IMPAIRMENT | 1 | 1 | 0 | 0 | 0 | 1 | 1 | 0 | 0 | 0 |
| **GASTROINTESTINAL DISORDERS** | | | | | | | | | | |
| ***ANAL AND RECTAL CONDITIONS NEC*** | | | | | | | | | | |
| *ANAL AND RECTAL SIGNS AND SYMPTOMS* | | | | | | | | | | |
| ANAL PARAESTHESIA | 0 | 0 | 0 | 0 | 0 | 0 | 0 | 0 | 0 | 0 |
| ***BENIGN NEOPLASMS GASTROINTESTINAL*** | | | | | | | | | | |
| *BENIGN ORAL CAVITY NEOPLASMS* | | | | | | | | | | |
| MOUTH CYST | 0 | 0 | 0 | 0 | 0 | 0 | 0 | 0 | 0 | 0 |
| ***DENTAL AND GINGIVAL CONDITIONS*** | | | | | | | | | | |
| *DENTAL DISORDERS NEC* | | | | | | | | | | |
| TEETHING | 0 | 0 | 0 | 0 | 0 | 0 | 0 | 0 | 0 | 0 |

| | | | | | | | | | | |
|---|---|---|---|---|---|---|---|---|---|---|
| *DENTAL PAIN AND SENSATION DISORDERS* | | | | | | | | | | |
| DENTAL PARAESTHESIA | 0 | 0 | 0 | 0 | 0 | 0 | 0 | 0 | 0 | 0 |
| TOOTHACHE | 1 | 0 | 1 | 0 | 0 | 0 | 0 | 0 | 0 | 0 |
| *GINGIVAL DISORDERS, SIGNS AND SYMPTOMS NEC* | | | | | | | | | | |
| GINGIVAL BLISTER | 0 | 0 | 0 | 0 | 0 | 0 | 0 | 0 | 0 | 0 |
| GINGIVAL PAIN | 1 | 1 | 0 | 0 | 0 | 1 | 1 | 0 | 0 | 0 |
| GINGIVAL SWELLING | 0 | 0 | 0 | 0 | 0 | 0 | 0 | 0 | 0 | 0 |
| ***GASTROINTESTINAL CONDITIONS NEC*** | | | | | | | | | | |
| *GASTROINTESTINAL MUCOSAL DYSTROPHIES AND SECRETION DISORDERS* | | | | | | | | | | |
| BARRETT'S OESOPHAGUS | 0 | 0 | 0 | 0 | 0 | 0 | 0 | 0 | 0 | 0 |
| ***GASTROINTESTINAL HAEMORRHAGES NEC*** | | | | | | | | | | |
| *INTESTINAL HAEMORRHAGES* | | | | | | | | | | |
| RECTAL HAEMORRHAGE | 0 | 0 | 0 | 0 | 0 | 0 | 0 | 0 | 0 | 0 |
| *NON-SITE SPECIFIC GASTROINTESTINAL HAEMORRHAGES* | | | | | | | | | | |
| HAEMATEMESIS | 0 | 0 | 0 | 0 | 0 | 0 | 0 | 0 | 0 | 0 |
| ***GASTROINTESTINAL INFLAMMATORY CONDITIONS*** | | | | | | | | | | |
| *COLITIS (EXCL INFECTIVE)* | | | | | | | | | | |
| COLITIS | 0 | 0 | 0 | 0 | 0 | 0 | 0 | 0 | 0 | 0 |
| COLITIS MICROSCOPIC | 0 | 0 | 0 | 0 | 0 | 0 | 0 | 0 | 0 | 0 |
| COLITIS ULCERATIVE | 0 | 0 | 0 | 0 | 0 | 0 | 0 | 0 | 0 | 0 |
| *GASTRITIS (EXCL INFECTIVE)* | | | | | | | | | | |
| GASTRITIS | 0 | 0 | 0 | 0 | 0 | 0 | 0 | 0 | 0 | 0 |
| REFLUX GASTRITIS | 0 | 0 | 0 | 0 | 0 | 0 | 0 | 0 | 0 | 0 |
| *GASTROINTESTINAL INFLAMMATORY DISORDERS NEC* | | | | | | | | | | |
| GASTROINTESTINAL TRACT IRRITATION | 0 | 0 | 0 | 0 | 0 | 0 | 0 | 0 | 0 | 0 |
| ***GASTROINTESTINAL MOTILITY AND DEFAECATION CONDITIONS*** | | | | | | | | | | |
| *DIARRHOEA (EXCL INFECTIVE)* | | | | | | | | | | |
| DIARRHOEA | 11 | 2 | 0 | 4 | 5 | 9 | 2 | 0 | 3 | 4 |
| *GASTROINTESTINAL ATONIC AND HYPOMOTILITY DISORDERS NEC* | | | | | | | | | | |
| CONSTIPATION | 0 | 0 | 0 | 0 | 0 | 0 | 0 | 0 | 0 | 0 |
| GASTROOESOPHAGEAL REFLUX DISEASE | 0 | 0 | 0 | 0 | 0 | 0 | 0 | 0 | 0 | 0 |
| *GASTROINTESTINAL DYSKINETIC DISORDERS* | | | | | | | | | | |
| CHANGE OF BOWEL HABIT | 0 | 0 | 0 | 0 | 0 | 0 | 0 | 0 | 0 | 0 |
| *GASTROINTESTINAL SPASTIC AND HYPERMOTILITY DISORDERS* | | | | | | | | | | |
| IRRITABLE BOWEL SYNDROME | 0 | 0 | 0 | 0 | 0 | 0 | 0 | 0 | 0 | 0 |
| ***GASTROINTESTINAL SIGNS AND SYMPTOMS*** | | | | | | | | | | |

| | | | | | | | | | | |
|---|---|---|---|---|---|---|---|---|---|---|
| *DYSPEPTIC SIGNS AND SYMPTOMS* | | | | | | | | | | |
| DYSPEPSIA | 2 | 0 | 0 | 1 | 1 | 2 | 0 | 0 | 1 | 1 |
| ERUCTATION | 0 | 0 | 0 | 0 | 0 | 0 | 0 | 0 | 0 | 0 |
| *FAECAL ABNORMALITIES NEC* | | | | | | | | | | |
| FAECALOMA | 0 | 0 | 0 | 0 | 0 | 0 | 0 | 0 | 0 | 0 |
| FAECES DISCOLOURED | 0 | 0 | 0 | 0 | 0 | 0 | 0 | 0 | 0 | 0 |
| FAECES SOFT | 1 | 0 | 0 | 1 | 0 | 1 | 0 | 0 | 1 | 0 |
| *FLATULENCE, BLOATING AND DISTENSION* | | | | | | | | | | |
| ABDOMINAL DISTENSION | 1 | 0 | 0 | 1 | 0 | 1 | 0 | 0 | 1 | 0 |
| FLATULENCE | 0 | 0 | 0 | 0 | 0 | 0 | 0 | 0 | 0 | 0 |
| *GASTROINTESTINAL AND ABDOMINAL PAINS (EXCL ORAL AND THROAT)* | | | | | | | | | | |
| ABDOMINAL PAIN | 3 | 0 | 0 | 2 | 1 | 2 | 0 | 0 | 2 | 0 |
| ABDOMINAL PAIN LOWER | 0 | 0 | 0 | 0 | 0 | 0 | 0 | 0 | 0 | 0 |
| ABDOMINAL PAIN UPPER | 5 | 1 | 2 | 2 | 0 | 3 | 1 | 1 | 1 | 0 |
| GASTROINTESTINAL PAIN | 1 | 0 | 0 | 1 | 0 | 1 | 0 | 0 | 1 | 0 |
| *GASTROINTESTINAL SIGNS AND SYMPTOMS NEC* | | | | | | | | | | |
| ABDOMINAL DISCOMFORT | 0 | 0 | 0 | 0 | 0 | 0 | 0 | 0 | 0 | 0 |
| ABDOMINAL SYMPTOM | 0 | 0 | 0 | 0 | 0 | 0 | 0 | 0 | 0 | 0 |
| ACUTE ABDOMEN | 0 | 0 | 0 | 0 | 0 | 0 | 0 | 0 | 0 | 0 |
| ANAL INCONTINENCE | 0 | 0 | 0 | 0 | 0 | 0 | 0 | 0 | 0 | 0 |
| BREATH ODOUR | 0 | 0 | 0 | 0 | 0 | 0 | 0 | 0 | 0 | 0 |
| DYSPHAGIA | 0 | 0 | 0 | 0 | 0 | 0 | 0 | 0 | 0 | 0 |
| ODYNOPHAGIA | 0 | 0 | 0 | 0 | 0 | 0 | 0 | 0 | 0 | 0 |
| *NAUSEA AND VOMITING SYMPTOMS* | | | | | | | | | | |
| NAUSEA | 87 | 23 | 9 | 45 | 10 | 78 | 23 | 6 | 40 | 9 |
| RETCHING | 0 | 0 | 0 | 0 | 0 | 0 | 0 | 0 | 0 | 0 |
| VOMITING | 16 | 1 | 5 | 9 | 1 | 12 | 1 | 4 | 6 | 1 |
| VOMITING PROJECTILE | 0 | 0 | 0 | 0 | 0 | 0 | 0 | 0 | 0 | 0 |
| ***MALABSORPTION CONDITIONS*** | | | | | | | | | | |
| *MALABSORPTION SYNDROMES* | | | | | | | | | | |
| COELIAC DISEASE | 0 | 0 | 0 | 0 | 0 | 0 | 0 | 0 | 0 | 0 |
| ***ORAL SOFT TISSUE CONDITIONS*** | | | | | | | | | | |
| *ORAL SOFT TISSUE DISORDERS NEC* | | | | | | | | | | |
| CHEILITIS | 0 | 0 | 0 | 0 | 0 | 0 | 0 | 0 | 0 | 0 |
| LIP BLISTER | 0 | 0 | 0 | 0 | 0 | 0 | 0 | 0 | 0 | 0 |

| | | | | | | | | | | |
|---|---|---|---|---|---|---|---|---|---|---|
| ORAL LICHEN PLANUS | 0 | 0 | 0 | 0 | 0 | 0 | 0 | 0 | 0 | 0 |
| *ORAL SOFT TISSUE INFECTIONS* | | | | | | | | | | |
| ANGULAR CHEILITIS | 0 | 0 | 0 | 0 | 0 | 0 | 0 | 0 | 0 | 0 |
| *ORAL SOFT TISSUE SIGNS AND SYMPTOMS* | | | | | | | | | | |
| HYPOAESTHESIA ORAL | 0 | 0 | 0 | 0 | 0 | 0 | 0 | 0 | 0 | 0 |
| LIP PAIN | 0 | 0 | 0 | 0 | 0 | 0 | 0 | 0 | 0 | 0 |
| ORAL DISCOMFORT | 0 | 0 | 0 | 0 | 0 | 0 | 0 | 0 | 0 | 0 |
| ORAL MUCOSAL ROUGHENING | 0 | 0 | 0 | 0 | 0 | 0 | 0 | 0 | 0 | 0 |
| ORAL PAIN | 0 | 0 | 0 | 0 | 0 | 0 | 0 | 0 | 0 | 0 |
| PARAESTHESIA ORAL | 0 | 0 | 0 | 0 | 0 | 0 | 0 | 0 | 0 | 0 |
| LIP SWELLING | 1 | 0 | 1 | 0 | 0 | 1 | 0 | 1 | 0 | 0 |
| MOUTH SWELLING | 0 | 0 | 0 | 0 | 0 | 0 | 0 | 0 | 0 | 0 |
| *STOMATITIS AND ULCERATION* | | | | | | | | | | |
| APHTHOUS ULCER | 0 | 0 | 0 | 0 | 0 | 0 | 0 | 0 | 0 | 0 |
| LIP ULCERATION | 0 | 0 | 0 | 0 | 0 | 0 | 0 | 0 | 0 | 0 |
| MOUTH ULCERATION | 1 | 0 | 0 | 1 | 0 | 1 | 0 | 0 | 1 | 0 |
| STOMATITIS | 0 | 0 | 0 | 0 | 0 | 0 | 0 | 0 | 0 | 0 |
| ***SALIVARY GLAND CONDITIONS*** | | | | | | | | | | |
| *ORAL DRYNESS AND SALIVA ALTERED* | | | | | | | | | | |
| DRY MOUTH | 3 | 0 | 0 | 0 | 3 | 2 | 0 | 0 | 0 | 2 |
| LIP DRY | 1 | 0 | 0 | 1 | 0 | 1 | 0 | 0 | 1 | 0 |
| SALIVARY HYPOSECRETION | 0 | 0 | 0 | 0 | 0 | 0 | 0 | 0 | 0 | 0 |
| ***TONGUE CONDITIONS*** | | | | | | | | | | |
| *TONGUE SIGNS AND SYMPTOMS* | | | | | | | | | | |
| GLOSSODYNIA | 1 | 0 | 0 | 1 | 0 | 0 | 0 | 0 | 0 | 0 |
| SWOLLEN TONGUE | 1 | 0 | 0 | 1 | 0 | 1 | 0 | 0 | 1 | 0 |
| TONGUE COATED | 0 | 0 | 0 | 0 | 0 | 0 | 0 | 0 | 0 | 0 |
| TONGUE DISCOMFORT | 0 | 0 | 0 | 0 | 0 | 0 | 0 | 0 | 0 | 0 |
| TONGUE OEDEMA | 0 | 0 | 0 | 0 | 0 | 0 | 0 | 0 | 0 | 0 |
| TONGUE SPASM | 1 | 0 | 0 | 1 | 0 | 1 | 0 | 0 | 1 | 0 |
| **GENERAL DISORDERS AND ADMINISTRATION SITE CONDITIONS** | | | | | | | | | | |
| ***ADMINISTRATION SITE REACTIONS*** | | | | | | | | | | |
| *ADMINISTRATION SITE REACTIONS NEC* | | | | | | | | | | |
| ADMINISTRATION SITE BRUISE | 0 | 0 | 0 | 0 | 0 | 0 | 0 | 0 | 0 | 0 |
| ADMINISTRATION SITE PAIN | 0 | 0 | 0 | 0 | 0 | 0 | 0 | 0 | 0 | 0 |

| | | | | | | | | | | |
|---|---|---|---|---|---|---|---|---|---|---|
| PUNCTURE SITE BRUISE | 1 | 0 | 0 | 1 | 0 | 1 | 0 | 0 | 1 | 0 |
| PUNCTURE SITE PAIN | 0 | 0 | 0 | 0 | 0 | 0 | 0 | 0 | 0 | 0 |
| *APPLICATION SITE REACTIONS* | | | | | | | | | | |
| APPLICATION SITE BRUISE | 1 | 0 | 0 | 0 | 1 | 1 | 0 | 0 | 0 | 1 |
| APPLICATION SITE ERYTHEMA | 0 | 0 | 0 | 0 | 0 | 0 | 0 | 0 | 0 | 0 |
| APPLICATION SITE PAIN | 2 | 0 | 0 | 2 | 0 | 2 | 0 | 0 | 2 | 0 |
| *IMPLANT AND CATHETER SITE REACTIONS* | | | | | | | | | | |
| IMPLANT SITE PAIN | 0 | 0 | 0 | 0 | 0 | 0 | 0 | 0 | 0 | 0 |
| IMPLANT SITE WARMTH | 0 | 0 | 0 | 0 | 0 | 0 | 0 | 0 | 0 | 0 |
| *INFUSION SITE REACTIONS* | | | | | | | | | | |
| INFUSION SITE PAIN | 0 | 0 | 0 | 0 | 0 | 0 | 0 | 0 | 0 | 0 |
| INFUSION SITE SCAB | 0 | 0 | 0 | 0 | 0 | 0 | 0 | 0 | 0 | 0 |
| INFUSION SITE WARMTH | 0 | 0 | 0 | 0 | 0 | 0 | 0 | 0 | 0 | 0 |
| *INJECTION SITE REACTIONS* | | | | | | | | | | |
| INJECTION SITE BRUISING | 2 | 0 | 0 | 2 | 0 | 2 | 0 | 0 | 2 | 0 |
| INJECTION SITE DISCOMFORT | 0 | 0 | 0 | 0 | 0 | 0 | 0 | 0 | 0 | 0 |
| INJECTION SITE ERYTHEMA | 5 | 2 | 1 | 1 | 1 | 4 | 2 | 1 | 1 | 0 |
| INJECTION SITE HYPERSENSITIVITY | 0 | 0 | 0 | 0 | 0 | 0 | 0 | 0 | 0 | 0 |
| INJECTION SITE INFLAMMATION | 0 | 0 | 0 | 0 | 0 | 0 | 0 | 0 | 0 | 0 |
| INJECTION SITE INJURY | 0 | 0 | 0 | 0 | 0 | 0 | 0 | 0 | 0 | 0 |
| INJECTION SITE IRRITATION | 0 | 0 | 0 | 0 | 0 | 0 | 0 | 0 | 0 | 0 |
| INJECTION SITE JOINT PAIN | 1 | 0 | 0 | 0 | 1 | 0 | 0 | 0 | 0 | 0 |
| INJECTION SITE MASS | 4 | 2 | 1 | 0 | 1 | 4 | 2 | 1 | 0 | 1 |
| INJECTION SITE NODULE | 0 | 0 | 0 | 0 | 0 | 0 | 0 | 0 | 0 | 0 |
| INJECTION SITE OEDEMA | 0 | 0 | 0 | 0 | 0 | 0 | 0 | 0 | 0 | 0 |
| INJECTION SITE PAIN | 49 | 15 | 4 | 23 | 7 | 46 | 15 | 2 | 22 | 7 |
| INJECTION SITE PAPULE | 0 | 0 | 0 | 0 | 0 | 0 | 0 | 0 | 0 | 0 |
| INJECTION SITE PARAESTHESIA | 0 | 0 | 0 | 0 | 0 | 0 | 0 | 0 | 0 | 0 |
| INJECTION SITE PRURITUS | 3 | 2 | 0 | 1 | 0 | 3 | 2 | 0 | 1 | 0 |
| INJECTION SITE RASH | 3 | 2 | 1 | 0 | 0 | 3 | 2 | 1 | 0 | 0 |
| INJECTION SITE REACTION | 0 | 0 | 0 | 0 | 0 | 0 | 0 | 0 | 0 | 0 |
| INJECTION SITE SCAB | 0 | 0 | 0 | 0 | 0 | 0 | 0 | 0 | 0 | 0 |
| INJECTION SITE SWELLING | 2 | 0 | 0 | 2 | 0 | 2 | 0 | 0 | 2 | 0 |
| INJECTION SITE URTICARIA | 3 | 0 | 0 | 0 | 3 | 3 | 0 | 0 | 0 | 3 |
| INJECTION SITE WARMTH | 4 | 2 | 0 | 1 | 1 | 4 | 2 | 0 | 1 | 1 |

| | | | | | | | | | | |
|---|---|---|---|---|---|---|---|---|---|---|
| *INSTILLATION SITE REACTIONS* | | | | | | | | | | |
| INSTILLATION SITE PRURITUS | 0 | 0 | 0 | 0 | 0 | 0 | 0 | 0 | 0 | 0 |
| INSTILLATION SITE WARMTH | 0 | 0 | 0 | 0 | 0 | 0 | 0 | 0 | 0 | 0 |
| *VACCINATION SITE REACTIONS* | | | | | | | | | | |
| SHOULDER INJURY RELATED TO VACCINE ADMINISTRATION | 0 | 0 | 0 | 0 | 0 | 0 | 0 | 0 | 0 | 0 |
| VACCINATION SITE BRUISING | 0 | 0 | 0 | 0 | 0 | 0 | 0 | 0 | 0 | 0 |
| VACCINATION SITE DISCOMFORT | 0 | 0 | 0 | 0 | 0 | 0 | 0 | 0 | 0 | 0 |
| VACCINATION SITE ERYTHEMA | 0 | 0 | 0 | 0 | 0 | 0 | 0 | 0 | 0 | 0 |
| VACCINATION SITE JOINT ERYTHEMA | 0 | 0 | 0 | 0 | 0 | 0 | 0 | 0 | 0 | 0 |
| VACCINATION SITE JOINT PAIN | 1 | 0 | 0 | 1 | 0 | 1 | 0 | 0 | 1 | 0 |
| VACCINATION SITE MASS | 1 | 1 | 0 | 0 | 0 | 1 | 1 | 0 | 0 | 0 |
| VACCINATION SITE PAIN | 5 | 0 | 0 | 4 | 1 | 5 | 0 | 0 | 4 | 1 |
| VACCINATION SITE RASH | 0 | 0 | 0 | 0 | 0 | 0 | 0 | 0 | 0 | 0 |
| VACCINATION SITE SWELLING | 0 | 0 | 0 | 0 | 0 | 0 | 0 | 0 | 0 | 0 |
| VACCINATION SITE WARMTH | 0 | 0 | 0 | 0 | 0 | 0 | 0 | 0 | 0 | 0 |
| ***BODY TEMPERATURE CONDITIONS*** | | | | | | | | | | |
| *BODY TEMPERATURE ALTERED* | | | | | | | | | | |
| HYPERTHERMIA | 0 | 0 | 0 | 0 | 0 | 0 | 0 | 0 | 0 | 0 |
| HYPOTHERMIA | 0 | 0 | 0 | 0 | 0 | 0 | 0 | 0 | 0 | 0 |
| *FEBRILE DISORDERS* | | | | | | | | | | |
| PYREXIA | 176 | 33 | 48 | 75 | 20 | 155 | 33 | 39 | 64 | 19 |
| ***FATAL OUTCOMES*** | | | | | | | | | | |
| *DEATH AND SUDDEN DEATH* | | | | | | | | | | |
| DEATH | 0 | 0 | 0 | 0 | 0 | 0 | 0 | 0 | 0 | 0 |
| ***GENERAL SYSTEM DISORDERS NEC*** | | | | | | | | | | |
| *ADVERSE EFFECT ABSENT* | | | | | | | | | | |
| NO ADVERSE EVENT | 0 | 0 | 0 | 0 | 0 | 0 | 0 | 0 | 0 | 0 |
| *ASTHENIC CONDITIONS* | | | | | | | | | | |
| ASTHENIA | 10 | 5 | 1 | 2 | 2 | 7 | 5 | 0 | 1 | 1 |
| CHRONIC FATIGUE SYNDROME | 0 | 0 | 0 | 0 | 0 | 0 | 0 | 0 | 0 | 0 |
| DECREASED ACTIVITY | 0 | 0 | 0 | 0 | 0 | 0 | 0 | 0 | 0 | 0 |
| FATIGUE | 278 | 63 | 46 | 121 | 48 | 245 | 63 | 37 | 106 | 39 |
| MALAISE | 33 | 4 | 3 | 18 | 8 | 27 | 4 | 3 | 15 | 5 |
| SLUGGISHNESS | 1 | 0 | 0 | 0 | 1 | 1 | 0 | 0 | 0 | 1 |
| *FEELINGS AND SENSATIONS NEC* | | | | | | | | | | |

| | | | | | | | | | | |
|---|---|---|---|---|---|---|---|---|---|---|
| CHILLS | 104 | 10 | 22 | 62 | 10 | 92 | 10 | 17 | 57 | 8 |
| FEELING ABNORMAL | 6 | 1 | 1 | 1 | 3 | 6 | 1 | 1 | 1 | 3 |
| FEELING COLD | 19 | 0 | 3 | 11 | 5 | 12 | 0 | 0 | 8 | 4 |
| FEELING HOT | 5 | 1 | 1 | 2 | 1 | 4 | 1 | 1 | 2 | 0 |
| FEELING JITTERY | 0 | 0 | 0 | 0 | 0 | 0 | 0 | 0 | 0 | 0 |
| FEELING OF BODY TEMPERATURE CHANGE | 4 | 0 | 2 | 2 | 0 | 4 | 0 | 2 | 2 | 0 |
| FEELING OF RELAXATION | 0 | 0 | 0 | 0 | 0 | 0 | 0 | 0 | 0 | 0 |
| HANGOVER | 0 | 0 | 0 | 0 | 0 | 0 | 0 | 0 | 0 | 0 |
| HUNGER | 1 | 0 | 1 | 0 | 0 | 1 | 0 | 1 | 0 | 0 |
| SENSATION OF BLOOD FLOW | 0 | 0 | 0 | 0 | 0 | 0 | 0 | 0 | 0 | 0 |
| THIRST | 0 | 0 | 0 | 0 | 0 | 0 | 0 | 0 | 0 | 0 |
| *GAIT DISTURBANCES* | | | | | | | | | | |
| GAIT DISTURBANCE | 0 | 0 | 0 | 0 | 0 | 0 | 0 | 0 | 0 | 0 |
| GAIT INABILITY | 0 | 0 | 0 | 0 | 0 | 0 | 0 | 0 | 0 | 0 |
| *GENERAL SIGNS AND SYMPTOMS NEC* | | | | | | | | | | |
| CRYING | 0 | 0 | 0 | 0 | 0 | 0 | 0 | 0 | 0 | 0 |
| ENERGY INCREASED | 0 | 0 | 0 | 0 | 0 | 0 | 0 | 0 | 0 | 0 |
| EXERCISE TOLERANCE DECREASED | 0 | 0 | 0 | 0 | 0 | 0 | 0 | 0 | 0 | 0 |
| GENERAL SYMPTOM | 0 | 0 | 0 | 0 | 0 | 0 | 0 | 0 | 0 | 0 |
| ILLNESS | 14 | 5 | 4 | 5 | 0 | 13 | 5 | 3 | 5 | 0 |
| INFLUENZA LIKE ILLNESS | 45 | 1 | 2 | 35 | 7 | 33 | 1 | 1 | 25 | 6 |
| LOCAL REACTION | 0 | 0 | 0 | 0 | 0 | 0 | 0 | 0 | 0 | 0 |
| PERIPHERAL SWELLING | 30 | 5 | 3 | 18 | 4 | 26 | 5 | 3 | 15 | 3 |
| SWELLING | 6 | 3 | 0 | 3 | 0 | 5 | 3 | 0 | 2 | 0 |
| SWELLING FACE | 0 | 0 | 0 | 0 | 0 | 0 | 0 | 0 | 0 | 0 |
| TISSUE IRRITATION | 0 | 0 | 0 | 0 | 0 | 0 | 0 | 0 | 0 | 0 |
| *INFLAMMATIONS* | | | | | | | | | | |
| INFLAMMATION | 0 | 0 | 0 | 0 | 0 | 0 | 0 | 0 | 0 | 0 |
| SYSTEMIC INFLAMMATORY RESPONSE SYNDROME | 0 | 0 | 0 | 0 | 0 | 0 | 0 | 0 | 0 | 0 |
| *OEDEMA NEC* | | | | | | | | | | |
| OEDEMA | 0 | 0 | 0 | 0 | 0 | 0 | 0 | 0 | 0 | 0 |
| OEDEMA PERIPHERAL | 0 | 0 | 0 | 0 | 0 | 0 | 0 | 0 | 0 | 0 |
| *PAIN AND DISCOMFORT NEC* | | | | | | | | | | |
| AXILLARY PAIN | 14 | 7 | 1 | 6 | 0 | 14 | 7 | 1 | 6 | 0 |

| | | | | | | | | | | |
|---|---|---|---|---|---|---|---|---|---|---|
| CHEST DISCOMFORT | 4 | 0 | 0 | 4 | 0 | 4 | 0 | 0 | 4 | 0 |
| CHEST PAIN | 7 | 1 | 0 | 4 | 2 | 5 | 1 | 0 | 3 | 1 |
| DISCOMFORT | 3 | 0 | 0 | 2 | 1 | 1 | 0 | 0 | 1 | 0 |
| FACIAL PAIN | 0 | 0 | 0 | 0 | 0 | 0 | 0 | 0 | 0 | 0 |
| HERNIA PAIN | 0 | 0 | 0 | 0 | 0 | 0 | 0 | 0 | 0 | 0 |
| INFLAMMATORY PAIN | 0 | 0 | 0 | 0 | 0 | 0 | 0 | 0 | 0 | 0 |
| NON-CARDIAC CHEST PAIN | 0 | 0 | 0 | 0 | 0 | 0 | 0 | 0 | 0 | 0 |
| PAIN | 75 | 20 | 18 | 28 | 9 | 63 | 20 | 15 | 21 | 7 |
| TENDERNESS | 17 | 4 | 0 | 11 | 2 | 15 | 4 | 0 | 9 | 2 |
| ***THERAPEUTIC AND NONTHERAPEUTIC EFFECTS (EXCL TOXICITY)*** | | | | | | | | | | |
| *THERAPEUTIC AND NONTHERAPEUTIC RESPONSES* | | | | | | | | | | |
| ADVERSE DRUG REACTION | 0 | 0 | 0 | 0 | 0 | 0 | 0 | 0 | 0 | 0 |
| ADVERSE EVENT | 0 | 0 | 0 | 0 | 0 | 0 | 0 | 0 | 0 | 0 |
| ADVERSE REACTION | 0 | 0 | 0 | 0 | 0 | 0 | 0 | 0 | 0 | 0 |
| IMMEDIATE POST-INJECTION REACTION | 0 | 0 | 0 | 0 | 0 | 0 | 0 | 0 | 0 | 0 |
| **HEPATOBILIARY DISORDERS** | | | | | | | | | | |
| ***HEPATIC AND HEPATOBILIARY DISORDERS*** | | | | | | | | | | |
| *HEPATOBILIARY SIGNS AND SYMPTOMS* | | | | | | | | | | |
| HEPATIC PAIN | 0 | 0 | 0 | 0 | 0 | 0 | 0 | 0 | 0 | 0 |
| *HEPATOCELLULAR DAMAGE AND HEPATITIS NEC* | | | | | | | | | | |
| LIVER INJURY | 0 | 0 | 0 | 0 | 0 | 0 | 0 | 0 | 0 | 0 |
| **IMMUNE SYSTEM DISORDERS** | | | | | | | | | | |
| ***ALLERGIC CONDITIONS*** | | | | | | | | | | |
| *ALLERGIC CONDITIONS NEC* | | | | | | | | | | |
| HYPERSENSITIVITY | 0 | 0 | 0 | 0 | 0 | 0 | 0 | 0 | 0 | 0 |
| MULTIPLE ALLERGIES | 0 | 0 | 0 | 0 | 0 | 0 | 0 | 0 | 0 | 0 |
| *ALLERGIES TO FOODS, FOOD ADDITIVES, DRUGS AND OTHER CHEMICALS* | | | | | | | | | | |
| ALLERGY TO CHEMICALS | 0 | 0 | 0 | 0 | 0 | 0 | 0 | 0 | 0 | 0 |
| *ANAPHYLACTIC AND ANAPHYLACTOID RESPONSES* | | | | | | | | | | |
| ANAPHYLACTIC REACTION | 0 | 0 | 0 | 0 | 0 | 0 | 0 | 0 | 0 | 0 |
| *ATOPIC DISORDERS* | | | | | | | | | | |
| SEASONAL ALLERGY | 0 | 0 | 0 | 0 | 0 | 0 | 0 | 0 | 0 | 0 |
| ***IMMUNE DISORDERS NEC*** | | | | | | | | | | |
| *IMMUNE AND ASSOCIATED CONDITIONS NEC* | | | | | | | | | | |

| | | | | | | | | | | |
|---|---|---|---|---|---|---|---|---|---|---|
| BACILLE CALMETTE-GUERIN SCAR REACTIVATION | 0 | 0 | 0 | 0 | 0 | 0 | 0 | 0 | 0 | 0 |
| SENSITISATION | 0 | 0 | 0 | 0 | 0 | 0 | 0 | 0 | 0 | 0 |
| **INFECTIONS AND INFESTATIONS** | | | | | | | | | | |
| ***BACTERIAL INFECTIOUS DISORDERS*** | | | | | | | | | | |
| *BACTERIAL INFECTIONS NEC* | | | | | | | | | | |
| CELLULITIS | 2 | 1 | 1 | 0 | 0 | 1 | 1 | 0 | 0 | 0 |
| PERIORBITAL CELLULITIS | 0 | 0 | 0 | 0 | 0 | 0 | 0 | 0 | 0 | 0 |
| *HELICOBACTER INFECTIONS* | | | | | | | | | | |
| HELICOBACTER GASTRITIS | 0 | 0 | 0 | 0 | 0 | 0 | 0 | 0 | 0 | 0 |
| *STAPHYLOCOCCAL INFECTIONS* | | | | | | | | | | |
| FURUNCLE | 0 | 0 | 0 | 0 | 0 | 0 | 0 | 0 | 0 | 0 |
| ***FUNGAL INFECTIOUS DISORDERS*** | | | | | | | | | | |
| *CANDIDA INFECTIONS* | | | | | | | | | | |
| CANDIDA INFECTION | 0 | 0 | 0 | 0 | 0 | 0 | 0 | 0 | 0 | 0 |
| VULVOVAGINAL CANDIDIASIS | 0 | 0 | 0 | 0 | 0 | 0 | 0 | 0 | 0 | 0 |
| ***INFECTIONS - PATHOGEN UNSPECIFIED*** | | | | | | | | | | |
| *ABDOMINAL AND GASTROINTESTINAL INFECTIONS* | | | | | | | | | | |
| APPENDICITIS | 0 | 0 | 0 | 0 | 0 | 0 | 0 | 0 | 0 | 0 |
| DIARRHOEA INFECTIOUS | 1 | 0 | 0 | 1 | 0 | 1 | 0 | 0 | 1 | 0 |
| GASTROINTESTINAL INFECTION | 0 | 0 | 0 | 0 | 0 | 0 | 0 | 0 | 0 | 0 |
| *BREAST INFECTIONS* | | | | | | | | | | |
| MASTITIS | 1 | 0 | 0 | 1 | 0 | 1 | 0 | 0 | 1 | 0 |
| *CENTRAL NERVOUS SYSTEM AND SPINAL INFECTIONS* | | | | | | | | | | |
| MYELITIS | 0 | 0 | 0 | 0 | 0 | 0 | 0 | 0 | 0 | 0 |
| *DENTAL AND ORAL SOFT TISSUE INFECTIONS* | | | | | | | | | | |
| ABSCESS ORAL | 0 | 0 | 0 | 0 | 0 | 0 | 0 | 0 | 0 | 0 |
| ORAL PUSTULE | 0 | 0 | 0 | 0 | 0 | 0 | 0 | 0 | 0 | 0 |
| PERICORONITIS | 0 | 0 | 0 | 0 | 0 | 0 | 0 | 0 | 0 | 0 |
| TOOTH ABSCESS | 0 | 0 | 0 | 0 | 0 | 0 | 0 | 0 | 0 | 0 |
| *EAR INFECTIONS* | | | | | | | | | | |
| EAR INFECTION | 0 | 0 | 0 | 0 | 0 | 0 | 0 | 0 | 0 | 0 |
| LABYRINTHITIS | 0 | 0 | 0 | 0 | 0 | 0 | 0 | 0 | 0 | 0 |
| OTITIS EXTERNA | 0 | 0 | 0 | 0 | 0 | 0 | 0 | 0 | 0 | 0 |
| *EYE AND EYELID INFECTIONS* | | | | | | | | | | |
| CONJUNCTIVITIS | 1 | 0 | 1 | 0 | 0 | 0 | 0 | 0 | 0 | 0 |

| | | | | | | | | | | |
|---|---|---|---|---|---|---|---|---|---|---|
| HORDEOLUM | 0 | 0 | 0 | 0 | 0 | 0 | 0 | 0 | 0 | 0 |
| *INFECTIONS NEC* | | | | | | | | | | |
| ABSCESS | 0 | 0 | 0 | 0 | 0 | 0 | 0 | 0 | 0 | 0 |
| INFECTION | 1 | 1 | 0 | 0 | 0 | 1 | 1 | 0 | 0 | 0 |
| INJECTION SITE INFECTION | 0 | 0 | 0 | 0 | 0 | 0 | 0 | 0 | 0 | 0 |
| LOCALISED INFECTION | 1 | 1 | 0 | 0 | 0 | 1 | 1 | 0 | 0 | 0 |
| WOUND INFECTION | 0 | 0 | 0 | 0 | 0 | 0 | 0 | 0 | 0 | 0 |
| *LOWER RESPIRATORY TRACT AND LUNG INFECTIONS* | | | | | | | | | | |
| LOWER RESPIRATORY TRACT INFECTION | 1 | 1 | 0 | 0 | 0 | 1 | 1 | 0 | 0 | 0 |
| PNEUMONIA | 0 | 0 | 0 | 0 | 0 | 0 | 0 | 0 | 0 | 0 |
| *MALE REPRODUCTIVE TRACT INFECTIONS* | | | | | | | | | | |
| ORCHITIS | 0 | 0 | 0 | 0 | 0 | 0 | 0 | 0 | 0 | 0 |
| *SEPSIS, BACTERAEMIA, VIRAEMIA AND FUNGAEMIA NEC* | | | | | | | | | | |
| SEPTIC RASH | 0 | 0 | 0 | 0 | 0 | 0 | 0 | 0 | 0 | 0 |
| *SKIN STRUCTURES AND SOFT TISSUE INFECTIONS* | | | | | | | | | | |
| INFECTED DERMAL CYST | 0 | 0 | 0 | 0 | 0 | 0 | 0 | 0 | 0 | 0 |
| INJECTION SITE PUSTULE | 0 | 0 | 0 | 0 | 0 | 0 | 0 | 0 | 0 | 0 |
| SKIN INFECTION | 0 | 0 | 0 | 0 | 0 | 0 | 0 | 0 | 0 | 0 |
| *UPPER RESPIRATORY TRACT INFECTIONS* | | | | | | | | | | |
| LARYNGITIS | 1 | 1 | 0 | 0 | 0 | 1 | 1 | 0 | 0 | 0 |
| NASOPHARYNGITIS | 18 | 2 | 2 | 13 | 1 | 16 | 2 | 2 | 11 | 1 |
| PHARYNGITIS | 0 | 0 | 0 | 0 | 0 | 0 | 0 | 0 | 0 | 0 |
| RHINITIS | 0 | 0 | 0 | 0 | 0 | 0 | 0 | 0 | 0 | 0 |
| SINUSITIS | 0 | 0 | 0 | 0 | 0 | 0 | 0 | 0 | 0 | 0 |
| TONSILLITIS | 0 | 0 | 0 | 0 | 0 | 0 | 0 | 0 | 0 | 0 |
| *URINARY TRACT INFECTIONS* | | | | | | | | | | |
| CYSTITIS | 0 | 0 | 0 | 0 | 0 | 0 | 0 | 0 | 0 | 0 |
| URINARY TRACT INFECTION | 0 | 0 | 0 | 0 | 0 | 0 | 0 | 0 | 0 | 0 |
| ***PROTOZOAL INFECTIOUS DISORDERS*** | | | | | | | | | | |
| *TRYPANOSOMAL INFECTIONS* | | | | | | | | | | |
| AFRICAN TRYPANOSOMIASIS | 0 | 0 | 0 | 0 | 0 | 0 | 0 | 0 | 0 | 0 |
| ***RICKETTSIAL INFECTIOUS DISORDERS*** | | | | | | | | | | |
| *COXIELLA INFECTIONS* | | | | | | | | | | |
| Q FEVER | 0 | 0 | 0 | 0 | 0 | 0 | 0 | 0 | 0 | 0 |
| ***VIRAL INFECTIOUS DISORDERS*** | | | | | | | | | | |
| *CORONAVIRUS INFECTIONS* | | | | | | | | | | |

| | | | | | | | | | | |
|---|---|---|---|---|---|---|---|---|---|---|
| COVID-19 | 3 | 0 | 0 | 0 | 3 | 2 | 0 | 0 | 0 | 2 |
| *HERPES VIRAL INFECTIONS* | | | | | | | | | | |
| GENITAL HERPES | 0 | 0 | 0 | 0 | 0 | 0 | 0 | 0 | 0 | 0 |
| HERPES OPHTHALMIC | 0 | 0 | 0 | 0 | 0 | 0 | 0 | 0 | 0 | 0 |
| HERPES SIMPLEX | 0 | 0 | 0 | 0 | 0 | 0 | 0 | 0 | 0 | 0 |
| HERPES ZOSTER | 0 | 0 | 0 | 0 | 0 | 0 | 0 | 0 | 0 | 0 |
| NASAL HERPES | 0 | 0 | 0 | 0 | 0 | 0 | 0 | 0 | 0 | 0 |
| OPHTHALMIC HERPES ZOSTER | 0 | 0 | 0 | 0 | 0 | 0 | 0 | 0 | 0 | 0 |
| ORAL HERPES | 1 | 1 | 0 | 0 | 0 | 1 | 1 | 0 | 0 | 0 |
| *INFLUENZA VIRAL INFECTIONS* | | | | | | | | | | |
| H2N2 INFLUENZA | 0 | 0 | 0 | 0 | 0 | 0 | 0 | 0 | 0 | 0 |
| INFLUENZA | 47 | 2 | 10 | 29 | 6 | 36 | 2 | 8 | 21 | 5 |
| *RETROVIRAL INFECTIONS* | | | | | | | | | | |
| AIDS RELATED COMPLEX | 0 | 0 | 0 | 0 | 0 | 0 | 0 | 0 | 0 | 0 |
| *VIRAL INFECTIONS NEC* | | | | | | | | | | |
| GASTROENTERITIS VIRAL | 0 | 0 | 0 | 0 | 0 | 0 | 0 | 0 | 0 | 0 |
| SWEATING FEVER | 0 | 0 | 0 | 0 | 0 | 0 | 0 | 0 | 0 | 0 |
| VESTIBULAR NEURONITIS | 1 | 0 | 0 | 0 | 1 | 0 | 0 | 0 | 0 | 0 |
| VIRAL DIARRHOEA | 0 | 0 | 0 | 0 | 0 | 0 | 0 | 0 | 0 | 0 |
| VIRAL RASH | 0 | 0 | 0 | 0 | 0 | 0 | 0 | 0 | 0 | 0 |
| VIRAL UPPER RESPIRATORY TRACT INFECTION | 0 | 0 | 0 | 0 | 0 | 0 | 0 | 0 | 0 | 0 |
| **INJURY, POISONING AND PROCEDURAL COMPLICATIONS** | | | | | | | | | | |
| ***BONE AND JOINT INJURIES*** | | | | | | | | | | |
| *FRACTURES AND DISLOCATIONS NEC* | | | | | | | | | | |
| JOINT DISLOCATION | 0 | 0 | 0 | 0 | 0 | 0 | 0 | 0 | 0 | 0 |
| ***EXPOSURES, CHEMICAL INJURIES AND POISONING*** | | | | | | | | | | |
| *POISONING AND TOXICITY* | | | | | | | | | | |
| SYSTEMIC TOXICITY | 0 | 0 | 0 | 0 | 0 | 0 | 0 | 0 | 0 | 0 |
| TOXICITY TO VARIOUS AGENTS | 0 | 0 | 0 | 0 | 0 | 0 | 0 | 0 | 0 | 0 |
| ***INJURIES BY PHYSICAL AGENTS*** | | | | | | | | | | |
| *CONDITIONS CAUSED BY COLD* | | | | | | | | | | |
| CHILLBLAINS | 0 | 0 | 0 | 0 | 0 | 0 | 0 | 0 | 0 | 0 |
| *HEAT INJURIES (EXCL THERMAL BURNS)* | | | | | | | | | | |
| HEAT EXHAUSTION | 0 | 0 | 0 | 0 | 0 | 0 | 0 | 0 | 0 | 0 |
| HEAT OEDEMA | 0 | 0 | 0 | 0 | 0 | 0 | 0 | 0 | 0 | 0 |
| *RADIATION INJURIES* | | | | | | | | | | |

| | | | | | | | | | | |
|---|---|---|---|---|---|---|---|---|---|---|
| SUNBURN | 0 | 0 | 0 | 0 | 0 | 0 | 0 | 0 | 0 | 0 |
| *THERMAL BURNS* | | | | | | | | | | |
| THERMAL BURN | 0 | 0 | 0 | 0 | 0 | 0 | 0 | 0 | 0 | 0 |
| THERMAL BURNS OF EYE | 0 | 0 | 0 | 0 | 0 | 0 | 0 | 0 | 0 | 0 |
| ***INJURIES NEC*** | | | | | | | | | | |
| *CHEST AND RESPIRATORY TRACT INJURIES NEC* | | | | | | | | | | |
| CHEST CRUSHING | 0 | 0 | 0 | 0 | 0 | 0 | 0 | 0 | 0 | 0 |
| *EYE INJURIES NEC* | | | | | | | | | | |
| EYE CONTUSION | 0 | 0 | 0 | 0 | 0 | 0 | 0 | 0 | 0 | 0 |
| *MUSCLE, TENDON AND LIGAMENT INJURIES* | | | | | | | | | | |
| LIGAMENT SPRAIN | 0 | 0 | 0 | 0 | 0 | 0 | 0 | 0 | 0 | 0 |
| MUSCLE INJURY | 0 | 0 | 0 | 0 | 0 | 0 | 0 | 0 | 0 | 0 |
| MUSCLE STRAIN | 0 | 0 | 0 | 0 | 0 | 0 | 0 | 0 | 0 | 0 |
| *NERVE INJURIES NEC* | | | | | | | | | | |
| NERVE INJURY | 0 | 0 | 0 | 0 | 0 | 0 | 0 | 0 | 0 | 0 |
| *NON-SITE SPECIFIC INJURIES NEC* | | | | | | | | | | |
| ARTHROPOD STING | 0 | 0 | 0 | 0 | 0 | 0 | 0 | 0 | 0 | 0 |
| BITE | 0 | 0 | 0 | 0 | 0 | 0 | 0 | 0 | 0 | 0 |
| FALL | 1 | 0 | 0 | 1 | 0 | 1 | 0 | 0 | 1 | 0 |
| INFLAMMATION OF WOUND | 0 | 0 | 0 | 0 | 0 | 0 | 0 | 0 | 0 | 0 |
| TISSUE INJURY | 0 | 0 | 0 | 0 | 0 | 0 | 0 | 0 | 0 | 0 |
| WOUND COMPLICATION | 0 | 0 | 0 | 0 | 0 | 0 | 0 | 0 | 0 | 0 |
| WOUND SECRETION | 0 | 0 | 0 | 0 | 0 | 0 | 0 | 0 | 0 | 0 |
| *SITE SPECIFIC INJURIES NEC* | | | | | | | | | | |
| LIMB INJURY | 0 | 0 | 0 | 0 | 0 | 0 | 0 | 0 | 0 | 0 |
| *SKIN INJURIES NEC* | | | | | | | | | | |
| CONTUSION | 7 | 3 | 0 | 4 | 0 | 6 | 3 | 0 | 3 | 0 |
| SCAR | 0 | 0 | 0 | 0 | 0 | 0 | 0 | 0 | 0 | 0 |
| SKIN WOUND | 0 | 0 | 0 | 0 | 0 | 0 | 0 | 0 | 0 | 0 |
| ***PROCEDURAL RELATED INJURIES AND COMPLICATIONS NEC*** | | | | | | | | | | |
| *CARDIAC AND VASCULAR PROCEDURAL COMPLICATIONS* | | | | | | | | | | |
| CARDIAC PROCEDURE COMPLICATION | 0 | 0 | 0 | 0 | 0 | 0 | 0 | 0 | 0 | 0 |
| *GASTROINTESTINAL AND HEPATOBILIARY PROCEDURAL COMPLICATIONS* | | | | | | | | | | |
| PROCEDURAL NAUSEA | 0 | 0 | 0 | 0 | 0 | 0 | 0 | 0 | 0 | 0 |
| *NEUROLOGICAL AND PSYCHIATRIC PROCEDURAL COMPLICATIONS* | | | | | | | | | | |
| PROCEDURAL DIZZINESS | 0 | 0 | 0 | 0 | 0 | 0 | 0 | 0 | 0 | 0 |

| | | | | | | | | | | |
|---|---|---|---|---|---|---|---|---|---|---|
| *NON-SITE SPECIFIC PROCEDURAL COMPLICATIONS* | | | | | | | | | | |
| INCISION SITE PAIN | 0 | 0 | 0 | 0 | 0 | 0 | 0 | 0 | 0 | 0 |
| INCISION SITE SWELLING | 0 | 0 | 0 | 0 | 0 | 0 | 0 | 0 | 0 | 0 |
| INJECTION RELATED REACTION | 4 | 0 | 0 | 1 | 3 | 4 | 0 | 0 | 1 | 3 |
| POST PROCEDURAL COMPLICATION | 2 | 0 | 0 | 1 | 1 | 0 | 0 | 0 | 0 | 0 |
| PROCEDURAL PAIN | 0 | 0 | 0 | 0 | 0 | 0 | 0 | 0 | 0 | 0 |
| *VACCINATION RELATED COMPLICATIONS* | | | | | | | | | | |
| IMMUNISATION REACTION | 0 | 0 | 0 | 0 | 0 | 0 | 0 | 0 | 0 | 0 |
| **INVESTIGATIONS** | | | | | | | | | | |
| ***CARDIAC AND VASCULAR INVESTIGATIONS (EXCL ENZYME TESTS)*** | | | | | | | | | | |
| *HEART RATE AND PULSE INVESTIGATIONS* | | | | | | | | | | |
| HEART RATE | 1 | 0 | 0 | 1 | 0 | 1 | 0 | 0 | 1 | 0 |
| HEART RATE DECREASED | 0 | 0 | 0 | 0 | 0 | 0 | 0 | 0 | 0 | 0 |
| HEART RATE INCREASED | 3 | 2 | 0 | 1 | 0 | 3 | 2 | 0 | 1 | 0 |
| HEART RATE IRREGULAR | 0 | 0 | 0 | 0 | 0 | 0 | 0 | 0 | 0 | 0 |
| *VASCULAR TESTS NEC (INCL BLOOD PRESSURE)* | | | | | | | | | | |
| BLOOD PRESSURE DECREASED | 0 | 0 | 0 | 0 | 0 | 0 | 0 | 0 | 0 | 0 |
| BLOOD PRESSURE INCREASED | 0 | 0 | 0 | 0 | 0 | 0 | 0 | 0 | 0 | 0 |
| BLOOD PRESSURE MEASUREMENT | 1 | 0 | 0 | 1 | 0 | 0 | 0 | 0 | 0 | 0 |
| ***ENDOCRINE INVESTIGATIONS (INCL SEX HORMONES)*** | | | | | | | | | | |
| *ENDOCRINE ANALYSES AND IMAGING NEC* | | | | | | | | | | |
| HORMONE LEVEL ABNORMAL | 0 | 0 | 0 | 0 | 0 | 0 | 0 | 0 | 0 | 0 |
| *PITUITARY ANALYSES ANTERIOR* | | | | | | | | | | |
| BLOOD FOLLICLE STIMULATING HORMONE INCREASED | 1 | 0 | 0 | 1 | 0 | 1 | 0 | 0 | 1 | 0 |
| BLOOD LUTEINISING HORMONE | 0 | 0 | 0 | 0 | 0 | 0 | 0 | 0 | 0 | 0 |
| *THYROID ANALYSES* | | | | | | | | | | |
| TRI-IODOTHYRONINE | 0 | 0 | 0 | 0 | 0 | 0 | 0 | 0 | 0 | 0 |
| ***HAEMATOLOGY INVESTIGATIONS (INCL BLOOD GROUPS)*** | | | | | | | | | | |
| *COAGULATION AND BLEEDING ANALYSES* | | | | | | | | | | |
| BLEEDING TIME | 0 | 0 | 0 | 0 | 0 | 0 | 0 | 0 | 0 | 0 |
| INTERNATIONAL NORMALISED RATIO DECREASED | 0 | 0 | 0 | 0 | 0 | 0 | 0 | 0 | 0 | 0 |
| *PLATELET ANALYSES* | | | | | | | | | | |
| PLATELET COUNT INCREASED | 0 | 0 | 0 | 0 | 0 | 0 | 0 | 0 | 0 | 0 |
| *RED BLOOD CELL ANALYSES* | | | | | | | | | | |
| HAEMOGLOBIN | 0 | 0 | 0 | 0 | 0 | 0 | 0 | 0 | 0 | 0 |

| | | | | | | | | | | |
|---|---|---|---|---|---|---|---|---|---|---|
| ***METABOLIC, NUTRITIONAL AND BLOOD GAS INVESTIGATIONS*** | | | | | | | | | | |
| *BLOOD GAS AND ACID BASE ANALYSES* | | | | | | | | | | |
| OXYGEN SATURATION DECREASED | 0 | 0 | 0 | 0 | 0 | 0 | 0 | 0 | 0 | 0 |
| *CARBOHYDRATE TOLERANCE ANALYSES (INCL DIABETES)* | | | | | | | | | | |
| BLOOD GLUCOSE | 0 | 0 | 0 | 0 | 0 | 0 | 0 | 0 | 0 | 0 |
| BLOOD GLUCOSE ABNORMAL | 0 | 0 | 0 | 0 | 0 | 0 | 0 | 0 | 0 | 0 |
| BLOOD GLUCOSE DECREASED | 0 | 0 | 0 | 0 | 0 | 0 | 0 | 0 | 0 | 0 |
| BLOOD GLUCOSE INCREASED | 0 | 0 | 0 | 0 | 0 | 0 | 0 | 0 | 0 | 0 |
| ***MICROBIOLOGY AND SEROLOGY INVESTIGATIONS*** | | | | | | | | | | |
| *VIRUS IDENTIFICATION AND SEROLOGY* | | | | | | | | | | |
| SARS-COV-2 TEST | 0 | 0 | 0 | 0 | 0 | 0 | 0 | 0 | 0 | 0 |
| SARS-COV-2 TEST POSITIVE | 0 | 0 | 0 | 0 | 0 | 0 | 0 | 0 | 0 | 0 |
| ***MUSCULOSKELETAL AND SOFT TISSUE INVESTIGATIONS (EXCL ENZYME TESTS)*** | | | | | | | | | | |
| *MUSCULOSKELETAL AND SOFT TISSUE IMAGING PROCEDURES* | | | | | | | | | | |
| BONE SCAN | 0 | 0 | 0 | 0 | 0 | 0 | 0 | 0 | 0 | 0 |
| ***NEUROLOGICAL, SPECIAL SENSES AND PSYCHIATRIC INVESTIGATIONS*** | | | | | | | | | | |
| *CENTRAL NERVOUS SYSTEM IMAGING PROCEDURES* | | | | | | | | | | |
| MAGNETIC RESONANCE IMAGING HEAD | 0 | 0 | 0 | 0 | 0 | 0 | 0 | 0 | 0 | 0 |
| SCAN BRAIN | 0 | 0 | 0 | 0 | 0 | 0 | 0 | 0 | 0 | 0 |
| ***PHYSICAL EXAMINATION AND ORGAN SYSTEM STATUS TOPICS*** | | | | | | | | | | |
| *PHYSICAL EXAMINATION PROCEDURES AND ORGAN SYSTEM STATUS* | | | | | | | | | | |
| BODY TEMPERATURE | 2 | 1 | 0 | 1 | 0 | 2 | 1 | 0 | 1 | 0 |
| BODY TEMPERATURE ABNORMAL | 1 | 0 | 0 | 1 | 0 | 1 | 0 | 0 | 1 | 0 |
| BODY TEMPERATURE DECREASED | 0 | 0 | 0 | 0 | 0 | 0 | 0 | 0 | 0 | 0 |
| BODY TEMPERATURE FLUCTUATION | 2 | 0 | 1 | 1 | 0 | 2 | 0 | 1 | 1 | 0 |
| BODY TEMPERATURE INCREASED | 7 | 2 | 0 | 5 | 0 | 7 | 2 | 0 | 5 | 0 |
| GRIP STRENGTH DECREASED | 0 | 0 | 0 | 0 | 0 | 0 | 0 | 0 | 0 | 0 |
| HEAD LAG | 1 | 1 | 0 | 0 | 0 | 1 | 1 | 0 | 0 | 0 |
| LEFT-HANDEDNESS | 0 | 0 | 0 | 0 | 0 | 0 | 0 | 0 | 0 | 0 |
| LYMPH NODE PALPABLE | 1 | 0 | 0 | 1 | 0 | 1 | 0 | 0 | 1 | 0 |
| RESPIRATORY RATE DECREASED | 0 | 0 | 0 | 0 | 0 | 0 | 0 | 0 | 0 | 0 |
| SKIN TEMPERATURE | 0 | 0 | 0 | 0 | 0 | 0 | 0 | 0 | 0 | 0 |
| WEIGHT DECREASED | 0 | 0 | 0 | 0 | 0 | 0 | 0 | 0 | 0 | 0 |
| WEIGHT INCREASED | 0 | 0 | 0 | 0 | 0 | 0 | 0 | 0 | 0 | 0 |
| ***RENAL AND URINARY TRACT INVESTIGATIONS AND URINALYSES*** | | | | | | | | | | |
| *URINALYSIS NEC* | | | | | | | | | | |

| | | | | | | | | | | |
|---|---|---|---|---|---|---|---|---|---|---|
| BLOOD URINE | 0 | 0 | 0 | 0 | 0 | 0 | 0 | 0 | 0 | 0 |
| NITRITE URINE PRESENT | 0 | 0 | 0 | 0 | 0 | 0 | 0 | 0 | 0 | 0 |
| PH URINE | 0 | 0 | 0 | 0 | 0 | 0 | 0 | 0 | 0 | 0 |
| *URINARY TRACT FUNCTION ANALYSES NEC* | | | | | | | | | | |
| URINE OUTPUT | 0 | 0 | 0 | 0 | 0 | 0 | 0 | 0 | 0 | 0 |
| URINE OUTPUT INCREASED | 0 | 0 | 0 | 0 | 0 | 0 | 0 | 0 | 0 | 0 |
| ***REPRODUCTIVE ORGAN AND BREAST INVESTIGATIONS (EXCL HORMONE ANALYSES)*** | | | | | | | | | | |
| *REPRODUCTIVE ORGAN AND BREAST IMAGING PROCEDURES* | | | | | | | | | | |
| BREAST SCAN | 0 | 0 | 0 | 0 | 0 | 0 | 0 | 0 | 0 | 0 |
| ***RESPIRATORY AND PULMONARY INVESTIGATIONS (EXCL BLOOD GASES)*** | | | | | | | | | | |
| *RESPIRATORY AND PULMONARY FUNCTION DIAGNOSTIC PROCEDURES* | | | | | | | | | | |
| FORCED EXPIRATORY VOLUME DECREASED | 0 | 0 | 0 | 0 | 0 | 0 | 0 | 0 | 0 | 0 |
| FORCED EXPIRATORY VOLUME INCREASED | 0 | 0 | 0 | 0 | 0 | 0 | 0 | 0 | 0 | 0 |
| ***WATER, ELECTROLYTE AND MINERAL INVESTIGATIONS*** | | | | | | | | | | |
| *WATER AND ELECTROLYTE ANALYSES NEC* | | | | | | | | | | |
| VOLUME BLOOD | 0 | 0 | 0 | 0 | 0 | 0 | 0 | 0 | 0 | 0 |
| **METABOLISM AND NUTRITION DISORDERS** | | | | | | | | | | |
| ***APPETITE AND GENERAL NUTRITIONAL DISORDERS*** | | | | | | | | | | |
| *APPETITE DISORDERS* | | | | | | | | | | |
| APPETITE DISORDER | 0 | 0 | 0 | 0 | 0 | 0 | 0 | 0 | 0 | 0 |
| DECREASED APPETITE | 8 | 2 | 1 | 4 | 1 | 8 | 2 | 1 | 4 | 1 |
| FOOD CRAVING | 0 | 0 | 0 | 0 | 0 | 0 | 0 | 0 | 0 | 0 |
| FOOD REFUSAL | 1 | 0 | 0 | 1 | 0 | 0 | 0 | 0 | 0 | 0 |
| INCREASED APPETITE | 0 | 0 | 0 | 0 | 0 | 0 | 0 | 0 | 0 | 0 |
| *GENERAL NUTRITIONAL DISORDERS NEC* | | | | | | | | | | |
| FOOD AVERSION | 0 | 0 | 0 | 0 | 0 | 0 | 0 | 0 | 0 | 0 |
| ***ELECTROLYTE AND FLUID BALANCE CONDITIONS*** | | | | | | | | | | |
| *TOTAL FLUID VOLUME DECREASED* | | | | | | | | | | |
| DEHYDRATION | 1 | 1 | 0 | 0 | 0 | 1 | 1 | 0 | 0 | 0 |
| *TOTAL FLUID VOLUME INCREASED* | | | | | | | | | | |
| FLUID RETENTION | 0 | 0 | 0 | 0 | 0 | 0 | 0 | 0 | 0 | 0 |
| ***FOOD INTOLERANCE SYNDROMES*** | | | | | | | | | | |
| *FOOD MALABSORPTION AND INTOLERANCE SYNDROMES (EXCL SUGAR INTOLERANCE)* | | | | | | | | | | |
| ALCOHOL INTOLERANCE | 0 | 0 | 0 | 0 | 0 | 0 | 0 | 0 | 0 | 0 |
| ***GLUCOSE METABOLISM DISORDERS (INCL DIABETES MELLITUS)*** | | | | | | | | | | |
| *DIABETES MELLITUS (INCL SUBTYPES)* | | | | | | | | | | |

| | | | | | | | | | | |
|---|---|---|---|---|---|---|---|---|---|---|
| DIABETES MELLITUS | 0 | 0 | 0 | 0 | 0 | 0 | 0 | 0 | 0 | 0 |
| DIABETES MELLITUS INADEQUATE CONTROL | 0 | 0 | 0 | 0 | 0 | 0 | 0 | 0 | 0 | 0 |
| *HYPERGLYCAEMIC CONDITIONS NEC* | | | | | | | | | | |
| HYPERGLYCAEMIA | 0 | 0 | 0 | 0 | 0 | 0 | 0 | 0 | 0 | 0 |
| *HYPOGLYCAEMIC CONDITIONS NEC* | | | | | | | | | | |
| HYPOGLYCAEMIA | 0 | 0 | 0 | 0 | 0 | 0 | 0 | 0 | 0 | 0 |
| ***PURINE AND PYRIMIDINE METABOLISM DISORDERS*** | | | | | | | | | | |
| *DISORDERS OF PURINE METABOLISM* | | | | | | | | | | |
| GOUT | 0 | 0 | 0 | 0 | 0 | 0 | 0 | 0 | 0 | 0 |
| **MUSCULOSKELETAL AND CONNECTIVE TISSUE DISORDERS** | | | | | | | | | | |
| ***BONE DISORDERS (EXCL CONGENITAL AND FRACTURES)*** | | | | | | | | | | |
| *BONE DISORDERS NEC* | | | | | | | | | | |
| JAW DISORDER | 0 | 0 | 0 | 0 | 0 | 0 | 0 | 0 | 0 | 0 |
| OSTEITIS | 0 | 0 | 0 | 0 | 0 | 0 | 0 | 0 | 0 | 0 |
| *BONE RELATED SIGNS AND SYMPTOMS* | | | | | | | | | | |
| BONE PAIN | 1 | 0 | 0 | 0 | 1 | 0 | 0 | 0 | 0 | 0 |
| BONE SWELLING | 0 | 0 | 0 | 0 | 0 | 0 | 0 | 0 | 0 | 0 |
| PAIN IN JAW | 1 | 0 | 1 | 0 | 0 | 1 | 0 | 1 | 0 | 0 |
| PUBIC PAIN | 0 | 0 | 0 | 0 | 0 | 0 | 0 | 0 | 0 | 0 |
| SPINAL PAIN | 0 | 0 | 0 | 0 | 0 | 0 | 0 | 0 | 0 | 0 |
| ***CONNECTIVE TISSUE DISORDERS (EXCL CONGENITAL)*** | | | | | | | | | | |
| *CONNECTIVE TISSUE DISORDERS NEC* | | | | | | | | | | |
| POLYMYALGIA RHEUMATICA | 1 | 0 | 0 | 0 | 1 | 1 | 0 | 0 | 0 | 1 |
| *LUPUS ERYTHEMATOSUS (INCL SUBTYPES)* | | | | | | | | | | |
| SYSTEMIC LUPUS ERYTHEMATOSUS | 0 | 0 | 0 | 0 | 0 | 0 | 0 | 0 | 0 | 0 |
| ***JOINT DISORDERS*** | | | | | | | | | | |
| *ARTHROPATHIES NEC* | | | | | | | | | | |
| ARTHRITIS | 0 | 0 | 0 | 0 | 0 | 0 | 0 | 0 | 0 | 0 |
| ARTHROPATHY | 0 | 0 | 0 | 0 | 0 | 0 | 0 | 0 | 0 | 0 |
| POLYARTHRITIS | 0 | 0 | 0 | 0 | 0 | 0 | 0 | 0 | 0 | 0 |
| RHEUMATIC FEVER | 0 | 0 | 0 | 0 | 0 | 0 | 0 | 0 | 0 | 0 |
| *JOINT RELATED DISORDERS NEC* | | | | | | | | | | |
| JOINT LOCK | 0 | 0 | 0 | 0 | 0 | 0 | 0 | 0 | 0 | 0 |
| PERIARTHRITIS | 1 | 0 | 0 | 1 | 0 | 1 | 0 | 0 | 1 | 0 |
| TEMPOROMANDIBULAR PAIN AND DYSFUNCTION SYNDROME | 0 | 0 | 0 | 0 | 0 | 0 | 0 | 0 | 0 | 0 |

| | | | | | | | | | | |
|---|---|---|---|---|---|---|---|---|---|---|
| *JOINT RELATED SIGNS AND SYMPTOMS* | | | | | | | | | | |
| ARTHRALGIA | 84 | 16 | 9 | 47 | 12 | 70 | 16 | 8 | 37 | 9 |
| JAW CLICKING | 0 | 0 | 0 | 0 | 0 | 0 | 0 | 0 | 0 | 0 |
| JOINT STIFFNESS | 3 | 1 | 0 | 1 | 1 | 3 | 1 | 0 | 1 | 1 |
| JOINT SWELLING | 0 | 0 | 0 | 0 | 0 | 0 | 0 | 0 | 0 | 0 |
| *OSTEOARTHROPATHIES* | | | | | | | | | | |
| OSTEOARTHRITIS | 0 | 0 | 0 | 0 | 0 | 0 | 0 | 0 | 0 | 0 |
| *PSORIATIC ARTHROPATHIES* | | | | | | | | | | |
| PSORIATIC ARTHROPATHY | 0 | 0 | 0 | 0 | 0 | 0 | 0 | 0 | 0 | 0 |
| *RHEUMATOID ARTHROPATHIES* | | | | | | | | | | |
| RHEUMATOID ARTHRITIS | 0 | 0 | 0 | 0 | 0 | 0 | 0 | 0 | 0 | 0 |
| ***MUSCLE DISORDERS*** | | | | | | | | | | |
| *MUSCLE INFECTIONS AND INFLAMMATIONS* | | | | | | | | | | |
| MYOSITIS | 0 | 0 | 0 | 0 | 0 | 0 | 0 | 0 | 0 | 0 |
| *MUSCLE PAINS* | | | | | | | | | | |
| FIBROMYALGIA | 0 | 0 | 0 | 0 | 0 | 0 | 0 | 0 | 0 | 0 |
| MYALGIA | 112 | 21 | 17 | 51 | 23 | 92 | 21 | 11 | 40 | 20 |
| *MUSCLE RELATED SIGNS AND SYMPTOMS NEC* | | | | | | | | | | |
| MUSCLE ATROPHY | 0 | 0 | 0 | 0 | 0 | 0 | 0 | 0 | 0 | 0 |
| MUSCLE DISCOMFORT | 0 | 0 | 0 | 0 | 0 | 0 | 0 | 0 | 0 | 0 |
| MUSCLE FATIGUE | 4 | 2 | 0 | 2 | 0 | 3 | 2 | 0 | 1 | 0 |
| MUSCLE FIBROSIS | 0 | 0 | 0 | 0 | 0 | 0 | 0 | 0 | 0 | 0 |
| MUSCLE MASS | 0 | 0 | 0 | 0 | 0 | 0 | 0 | 0 | 0 | 0 |
| MUSCLE SPASMS | 2 | 0 | 0 | 1 | 1 | 2 | 0 | 0 | 1 | 1 |
| MUSCLE TIGHTNESS | 0 | 0 | 0 | 0 | 0 | 0 | 0 | 0 | 0 | 0 |
| MUSCLE TWITCHING | 0 | 0 | 0 | 0 | 0 | 0 | 0 | 0 | 0 | 0 |
| *MUSCLE TONE ABNORMALITIES* | | | | | | | | | | |
| MUSCLE RIGIDITY | 0 | 0 | 0 | 0 | 0 | 0 | 0 | 0 | 0 | 0 |
| TRISMUS | 1 | 0 | 1 | 0 | 0 | 1 | 0 | 1 | 0 | 0 |
| *MUSCLE WEAKNESS CONDITIONS* | | | | | | | | | | |
| MUSCULAR WEAKNESS | 4 | 2 | 1 | 1 | 0 | 4 | 2 | 1 | 1 | 0 |
| ***MUSCULOSKELETAL AND CONNECTIVE TISSUE DISORDERS NEC*** | | | | | | | | | | |
| *MUSCULOSKELETAL AND CONNECTIVE TISSUE CONDITIONS NEC* | | | | | | | | | | |
| MOBILITY DECREASED | 1 | 0 | 0 | 0 | 1 | 1 | 0 | 0 | 0 | 1 |
| MUSCULOSKELETAL STIFFNESS | 17 | 3 | 3 | 5 | 6 | 15 | 3 | 3 | 4 | 5 |

| | | | | | | | | | | |
|---|---|---|---|---|---|---|---|---|---|---|
| BACK PAIN | 17 | 3 | 5 | 9 | 0 | 15 | 3 | 5 | 7 | 0 |
| FLANK PAIN | 0 | 0 | 0 | 0 | 0 | 0 | 0 | 0 | 0 | 0 |
| LIMB DISCOMFORT | 43 | 13 | 3 | 25 | 2 | 42 | 13 | 3 | 24 | 2 |
| MUSCULOSKELETAL CHEST PAIN | 0 | 0 | 0 | 0 | 0 | 0 | 0 | 0 | 0 | 0 |
| MUSCULOSKELETAL DISCOMFORT | 3 | 0 | 0 | 3 | 0 | 3 | 0 | 0 | 3 | 0 |
| MUSCULOSKELETAL PAIN | 0 | 0 | 0 | 0 | 0 | 0 | 0 | 0 | 0 | 0 |
| NECK PAIN | 7 | 2 | 1 | 2 | 2 | 7 | 2 | 1 | 2 | 2 |
| PAIN IN EXTREMITY | 399 | 141 | 26 | 179 | 53 | 354 | 141 | 17 | 155 | 41 |
| *SOFT TISSUE DISORDERS NEC* | | | | | | | | | | |
| AXILLARY MASS | 0 | 0 | 0 | 0 | 0 | 0 | 0 | 0 | 0 | 0 |
| GROIN PAIN | 0 | 0 | 0 | 0 | 0 | 0 | 0 | 0 | 0 | 0 |
| ***SYNOVIAL AND BURSAL DISORDERS*** | | | | | | | | | | |
| *SYNOVIAL DISORDERS* | | | | | | | | | | |
| SYNOVITIS | 0 | 0 | 0 | 0 | 0 | 0 | 0 | 0 | 0 | 0 |
| ***TENDON, LIGAMENT AND CARTILAGE DISORDERS*** | | | | | | | | | | |
| *CARTILAGE DISORDERS* | | | | | | | | | | |
| COSTOCHONDRITIS | 0 | 0 | 0 | 0 | 0 | 0 | 0 | 0 | 0 | 0 |
| *TENDON DISORDERS* | | | | | | | | | | |
| TENDONITIS | 0 | 0 | 0 | 0 | 0 | 0 | 0 | 0 | 0 | 0 |
| TENOSYNOVITIS | 0 | 0 | 0 | 0 | 0 | 0 | 0 | 0 | 0 | 0 |
| TRIGGER FINGER | 1 | 0 | 0 | 1 | 0 | 1 | 0 | 0 | 1 | 0 |
| **NEOPLASMS BENIGN, MALIGNANT AND UNSPECIFIED (INCL CYSTS AND POLYPS)** | | | | | | | | | | |
| ***CUTANEOUS NEOPLASMS BENIGN*** | | | | | | | | | | |
| *SKIN NEOPLASMS BENIGN* | | | | | | | | | | |
| MELANOCYTIC NAEVUS | 0 | 0 | 0 | 0 | 0 | 0 | 0 | 0 | 0 | 0 |
| SEBORRHOEIC KERATOSIS | 1 | 0 | 0 | 1 | 0 | 1 | 0 | 0 | 1 | 0 |
| SKIN PAPILLOMA | 0 | 0 | 0 | 0 | 0 | 0 | 0 | 0 | 0 | 0 |
| **NERVOUS SYSTEM DISORDERS** | | | | | | | | | | |
| ***CENTRAL NERVOUS SYSTEM INFECTIONS AND INFLAMMATIONS*** | | | | | | | | | | |
| *MYELITIS (INCL INFECTIVE)* | | | | | | | | | | |
| MYELITIS TRANSVERSE | 0 | 0 | 0 | 0 | 0 | 0 | 0 | 0 | 0 | 0 |
| ***CENTRAL NERVOUS SYSTEM VASCULAR DISORDERS*** | | | | | | | | | | |
| *CENTRAL NERVOUS SYSTEM HAEMORRHAGES AND CEREBROVASCULAR ACCIDENTS* | | | | | | | | | | |
| CEREBRAL HAEMORRHAGE | 0 | 0 | 0 | 0 | 0 | 0 | 0 | 0 | 0 | 0 |
| CEREBROVASCULAR ACCIDENT | 0 | 0 | 0 | 0 | 0 | 0 | 0 | 0 | 0 | 0 |
| *TRANSIENT CEREBROVASCULAR EVENTS* | | | | | | | | | | |

| | | | | | | | | | | |
|---|---|---|---|---|---|---|---|---|---|---|
| TRANSIENT ISCHAEMIC ATTACK | 0 | 0 | 0 | 0 | 0 | 0 | 0 | 0 | 0 | 0 |
| ***CRANIAL NERVE DISORDERS (EXCL NEOPLASMS)*** | | | | | | | | | | |
| *FACIAL CRANIAL NERVE DISORDERS* | | | | | | | | | | |
| BELL'S PALSY | 0 | 0 | 0 | 0 | 0 | 0 | 0 | 0 | 0 | 0 |
| FACIAL PARALYSIS | 0 | 0 | 0 | 0 | 0 | 0 | 0 | 0 | 0 | 0 |
| FACIAL PARESIS | 1 | 0 | 0 | 1 | 0 | 1 | 0 | 0 | 1 | 0 |
| *OLFACTORY NERVE DISORDERS* | | | | | | | | | | |
| ANOSMIA | 1 | 0 | 0 | 1 | 0 | 1 | 0 | 0 | 1 | 0 |
| HYPOSMIA | 0 | 0 | 0 | 0 | 0 | 0 | 0 | 0 | 0 | 0 |
| PAROSMIA | 0 | 0 | 0 | 0 | 0 | 0 | 0 | 0 | 0 | 0 |
| *TRIGEMINAL DISORDERS* | | | | | | | | | | |
| TRIGEMINAL NEURALGIA | 0 | 0 | 0 | 0 | 0 | 0 | 0 | 0 | 0 | 0 |
| TRIGEMINAL NEURITIS | 0 | 0 | 0 | 0 | 0 | 0 | 0 | 0 | 0 | 0 |
| ***DEMYELINATING DISORDERS*** | | | | | | | | | | |
| *MULTIPLE SCLEROSIS ACUTE AND PROGRESSIVE* | | | | | | | | | | |
| MULTIPLE SCLEROSIS RELAPSE | 0 | 0 | 0 | 0 | 0 | 0 | 0 | 0 | 0 | 0 |
| ***HEADACHES*** | | | | | | | | | | |
| *HEADACHES NEC* | | | | | | | | | | |
| CLUSTER HEADACHE | 1 | 0 | 0 | 0 | 1 | 1 | 0 | 0 | 0 | 1 |
| COLD-STIMULUS HEADACHE | 0 | 0 | 0 | 0 | 0 | 0 | 0 | 0 | 0 | 0 |
| DRUG WITHDRAWAL HEADACHE | 0 | 0 | 0 | 0 | 0 | 0 | 0 | 0 | 0 | 0 |
| HEADACHE | 263 | 57 | 41 | 130 | 35 | 221 | 57 | 28 | 106 | 30 |
| PRIMARY STABBING HEADACHE | 0 | 0 | 0 | 0 | 0 | 0 | 0 | 0 | 0 | 0 |
| SINUS HEADACHE | 5 | 1 | 1 | 1 | 2 | 3 | 1 | 1 | 1 | 0 |
| TENSION HEADACHE | 10 | 3 | 2 | 1 | 4 | 10 | 3 | 2 | 1 | 4 |
| THUNDERCLAP HEADACHE | 0 | 0 | 0 | 0 | 0 | 0 | 0 | 0 | 0 | 0 |
| VASCULAR HEADACHE | 0 | 0 | 0 | 0 | 0 | 0 | 0 | 0 | 0 | 0 |
| *MIGRAINE HEADACHES* | | | | | | | | | | |
| MIGRAINE | 6 | 3 | 0 | 3 | 0 | 6 | 3 | 0 | 3 | 0 |
| MIGRAINE WITH AURA | 1 | 0 | 1 | 0 | 0 | 1 | 0 | 1 | 0 | 0 |
| RETINAL MIGRAINE | 0 | 0 | 0 | 0 | 0 | 0 | 0 | 0 | 0 | 0 |
| TYPICAL AURA WITHOUT HEADACHE | 0 | 0 | 0 | 0 | 0 | 0 | 0 | 0 | 0 | 0 |
| ***MENTAL IMPAIRMENT DISORDERS*** | | | | | | | | | | |
| *INTELLECTUAL DISABILITIES* | | | | | | | | | | |
| INTELLECTUAL DISABILITY | 1 | 0 | 0 | 1 | 0 | 1 | 0 | 0 | 1 | 0 |

| | | | | | | | | | | |
|---|---|---|---|---|---|---|---|---|---|---|
| *MEMORY LOSS (EXCL DEMENTIA)* | | | | | | | | | | |
| AMNESIA | 0 | 0 | 0 | 0 | 0 | 0 | 0 | 0 | 0 | 0 |
| MEMORY IMPAIRMENT | 1 | 0 | 0 | 1 | 0 | 1 | 0 | 0 | 1 | 0 |
| *MENTAL IMPAIRMENT (EXCL DEMENTIA AND MEMORY LOSS)* | | | | | | | | | | |
| COGNITIVE DISORDER | 0 | 0 | 0 | 0 | 0 | 0 | 0 | 0 | 0 | 0 |
| DISTURBANCE IN ATTENTION | 0 | 0 | 0 | 0 | 0 | 0 | 0 | 0 | 0 | 0 |
| MENTAL IMPAIRMENT | 0 | 0 | 0 | 0 | 0 | 0 | 0 | 0 | 0 | 0 |
| ***MOVEMENT DISORDERS (INCL PARKINSONISM)*** | | | | | | | | | | |
| *DYSKINESIAS AND MOVEMENT DISORDERS NEC* | | | | | | | | | | |
| BRADYKINESIA | 2 | 1 | 0 | 1 | 0 | 2 | 1 | 0 | 1 | 0 |
| CLUMSINESS | 0 | 0 | 0 | 0 | 0 | 0 | 0 | 0 | 0 | 0 |
| DYSKINESIA | 0 | 0 | 0 | 0 | 0 | 0 | 0 | 0 | 0 | 0 |
| EXTRAPYRAMIDAL DISORDER | 0 | 0 | 0 | 0 | 0 | 0 | 0 | 0 | 0 | 0 |
| PSYCHOMOTOR HYPERACTIVITY | 0 | 0 | 0 | 0 | 0 | 0 | 0 | 0 | 0 | 0 |
| *PARALYSIS AND PARESIS (EXCL CRANIAL NERVE)* | | | | | | | | | | |
| HEMIPLEGIA | 1 | 0 | 0 | 1 | 0 | 1 | 0 | 0 | 1 | 0 |
| MONOPARESIS | 1 | 0 | 0 | 1 | 0 | 0 | 0 | 0 | 0 | 0 |
| MONOPLEGIA | 0 | 0 | 0 | 0 | 0 | 0 | 0 | 0 | 0 | 0 |
| PARALYSIS | 1 | 0 | 0 | 1 | 0 | 1 | 0 | 0 | 1 | 0 |
| *PARKINSON'S DISEASE AND PARKINSONISM* | | | | | | | | | | |
| FREEZING PHENOMENON | 1 | 0 | 0 | 1 | 0 | 1 | 0 | 0 | 1 | 0 |
| PARKINSON'S DISEASE | 0 | 0 | 0 | 0 | 0 | 0 | 0 | 0 | 0 | 0 |
| *TREMOR (EXCL CONGENITAL)* | | | | | | | | | | |
| TREMOR | 7 | 1 | 3 | 2 | 1 | 6 | 1 | 3 | 1 | 1 |
| ***NEUROLOGICAL DISORDERS NEC*** | | | | | | | | | | |
| *COMA STATES* | | | | | | | | | | |
| DIABETIC HYPERGLYCAEMIC COMA | 0 | 0 | 0 | 0 | 0 | 0 | 0 | 0 | 0 | 0 |
| *COORDINATION AND BALANCE DISTURBANCES* | | | | | | | | | | |
| BALANCE DISORDER | 1 | 0 | 0 | 0 | 1 | 1 | 0 | 0 | 0 | 1 |
| COORDINATION ABNORMAL | 0 | 0 | 0 | 0 | 0 | 0 | 0 | 0 | 0 | 0 |
| DYSSTASIA | 0 | 0 | 0 | 0 | 0 | 0 | 0 | 0 | 0 | 0 |
| VESTIBULAR NYSTAGMUS | 0 | 0 | 0 | 0 | 0 | 0 | 0 | 0 | 0 | 0 |
| *DISTURBANCES IN CONSCIOUSNESS NEC* | | | | | | | | | | |
| DEPRESSED LEVEL OF CONSCIOUSNESS | 0 | 0 | 0 | 0 | 0 | 0 | 0 | 0 | 0 | 0 |
| LETHARGY | 23 | 4 | 2 | 11 | 6 | 19 | 4 | 2 | 11 | 2 |

| | | | | | | | | | | |
|---|---|---|---|---|---|---|---|---|---|---|
| LOSS OF CONSCIOUSNESS | 1 | 0 | 0 | 1 | 0 | 1 | 0 | 0 | 1 | 0 |
| SEDATION | 1 | 0 | 0 | 0 | 1 | 1 | 0 | 0 | 0 | 1 |
| SOMNOLENCE | 14 | 4 | 5 | 5 | 0 | 12 | 4 | 5 | 3 | 0 |
| SYNCOPE | 5 | 2 | 0 | 3 | 0 | 4 | 2 | 0 | 2 | 0 |
| *NERVOUS SYSTEM DISORDERS NEC* | | | | | | | | | | |
| NERVOUS SYSTEM DISORDER | 0 | 0 | 0 | 0 | 0 | 0 | 0 | 0 | 0 | 0 |
| *NEUROLOGICAL SIGNS AND SYMPTOMS NEC* | | | | | | | | | | |
| AGITATION NEONATAL | 0 | 0 | 0 | 0 | 0 | 0 | 0 | 0 | 0 | 0 |
| BRAIN FOG | 0 | 0 | 0 | 0 | 0 | 0 | 0 | 0 | 0 | 0 |
| DIZZINESS | 40 | 11 | 5 | 16 | 8 | 31 | 11 | 2 | 12 | 6 |
| DIZZINESS EXERTIONAL | 1 | 1 | 0 | 0 | 0 | 1 | 1 | 0 | 0 | 0 |
| DIZZINESS POSTURAL | 3 | 1 | 0 | 1 | 1 | 3 | 1 | 0 | 1 | 1 |
| HEAD DISCOMFORT | 2 | 0 | 2 | 0 | 0 | 2 | 0 | 2 | 0 | 0 |
| INFANT IRRITABILITY | 0 | 0 | 0 | 0 | 0 | 0 | 0 | 0 | 0 | 0 |
| MENINGISM | 0 | 0 | 0 | 0 | 0 | 0 | 0 | 0 | 0 | 0 |
| MYOCLONUS | 0 | 0 | 0 | 0 | 0 | 0 | 0 | 0 | 0 | 0 |
| NEUROLOGICAL SYMPTOM | 0 | 0 | 0 | 0 | 0 | 0 | 0 | 0 | 0 | 0 |
| PERSISTENT POSTURAL-PERCEPTUAL DIZZINESS | 0 | 0 | 0 | 0 | 0 | 0 | 0 | 0 | 0 | 0 |
| PRESYNCOPE | 2 | 0 | 0 | 2 | 0 | 2 | 0 | 0 | 2 | 0 |
| *PARAESTHESIAS AND DYSAESTHESIAS* | | | | | | | | | | |
| BURNING FEET SYNDROME | 0 | 0 | 0 | 0 | 0 | 0 | 0 | 0 | 0 | 0 |
| BURNING SENSATION | 0 | 0 | 0 | 0 | 0 | 0 | 0 | 0 | 0 | 0 |
| FORMICATION | 0 | 0 | 0 | 0 | 0 | 0 | 0 | 0 | 0 | 0 |
| HYPERAESTHESIA | 0 | 0 | 0 | 0 | 0 | 0 | 0 | 0 | 0 | 0 |
| HYPOAESTHESIA | 2 | 2 | 0 | 0 | 0 | 2 | 2 | 0 | 0 | 0 |
| PARAESTHESIA | 4 | 1 | 1 | 1 | 1 | 4 | 1 | 1 | 1 | 1 |
| REVERSED HOT-COLD SENSATION | 0 | 0 | 0 | 0 | 0 | 0 | 0 | 0 | 0 | 0 |
| *SENSORY ABNORMALITIES NEC* | | | | | | | | | | |
| AGEUSIA | 1 | 0 | 0 | 1 | 0 | 1 | 0 | 0 | 1 | 0 |
| ALLODYNIA | 1 | 0 | 1 | 0 | 0 | 0 | 0 | 0 | 0 | 0 |
| DYSGEUSIA | 3 | 0 | 0 | 2 | 1 | 3 | 0 | 0 | 2 | 1 |
| HYPOGEUSIA | 0 | 0 | 0 | 0 | 0 | 0 | 0 | 0 | 0 | 0 |
| NEURALGIA | 0 | 0 | 0 | 0 | 0 | 0 | 0 | 0 | 0 | 0 |
| POST HERPETIC NEURALGIA | 0 | 0 | 0 | 0 | 0 | 0 | 0 | 0 | 0 | 0 |
| RESTLESS ARM SYNDROME | 0 | 0 | 0 | 0 | 0 | 0 | 0 | 0 | 0 | 0 |

| | | | | | | | | | | |
|---|---|---|---|---|---|---|---|---|---|---|
| RESTLESS LEGS SYNDROME | 0 | 0 | 0 | 0 | 0 | 0 | 0 | 0 | 0 | 0 |
| SENSORY LOSS | 0 | 0 | 0 | 0 | 0 | 0 | 0 | 0 | 0 | 0 |
| TASTE DISORDER | 1 | 1 | 0 | 0 | 0 | 1 | 1 | 0 | 0 | 0 |
| *SPEECH AND LANGUAGE ABNORMALITIES* | | | | | | | | | | |
| DYSARTHRIA | 1 | 1 | 0 | 0 | 0 | 1 | 1 | 0 | 0 | 0 |
| SPEECH DISORDER DEVELOPMENTAL | 1 | 0 | 0 | 1 | 0 | 1 | 0 | 0 | 1 | 0 |
| ***NEUROLOGICAL DISORDERS OF THE EYE*** | | | | | | | | | | |
| *NEUROLOGIC VISUAL PROBLEMS NEC* | | | | | | | | | | |
| TUNNEL VISION | 0 | 0 | 0 | 0 | 0 | 0 | 0 | 0 | 0 | 0 |
| ***NEUROMUSCULAR DISORDERS*** | | | | | | | | | | |
| *MUSCLE TONE ABNORMAL* | | | | | | | | | | |
| HYPOTONIA | 0 | 0 | 0 | 0 | 0 | 0 | 0 | 0 | 0 | 0 |
| STIFF LEG SYNDROME | 0 | 0 | 0 | 0 | 0 | 0 | 0 | 0 | 0 | 0 |
| *NEUROMUSCULAR DISORDERS NEC* | | | | | | | | | | |
| MUSCLE SPASTICITY | 0 | 0 | 0 | 0 | 0 | 0 | 0 | 0 | 0 | 0 |
| ***PERIPHERAL NEUROPATHIES*** | | | | | | | | | | |
| *ACUTE POLYNEUROPATHIES* | | | | | | | | | | |
| GUILLAIN-BARRE SYNDROME | 0 | 0 | 0 | 0 | 0 | 0 | 0 | 0 | 0 | 0 |
| *PERIPHERAL NEUROPATHIES NEC* | | | | | | | | | | |
| AXONAL NEUROPATHY | 0 | 0 | 0 | 0 | 0 | 0 | 0 | 0 | 0 | 0 |
| NEUROPATHY PERIPHERAL | 1 | 0 | 0 | 1 | 0 | 1 | 0 | 0 | 1 | 0 |
| ***SEIZURES (INCL SUBTYPES)*** | | | | | | | | | | |
| *SEIZURES AND SEIZURE DISORDERS NEC* | | | | | | | | | | |
| EPILEPSY | 1 | 0 | 0 | 1 | 0 | 1 | 0 | 0 | 1 | 0 |
| SEIZURE | 1 | 0 | 0 | 1 | 0 | 1 | 0 | 0 | 1 | 0 |
| ***SLEEP DISTURBANCES (INCL SUBTYPES)*** | | | | | | | | | | |
| *DISTURBANCES IN SLEEP PHASE RHYTHM* | | | | | | | | | | |
| CIRCADIAN RHYTHM SLEEP DISORDER | 0 | 0 | 0 | 0 | 0 | 0 | 0 | 0 | 0 | 0 |
| *SLEEP DISTURBANCES NEC* | | | | | | | | | | |
| SLEEP DEFICIT | 0 | 0 | 0 | 0 | 0 | 0 | 0 | 0 | 0 | 0 |
| ***SPINAL CORD AND NERVE ROOT DISORDERS*** | | | | | | | | | | |
| *CERVICAL SPINAL CORD AND NERVE ROOT DISORDERS* | | | | | | | | | | |
| CERVICOBRACHIAL SYNDROME | 1 | 0 | 0 | 1 | 0 | 1 | 0 | 0 | 1 | 0 |
| *LUMBAR SPINAL CORD AND NERVE ROOT DISORDERS* | | | | | | | | | | |
| CAUDA EQUINA SYNDROME | 1 | 0 | 0 | 1 | 0 | 1 | 0 | 0 | 1 | 0 |
| SCIATICA | 0 | 0 | 0 | 0 | 0 | 0 | 0 | 0 | 0 | 0 |

| | | | | | | | | | | |
|---|---|---|---|---|---|---|---|---|---|---|
| **PREGNANCY, PUERPERIUM AND PERINATAL CONDITIONS** | | | | | | | | | | |
| ***ABORTIONS AND STILLBIRTH*** | | | | | | | | | | |
| *ABORTIONS SPONTANEOUS* | | | | | | | | | | |
| ABORTION SPONTANEOUS | 1 | 1 | 0 | 0 | 0 | 1 | 1 | 0 | 0 | 0 |
| *STILLBIRTH AND FOETAL DEATH* | | | | | | | | | | |
| FOETAL DEATH | 0 | 0 | 0 | 0 | 0 | 0 | 0 | 0 | 0 | 0 |
| ***FOETAL COMPLICATIONS*** | | | | | | | | | | |
| *FOETAL COMPLICATIONS NEC* | | | | | | | | | | |
| FOETAL DISORDER | 0 | 0 | 0 | 0 | 0 | 0 | 0 | 0 | 0 | 0 |
| FOETAL HYPOKINESIA | 0 | 0 | 0 | 0 | 0 | 0 | 0 | 0 | 0 | 0 |
| *FOETAL GROWTH COMPLICATIONS* | | | | | | | | | | |
| FOETAL MACROSOMIA | 0 | 0 | 0 | 0 | 0 | 0 | 0 | 0 | 0 | 0 |
| ***MATERNAL COMPLICATIONS OF PREGNANCY*** | | | | | | | | | | |
| *MATERNAL COMPLICATIONS OF PREGNANCY NEC* | | | | | | | | | | |
| MORNING SICKNESS | 0 | 0 | 0 | 0 | 0 | 0 | 0 | 0 | 0 | 0 |
| ***PLACENTAL, AMNIOTIC AND CAVITY DISORDERS (EXCL HAEMORRHAGES)*** | | | | | | | | | | |
| *PLACENTAL ABNORMALITIES (EXCL NEOPLASMS)* | | | | | | | | | | |
| PLACENTAL INFARCTION | 0 | 0 | 0 | 0 | 0 | 0 | 0 | 0 | 0 | 0 |
| ***PREGNANCY, LABOUR, DELIVERY AND POSTPARTUM CONDITIONS*** | | | | | | | | | | |
| *NORMAL PREGNANCY, LABOUR AND DELIVERY* | | | | | | | | | | |
| PREGNANCY | 0 | 0 | 0 | 0 | 0 | 0 | 0 | 0 | 0 | 0 |
| UTERINE CONTRACTIONS DURING PREGNANCY | 0 | 0 | 0 | 0 | 0 | 0 | 0 | 0 | 0 | 0 |
| **PRODUCT ISSUES** | | | | | | | | | | |
| ***DEVICE ISSUES*** | | | | | | | | | | |
| *DEVICE MALFUNCTION EVENTS NEC* | | | | | | | | | | |
| OVERSENSING | 0 | 0 | 0 | 0 | 0 | 0 | 0 | 0 | 0 | 0 |
| **PSYCHIATRIC DISORDERS** | | | | | | | | | | |
| ***ANXIETY DISORDERS AND SYMPTOMS*** | | | | | | | | | | |
| *ANXIETY SYMPTOMS* | | | | | | | | | | |
| AGITATION | 0 | 0 | 0 | 0 | 0 | 0 | 0 | 0 | 0 | 0 |
| ANXIETY | 2 | 0 | 0 | 2 | 0 | 2 | 0 | 0 | 2 | 0 |
| NERVOUSNESS | 0 | 0 | 0 | 0 | 0 | 0 | 0 | 0 | 0 | 0 |
| TENSION | 0 | 0 | 0 | 0 | 0 | 0 | 0 | 0 | 0 | 0 |
| *PANIC ATTACKS AND DISORDERS* | | | | | | | | | | |
| PANIC ATTACK | 0 | 0 | 0 | 0 | 0 | 0 | 0 | 0 | 0 | 0 |
| ***CHANGES IN PHYSICAL ACTIVITY*** | | | | | | | | | | |

| | | | | | | | | | | |
|---|---|---|---|---|---|---|---|---|---|---|
| *INCREASED PHYSICAL ACTIVITY LEVELS* | | | | | | | | | | |
| RESTLESSNESS | 1 | 0 | 0 | 0 | 1 | 1 | 0 | 0 | 0 | 1 |
| *STEREOTYPIES AND AUTOMATISMS* | | | | | | | | | | |
| BRUXISM | 0 | 0 | 0 | 0 | 0 | 0 | 0 | 0 | 0 | 0 |
| ***COGNITIVE AND ATTENTION DISORDERS AND DISTURBANCES*** | | | | | | | | | | |
| *COGNITIVE AND ATTENTION DISORDERS AND DISTURBANCES NEC* | | | | | | | | | | |
| DAYDREAMING | 0 | 0 | 0 | 0 | 0 | 0 | 0 | 0 | 0 | 0 |
| MENTAL FATIGUE | 1 | 1 | 0 | 0 | 0 | 1 | 1 | 0 | 0 | 0 |
| ***COMMUNICATION DISORDERS AND DISTURBANCES*** | | | | | | | | | | |
| *SPEECH ARTICULATION AND RHYTHM DISTURBANCES* | | | | | | | | | | |
| DYSPHEMIA | 0 | 0 | 0 | 0 | 0 | 0 | 0 | 0 | 0 | 0 |
| ***DELIRIA (INCL CONFUSION)*** | | | | | | | | | | |
| *CONFUSION AND DISORIENTATION* | | | | | | | | | | |
| CONFUSIONAL STATE | 3 | 1 | 1 | 1 | 0 | 2 | 1 | 1 | 0 | 0 |
| DISORIENTATION | 0 | 0 | 0 | 0 | 0 | 0 | 0 | 0 | 0 | 0 |
| *DELIRIA* | | | | | | | | | | |
| DELIRIUM | 0 | 0 | 0 | 0 | 0 | 0 | 0 | 0 | 0 | 0 |
| ***DEPRESSED MOOD DISORDERS AND DISTURBANCES*** | | | | | | | | | | |
| *DEPRESSIVE DISORDERS* | | | | | | | | | | |
| DEPRESSION | 0 | 0 | 0 | 0 | 0 | 0 | 0 | 0 | 0 | 0 |
| *MOOD ALTERATIONS WITH DEPRESSIVE SYMPTOMS* | | | | | | | | | | |
| DEPRESSED MOOD | 0 | 0 | 0 | 0 | 0 | 0 | 0 | 0 | 0 | 0 |
| TEARFULNESS | 0 | 0 | 0 | 0 | 0 | 0 | 0 | 0 | 0 | 0 |
| ***DISSOCIATIVE DISORDERS*** | | | | | | | | | | |
| *DISSOCIATIVE STATES* | | | | | | | | | | |
| DISSOCIATION | 1 | 0 | 0 | 1 | 0 | 1 | 0 | 0 | 1 | 0 |
| ***DISTURBANCES IN THINKING AND PERCEPTION*** | | | | | | | | | | |
| *DELUSIONAL SYMPTOMS* | | | | | | | | | | |
| DELUSION | 0 | 0 | 0 | 0 | 0 | 0 | 0 | 0 | 0 | 0 |
| *HALLUCINATIONS (EXCL SLEEP-RELATED)* | | | | | | | | | | |
| HALLUCINATION | 0 | 0 | 0 | 0 | 0 | 0 | 0 | 0 | 0 | 0 |
| *PERCEPTION DISTURBANCES NEC* | | | | | | | | | | |
| DEREALISATION | 0 | 0 | 0 | 0 | 0 | 0 | 0 | 0 | 0 | 0 |
| *THINKING DISTURBANCES* | | | | | | | | | | |
| BRADYPHRENIA | 2 | 1 | 0 | 1 | 0 | 2 | 1 | 0 | 1 | 0 |
| THOUGHT BLOCKING | 0 | 0 | 0 | 0 | 0 | 0 | 0 | 0 | 0 | 0 |

| | | | | | | | | | | |
|---|---|---|---|---|---|---|---|---|---|---|
| ***MOOD DISORDERS AND DISTURBANCES NEC*** | | | | | | | | | | |
| *AFFECT ALTERATIONS NEC* | | | | | | | | | | |
| AFFECT LABILITY | 1 | 0 | 1 | 0 | 0 | 1 | 0 | 1 | 0 | 0 |
| INAPPROPRIATE AFFECT | 0 | 0 | 0 | 0 | 0 | 0 | 0 | 0 | 0 | 0 |
| *EMOTIONAL AND MOOD DISTURBANCES NEC* | | | | | | | | | | |
| ANGER | 0 | 0 | 0 | 0 | 0 | 0 | 0 | 0 | 0 | 0 |
| EMOTIONAL DISORDER | 0 | 0 | 0 | 0 | 0 | 0 | 0 | 0 | 0 | 0 |
| EMOTIONAL DISTRESS | 0 | 0 | 0 | 0 | 0 | 0 | 0 | 0 | 0 | 0 |
| EUPHORIC MOOD | 0 | 0 | 0 | 0 | 0 | 0 | 0 | 0 | 0 | 0 |
| IRRITABILITY | 0 | 0 | 0 | 0 | 0 | 0 | 0 | 0 | 0 | 0 |
| MOOD ALTERED | 1 | 0 | 0 | 1 | 0 | 1 | 0 | 0 | 1 | 0 |
| *FLUCTUATING MOOD SYMPTOMS* | | | | | | | | | | |
| MOOD SWINGS | 0 | 0 | 0 | 0 | 0 | 0 | 0 | 0 | 0 | 0 |
| *MOOD DISORDERS NEC* | | | | | | | | | | |
| APATHY | 0 | 0 | 0 | 0 | 0 | 0 | 0 | 0 | 0 | 0 |
| LISTLESS | 0 | 0 | 0 | 0 | 0 | 0 | 0 | 0 | 0 | 0 |
| ***PERSONALITY DISORDERS AND DISTURBANCES IN BEHAVIOUR*** | | | | | | | | | | |
| *BEHAVIOUR AND SOCIALISATION DISTURBANCES* | | | | | | | | | | |
| AGGRESSION | 0 | 0 | 0 | 0 | 0 | 0 | 0 | 0 | 0 | 0 |
| ***PSYCHIATRIC AND BEHAVIOURAL SYMPTOMS NEC*** | | | | | | | | | | |
| *PSYCHIATRIC SYMPTOMS NEC* | | | | | | | | | | |
| PSYCHIATRIC SYMPTOM | 0 | 0 | 0 | 0 | 0 | 0 | 0 | 0 | 0 | 0 |
| ***PSYCHIATRIC DISORDERS NEC*** | | | | | | | | | | |
| *MENTAL DISORDERS NEC* | | | | | | | | | | |
| MENTAL DISORDER | 0 | 0 | 0 | 0 | 0 | 0 | 0 | 0 | 0 | 0 |
| ***SCHIZOPHRENIA AND OTHER PSYCHOTIC DISORDERS*** | | | | | | | | | | |
| *PSYCHOTIC DISORDER NEC* | | | | | | | | | | |
| PSYCHOTIC DISORDER | 0 | 0 | 0 | 0 | 0 | 0 | 0 | 0 | 0 | 0 |
| ***SEXUAL DYSFUNCTIONS, DISTURBANCES AND GENDER IDENTITY DISORDERS*** | | | | | | | | | | |
| *SEXUAL DESIRE DISORDERS* | | | | | | | | | | |
| LOSS OF LIBIDO | 0 | 0 | 0 | 0 | 0 | 0 | 0 | 0 | 0 | 0 |
| ***SLEEP DISORDERS AND DISTURBANCES*** | | | | | | | | | | |
| *DISTURBANCES IN INITIATING AND MAINTAINING SLEEP* | | | | | | | | | | |
| INITIAL INSOMNIA | 0 | 0 | 0 | 0 | 0 | 0 | 0 | 0 | 0 | 0 |
| INSOMNIA | 5 | 1 | 1 | 3 | 0 | 4 | 1 | 1 | 2 | 0 |
| *DYSSOMNIAS* | | | | | | | | | | |

| | | | | | | | | | | |
|---|---|---|---|---|---|---|---|---|---|---|
| BREATHING-RELATED SLEEP DISORDER | 0 | 0 | 0 | 0 | 0 | 0 | 0 | 0 | 0 | 0 |
| POOR QUALITY SLEEP | 0 | 0 | 0 | 0 | 0 | 0 | 0 | 0 | 0 | 0 |
| *PARASOMNIAS* | | | | | | | | | | |
| ABNORMAL DREAMS | 1 | 0 | 0 | 1 | 0 | 1 | 0 | 0 | 1 | 0 |
| EXPLODING HEAD SYNDROME | 0 | 0 | 0 | 0 | 0 | 0 | 0 | 0 | 0 | 0 |
| NIGHTMARE | 1 | 1 | 0 | 0 | 0 | 1 | 1 | 0 | 0 | 0 |
| *SLEEP DISORDERS NEC* | | | | | | | | | | |
| SLEEP DISORDER | 0 | 0 | 0 | 0 | 0 | 0 | 0 | 0 | 0 | 0 |
| ***SOMATIC SYMPTOM AND RELATED DISORDERS*** | | | | | | | | | | |
| *SOMATIC SYMPTOM DISORDERS* | | | | | | | | | | |
| HABIT COUGH | 0 | 0 | 0 | 0 | 0 | 0 | 0 | 0 | 0 | 0 |
| ***SUICIDAL AND SELF-INJURIOUS BEHAVIOURS NEC*** | | | | | | | | | | |
| *SUICIDAL AND SELF-INJURIOUS BEHAVIOUR* | | | | | | | | | | |
| SUICIDAL IDEATION | 0 | 0 | 0 | 0 | 0 | 0 | 0 | 0 | 0 | 0 |
| **RENAL AND URINARY DISORDERS** | | | | | | | | | | |
| ***RENAL DISORDERS (EXCL NEPHROPATHIES)*** | | | | | | | | | | |
| *RENAL FAILURE AND IMPAIRMENT* | | | | | | | | | | |
| RENAL FAILURE | 0 | 0 | 0 | 0 | 0 | 0 | 0 | 0 | 0 | 0 |
| ***URINARY TRACT SIGNS AND SYMPTOMS*** | | | | | | | | | | |
| *BLADDER AND URETHRAL SYMPTOMS* | | | | | | | | | | |
| BLADDER PAIN | 0 | 0 | 0 | 0 | 0 | 0 | 0 | 0 | 0 | 0 |
| MICTURITION URGENCY | 1 | 0 | 0 | 1 | 0 | 1 | 0 | 0 | 1 | 0 |
| POLLAKIURIA | 0 | 0 | 0 | 0 | 0 | 0 | 0 | 0 | 0 | 0 |
| URINARY INCONTINENCE | 0 | 0 | 0 | 0 | 0 | 0 | 0 | 0 | 0 | 0 |
| *URINARY ABNORMALITIES* | | | | | | | | | | |
| HAEMATURIA | 0 | 0 | 0 | 0 | 0 | 0 | 0 | 0 | 0 | 0 |
| URINE ABNORMALITY | 1 | 0 | 0 | 1 | 0 | 1 | 0 | 0 | 1 | 0 |
| URINE ODOUR ABNORMAL | 0 | 0 | 0 | 0 | 0 | 0 | 0 | 0 | 0 | 0 |
| *URINARY TRACT SIGNS AND SYMPTOMS NEC* | | | | | | | | | | |
| POLYURIA | 1 | 0 | 0 | 0 | 1 | 1 | 0 | 0 | 0 | 1 |
| RENAL PAIN | 0 | 0 | 0 | 0 | 0 | 0 | 0 | 0 | 0 | 0 |
| **REPRODUCTIVE SYSTEM AND BREAST DISORDERS** | | | | | | | | | | |
| ***BREAST DISORDERS*** | | | | | | | | | | |
| *BREAST DISORDERS NEC* | | | | | | | | | | |
| BREAST MASS | 0 | 0 | 0 | 0 | 0 | 0 | 0 | 0 | 0 | 0 |
| NIPPLE ENLARGEMENT | 0 | 0 | 0 | 0 | 0 | 0 | 0 | 0 | 0 | 0 |

| | | | | | | | | | | |
|---|---|---|---|---|---|---|---|---|---|---|
| *BREAST SIGNS AND SYMPTOMS* | | | | | | | | | | |
| BREAST DISCHARGE | 0 | 0 | 0 | 0 | 0 | 0 | 0 | 0 | 0 | 0 |
| BREAST PAIN | 2 | 0 | 1 | 1 | 0 | 2 | 0 | 1 | 1 | 0 |
| BREAST SWELLING | 0 | 0 | 0 | 0 | 0 | 0 | 0 | 0 | 0 | 0 |
| BREAST TENDERNESS | 0 | 0 | 0 | 0 | 0 | 0 | 0 | 0 | 0 | 0 |
| NIPPLE PAIN | 0 | 0 | 0 | 0 | 0 | 0 | 0 | 0 | 0 | 0 |
| ***MENOPAUSE RELATED CONDITIONS*** | | | | | | | | | | |
| *MENOPAUSAL EFFECTS ON THE GENITOURINARY TRACT* | | | | | | | | | | |
| POSTMENOPAUSAL HAEMORRHAGE | 0 | 0 | 0 | 0 | 0 | 0 | 0 | 0 | 0 | 0 |
| ***MENSTRUAL CYCLE AND UTERINE BLEEDING DISORDERS*** | | | | | | | | | | |
| *MENSTRUATION AND UTERINE BLEEDING NEC* | | | | | | | | | | |
| DYSMENORRHOEA | 2 | 1 | 1 | 0 | 0 | 1 | 1 | 0 | 0 | 0 |
| INTERMENSTRUAL BLEEDING | 2 | 0 | 0 | 2 | 0 | 1 | 0 | 0 | 1 | 0 |
| MENSTRUAL DISORDER | 2 | 0 | 0 | 2 | 0 | 2 | 0 | 0 | 2 | 0 |
| MENSTRUATION IRREGULAR | 5 | 2 | 2 | 1 | 0 | 4 | 2 | 1 | 1 | 0 |
| PREMENSTRUAL PAIN | 1 | 1 | 0 | 0 | 0 | 1 | 1 | 0 | 0 | 0 |
| RETROGRADE MENSTRUATION | 0 | 0 | 0 | 0 | 0 | 0 | 0 | 0 | 0 | 0 |
| *MENSTRUATION WITH DECREASED BLEEDING* | | | | | | | | | | |
| AMENORRHOEA | 0 | 0 | 0 | 0 | 0 | 0 | 0 | 0 | 0 | 0 |
| HYPOMENORRHOEA | 0 | 0 | 0 | 0 | 0 | 0 | 0 | 0 | 0 | 0 |
| MENSTRUATION DELAYED | 1 | 0 | 1 | 0 | 0 | 1 | 0 | 1 | 0 | 0 |
| OLIGOMENORRHOEA | 0 | 0 | 0 | 0 | 0 | 0 | 0 | 0 | 0 | 0 |
| *MENSTRUATION WITH INCREASED BLEEDING* | | | | | | | | | | |
| HEAVY MENSTRUAL BLEEDING | 7 | 4 | 1 | 2 | 0 | 6 | 4 | 0 | 2 | 0 |
| POLYMENORRHOEA | 0 | 0 | 0 | 0 | 0 | 0 | 0 | 0 | 0 | 0 |
| ***PENILE AND SCROTAL DISORDERS (EXCL INFECTIONS AND INFLAMMATIONS)*** | | | | | | | | | | |
| *SCROTAL DISORDERS NEC* | | | | | | | | | | |
| SCROTAL SWELLING | 0 | 0 | 0 | 0 | 0 | 0 | 0 | 0 | 0 | 0 |
| ***REPRODUCTIVE TRACT DISORDERS NEC*** | | | | | | | | | | |
| *REPRODUCTIVE TRACT DISORDERS NEC (EXCL NEOPLASMS)* | | | | | | | | | | |
| GENITAL LESION | 0 | 0 | 0 | 0 | 0 | 0 | 0 | 0 | 0 | 0 |
| *REPRODUCTIVE TRACT SIGNS AND SYMPTOMS NEC* | | | | | | | | | | |
| GENITAL DISCOMFORT | 0 | 0 | 0 | 0 | 0 | 0 | 0 | 0 | 0 | 0 |
| GENITAL PAIN | 0 | 0 | 0 | 0 | 0 | 0 | 0 | 0 | 0 | 0 |
| ***UTERINE, PELVIC AND BROAD LIGAMENT DISORDERS*** | | | | | | | | | | |
| *PELVIS AND BROAD LIGAMENT DISORDERS NEC* | | | | | | | | | | |

| | | | | | | | | | | |
|---|---|---|---|---|---|---|---|---|---|---|
| ADNEXA UTERI PAIN | 0 | 0 | 0 | 0 | 0 | 0 | 0 | 0 | 0 | 0 |
| ***VULVOVAGINAL DISORDERS (EXCL INFECTIONS AND INFLAMMATIONS)*** | | | | | | | | | | |
| *VULVOVAGINAL DISORDERS NEC* | | | | | | | | | | |
| VAGINAL HAEMORRHAGE | 0 | 0 | 0 | 0 | 0 | 0 | 0 | 0 | 0 | 0 |
| *VULVOVAGINAL SIGNS AND SYMPTOMS* | | | | | | | | | | |
| VAGINAL DISCHARGE | 0 | 0 | 0 | 0 | 0 | 0 | 0 | 0 | 0 | 0 |
| **RESPIRATORY, THORACIC AND MEDIASTINAL DISORDERS** | | | | | | | | | | |
| ***BRONCHIAL DISORDERS (EXCL NEOPLASMS)*** | | | | | | | | | | |
| *BRONCHOSPASM AND OBSTRUCTION* | | | | | | | | | | |
| ASTHMA | 0 | 0 | 0 | 0 | 0 | 0 | 0 | 0 | 0 | 0 |
| WHEEZING | 1 | 0 | 1 | 0 | 0 | 1 | 0 | 1 | 0 | 0 |
| ***LOWER RESPIRATORY TRACT DISORDERS (EXCL OBSTRUCTION AND INFECTION)*** | | | | | | | | | | |
| *PULMONARY OEDEMAS* | | | | | | | | | | |
| PULMONARY CONGESTION | 0 | 0 | 0 | 0 | 0 | 0 | 0 | 0 | 0 | 0 |
| ***PULMONARY VASCULAR DISORDERS*** | | | | | | | | | | |
| *PULMONARY THROMBOTIC AND EMBOLIC CONDITIONS* | | | | | | | | | | |
| PULMONARY EMBOLISM | 0 | 0 | 0 | 0 | 0 | 0 | 0 | 0 | 0 | 0 |
| ***RESPIRATORY DISORDERS NEC*** | | | | | | | | | | |
| *BREATHING ABNORMALITIES* | | | | | | | | | | |
| DYSPNOEA | 10 | 2 | 2 | 3 | 3 | 9 | 2 | 2 | 2 | 3 |
| HYPERVENTILATION | 0 | 0 | 0 | 0 | 0 | 0 | 0 | 0 | 0 | 0 |
| HYPOPNOEA | 1 | 0 | 1 | 0 | 0 | 1 | 0 | 1 | 0 | 0 |
| IRREGULAR BREATHING | 0 | 0 | 0 | 0 | 0 | 0 | 0 | 0 | 0 | 0 |
| MOUTH BREATHING | 0 | 0 | 0 | 0 | 0 | 0 | 0 | 0 | 0 | 0 |
| RESPIRATION ABNORMAL | 0 | 0 | 0 | 0 | 0 | 0 | 0 | 0 | 0 | 0 |
| RESPIRATORY ARREST | 0 | 0 | 0 | 0 | 0 | 0 | 0 | 0 | 0 | 0 |
| RESPIRATORY FATIGUE | 0 | 0 | 0 | 0 | 0 | 0 | 0 | 0 | 0 | 0 |
| SLEEP APNOEA SYNDROME | 0 | 0 | 0 | 0 | 0 | 0 | 0 | 0 | 0 | 0 |
| *COUGHING AND ASSOCIATED SYMPTOMS* | | | | | | | | | | |
| COUGH | 10 | 3 | 0 | 5 | 2 | 8 | 3 | 0 | 4 | 1 |
| PRODUCTIVE COUGH | 0 | 0 | 0 | 0 | 0 | 0 | 0 | 0 | 0 | 0 |
| *RESPIRATORY TRACT DISORDERS NEC* | | | | | | | | | | |
| RESPIRATORY TRACT IRRITATION | 0 | 0 | 0 | 0 | 0 | 0 | 0 | 0 | 0 | 0 |
| ***RESPIRATORY TRACT SIGNS AND SYMPTOMS*** | | | | | | | | | | |
| *LOWER RESPIRATORY TRACT SIGNS AND SYMPTOMS* | | | | | | | | | | |
| PULMONARY PAIN | 0 | 0 | 0 | 0 | 0 | 0 | 0 | 0 | 0 | 0 |

| | | | | | | | | | | |
|---|---|---|---|---|---|---|---|---|---|---|
| *RESPIRATORY SIGNS AND SYMPTOMS NEC* | | | | | | | | | | |
| RESPIRATORY SYMPTOM | 0 | 0 | 0 | 0 | 0 | 0 | 0 | 0 | 0 | 0 |
| *UPPER RESPIRATORY TRACT SIGNS AND SYMPTOMS* | | | | | | | | | | |
| APHONIA | 0 | 0 | 0 | 0 | 0 | 0 | 0 | 0 | 0 | 0 |
| CATARRH | 2 | 0 | 0 | 2 | 0 | 1 | 0 | 0 | 1 | 0 |
| DRY THROAT | 0 | 0 | 0 | 0 | 0 | 0 | 0 | 0 | 0 | 0 |
| DYSPHONIA | 0 | 0 | 0 | 0 | 0 | 0 | 0 | 0 | 0 | 0 |
| INCREASED UPPER AIRWAY SECRETION | 0 | 0 | 0 | 0 | 0 | 0 | 0 | 0 | 0 | 0 |
| INCREASED VISCOSITY OF UPPER RESPIRATORY SE | 0 | 0 | 0 | 0 | 0 | 0 | 0 | 0 | 0 | 0 |
| NASAL DISCOMFORT | 0 | 0 | 0 | 0 | 0 | 0 | 0 | 0 | 0 | 0 |
| OROPHARYNGEAL DISCOMFORT | 0 | 0 | 0 | 0 | 0 | 0 | 0 | 0 | 0 | 0 |
| OROPHARYNGEAL PAIN | 13 | 6 | 1 | 4 | 2 | 13 | 6 | 1 | 4 | 2 |
| PARANASAL SINUS DISCOMFORT | 0 | 0 | 0 | 0 | 0 | 0 | 0 | 0 | 0 | 0 |
| RHINALGIA | 0 | 0 | 0 | 0 | 0 | 0 | 0 | 0 | 0 | 0 |
| RHINORRHOEA | 6 | 0 | 0 | 5 | 1 | 5 | 0 | 0 | 5 | 0 |
| SINUS PAIN | 2 | 1 | 1 | 0 | 0 | 2 | 1 | 1 | 0 | 0 |
| SNEEZING | 1 | 0 | 0 | 1 | 0 | 1 | 0 | 0 | 1 | 0 |
| THROAT CLEARING | 0 | 0 | 0 | 0 | 0 | 0 | 0 | 0 | 0 | 0 |
| THROAT IRRITATION | 1 | 1 | 0 | 0 | 0 | 1 | 1 | 0 | 0 | 0 |
| THROAT TIGHTNESS | 0 | 0 | 0 | 0 | 0 | 0 | 0 | 0 | 0 | 0 |
| UPPER-AIRWAY COUGH SYNDROME | 0 | 0 | 0 | 0 | 0 | 0 | 0 | 0 | 0 | 0 |
| YAWNING | 0 | 0 | 0 | 0 | 0 | 0 | 0 | 0 | 0 | 0 |
| ***UPPER RESPIRATORY TRACT DISORDERS (EXCL INFECTIONS)*** | | | | | | | | | | |
| *NASAL CONGESTION AND INFLAMMATIONS* | | | | | | | | | | |
| NASAL CONGESTION | 2 | 0 | 1 | 1 | 0 | 2 | 0 | 1 | 1 | 0 |
| RHINITIS ALLERGIC | 0 | 0 | 0 | 0 | 0 | 0 | 0 | 0 | 0 | 0 |
| RHINITIS ATROPHIC | 0 | 0 | 0 | 0 | 0 | 0 | 0 | 0 | 0 | 0 |
| *NASAL DISORDERS NEC* | | | | | | | | | | |
| EPISTAXIS | 1 | 1 | 0 | 0 | 0 | 1 | 1 | 0 | 0 | 0 |
| NASAL DRYNESS | 0 | 0 | 0 | 0 | 0 | 0 | 0 | 0 | 0 | 0 |
| *PARANASAL SINUS DISORDERS (EXCL INFECTIONS AND NEOPLASMS)* | | | | | | | | | | |
| SINUS CONGESTION | 0 | 0 | 0 | 0 | 0 | 0 | 0 | 0 | 0 | 0 |
| *PHARYNGEAL DISORDERS (EXCL INFECTIONS AND NEOPLASMS)* | | | | | | | | | | |
| PHARYNGEAL SWELLING | 1 | 0 | 1 | 0 | 0 | 1 | 0 | 1 | 0 | 0 |
| PHARYNGEAL ULCERATION | 0 | 0 | 0 | 0 | 0 | 0 | 0 | 0 | 0 | 0 |

| | | | | | | | | | | |
|---|---|---|---|---|---|---|---|---|---|---|
| TONSILLAR ERYTHEMA | 0 | 0 | 0 | 0 | 0 | 0 | 0 | 0 | 0 | 0 |
| *TRACHEAL DISORDERS (EXCL INFECTIONS AND NEOPLASMS)* | | | | | | | | | | |
| TRACHEAL PAIN | 0 | 0 | 0 | 0 | 0 | 0 | 0 | 0 | 0 | 0 |
| **SKIN AND SUBCUTANEOUS TISSUE DISORDERS** | | | | | | | | | | |
| ***ANGIOEDEMA AND URTICARIA*** | | | | | | | | | | |
| *ANGIOEDEMAS* | | | | | | | | | | |
| ANGIOEDEMA | 0 | 0 | 0 | 0 | 0 | 0 | 0 | 0 | 0 | 0 |
| *URTICARIAS* | | | | | | | | | | |
| COLD URTICARIA | 0 | 0 | 0 | 0 | 0 | 0 | 0 | 0 | 0 | 0 |
| SOLAR URTICARIA | 0 | 0 | 0 | 0 | 0 | 0 | 0 | 0 | 0 | 0 |
| URTICARIA | 2 | 0 | 0 | 2 | 0 | 2 | 0 | 0 | 2 | 0 |
| URTICARIA CHRONIC | 0 | 0 | 0 | 0 | 0 | 0 | 0 | 0 | 0 | 0 |
| ***CORNIFICATION AND DYSTROPHIC SKIN DISORDERS*** | | | | | | | | | | |
| *SKIN DYSTROPHIES* | | | | | | | | | | |
| HYPERTROPHIC SCAR | 0 | 0 | 0 | 0 | 0 | 0 | 0 | 0 | 0 | 0 |
| ***CUTANEOUS NEOPLASMS BENIGN*** | | | | | | | | | | |
| *SKIN CYSTS AND POLYPS* | | | | | | | | | | |
| DERMAL CYST | 0 | 0 | 0 | 0 | 0 | 0 | 0 | 0 | 0 | 0 |
| ***EPIDERMAL AND DERMAL CONDITIONS*** | | | | | | | | | | |
| *BULLOUS CONDITIONS* | | | | | | | | | | |
| BLISTER | 0 | 0 | 0 | 0 | 0 | 0 | 0 | 0 | 0 | 0 |
| BLOOD BLISTER | 0 | 0 | 0 | 0 | 0 | 0 | 0 | 0 | 0 | 0 |
| PEMPHIGOID | 0 | 0 | 0 | 0 | 0 | 0 | 0 | 0 | 0 | 0 |
| TOXIC EPIDERMAL NECROLYSIS | 0 | 0 | 0 | 0 | 0 | 0 | 0 | 0 | 0 | 0 |
| *DERMAL AND EPIDERMAL CONDITIONS NEC* | | | | | | | | | | |
| DRY SKIN | 1 | 0 | 0 | 1 | 0 | 1 | 0 | 0 | 1 | 0 |
| PAIN OF SKIN | 2 | 0 | 0 | 1 | 1 | 1 | 0 | 0 | 1 | 0 |
| SENSITIVE SKIN | 1 | 1 | 0 | 0 | 0 | 1 | 1 | 0 | 0 | 0 |
| SKIN BURNING SENSATION | 0 | 0 | 0 | 0 | 0 | 0 | 0 | 0 | 0 | 0 |
| SKIN DISCOLOURATION | 0 | 0 | 0 | 0 | 0 | 0 | 0 | 0 | 0 | 0 |
| SKIN FRAGILITY | 0 | 0 | 0 | 0 | 0 | 0 | 0 | 0 | 0 | 0 |
| SKIN LESION | 0 | 0 | 0 | 0 | 0 | 0 | 0 | 0 | 0 | 0 |
| SKIN ODOUR ABNORMAL | 0 | 0 | 0 | 0 | 0 | 0 | 0 | 0 | 0 | 0 |
| SKIN REACTION | 1 | 1 | 0 | 0 | 0 | 1 | 1 | 0 | 0 | 0 |
| SKIN SENSITISATION | 0 | 0 | 0 | 0 | 0 | 0 | 0 | 0 | 0 | 0 |

| | | | | | | | | | | |
|---|---|---|---|---|---|---|---|---|---|---|
| SKIN WARM | 2 | 0 | 1 | 1 | 0 | 1 | 0 | 0 | 1 | 0 |
| *DERMATITIS AND ECZEMA* | | | | | | | | | | |
| DERMATITIS | 0 | 0 | 0 | 0 | 0 | 0 | 0 | 0 | 0 | 0 |
| DERMATITIS ALLERGIC | 0 | 0 | 0 | 0 | 0 | 0 | 0 | 0 | 0 | 0 |
| DERMATITIS ATOPIC | 0 | 0 | 0 | 0 | 0 | 0 | 0 | 0 | 0 | 0 |
| DERMATITIS CONTACT | 0 | 0 | 0 | 0 | 0 | 0 | 0 | 0 | 0 | 0 |
| ECZEMA | 0 | 0 | 0 | 0 | 0 | 0 | 0 | 0 | 0 | 0 |
| ECZEMA ASTEATOTIC | 0 | 0 | 0 | 0 | 0 | 0 | 0 | 0 | 0 | 0 |
| NEURODERMATITIS | 0 | 0 | 0 | 0 | 0 | 0 | 0 | 0 | 0 | 0 |
| SEBORRHOEIC DERMATITIS | 0 | 0 | 0 | 0 | 0 | 0 | 0 | 0 | 0 | 0 |
| SKIN IRRITATION | 0 | 0 | 0 | 0 | 0 | 0 | 0 | 0 | 0 | 0 |
| *ERYTHEMAS* | | | | | | | | | | |
| ERYTHEMA | 4 | 1 | 0 | 2 | 1 | 3 | 1 | 0 | 2 | 0 |
| *EXFOLIATIVE CONDITIONS* | | | | | | | | | | |
| SKIN EXFOLIATION | 1 | 0 | 0 | 0 | 1 | 0 | 0 | 0 | 0 | 0 |
| *PAPULOSQUAMOUS CONDITIONS* | | | | | | | | | | |
| LICHEN PLANUS | 0 | 0 | 0 | 0 | 0 | 0 | 0 | 0 | 0 | 0 |
| PITYRIASIS ROSEA | 0 | 0 | 0 | 0 | 0 | 0 | 0 | 0 | 0 | 0 |
| *PHOTOSENSITIVITY AND PHOTODERMATOSIS CONDITIONS* | | | | | | | | | | |
| PHOTOSENSITIVITY REACTION | 0 | 0 | 0 | 0 | 0 | 0 | 0 | 0 | 0 | 0 |
| *PRURITUS NEC* | | | | | | | | | | |
| PRURITUS | 11 | 4 | 3 | 2 | 2 | 11 | 4 | 3 | 2 | 2 |
| *PSORIATIC CONDITIONS* | | | | | | | | | | |
| PSORIASIS | 1 | 0 | 1 | 0 | 0 | 1 | 0 | 1 | 0 | 0 |
| *RASHES, ERUPTIONS AND EXANTHEMS NEC* | | | | | | | | | | |
| RASH | 33 | 15 | 4 | 12 | 2 | 26 | 15 | 1 | 8 | 2 |
| RASH ERYTHEMATOUS | 5 | 3 | 0 | 2 | 0 | 5 | 3 | 0 | 2 | 0 |
| RASH MACULAR | 1 | 1 | 0 | 0 | 0 | 1 | 1 | 0 | 0 | 0 |
| RASH MORBILLIFORM | 0 | 0 | 0 | 0 | 0 | 0 | 0 | 0 | 0 | 0 |
| RASH PAPULAR | 1 | 1 | 0 | 0 | 0 | 1 | 1 | 0 | 0 | 0 |
| RASH PRURITIC | 6 | 1 | 0 | 4 | 1 | 6 | 1 | 0 | 4 | 1 |
| *SKIN INJURIES AND MECHANICAL DERMATOSES* | | | | | | | | | | |
| DECUBITUS ULCER | 0 | 0 | 0 | 0 | 0 | 0 | 0 | 0 | 0 | 0 |
| NEEDLE TRACK MARKS | 0 | 0 | 0 | 0 | 0 | 0 | 0 | 0 | 0 | 0 |
| ***SKIN AND SUBCUTANEOUS TISSUE DISORDERS NEC*** | | | | | | | | | | |

| | | | | | | | | | | |
|---|---|---|---|---|---|---|---|---|---|---|
| *SKIN AND SUBCUTANEOUS TISSUE ULCERATIONS* | | | | | | | | | | |
| SKIN EROSION | 0 | 0 | 0 | 0 | 0 | 0 | 0 | 0 | 0 | 0 |
| ***SKIN APPENDAGE CONDITIONS*** | | | | | | | | | | |
| *ACNES* | | | | | | | | | | |
| ACNE | 0 | 0 | 0 | 0 | 0 | 0 | 0 | 0 | 0 | 0 |
| ACNE CYSTIC | 0 | 0 | 0 | 0 | 0 | 0 | 0 | 0 | 0 | 0 |
| *ALOPECIAS* | | | | | | | | | | |
| ALOPECIA | 1 | 0 | 0 | 1 | 0 | 1 | 0 | 0 | 1 | 0 |
| *APOCRINE AND ECCRINE GLAND DISORDERS* | | | | | | | | | | |
| COLD SWEAT | 5 | 3 | 1 | 1 | 0 | 5 | 3 | 1 | 1 | 0 |
| HYPERHIDROSIS | 10 | 1 | 4 | 4 | 1 | 9 | 1 | 3 | 4 | 1 |
| MILIARIA | 0 | 0 | 0 | 0 | 0 | 0 | 0 | 0 | 0 | 0 |
| NIGHT SWEATS | 5 | 1 | 2 | 2 | 0 | 5 | 1 | 2 | 2 | 0 |
| *HYPERTRICHOSES* | | | | | | | | | | |
| HIRSUTISM | 0 | 0 | 0 | 0 | 0 | 0 | 0 | 0 | 0 | 0 |
| *NAIL AND NAIL BED CONDITIONS (EXCL INFECTIONS AND INFESTATIONS)* | | | | | | | | | | |
| NAIL DISCOLOURATION | 0 | 0 | 0 | 0 | 0 | 0 | 0 | 0 | 0 | 0 |
| ONYCHOCLASIS | 0 | 0 | 0 | 0 | 0 | 0 | 0 | 0 | 0 | 0 |
| *PILAR DISORDERS NEC* | | | | | | | | | | |
| PILOERECTION | 0 | 0 | 0 | 0 | 0 | 0 | 0 | 0 | 0 | 0 |
| ***SKIN VASCULAR ABNORMALITIES*** | | | | | | | | | | |
| *PURPURA AND RELATED CONDITIONS* | | | | | | | | | | |
| PETECHIAE | 0 | 0 | 0 | 0 | 0 | 0 | 0 | 0 | 0 | 0 |
| PURPURA | 1 | 0 | 0 | 0 | 1 | 1 | 0 | 0 | 0 | 1 |
| *SKIN HAEMORRHAGES* | | | | | | | | | | |
| SKIN HAEMORRHAGE | 0 | 0 | 0 | 0 | 0 | 0 | 0 | 0 | 0 | 0 |
| **SOCIAL CIRCUMSTANCES** | | | | | | | | | | |
| ***ECONOMIC AND HOUSING ISSUES*** | | | | | | | | | | |
| *EMPLOYMENT ISSUES* | | | | | | | | | | |
| RETIREMENT | 0 | 0 | 0 | 0 | 0 | 0 | 0 | 0 | 0 | 0 |
| ***ENVIRONMENTAL ISSUES*** | | | | | | | | | | |
| *NON-OCCUPATIONAL AND UNSPECIFIED ENVIRONMENTAL PROBLEMS* | | | | | | | | | | |
| POLLUTION | 0 | 0 | 0 | 0 | 0 | 0 | 0 | 0 | 0 | 0 |
| **SURGICAL AND MEDICAL PROCEDURES** | | | | | | | | | | |
| ***BONE AND JOINT THERAPEUTIC PROCEDURES*** | | | | | | | | | | |
| *JOINT THERAPEUTIC PROCEDURES* | | | | | | | | | | |

| | | | | | | | | | | |
|---|---|---|---|---|---|---|---|---|---|---|
| KNEE OPERATION | 0 | 0 | 0 | 0 | 0 | 0 | 0 | 0 | 0 | 0 |
| ***BREAST THERAPEUTIC PROCEDURES*** | | | | | | | | | | |
| *MASTECTOMIES* | | | | | | | | | | |
| BREAST CONSERVING SURGERY | 0 | 0 | 0 | 0 | 0 | 0 | 0 | 0 | 0 | 0 |
| ***GASTROINTESTINAL THERAPEUTIC PROCEDURES*** | | | | | | | | | | |
| *GASTROINTESTINAL THERAPEUTIC PROCEDURES NEC* | | | | | | | | | | |
| PROPHYLAXIS OF NAUSEA AND VOMITING | 0 | 0 | 0 | 0 | 0 | 0 | 0 | 0 | 0 | 0 |
| ***HEAD AND NECK THERAPEUTIC PROCEDURES*** | | | | | | | | | | |
| *DENTAL AND GINGIVAL THERAPEUTIC PROCEDURES* | | | | | | | | | | |
| DENTAL CARE | 0 | 0 | 0 | 0 | 0 | 0 | 0 | 0 | 0 | 0 |
| ***NERVOUS SYSTEM, SKULL AND SPINE THERAPEUTIC PROCEDURES*** | | | | | | | | | | |
| *SKULL AND BRAIN THERAPEUTIC PROCEDURES* | | | | | | | | | | |
| BRAIN TUMOUR OPERATION | 0 | 0 | 0 | 0 | 0 | 0 | 0 | 0 | 0 | 0 |
| ***OBSTETRIC AND GYNAECOLOGICAL THERAPEUTIC PROCEDURES*** | | | | | | | | | | |
| *FERTILITY AND FERTILISATION INTERVENTIONS FEMALE* | | | | | | | | | | |
| ENDOMETRIAL SCRATCHING | 0 | 0 | 0 | 0 | 0 | 0 | 0 | 0 | 0 | 0 |
| ***RESPIRATORY TRACT THERAPEUTIC PROCEDURES*** | | | | | | | | | | |
| *RESPIRATORY TRACT THERAPEUTIC PROCEDURES NEC* | | | | | | | | | | |
| OXYGEN THERAPY | 0 | 0 | 0 | 0 | 0 | 0 | 0 | 0 | 0 | 0 |
| ***THERAPEUTIC PROCEDURES AND SUPPORTIVE CARE NEC*** | | | | | | | | | | |
| *ANAESTHESIA AND ALLIED PROCEDURES* | | | | | | | | | | |
| NERVE BLOCK | 0 | 0 | 0 | 0 | 0 | 0 | 0 | 0 | 0 | 0 |
| *DIETARY AND NUTRITIONAL THERAPIES* | | | | | | | | | | |
| NOTHING BY MOUTH ORDER | 0 | 0 | 0 | 0 | 0 | 0 | 0 | 0 | 0 | 0 |
| *IMMUNISATIONS* | | | | | | | | | | |
| COVID-19 IMMUNISATION | 0 | 0 | 0 | 0 | 0 | 0 | 0 | 0 | 0 | 0 |
| *THERAPEUTIC PROCEDURES NEC* | | | | | | | | | | |
| INJECTION | 0 | 0 | 0 | 0 | 0 | 0 | 0 | 0 | 0 | 0 |
| LOCALISED ALTERNATING HOT AND COLD THERAPY | 0 | 0 | 0 | 0 | 0 | 0 | 0 | 0 | 0 | 0 |
| MASS EXCISION | 0 | 0 | 0 | 0 | 0 | 0 | 0 | 0 | 0 | 0 |
| REINFUSION | 1 | 0 | 0 | 1 | 0 | 1 | 0 | 0 | 1 | 0 |
| **VASCULAR DISORDERS** | | | | | | | | | | |
| ***ARTERIOSCLEROSIS, STENOSIS, VASCULAR INSUFFICIENCY AND NECROSIS*** | | | | | | | | | | |
| *NON-SITE SPECIFIC NECROSIS AND VASCULAR INSUFFICIENCY NEC* | | | | | | | | | | |
| VASOSPASM | 0 | 0 | 0 | 0 | 0 | 0 | 0 | 0 | 0 | 0 |
| *PERIPHERAL VASOCONSTRICTION, NECROSIS AND VASCULAR INSUFFICIENCY* | | | | | | | | | | |

| | | | | | | | | | | |
|---|---|---|---|---|---|---|---|---|---|---|
| PERIPHERAL COLDNESS | 2 | 0 | 0 | 2 | 0 | 2 | 0 | 0 | 2 | 0 |
| RAYNAUD'S PHENOMENON | 0 | 0 | 0 | 0 | 0 | 0 | 0 | 0 | 0 | 0 |
| ***DECREASED AND NONSPECIFIC BLOOD PRESSURE DISORDERS AND SHOCK*** | | | | | | | | | | |
| *BLOOD PRESSURE DISORDERS NEC* | | | | | | | | | | |
| BLOOD PRESSURE FLUCTUATION | 0 | 0 | 0 | 0 | 0 | 0 | 0 | 0 | 0 | 0 |
| *VASCULAR HYPOTENSIVE DISORDERS* | | | | | | | | | | |
| CAPILLARY LEAK SYNDROME | 0 | 0 | 0 | 0 | 0 | 0 | 0 | 0 | 0 | 0 |
| HYPOTENSION | 1 | 0 | 0 | 1 | 0 | 0 | 0 | 0 | 0 | 0 |
| ORTHOSTATIC HYPOTENSION | 0 | 0 | 0 | 0 | 0 | 0 | 0 | 0 | 0 | 0 |
| ***EMBOLISM AND THROMBOSIS*** | | | | | | | | | | |
| *NON-SITE SPECIFIC EMBOLISM AND THROMBOSIS* | | | | | | | | | | |
| EMBOLISM | 1 | 1 | 0 | 0 | 0 | 1 | 1 | 0 | 0 | 0 |
| THROMBOSIS | 0 | 0 | 0 | 0 | 0 | 0 | 0 | 0 | 0 | 0 |
| VENOUS THROMBOSIS | 0 | 0 | 0 | 0 | 0 | 0 | 0 | 0 | 0 | 0 |
| *PERIPHERAL EMBOLISM AND THROMBOSIS* | | | | | | | | | | |
| BLUE TOE SYNDROME | 0 | 0 | 0 | 0 | 0 | 0 | 0 | 0 | 0 | 0 |
| DEEP VEIN THROMBOSIS | 0 | 0 | 0 | 0 | 0 | 0 | 0 | 0 | 0 | 0 |
| SUPERFICIAL VEIN THROMBOSIS | 0 | 0 | 0 | 0 | 0 | 0 | 0 | 0 | 0 | 0 |
| ***LYMPHATIC VESSEL DISORDERS*** | | | | | | | | | | |
| *LYMPHOEDEMAS* | | | | | | | | | | |
| LYMPHOEDEMA | 0 | 0 | 0 | 0 | 0 | 0 | 0 | 0 | 0 | 0 |
| ***VASCULAR DISORDERS NEC*** | | | | | | | | | | |
| *NON-SITE SPECIFIC VASCULAR DISORDERS NEC* | | | | | | | | | | |
| VASCULAR PAIN | 0 | 0 | 0 | 0 | 0 | 0 | 0 | 0 | 0 | 0 |
| VEIN DISCOLOURATION | 0 | 0 | 0 | 0 | 0 | 0 | 0 | 0 | 0 | 0 |
| VEIN RUPTURE | 0 | 0 | 0 | 0 | 0 | 0 | 0 | 0 | 0 | 0 |
| *PERIPHERAL VASCULAR DISORDERS NEC* | | | | | | | | | | |
| FLUSHING | 0 | 0 | 0 | 0 | 0 | 0 | 0 | 0 | 0 | 0 |
| HOT FLUSH | 4 | 1 | 0 | 3 | 0 | 2 | 1 | 0 | 1 | 0 |
| *SITE SPECIFIC VASCULAR DISORDERS NEC* | | | | | | | | | | |
| PALLOR | 1 | 0 | 1 | 0 | 0 | 0 | 0 | 0 | 0 | 0 |
| ***VASCULAR HAEMORRHAGIC DISORDERS*** | | | | | | | | | | |
| *HAEMORRHAGES NEC* | | | | | | | | | | |
| HAEMATOMA | 0 | 0 | 0 | 0 | 0 | 0 | 0 | 0 | 0 | 0 |
| HAEMORRHAGE | 0 | 0 | 0 | 0 | 0 | 0 | 0 | 0 | 0 | 0 |
| ***VASCULAR HYPERTENSIVE DISORDERS*** | | | | | | | | | | |

| | | | | | | | | | | |
|---|---|---|---|---|---|---|---|---|---|---|
| *ACCELERATED AND MALIGNANT HYPERTENSION* | | | | | | | | | | |
| HYPERTENSIVE URGENCY | 0 | 0 | 0 | 0 | 0 | 0 | 0 | 0 | 0 | 0 |
| *VASCULAR HYPERTENSIVE DISORDERS NEC* | | | | | | | | | | |
| HYPERTENSION | 0 | 0 | 0 | 0 | 0 | 0 | 0 | 0 | 0 | 0 |
| SYSTOLIC HYPERTENSION | 0 | 0 | 0 | 0 | 0 | 0 | 0 | 0 | 0 | 0 |
| ***VASCULAR INFECTIONS AND INFLAMMATIONS*** | | | | | | | | | | |
| *ARTERIAL INFECTIONS AND INFLAMMATIONS* | | | | | | | | | | |
| GIANT CELL ARTERITIS | 0 | 0 | 0 | 0 | 0 | 0 | 0 | 0 | 0 | 0 |
| *PHLEBITIS NEC* | | | | | | | | | | |
| PHLEBITIS | 0 | 0 | 0 | 0 | 0 | 0 | 0 | 0 | 0 | 0 |
| *VASCULITIDES NEC* | | | | | | | | | | |
| VASCULITIS | 1 | 0 | 0 | 0 | 1 | 1 | 0 | 0 | 0 | 1 |
| ***VENOUS VARICES*** | | | | | | | | | | |
| *VARICOSE VEINS NEC* | | | | | | | | | | |
| SPIDER VEIN | 0 | 0 | 0 | 0 | 0 | 0 | 0 | 0 | 0 | 0 |
| VARICOSE VEIN | 0 | 0 | 0 | 0 | 0 | 0 | 0 | 0 | 0 | 0 |
| **TOTAL ADR EVENTS** | ***2581*** | ***623*** | ***367*** | ***1227*** | ***364*** | ***2230*** | ***623*** | ***281*** | ***1034*** | ***292*** |

**TABLE S1-5. Others/Unknown COVID-19 vaccine: ADR listings for events reported in the YCVM in (a) those reporting any vaccination dose and (b) in those who had reported a 1st dose vaccination and any subsequent**

| | Patients with any vaccination dose: ADR Counts | | | | | Patients reporting a 1st dose: ADR Counts | | | | |
|---|---|---|---|---|---|---|---|---|---|---|
| **REACTION TERM (SOC, *HLGT*, *HLT*, PT)** | All doses | 1st dose | 2nd dose | 3rd dose | Other doses | All doses | 1st dose | 2nd dose | 3rd dose | Other doses |
| ***(freetext)*** | 2 | 2 | 0 | 0 | 0 | 2 | 2 | 0 | 0 | 0 |
| **BLOOD AND LYMPHATIC SYSTEM DISORDERS** | | | | | | | | | | |
| ***COAGULOPATHIES AND BLEEDING DIATHESES (EXCL THROMBOCYTOPENIC)*** | | | | | | | | | | |
| *BLEEDING TENDENCIES* | | | | | | | | | | |
| INCREASED TENDENCY TO BRUISE | 0 | 0 | 0 | 0 | 0 | 0 | 0 | 0 | 0 | 0 |
| ***HAEMOGLOBINOPATHIES*** | | | | | | | | | | |
| *SICKLE CELL TRAIT AND DISORDERS* | | | | | | | | | | |
| SICKLE CELL ANAEMIA WITH CRISIS | 0 | 0 | 0 | 0 | 0 | 0 | 0 | 0 | 0 | 0 |
| ***PLATELET DISORDERS*** | | | | | | | | | | |
| *THROMBOCYTOPENIAS* | | | | | | | | | | |
| IMMUNE THROMBOCYTOPENIA | 0 | 0 | 0 | 0 | 0 | 0 | 0 | 0 | 0 | 0 |
| ***SPLEEN, LYMPHATIC AND RETICULOENDOTHELIAL SYSTEM DISORDERS*** | | | | | | | | | | |
| *LYMPHATIC SYSTEM DISORDERS NEC* | | | | | | | | | | |
| LYMPH NODE PAIN | 0 | 0 | 0 | 0 | 0 | 0 | 0 | 0 | 0 | 0 |
| LYMPHADENITIS | 0 | 0 | 0 | 0 | 0 | 0 | 0 | 0 | 0 | 0 |
| LYMPHADENOPATHY | 0 | 0 | 0 | 0 | 0 | 0 | 0 | 0 | 0 | 0 |
| **CARDIAC DISORDERS** | | | | | | | | | | |
| ***CARDIAC ARRHYTHMIAS*** | | | | | | | | | | |
| *RATE AND RHYTHM DISORDERS NEC* | | | | | | | | | | |
| ARRHYTHMIA | 0 | 0 | 0 | 0 | 0 | 0 | 0 | 0 | 0 | 0 |
| CARDIAC FLUTTER | 0 | 0 | 0 | 0 | 0 | 0 | 0 | 0 | 0 | 0 |
| EXTRASYSTOLES | 0 | 0 | 0 | 0 | 0 | 0 | 0 | 0 | 0 | 0 |
| TACHYCARDIA | 0 | 0 | 0 | 0 | 0 | 0 | 0 | 0 | 0 | 0 |
| *SUPRAVENTRICULAR ARRHYTHMIAS* | | | | | | | | | | |
| ATRIAL FIBRILLATION | 0 | 0 | 0 | 0 | 0 | 0 | 0 | 0 | 0 | 0 |
| SUPRAVENTRICULAR TACHYCARDIA | 0 | 0 | 0 | 0 | 0 | 0 | 0 | 0 | 0 | 0 |

| | | | | | | | | | | |
|---|---|---|---|---|---|---|---|---|---|---|
| ***CARDIAC DISORDERS, SIGNS AND SYMPTOMS NEC*** | | | | | | | | | | |
| *CARDIAC DISORDERS NEC* | | | | | | | | | | |
| CARDIOVASCULAR DISORDER | 0 | 0 | 0 | 0 | 0 | 0 | 0 | 0 | 0 | 0 |
| *CARDIAC SIGNS AND SYMPTOMS NEC* | | | | | | | | | | |
| PALPITATIONS | 1 | 1 | 0 | 0 | 0 | 1 | 1 | 0 | 0 | 0 |
| ***CORONARY ARTERY DISORDERS*** | | | | | | | | | | |
| *ISCHAEMIC CORONARY ARTERY DISORDERS* | | | | | | | | | | |
| ANGINA PECTORIS | 0 | 0 | 0 | 0 | 0 | 0 | 0 | 0 | 0 | 0 |
| MYOCARDIAL INFARCTION | 0 | 0 | 0 | 0 | 0 | 0 | 0 | 0 | 0 | 0 |
| ***HEART FAILURES*** | | | | | | | | | | |
| *HEART FAILURES NEC* | | | | | | | | | | |
| CARDIAC FAILURE | 0 | 0 | 0 | 0 | 0 | 0 | 0 | 0 | 0 | 0 |
| ***MYOCARDIAL DISORDERS*** | | | | | | | | | | |
| *NONINFECTIOUS MYOCARDITIS* | | | | | | | | | | |
| MYOCARDITIS | 0 | 0 | 0 | 0 | 0 | 0 | 0 | 0 | 0 | 0 |
| ***PERICARDIAL DISORDERS*** | | | | | | | | | | |
| *NONINFECTIOUS PERICARDITIS* | | | | | | | | | | |
| PERICARDITIS | 0 | 0 | 0 | 0 | 0 | 0 | 0 | 0 | 0 | 0 |
| **CONGENITAL, FAMILIAL AND GENETIC DISORDERS** | | | | | | | | | | |
| ***CARDIAC AND VASCULAR DISORDERS CONGENITAL*** | | | | | | | | | | |
| *CARDIAC DISORDERS CONGENITAL NEC* | | | | | | | | | | |
| HEART DISEASE CONGENITAL | 0 | 0 | 0 | 0 | 0 | 0 | 0 | 0 | 0 | 0 |
| ***METABOLIC AND NUTRITIONAL DISORDERS CONGENITAL*** | | | | | | | | | | |
| *INBORN ERRORS OF AMINO ACID METABOLISM* | | | | | | | | | | |
| HYPERGLYCINAEMIA | 0 | 0 | 0 | 0 | 0 | 0 | 0 | 0 | 0 | 0 |
| *INBORN ERRORS OF STEROID SYNTHESIS* | | | | | | | | | | |
| 11-BETA-HYDROXYLASE DEFICIENCY | 1 | 1 | 0 | 0 | 0 | 1 | 1 | 0 | 0 | 0 |
| ***NEUROLOGICAL DISORDERS CONGENITAL*** | | | | | | | | | | |
| *PERIPHERAL NERVOUS SYSTEM DISORDERS CONGENITAL NEC* | | | | | | | | | | |
| PAROXYSMAL EXTREME PAIN DISORDER | 0 | 0 | 0 | 0 | 0 | 0 | 0 | 0 | 0 | 0 |
| **EAR AND LABYRINTH DISORDERS** | | | | | | | | | | |

| | | | | | | | | | | |
|---|---|---|---|---|---|---|---|---|---|---|
| ***AURAL DISORDERS NEC*** | | | | | | | | | | |
| *EAR DISORDERS NEC* | | | | | | | | | | |
| EAR DISCOMFORT | 0 | 0 | 0 | 0 | 0 | 0 | 0 | 0 | 0 | 0 |
| EAR DISORDER | 0 | 0 | 0 | 0 | 0 | 0 | 0 | 0 | 0 | 0 |
| EAR PAIN | 0 | 0 | 0 | 0 | 0 | 0 | 0 | 0 | 0 | 0 |
| EAR SWELLING | 0 | 0 | 0 | 0 | 0 | 0 | 0 | 0 | 0 | 0 |
| ***EXTERNAL EAR DISORDERS (EXCL CONGENITAL)*** | | | | | | | | | | |
| *EXTERNAL EAR DISORDERS NEC* | | | | | | | | | | |
| EXCESSIVE CERUMEN PRODUCTION | 0 | 0 | 0 | 0 | 0 | 0 | 0 | 0 | 0 | 0 |
| ***HEARING DISORDERS*** | | | | | | | | | | |
| *HEARING LOSSES* | | | | | | | | | | |
| DEAFNESS | 0 | 0 | 0 | 0 | 0 | 0 | 0 | 0 | 0 | 0 |
| DEAFNESS NEUROSENSORY | 0 | 0 | 0 | 0 | 0 | 0 | 0 | 0 | 0 | 0 |
| DEAFNESS PERMANENT | 0 | 0 | 0 | 0 | 0 | 0 | 0 | 0 | 0 | 0 |
| DEAFNESS UNILATERAL | 0 | 0 | 0 | 0 | 0 | 0 | 0 | 0 | 0 | 0 |
| HYPOACUSIS | 0 | 0 | 0 | 0 | 0 | 0 | 0 | 0 | 0 | 0 |
| SUDDEN HEARING LOSS | 0 | 0 | 0 | 0 | 0 | 0 | 0 | 0 | 0 | 0 |
| *HYPERACUSIA* | | | | | | | | | | |
| HYPERACUSIS | 0 | 0 | 0 | 0 | 0 | 0 | 0 | 0 | 0 | 0 |
| ***INNER EAR AND VIIITH CRANIAL NERVE DISORDERS*** | | | | | | | | | | |
| *INNER EAR SIGNS AND SYMPTOMS* | | | | | | | | | | |
| MOTION SICKNESS | 0 | 0 | 0 | 0 | 0 | 0 | 0 | 0 | 0 | 0 |
| TINNITUS | 0 | 0 | 0 | 0 | 0 | 0 | 0 | 0 | 0 | 0 |
| VERTIGO | 1 | 0 | 0 | 1 | 0 | 1 | 0 | 0 | 1 | 0 |
| VERTIGO LABYRINTHINE | 0 | 0 | 0 | 0 | 0 | 0 | 0 | 0 | 0 | 0 |
| VERTIGO POSITIONAL | 0 | 0 | 0 | 0 | 0 | 0 | 0 | 0 | 0 | 0 |
| **ENDOCRINE DISORDERS** | | | | | | | | | | |
| ***THYROID GLAND DISORDERS*** | | | | | | | | | | |
| *THYROID HYPERFUNCTION DISORDERS* | | | | | | | | | | |
| HYPERTHYROIDISM | 0 | 0 | 0 | 0 | 0 | 0 | 0 | 0 | 0 | 0 |
| **EYE DISORDERS** | | | | | | | | | | |
| ***EYE DISORDERS NEC*** | | | | | | | | | | |
| *LACRIMATION DISORDERS* | | | | | | | | | | |
| DRY EYE | 0 | 0 | 0 | 0 | 0 | 0 | 0 | 0 | 0 | 0 |
| LACRIMATION INCREASED | 0 | 0 | 0 | 0 | 0 | 0 | 0 | 0 | 0 | 0 |
| *OCULAR DISORDERS NEC* | | | | | | | | | | |

| | | | | | | | | | | |
|---|---|---|---|---|---|---|---|---|---|---|
| EYE OEDEMA | 0 | 0 | 0 | 0 | 0 | 0 | 0 | 0 | 0 | 0 |
| EYE PAIN | 0 | 0 | 0 | 0 | 0 | 0 | 0 | 0 | 0 | 0 |
| EYE SWELLING | 0 | 0 | 0 | 0 | 0 | 0 | 0 | 0 | 0 | 0 |
| EYE ULCER | 0 | 0 | 0 | 0 | 0 | 0 | 0 | 0 | 0 | 0 |
| EYELID PAIN | 0 | 0 | 0 | 0 | 0 | 0 | 0 | 0 | 0 | 0 |
| OCULAR DISCOMFORT | 0 | 0 | 0 | 0 | 0 | 0 | 0 | 0 | 0 | 0 |
| PERIORBITAL DISCOMFORT | 0 | 0 | 0 | 0 | 0 | 0 | 0 | 0 | 0 | 0 |
| PERIORBITAL SWELLING | 0 | 0 | 0 | 0 | 0 | 0 | 0 | 0 | 0 | 0 |
| ***OCULAR HAEMORRHAGES AND VASCULAR DISORDERS NEC*** | | | | | | | | | | |
| *CONJUNCTIVAL AND CORNEAL BLEEDING AND VASCULAR DISORDERS* | | | | | | | | | | |
| CONJUNCTIVAL HAEMORRHAGE | 0 | 0 | 0 | 0 | 0 | 0 | 0 | 0 | 0 | 0 |
| *LID BLEEDING AND VASCULAR DISORDERS* | | | | | | | | | | |
| EYELID BLEEDING | 0 | 0 | 0 | 0 | 0 | 0 | 0 | 0 | 0 | 0 |
| ***OCULAR INFECTIONS, IRRITATIONS AND INFLAMMATIONS*** | | | | | | | | | | |
| *LID, LASH AND LACRIMAL INFECTIONS, IRRITATIONS AND INFLAMMATIONS* | | | | | | | | | | |
| BLEPHARITIS | 0 | 0 | 0 | 0 | 0 | 0 | 0 | 0 | 0 | 0 |
| ERYTHEMA OF EYELID | 0 | 0 | 0 | 0 | 0 | 0 | 0 | 0 | 0 | 0 |
| EYELID IRRITATION | 0 | 0 | 0 | 0 | 0 | 0 | 0 | 0 | 0 | 0 |
| *OCULAR INFECTIONS, INFLAMMATIONS AND ASSOCIATED MANIFESTATIONS* | | | | | | | | | | |
| EYE DISCHARGE | 0 | 0 | 0 | 0 | 0 | 0 | 0 | 0 | 0 | 0 |
| EYE IRRITATION | 0 | 0 | 0 | 0 | 0 | 0 | 0 | 0 | 0 | 0 |
| EYE PRURITUS | 0 | 0 | 0 | 0 | 0 | 0 | 0 | 0 | 0 | 0 |
| LIMBAL SWELLING | 0 | 0 | 0 | 0 | 0 | 0 | 0 | 0 | 0 | 0 |
| OCULAR HYPERAEMIA | 0 | 0 | 0 | 0 | 0 | 0 | 0 | 0 | 0 | 0 |
| ***OCULAR NEUROMUSCULAR DISORDERS*** | | | | | | | | | | |
| *EYELID MOVEMENT DISORDERS* | | | | | | | | | | |
| BLEPHAROSPASM | 0 | 0 | 0 | 0 | 0 | 0 | 0 | 0 | 0 | 0 |
| ***OCULAR SENSORY SYMPTOMS NEC*** | | | | | | | | | | |
| *OCULAR SENSATION DISORDERS* | | | | | | | | | | |
| ABNORMAL SENSATION IN EYE | 0 | 0 | 0 | 0 | 0 | 0 | 0 | 0 | 0 | 0 |
| ASTHENOPIA | 0 | 0 | 0 | 0 | 0 | 0 | 0 | 0 | 0 | 0 |
| FOREIGN BODY SENSATION IN EYES | 0 | 0 | 0 | 0 | 0 | 0 | 0 | 0 | 0 | 0 |
| PHOTOPHOBIA | 0 | 0 | 0 | 0 | 0 | 0 | 0 | 0 | 0 | 0 |

| | | | | | | | | | | |
|---|---|---|---|---|---|---|---|---|---|---|
| ***OCULAR STRUCTURAL CHANGE, DEPOSIT AND DEGENERATION NEC*** | | | | | | | | | | |
| *CHOROID AND VITREOUS STRUCTURAL CHANGE, DEPOSIT AND DEGENERATION* | | | | | | | | | | |
| VITREOUS DETACHMENT | 0 | 0 | 0 | 0 | 0 | 0 | 0 | 0 | 0 | 0 |
| VITREOUS FLOATERS | 0 | 0 | 0 | 0 | 0 | 0 | 0 | 0 | 0 | 0 |
| ***RETINA, CHOROID AND VITREOUS HAEMORRHAGES AND VASCULAR DISORDERS*** | | | | | | | | | | |
| *RETINAL BLEEDING AND VASCULAR DISORDERS (EXCL RETINOPATHY)* | | | | | | | | | | |
| RETINAL VEIN OCCLUSION | 0 | 0 | 0 | 0 | 0 | 0 | 0 | 0 | 0 | 0 |
| *RETINOPATHIES NEC* | | | | | | | | | | |
| RETINAL EXUDATES | 0 | 0 | 0 | 0 | 0 | 0 | 0 | 0 | 0 | 0 |
| ***VISION DISORDERS*** | | | | | | | | | | |
| *VISUAL DISORDERS NEC* | | | | | | | | | | |
| DIPLOPIA | 0 | 0 | 0 | 0 | 0 | 0 | 0 | 0 | 0 | 0 |
| HALO VISION | 0 | 0 | 0 | 0 | 0 | 0 | 0 | 0 | 0 | 0 |
| METAMORPHOPSIA | 0 | 0 | 0 | 0 | 0 | 0 | 0 | 0 | 0 | 0 |
| PHOTOPSIA | 0 | 0 | 0 | 0 | 0 | 0 | 0 | 0 | 0 | 0 |
| VISION BLURRED | 0 | 0 | 0 | 0 | 0 | 0 | 0 | 0 | 0 | 0 |
| *VISUAL IMPAIRMENT AND BLINDNESS (EXCL COLOUR BLINDNESS)* | | | | | | | | | | |
| BLINDNESS | 0 | 0 | 0 | 0 | 0 | 0 | 0 | 0 | 0 | 0 |
| BLINDNESS TRANSIENT | 0 | 0 | 0 | 0 | 0 | 0 | 0 | 0 | 0 | 0 |
| SUDDEN VISUAL LOSS | 0 | 0 | 0 | 0 | 0 | 0 | 0 | 0 | 0 | 0 |
| VISUAL IMPAIRMENT | 0 | 0 | 0 | 0 | 0 | 0 | 0 | 0 | 0 | 0 |
| **GASTROINTESTINAL DISORDERS** | | | | | | | | | | |
| ***ANAL AND RECTAL CONDITIONS NEC*** | | | | | | | | | | |
| *ANAL AND RECTAL SIGNS AND SYMPTOMS* | | | | | | | | | | |
| ANAL PARAESTHESIA | 0 | 0 | 0 | 0 | 0 | 0 | 0 | 0 | 0 | 0 |
| ***BENIGN NEOPLASMS GASTROINTESTINAL*** | | | | | | | | | | |
| *BENIGN ORAL CAVITY NEOPLASMS* | | | | | | | | | | |
| MOUTH CYST | 0 | 0 | 0 | 0 | 0 | 0 | 0 | 0 | 0 | 0 |
| ***DENTAL AND GINGIVAL CONDITIONS*** | | | | | | | | | | |
| *DENTAL DISORDERS NEC* | | | | | | | | | | |
| TEETHING | 0 | 0 | 0 | 0 | 0 | 0 | 0 | 0 | 0 | 0 |

| | | | | | | | | | | |
|---|---|---|---|---|---|---|---|---|---|---|
| *DENTAL PAIN AND SENSATION DISORDERS* | | | | | | | | | | |
| DENTAL PARAESTHESIA | 0 | 0 | 0 | 0 | 0 | 0 | 0 | 0 | 0 | 0 |
| TOOTHACHE | 0 | 0 | 0 | 0 | 0 | 0 | 0 | 0 | 0 | 0 |
| *GINGIVAL DISORDERS, SIGNS AND SYMPTOMS NEC* | | | | | | | | | | |
| GINGIVAL BLISTER | 0 | 0 | 0 | 0 | 0 | 0 | 0 | 0 | 0 | 0 |
| GINGIVAL PAIN | 0 | 0 | 0 | 0 | 0 | 0 | 0 | 0 | 0 | 0 |
| GINGIVAL SWELLING | 0 | 0 | 0 | 0 | 0 | 0 | 0 | 0 | 0 | 0 |
| ***GASTROINTESTINAL CONDITIONS NEC*** | | | | | | | | | | |
| *GASTROINTESTINAL MUCOSAL DYSTROPHIES AND SECRETION DISORDERS* | | | | | | | | | | |
| BARRETT'S OESOPHAGUS | 0 | 0 | 0 | 0 | 0 | 0 | 0 | 0 | 0 | 0 |
| ***GASTROINTESTINAL HAEMORRHAGES NEC*** | | | | | | | | | | |
| *INTESTINAL HAEMORRHAGES* | | | | | | | | | | |
| RECTAL HAEMORRHAGE | 0 | 0 | 0 | 0 | 0 | 0 | 0 | 0 | 0 | 0 |
| *NON-SITE SPECIFIC GASTROINTESTINAL HAEMORRHAGES* | | | | | | | | | | |
| HAEMATEMESIS | 0 | 0 | 0 | 0 | 0 | 0 | 0 | 0 | 0 | 0 |
| ***GASTROINTESTINAL INFLAMMATORY CONDITIONS*** | | | | | | | | | | |
| *COLITIS (EXCL INFECTIVE)* | | | | | | | | | | |
| COLITIS | 0 | 0 | 0 | 0 | 0 | 0 | 0 | 0 | 0 | 0 |
| COLITIS MICROSCOPIC | 0 | 0 | 0 | 0 | 0 | 0 | 0 | 0 | 0 | 0 |
| COLITIS ULCERATIVE | 0 | 0 | 0 | 0 | 0 | 0 | 0 | 0 | 0 | 0 |
| *GASTRITIS (EXCL INFECTIVE)* | | | | | | | | | | |
| GASTRITIS | 0 | 0 | 0 | 0 | 0 | 0 | 0 | 0 | 0 | 0 |
| REFLUX GASTRITIS | 0 | 0 | 0 | 0 | 0 | 0 | 0 | 0 | 0 | 0 |
| *GASTROINTESTINAL INFLAMMATORY DISORDERS NEC* | | | | | | | | | | |
| GASTROINTESTINAL TRACT IRRITATION | 0 | 0 | 0 | 0 | 0 | 0 | 0 | 0 | 0 | 0 |
| ***GASTROINTESTINAL MOTILITY AND DEFAECATION CONDITIONS*** | | | | | | | | | | |
| *DIARRHOEA (EXCL INFECTIVE)* | | | | | | | | | | |
| DIARRHOEA | 2 | 2 | 0 | 0 | 0 | 2 | 2 | 0 | 0 | 0 |
| *GASTROINTESTINAL ATONIC AND HYPOMOTILITY DISORDERS NEC* | | | | | | | | | | |
| CONSTIPATION | 1 | 1 | 0 | 0 | 0 | 1 | 1 | 0 | 0 | 0 |
| GASTROOESOPHAGEAL REFLUX DISEASE | 0 | 0 | 0 | 0 | 0 | 0 | 0 | 0 | 0 | 0 |
| *GASTROINTESTINAL DYSKINETIC DISORDERS* | | | | | | | | | | |
| CHANGE OF BOWEL HABIT | 0 | 0 | 0 | 0 | 0 | 0 | 0 | 0 | 0 | 0 |
| *GASTROINTESTINAL SPASTIC AND HYPERMOTILITY DISORDERS* | | | | | | | | | | |
| IRRITABLE BOWEL SYNDROME | 0 | 0 | 0 | 0 | 0 | 0 | 0 | 0 | 0 | 0 |
| ***GASTROINTESTINAL SIGNS AND SYMPTOMS*** | | | | | | | | | | |

| | | | | | | | | | | |
|---|---|---|---|---|---|---|---|---|---|---|
| *DYSPEPTIC SIGNS AND SYMPTOMS* | | | | | | | | | | |
| DYSPEPSIA | 1 | 1 | 0 | 0 | 0 | 1 | 1 | 0 | 0 | 0 |
| ERUCTATION | 0 | 0 | 0 | 0 | 0 | 0 | 0 | 0 | 0 | 0 |
| *FAECAL ABNORMALITIES NEC* | | | | | | | | | | |
| FAECALOMA | 0 | 0 | 0 | 0 | 0 | 0 | 0 | 0 | 0 | 0 |
| FAECES DISCOLOURED | 0 | 0 | 0 | 0 | 0 | 0 | 0 | 0 | 0 | 0 |
| FAECES SOFT | 0 | 0 | 0 | 0 | 0 | 0 | 0 | 0 | 0 | 0 |
| *FLATULENCE, BLOATING AND DISTENSION* | | | | | | | | | | |
| ABDOMINAL DISTENSION | 0 | 0 | 0 | 0 | 0 | 0 | 0 | 0 | 0 | 0 |
| FLATULENCE | 1 | 1 | 0 | 0 | 0 | 1 | 1 | 0 | 0 | 0 |
| *GASTROINTESTINAL AND ABDOMINAL PAINS (EXCL ORAL AND THROAT)* | | | | | | | | | | |
| ABDOMINAL PAIN | 2 | 2 | 0 | 0 | 0 | 2 | 2 | 0 | 0 | 0 |
| ABDOMINAL PAIN LOWER | 0 | 0 | 0 | 0 | 0 | 0 | 0 | 0 | 0 | 0 |
| ABDOMINAL PAIN UPPER | 0 | 0 | 0 | 0 | 0 | 0 | 0 | 0 | 0 | 0 |
| GASTROINTESTINAL PAIN | 0 | 0 | 0 | 0 | 0 | 0 | 0 | 0 | 0 | 0 |
| *GASTROINTESTINAL SIGNS AND SYMPTOMS NEC* | | | | | | | | | | |
| ABDOMINAL DISCOMFORT | 0 | 0 | 0 | 0 | 0 | 0 | 0 | 0 | 0 | 0 |
| ABDOMINAL SYMPTOM | 0 | 0 | 0 | 0 | 0 | 0 | 0 | 0 | 0 | 0 |
| ACUTE ABDOMEN | 0 | 0 | 0 | 0 | 0 | 0 | 0 | 0 | 0 | 0 |
| ANAL INCONTINENCE | 0 | 0 | 0 | 0 | 0 | 0 | 0 | 0 | 0 | 0 |
| BREATH ODOUR | 0 | 0 | 0 | 0 | 0 | 0 | 0 | 0 | 0 | 0 |
| DYSPHAGIA | 0 | 0 | 0 | 0 | 0 | 0 | 0 | 0 | 0 | 0 |
| ODYNOPHAGIA | 0 | 0 | 0 | 0 | 0 | 0 | 0 | 0 | 0 | 0 |
| *NAUSEA AND VOMITING SYMPTOMS* | | | | | | | | | | |
| NAUSEA | 9 | 7 | 0 | 2 | 0 | 9 | 7 | 0 | 2 | 0 |
| RETCHING | 0 | 0 | 0 | 0 | 0 | 0 | 0 | 0 | 0 | 0 |
| VOMITING | 2 | 1 | 0 | 0 | 1 | 2 | 1 | 0 | 0 | 1 |
| VOMITING PROJECTILE | 0 | 0 | 0 | 0 | 0 | 0 | 0 | 0 | 0 | 0 |
| ***MALABSORPTION CONDITIONS*** | | | | | | | | | | |
| *MALABSORPTION SYNDROMES* | | | | | | | | | | |
| COELIAC DISEASE | 0 | 0 | 0 | 0 | 0 | 0 | 0 | 0 | 0 | 0 |
| ***ORAL SOFT TISSUE CONDITIONS*** | | | | | | | | | | |
| *ORAL SOFT TISSUE DISORDERS NEC* | | | | | | | | | | |
| CHEILITIS | 0 | 0 | 0 | 0 | 0 | 0 | 0 | 0 | 0 | 0 |
| LIP BLISTER | 0 | 0 | 0 | 0 | 0 | 0 | 0 | 0 | 0 | 0 |

| | | | | | | | | | | |
|---|---|---|---|---|---|---|---|---|---|---|
| ORAL LICHEN PLANUS | 0 | 0 | 0 | 0 | 0 | 0 | 0 | 0 | 0 | 0 |
| *ORAL SOFT TISSUE INFECTIONS* | | | | | | | | | | |
| ANGULAR CHEILITIS | 0 | 0 | 0 | 0 | 0 | 0 | 0 | 0 | 0 | 0 |
| *ORAL SOFT TISSUE SIGNS AND SYMPTOMS* | | | | | | | | | | |
| HYPOAESTHESIA ORAL | 1 | 1 | 0 | 0 | 0 | 1 | 1 | 0 | 0 | 0 |
| LIP PAIN | 0 | 0 | 0 | 0 | 0 | 0 | 0 | 0 | 0 | 0 |
| ORAL DISCOMFORT | 0 | 0 | 0 | 0 | 0 | 0 | 0 | 0 | 0 | 0 |
| ORAL MUCOSAL ROUGHENING | 0 | 0 | 0 | 0 | 0 | 0 | 0 | 0 | 0 | 0 |
| ORAL PAIN | 0 | 0 | 0 | 0 | 0 | 0 | 0 | 0 | 0 | 0 |
| PARAESTHESIA ORAL | 0 | 0 | 0 | 0 | 0 | 0 | 0 | 0 | 0 | 0 |
| LIP SWELLING | 0 | 0 | 0 | 0 | 0 | 0 | 0 | 0 | 0 | 0 |
| MOUTH SWELLING | 0 | 0 | 0 | 0 | 0 | 0 | 0 | 0 | 0 | 0 |
| *STOMATITIS AND ULCERATION* | | | | | | | | | | |
| APHTHOUS ULCER | 0 | 0 | 0 | 0 | 0 | 0 | 0 | 0 | 0 | 0 |
| LIP ULCERATION | 0 | 0 | 0 | 0 | 0 | 0 | 0 | 0 | 0 | 0 |
| MOUTH ULCERATION | 0 | 0 | 0 | 0 | 0 | 0 | 0 | 0 | 0 | 0 |
| STOMATITIS | 0 | 0 | 0 | 0 | 0 | 0 | 0 | 0 | 0 | 0 |
| ***SALIVARY GLAND CONDITIONS*** | | | | | | | | | | |
| *ORAL DRYNESS AND SALIVA ALTERED* | | | | | | | | | | |
| DRY MOUTH | 0 | 0 | 0 | 0 | 0 | 0 | 0 | 0 | 0 | 0 |
| LIP DRY | 0 | 0 | 0 | 0 | 0 | 0 | 0 | 0 | 0 | 0 |
| SALIVARY HYPOSECRETION | 0 | 0 | 0 | 0 | 0 | 0 | 0 | 0 | 0 | 0 |
| ***TONGUE CONDITIONS*** | | | | | | | | | | |
| *TONGUE SIGNS AND SYMPTOMS* | | | | | | | | | | |
| GLOSSODYNIA | 0 | 0 | 0 | 0 | 0 | 0 | 0 | 0 | 0 | 0 |
| SWOLLEN TONGUE | 0 | 0 | 0 | 0 | 0 | 0 | 0 | 0 | 0 | 0 |
| TONGUE COATED | 0 | 0 | 0 | 0 | 0 | 0 | 0 | 0 | 0 | 0 |
| TONGUE DISCOMFORT | 0 | 0 | 0 | 0 | 0 | 0 | 0 | 0 | 0 | 0 |
| TONGUE OEDEMA | 0 | 0 | 0 | 0 | 0 | 0 | 0 | 0 | 0 | 0 |
| TONGUE SPASM | 0 | 0 | 0 | 0 | 0 | 0 | 0 | 0 | 0 | 0 |
| **GENERAL DISORDERS AND ADMINISTRATION SITE CONDITIONS** | | | | | | | | | | |
| ***ADMINISTRATION SITE REACTIONS*** | | | | | | | | | | |
| *ADMINISTRATION SITE REACTIONS NEC* | | | | | | | | | | |
| ADMINISTRATION SITE BRUISE | 0 | 0 | 0 | 0 | 0 | 0 | 0 | 0 | 0 | 0 |
| ADMINISTRATION SITE PAIN | 0 | 0 | 0 | 0 | 0 | 0 | 0 | 0 | 0 | 0 |

| | | | | | | | | | | |
|---|---|---|---|---|---|---|---|---|---|---|
| PUNCTURE SITE BRUISE | 0 | 0 | 0 | 0 | 0 | 0 | 0 | 0 | 0 | 0 |
| PUNCTURE SITE PAIN | 0 | 0 | 0 | 0 | 0 | 0 | 0 | 0 | 0 | 0 |
| *APPLICATION SITE REACTIONS* | | | | | | | | | | |
| APPLICATION SITE BRUISE | 0 | 0 | 0 | 0 | 0 | 0 | 0 | 0 | 0 | 0 |
| APPLICATION SITE ERYTHEMA | 0 | 0 | 0 | 0 | 0 | 0 | 0 | 0 | 0 | 0 |
| APPLICATION SITE PAIN | 0 | 0 | 0 | 0 | 0 | 0 | 0 | 0 | 0 | 0 |
| *IMPLANT AND CATHETER SITE REACTIONS* | | | | | | | | | | |
| IMPLANT SITE PAIN | 0 | 0 | 0 | 0 | 0 | 0 | 0 | 0 | 0 | 0 |
| IMPLANT SITE WARMTH | 0 | 0 | 0 | 0 | 0 | 0 | 0 | 0 | 0 | 0 |
| *INFUSION SITE REACTIONS* | | | | | | | | | | |
| INFUSION SITE PAIN | 0 | 0 | 0 | 0 | 0 | 0 | 0 | 0 | 0 | 0 |
| INFUSION SITE SCAB | 0 | 0 | 0 | 0 | 0 | 0 | 0 | 0 | 0 | 0 |
| INFUSION SITE WARMTH | 0 | 0 | 0 | 0 | 0 | 0 | 0 | 0 | 0 | 0 |
| *INJECTION SITE REACTIONS* | | | | | | | | | | |
| INJECTION SITE BRUISING | 1 | 1 | 0 | 0 | 0 | 1 | 1 | 0 | 0 | 0 |
| INJECTION SITE DISCOMFORT | 0 | 0 | 0 | 0 | 0 | 0 | 0 | 0 | 0 | 0 |
| INJECTION SITE ERYTHEMA | 0 | 0 | 0 | 0 | 0 | 0 | 0 | 0 | 0 | 0 |
| INJECTION SITE HYPERSENSITIVITY | 0 | 0 | 0 | 0 | 0 | 0 | 0 | 0 | 0 | 0 |
| INJECTION SITE INFLAMMATION | 0 | 0 | 0 | 0 | 0 | 0 | 0 | 0 | 0 | 0 |
| INJECTION SITE INJURY | 0 | 0 | 0 | 0 | 0 | 0 | 0 | 0 | 0 | 0 |
| INJECTION SITE IRRITATION | 0 | 0 | 0 | 0 | 0 | 0 | 0 | 0 | 0 | 0 |
| INJECTION SITE JOINT PAIN | 0 | 0 | 0 | 0 | 0 | 0 | 0 | 0 | 0 | 0 |
| INJECTION SITE MASS | 0 | 0 | 0 | 0 | 0 | 0 | 0 | 0 | 0 | 0 |
| INJECTION SITE NODULE | 0 | 0 | 0 | 0 | 0 | 0 | 0 | 0 | 0 | 0 |
| INJECTION SITE OEDEMA | 0 | 0 | 0 | 0 | 0 | 0 | 0 | 0 | 0 | 0 |
| INJECTION SITE PAIN | 3 | 3 | 0 | 0 | 0 | 3 | 3 | 0 | 0 | 0 |
| INJECTION SITE PAPULE | 0 | 0 | 0 | 0 | 0 | 0 | 0 | 0 | 0 | 0 |
| INJECTION SITE PARAESTHESIA | 0 | 0 | 0 | 0 | 0 | 0 | 0 | 0 | 0 | 0 |
| INJECTION SITE PRURITUS | 1 | 1 | 0 | 0 | 0 | 1 | 1 | 0 | 0 | 0 |
| INJECTION SITE RASH | 0 | 0 | 0 | 0 | 0 | 0 | 0 | 0 | 0 | 0 |
| INJECTION SITE REACTION | 0 | 0 | 0 | 0 | 0 | 0 | 0 | 0 | 0 | 0 |
| INJECTION SITE SCAB | 0 | 0 | 0 | 0 | 0 | 0 | 0 | 0 | 0 | 0 |
| INJECTION SITE SWELLING | 0 | 0 | 0 | 0 | 0 | 0 | 0 | 0 | 0 | 0 |
| INJECTION SITE URTICARIA | 0 | 0 | 0 | 0 | 0 | 0 | 0 | 0 | 0 | 0 |
| INJECTION SITE WARMTH | 0 | 0 | 0 | 0 | 0 | 0 | 0 | 0 | 0 | 0 |

| | | | | | | | | | | |
|---|---|---|---|---|---|---|---|---|---|---|
| *INSTILLATION SITE REACTIONS* | | | | | | | | | | |
| INSTILLATION SITE PRURITUS | 0 | 0 | 0 | 0 | 0 | 0 | 0 | 0 | 0 | 0 |
| INSTILLATION SITE WARMTH | 0 | 0 | 0 | 0 | 0 | 0 | 0 | 0 | 0 | 0 |
| *VACCINATION SITE REACTIONS* | | | | | | | | | | |
| SHOULDER INJURY RELATED TO VACCINE ADMINISTRATION | 0 | 0 | 0 | 0 | 0 | 0 | 0 | 0 | 0 | 0 |
| VACCINATION SITE BRUISING | 0 | 0 | 0 | 0 | 0 | 0 | 0 | 0 | 0 | 0 |
| VACCINATION SITE DISCOMFORT | 0 | 0 | 0 | 0 | 0 | 0 | 0 | 0 | 0 | 0 |
| VACCINATION SITE ERYTHEMA | 0 | 0 | 0 | 0 | 0 | 0 | 0 | 0 | 0 | 0 |
| VACCINATION SITE JOINT ERYTHEMA | 0 | 0 | 0 | 0 | 0 | 0 | 0 | 0 | 0 | 0 |
| VACCINATION SITE JOINT PAIN | 0 | 0 | 0 | 0 | 0 | 0 | 0 | 0 | 0 | 0 |
| VACCINATION SITE MASS | 0 | 0 | 0 | 0 | 0 | 0 | 0 | 0 | 0 | 0 |
| VACCINATION SITE PAIN | 0 | 0 | 0 | 0 | 0 | 0 | 0 | 0 | 0 | 0 |
| VACCINATION SITE RASH | 0 | 0 | 0 | 0 | 0 | 0 | 0 | 0 | 0 | 0 |
| VACCINATION SITE SWELLING | 0 | 0 | 0 | 0 | 0 | 0 | 0 | 0 | 0 | 0 |
| VACCINATION SITE WARMTH | 0 | 0 | 0 | 0 | 0 | 0 | 0 | 0 | 0 | 0 |
| ***BODY TEMPERATURE CONDITIONS*** | | | | | | | | | | |
| *BODY TEMPERATURE ALTERED* | | | | | | | | | | |
| HYPERTHERMIA | 0 | 0 | 0 | 0 | 0 | 0 | 0 | 0 | 0 | 0 |
| HYPOTHERMIA | 0 | 0 | 0 | 0 | 0 | 0 | 0 | 0 | 0 | 0 |
| *FEBRILE DISORDERS* | | | | | | | | | | |
| PYREXIA | 16 | 12 | 3 | 1 | 0 | 13 | 12 | 0 | 1 | 0 |
| ***FATAL OUTCOMES*** | | | | | | | | | | |
| *DEATH AND SUDDEN DEATH* | | | | | | | | | | |
| DEATH | 0 | 0 | 0 | 0 | 0 | 0 | 0 | 0 | 0 | 0 |
| ***GENERAL SYSTEM DISORDERS NEC*** | | | | | | | | | | |
| *ADVERSE EFFECT ABSENT* | | | | | | | | | | |
| NO ADVERSE EVENT | 0 | 0 | 0 | 0 | 0 | 0 | 0 | 0 | 0 | 0 |
| *ASTHENIC CONDITIONS* | | | | | | | | | | |
| ASTHENIA | 3 | 3 | 0 | 0 | 0 | 3 | 3 | 0 | 0 | 0 |
| CHRONIC FATIGUE SYNDROME | 0 | 0 | 0 | 0 | 0 | 0 | 0 | 0 | 0 | 0 |
| DECREASED ACTIVITY | 0 | 0 | 0 | 0 | 0 | 0 | 0 | 0 | 0 | 0 |
| FATIGUE | 42 | 24 | 7 | 9 | 2 | 38 | 24 | 5 | 9 | 0 |
| MALAISE | 4 | 4 | 0 | 0 | 0 | 4 | 4 | 0 | 0 | 0 |
| SLUGGISHNESS | 0 | 0 | 0 | 0 | 0 | 0 | 0 | 0 | 0 | 0 |
| *FEELINGS AND SENSATIONS NEC* | | | | | | | | | | |

| | | | | | | | | | | |
|---|---|---|---|---|---|---|---|---|---|---|
| CHILLS | 10 | 8 | 0 | 2 | 0 | 10 | 8 | 0 | 2 | 0 |
| FEELING ABNORMAL | 1 | 1 | 0 | 0 | 0 | 1 | 1 | 0 | 0 | 0 |
| FEELING COLD | 4 | 2 | 1 | 1 | 0 | 4 | 2 | 1 | 1 | 0 |
| FEELING HOT | 2 | 1 | 0 | 1 | 0 | 2 | 1 | 0 | 1 | 0 |
| FEELING JITTERY | 0 | 0 | 0 | 0 | 0 | 0 | 0 | 0 | 0 | 0 |
| FEELING OF BODY TEMPERATURE CHANGE | 0 | 0 | 0 | 0 | 0 | 0 | 0 | 0 | 0 | 0 |
| FEELING OF RELAXATION | 0 | 0 | 0 | 0 | 0 | 0 | 0 | 0 | 0 | 0 |
| HANGOVER | 0 | 0 | 0 | 0 | 0 | 0 | 0 | 0 | 0 | 0 |
| HUNGER | 0 | 0 | 0 | 0 | 0 | 0 | 0 | 0 | 0 | 0 |
| SENSATION OF BLOOD FLOW | 0 | 0 | 0 | 0 | 0 | 0 | 0 | 0 | 0 | 0 |
| THIRST | 0 | 0 | 0 | 0 | 0 | 0 | 0 | 0 | 0 | 0 |
| *GAIT DISTURBANCES* | | | | | | | | | | |
| GAIT DISTURBANCE | 1 | 0 | 1 | 0 | 0 | 1 | 0 | 1 | 0 | 0 |
| GAIT INABILITY | 0 | 0 | 0 | 0 | 0 | 0 | 0 | 0 | 0 | 0 |
| *GENERAL SIGNS AND SYMPTOMS NEC* | | | | | | | | | | |
| CRYING | 0 | 0 | 0 | 0 | 0 | 0 | 0 | 0 | 0 | 0 |
| ENERGY INCREASED | 0 | 0 | 0 | 0 | 0 | 0 | 0 | 0 | 0 | 0 |
| EXERCISE TOLERANCE DECREASED | 0 | 0 | 0 | 0 | 0 | 0 | 0 | 0 | 0 | 0 |
| GENERAL SYMPTOM | 0 | 0 | 0 | 0 | 0 | 0 | 0 | 0 | 0 | 0 |
| ILLNESS | 2 | 1 | 0 | 1 | 0 | 1 | 1 | 0 | 0 | 0 |
| INFLUENZA LIKE ILLNESS | 8 | 6 | 0 | 2 | 0 | 8 | 6 | 0 | 2 | 0 |
| LOCAL REACTION | 0 | 0 | 0 | 0 | 0 | 0 | 0 | 0 | 0 | 0 |
| PERIPHERAL SWELLING | 1 | 1 | 0 | 0 | 0 | 1 | 1 | 0 | 0 | 0 |
| SWELLING | 2 | 1 | 0 | 1 | 0 | 2 | 1 | 0 | 1 | 0 |
| SWELLING FACE | 0 | 0 | 0 | 0 | 0 | 0 | 0 | 0 | 0 | 0 |
| TISSUE IRRITATION | 0 | 0 | 0 | 0 | 0 | 0 | 0 | 0 | 0 | 0 |
| *INFLAMMATIONS* | | | | | | | | | | |
| INFLAMMATION | 1 | 0 | 1 | 0 | 0 | 1 | 0 | 1 | 0 | 0 |
| SYSTEMIC INFLAMMATORY RESPONSE SYNDROME | 0 | 0 | 0 | 0 | 0 | 0 | 0 | 0 | 0 | 0 |
| *OEDEMA NEC* | | | | | | | | | | |
| OEDEMA | 0 | 0 | 0 | 0 | 0 | 0 | 0 | 0 | 0 | 0 |
| OEDEMA PERIPHERAL | 0 | 0 | 0 | 0 | 0 | 0 | 0 | 0 | 0 | 0 |
| *PAIN AND DISCOMFORT NEC* | | | | | | | | | | |
| AXILLARY PAIN | 0 | 0 | 0 | 0 | 0 | 0 | 0 | 0 | 0 | 0 |

| | | | | | | | | | | |
|---|---|---|---|---|---|---|---|---|---|---|
| CHEST DISCOMFORT | 0 | 0 | 0 | 0 | 0 | 0 | 0 | 0 | 0 | 0 |
| CHEST PAIN | 1 | 0 | 1 | 0 | 0 | 1 | 0 | 1 | 0 | 0 |
| DISCOMFORT | 0 | 0 | 0 | 0 | 0 | 0 | 0 | 0 | 0 | 0 |
| FACIAL PAIN | 0 | 0 | 0 | 0 | 0 | 0 | 0 | 0 | 0 | 0 |
| HERNIA PAIN | 0 | 0 | 0 | 0 | 0 | 0 | 0 | 0 | 0 | 0 |
| INFLAMMATORY PAIN | 0 | 0 | 0 | 0 | 0 | 0 | 0 | 0 | 0 | 0 |
| NON-CARDIAC CHEST PAIN | 0 | 0 | 0 | 0 | 0 | 0 | 0 | 0 | 0 | 0 |
| PAIN | 10 | 5 | 2 | 3 | 0 | 9 | 5 | 1 | 3 | 0 |
| TENDERNESS | 4 | 3 | 0 | 0 | 1 | 4 | 3 | 0 | 0 | 1 |
| ***THERAPEUTIC AND NONTHERAPEUTIC EFFECTS (EXCL TOXICITY)*** | | | | | | | | | | |
| *THERAPEUTIC AND NONTHERAPEUTIC RESPONSES* | | | | | | | | | | |
| ADVERSE DRUG REACTION | 2 | 2 | 0 | 0 | 0 | 2 | 2 | 0 | 0 | 0 |
| ADVERSE EVENT | 0 | 0 | 0 | 0 | 0 | 0 | 0 | 0 | 0 | 0 |
| ADVERSE REACTION | 0 | 0 | 0 | 0 | 0 | 0 | 0 | 0 | 0 | 0 |
| IMMEDIATE POST-INJECTION REACTION | 0 | 0 | 0 | 0 | 0 | 0 | 0 | 0 | 0 | 0 |
| **HEPATOBILIARY DISORDERS** | | | | | | | | | | |
| ***HEPATIC AND HEPATOBILIARY DISORDERS*** | | | | | | | | | | |
| *HEPATOBILIARY SIGNS AND SYMPTOMS* | | | | | | | | | | |
| HEPATIC PAIN | 0 | 0 | 0 | 0 | 0 | 0 | 0 | 0 | 0 | 0 |
| *HEPATOCELLULAR DAMAGE AND HEPATITIS NEC* | | | | | | | | | | |
| LIVER INJURY | 0 | 0 | 0 | 0 | 0 | 0 | 0 | 0 | 0 | 0 |
| **IMMUNE SYSTEM DISORDERS** | | | | | | | | | | |
| ***ALLERGIC CONDITIONS*** | | | | | | | | | | |
| *ALLERGIC CONDITIONS NEC* | | | | | | | | | | |
| HYPERSENSITIVITY | 0 | 0 | 0 | 0 | 0 | 0 | 0 | 0 | 0 | 0 |
| MULTIPLE ALLERGIES | 0 | 0 | 0 | 0 | 0 | 0 | 0 | 0 | 0 | 0 |
| *ALLERGIES TO FOODS, FOOD ADDITIVES, DRUGS AND OTHER CHEMICALS* | | | | | | | | | | |
| ALLERGY TO CHEMICALS | 0 | 0 | 0 | 0 | 0 | 0 | 0 | 0 | 0 | 0 |
| *ANAPHYLACTIC AND ANAPHYLACTOID RESPONSES* | | | | | | | | | | |
| ANAPHYLACTIC REACTION | 0 | 0 | 0 | 0 | 0 | 0 | 0 | 0 | 0 | 0 |
| *ATOPIC DISORDERS* | | | | | | | | | | |
| SEASONAL ALLERGY | 0 | 0 | 0 | 0 | 0 | 0 | 0 | 0 | 0 | 0 |
| ***IMMUNE DISORDERS NEC*** | | | | | | | | | | |
| *IMMUNE AND ASSOCIATED CONDITIONS NEC* | | | | | | | | | | |
| BACILLE CALMETTE-GUERIN SCAR REACTIVATION | 0 | 0 | 0 | 0 | 0 | 0 | 0 | 0 | 0 | 0 |

| | | | | | | | | | | |
|---|---|---|---|---|---|---|---|---|---|---|
| SENSITISATION | 0 | 0 | 0 | 0 | 0 | 0 | 0 | 0 | 0 | 0 |
| **INFECTIONS AND INFESTATIONS** | | | | | | | | | | |
| ***BACTERIAL INFECTIOUS DISORDERS*** | | | | | | | | | | |
| *BACTERIAL INFECTIONS NEC* | | | | | | | | | | |
| CELLULITIS | 0 | 0 | 0 | 0 | 0 | 0 | 0 | 0 | 0 | 0 |
| PERIORBITAL CELLULITIS | 0 | 0 | 0 | 0 | 0 | 0 | 0 | 0 | 0 | 0 |
| *HELICOBACTER INFECTIONS* | | | | | | | | | | |
| HELICOBACTER GASTRITIS | 0 | 0 | 0 | 0 | 0 | 0 | 0 | 0 | 0 | 0 |
| *STAPHYLOCOCCAL INFECTIONS* | | | | | | | | | | |
| FURUNCLE | 0 | 0 | 0 | 0 | 0 | 0 | 0 | 0 | 0 | 0 |
| ***FUNGAL INFECTIOUS DISORDERS*** | | | | | | | | | | |
| *CANDIDA INFECTIONS* | | | | | | | | | | |
| CANDIDA INFECTION | 0 | 0 | 0 | 0 | 0 | 0 | 0 | 0 | 0 | 0 |
| VULVOVAGINAL CANDIDIASIS | 0 | 0 | 0 | 0 | 0 | 0 | 0 | 0 | 0 | 0 |
| ***INFECTIONS - PATHOGEN UNSPECIFIED*** | | | | | | | | | | |
| *ABDOMINAL AND GASTROINTESTINAL INFECTIONS* | | | | | | | | | | |
| APPENDICITIS | 0 | 0 | 0 | 0 | 0 | 0 | 0 | 0 | 0 | 0 |
| DIARRHOEA INFECTIOUS | 0 | 0 | 0 | 0 | 0 | 0 | 0 | 0 | 0 | 0 |
| GASTROINTESTINAL INFECTION | 1 | 1 | 0 | 0 | 0 | 1 | 1 | 0 | 0 | 0 |
| *BREAST INFECTIONS* | | | | | | | | | | |
| MASTITIS | 0 | 0 | 0 | 0 | 0 | 0 | 0 | 0 | 0 | 0 |
| *CENTRAL NERVOUS SYSTEM AND SPINAL INFECTIONS* | | | | | | | | | | |
| MYELITIS | 0 | 0 | 0 | 0 | 0 | 0 | 0 | 0 | 0 | 0 |
| *DENTAL AND ORAL SOFT TISSUE INFECTIONS* | | | | | | | | | | |
| ABSCESS ORAL | 0 | 0 | 0 | 0 | 0 | 0 | 0 | 0 | 0 | 0 |
| ORAL PUSTULE | 0 | 0 | 0 | 0 | 0 | 0 | 0 | 0 | 0 | 0 |
| PERICORONITIS | 0 | 0 | 0 | 0 | 0 | 0 | 0 | 0 | 0 | 0 |
| TOOTH ABSCESS | 0 | 0 | 0 | 0 | 0 | 0 | 0 | 0 | 0 | 0 |
| *EAR INFECTIONS* | | | | | | | | | | |
| EAR INFECTION | 0 | 0 | 0 | 0 | 0 | 0 | 0 | 0 | 0 | 0 |
| LABYRINTHITIS | 0 | 0 | 0 | 0 | 0 | 0 | 0 | 0 | 0 | 0 |
| OTITIS EXTERNA | 0 | 0 | 0 | 0 | 0 | 0 | 0 | 0 | 0 | 0 |
| *EYE AND EYELID INFECTIONS* | | | | | | | | | | |
| CONJUNCTIVITIS | 0 | 0 | 0 | 0 | 0 | 0 | 0 | 0 | 0 | 0 |
| HORDEOLUM | 0 | 0 | 0 | 0 | 0 | 0 | 0 | 0 | 0 | 0 |
| *INFECTIONS NEC* | | | | | | | | | | |

| | | | | | | | | | | |
|---|---|---|---|---|---|---|---|---|---|---|
| ABSCESS | 0 | 0 | 0 | 0 | 0 | 0 | 0 | 0 | 0 | 0 |
| INFECTION | 0 | 0 | 0 | 0 | 0 | 0 | 0 | 0 | 0 | 0 |
| INJECTION SITE INFECTION | 0 | 0 | 0 | 0 | 0 | 0 | 0 | 0 | 0 | 0 |
| LOCALISED INFECTION | 0 | 0 | 0 | 0 | 0 | 0 | 0 | 0 | 0 | 0 |
| WOUND INFECTION | 0 | 0 | 0 | 0 | 0 | 0 | 0 | 0 | 0 | 0 |
| *LOWER RESPIRATORY TRACT AND LUNG INFECTIONS* | | | | | | | | | | |
| LOWER RESPIRATORY TRACT INFECTION | 0 | 0 | 0 | 0 | 0 | 0 | 0 | 0 | 0 | 0 |
| PNEUMONIA | 0 | 0 | 0 | 0 | 0 | 0 | 0 | 0 | 0 | 0 |
| *MALE REPRODUCTIVE TRACT INFECTIONS* | | | | | | | | | | |
| ORCHITIS | 0 | 0 | 0 | 0 | 0 | 0 | 0 | 0 | 0 | 0 |
| *SEPSIS, BACTERAEMIA, VIRAEMIA AND FUNGAEMIA NEC* | | | | | | | | | | |
| SEPTIC RASH | 0 | 0 | 0 | 0 | 0 | 0 | 0 | 0 | 0 | 0 |
| *SKIN STRUCTURES AND SOFT TISSUE INFECTIONS* | | | | | | | | | | |
| INFECTED DERMAL CYST | 0 | 0 | 0 | 0 | 0 | 0 | 0 | 0 | 0 | 0 |
| INJECTION SITE PUSTULE | 0 | 0 | 0 | 0 | 0 | 0 | 0 | 0 | 0 | 0 |
| SKIN INFECTION | 0 | 0 | 0 | 0 | 0 | 0 | 0 | 0 | 0 | 0 |
| *UPPER RESPIRATORY TRACT INFECTIONS* | | | | | | | | | | |
| LARYNGITIS | 0 | 0 | 0 | 0 | 0 | 0 | 0 | 0 | 0 | 0 |
| NASOPHARYNGITIS | 3 | 3 | 0 | 0 | 0 | 3 | 3 | 0 | 0 | 0 |
| PHARYNGITIS | 0 | 0 | 0 | 0 | 0 | 0 | 0 | 0 | 0 | 0 |
| RHINITIS | 0 | 0 | 0 | 0 | 0 | 0 | 0 | 0 | 0 | 0 |
| SINUSITIS | 0 | 0 | 0 | 0 | 0 | 0 | 0 | 0 | 0 | 0 |
| TONSILLITIS | 0 | 0 | 0 | 0 | 0 | 0 | 0 | 0 | 0 | 0 |
| *URINARY TRACT INFECTIONS* | | | | | | | | | | |
| CYSTITIS | 0 | 0 | 0 | 0 | 0 | 0 | 0 | 0 | 0 | 0 |
| URINARY TRACT INFECTION | 0 | 0 | 0 | 0 | 0 | 0 | 0 | 0 | 0 | 0 |
| ***PROTOZOAL INFECTIOUS DISORDERS*** | | | | | | | | | | |
| *TRYPANOSOMAL INFECTIONS* | | | | | | | | | | |
| AFRICAN TRYPANOSOMIASIS | 0 | 0 | 0 | 0 | 0 | 0 | 0 | 0 | 0 | 0 |
| ***RICKETTSIAL INFECTIOUS DISORDERS*** | | | | | | | | | | |
| *COXIELLA INFECTIONS* | | | | | | | | | | |
| Q FEVER | 0 | 0 | 0 | 0 | 0 | 0 | 0 | 0 | 0 | 0 |
| ***VIRAL INFECTIOUS DISORDERS*** | | | | | | | | | | |
| *CORONAVIRUS INFECTIONS* | | | | | | | | | | |
| COVID-19 | 0 | 0 | 0 | 0 | 0 | 0 | 0 | 0 | 0 | 0 |
| *HERPES VIRAL INFECTIONS* | | | | | | | | | | |

| | | | | | | | | | | |
|---|---|---|---|---|---|---|---|---|---|---|
| GENITAL HERPES | 0 | 0 | 0 | 0 | 0 | 0 | 0 | 0 | 0 | 0 |
| HERPES OPHTHALMIC | 0 | 0 | 0 | 0 | 0 | 0 | 0 | 0 | 0 | 0 |
| HERPES SIMPLEX | 0 | 0 | 0 | 0 | 0 | 0 | 0 | 0 | 0 | 0 |
| HERPES ZOSTER | 0 | 0 | 0 | 0 | 0 | 0 | 0 | 0 | 0 | 0 |
| NASAL HERPES | 0 | 0 | 0 | 0 | 0 | 0 | 0 | 0 | 0 | 0 |
| OPHTHALMIC HERPES ZOSTER | 0 | 0 | 0 | 0 | 0 | 0 | 0 | 0 | 0 | 0 |
| ORAL HERPES | 0 | 0 | 0 | 0 | 0 | 0 | 0 | 0 | 0 | 0 |
| *INFLUENZA VIRAL INFECTIONS* | | | | | | | | | | |
| H2N2 INFLUENZA | 0 | 0 | 0 | 0 | 0 | 0 | 0 | 0 | 0 | 0 |
| INFLUENZA | 7 | 5 | 0 | 2 | 0 | 7 | 5 | 0 | 2 | 0 |
| *RETROVIRAL INFECTIONS* | | | | | | | | | | |
| AIDS RELATED COMPLEX | 0 | 0 | 0 | 0 | 0 | 0 | 0 | 0 | 0 | 0 |
| *VIRAL INFECTIONS NEC* | | | | | | | | | | |
| GASTROENTERITIS VIRAL | 0 | 0 | 0 | 0 | 0 | 0 | 0 | 0 | 0 | 0 |
| SWEATING FEVER | 0 | 0 | 0 | 0 | 0 | 0 | 0 | 0 | 0 | 0 |
| VESTIBULAR NEURONITIS | 0 | 0 | 0 | 0 | 0 | 0 | 0 | 0 | 0 | 0 |
| VIRAL DIARRHOEA | 0 | 0 | 0 | 0 | 0 | 0 | 0 | 0 | 0 | 0 |
| VIRAL RASH | 0 | 0 | 0 | 0 | 0 | 0 | 0 | 0 | 0 | 0 |
| VIRAL UPPER RESPIRATORY TRACT INFECTION | 0 | 0 | 0 | 0 | 0 | 0 | 0 | 0 | 0 | 0 |
| **INJURY, POISONING AND PROCEDURAL COMPLICATIONS** | | | | | | | | | | |
| ***BONE AND JOINT INJURIES*** | | | | | | | | | | |
| *FRACTURES AND DISLOCATIONS NEC* | | | | | | | | | | |
| JOINT DISLOCATION | 0 | 0 | 0 | 0 | 0 | 0 | 0 | 0 | 0 | 0 |
| ***EXPOSURES, CHEMICAL INJURIES AND POISONING*** | | | | | | | | | | |
| *POISONING AND TOXICITY* | | | | | | | | | | |
| SYSTEMIC TOXICITY | 0 | 0 | 0 | 0 | 0 | 0 | 0 | 0 | 0 | 0 |
| TOXICITY TO VARIOUS AGENTS | 0 | 0 | 0 | 0 | 0 | 0 | 0 | 0 | 0 | 0 |
| ***INJURIES BY PHYSICAL AGENTS*** | | | | | | | | | | |
| *CONDITIONS CAUSED BY COLD* | | | | | | | | | | |
| CHILLBLAINS | 0 | 0 | 0 | 0 | 0 | 0 | 0 | 0 | 0 | 0 |
| *HEAT INJURIES (EXCL THERMAL BURNS)* | | | | | | | | | | |
| HEAT EXHAUSTION | 0 | 0 | 0 | 0 | 0 | 0 | 0 | 0 | 0 | 0 |
| HEAT OEDEMA | 0 | 0 | 0 | 0 | 0 | 0 | 0 | 0 | 0 | 0 |
| *RADIATION INJURIES* | | | | | | | | | | |
| SUNBURN | 0 | 0 | 0 | 0 | 0 | 0 | 0 | 0 | 0 | 0 |
| *THERMAL BURNS* | | | | | | | | | | |

| | | | | | | | | | | |
|---|---|---|---|---|---|---|---|---|---|---|
| THERMAL BURN | 0 | 0 | 0 | 0 | 0 | 0 | 0 | 0 | 0 | 0 |
| THERMAL BURNS OF EYE | 0 | 0 | 0 | 0 | 0 | 0 | 0 | 0 | 0 | 0 |
| ***INJURIES NEC*** | | | | | | | | | | |
| *CHEST AND RESPIRATORY TRACT INJURIES NEC* | | | | | | | | | | |
| CHEST CRUSHING | 0 | 0 | 0 | 0 | 0 | 0 | 0 | 0 | 0 | 0 |
| *EYE INJURIES NEC* | | | | | | | | | | |
| EYE CONTUSION | 0 | 0 | 0 | 0 | 0 | 0 | 0 | 0 | 0 | 0 |
| *MUSCLE, TENDON AND LIGAMENT INJURIES* | | | | | | | | | | |
| LIGAMENT SPRAIN | 0 | 0 | 0 | 0 | 0 | 0 | 0 | 0 | 0 | 0 |
| MUSCLE INJURY | 0 | 0 | 0 | 0 | 0 | 0 | 0 | 0 | 0 | 0 |
| MUSCLE STRAIN | 0 | 0 | 0 | 0 | 0 | 0 | 0 | 0 | 0 | 0 |
| *NERVE INJURIES NEC* | | | | | | | | | | |
| NERVE INJURY | 0 | 0 | 0 | 0 | 0 | 0 | 0 | 0 | 0 | 0 |
| *NON-SITE SPECIFIC INJURIES NEC* | | | | | | | | | | |
| ARTHROPOD STING | 0 | 0 | 0 | 0 | 0 | 0 | 0 | 0 | 0 | 0 |
| BITE | 0 | 0 | 0 | 0 | 0 | 0 | 0 | 0 | 0 | 0 |
| FALL | 0 | 0 | 0 | 0 | 0 | 0 | 0 | 0 | 0 | 0 |
| INFLAMMATION OF WOUND | 0 | 0 | 0 | 0 | 0 | 0 | 0 | 0 | 0 | 0 |
| TISSUE INJURY | 0 | 0 | 0 | 0 | 0 | 0 | 0 | 0 | 0 | 0 |
| WOUND COMPLICATION | 0 | 0 | 0 | 0 | 0 | 0 | 0 | 0 | 0 | 0 |
| WOUND SECRETION | 0 | 0 | 0 | 0 | 0 | 0 | 0 | 0 | 0 | 0 |
| *SITE SPECIFIC INJURIES NEC* | | | | | | | | | | |
| LIMB INJURY | 0 | 0 | 0 | 0 | 0 | 0 | 0 | 0 | 0 | 0 |
| *SKIN INJURIES NEC* | | | | | | | | | | |
| CONTUSION | 0 | 0 | 0 | 0 | 0 | 0 | 0 | 0 | 0 | 0 |
| SCAR | 0 | 0 | 0 | 0 | 0 | 0 | 0 | 0 | 0 | 0 |
| SKIN WOUND | 0 | 0 | 0 | 0 | 0 | 0 | 0 | 0 | 0 | 0 |
| ***PROCEDURAL RELATED INJURIES AND COMPLICATIONS NEC*** | | | | | | | | | | |
| *CARDIAC AND VASCULAR PROCEDURAL COMPLICATIONS* | | | | | | | | | | |
| CARDIAC PROCEDURE COMPLICATION | 0 | 0 | 0 | 0 | 0 | 0 | 0 | 0 | 0 | 0 |
| *GASTROINTESTINAL AND HEPATOBILIARY PROCEDURAL COMPLICATIONS* | | | | | | | | | | |
| PROCEDURAL NAUSEA | 0 | 0 | 0 | 0 | 0 | 0 | 0 | 0 | 0 | 0 |
| *NEUROLOGICAL AND PSYCHIATRIC PROCEDURAL COMPLICATIONS* | | | | | | | | | | |
| PROCEDURAL DIZZINESS | 0 | 0 | 0 | 0 | 0 | 0 | 0 | 0 | 0 | 0 |
| *NON-SITE SPECIFIC PROCEDURAL COMPLICATIONS* | | | | | | | | | | |
| INCISION SITE PAIN | 0 | 0 | 0 | 0 | 0 | 0 | 0 | 0 | 0 | 0 |

| | | | | | | | | | | |
|---|---|---|---|---|---|---|---|---|---|---|
| INCISION SITE SWELLING | 0 | 0 | 0 | 0 | 0 | 0 | 0 | 0 | 0 | 0 |
| INJECTION RELATED REACTION | 1 | 1 | 0 | 0 | 0 | 1 | 1 | 0 | 0 | 0 |
| POST PROCEDURAL COMPLICATION | 3 | 3 | 0 | 0 | 0 | 3 | 3 | 0 | 0 | 0 |
| PROCEDURAL PAIN | 0 | 0 | 0 | 0 | 0 | 0 | 0 | 0 | 0 | 0 |
| *VACCINATION RELATED COMPLICATIONS* | | | | | | | | | | |
| IMMUNISATION REACTION | 0 | 0 | 0 | 0 | 0 | 0 | 0 | 0 | 0 | 0 |
| **INVESTIGATIONS** | | | | | | | | | | |
| ***CARDIAC AND VASCULAR INVESTIGATIONS (EXCL ENZYME TESTS)*** | | | | | | | | | | |
| *HEART RATE AND PULSE INVESTIGATIONS* | | | | | | | | | | |
| HEART RATE | 0 | 0 | 0 | 0 | 0 | 0 | 0 | 0 | 0 | 0 |
| HEART RATE DECREASED | 0 | 0 | 0 | 0 | 0 | 0 | 0 | 0 | 0 | 0 |
| HEART RATE INCREASED | 1 | 1 | 0 | 0 | 0 | 1 | 1 | 0 | 0 | 0 |
| HEART RATE IRREGULAR | 0 | 0 | 0 | 0 | 0 | 0 | 0 | 0 | 0 | 0 |
| *VASCULAR TESTS NEC (INCL BLOOD PRESSURE)* | | | | | | | | | | |
| BLOOD PRESSURE DECREASED | 0 | 0 | 0 | 0 | 0 | 0 | 0 | 0 | 0 | 0 |
| BLOOD PRESSURE INCREASED | 0 | 0 | 0 | 0 | 0 | 0 | 0 | 0 | 0 | 0 |
| BLOOD PRESSURE MEASUREMENT | 0 | 0 | 0 | 0 | 0 | 0 | 0 | 0 | 0 | 0 |
| ***ENDOCRINE INVESTIGATIONS (INCL SEX HORMONES)*** | | | | | | | | | | |
| *ENDOCRINE ANALYSES AND IMAGING NEC* | | | | | | | | | | |
| HORMONE LEVEL ABNORMAL | 0 | 0 | 0 | 0 | 0 | 0 | 0 | 0 | 0 | 0 |
| *PITUITARY ANALYSES ANTERIOR* | | | | | | | | | | |
| BLOOD FOLLICLE STIMULATING HORMONE INCREASED | 0 | 0 | 0 | 0 | 0 | 0 | 0 | 0 | 0 | 0 |
| BLOOD LUTEINISING HORMONE | 0 | 0 | 0 | 0 | 0 | 0 | 0 | 0 | 0 | 0 |
| *THYROID ANALYSES* | | | | | | | | | | |
| TRI-IODOTHYRONINE | 0 | 0 | 0 | 0 | 0 | 0 | 0 | 0 | 0 | 0 |
| ***HAEMATOLOGY INVESTIGATIONS (INCL BLOOD GROUPS)*** | | | | | | | | | | |
| *COAGULATION AND BLEEDING ANALYSES* | | | | | | | | | | |
| BLEEDING TIME | 0 | 0 | 0 | 0 | 0 | 0 | 0 | 0 | 0 | 0 |
| INTERNATIONAL NORMALISED RATIO DECREASED | 0 | 0 | 0 | 0 | 0 | 0 | 0 | 0 | 0 | 0 |
| *PLATELET ANALYSES* | | | | | | | | | | |
| PLATELET COUNT INCREASED | 0 | 0 | 0 | 0 | 0 | 0 | 0 | 0 | 0 | 0 |
| *RED BLOOD CELL ANALYSES* | | | | | | | | | | |
| HAEMOGLOBIN | 0 | 0 | 0 | 0 | 0 | 0 | 0 | 0 | 0 | 0 |
| ***METABOLIC, NUTRITIONAL AND BLOOD GAS INVESTIGATIONS*** | | | | | | | | | | |
| *BLOOD GAS AND ACID BASE ANALYSES* | | | | | | | | | | |

| | | | | | | | | | | |
|---|---|---|---|---|---|---|---|---|---|---|
| OXYGEN SATURATION DECREASED | 0 | 0 | 0 | 0 | 0 | 0 | 0 | 0 | 0 | 0 |
| *CARBOHYDRATE TOLERANCE ANALYSES (INCL DIABETES)* | | | | | | | | | | |
| BLOOD GLUCOSE | 0 | 0 | 0 | 0 | 0 | 0 | 0 | 0 | 0 | 0 |
| BLOOD GLUCOSE ABNORMAL | 0 | 0 | 0 | 0 | 0 | 0 | 0 | 0 | 0 | 0 |
| BLOOD GLUCOSE DECREASED | 0 | 0 | 0 | 0 | 0 | 0 | 0 | 0 | 0 | 0 |
| BLOOD GLUCOSE INCREASED | 0 | 0 | 0 | 0 | 0 | 0 | 0 | 0 | 0 | 0 |
| ***MICROBIOLOGY AND SEROLOGY INVESTIGATIONS*** | | | | | | | | | | |
| *VIRUS IDENTIFICATION AND SEROLOGY* | | | | | | | | | | |
| SARS-COV-2 TEST | 0 | 0 | 0 | 0 | 0 | 0 | 0 | 0 | 0 | 0 |
| SARS-COV-2 TEST POSITIVE | 0 | 0 | 0 | 0 | 0 | 0 | 0 | 0 | 0 | 0 |
| ***MUSCULOSKELETAL AND SOFT TISSUE INVESTIGATIONS (EXCL ENZYME TESTS)*** | | | | | | | | | | |
| *MUSCULOSKELETAL AND SOFT TISSUE IMAGING PROCEDURES* | | | | | | | | | | |
| BONE SCAN | 0 | 0 | 0 | 0 | 0 | 0 | 0 | 0 | 0 | 0 |
| ***NEUROLOGICAL, SPECIAL SENSES AND PSYCHIATRIC INVESTIGATIONS*** | | | | | | | | | | |
| *CENTRAL NERVOUS SYSTEM IMAGING PROCEDURES* | | | | | | | | | | |
| MAGNETIC RESONANCE IMAGING HEAD | 0 | 0 | 0 | 0 | 0 | 0 | 0 | 0 | 0 | 0 |
| SCAN BRAIN | 0 | 0 | 0 | 0 | 0 | 0 | 0 | 0 | 0 | 0 |
| ***PHYSICAL EXAMINATION AND ORGAN SYSTEM STATUS TOPICS*** | | | | | | | | | | |
| *PHYSICAL EXAMINATION PROCEDURES AND ORGAN SYSTEM STATUS* | | | | | | | | | | |
| BODY TEMPERATURE | 0 | 0 | 0 | 0 | 0 | 0 | 0 | 0 | 0 | 0 |
| BODY TEMPERATURE ABNORMAL | 0 | 0 | 0 | 0 | 0 | 0 | 0 | 0 | 0 | 0 |
| BODY TEMPERATURE DECREASED | 0 | 0 | 0 | 0 | 0 | 0 | 0 | 0 | 0 | 0 |
| BODY TEMPERATURE FLUCTUATION | 0 | 0 | 0 | 0 | 0 | 0 | 0 | 0 | 0 | 0 |
| BODY TEMPERATURE INCREASED | 1 | 1 | 0 | 0 | 0 | 1 | 1 | 0 | 0 | 0 |
| GRIP STRENGTH DECREASED | 0 | 0 | 0 | 0 | 0 | 0 | 0 | 0 | 0 | 0 |
| HEAD LAG | 0 | 0 | 0 | 0 | 0 | 0 | 0 | 0 | 0 | 0 |
| LEFT-HANDEDNESS | 0 | 0 | 0 | 0 | 0 | 0 | 0 | 0 | 0 | 0 |
| LYMPH NODE PALPABLE | 0 | 0 | 0 | 0 | 0 | 0 | 0 | 0 | 0 | 0 |
| RESPIRATORY RATE DECREASED | 0 | 0 | 0 | 0 | 0 | 0 | 0 | 0 | 0 | 0 |
| SKIN TEMPERATURE | 0 | 0 | 0 | 0 | 0 | 0 | 0 | 0 | 0 | 0 |
| WEIGHT DECREASED | 0 | 0 | 0 | 0 | 0 | 0 | 0 | 0 | 0 | 0 |
| WEIGHT INCREASED | 0 | 0 | 0 | 0 | 0 | 0 | 0 | 0 | 0 | 0 |
| ***RENAL AND URINARY TRACT INVESTIGATIONS AND URINALYSES*** | | | | | | | | | | |
| *URINALYSIS NEC* | | | | | | | | | | |
| BLOOD URINE | 0 | 0 | 0 | 0 | 0 | 0 | 0 | 0 | 0 | 0 |

| | | | | | | | | | | |
|---|---|---|---|---|---|---|---|---|---|---|
| NITRITE URINE PRESENT | 0 | 0 | 0 | 0 | 0 | 0 | 0 | 0 | 0 | 0 |
| PH URINE | 0 | 0 | 0 | 0 | 0 | 0 | 0 | 0 | 0 | 0 |
| *URINARY TRACT FUNCTION ANALYSES NEC* | | | | | | | | | | |
| URINE OUTPUT | 0 | 0 | 0 | 0 | 0 | 0 | 0 | 0 | 0 | 0 |
| URINE OUTPUT INCREASED | 0 | 0 | 0 | 0 | 0 | 0 | 0 | 0 | 0 | 0 |
| ***REPRODUCTIVE ORGAN AND BREAST INVESTIGATIONS (EXCL HORMONE ANALYSES)*** | | | | | | | | | | |
| *REPRODUCTIVE ORGAN AND BREAST IMAGING PROCEDURES* | | | | | | | | | | |
| BREAST SCAN | 0 | 0 | 0 | 0 | 0 | 0 | 0 | 0 | 0 | 0 |
| ***RESPIRATORY AND PULMONARY INVESTIGATIONS (EXCL BLOOD GASES)*** | | | | | | | | | | |
| *RESPIRATORY AND PULMONARY FUNCTION DIAGNOSTIC PROCEDURES* | | | | | | | | | | |
| FORCED EXPIRATORY VOLUME DECREASED | 0 | 0 | 0 | 0 | 0 | 0 | 0 | 0 | 0 | 0 |
| FORCED EXPIRATORY VOLUME INCREASED | 0 | 0 | 0 | 0 | 0 | 0 | 0 | 0 | 0 | 0 |
| ***WATER, ELECTROLYTE AND MINERAL INVESTIGATIONS*** | | | | | | | | | | |
| *WATER AND ELECTROLYTE ANALYSES NEC* | | | | | | | | | | |
| VOLUME BLOOD | 0 | 0 | 0 | 0 | 0 | 0 | 0 | 0 | 0 | 0 |
| **METABOLISM AND NUTRITION DISORDERS** | | | | | | | | | | |
| ***APPETITE AND GENERAL NUTRITIONAL DISORDERS*** | | | | | | | | | | |
| *APPETITE DISORDERS* | | | | | | | | | | |
| APPETITE DISORDER | 0 | 0 | 0 | 0 | 0 | 0 | 0 | 0 | 0 | 0 |
| DECREASED APPETITE | 0 | 0 | 0 | 0 | 0 | 0 | 0 | 0 | 0 | 0 |
| FOOD CRAVING | 0 | 0 | 0 | 0 | 0 | 0 | 0 | 0 | 0 | 0 |
| FOOD REFUSAL | 0 | 0 | 0 | 0 | 0 | 0 | 0 | 0 | 0 | 0 |
| INCREASED APPETITE | 0 | 0 | 0 | 0 | 0 | 0 | 0 | 0 | 0 | 0 |
| *GENERAL NUTRITIONAL DISORDERS NEC* | | | | | | | | | | |
| FOOD AVERSION | 0 | 0 | 0 | 0 | 0 | 0 | 0 | 0 | 0 | 0 |
| ***ELECTROLYTE AND FLUID BALANCE CONDITIONS*** | | | | | | | | | | |
| *TOTAL FLUID VOLUME DECREASED* | | | | | | | | | | |
| DEHYDRATION | 0 | 0 | 0 | 0 | 0 | 0 | 0 | 0 | 0 | 0 |
| *TOTAL FLUID VOLUME INCREASED* | | | | | | | | | | |
| FLUID RETENTION | 0 | 0 | 0 | 0 | 0 | 0 | 0 | 0 | 0 | 0 |
| ***FOOD INTOLERANCE SYNDROMES*** | | | | | | | | | | |
| *FOOD MALABSORPTION AND INTOLERANCE SYNDROMES (EXCL SUGAR INTOLERANCE)* | | | | | | | | | | |
| ALCOHOL INTOLERANCE | 0 | 0 | 0 | 0 | 0 | 0 | 0 | 0 | 0 | 0 |
| ***GLUCOSE METABOLISM DISORDERS (INCL DIABETES MELLITUS)*** | | | | | | | | | | |
| *DIABETES MELLITUS (INCL SUBTYPES)* | | | | | | | | | | |
| DIABETES MELLITUS | 0 | 0 | 0 | 0 | 0 | 0 | 0 | 0 | 0 | 0 |

| | | | | | | | | | | |
|---|---|---|---|---|---|---|---|---|---|---|
| DIABETES MELLITUS INADEQUATE CONTROL | 0 | 0 | 0 | 0 | 0 | 0 | 0 | 0 | 0 | 0 |
| *HYPERGLYCAEMIC CONDITIONS NEC* | | | | | | | | | | |
| HYPERGLYCAEMIA | 0 | 0 | 0 | 0 | 0 | 0 | 0 | 0 | 0 | 0 |
| *HYPOGLYCAEMIC CONDITIONS NEC* | | | | | | | | | | |
| HYPOGLYCAEMIA | 0 | 0 | 0 | 0 | 0 | 0 | 0 | 0 | 0 | 0 |
| ***PURINE AND PYRIMIDINE METABOLISM DISORDERS*** | | | | | | | | | | |
| *DISORDERS OF PURINE METABOLISM* | | | | | | | | | | |
| GOUT | 0 | 0 | 0 | 0 | 0 | 0 | 0 | 0 | 0 | 0 |
| **MUSCULOSKELETAL AND CONNECTIVE TISSUE DISORDERS** | | | | | | | | | | |
| ***BONE DISORDERS (EXCL CONGENITAL AND FRACTURES)*** | | | | | | | | | | |
| *BONE DISORDERS NEC* | | | | | | | | | | |
| JAW DISORDER | 0 | 0 | 0 | 0 | 0 | 0 | 0 | 0 | 0 | 0 |
| OSTEITIS | 0 | 0 | 0 | 0 | 0 | 0 | 0 | 0 | 0 | 0 |
| *BONE RELATED SIGNS AND SYMPTOMS* | | | | | | | | | | |
| BONE PAIN | 0 | 0 | 0 | 0 | 0 | 0 | 0 | 0 | 0 | 0 |
| BONE SWELLING | 0 | 0 | 0 | 0 | 0 | 0 | 0 | 0 | 0 | 0 |
| PAIN IN JAW | 0 | 0 | 0 | 0 | 0 | 0 | 0 | 0 | 0 | 0 |
| PUBIC PAIN | 0 | 0 | 0 | 0 | 0 | 0 | 0 | 0 | 0 | 0 |
| SPINAL PAIN | 0 | 0 | 0 | 0 | 0 | 0 | 0 | 0 | 0 | 0 |
| ***CONNECTIVE TISSUE DISORDERS (EXCL CONGENITAL)*** | | | | | | | | | | |
| *CONNECTIVE TISSUE DISORDERS NEC* | | | | | | | | | | |
| POLYMYALGIA RHEUMATICA | 0 | 0 | 0 | 0 | 0 | 0 | 0 | 0 | 0 | 0 |
| *LUPUS ERYTHEMATOSUS (INCL SUBTYPES)* | | | | | | | | | | |
| SYSTEMIC LUPUS ERYTHEMATOSUS | 0 | 0 | 0 | 0 | 0 | 0 | 0 | 0 | 0 | 0 |
| ***JOINT DISORDERS*** | | | | | | | | | | |
| *ARTHROPATHIES NEC* | | | | | | | | | | |
| ARTHRITIS | 0 | 0 | 0 | 0 | 0 | 0 | 0 | 0 | 0 | 0 |
| ARTHROPATHY | 0 | 0 | 0 | 0 | 0 | 0 | 0 | 0 | 0 | 0 |
| POLYARTHRITIS | 0 | 0 | 0 | 0 | 0 | 0 | 0 | 0 | 0 | 0 |
| RHEUMATIC FEVER | 0 | 0 | 0 | 0 | 0 | 0 | 0 | 0 | 0 | 0 |
| *JOINT RELATED DISORDERS NEC* | | | | | | | | | | |
| JOINT LOCK | 0 | 0 | 0 | 0 | 0 | 0 | 0 | 0 | 0 | 0 |
| PERIARTHRITIS | 0 | 0 | 0 | 0 | 0 | 0 | 0 | 0 | 0 | 0 |
| TEMPOROMANDIBULAR PAIN AND DYSFUNCTION SYNDROME | 0 | 0 | 0 | 0 | 0 | 0 | 0 | 0 | 0 | 0 |
| *JOINT RELATED SIGNS AND SYMPTOMS* | | | | | | | | | | |

| | | | | | | | | | | |
|---|---|---|---|---|---|---|---|---|---|---|
| ARTHRALGIA | 12 | 6 | 4 | 1 | 1 | 11 | 6 | 3 | 1 | 1 |
| JAW CLICKING | 0 | 0 | 0 | 0 | 0 | 0 | 0 | 0 | 0 | 0 |
| JOINT STIFFNESS | 0 | 0 | 0 | 0 | 0 | 0 | 0 | 0 | 0 | 0 |
| JOINT SWELLING | 0 | 0 | 0 | 0 | 0 | 0 | 0 | 0 | 0 | 0 |
| *OSTEOARTHROPATHIES* | | | | | | | | | | |
| OSTEOARTHRITIS | 0 | 0 | 0 | 0 | 0 | 0 | 0 | 0 | 0 | 0 |
| *PSORIATIC ARTHROPATHIES* | | | | | | | | | | |
| PSORIATIC ARTHROPATHY | 0 | 0 | 0 | 0 | 0 | 0 | 0 | 0 | 0 | 0 |
| *RHEUMATOID ARTHROPATHIES* | | | | | | | | | | |
| RHEUMATOID ARTHRITIS | 0 | 0 | 0 | 0 | 0 | 0 | 0 | 0 | 0 | 0 |
| ***MUSCLE DISORDERS*** | | | | | | | | | | |
| *MUSCLE INFECTIONS AND INFLAMMATIONS* | | | | | | | | | | |
| MYOSITIS | 0 | 0 | 0 | 0 | 0 | 0 | 0 | 0 | 0 | 0 |
| *MUSCLE PAINS* | | | | | | | | | | |
| FIBROMYALGIA | 0 | 0 | 0 | 0 | 0 | 0 | 0 | 0 | 0 | 0 |
| MYALGIA | 12 | 10 | 2 | 0 | 0 | 10 | 10 | 0 | 0 | 0 |
| *MUSCLE RELATED SIGNS AND SYMPTOMS NEC* | | | | | | | | | | |
| MUSCLE ATROPHY | 0 | 0 | 0 | 0 | 0 | 0 | 0 | 0 | 0 | 0 |
| MUSCLE DISCOMFORT | 0 | 0 | 0 | 0 | 0 | 0 | 0 | 0 | 0 | 0 |
| MUSCLE FATIGUE | 6 | 5 | 1 | 0 | 0 | 5 | 5 | 0 | 0 | 0 |
| MUSCLE FIBROSIS | 0 | 0 | 0 | 0 | 0 | 0 | 0 | 0 | 0 | 0 |
| MUSCLE MASS | 0 | 0 | 0 | 0 | 0 | 0 | 0 | 0 | 0 | 0 |
| MUSCLE SPASMS | 1 | 1 | 0 | 0 | 0 | 1 | 1 | 0 | 0 | 0 |
| MUSCLE TIGHTNESS | 0 | 0 | 0 | 0 | 0 | 0 | 0 | 0 | 0 | 0 |
| MUSCLE TWITCHING | 0 | 0 | 0 | 0 | 0 | 0 | 0 | 0 | 0 | 0 |
| *MUSCLE TONE ABNORMALITIES* | | | | | | | | | | |
| MUSCLE RIGIDITY | 0 | 0 | 0 | 0 | 0 | 0 | 0 | 0 | 0 | 0 |
| TRISMUS | 0 | 0 | 0 | 0 | 0 | 0 | 0 | 0 | 0 | 0 |
| *MUSCLE WEAKNESS CONDITIONS* | | | | | | | | | | |
| MUSCULAR WEAKNESS | 1 | 1 | 0 | 0 | 0 | 1 | 1 | 0 | 0 | 0 |
| ***MUSCULOSKELETAL AND CONNECTIVE TISSUE DISORDERS NEC*** | | | | | | | | | | |
| *MUSCULOSKELETAL AND CONNECTIVE TISSUE CONDITIONS NEC* | | | | | | | | | | |
| MOBILITY DECREASED | 0 | 0 | 0 | 0 | 0 | 0 | 0 | 0 | 0 | 0 |
| MUSCULOSKELETAL STIFFNESS | 1 | 1 | 0 | 0 | 0 | 1 | 1 | 0 | 0 | 0 |
| BACK PAIN | 2 | 2 | 0 | 0 | 0 | 2 | 2 | 0 | 0 | 0 |

| | | | | | | | | | | |
|---|---|---|---|---|---|---|---|---|---|---|
| FLANK PAIN | 0 | 0 | 0 | 0 | 0 | 0 | 0 | 0 | 0 | 0 |
| LIMB DISCOMFORT | 3 | 1 | 0 | 2 | 0 | 3 | 1 | 0 | 2 | 0 |
| MUSCULOSKELETAL CHEST PAIN | 0 | 0 | 0 | 0 | 0 | 0 | 0 | 0 | 0 | 0 |
| MUSCULOSKELETAL DISCOMFORT | 0 | 0 | 0 | 0 | 0 | 0 | 0 | 0 | 0 | 0 |
| MUSCULOSKELETAL PAIN | 0 | 0 | 0 | 0 | 0 | 0 | 0 | 0 | 0 | 0 |
| NECK PAIN | 2 | 2 | 0 | 0 | 0 | 2 | 2 | 0 | 0 | 0 |
| PAIN IN EXTREMITY | 29 | 9 | 5 | 8 | 7 | 24 | 9 | 4 | 8 | 3 |
| *SOFT TISSUE DISORDERS NEC* | | | | | | | | | | |
| AXILLARY MASS | 0 | 0 | 0 | 0 | 0 | 0 | 0 | 0 | 0 | 0 |
| GROIN PAIN | 0 | 0 | 0 | 0 | 0 | 0 | 0 | 0 | 0 | 0 |
| ***SYNOVIAL AND BURSAL DISORDERS*** | | | | | | | | | | |
| *SYNOVIAL DISORDERS* | | | | | | | | | | |
| SYNOVITIS | 0 | 0 | 0 | 0 | 0 | 0 | 0 | 0 | 0 | 0 |
| ***TENDON, LIGAMENT AND CARTILAGE DISORDERS*** | | | | | | | | | | |
| *CARTILAGE DISORDERS* | | | | | | | | | | |
| COSTOCHONDRITIS | 0 | 0 | 0 | 0 | 0 | 0 | 0 | 0 | 0 | 0 |
| *TENDON DISORDERS* | | | | | | | | | | |
| TENDONITIS | 0 | 0 | 0 | 0 | 0 | 0 | 0 | 0 | 0 | 0 |
| TENOSYNOVITIS | 0 | 0 | 0 | 0 | 0 | 0 | 0 | 0 | 0 | 0 |
| TRIGGER FINGER | 0 | 0 | 0 | 0 | 0 | 0 | 0 | 0 | 0 | 0 |
| **NEOPLASMS BENIGN, MALIGNANT AND UNSPECIFIED (INCL CYSTS AND POLYPS)** | | | | | | | | | | |
| ***CUTANEOUS NEOPLASMS BENIGN*** | | | | | | | | | | |
| *SKIN NEOPLASMS BENIGN* | | | | | | | | | | |
| MELANOCYTIC NAEVUS | 0 | 0 | 0 | 0 | 0 | 0 | 0 | 0 | 0 | 0 |
| SEBORRHOEIC KERATOSIS | 0 | 0 | 0 | 0 | 0 | 0 | 0 | 0 | 0 | 0 |
| SKIN PAPILLOMA | 0 | 0 | 0 | 0 | 0 | 0 | 0 | 0 | 0 | 0 |
| **NERVOUS SYSTEM DISORDERS** | | | | | | | | | | |
| ***CENTRAL NERVOUS SYSTEM INFECTIONS AND INFLAMMATIONS*** | | | | | | | | | | |
| *MYELITIS (INCL INFECTIVE)* | | | | | | | | | | |
| MYELITIS TRANSVERSE | 0 | 0 | 0 | 0 | 0 | 0 | 0 | 0 | 0 | 0 |
| ***CENTRAL NERVOUS SYSTEM VASCULAR DISORDERS*** | | | | | | | | | | |
| *CENTRAL NERVOUS SYSTEM HAEMORRHAGES AND CEREBROVASCULAR ACCIDENTS* | | | | | | | | | | |
| CEREBRAL HAEMORRHAGE | 0 | 0 | 0 | 0 | 0 | 0 | 0 | 0 | 0 | 0 |
| CEREBROVASCULAR ACCIDENT | 0 | 0 | 0 | 0 | 0 | 0 | 0 | 0 | 0 | 0 |
| *TRANSIENT CEREBROVASCULAR EVENTS* | | | | | | | | | | |
| TRANSIENT ISCHAEMIC ATTACK | 0 | 0 | 0 | 0 | 0 | 0 | 0 | 0 | 0 | 0 |

| | | | | | | | | | | |
|---|---|---|---|---|---|---|---|---|---|---|
| ***CRANIAL NERVE DISORDERS (EXCL NEOPLASMS)*** | | | | | | | | | | |
| *FACIAL CRANIAL NERVE DISORDERS* | | | | | | | | | | |
| BELL'S PALSY | 0 | 0 | 0 | 0 | 0 | 0 | 0 | 0 | 0 | 0 |
| FACIAL PARALYSIS | 0 | 0 | 0 | 0 | 0 | 0 | 0 | 0 | 0 | 0 |
| FACIAL PARESIS | 0 | 0 | 0 | 0 | 0 | 0 | 0 | 0 | 0 | 0 |
| *OLFACTORY NERVE DISORDERS* | | | | | | | | | | |
| ANOSMIA | 0 | 0 | 0 | 0 | 0 | 0 | 0 | 0 | 0 | 0 |
| HYPOSMIA | 0 | 0 | 0 | 0 | 0 | 0 | 0 | 0 | 0 | 0 |
| PAROSMIA | 0 | 0 | 0 | 0 | 0 | 0 | 0 | 0 | 0 | 0 |
| *TRIGEMINAL DISORDERS* | | | | | | | | | | |
| TRIGEMINAL NEURALGIA | 0 | 0 | 0 | 0 | 0 | 0 | 0 | 0 | 0 | 0 |
| TRIGEMINAL NEURITIS | 0 | 0 | 0 | 0 | 0 | 0 | 0 | 0 | 0 | 0 |
| ***DEMYELINATING DISORDERS*** | | | | | | | | | | |
| *MULTIPLE SCLEROSIS ACUTE AND PROGRESSIVE* | | | | | | | | | | |
| MULTIPLE SCLEROSIS RELAPSE | 0 | 0 | 0 | 0 | 0 | 0 | 0 | 0 | 0 | 0 |
| ***HEADACHES*** | | | | | | | | | | |
| *HEADACHES NEC* | | | | | | | | | | |
| CLUSTER HEADACHE | 0 | 0 | 0 | 0 | 0 | 0 | 0 | 0 | 0 | 0 |
| COLD-STIMULUS HEADACHE | 0 | 0 | 0 | 0 | 0 | 0 | 0 | 0 | 0 | 0 |
| DRUG WITHDRAWAL HEADACHE | 0 | 0 | 0 | 0 | 0 | 0 | 0 | 0 | 0 | 0 |
| HEADACHE | 37 | 26 | 3 | 8 | 0 | 36 | 26 | 2 | 8 | 0 |
| PRIMARY STABBING HEADACHE | 0 | 0 | 0 | 0 | 0 | 0 | 0 | 0 | 0 | 0 |
| SINUS HEADACHE | 2 | 1 | 1 | 0 | 0 | 2 | 1 | 1 | 0 | 0 |
| TENSION HEADACHE | 5 | 2 | 0 | 3 | 0 | 5 | 2 | 0 | 3 | 0 |
| THUNDERCLAP HEADACHE | 0 | 0 | 0 | 0 | 0 | 0 | 0 | 0 | 0 | 0 |
| VASCULAR HEADACHE | 0 | 0 | 0 | 0 | 0 | 0 | 0 | 0 | 0 | 0 |
| *MIGRAINE HEADACHES* | | | | | | | | | | |
| MIGRAINE | 3 | 3 | 0 | 0 | 0 | 3 | 3 | 0 | 0 | 0 |
| MIGRAINE WITH AURA | 0 | 0 | 0 | 0 | 0 | 0 | 0 | 0 | 0 | 0 |
| RETINAL MIGRAINE | 0 | 0 | 0 | 0 | 0 | 0 | 0 | 0 | 0 | 0 |
| TYPICAL AURA WITHOUT HEADACHE | 0 | 0 | 0 | 0 | 0 | 0 | 0 | 0 | 0 | 0 |
| ***MENTAL IMPAIRMENT DISORDERS*** | | | | | | | | | | |
| *INTELLECTUAL DISABILITIES* | | | | | | | | | | |
| INTELLECTUAL DISABILITY | 0 | 0 | 0 | 0 | 0 | 0 | 0 | 0 | 0 | 0 |
| *MEMORY LOSS (EXCL DEMENTIA)* | | | | | | | | | | |

| | | | | | | | | | | |
|---|---|---|---|---|---|---|---|---|---|---|
| AMNESIA | 0 | 0 | 0 | 0 | 0 | 0 | 0 | 0 | 0 | 0 |
| MEMORY IMPAIRMENT | 0 | 0 | 0 | 0 | 0 | 0 | 0 | 0 | 0 | 0 |
| *MENTAL IMPAIRMENT (EXCL DEMENTIA AND MEMORY LOSS)* | | | | | | | | | | |
| COGNITIVE DISORDER | 0 | 0 | 0 | 0 | 0 | 0 | 0 | 0 | 0 | 0 |
| DISTURBANCE IN ATTENTION | 0 | 0 | 0 | 0 | 0 | 0 | 0 | 0 | 0 | 0 |
| MENTAL IMPAIRMENT | 0 | 0 | 0 | 0 | 0 | 0 | 0 | 0 | 0 | 0 |
| ***MOVEMENT DISORDERS (INCL PARKINSONISM)*** | | | | | | | | | | |
| *DYSKINESIAS AND MOVEMENT DISORDERS NEC* | | | | | | | | | | |
| BRADYKINESIA | 0 | 0 | 0 | 0 | 0 | 0 | 0 | 0 | 0 | 0 |
| CLUMSINESS | 0 | 0 | 0 | 0 | 0 | 0 | 0 | 0 | 0 | 0 |
| DYSKINESIA | 0 | 0 | 0 | 0 | 0 | 0 | 0 | 0 | 0 | 0 |
| EXTRAPYRAMIDAL DISORDER | 0 | 0 | 0 | 0 | 0 | 0 | 0 | 0 | 0 | 0 |
| PSYCHOMOTOR HYPERACTIVITY | 0 | 0 | 0 | 0 | 0 | 0 | 0 | 0 | 0 | 0 |
| *PARALYSIS AND PARESIS (EXCL CRANIAL NERVE)* | | | | | | | | | | |
| HEMIPLEGIA | 0 | 0 | 0 | 0 | 0 | 0 | 0 | 0 | 0 | 0 |
| MONOPARESIS | 0 | 0 | 0 | 0 | 0 | 0 | 0 | 0 | 0 | 0 |
| MONOPLEGIA | 0 | 0 | 0 | 0 | 0 | 0 | 0 | 0 | 0 | 0 |
| PARALYSIS | 0 | 0 | 0 | 0 | 0 | 0 | 0 | 0 | 0 | 0 |
| *PARKINSON'S DISEASE AND PARKINSONISM* | | | | | | | | | | |
| FREEZING PHENOMENON | 0 | 0 | 0 | 0 | 0 | 0 | 0 | 0 | 0 | 0 |
| PARKINSON'S DISEASE | 0 | 0 | 0 | 0 | 0 | 0 | 0 | 0 | 0 | 0 |
| *TREMOR (EXCL CONGENITAL)* | | | | | | | | | | |
| TREMOR | 3 | 2 | 0 | 1 | 0 | 3 | 2 | 0 | 1 | 0 |
| ***NEUROLOGICAL DISORDERS NEC*** | | | | | | | | | | |
| *COMA STATES* | | | | | | | | | | |
| DIABETIC HYPERGLYCAEMIC COMA | 0 | 0 | 0 | 0 | 0 | 0 | 0 | 0 | 0 | 0 |
| *COORDINATION AND BALANCE DISTURBANCES* | | | | | | | | | | |
| BALANCE DISORDER | 0 | 0 | 0 | 0 | 0 | 0 | 0 | 0 | 0 | 0 |
| COORDINATION ABNORMAL | 0 | 0 | 0 | 0 | 0 | 0 | 0 | 0 | 0 | 0 |
| DYSSTASIA | 0 | 0 | 0 | 0 | 0 | 0 | 0 | 0 | 0 | 0 |
| VESTIBULAR NYSTAGMUS | 0 | 0 | 0 | 0 | 0 | 0 | 0 | 0 | 0 | 0 |
| *DISTURBANCES IN CONSCIOUSNESS NEC* | | | | | | | | | | |
| DEPRESSED LEVEL OF CONSCIOUSNESS | 0 | 0 | 0 | 0 | 0 | 0 | 0 | 0 | 0 | 0 |
| LETHARGY | 1 | 0 | 1 | 0 | 0 | 1 | 0 | 1 | 0 | 0 |
| LOSS OF CONSCIOUSNESS | 0 | 0 | 0 | 0 | 0 | 0 | 0 | 0 | 0 | 0 |

| | | | | | | | | | | |
|---|---|---|---|---|---|---|---|---|---|---|
| SEDATION | 0 | 0 | 0 | 0 | 0 | 0 | 0 | 0 | 0 | 0 |
| SOMNOLENCE | 2 | 0 | 2 | 0 | 0 | 2 | 0 | 2 | 0 | 0 |
| SYNCOPE | 0 | 0 | 0 | 0 | 0 | 0 | 0 | 0 | 0 | 0 |
| *NERVOUS SYSTEM DISORDERS NEC* | | | | | | | | | | |
| NERVOUS SYSTEM DISORDER | 0 | 0 | 0 | 0 | 0 | 0 | 0 | 0 | 0 | 0 |
| *NEUROLOGICAL SIGNS AND SYMPTOMS NEC* | | | | | | | | | | |
| AGITATION NEONATAL | 0 | 0 | 0 | 0 | 0 | 0 | 0 | 0 | 0 | 0 |
| BRAIN FOG | 1 | 1 | 0 | 0 | 0 | 1 | 1 | 0 | 0 | 0 |
| DIZZINESS | 2 | 1 | 0 | 1 | 0 | 2 | 1 | 0 | 1 | 0 |
| DIZZINESS EXERTIONAL | 0 | 0 | 0 | 0 | 0 | 0 | 0 | 0 | 0 | 0 |
| DIZZINESS POSTURAL | 3 | 2 | 1 | 0 | 0 | 3 | 2 | 1 | 0 | 0 |
| HEAD DISCOMFORT | 0 | 0 | 0 | 0 | 0 | 0 | 0 | 0 | 0 | 0 |
| INFANT IRRITABILITY | 0 | 0 | 0 | 0 | 0 | 0 | 0 | 0 | 0 | 0 |
| MENINGISM | 0 | 0 | 0 | 0 | 0 | 0 | 0 | 0 | 0 | 0 |
| MYOCLONUS | 0 | 0 | 0 | 0 | 0 | 0 | 0 | 0 | 0 | 0 |
| NEUROLOGICAL SYMPTOM | 0 | 0 | 0 | 0 | 0 | 0 | 0 | 0 | 0 | 0 |
| PERSISTENT POSTURAL-PERCEPTUAL DIZZINESS | 0 | 0 | 0 | 0 | 0 | 0 | 0 | 0 | 0 | 0 |
| PRESYNCOPE | 0 | 0 | 0 | 0 | 0 | 0 | 0 | 0 | 0 | 0 |
| *PARAESTHESIAS AND DYSAESTHESIAS* | | | | | | | | | | |
| BURNING FEET SYNDROME | 0 | 0 | 0 | 0 | 0 | 0 | 0 | 0 | 0 | 0 |
| BURNING SENSATION | 0 | 0 | 0 | 0 | 0 | 0 | 0 | 0 | 0 | 0 |
| FORMICATION | 0 | 0 | 0 | 0 | 0 | 0 | 0 | 0 | 0 | 0 |
| HYPERAESTHESIA | 0 | 0 | 0 | 0 | 0 | 0 | 0 | 0 | 0 | 0 |
| HYPOAESTHESIA | 0 | 0 | 0 | 0 | 0 | 0 | 0 | 0 | 0 | 0 |
| PARAESTHESIA | 2 | 2 | 0 | 0 | 0 | 2 | 2 | 0 | 0 | 0 |
| REVERSED HOT-COLD SENSATION | 0 | 0 | 0 | 0 | 0 | 0 | 0 | 0 | 0 | 0 |
| *SENSORY ABNORMALITIES NEC* | | | | | | | | | | |
| AGEUSIA | 0 | 0 | 0 | 0 | 0 | 0 | 0 | 0 | 0 | 0 |
| ALLODYNIA | 0 | 0 | 0 | 0 | 0 | 0 | 0 | 0 | 0 | 0 |
| DYSGEUSIA | 0 | 0 | 0 | 0 | 0 | 0 | 0 | 0 | 0 | 0 |
| HYPOGEUSIA | 0 | 0 | 0 | 0 | 0 | 0 | 0 | 0 | 0 | 0 |
| NEURALGIA | 0 | 0 | 0 | 0 | 0 | 0 | 0 | 0 | 0 | 0 |
| POST HERPETIC NEURALGIA | 0 | 0 | 0 | 0 | 0 | 0 | 0 | 0 | 0 | 0 |
| RESTLESS ARM SYNDROME | 0 | 0 | 0 | 0 | 0 | 0 | 0 | 0 | 0 | 0 |
| RESTLESS LEGS SYNDROME | 0 | 0 | 0 | 0 | 0 | 0 | 0 | 0 | 0 | 0 |

| | | | | | | | | | | |
|---|---|---|---|---|---|---|---|---|---|---|
| SENSORY LOSS | 0 | 0 | 0 | 0 | 0 | 0 | 0 | 0 | 0 | 0 |
| TASTE DISORDER | 0 | 0 | 0 | 0 | 0 | 0 | 0 | 0 | 0 | 0 |
| *SPEECH AND LANGUAGE ABNORMALITIES* | | | | | | | | | | |
| DYSARTHRIA | 0 | 0 | 0 | 0 | 0 | 0 | 0 | 0 | 0 | 0 |
| SPEECH DISORDER DEVELOPMENTAL | 0 | 0 | 0 | 0 | 0 | 0 | 0 | 0 | 0 | 0 |
| ***NEUROLOGICAL DISORDERS OF THE EYE*** | | | | | | | | | | |
| *NEUROLOGIC VISUAL PROBLEMS NEC* | | | | | | | | | | |
| TUNNEL VISION | 0 | 0 | 0 | 0 | 0 | 0 | 0 | 0 | 0 | 0 |
| ***NEUROMUSCULAR DISORDERS*** | | | | | | | | | | |
| *MUSCLE TONE ABNORMAL* | | | | | | | | | | |
| HYPOTONIA | 0 | 0 | 0 | 0 | 0 | 0 | 0 | 0 | 0 | 0 |
| STIFF LEG SYNDROME | 0 | 0 | 0 | 0 | 0 | 0 | 0 | 0 | 0 | 0 |
| *NEUROMUSCULAR DISORDERS NEC* | | | | | | | | | | |
| MUSCLE SPASTICITY | 0 | 0 | 0 | 0 | 0 | 0 | 0 | 0 | 0 | 0 |
| ***PERIPHERAL NEUROPATHIES*** | | | | | | | | | | |
| *ACUTE POLYNEUROPATHIES* | | | | | | | | | | |
| GUILLAIN-BARRE SYNDROME | 0 | 0 | 0 | 0 | 0 | 0 | 0 | 0 | 0 | 0 |
| *PERIPHERAL NEUROPATHIES NEC* | | | | | | | | | | |
| AXONAL NEUROPATHY | 1 | 1 | 0 | 0 | 0 | 1 | 1 | 0 | 0 | 0 |
| NEUROPATHY PERIPHERAL | 0 | 0 | 0 | 0 | 0 | 0 | 0 | 0 | 0 | 0 |
| ***SEIZURES (INCL SUBTYPES)*** | | | | | | | | | | |
| *SEIZURES AND SEIZURE DISORDERS NEC* | | | | | | | | | | |
| EPILEPSY | 0 | 0 | 0 | 0 | 0 | 0 | 0 | 0 | 0 | 0 |
| SEIZURE | 0 | 0 | 0 | 0 | 0 | 0 | 0 | 0 | 0 | 0 |
| ***SLEEP DISTURBANCES (INCL SUBTYPES)*** | | | | | | | | | | |
| *DISTURBANCES IN SLEEP PHASE RHYTHM* | | | | | | | | | | |
| CIRCADIAN RHYTHM SLEEP DISORDER | 0 | 0 | 0 | 0 | 0 | 0 | 0 | 0 | 0 | 0 |
| *SLEEP DISTURBANCES NEC* | | | | | | | | | | |
| SLEEP DEFICIT | 0 | 0 | 0 | 0 | 0 | 0 | 0 | 0 | 0 | 0 |
| ***SPINAL CORD AND NERVE ROOT DISORDERS*** | | | | | | | | | | |
| *CERVICAL SPINAL CORD AND NERVE ROOT DISORDERS* | | | | | | | | | | |
| CERVICOBRACHIAL SYNDROME | 0 | 0 | 0 | 0 | 0 | 0 | 0 | 0 | 0 | 0 |
| *LUMBAR SPINAL CORD AND NERVE ROOT DISORDERS* | | | | | | | | | | |
| CAUDA EQUINA SYNDROME | 0 | 0 | 0 | 0 | 0 | 0 | 0 | 0 | 0 | 0 |
| SCIATICA | 0 | 0 | 0 | 0 | 0 | 0 | 0 | 0 | 0 | 0 |
| **PREGNANCY, PUERPERIUM AND PERINATAL CONDITIONS** | | | | | | | | | | |

| | | | | | | | | | | |
|---|---|---|---|---|---|---|---|---|---|---|
| ***ABORTIONS AND STILLBIRTH*** | | | | | | | | | | |
| *ABORTIONS SPONTANEOUS* | | | | | | | | | | |
| ABORTION SPONTANEOUS | 0 | 0 | 0 | 0 | 0 | 0 | 0 | 0 | 0 | 0 |
| *STILLBIRTH AND FOETAL DEATH* | | | | | | | | | | |
| FOETAL DEATH | 0 | 0 | 0 | 0 | 0 | 0 | 0 | 0 | 0 | 0 |
| ***FOETAL COMPLICATIONS*** | | | | | | | | | | |
| *FOETAL COMPLICATIONS NEC* | | | | | | | | | | |
| FOETAL DISORDER | 1 | 1 | 0 | 0 | 0 | 1 | 1 | 0 | 0 | 0 |
| FOETAL HYPOKINESIA | 0 | 0 | 0 | 0 | 0 | 0 | 0 | 0 | 0 | 0 |
| *FOETAL GROWTH COMPLICATIONS* | | | | | | | | | | |
| FOETAL MACROSOMIA | 0 | 0 | 0 | 0 | 0 | 0 | 0 | 0 | 0 | 0 |
| ***MATERNAL COMPLICATIONS OF PREGNANCY*** | | | | | | | | | | |
| *MATERNAL COMPLICATIONS OF PREGNANCY NEC* | | | | | | | | | | |
| MORNING SICKNESS | 0 | 0 | 0 | 0 | 0 | 0 | 0 | 0 | 0 | 0 |
| ***PLACENTAL, AMNIOTIC AND CAVITY DISORDERS (EXCL HAEMORRHAGES)*** | | | | | | | | | | |
| *PLACENTAL ABNORMALITIES (EXCL NEOPLASMS)* | | | | | | | | | | |
| PLACENTAL INFARCTION | 0 | 0 | 0 | 0 | 0 | 0 | 0 | 0 | 0 | 0 |
| ***PREGNANCY, LABOUR, DELIVERY AND POSTPARTUM CONDITIONS*** | | | | | | | | | | |
| *NORMAL PREGNANCY, LABOUR AND DELIVERY* | | | | | | | | | | |
| PREGNANCY | 0 | 0 | 0 | 0 | 0 | 0 | 0 | 0 | 0 | 0 |
| UTERINE CONTRACTIONS DURING PREGNANCY | 0 | 0 | 0 | 0 | 0 | 0 | 0 | 0 | 0 | 0 |
| **PRODUCT ISSUES** | | | | | | | | | | |
| ***DEVICE ISSUES*** | | | | | | | | | | |
| *DEVICE MALFUNCTION EVENTS NEC* | | | | | | | | | | |
| OVERSENSING | 0 | 0 | 0 | 0 | 0 | 0 | 0 | 0 | 0 | 0 |
| **PSYCHIATRIC DISORDERS** | | | | | | | | | | |
| ***ANXIETY DISORDERS AND SYMPTOMS*** | | | | | | | | | | |
| *ANXIETY SYMPTOMS* | | | | | | | | | | |
| AGITATION | 0 | 0 | 0 | 0 | 0 | 0 | 0 | 0 | 0 | 0 |
| ANXIETY | 0 | 0 | 0 | 0 | 0 | 0 | 0 | 0 | 0 | 0 |
| NERVOUSNESS | 0 | 0 | 0 | 0 | 0 | 0 | 0 | 0 | 0 | 0 |
| TENSION | 0 | 0 | 0 | 0 | 0 | 0 | 0 | 0 | 0 | 0 |
| *PANIC ATTACKS AND DISORDERS* | | | | | | | | | | |
| PANIC ATTACK | 0 | 0 | 0 | 0 | 0 | 0 | 0 | 0 | 0 | 0 |
| ***CHANGES IN PHYSICAL ACTIVITY*** | | | | | | | | | | |
| *INCREASED PHYSICAL ACTIVITY LEVELS* | | | | | | | | | | |

| | | | | | | | | | | |
|---|---|---|---|---|---|---|---|---|---|---|
| RESTLESSNESS | 0 | 0 | 0 | 0 | 0 | 0 | 0 | 0 | 0 | 0 |
| *STEREOTYPIES AND AUTOMATISMS* | | | | | | | | | | |
| BRUXISM | 0 | 0 | 0 | 0 | 0 | 0 | 0 | 0 | 0 | 0 |
| ***COGNITIVE AND ATTENTION DISORDERS AND DISTURBANCES*** | | | | | | | | | | |
| *COGNITIVE AND ATTENTION DISORDERS AND DISTURBANCES NEC* | | | | | | | | | | |
| DAYDREAMING | 0 | 0 | 0 | 0 | 0 | 0 | 0 | 0 | 0 | 0 |
| MENTAL FATIGUE | 1 | 1 | 0 | 0 | 0 | 1 | 1 | 0 | 0 | 0 |
| ***COMMUNICATION DISORDERS AND DISTURBANCES*** | | | | | | | | | | |
| *SPEECH ARTICULATION AND RHYTHM DISTURBANCES* | | | | | | | | | | |
| DYSPHEMIA | 0 | 0 | 0 | 0 | 0 | 0 | 0 | 0 | 0 | 0 |
| ***DELIRIA (INCL CONFUSION)*** | | | | | | | | | | |
| *CONFUSION AND DISORIENTATION* | | | | | | | | | | |
| CONFUSIONAL STATE | 0 | 0 | 0 | 0 | 0 | 0 | 0 | 0 | 0 | 0 |
| DISORIENTATION | 0 | 0 | 0 | 0 | 0 | 0 | 0 | 0 | 0 | 0 |
| *DELIRIA* | | | | | | | | | | |
| DELIRIUM | 0 | 0 | 0 | 0 | 0 | 0 | 0 | 0 | 0 | 0 |
| ***DEPRESSED MOOD DISORDERS AND DISTURBANCES*** | | | | | | | | | | |
| *DEPRESSIVE DISORDERS* | | | | | | | | | | |
| DEPRESSION | 0 | 0 | 0 | 0 | 0 | 0 | 0 | 0 | 0 | 0 |
| *MOOD ALTERATIONS WITH DEPRESSIVE SYMPTOMS* | | | | | | | | | | |
| DEPRESSED MOOD | 1 | 1 | 0 | 0 | 0 | 1 | 1 | 0 | 0 | 0 |
| TEARFULNESS | 0 | 0 | 0 | 0 | 0 | 0 | 0 | 0 | 0 | 0 |
| ***DISSOCIATIVE DISORDERS*** | | | | | | | | | | |
| *DISSOCIATIVE STATES* | | | | | | | | | | |
| DISSOCIATION | 0 | 0 | 0 | 0 | 0 | 0 | 0 | 0 | 0 | 0 |
| ***DISTURBANCES IN THINKING AND PERCEPTION*** | | | | | | | | | | |
| *DELUSIONAL SYMPTOMS* | | | | | | | | | | |
| DELUSION | 0 | 0 | 0 | 0 | 0 | 0 | 0 | 0 | 0 | 0 |
| *HALLUCINATIONS (EXCL SLEEP-RELATED)* | | | | | | | | | | |
| HALLUCINATION | 0 | 0 | 0 | 0 | 0 | 0 | 0 | 0 | 0 | 0 |
| *PERCEPTION DISTURBANCES NEC* | | | | | | | | | | |
| DEREALISATION | 0 | 0 | 0 | 0 | 0 | 0 | 0 | 0 | 0 | 0 |
| *THINKING DISTURBANCES* | | | | | | | | | | |
| BRADYPHRENIA | 0 | 0 | 0 | 0 | 0 | 0 | 0 | 0 | 0 | 0 |
| THOUGHT BLOCKING | 0 | 0 | 0 | 0 | 0 | 0 | 0 | 0 | 0 | 0 |
| ***MOOD DISORDERS AND DISTURBANCES NEC*** | | | | | | | | | | |

| | | | | | | | | | | |
|---|---|---|---|---|---|---|---|---|---|---|
| *AFFECT ALTERATIONS NEC* | | | | | | | | | | |
| AFFECT LABILITY | 0 | 0 | 0 | 0 | 0 | 0 | 0 | 0 | 0 | 0 |
| INAPPROPRIATE AFFECT | 0 | 0 | 0 | 0 | 0 | 0 | 0 | 0 | 0 | 0 |
| *EMOTIONAL AND MOOD DISTURBANCES NEC* | | | | | | | | | | |
| ANGER | 0 | 0 | 0 | 0 | 0 | 0 | 0 | 0 | 0 | 0 |
| EMOTIONAL DISORDER | 0 | 0 | 0 | 0 | 0 | 0 | 0 | 0 | 0 | 0 |
| EMOTIONAL DISTRESS | 0 | 0 | 0 | 0 | 0 | 0 | 0 | 0 | 0 | 0 |
| EUPHORIC MOOD | 0 | 0 | 0 | 0 | 0 | 0 | 0 | 0 | 0 | 0 |
| IRRITABILITY | 0 | 0 | 0 | 0 | 0 | 0 | 0 | 0 | 0 | 0 |
| MOOD ALTERED | 0 | 0 | 0 | 0 | 0 | 0 | 0 | 0 | 0 | 0 |
| *FLUCTUATING MOOD SYMPTOMS* | | | | | | | | | | |
| MOOD SWINGS | 0 | 0 | 0 | 0 | 0 | 0 | 0 | 0 | 0 | 0 |
| *MOOD DISORDERS NEC* | | | | | | | | | | |
| APATHY | 0 | 0 | 0 | 0 | 0 | 0 | 0 | 0 | 0 | 0 |
| LISTLESS | 0 | 0 | 0 | 0 | 0 | 0 | 0 | 0 | 0 | 0 |
| ***PERSONALITY DISORDERS AND DISTURBANCES IN BEHAVIOUR*** | | | | | | | | | | |
| *BEHAVIOUR AND SOCIALISATION DISTURBANCES* | | | | | | | | | | |
| AGGRESSION | 0 | 0 | 0 | 0 | 0 | 0 | 0 | 0 | 0 | 0 |
| ***PSYCHIATRIC AND BEHAVIOURAL SYMPTOMS NEC*** | | | | | | | | | | |
| *PSYCHIATRIC SYMPTOMS NEC* | | | | | | | | | | |
| PSYCHIATRIC SYMPTOM | 0 | 0 | 0 | 0 | 0 | 0 | 0 | 0 | 0 | 0 |
| ***PSYCHIATRIC DISORDERS NEC*** | | | | | | | | | | |
| *MENTAL DISORDERS NEC* | | | | | | | | | | |
| MENTAL DISORDER | 0 | 0 | 0 | 0 | 0 | 0 | 0 | 0 | 0 | 0 |
| ***SCHIZOPHRENIA AND OTHER PSYCHOTIC DISORDERS*** | | | | | | | | | | |
| *PSYCHOTIC DISORDER NEC* | | | | | | | | | | |
| PSYCHOTIC DISORDER | 0 | 0 | 0 | 0 | 0 | 0 | 0 | 0 | 0 | 0 |
| ***SEXUAL DYSFUNCTIONS, DISTURBANCES AND GENDER IDENTITY DISORDERS*** | | | | | | | | | | |
| *SEXUAL DESIRE DISORDERS* | | | | | | | | | | |
| LOSS OF LIBIDO | 0 | 0 | 0 | 0 | 0 | 0 | 0 | 0 | 0 | 0 |
| ***SLEEP DISORDERS AND DISTURBANCES*** | | | | | | | | | | |
| *DISTURBANCES IN INITIATING AND MAINTAINING SLEEP* | | | | | | | | | | |
| INITIAL INSOMNIA | 0 | 0 | 0 | 0 | 0 | 0 | 0 | 0 | 0 | 0 |
| INSOMNIA | 0 | 0 | 0 | 0 | 0 | 0 | 0 | 0 | 0 | 0 |
| *DYSSOMNIAS* | | | | | | | | | | |
| BREATHING-RELATED SLEEP DISORDER | 0 | 0 | 0 | 0 | 0 | 0 | 0 | 0 | 0 | 0 |

| | | | | | | | | | | |
|---|---|---|---|---|---|---|---|---|---|---|
| POOR QUALITY SLEEP | 0 | 0 | 0 | 0 | 0 | 0 | 0 | 0 | 0 | 0 |
| *PARASOMNIAS* | | | | | | | | | | |
| ABNORMAL DREAMS | 0 | 0 | 0 | 0 | 0 | 0 | 0 | 0 | 0 | 0 |
| EXPLODING HEAD SYNDROME | 0 | 0 | 0 | 0 | 0 | 0 | 0 | 0 | 0 | 0 |
| NIGHTMARE | 0 | 0 | 0 | 0 | 0 | 0 | 0 | 0 | 0 | 0 |
| *SLEEP DISORDERS NEC* | | | | | | | | | | |
| SLEEP DISORDER | 0 | 0 | 0 | 0 | 0 | 0 | 0 | 0 | 0 | 0 |
| ***SOMATIC SYMPTOM AND RELATED DISORDERS*** | | | | | | | | | | |
| *SOMATIC SYMPTOM DISORDERS* | | | | | | | | | | |
| HABIT COUGH | 0 | 0 | 0 | 0 | 0 | 0 | 0 | 0 | 0 | 0 |
| ***SUICIDAL AND SELF-INJURIOUS BEHAVIOURS NEC*** | | | | | | | | | | |
| *SUICIDAL AND SELF-INJURIOUS BEHAVIOUR* | | | | | | | | | | |
| SUICIDAL IDEATION | 0 | 0 | 0 | 0 | 0 | 0 | 0 | 0 | 0 | 0 |
| **RENAL AND URINARY DISORDERS** | | | | | | | | | | |
| ***RENAL DISORDERS (EXCL NEPHROPATHIES)*** | | | | | | | | | | |
| *RENAL FAILURE AND IMPAIRMENT* | | | | | | | | | | |
| RENAL FAILURE | 0 | 0 | 0 | 0 | 0 | 0 | 0 | 0 | 0 | 0 |
| ***URINARY TRACT SIGNS AND SYMPTOMS*** | | | | | | | | | | |
| *BLADDER AND URETHRAL SYMPTOMS* | | | | | | | | | | |
| BLADDER PAIN | 0 | 0 | 0 | 0 | 0 | 0 | 0 | 0 | 0 | 0 |
| MICTURITION URGENCY | 0 | 0 | 0 | 0 | 0 | 0 | 0 | 0 | 0 | 0 |
| POLLAKIURIA | 0 | 0 | 0 | 0 | 0 | 0 | 0 | 0 | 0 | 0 |
| URINARY INCONTINENCE | 0 | 0 | 0 | 0 | 0 | 0 | 0 | 0 | 0 | 0 |
| *URINARY ABNORMALITIES* | | | | | | | | | | |
| HAEMATURIA | 0 | 0 | 0 | 0 | 0 | 0 | 0 | 0 | 0 | 0 |
| URINE ABNORMALITY | 0 | 0 | 0 | 0 | 0 | 0 | 0 | 0 | 0 | 0 |
| URINE ODOUR ABNORMAL | 0 | 0 | 0 | 0 | 0 | 0 | 0 | 0 | 0 | 0 |
| *URINARY TRACT SIGNS AND SYMPTOMS NEC* | | | | | | | | | | |
| POLYURIA | 0 | 0 | 0 | 0 | 0 | 0 | 0 | 0 | 0 | 0 |
| RENAL PAIN | 0 | 0 | 0 | 0 | 0 | 0 | 0 | 0 | 0 | 0 |
| **REPRODUCTIVE SYSTEM AND BREAST DISORDERS** | | | | | | | | | | |
| ***BREAST DISORDERS*** | | | | | | | | | | |
| *BREAST DISORDERS NEC* | | | | | | | | | | |
| BREAST MASS | 0 | 0 | 0 | 0 | 0 | 0 | 0 | 0 | 0 | 0 |
| NIPPLE ENLARGEMENT | 0 | 0 | 0 | 0 | 0 | 0 | 0 | 0 | 0 | 0 |
| *BREAST SIGNS AND SYMPTOMS* | | | | | | | | | | |

| | | | | | | | | | | |
|---|---|---|---|---|---|---|---|---|---|---|
| BREAST DISCHARGE | 0 | 0 | 0 | 0 | 0 | 0 | 0 | 0 | 0 | 0 |
| BREAST PAIN | 0 | 0 | 0 | 0 | 0 | 0 | 0 | 0 | 0 | 0 |
| BREAST SWELLING | 0 | 0 | 0 | 0 | 0 | 0 | 0 | 0 | 0 | 0 |
| BREAST TENDERNESS | 0 | 0 | 0 | 0 | 0 | 0 | 0 | 0 | 0 | 0 |
| NIPPLE PAIN | 0 | 0 | 0 | 0 | 0 | 0 | 0 | 0 | 0 | 0 |
| ***MENOPAUSE RELATED CONDITIONS*** | | | | | | | | | | |
| *MENOPAUSAL EFFECTS ON THE GENITOURINARY TRACT* | | | | | | | | | | |
| POSTMENOPAUSAL HAEMORRHAGE | 0 | 0 | 0 | 0 | 0 | 0 | 0 | 0 | 0 | 0 |
| ***MENSTRUAL CYCLE AND UTERINE BLEEDING DISORDERS*** | | | | | | | | | | |
| *MENSTRUATION AND UTERINE BLEEDING NEC* | | | | | | | | | | |
| DYSMENORRHOEA | 0 | 0 | 0 | 0 | 0 | 0 | 0 | 0 | 0 | 0 |
| INTERMENSTRUAL BLEEDING | 0 | 0 | 0 | 0 | 0 | 0 | 0 | 0 | 0 | 0 |
| MENSTRUAL DISORDER | 0 | 0 | 0 | 0 | 0 | 0 | 0 | 0 | 0 | 0 |
| MENSTRUATION IRREGULAR | 0 | 0 | 0 | 0 | 0 | 0 | 0 | 0 | 0 | 0 |
| PREMENSTRUAL PAIN | 0 | 0 | 0 | 0 | 0 | 0 | 0 | 0 | 0 | 0 |
| RETROGRADE MENSTRUATION | 0 | 0 | 0 | 0 | 0 | 0 | 0 | 0 | 0 | 0 |
| *MENSTRUATION WITH DECREASED BLEEDING* | | | | | | | | | | |
| AMENORRHOEA | 0 | 0 | 0 | 0 | 0 | 0 | 0 | 0 | 0 | 0 |
| HYPOMENORRHOEA | 0 | 0 | 0 | 0 | 0 | 0 | 0 | 0 | 0 | 0 |
| MENSTRUATION DELAYED | 0 | 0 | 0 | 0 | 0 | 0 | 0 | 0 | 0 | 0 |
| OLIGOMENORRHOEA | 0 | 0 | 0 | 0 | 0 | 0 | 0 | 0 | 0 | 0 |
| *MENSTRUATION WITH INCREASED BLEEDING* | | | | | | | | | | |
| HEAVY MENSTRUAL BLEEDING | 1 | 0 | 1 | 0 | 0 | 1 | 0 | 1 | 0 | 0 |
| POLYMENORRHOEA | 0 | 0 | 0 | 0 | 0 | 0 | 0 | 0 | 0 | 0 |
| ***PENILE AND SCROTAL DISORDERS (EXCL INFECTIONS AND INFLAMMATIONS)*** | | | | | | | | | | |
| *SCROTAL DISORDERS NEC* | | | | | | | | | | |
| SCROTAL SWELLING | 0 | 0 | 0 | 0 | 0 | 0 | 0 | 0 | 0 | 0 |
| ***REPRODUCTIVE TRACT DISORDERS NEC*** | | | | | | | | | | |
| *REPRODUCTIVE TRACT DISORDERS NEC (EXCL NEOPLASMS)* | | | | | | | | | | |
| GENITAL LESION | 0 | 0 | 0 | 0 | 0 | 0 | 0 | 0 | 0 | 0 |
| *REPRODUCTIVE TRACT SIGNS AND SYMPTOMS NEC* | | | | | | | | | | |
| GENITAL DISCOMFORT | 0 | 0 | 0 | 0 | 0 | 0 | 0 | 0 | 0 | 0 |
| GENITAL PAIN | 0 | 0 | 0 | 0 | 0 | 0 | 0 | 0 | 0 | 0 |
| ***UTERINE, PELVIC AND BROAD LIGAMENT DISORDERS*** | | | | | | | | | | |
| *PELVIS AND BROAD LIGAMENT DISORDERS NEC* | | | | | | | | | | |
| ADNEXA UTERI PAIN | 0 | 0 | 0 | 0 | 0 | 0 | 0 | 0 | 0 | 0 |

| | | | | | | | | | | |
|---|---|---|---|---|---|---|---|---|---|---|
| ***VULVOVAGINAL DISORDERS (EXCL INFECTIONS AND INFLAMMATIONS)*** | | | | | | | | | | |
| *VULVOVAGINAL DISORDERS NEC* | | | | | | | | | | |
| VAGINAL HAEMORRHAGE | 0 | 0 | 0 | 0 | 0 | 0 | 0 | 0 | 0 | 0 |
| *VULVOVAGINAL SIGNS AND SYMPTOMS* | | | | | | | | | | |
| VAGINAL DISCHARGE | 0 | 0 | 0 | 0 | 0 | 0 | 0 | 0 | 0 | 0 |
| **RESPIRATORY, THORACIC AND MEDIASTINAL DISORDERS** | | | | | | | | | | |
| ***BRONCHIAL DISORDERS (EXCL NEOPLASMS)*** | | | | | | | | | | |
| *BRONCHOSPASM AND OBSTRUCTION* | | | | | | | | | | |
| ASTHMA | 0 | 0 | 0 | 0 | 0 | 0 | 0 | 0 | 0 | 0 |
| WHEEZING | 0 | 0 | 0 | 0 | 0 | 0 | 0 | 0 | 0 | 0 |
| ***LOWER RESPIRATORY TRACT DISORDERS (EXCL OBSTRUCTION AND INFECTION)*** | | | | | | | | | | |
| *PULMONARY OEDEMAS* | | | | | | | | | | |
| PULMONARY CONGESTION | 0 | 0 | 0 | 0 | 0 | 0 | 0 | 0 | 0 | 0 |
| ***PULMONARY VASCULAR DISORDERS*** | | | | | | | | | | |
| *PULMONARY THROMBOTIC AND EMBOLIC CONDITIONS* | | | | | | | | | | |
| PULMONARY EMBOLISM | 0 | 0 | 0 | 0 | 0 | 0 | 0 | 0 | 0 | 0 |
| ***RESPIRATORY DISORDERS NEC*** | | | | | | | | | | |
| *BREATHING ABNORMALITIES* | | | | | | | | | | |
| DYSPNOEA | 1 | 1 | 0 | 0 | 0 | 1 | 1 | 0 | 0 | 0 |
| HYPERVENTILATION | 0 | 0 | 0 | 0 | 0 | 0 | 0 | 0 | 0 | 0 |
| HYPOPNOEA | 0 | 0 | 0 | 0 | 0 | 0 | 0 | 0 | 0 | 0 |
| IRREGULAR BREATHING | 0 | 0 | 0 | 0 | 0 | 0 | 0 | 0 | 0 | 0 |
| MOUTH BREATHING | 0 | 0 | 0 | 0 | 0 | 0 | 0 | 0 | 0 | 0 |
| RESPIRATION ABNORMAL | 0 | 0 | 0 | 0 | 0 | 0 | 0 | 0 | 0 | 0 |
| RESPIRATORY ARREST | 0 | 0 | 0 | 0 | 0 | 0 | 0 | 0 | 0 | 0 |
| RESPIRATORY FATIGUE | 0 | 0 | 0 | 0 | 0 | 0 | 0 | 0 | 0 | 0 |
| SLEEP APNOEA SYNDROME | 0 | 0 | 0 | 0 | 0 | 0 | 0 | 0 | 0 | 0 |
| *COUGHING AND ASSOCIATED SYMPTOMS* | | | | | | | | | | |
| COUGH | 1 | 1 | 0 | 0 | 0 | 1 | 1 | 0 | 0 | 0 |
| PRODUCTIVE COUGH | 0 | 0 | 0 | 0 | 0 | 0 | 0 | 0 | 0 | 0 |
| *RESPIRATORY TRACT DISORDERS NEC* | | | | | | | | | | |
| RESPIRATORY TRACT IRRITATION | 0 | 0 | 0 | 0 | 0 | 0 | 0 | 0 | 0 | 0 |
| ***RESPIRATORY TRACT SIGNS AND SYMPTOMS*** | | | | | | | | | | |
| *LOWER RESPIRATORY TRACT SIGNS AND SYMPTOMS* | | | | | | | | | | |
| PULMONARY PAIN | 0 | 0 | 0 | 0 | 0 | 0 | 0 | 0 | 0 | 0 |
| *RESPIRATORY SIGNS AND SYMPTOMS NEC* | | | | | | | | | | |

| | | | | | | | | | | |
|---|---|---|---|---|---|---|---|---|---|---|
| RESPIRATORY SYMPTOM | 0 | 0 | 0 | 0 | 0 | 0 | 0 | 0 | 0 | 0 |
| *UPPER RESPIRATORY TRACT SIGNS AND SYMPTOMS* | | | | | | | | | | |
| APHONIA | 0 | 0 | 0 | 0 | 0 | 0 | 0 | 0 | 0 | 0 |
| CATARRH | 0 | 0 | 0 | 0 | 0 | 0 | 0 | 0 | 0 | 0 |
| DRY THROAT | 0 | 0 | 0 | 0 | 0 | 0 | 0 | 0 | 0 | 0 |
| DYSPHONIA | 0 | 0 | 0 | 0 | 0 | 0 | 0 | 0 | 0 | 0 |
| INCREASED UPPER AIRWAY SECRETION | 0 | 0 | 0 | 0 | 0 | 0 | 0 | 0 | 0 | 0 |
| INCREASED VISCOSITY OF UPPER RESPIRATORY SE( | 0 | 0 | 0 | 0 | 0 | 0 | 0 | 0 | 0 | 0 |
| NASAL DISCOMFORT | 0 | 0 | 0 | 0 | 0 | 0 | 0 | 0 | 0 | 0 |
| OROPHARYNGEAL DISCOMFORT | 0 | 0 | 0 | 0 | 0 | 0 | 0 | 0 | 0 | 0 |
| OROPHARYNGEAL PAIN | 1 | 1 | 0 | 0 | 0 | 1 | 1 | 0 | 0 | 0 |
| PARANASAL SINUS DISCOMFORT | 0 | 0 | 0 | 0 | 0 | 0 | 0 | 0 | 0 | 0 |
| RHINALGIA | 1 | 1 | 0 | 0 | 0 | 1 | 1 | 0 | 0 | 0 |
| RHINORRHOEA | 1 | 0 | 0 | 1 | 0 | 1 | 0 | 0 | 1 | 0 |
| SINUS PAIN | 1 | 0 | 0 | 1 | 0 | 1 | 0 | 0 | 1 | 0 |
| SNEEZING | 0 | 0 | 0 | 0 | 0 | 0 | 0 | 0 | 0 | 0 |
| THROAT CLEARING | 0 | 0 | 0 | 0 | 0 | 0 | 0 | 0 | 0 | 0 |
| THROAT IRRITATION | 0 | 0 | 0 | 0 | 0 | 0 | 0 | 0 | 0 | 0 |
| THROAT TIGHTNESS | 0 | 0 | 0 | 0 | 0 | 0 | 0 | 0 | 0 | 0 |
| UPPER-AIRWAY COUGH SYNDROME | 0 | 0 | 0 | 0 | 0 | 0 | 0 | 0 | 0 | 0 |
| YAWNING | 0 | 0 | 0 | 0 | 0 | 0 | 0 | 0 | 0 | 0 |
| ***UPPER RESPIRATORY TRACT DISORDERS (EXCL INFECTIONS)*** | | | | | | | | | | |
| *NASAL CONGESTION AND INFLAMMATIONS* | | | | | | | | | | |
| NASAL CONGESTION | 0 | 0 | 0 | 0 | 0 | 0 | 0 | 0 | 0 | 0 |
| RHINITIS ALLERGIC | 0 | 0 | 0 | 0 | 0 | 0 | 0 | 0 | 0 | 0 |
| RHINITIS ATROPHIC | 0 | 0 | 0 | 0 | 0 | 0 | 0 | 0 | 0 | 0 |
| *NASAL DISORDERS NEC* | | | | | | | | | | |
| EPISTAXIS | 1 | 0 | 1 | 0 | 0 | 1 | 0 | 1 | 0 | 0 |
| NASAL DRYNESS | 0 | 0 | 0 | 0 | 0 | 0 | 0 | 0 | 0 | 0 |
| *PARANASAL SINUS DISORDERS (EXCL INFECTIONS AND NEOPLASMS)* | | | | | | | | | | |
| SINUS CONGESTION | 0 | 0 | 0 | 0 | 0 | 0 | 0 | 0 | 0 | 0 |
| *PHARYNGEAL DISORDERS (EXCL INFECTIONS AND NEOPLASMS)* | | | | | | | | | | |
| PHARYNGEAL SWELLING | 0 | 0 | 0 | 0 | 0 | 0 | 0 | 0 | 0 | 0 |
| PHARYNGEAL ULCERATION | 0 | 0 | 0 | 0 | 0 | 0 | 0 | 0 | 0 | 0 |
| TONSILLAR ERYTHEMA | 0 | 0 | 0 | 0 | 0 | 0 | 0 | 0 | 0 | 0 |

| | | | | | | | | | | |
|---|---|---|---|---|---|---|---|---|---|---|
| *TRACHEAL DISORDERS (EXCL INFECTIONS AND NEOPLASMS)* | | | | | | | | | | |
| TRACHEAL PAIN | 0 | 0 | 0 | 0 | 0 | 0 | 0 | 0 | 0 | 0 |
| **SKIN AND SUBCUTANEOUS TISSUE DISORDERS** | | | | | | | | | | |
| ***ANGIOEDEMA AND URTICARIA*** | | | | | | | | | | |
| *ANGIOEDEMAS* | | | | | | | | | | |
| ANGIOEDEMA | 0 | 0 | 0 | 0 | 0 | 0 | 0 | 0 | 0 | 0 |
| *URTICARIAS* | | | | | | | | | | |
| COLD URTICARIA | 0 | 0 | 0 | 0 | 0 | 0 | 0 | 0 | 0 | 0 |
| SOLAR URTICARIA | 0 | 0 | 0 | 0 | 0 | 0 | 0 | 0 | 0 | 0 |
| URTICARIA | 0 | 0 | 0 | 0 | 0 | 0 | 0 | 0 | 0 | 0 |
| URTICARIA CHRONIC | 0 | 0 | 0 | 0 | 0 | 0 | 0 | 0 | 0 | 0 |
| ***CORNIFICATION AND DYSTROPHIC SKIN DISORDERS*** | | | | | | | | | | |
| *SKIN DYSTROPHIES* | | | | | | | | | | |
| HYPERTROPHIC SCAR | 0 | 0 | 0 | 0 | 0 | 0 | 0 | 0 | 0 | 0 |
| ***CUTANEOUS NEOPLASMS BENIGN*** | | | | | | | | | | |
| *SKIN CYSTS AND POLYPS* | | | | | | | | | | |
| DERMAL CYST | 0 | 0 | 0 | 0 | 0 | 0 | 0 | 0 | 0 | 0 |
| ***EPIDERMAL AND DERMAL CONDITIONS*** | | | | | | | | | | |
| *BULLOUS CONDITIONS* | | | | | | | | | | |
| BLISTER | 0 | 0 | 0 | 0 | 0 | 0 | 0 | 0 | 0 | 0 |
| BLOOD BLISTER | 0 | 0 | 0 | 0 | 0 | 0 | 0 | 0 | 0 | 0 |
| PEMPHIGOID | 0 | 0 | 0 | 0 | 0 | 0 | 0 | 0 | 0 | 0 |
| TOXIC EPIDERMAL NECROLYSIS | 0 | 0 | 0 | 0 | 0 | 0 | 0 | 0 | 0 | 0 |
| *DERMAL AND EPIDERMAL CONDITIONS NEC* | | | | | | | | | | |
| DRY SKIN | 0 | 0 | 0 | 0 | 0 | 0 | 0 | 0 | 0 | 0 |
| PAIN OF SKIN | 0 | 0 | 0 | 0 | 0 | 0 | 0 | 0 | 0 | 0 |
| SENSITIVE SKIN | 0 | 0 | 0 | 0 | 0 | 0 | 0 | 0 | 0 | 0 |
| SKIN BURNING SENSATION | 0 | 0 | 0 | 0 | 0 | 0 | 0 | 0 | 0 | 0 |
| SKIN DISCOLOURATION | 0 | 0 | 0 | 0 | 0 | 0 | 0 | 0 | 0 | 0 |
| SKIN FRAGILITY | 0 | 0 | 0 | 0 | 0 | 0 | 0 | 0 | 0 | 0 |
| SKIN LESION | 0 | 0 | 0 | 0 | 0 | 0 | 0 | 0 | 0 | 0 |
| SKIN ODOUR ABNORMAL | 0 | 0 | 0 | 0 | 0 | 0 | 0 | 0 | 0 | 0 |
| SKIN REACTION | 0 | 0 | 0 | 0 | 0 | 0 | 0 | 0 | 0 | 0 |
| SKIN SENSITISATION | 0 | 0 | 0 | 0 | 0 | 0 | 0 | 0 | 0 | 0 |
| SKIN WARM | 0 | 0 | 0 | 0 | 0 | 0 | 0 | 0 | 0 | 0 |
| *DERMATITIS AND ECZEMA* | | | | | | | | | | |

| | | | | | | | | | | |
|---|---|---|---|---|---|---|---|---|---|---|
| DERMATITIS | 0 | 0 | 0 | 0 | 0 | 0 | 0 | 0 | 0 | 0 |
| DERMATITIS ALLERGIC | 1 | 1 | 0 | 0 | 0 | 1 | 1 | 0 | 0 | 0 |
| DERMATITIS ATOPIC | 0 | 0 | 0 | 0 | 0 | 0 | 0 | 0 | 0 | 0 |
| DERMATITIS CONTACT | 0 | 0 | 0 | 0 | 0 | 0 | 0 | 0 | 0 | 0 |
| ECZEMA | 0 | 0 | 0 | 0 | 0 | 0 | 0 | 0 | 0 | 0 |
| ECZEMA ASTEATOTIC | 0 | 0 | 0 | 0 | 0 | 0 | 0 | 0 | 0 | 0 |
| NEURODERMATITIS | 0 | 0 | 0 | 0 | 0 | 0 | 0 | 0 | 0 | 0 |
| SEBORRHOEIC DERMATITIS | 0 | 0 | 0 | 0 | 0 | 0 | 0 | 0 | 0 | 0 |
| SKIN IRRITATION | 1 | 1 | 0 | 0 | 0 | 1 | 1 | 0 | 0 | 0 |
| *ERYTHEMAS* | | | | | | | | | | |
| ERYTHEMA | 0 | 0 | 0 | 0 | 0 | 0 | 0 | 0 | 0 | 0 |
| *EXFOLIATIVE CONDITIONS* | | | | | | | | | | |
| SKIN EXFOLIATION | 0 | 0 | 0 | 0 | 0 | 0 | 0 | 0 | 0 | 0 |
| *PAPULOSQUAMOUS CONDITIONS* | | | | | | | | | | |
| LICHEN PLANUS | 0 | 0 | 0 | 0 | 0 | 0 | 0 | 0 | 0 | 0 |
| PITYRIASIS ROSEA | 0 | 0 | 0 | 0 | 0 | 0 | 0 | 0 | 0 | 0 |
| *PHOTOSENSITIVITY AND PHOTODERMATOSIS CONDITIONS* | | | | | | | | | | |
| PHOTOSENSITIVITY REACTION | 0 | 0 | 0 | 0 | 0 | 0 | 0 | 0 | 0 | 0 |
| *PRURITUS NEC* | | | | | | | | | | |
| PRURITUS | 0 | 0 | 0 | 0 | 0 | 0 | 0 | 0 | 0 | 0 |
| *PSORIATIC CONDITIONS* | | | | | | | | | | |
| PSORIASIS | 0 | 0 | 0 | 0 | 0 | 0 | 0 | 0 | 0 | 0 |
| *RASHES, ERUPTIONS AND EXANTHEMS NEC* | | | | | | | | | | |
| RASH | 2 | 1 | 1 | 0 | 0 | 2 | 1 | 1 | 0 | 0 |
| RASH ERYTHEMATOUS | 0 | 0 | 0 | 0 | 0 | 0 | 0 | 0 | 0 | 0 |
| RASH MACULAR | 0 | 0 | 0 | 0 | 0 | 0 | 0 | 0 | 0 | 0 |
| RASH MORBILLIFORM | 0 | 0 | 0 | 0 | 0 | 0 | 0 | 0 | 0 | 0 |
| RASH PAPULAR | 0 | 0 | 0 | 0 | 0 | 0 | 0 | 0 | 0 | 0 |
| RASH PRURITIC | 2 | 1 | 0 | 0 | 1 | 1 | 1 | 0 | 0 | 0 |
| *SKIN INJURIES AND MECHANICAL DERMATOSES* | | | | | | | | | | |
| DECUBITUS ULCER | 0 | 0 | 0 | 0 | 0 | 0 | 0 | 0 | 0 | 0 |
| NEEDLE TRACK MARKS | 0 | 0 | 0 | 0 | 0 | 0 | 0 | 0 | 0 | 0 |
| ***SKIN AND SUBCUTANEOUS TISSUE DISORDERS NEC*** | | | | | | | | | | |
| *SKIN AND SUBCUTANEOUS TISSUE ULCERATIONS* | | | | | | | | | | |
| SKIN EROSION | 0 | 0 | 0 | 0 | 0 | 0 | 0 | 0 | 0 | 0 |

| | | | | | | | | | | |
|---|---|---|---|---|---|---|---|---|---|---|
| ***SKIN APPENDAGE CONDITIONS*** | | | | | | | | | | |
| *ACNES* | | | | | | | | | | |
| ACNE | 0 | 0 | 0 | 0 | 0 | 0 | 0 | 0 | 0 | 0 |
| ACNE CYSTIC | 0 | 0 | 0 | 0 | 0 | 0 | 0 | 0 | 0 | 0 |
| *ALOPECIAS* | | | | | | | | | | |
| ALOPECIA | 0 | 0 | 0 | 0 | 0 | 0 | 0 | 0 | 0 | 0 |
| *APOCRINE AND ECCRINE GLAND DISORDERS* | | | | | | | | | | |
| COLD SWEAT | 1 | 1 | 0 | 0 | 0 | 1 | 1 | 0 | 0 | 0 |
| HYPERHIDROSIS | 2 | 2 | 0 | 0 | 0 | 2 | 2 | 0 | 0 | 0 |
| MILIARIA | 0 | 0 | 0 | 0 | 0 | 0 | 0 | 0 | 0 | 0 |
| NIGHT SWEATS | 0 | 0 | 0 | 0 | 0 | 0 | 0 | 0 | 0 | 0 |
| *HYPERTRICHOSES* | | | | | | | | | | |
| HIRSUTISM | 0 | 0 | 0 | 0 | 0 | 0 | 0 | 0 | 0 | 0 |
| *NAIL AND NAIL BED CONDITIONS (EXCL INFECTIONS AND INFESTATIONS)* | | | | | | | | | | |
| NAIL DISCOLOURATION | 0 | 0 | 0 | 0 | 0 | 0 | 0 | 0 | 0 | 0 |
| ONYCHOCLASIS | 0 | 0 | 0 | 0 | 0 | 0 | 0 | 0 | 0 | 0 |
| *PILAR DISORDERS NEC* | | | | | | | | | | |
| PILOERECTION | 0 | 0 | 0 | 0 | 0 | 0 | 0 | 0 | 0 | 0 |
| ***SKIN VASCULAR ABNORMALITIES*** | | | | | | | | | | |
| *PURPURA AND RELATED CONDITIONS* | | | | | | | | | | |
| PETECHIAE | 0 | 0 | 0 | 0 | 0 | 0 | 0 | 0 | 0 | 0 |
| PURPURA | 0 | 0 | 0 | 0 | 0 | 0 | 0 | 0 | 0 | 0 |
| *SKIN HAEMORRHAGES* | | | | | | | | | | |
| SKIN HAEMORRHAGE | 0 | 0 | 0 | 0 | 0 | 0 | 0 | 0 | 0 | 0 |
| **SOCIAL CIRCUMSTANCES** | | | | | | | | | | |
| ***ECONOMIC AND HOUSING ISSUES*** | | | | | | | | | | |
| *EMPLOYMENT ISSUES* | | | | | | | | | | |
| RETIREMENT | 0 | 0 | 0 | 0 | 0 | 0 | 0 | 0 | 0 | 0 |
| ***ENVIRONMENTAL ISSUES*** | | | | | | | | | | |
| *NON-OCCUPATIONAL AND UNSPECIFIED ENVIRONMENTAL PROBLEMS* | | | | | | | | | | |
| POLLUTION | 0 | 0 | 0 | 0 | 0 | 0 | 0 | 0 | 0 | 0 |
| **SURGICAL AND MEDICAL PROCEDURES** | | | | | | | | | | |
| ***BONE AND JOINT THERAPEUTIC PROCEDURES*** | | | | | | | | | | |
| *JOINT THERAPEUTIC PROCEDURES* | | | | | | | | | | |
| KNEE OPERATION | 0 | 0 | 0 | 0 | 0 | 0 | 0 | 0 | 0 | 0 |
| ***BREAST THERAPEUTIC PROCEDURES*** | | | | | | | | | | |

| | | | | | | | | | | |
|---|---|---|---|---|---|---|---|---|---|---|
| *MASTECTOMIES* | | | | | | | | | | |
| BREAST CONSERVING SURGERY | 0 | 0 | 0 | 0 | 0 | 0 | 0 | 0 | 0 | 0 |
| ***GASTROINTESTINAL THERAPEUTIC PROCEDURES*** | | | | | | | | | | |
| *GASTROINTESTINAL THERAPEUTIC PROCEDURES NEC* | | | | | | | | | | |
| PROPHYLAXIS OF NAUSEA AND VOMITING | 0 | 0 | 0 | 0 | 0 | 0 | 0 | 0 | 0 | 0 |
| ***HEAD AND NECK THERAPEUTIC PROCEDURES*** | | | | | | | | | | |
| *DENTAL AND GINGIVAL THERAPEUTIC PROCEDURES* | | | | | | | | | | |
| DENTAL CARE | 0 | 0 | 0 | 0 | 0 | 0 | 0 | 0 | 0 | 0 |
| ***NERVOUS SYSTEM, SKULL AND SPINE THERAPEUTIC PROCEDURES*** | | | | | | | | | | |
| *SKULL AND BRAIN THERAPEUTIC PROCEDURES* | | | | | | | | | | |
| BRAIN TUMOUR OPERATION | 0 | 0 | 0 | 0 | 0 | 0 | 0 | 0 | 0 | 0 |
| ***OBSTETRIC AND GYNAECOLOGICAL THERAPEUTIC PROCEDURES*** | | | | | | | | | | |
| *FERTILITY AND FERTILISATION INTERVENTIONS FEMALE* | | | | | | | | | | |
| ENDOMETRIAL SCRATCHING | 0 | 0 | 0 | 0 | 0 | 0 | 0 | 0 | 0 | 0 |
| ***RESPIRATORY TRACT THERAPEUTIC PROCEDURES*** | | | | | | | | | | |
| *RESPIRATORY TRACT THERAPEUTIC PROCEDURES NEC* | | | | | | | | | | |
| OXYGEN THERAPY | 0 | 0 | 0 | 0 | 0 | 0 | 0 | 0 | 0 | 0 |
| ***THERAPEUTIC PROCEDURES AND SUPPORTIVE CARE NEC*** | | | | | | | | | | |
| *ANAESTHESIA AND ALLIED PROCEDURES* | | | | | | | | | | |
| NERVE BLOCK | 0 | 0 | 0 | 0 | 0 | 0 | 0 | 0 | 0 | 0 |
| *DIETARY AND NUTRITIONAL THERAPIES* | | | | | | | | | | |
| NOTHING BY MOUTH ORDER | 0 | 0 | 0 | 0 | 0 | 0 | 0 | 0 | 0 | 0 |
| *IMMUNISATIONS* | | | | | | | | | | |
| COVID-19 IMMUNISATION | 0 | 0 | 0 | 0 | 0 | 0 | 0 | 0 | 0 | 0 |
| *THERAPEUTIC PROCEDURES NEC* | | | | | | | | | | |
| INJECTION | 0 | 0 | 0 | 0 | 0 | 0 | 0 | 0 | 0 | 0 |
| LOCALISED ALTERNATING HOT AND COLD THERAPY | 0 | 0 | 0 | 0 | 0 | 0 | 0 | 0 | 0 | 0 |
| MASS EXCISION | 0 | 0 | 0 | 0 | 0 | 0 | 0 | 0 | 0 | 0 |
| REINFUSION | 0 | 0 | 0 | 0 | 0 | 0 | 0 | 0 | 0 | 0 |
| **VASCULAR DISORDERS** | | | | | | | | | | |
| ***ARTERIOSCLEROSIS, STENOSIS, VASCULAR INSUFFICIENCY AND NECROSIS*** | | | | | | | | | | |
| *NON-SITE SPECIFIC NECROSIS AND VASCULAR INSUFFICIENCY NEC* | | | | | | | | | | |
| VASOSPASM | 0 | 0 | 0 | 0 | 0 | 0 | 0 | 0 | 0 | 0 |
| *PERIPHERAL VASOCONSTRICTION, NECROSIS AND VASCULAR INSUFFICIENCY* | | | | | | | | | | |
| PERIPHERAL COLDNESS | 0 | 0 | 0 | 0 | 0 | 0 | 0 | 0 | 0 | 0 |

| | | | | | | | | | | |
|---|---|---|---|---|---|---|---|---|---|---|
| RAYNAUD'S PHENOMENON | 0 | 0 | 0 | 0 | 0 | 0 | 0 | 0 | 0 | 0 |
| ***DECREASED AND NONSPECIFIC BLOOD PRESSURE DISORDERS AND SHOCK*** | | | | | | | | | | |
| *BLOOD PRESSURE DISORDERS NEC* | | | | | | | | | | |
| BLOOD PRESSURE FLUCTUATION | 0 | 0 | 0 | 0 | 0 | 0 | 0 | 0 | 0 | 0 |
| *VASCULAR HYPOTENSIVE DISORDERS* | | | | | | | | | | |
| CAPILLARY LEAK SYNDROME | 0 | 0 | 0 | 0 | 0 | 0 | 0 | 0 | 0 | 0 |
| HYPOTENSION | 0 | 0 | 0 | 0 | 0 | 0 | 0 | 0 | 0 | 0 |
| ORTHOSTATIC HYPOTENSION | 0 | 0 | 0 | 0 | 0 | 0 | 0 | 0 | 0 | 0 |
| ***EMBOLISM AND THROMBOSIS*** | | | | | | | | | | |
| *NON-SITE SPECIFIC EMBOLISM AND THROMBOSIS* | | | | | | | | | | |
| EMBOLISM | 0 | 0 | 0 | 0 | 0 | 0 | 0 | 0 | 0 | 0 |
| THROMBOSIS | 0 | 0 | 0 | 0 | 0 | 0 | 0 | 0 | 0 | 0 |
| VENOUS THROMBOSIS | 0 | 0 | 0 | 0 | 0 | 0 | 0 | 0 | 0 | 0 |
| *PERIPHERAL EMBOLISM AND THROMBOSIS* | | | | | | | | | | |
| BLUE TOE SYNDROME | 0 | 0 | 0 | 0 | 0 | 0 | 0 | 0 | 0 | 0 |
| DEEP VEIN THROMBOSIS | 0 | 0 | 0 | 0 | 0 | 0 | 0 | 0 | 0 | 0 |
| SUPERFICIAL VEIN THROMBOSIS | 0 | 0 | 0 | 0 | 0 | 0 | 0 | 0 | 0 | 0 |
| ***LYMPHATIC VESSEL DISORDERS*** | | | | | | | | | | |
| *LYMPHOEDEMAS* | | | | | | | | | | |
| LYMPHOEDEMA | 0 | 0 | 0 | 0 | 0 | 0 | 0 | 0 | 0 | 0 |
| ***VASCULAR DISORDERS NEC*** | | | | | | | | | | |
| *NON-SITE SPECIFIC VASCULAR DISORDERS NEC* | | | | | | | | | | |
| VASCULAR PAIN | 0 | 0 | 0 | 0 | 0 | 0 | 0 | 0 | 0 | 0 |
| VEIN DISCOLOURATION | 0 | 0 | 0 | 0 | 0 | 0 | 0 | 0 | 0 | 0 |
| VEIN RUPTURE | 0 | 0 | 0 | 0 | 0 | 0 | 0 | 0 | 0 | 0 |
| *PERIPHERAL VASCULAR DISORDERS NEC* | | | | | | | | | | |
| FLUSHING | 0 | 0 | 0 | 0 | 0 | 0 | 0 | 0 | 0 | 0 |
| HOT FLUSH | 2 | 1 | 0 | 1 | 0 | 2 | 1 | 0 | 1 | 0 |
| *SITE SPECIFIC VASCULAR DISORDERS NEC* | | | | | | | | | | |
| PALLOR | 0 | 0 | 0 | 0 | 0 | 0 | 0 | 0 | 0 | 0 |
| ***VASCULAR HAEMORRHAGIC DISORDERS*** | | | | | | | | | | |
| *HAEMORRHAGES NEC* | | | | | | | | | | |
| HAEMATOMA | 0 | 0 | 0 | 0 | 0 | 0 | 0 | 0 | 0 | 0 |
| HAEMORRHAGE | 0 | 0 | 0 | 0 | 0 | 0 | 0 | 0 | 0 | 0 |
| ***VASCULAR HYPERTENSIVE DISORDERS*** | | | | | | | | | | |
| *ACCELERATED AND MALIGNANT HYPERTENSION* | | | | | | | | | | |

| | | | | | | | | | | |
|---|---|---|---|---|---|---|---|---|---|---|
| HYPERTENSIVE URGENCY | 0 | 0 | 0 | 0 | 0 | 0 | 0 | 0 | 0 | 0 |
| *VASCULAR HYPERTENSIVE DISORDERS NEC* | | | | | | | | | | |
| HYPERTENSION | 1 | 1 | 0 | 0 | 0 | 1 | 1 | 0 | 0 | 0 |
| SYSTOLIC HYPERTENSION | 0 | 0 | 0 | 0 | 0 | 0 | 0 | 0 | 0 | 0 |
| ***VASCULAR INFECTIONS AND INFLAMMATIONS*** | | | | | | | | | | |
| *ARTERIAL INFECTIONS AND INFLAMMATIONS* | | | | | | | | | | |
| GIANT CELL ARTERITIS | 0 | 0 | 0 | 0 | 0 | 0 | 0 | 0 | 0 | 0 |
| *PHLEBITIS NEC* | | | | | | | | | | |
| PHLEBITIS | 0 | 0 | 0 | 0 | 0 | 0 | 0 | 0 | 0 | 0 |
| *VASCULITIDES NEC* | | | | | | | | | | |
| VASCULITIS | 0 | 0 | 0 | 0 | 0 | 0 | 0 | 0 | 0 | 0 |
| ***VENOUS VARICES*** | | | | | | | | | | |
| *VARICOSE VEINS NEC* | | | | | | | | | | |
| SPIDER VEIN | 0 | 0 | 0 | 0 | 0 | 0 | 0 | 0 | 0 | 0 |
| VARICOSE VEIN | 0 | 0 | 0 | 0 | 0 | 0 | 0 | 0 | 0 | 0 |
| **TOTAL ADR EVENTS** | ***314*** | ***209*** | ***39*** | ***53*** | ***13*** | ***294*** | ***209*** | ***27*** | ***52*** | ***6*** |

**TABLE S1-6. COVID-19 vaccine ADR listings for events reported in the YCVM with no dose identification information in (a) those reporting any vaccination dose and (b) in those who had reported a 1st dose vaccination and any subsequent doses.**

| | Patients with any vaccination dose: ADR Counts | Patients reporting a 1st dose: ADR Counts |
|---|---|---|
| **REACTION TERM (SOC, *HLGT*, *HLT*, PT)** | Unknown | Unknown |
| ***(freetext)*** | 3 | 3 |
| **BLOOD AND LYMPHATIC SYSTEM DISORDERS** | | |
| ***COAGULOPATHIES AND BLEEDING DIATHESES (EXCL THROMBOCYTOPENIC)*** | | |
| *BLEEDING TENDENCIES* | | |
| INCREASED TENDENCY TO BRUISE | 1 | 1 |
| ***HAEMOGLOBINOPATHIES*** | | |
| *SICKLE CELL TRAIT AND DISORDERS* | | |
| SICKLE CELL ANAEMIA WITH CRISIS | 0 | 0 |
| ***PLATELET DISORDERS*** | | |
| *THROMBOCYTOPENIAS* | | |
| IMMUNE THROMBOCYTOPENIA | 0 | 0 |
| ***SPLEEN, LYMPHATIC AND RETICULOENDOTHELIAL SYSTEM DISORDERS*** | | |
| *LYMPHATIC SYSTEM DISORDERS NEC* | | |
| LYMPH NODE PAIN | 4 | 4 |
| LYMPHADENITIS | 0 | 0 |
| LYMPHADENOPATHY | 9 | 8 |
| **CARDIAC DISORDERS** | | |
| ***CARDIAC ARRHYTHMIAS*** | | |
| *RATE AND RHYTHM DISORDERS NEC* | | |
| ARRHYTHMIA | 0 | 0 |
| CARDIAC FLUTTER | 0 | 0 |
| EXTRASYSTOLES | 0 | 0 |
| TACHYCARDIA | 0 | 0 |
| *SUPRAVENTRICULAR ARRHYTHMIAS* | | |
| ATRIAL FIBRILLATION | 0 | 0 |
| SUPRAVENTRICULAR TACHYCARDIA | 0 | 0 |
| ***CARDIAC DISORDERS, SIGNS AND SYMPTOMS NEC*** | | |
| *CARDIAC DISORDERS NEC* | | |
| CARDIOVASCULAR DISORDER | 0 | 0 |
| *CARDIAC SIGNS AND SYMPTOMS NEC* | | |
| PALPITATIONS | 10 | 8 |
| ***CORONARY ARTERY DISORDERS*** | | |
| *ISCHAEMIC CORONARY ARTERY DISORDERS* | | |
| ANGINA PECTORIS | 0 | 0 |
| MYOCARDIAL INFARCTION | 0 | 0 |
| ***HEART FAILURES*** | | |
| *HEART FAILURES NEC* | | |
| CARDIAC FAILURE | 0 | 0 |
| ***MYOCARDIAL DISORDERS*** | | |
| *NONINFECTIOUS MYOCARDITIS* | | |

| | | |
|---|---|---|
| MYOCARDITIS | 2 | 2 |
| ***PERICARDIAL DISORDERS*** | | |
| *NONINFECTIOUS PERICARDITIS* | | |
| PERICARDITIS | 0 | 0 |
| **CONGENITAL, FAMILIAL AND GENETIC DISORDERS** | | |
| ***CARDIAC AND VASCULAR DISORDERS CONGENITAL*** | | |
| *CARDIAC DISORDERS CONGENITAL NEC* | | |
| HEART DISEASE CONGENITAL | 0 | 0 |
| ***METABOLIC AND NUTRITIONAL DISORDERS CONGENITAL*** | | |
| *INBORN ERRORS OF AMINO ACID METABOLISM* | | |
| HYPERGLYCINAEMIA | 0 | 0 |
| *INBORN ERRORS OF STEROID SYNTHESIS* | | |
| 11-BETA-HYDROXYLASE DEFICIENCY | 0 | 0 |
| ***NEUROLOGICAL DISORDERS CONGENITAL*** | | |
| *PERIPHERAL NERVOUS SYSTEM DISORDERS CONGENITAL NEC* | | |
| PAROXYSMAL EXTREME PAIN DISORDER | 0 | 0 |
| **EAR AND LABYRINTH DISORDERS** | | |
| ***AURAL DISORDERS NEC*** | | |
| *EAR DISORDERS NEC* | | |
| EAR DISCOMFORT | 0 | 0 |
| EAR DISORDER | 0 | 0 |
| EAR PAIN | 2 | 2 |
| EAR SWELLING | 0 | 0 |
| ***EXTERNAL EAR DISORDERS (EXCL CONGENITAL)*** | | |
| *EXTERNAL EAR DISORDERS NEC* | | |
| EXCESSIVE CERUMEN PRODUCTION | 0 | 0 |
| ***HEARING DISORDERS*** | | |
| *HEARING LOSSES* | | |
| DEAFNESS | 0 | 0 |
| DEAFNESS NEUROSENSORY | 0 | 0 |
| DEAFNESS PERMANENT | 0 | 0 |
| DEAFNESS UNILATERAL | 0 | 0 |
| HYPOACUSIS | 0 | 0 |
| SUDDEN HEARING LOSS | 0 | 0 |
| *HYPERACUSIA* | | |
| HYPERACUSIS | 0 | 0 |
| ***INNER EAR AND VIIITH CRANIAL NERVE DISORDERS*** | | |
| *INNER EAR SIGNS AND SYMPTOMS* | | |
| MOTION SICKNESS | 0 | 0 |
| TINNITUS | 7 | 7 |
| VERTIGO | 3 | 3 |
| VERTIGO LABYRINTHINE | 0 | 0 |
| VERTIGO POSITIONAL | 0 | 0 |
| **ENDOCRINE DISORDERS** | | |
| ***THYROID GLAND DISORDERS*** | | |
| *THYROID HYPERFUNCTION DISORDERS* | | |
| HYPERTHYROIDISM | 0 | 0 |
| **EYE DISORDERS** | | |
| ***EYE DISORDERS NEC*** | | |

| | | |
|---|---|---|
| *LACRIMATION DISORDERS* | | |
| DRY EYE | 0 | 0 |
| LACRIMATION INCREASED | 1 | 1 |
| *OCULAR DISORDERS NEC* | | |
| EYE OEDEMA | 0 | 0 |
| EYE PAIN | 9 | 9 |
| EYE SWELLING | 0 | 0 |
| EYE ULCER | 0 | 0 |
| EYELID PAIN | 0 | 0 |
| OCULAR DISCOMFORT | 0 | 0 |
| PERIORBITAL DISCOMFORT | 0 | 0 |
| PERIORBITAL SWELLING | 0 | 0 |
| ***OCULAR HAEMORRHAGES AND VASCULAR DISORDERS NEC*** | | |
| *CONJUNCTIVAL AND CORNEAL BLEEDING AND VASCULAR DISORDERS* | | |
| CONJUNCTIVAL HAEMORRHAGE | 1 | 1 |
| *LID BLEEDING AND VASCULAR DISORDERS* | | |
| EYELID BLEEDING | 0 | 0 |
| ***OCULAR INFECTIONS, IRRITATIONS AND INFLAMMATIONS*** | | |
| *LID, LASH AND LACRIMAL INFECTIONS, IRRITATIONS AND INFLAMMATIONS* | | |
| BLEPHARITIS | 0 | 0 |
| ERYTHEMA OF EYELID | 0 | 0 |
| EYELID IRRITATION | 0 | 0 |
| *OCULAR INFECTIONS, INFLAMMATIONS AND ASSOCIATED MANIFESTATIONS* | | |
| EYE DISCHARGE | 0 | 0 |
| EYE IRRITATION | 0 | 0 |
| EYE PRURITUS | 0 | 0 |
| LIMBAL SWELLING | 0 | 0 |
| OCULAR HYPERAEMIA | 0 | 0 |
| ***OCULAR NEUROMUSCULAR DISORDERS*** | | |
| *EYELID MOVEMENT DISORDERS* | | |
| BLEPHAROSPASM | 0 | 0 |
| ***OCULAR SENSORY SYMPTOMS NEC*** | | |
| *OCULAR SENSATION DISORDERS* | | |
| ABNORMAL SENSATION IN EYE | 0 | 0 |
| ASTHENOPIA | 2 | 2 |
| FOREIGN BODY SENSATION IN EYES | 0 | 0 |
| PHOTOPHOBIA | 2 | 2 |
| ***OCULAR STRUCTURAL CHANGE, DEPOSIT AND DEGENERATION NEC*** | | |
| *CHOROID AND VITREOUS STRUCTURAL CHANGE, DEPOSIT AND DEGENERATION* | | |
| VITREOUS DETACHMENT | 0 | 0 |
| VITREOUS FLOATERS | 0 | 0 |
| ***RETINA, CHOROID AND VITREOUS HAEMORRHAGES AND VASCULAR DISORDERS*** | | |
| *RETINAL BLEEDING AND VASCULAR DISORDERS (EXCL RETINOPATHY)* | | |
| RETINAL VEIN OCCLUSION | 0 | 0 |
| *RETINOPATHIES NEC* | | |
| RETINAL EXUDATES | 0 | 0 |

| | | |
|---|---|---|
| ***VISION DISORDERS*** | | |
| *VISUAL DISORDERS NEC* | | |
| DIPLOPIA | 1 | 1 |
| HALO VISION | 0 | 0 |
| METAMORPHOPSIA | 0 | 0 |
| PHOTOPSIA | 2 | 2 |
| VISION BLURRED | 1 | 0 |
| *VISUAL IMPAIRMENT AND BLINDNESS (EXCL COLOUR BLINDNESS)* | | |
| BLINDNESS | 0 | 0 |
| BLINDNESS TRANSIENT | 0 | 0 |
| SUDDEN VISUAL LOSS | 0 | 0 |
| VISUAL IMPAIRMENT | 0 | 0 |
| **GASTROINTESTINAL DISORDERS** | | |
| ***ANAL AND RECTAL CONDITIONS NEC*** | | |
| *ANAL AND RECTAL SIGNS AND SYMPTOMS* | | |
| ANAL PARAESTHESIA | 0 | 0 |
| ***BENIGN NEOPLASMS GASTROINTESTINAL*** | | |
| *BENIGN ORAL CAVITY NEOPLASMS* | | |
| MOUTH CYST | 0 | 0 |
| ***DENTAL AND GINGIVAL CONDITIONS*** | | |
| *DENTAL DISORDERS NEC* | | |
| TEETHING | 0 | 0 |
| *DENTAL PAIN AND SENSATION DISORDERS* | | |
| DENTAL PARAESTHESIA | 0 | 0 |
| TOOTHACHE | 0 | 0 |
| *GINGIVAL DISORDERS, SIGNS AND SYMPTOMS NEC* | | |
| GINGIVAL BLISTER | 0 | 0 |
| GINGIVAL PAIN | 0 | 0 |
| GINGIVAL SWELLING | 0 | 0 |
| ***GASTROINTESTINAL CONDITIONS NEC*** | | |
| *GASTROINTESTINAL MUCOSAL DYSTROPHIES AND SECRETION DISORDERS* | | |
| BARRETT'S OESOPHAGUS | 0 | 0 |
| ***GASTROINTESTINAL HAEMORRHAGES NEC*** | | |
| *INTESTINAL HAEMORRHAGES* | | |
| RECTAL HAEMORRHAGE | 0 | 0 |
| *NON-SITE SPECIFIC GASTROINTESTINAL HAEMORRHAGES* | | |
| HAEMATEMESIS | 0 | 0 |
| ***GASTROINTESTINAL INFLAMMATORY CONDITIONS*** | | |
| *COLITIS (EXCL INFECTIVE)* | | |
| COLITIS | 0 | 0 |
| COLITIS MICROSCOPIC | 0 | 0 |
| COLITIS ULCERATIVE | 0 | 0 |
| *GASTRITIS (EXCL INFECTIVE)* | | |
| GASTRITIS | 0 | 0 |
| REFLUX GASTRITIS | 0 | 0 |
| *GASTROINTESTINAL INFLAMMATORY DISORDERS NEC* | | |
| GASTROINTESTINAL TRACT IRRITATION | 0 | 0 |
| ***GASTROINTESTINAL MOTILITY AND DEFAECATION CONDITIONS*** | | |
| *DIARRHOEA (EXCL INFECTIVE)* | | |
| DIARRHOEA | 13 | 12 |
| *GASTROINTESTINAL ATONIC AND HYPOMOTILITY DISORDERS NEC* | | |
| CONSTIPATION | 0 | 0 |

| | | |
|---|---|---|
| GASTROOESOPHAGEAL REFLUX DISEASE | 1 | 1 |
| *GASTROINTESTINAL DYSKINETIC DISORDERS* | | |
| CHANGE OF BOWEL HABIT | 0 | 0 |
| *GASTROINTESTINAL SPASTIC AND HYPERMOTILITY DISORDERS* | | |
| IRRITABLE BOWEL SYNDROME | 0 | 0 |
| ***GASTROINTESTINAL SIGNS AND SYMPTOMS*** | | |
| *DYSPEPTIC SIGNS AND SYMPTOMS* | | |
| DYSPEPSIA | 2 | 1 |
| ERUCTATION | 0 | 0 |
| *FAECAL ABNORMALITIES NEC* | | |
| FAECALOMA | 0 | 0 |
| FAECES DISCOLOURED | 0 | 0 |
| FAECES SOFT | 0 | 0 |
| *FLATULENCE, BLOATING AND DISTENSION* | | |
| ABDOMINAL DISTENSION | 0 | 0 |
| FLATULENCE | 0 | 0 |
| *GASTROINTESTINAL AND ABDOMINAL PAINS (EXCL ORAL AND THROAT)* | | |
| ABDOMINAL PAIN | 3 | 3 |
| ABDOMINAL PAIN LOWER | 0 | 0 |
| ABDOMINAL PAIN UPPER | 10 | 10 |
| GASTROINTESTINAL PAIN | 0 | 0 |
| *GASTROINTESTINAL SIGNS AND SYMPTOMS NEC* | | |
| ABDOMINAL DISCOMFORT | 4 | 3 |
| ABDOMINAL SYMPTOM | 0 | 0 |
| ACUTE ABDOMEN | 0 | 0 |
| ANAL INCONTINENCE | 0 | 0 |
| BREATH ODOUR | 0 | 0 |
| DYSPHAGIA | 0 | 0 |
| ODYNOPHAGIA | 0 | 0 |
| *NAUSEA AND VOMITING SYMPTOMS* | | |
| NAUSEA | 79 | 75 |
| RETCHING | 0 | 0 |
| VOMITING | 17 | 17 |
| VOMITING PROJECTILE | 0 | 0 |
| ***MALABSORPTION CONDITIONS*** | | |
| *MALABSORPTION SYNDROMES* | | |
| COELIAC DISEASE | 0 | 0 |
| ***ORAL SOFT TISSUE CONDITIONS*** | | |
| *ORAL SOFT TISSUE DISORDERS NEC* | | |
| CHEILITIS | 0 | 0 |
| LIP BLISTER | 0 | 0 |
| ORAL LICHEN PLANUS | 0 | 0 |
| *ORAL SOFT TISSUE INFECTIONS* | | |
| ANGULAR CHEILITIS | 0 | 0 |
| *ORAL SOFT TISSUE SIGNS AND SYMPTOMS* | | |
| HYPOAESTHESIA ORAL | 0 | 0 |
| LIP PAIN | 0 | 0 |
| ORAL DISCOMFORT | 0 | 0 |
| ORAL MUCOSAL ROUGHENING | 0 | 0 |
| ORAL PAIN | 0 | 0 |
| PARAESTHESIA ORAL | 1 | 1 |
| LIP SWELLING | 2 | 2 |

| | | |
|---|---|---|
| MOUTH SWELLING | 0 | 0 |
| *STOMATITIS AND ULCERATION* | | |
| APHTHOUS ULCER | 0 | 0 |
| LIP ULCERATION | 0 | 0 |
| MOUTH ULCERATION | 0 | 0 |
| STOMATITIS | 0 | 0 |
| ***SALIVARY GLAND CONDITIONS*** | | |
| *ORAL DRYNESS AND SALIVA ALTERED* | | |
| DRY MOUTH | 7 | 7 |
| LIP DRY | 1 | 1 |
| SALIVARY HYPOSECRETION | 0 | 0 |
| ***TONGUE CONDITIONS*** | | |
| *TONGUE SIGNS AND SYMPTOMS* | | |
| GLOSSODYNIA | 0 | 0 |
| SWOLLEN TONGUE | 1 | 1 |
| TONGUE COATED | 0 | 0 |
| TONGUE DISCOMFORT | 0 | 0 |
| TONGUE OEDEMA | 0 | 0 |
| TONGUE SPASM | 0 | 0 |
| **GENERAL DISORDERS AND ADMINISTRATION SITE CONDITIONS** | | |
| ***ADMINISTRATION SITE REACTIONS*** | | |
| *ADMINISTRATION SITE REACTIONS NEC* | | |
| ADMINISTRATION SITE BRUISE | 0 | 0 |
| ADMINISTRATION SITE PAIN | 0 | 0 |
| PUNCTURE SITE BRUISE | 3 | 3 |
| PUNCTURE SITE PAIN | 1 | 1 |
| *APPLICATION SITE REACTIONS* | | |
| APPLICATION SITE BRUISE | 0 | 0 |
| APPLICATION SITE ERYTHEMA | 0 | 0 |
| APPLICATION SITE PAIN | 0 | 0 |
| *IMPLANT AND CATHETER SITE REACTIONS* | | |
| IMPLANT SITE PAIN | 0 | 0 |
| IMPLANT SITE WARMTH | 0 | 0 |
| *INFUSION SITE REACTIONS* | | |
| INFUSION SITE PAIN | 0 | 0 |
| INFUSION SITE SCAB | 0 | 0 |
| INFUSION SITE WARMTH | 0 | 0 |
| *INJECTION SITE REACTIONS* | | |
| INJECTION SITE BRUISING | 0 | 0 |
| INJECTION SITE DISCOMFORT | 0 | 0 |
| INJECTION SITE ERYTHEMA | 1 | 1 |
| INJECTION SITE HYPERSENSITIVITY | 0 | 0 |
| INJECTION SITE INFLAMMATION | 1 | 1 |
| INJECTION SITE INJURY | 0 | 0 |
| INJECTION SITE IRRITATION | 0 | 0 |
| INJECTION SITE JOINT PAIN | 0 | 0 |
| INJECTION SITE MASS | 2 | 2 |
| INJECTION SITE NODULE | 0 | 0 |
| INJECTION SITE OEDEMA | 0 | 0 |
| INJECTION SITE PAIN | 21 | 21 |
| INJECTION SITE PAPULE | 0 | 0 |
| INJECTION SITE PARAESTHESIA | 0 | 0 |

| | | |
|---|---|---|
| INJECTION SITE PRURITUS | 0 | 0 |
| INJECTION SITE RASH | 0 | 0 |
| INJECTION SITE REACTION | 0 | 0 |
| INJECTION SITE SCAB | 0 | 0 |
| INJECTION SITE SWELLING | 2 | 2 |
| INJECTION SITE URTICARIA | 0 | 0 |
| INJECTION SITE WARMTH | 0 | 0 |
| *INSTILLATION SITE REACTIONS* | | |
| INSTILLATION SITE PRURITUS | 0 | 0 |
| INSTILLATION SITE WARMTH | 1 | 1 |
| *VACCINATION SITE REACTIONS* | | |
| SHOULDER INJURY RELATED TO VACCINE ADMINISTRATION | 0 | 0 |
| VACCINATION SITE BRUISING | 0 | 0 |
| VACCINATION SITE DISCOMFORT | 0 | 0 |
| VACCINATION SITE ERYTHEMA | 1 | 1 |
| VACCINATION SITE JOINT ERYTHEMA | 0 | 0 |
| VACCINATION SITE JOINT PAIN | 5 | 5 |
| VACCINATION SITE MASS | 1 | 1 |
| VACCINATION SITE PAIN | 14 | 13 |
| VACCINATION SITE RASH | 1 | 1 |
| VACCINATION SITE SWELLING | 2 | 2 |
| VACCINATION SITE WARMTH | 1 | 1 |
| ***BODY TEMPERATURE CONDITIONS*** | | |
| *BODY TEMPERATURE ALTERED* | | |
| HYPERTHERMIA | 0 | 0 |
| HYPOTHERMIA | 0 | 0 |
| *FEBRILE DISORDERS* | | |
| PYREXIA | 100 | 99 |
| ***FATAL OUTCOMES*** | | |
| *DEATH AND SUDDEN DEATH* | | |
| DEATH | 0 | 0 |
| ***GENERAL SYSTEM DISORDERS NEC*** | | |
| *ADVERSE EFFECT ABSENT* | | |
| NO ADVERSE EVENT | 1 | 1 |
| *ASTHENIC CONDITIONS* | | |
| ASTHENIA | 9 | 8 |
| CHRONIC FATIGUE SYNDROME | 0 | 0 |
| DECREASED ACTIVITY | 0 | 0 |
| FATIGUE | 257 | 245 |
| MALAISE | 24 | 23 |
| SLUGGISHNESS | 0 | 0 |
| *FEELINGS AND SENSATIONS NEC* | | |
| CHILLS | 83 | 79 |
| FEELING ABNORMAL | 9 | 9 |
| FEELING COLD | 12 | 11 |
| FEELING HOT | 4 | 4 |
| FEELING JITTERY | 0 | 0 |
| FEELING OF BODY TEMPERATURE CHANGE | 3 | 3 |
| FEELING OF RELAXATION | 0 | 0 |
| HANGOVER | 2 | 2 |
| HUNGER | 2 | 2 |

| | | |
|---|---|---|
| SENSATION OF BLOOD FLOW | 0 | 0 |
| THIRST | 1 | 1 |
| *GAIT DISTURBANCES* | | |
| GAIT DISTURBANCE | 3 | 3 |
| GAIT INABILITY | 0 | 0 |
| *GENERAL SIGNS AND SYMPTOMS NEC* | | |
| CRYING | 0 | 0 |
| ENERGY INCREASED | 0 | 0 |
| EXERCISE TOLERANCE DECREASED | 1 | 1 |
| GENERAL SYMPTOM | 0 | 0 |
| ILLNESS | 11 | 11 |
| INFLUENZA LIKE ILLNESS | 37 | 35 |
| LOCAL REACTION | 0 | 0 |
| PERIPHERAL SWELLING | 12 | 12 |
| SWELLING | 4 | 3 |
| SWELLING FACE | 2 | 2 |
| TISSUE IRRITATION | 0 | 0 |
| *INFLAMMATIONS* | | |
| INFLAMMATION | 0 | 0 |
| SYSTEMIC INFLAMMATORY RESPONSE SYNDROME | 0 | 0 |
| *OEDEMA NEC* | | |
| OEDEMA | 0 | 0 |
| OEDEMA PERIPHERAL | 0 | 0 |
| *PAIN AND DISCOMFORT NEC* | | |
| AXILLARY PAIN | 1 | 1 |
| CHEST DISCOMFORT | 2 | 2 |
| CHEST PAIN | 6 | 5 |
| DISCOMFORT | 2 | 2 |
| FACIAL PAIN | 0 | 0 |
| HERNIA PAIN | 0 | 0 |
| INFLAMMATORY PAIN | 0 | 0 |
| NON-CARDIAC CHEST PAIN | 0 | 0 |
| PAIN | 49 | 47 |
| TENDERNESS | 20 | 19 |
| ***THERAPEUTIC AND NONTHERAPEUTIC EFFECTS (EXCL TOXICITY)*** | | |
| *THERAPEUTIC AND NONTHERAPEUTIC RESPONSES* | | |
| ADVERSE DRUG REACTION | 0 | 0 |
| ADVERSE EVENT | 5 | 4 |
| ADVERSE REACTION | 0 | 0 |
| IMMEDIATE POST-INJECTION REACTION | 0 | 0 |
| **HEPATOBILIARY DISORDERS** | | |
| ***HEPATIC AND HEPATOBILIARY DISORDERS*** | | |
| *HEPATOBILIARY SIGNS AND SYMPTOMS* | | |
| HEPATIC PAIN | 0 | 0 |
| *HEPATOCELLULAR DAMAGE AND HEPATITIS NEC* | | |
| LIVER INJURY | 0 | 0 |
| **IMMUNE SYSTEM DISORDERS** | | |
| ***ALLERGIC CONDITIONS*** | | |
| *ALLERGIC CONDITIONS NEC* | | |
| HYPERSENSITIVITY | 0 | 0 |
| MULTIPLE ALLERGIES | 0 | 0 |

| | | |
|---|---|---|
| *ALLERGIES TO FOODS, FOOD ADDITIVES, DRUGS AND OTHER CHEMICALS* | | |
| ALLERGY TO CHEMICALS | 0 | 0 |
| *ANAPHYLACTIC AND ANAPHYLACTOID RESPONSES* | | |
| ANAPHYLACTIC REACTION | 0 | 0 |
| *ATOPIC DISORDERS* | | |
| SEASONAL ALLERGY | 0 | 0 |
| ***IMMUNE DISORDERS NEC*** | | |
| *IMMUNE AND ASSOCIATED CONDITIONS NEC* | | |
| BACILLE CALMETTE-GUERIN SCAR REACTIVATION | | |
| SENSITISATION | 0 | 0 |
| **INFECTIONS AND INFESTATIONS** | | |
| ***BACTERIAL INFECTIOUS DISORDERS*** | | |
| *BACTERIAL INFECTIONS NEC* | | |
| CELLULITIS | 0 | 0 |
| PERIORBITAL CELLULITIS | 0 | 0 |
| *HELICOBACTER INFECTIONS* | | |
| HELICOBACTER GASTRITIS | 0 | 0 |
| *STAPHYLOCOCCAL INFECTIONS* | | |
| FURUNCLE | 1 | 1 |
| ***FUNGAL INFECTIOUS DISORDERS*** | | |
| *CANDIDA INFECTIONS* | | |
| CANDIDA INFECTION | 0 | 0 |
| VULVOVAGINAL CANDIDIASIS | 0 | 0 |
| ***INFECTIONS - PATHOGEN UNSPECIFIED*** | | |
| *ABDOMINAL AND GASTROINTESTINAL INFECTIONS* | | |
| APPENDICITIS | 0 | 0 |
| DIARRHOEA INFECTIOUS | 0 | 0 |
| GASTROINTESTINAL INFECTION | 0 | 0 |
| *BREAST INFECTIONS* | | |
| MASTITIS | 0 | 0 |
| *CENTRAL NERVOUS SYSTEM AND SPINAL INFECTIONS* | | |
| MYELITIS | 0 | 0 |
| *DENTAL AND ORAL SOFT TISSUE INFECTIONS* | | |
| ABSCESS ORAL | 0 | 0 |
| ORAL PUSTULE | 0 | 0 |
| PERICORONITIS | 0 | 0 |
| TOOTH ABSCESS | 0 | 0 |
| *EAR INFECTIONS* | | |
| EAR INFECTION | 1 | 1 |
| LABYRINTHITIS | 0 | 0 |
| OTITIS EXTERNA | 0 | 0 |
| *EYE AND EYELID INFECTIONS* | | |
| CONJUNCTIVITIS | 0 | 0 |
| HORDEOLUM | 0 | 0 |
| *INFECTIONS NEC* | | |
| ABSCESS | 0 | 0 |
| INFECTION | 1 | 1 |
| INJECTION SITE INFECTION | 0 | 0 |
| LOCALISED INFECTION | 1 | 0 |
| WOUND INFECTION | 0 | 0 |
| *LOWER RESPIRATORY TRACT AND LUNG INFECTIONS* | | |
| LOWER RESPIRATORY TRACT INFECTION | 1 | 1 |

| | | |
|---|---|---|
| PNEUMONIA | 2 | 2 |
| *MALE REPRODUCTIVE TRACT INFECTIONS* | | |
| ORCHITIS | 0 | 0 |
| *SEPSIS, BACTERAEMIA, VIRAEMIA AND FUNGAEMIA NEC* | | |
| SEPTIC RASH | 0 | 0 |
| *SKIN STRUCTURES AND SOFT TISSUE INFECTIONS* | | |
| INFECTED DERMAL CYST | 0 | 0 |
| INJECTION SITE PUSTULE | 0 | 0 |
| SKIN INFECTION | 0 | 0 |
| *UPPER RESPIRATORY TRACT INFECTIONS* | | |
| LARYNGITIS | 0 | 0 |
| NASOPHARYNGITIS | 9 | 8 |
| PHARYNGITIS | 0 | 0 |
| RHINITIS | 1 | 1 |
| SINUSITIS | 1 | 1 |
| TONSILLITIS | 1 | 1 |
| *URINARY TRACT INFECTIONS* | | |
| CYSTITIS | 0 | 0 |
| URINARY TRACT INFECTION | 0 | 0 |
| ***PROTOZOAL INFECTIOUS DISORDERS*** | | |
| *TRYPANOSOMAL INFECTIONS* | | |
| AFRICAN TRYPANOSOMIASIS | 0 | 0 |
| ***RICKETTSIAL INFECTIOUS DISORDERS*** | | |
| *COXIELLA INFECTIONS* | | |
| Q FEVER | 0 | 0 |
| ***VIRAL INFECTIOUS DISORDERS*** | | |
| *CORONAVIRUS INFECTIONS* | | |
| COVID-19 | 1 | 0 |
| *HERPES VIRAL INFECTIONS* | | |
| GENITAL HERPES | 0 | 0 |
| HERPES OPHTHALMIC | 0 | 0 |
| HERPES SIMPLEX | 0 | 0 |
| HERPES ZOSTER | 2 | 2 |
| NASAL HERPES | 0 | 0 |
| OPHTHALMIC HERPES ZOSTER | 0 | 0 |
| ORAL HERPES | 1 | 1 |
| *INFLUENZA VIRAL INFECTIONS* | | |
| H2N2 INFLUENZA | 0 | 0 |
| INFLUENZA | 22 | 21 |
| *RETROVIRAL INFECTIONS* | | |
| AIDS RELATED COMPLEX | 0 | 0 |
| *VIRAL INFECTIONS NEC* | | |
| GASTROENTERITIS VIRAL | 0 | 0 |
| SWEATING FEVER | 2 | 2 |
| VESTIBULAR NEURONITIS | 0 | 0 |
| VIRAL DIARRHOEA | 0 | 0 |
| VIRAL RASH | 0 | 0 |
| VIRAL UPPER RESPIRATORY TRACT INFECTION | 0 | 0 |
| **INJURY, POISONING AND PROCEDURAL COMPLICATIONS** | | |
| ***BONE AND JOINT INJURIES*** | | |
| *FRACTURES AND DISLOCATIONS NEC* | | |
| JOINT DISLOCATION | 0 | 0 |

| | | |
|---|---|---|
| ***EXPOSURES, CHEMICAL INJURIES AND POISONING*** | | |
| *POISONING AND TOXICITY* | | |
| SYSTEMIC TOXICITY | 0 | 0 |
| TOXICITY TO VARIOUS AGENTS | 0 | 0 |
| ***INJURIES BY PHYSICAL AGENTS*** | | |
| *CONDITIONS CAUSED BY COLD* | | |
| CHILLBLAINS | 1 | 1 |
| *HEAT INJURIES (EXCL THERMAL BURNS)* | | |
| HEAT EXHAUSTION | 0 | 0 |
| HEAT OEDEMA | 0 | 0 |
| *RADIATION INJURIES* | | |
| SUNBURN | 0 | 0 |
| *THERMAL BURNS* | | |
| THERMAL BURN | 0 | 0 |
| THERMAL BURNS OF EYE | 0 | 0 |
| ***INJURIES NEC*** | | |
| *CHEST AND RESPIRATORY TRACT INJURIES NEC* | | |
| CHEST CRUSHING | 0 | 0 |
| *EYE INJURIES NEC* | | |
| EYE CONTUSION | 0 | 0 |
| *MUSCLE, TENDON AND LIGAMENT INJURIES* | | |
| LIGAMENT SPRAIN | 0 | 0 |
| MUSCLE INJURY | 0 | 0 |
| MUSCLE STRAIN | 0 | 0 |
| *NERVE INJURIES NEC* | | |
| NERVE INJURY | 0 | 0 |
| *NON-SITE SPECIFIC INJURIES NEC* | | |
| ARTHROPOD STING | 0 | 0 |
| BITE | 0 | 0 |
| FALL | 0 | 0 |
| INFLAMMATION OF WOUND | 0 | 0 |
| TISSUE INJURY | 0 | 0 |
| WOUND COMPLICATION | 0 | 0 |
| WOUND SECRETION | 0 | 0 |
| *SITE SPECIFIC INJURIES NEC* | | |
| LIMB INJURY | 0 | 0 |
| *SKIN INJURIES NEC* | | |
| CONTUSION | 12 | 11 |
| SCAR | 0 | 0 |
| SKIN WOUND | 0 | 0 |
| ***PROCEDURAL RELATED INJURIES AND COMPLICATIONS NEC*** | | |
| *CARDIAC AND VASCULAR PROCEDURAL COMPLICATIONS* | | |
| CARDIAC PROCEDURE COMPLICATION | 0 | 0 |
| *GASTROINTESTINAL AND HEPATOBILIARY PROCEDURAL COMPLICATIONS* | | |
| PROCEDURAL NAUSEA | 0 | 0 |
| *NEUROLOGICAL AND PSYCHIATRIC PROCEDURAL COMPLICATIONS* | | |
| PROCEDURAL DIZZINESS | 0 | 0 |
| *NON-SITE SPECIFIC PROCEDURAL COMPLICATIONS* | | |
| INCISION SITE PAIN | 0 | 0 |
| INCISION SITE SWELLING | 0 | 0 |
| INJECTION RELATED REACTION | 0 | 0 |
| POST PROCEDURAL COMPLICATION | 0 | 0 |

| | | |
|---|---|---|
| PROCEDURAL PAIN | 1 | 1 |
| *VACCINATION RELATED COMPLICATIONS* | | |
| IMMUNISATION REACTION | 0 | 0 |
| **INVESTIGATIONS** | | |
| ***CARDIAC AND VASCULAR INVESTIGATIONS (EXCL ENZYME TESTS)*** | | |
| *HEART RATE AND PULSE INVESTIGATIONS* | | |
| HEART RATE | 3 | 3 |
| HEART RATE DECREASED | 1 | 1 |
| HEART RATE INCREASED | 3 | 3 |
| HEART RATE IRREGULAR | 0 | 0 |
| *VASCULAR TESTS NEC (INCL BLOOD PRESSURE)* | | |
| BLOOD PRESSURE DECREASED | 0 | 0 |
| BLOOD PRESSURE INCREASED | 0 | 0 |
| BLOOD PRESSURE MEASUREMENT | 1 | 1 |
| ***ENDOCRINE INVESTIGATIONS (INCL SEX HORMONES)*** | | |
| *ENDOCRINE ANALYSES AND IMAGING NEC* | | |
| HORMONE LEVEL ABNORMAL | 0 | 0 |
| *PITUITARY ANALYSES ANTERIOR* | | |
| BLOOD FOLLICLE STIMULATING HORMONE INCREASED | 0 | 0 |
| BLOOD LUTEINISING HORMONE | 0 | 0 |
| *THYROID ANALYSES* | | |
| TRI-IODOTHYRONINE | 0 | 0 |
| ***HAEMATOLOGY INVESTIGATIONS (INCL BLOOD GROUPS)*** | | |
| *COAGULATION AND BLEEDING ANALYSES* | | |
| BLEEDING TIME | 1 | 1 |
| INTERNATIONAL NORMALISED RATIO DECREASED | 0 | 0 |
| *PLATELET ANALYSES* | | |
| PLATELET COUNT INCREASED | 1 | 1 |
| *RED BLOOD CELL ANALYSES* | | |
| HAEMOGLOBIN | 0 | 0 |
| ***METABOLIC, NUTRITIONAL AND BLOOD GAS INVESTIGATIONS*** | | |
| *BLOOD GAS AND ACID BASE ANALYSES* | | |
| OXYGEN SATURATION DECREASED | 0 | 0 |
| *CARBOHYDRATE TOLERANCE ANALYSES (INCL DIABETES)* | | |
| BLOOD GLUCOSE | 0 | 0 |
| BLOOD GLUCOSE ABNORMAL | 0 | 0 |
| BLOOD GLUCOSE DECREASED | 0 | 0 |
| BLOOD GLUCOSE INCREASED | 0 | 0 |
| ***MICROBIOLOGY AND SEROLOGY INVESTIGATIONS*** | | |
| *VIRUS IDENTIFICATION AND SEROLOGY* | | |
| SARS-COV-2 TEST | 0 | 0 |
| SARS-COV-2 TEST POSITIVE | 0 | 0 |
| ***MUSCULOSKELETAL AND SOFT TISSUE INVESTIGATIONS (EXCL ENZYME TESTS)*** | | |
| *MUSCULOSKELETAL AND SOFT TISSUE IMAGING PROCEDURES* | | |
| BONE SCAN | 0 | 0 |
| ***NEUROLOGICAL, SPECIAL SENSES AND PSYCHIATRIC INVESTIGATIONS*** | | |
| *CENTRAL NERVOUS SYSTEM IMAGING PROCEDURES* | | |
| MAGNETIC RESONANCE IMAGING HEAD | 0 | 0 |
| SCAN BRAIN | 0 | 0 |
| ***PHYSICAL EXAMINATION AND ORGAN SYSTEM STATUS TOPICS*** | | |
| *PHYSICAL EXAMINATION PROCEDURES AND ORGAN SYSTEM STATUS* | | |
| BODY TEMPERATURE | 9 | 9 |

| | | |
|---|---|---|
| BODY TEMPERATURE ABNORMAL | 0 | 0 |
| BODY TEMPERATURE DECREASED | 0 | 0 |
| BODY TEMPERATURE FLUCTUATION | 0 | 0 |
| BODY TEMPERATURE INCREASED | 1 | 1 |
| GRIP STRENGTH DECREASED | 0 | 0 |
| HEAD LAG | 0 | 0 |
| LEFT-HANDEDNESS | 0 | 0 |
| LYMPH NODE PALPABLE | 0 | 0 |
| RESPIRATORY RATE DECREASED | 1 | 1 |
| SKIN TEMPERATURE | 0 | 0 |
| WEIGHT DECREASED | 0 | 0 |
| WEIGHT INCREASED | 1 | 1 |
| ***RENAL AND URINARY TRACT INVESTIGATIONS AND URINALYSES*** | | |
| *URINALYSIS NEC* | | |
| BLOOD URINE | 0 | 0 |
| NITRITE URINE PRESENT | 0 | 0 |
| PH URINE | 1 | 1 |
| *URINARY TRACT FUNCTION ANALYSES NEC* | | |
| URINE OUTPUT | 0 | 0 |
| URINE OUTPUT INCREASED | 0 | 0 |
| ***REPRODUCTIVE ORGAN AND BREAST INVESTIGATIONS (EXCL HORMONE ANALYSES)*** | | |
| *REPRODUCTIVE ORGAN AND BREAST IMAGING PROCEDURES* | | |
| BREAST SCAN | 0 | 0 |
| ***RESPIRATORY AND PULMONARY INVESTIGATIONS (EXCL BLOOD GASES)*** | | |
| *RESPIRATORY AND PULMONARY FUNCTION DIAGNOSTIC PROCEDURES* | | |
| FORCED EXPIRATORY VOLUME DECREASED | 0 | 0 |
| FORCED EXPIRATORY VOLUME INCREASED | 0 | 0 |
| ***WATER, ELECTROLYTE AND MINERAL INVESTIGATIONS*** | | |
| *WATER AND ELECTROLYTE ANALYSES NEC* | | |
| VOLUME BLOOD | 0 | 0 |
| **METABOLISM AND NUTRITION DISORDERS** | | |
| ***APPETITE AND GENERAL NUTRITIONAL DISORDERS*** | | |
| *APPETITE DISORDERS* | | |
| APPETITE DISORDER | 1 | 1 |
| DECREASED APPETITE | 15 | 14 |
| FOOD CRAVING | 0 | 0 |
| FOOD REFUSAL | 0 | 0 |
| INCREASED APPETITE | 1 | 1 |
| *GENERAL NUTRITIONAL DISORDERS NEC* | | |
| FOOD AVERSION | 0 | 0 |
| ***ELECTROLYTE AND FLUID BALANCE CONDITIONS*** | | |
| *TOTAL FLUID VOLUME DECREASED* | | |
| DEHYDRATION | 0 | 0 |
| *TOTAL FLUID VOLUME INCREASED* | | |
| FLUID RETENTION | 0 | 0 |
| ***FOOD INTOLERANCE SYNDROMES*** | | |
| *FOOD MALABSORPTION AND INTOLERANCE SYNDROMES (EXCL SUGAR INTOLERANCE)* | | |
| ALCOHOL INTOLERANCE | 0 | 0 |
| ***GLUCOSE METABOLISM DISORDERS (INCL DIABETES MELLITUS)*** | | |
| *DIABETES MELLITUS (INCL SUBTYPES)* | | |
| DIABETES MELLITUS | 0 | 0 |
| DIABETES MELLITUS INADEQUATE CONTROL | 1 | 1 |
| *HYPERGLYCAEMIC CONDITIONS NEC* | | |

| | | |
|---|---|---|
| HYPERGLYCAEMIA | 0 | 0 |
| *HYPOGLYCAEMIC CONDITIONS NEC* | | |
| HYPOGLYCAEMIA | 0 | 0 |
| ***PURINE AND PYRIMIDINE METABOLISM DISORDERS*** | | |
| *DISORDERS OF PURINE METABOLISM* | | |
| GOUT | 0 | 0 |
| **MUSCULOSKELETAL AND CONNECTIVE TISSUE DISORDERS** | | |
| ***BONE DISORDERS (EXCL CONGENITAL AND FRACTURES)*** | | |
| *BONE DISORDERS NEC* | | |
| JAW DISORDER | 0 | 0 |
| OSTEITIS | 0 | 0 |
| *BONE RELATED SIGNS AND SYMPTOMS* | | |
| BONE PAIN | 1 | 1 |
| BONE SWELLING | 1 | 1 |
| PAIN IN JAW | 0 | 0 |
| PUBIC PAIN | 0 | 0 |
| SPINAL PAIN | 0 | 0 |
| ***CONNECTIVE TISSUE DISORDERS (EXCL CONGENITAL)*** | | |
| *CONNECTIVE TISSUE DISORDERS NEC* | | |
| POLYMYALGIA RHEUMATICA | 0 | 0 |
| *LUPUS ERYTHEMATOSUS (INCL SUBTYPES)* | | |
| SYSTEMIC LUPUS ERYTHEMATOSUS | 0 | 0 |
| ***JOINT DISORDERS*** | | |
| *ARTHROPATHIES NEC* | | |
| ARTHRITIS | 1 | 0 |
| ARTHROPATHY | 0 | 0 |
| POLYARTHRITIS | 0 | 0 |
| RHEUMATIC FEVER | 0 | 0 |
| *JOINT RELATED DISORDERS NEC* | | |
| JOINT LOCK | 1 | 1 |
| PERIARTHRITIS | 0 | 0 |
| TEMPOROMANDIBULAR PAIN AND DYSFUNCTION SYNDROME | 0 | 0 |
| *JOINT RELATED SIGNS AND SYMPTOMS* | | |
| ARTHRALGIA | 60 | 54 |
| JAW CLICKING | 0 | 0 |
| JOINT STIFFNESS | 2 | 1 |
| JOINT SWELLING | 0 | 0 |
| *OSTEOARTHROPATHIES* | | |
| OSTEOARTHRITIS | 0 | 0 |
| *PSORIATIC ARTHROPATHIES* | | |
| PSORIATIC ARTHROPATHY | 0 | 0 |
| *RHEUMATOID ARTHROPATHIES* | | |
| RHEUMATOID ARTHRITIS | 0 | 0 |
| ***MUSCLE DISORDERS*** | | |
| *MUSCLE INFECTIONS AND INFLAMMATIONS* | | |
| MYOSITIS | 0 | 0 |
| *MUSCLE PAINS* | | |
| FIBROMYALGIA | 0 | 0 |
| MYALGIA | 65 | 61 |
| *MUSCLE RELATED SIGNS AND SYMPTOMS NEC* | | |
| MUSCLE ATROPHY | 1 | 1 |
| MUSCLE DISCOMFORT | 0 | 0 |

| | | |
|---|---|---|
| MUSCLE FATIGUE | 2 | 2 |
| MUSCLE FIBROSIS | 1 | 1 |
| MUSCLE MASS | 0 | 0 |
| MUSCLE SPASMS | 8 | 8 |
| MUSCLE TIGHTNESS | 0 | 0 |
| MUSCLE TWITCHING | 1 | 1 |
| *MUSCLE TONE ABNORMALITIES* | | |
| MUSCLE RIGIDITY | 0 | 0 |
| TRISMUS | 0 | 0 |
| *MUSCLE WEAKNESS CONDITIONS* | | |
| MUSCULAR WEAKNESS | 3 | 3 |
| ***MUSCULOSKELETAL AND CONNECTIVE TISSUE DISORDERS NEC*** | | |
| *MUSCULOSKELETAL AND CONNECTIVE TISSUE CONDITIONS NEC* | | |
| MOBILITY DECREASED | 0 | 0 |
| MUSCULOSKELETAL STIFFNESS | 3 | 3 |
| BACK PAIN | 13 | 12 |
| FLANK PAIN | 0 | 0 |
| LIMB DISCOMFORT | 28 | 28 |
| MUSCULOSKELETAL CHEST PAIN | 4 | 4 |
| MUSCULOSKELETAL DISCOMFORT | 0 | 0 |
| MUSCULOSKELETAL PAIN | 0 | 0 |
| NECK PAIN | 6 | 6 |
| PAIN IN EXTREMITY | 198 | 182 |
| *SOFT TISSUE DISORDERS NEC* | | |
| AXILLARY MASS | 0 | 0 |
| GROIN PAIN | 1 | 1 |
| ***SYNOVIAL AND BURSAL DISORDERS*** | | |
| *SYNOVIAL DISORDERS* | | |
| SYNOVITIS | 0 | 0 |
| ***TENDON, LIGAMENT AND CARTILAGE DISORDERS*** | | |
| *CARTILAGE DISORDERS* | | |
| COSTOCHONDRITIS | 0 | 0 |
| *TENDON DISORDERS* | | |
| TENDONITIS | 0 | 0 |
| TENOSYNOVITIS | 0 | 0 |
| TRIGGER FINGER | 0 | 0 |
| **NEOPLASMS BENIGN, MALIGNANT AND UNSPECIFIED (INCL CYSTS AND POLYPS)** | | |
| ***CUTANEOUS NEOPLASMS BENIGN*** | | |
| *SKIN NEOPLASMS BENIGN* | | |
| MELANOCYTIC NAEVUS | 1 | 1 |
| SEBORRHOEIC KERATOSIS | 0 | 0 |
| SKIN PAPILLOMA | 0 | 0 |
| **NERVOUS SYSTEM DISORDERS** | | |
| ***CENTRAL NERVOUS SYSTEM INFECTIONS AND INFLAMMATIONS*** | | |
| *MYELITIS (INCL INFECTIVE)* | | |
| MYELITIS TRANSVERSE | 0 | 0 |
| ***CENTRAL NERVOUS SYSTEM VASCULAR DISORDERS*** | | |
| *CENTRAL NERVOUS SYSTEM HAEMORRHAGES AND CEREBROVASCULAR ACCIDENTS* | | |
| CEREBRAL HAEMORRHAGE | 0 | 0 |
| CEREBROVASCULAR ACCIDENT | 0 | 0 |
| *TRANSIENT CEREBROVASCULAR EVENTS* | | |
| TRANSIENT ISCHAEMIC ATTACK | 0 | 0 |

| | | |
|---|---|---|
| ***CRANIAL NERVE DISORDERS (EXCL NEOPLASMS)*** | | |
| *FACIAL CRANIAL NERVE DISORDERS* | | |
| BELL'S PALSY | 0 | 0 |
| FACIAL PARALYSIS | 0 | 0 |
| FACIAL PARESIS | 0 | 0 |
| *OLFACTORY NERVE DISORDERS* | | |
| ANOSMIA | 2 | 2 |
| HYPOSMIA | 0 | 0 |
| PAROSMIA | 0 | 0 |
| *TRIGEMINAL DISORDERS* | | |
| TRIGEMINAL NEURALGIA | 0 | 0 |
| TRIGEMINAL NEURITIS | 0 | 0 |
| ***DEMYELINATING DISORDERS*** | | |
| *MULTIPLE SCLEROSIS ACUTE AND PROGRESSIVE* | | |
| MULTIPLE SCLEROSIS RELAPSE | 0 | 0 |
| ***HEADACHES*** | | |
| *HEADACHES NEC* | | |
| CLUSTER HEADACHE | 4 | 4 |
| COLD-STIMULUS HEADACHE | 0 | 0 |
| DRUG WITHDRAWAL HEADACHE | 0 | 0 |
| HEADACHE | 293 | 274 |
| PRIMARY STABBING HEADACHE | 0 | 0 |
| SINUS HEADACHE | 1 | 1 |
| TENSION HEADACHE | 2 | 2 |
| THUNDERCLAP HEADACHE | 0 | 0 |
| VASCULAR HEADACHE | 0 | 0 |
| *MIGRAINE HEADACHES* | | |
| MIGRAINE | 6 | 6 |
| MIGRAINE WITH AURA | 0 | 0 |
| RETINAL MIGRAINE | 0 | 0 |
| TYPICAL AURA WITHOUT HEADACHE | 0 | 0 |
| ***MENTAL IMPAIRMENT DISORDERS*** | | |
| *INTELLECTUAL DISABILITIES* | | |
| INTELLECTUAL DISABILITY | 0 | 0 |
| *MEMORY LOSS (EXCL DEMENTIA)* | | |
| AMNESIA | 1 | 1 |
| MEMORY IMPAIRMENT | 0 | 0 |
| *MENTAL IMPAIRMENT (EXCL DEMENTIA AND MEMORY LOSS)* | | |
| COGNITIVE DISORDER | 0 | 0 |
| DISTURBANCE IN ATTENTION | 1 | 1 |
| MENTAL IMPAIRMENT | 0 | 0 |
| ***MOVEMENT DISORDERS (INCL PARKINSONISM)*** | | |
| *DYSKINESIAS AND MOVEMENT DISORDERS NEC* | | |
| BRADYKINESIA | 0 | 0 |
| CLUMSINESS | 0 | 0 |
| DYSKINESIA | 0 | 0 |
| EXTRAPYRAMIDAL DISORDER | 0 | 0 |
| PSYCHOMOTOR HYPERACTIVITY | 0 | 0 |
| *PARALYSIS AND PARESIS (EXCL CRANIAL NERVE)* | | |
| HEMIPLEGIA | 1 | 1 |
| MONOPARESIS | 1 | 0 |
| MONOPLEGIA | 0 | 0 |

| | | |
|---|---|---|
| PARALYSIS | 1 | 1 |
| *PARKINSON'S DISEASE AND PARKINSONISM* | | |
| FREEZING PHENOMENON | 0 | 0 |
| PARKINSON'S DISEASE | 0 | 0 |
| *TREMOR (EXCL CONGENITAL)* | | |
| TREMOR | 22 | 21 |
| ***NEUROLOGICAL DISORDERS NEC*** | | |
| *COMA STATES* | | |
| DIABETIC HYPERGLYCAEMIC COMA | 1 | 0 |
| *COORDINATION AND BALANCE DISTURBANCES* | | |
| BALANCE DISORDER | 3 | 3 |
| COORDINATION ABNORMAL | 0 | 0 |
| DYSSTASIA | 0 | 0 |
| VESTIBULAR NYSTAGMUS | 0 | 0 |
| *DISTURBANCES IN CONSCIOUSNESS NEC* | | |
| DEPRESSED LEVEL OF CONSCIOUSNESS | 1 | 1 |
| LETHARGY | 19 | 17 |
| LOSS OF CONSCIOUSNESS | 1 | 1 |
| SEDATION | 0 | 0 |
| SOMNOLENCE | 12 | 10 |
| SYNCOPE | 4 | 4 |
| *NERVOUS SYSTEM DISORDERS NEC* | | |
| NERVOUS SYSTEM DISORDER | 1 | 1 |
| *NEUROLOGICAL SIGNS AND SYMPTOMS NEC* | | |
| AGITATION NEONATAL | 1 | 0 |
| BRAIN FOG | 5 | 5 |
| DIZZINESS | 40 | 39 |
| DIZZINESS EXERTIONAL | 0 | 0 |
| DIZZINESS POSTURAL | 5 | 4 |
| HEAD DISCOMFORT | 1 | 0 |
| INFANT IRRITABILITY | 0 | 0 |
| MENINGISM | 0 | 0 |
| MYOCLONUS | 0 | 0 |
| NEUROLOGICAL SYMPTOM | 0 | 0 |
| PERSISTENT POSTURAL-PERCEPTUAL DIZZINESS | 0 | 0 |
| PRESYNCOPE | 0 | 0 |
| *PARAESTHESIAS AND DYSAESTHESIAS* | | |
| BURNING FEET SYNDROME | 0 | 0 |
| BURNING SENSATION | 4 | 4 |
| FORMICATION | 0 | 0 |
| HYPERAESTHESIA | 0 | 0 |
| HYPOAESTHESIA | 3 | 2 |
| PARAESTHESIA | 11 | 11 |
| REVERSED HOT-COLD SENSATION | 0 | 0 |
| *SENSORY ABNORMALITIES NEC* | | |
| AGEUSIA | 4 | 4 |
| ALLODYNIA | 0 | 0 |
| DYSGEUSIA | 1 | 1 |
| HYPOGEUSIA | 0 | 0 |
| NEURALGIA | 1 | 1 |
| POST HERPETIC NEURALGIA | 0 | 0 |

| | | |
|---|---|---|
| RESTLESS ARM SYNDROME | 0 | 0 |
| RESTLESS LEGS SYNDROME | 1 | 1 |
| SENSORY LOSS | 0 | 0 |
| TASTE DISORDER | 1 | 1 |
| *SPEECH AND LANGUAGE ABNORMALITIES* | | |
| DYSARTHRIA | 0 | 0 |
| SPEECH DISORDER DEVELOPMENTAL | 0 | 0 |
| ***NEUROLOGICAL DISORDERS OF THE EYE*** | | |
| *NEUROLOGIC VISUAL PROBLEMS NEC* | | |
| TUNNEL VISION | 0 | 0 |
| ***NEUROMUSCULAR DISORDERS*** | | |
| *MUSCLE TONE ABNORMAL* | | |
| HYPOTONIA | 0 | 0 |
| STIFF LEG SYNDROME | 0 | 0 |
| *NEUROMUSCULAR DISORDERS NEC* | | |
| MUSCLE SPASTICITY | 0 | 0 |
| ***PERIPHERAL NEUROPATHIES*** | | |
| *ACUTE POLYNEUROPATHIES* | | |
| GUILLAIN-BARRE SYNDROME | 0 | 0 |
| *PERIPHERAL NEUROPATHIES NEC* | | |
| AXONAL NEUROPATHY | 0 | 0 |
| NEUROPATHY PERIPHERAL | 0 | 0 |
| ***SEIZURES (INCL SUBTYPES)*** | | |
| *SEIZURES AND SEIZURE DISORDERS NEC* | | |
| EPILEPSY | 0 | 0 |
| SEIZURE | 0 | 0 |
| ***SLEEP DISTURBANCES (INCL SUBTYPES)*** | | |
| *DISTURBANCES IN SLEEP PHASE RHYTHM* | | |
| CIRCADIAN RHYTHM SLEEP DISORDER | 0 | 0 |
| *SLEEP DISTURBANCES NEC* | | |
| SLEEP DEFICIT | 0 | 0 |
| ***SPINAL CORD AND NERVE ROOT DISORDERS*** | | |
| *CERVICAL SPINAL CORD AND NERVE ROOT DISORDERS* | | |
| CERVICOBRACHIAL SYNDROME | 0 | 0 |
| *LUMBAR SPINAL CORD AND NERVE ROOT DISORDERS* | | |
| CAUDA EQUINA SYNDROME | 0 | 0 |
| SCIATICA | 1 | 1 |
| **PREGNANCY, PUERPERIUM AND PERINATAL CONDITIONS** | | |
| ***ABORTIONS AND STILLBIRTH*** | | |
| *ABORTIONS SPONTANEOUS* | | |
| ABORTION SPONTANEOUS | 0 | 0 |
| *STILLBIRTH AND FOETAL DEATH* | | |
| FOETAL DEATH | 0 | 0 |
| ***FOETAL COMPLICATIONS*** | | |
| *FOETAL COMPLICATIONS NEC* | | |
| FOETAL DISORDER | 0 | 0 |
| FOETAL HYPOKINESIA | 0 | 0 |
| *FOETAL GROWTH COMPLICATIONS* | | |
| FOETAL MACROSOMIA | 0 | 0 |
| ***MATERNAL COMPLICATIONS OF PREGNANCY*** | | |
| *MATERNAL COMPLICATIONS OF PREGNANCY NEC* | | |
| MORNING SICKNESS | 1 | 0 |
| ***PLACENTAL, AMNIOTIC AND CAVITY DISORDERS (EXCL HAEMORRHAGES)*** | | |

| | | |
|---|---|---|
| *PLACENTAL ABNORMALITIES (EXCL NEOPLASMS)* | | |
| PLACENTAL INFARCTION | 0 | 0 |
| ***PREGNANCY, LABOUR, DELIVERY AND POSTPARTUM CONDITIONS*** | | |
| *NORMAL PREGNANCY, LABOUR AND DELIVERY* | | |
| PREGNANCY | 1 | 0 |
| UTERINE CONTRACTIONS DURING PREGNANCY | 0 | 0 |
| **PRODUCT ISSUES** | | |
| ***DEVICE ISSUES*** | | |
| *DEVICE MALFUNCTION EVENTS NEC* | | |
| OVERSENSING | 0 | 0 |
| **PSYCHIATRIC DISORDERS** | | |
| ***ANXIETY DISORDERS AND SYMPTOMS*** | | |
| *ANXIETY SYMPTOMS* | | |
| AGITATION | 0 | 0 |
| ANXIETY | 4 | 4 |
| NERVOUSNESS | 2 | 2 |
| TENSION | 0 | 0 |
| *PANIC ATTACKS AND DISORDERS* | | |
| PANIC ATTACK | 1 | 1 |
| ***CHANGES IN PHYSICAL ACTIVITY*** | | |
| *INCREASED PHYSICAL ACTIVITY LEVELS* | | |
| RESTLESSNESS | 1 | 1 |
| *STEREOTYPIES AND AUTOMATISMS* | | |
| BRUXISM | 0 | 0 |
| ***COGNITIVE AND ATTENTION DISORDERS AND DISTURBANCES*** | | |
| *COGNITIVE AND ATTENTION DISORDERS AND DISTURBANCES NEC* | | |
| DAYDREAMING | 0 | 0 |
| MENTAL FATIGUE | 3 | 3 |
| ***COMMUNICATION DISORDERS AND DISTURBANCES*** | | |
| *SPEECH ARTICULATION AND RHYTHM DISTURBANCES* | | |
| DYSPHEMIA | 0 | 0 |
| ***DELIRIA (INCL CONFUSION)*** | | |
| *CONFUSION AND DISORIENTATION* | | |
| CONFUSIONAL STATE | 0 | 0 |
| DISORIENTATION | 1 | 1 |
| *DELIRIA* | | |
| DELIRIUM | 1 | 1 |
| ***DEPRESSED MOOD DISORDERS AND DISTURBANCES*** | | |
| *DEPRESSIVE DISORDERS* | | |
| DEPRESSION | 0 | 0 |
| *MOOD ALTERATIONS WITH DEPRESSIVE SYMPTOMS* | | |
| DEPRESSED MOOD | 0 | 0 |
| TEARFULNESS | 0 | 0 |
| ***DISSOCIATIVE DISORDERS*** | | |
| *DISSOCIATIVE STATES* | | |
| DISSOCIATION | 0 | 0 |
| ***DISTURBANCES IN THINKING AND PERCEPTION*** | | |
| *DELUSIONAL SYMPTOMS* | | |
| DELUSION | 1 | 1 |
| *HALLUCINATIONS (EXCL SLEEP-RELATED)* | | |
| HALLUCINATION | 1 | 1 |
| *PERCEPTION DISTURBANCES NEC* | | |
| DEREALISATION | 0 | 0 |

| | | |
|---|---|---|
| *THINKING DISTURBANCES* | | |
| BRADYPHRENIA | 0 | 0 |
| THOUGHT BLOCKING | 0 | 0 |
| ***MOOD DISORDERS AND DISTURBANCES NEC*** | | |
| *AFFECT ALTERATIONS NEC* | | |
| AFFECT LABILITY | 0 | 0 |
| INAPPROPRIATE AFFECT | 0 | 0 |
| *EMOTIONAL AND MOOD DISTURBANCES NEC* | | |
| ANGER | 0 | 0 |
| EMOTIONAL DISORDER | 0 | 0 |
| EMOTIONAL DISTRESS | 0 | 0 |
| EUPHORIC MOOD | 0 | 0 |
| IRRITABILITY | 1 | 1 |
| MOOD ALTERED | 0 | 0 |
| *FLUCTUATING MOOD SYMPTOMS* | | |
| MOOD SWINGS | 0 | 0 |
| *MOOD DISORDERS NEC* | | |
| APATHY | 0 | 0 |
| LISTLESS | 0 | 0 |
| ***PERSONALITY DISORDERS AND DISTURBANCES IN BEHAVIOUR*** | | |
| *BEHAVIOUR AND SOCIALISATION DISTURBANCES* | | |
| AGGRESSION | 0 | 0 |
| ***PSYCHIATRIC AND BEHAVIOURAL SYMPTOMS NEC*** | | |
| *PSYCHIATRIC SYMPTOMS NEC* | | |
| PSYCHIATRIC SYMPTOM | 1 | 1 |
| ***PSYCHIATRIC DISORDERS NEC*** | | |
| *MENTAL DISORDERS NEC* | | |
| MENTAL DISORDER | 0 | 0 |
| ***SCHIZOPHRENIA AND OTHER PSYCHOTIC DISORDERS*** | | |
| *PSYCHOTIC DISORDER NEC* | | |
| PSYCHOTIC DISORDER | 0 | 0 |
| ***SEXUAL DYSFUNCTIONS, DISTURBANCES AND GENDER IDENTITY DISORDERS*** | | |
| *SEXUAL DESIRE DISORDERS* | | |
| LOSS OF LIBIDO | 0 | 0 |
| ***SLEEP DISORDERS AND DISTURBANCES*** | | |
| *DISTURBANCES IN INITIATING AND MAINTAINING SLEEP* | | |
| INITIAL INSOMNIA | 0 | 0 |
| INSOMNIA | 4 | 4 |
| *DYSSOMNIAS* | | |
| BREATHING-RELATED SLEEP DISORDER | 0 | 0 |
| POOR QUALITY SLEEP | 0 | 0 |
| *PARASOMNIAS* | | |
| ABNORMAL DREAMS | 3 | 3 |
| EXPLODING HEAD SYNDROME | 0 | 0 |
| NIGHTMARE | 1 | 1 |
| *SLEEP DISORDERS NEC* | | |
| SLEEP DISORDER | 0 | 0 |
| ***SOMATIC SYMPTOM AND RELATED DISORDERS*** | | |
| *SOMATIC SYMPTOM DISORDERS* | | |
| HABIT COUGH | 0 | 0 |
| ***SUICIDAL AND SELF-INJURIOUS BEHAVIOURS NEC*** | | |
| *SUICIDAL AND SELF-INJURIOUS BEHAVIOUR* | | |
| SUICIDAL IDEATION | 0 | 0 |

| | | |
|---|---|---|
| **RENAL AND URINARY DISORDERS** | | |
| ***RENAL DISORDERS (EXCL NEPHROPATHIES)*** | | |
| *RENAL FAILURE AND IMPAIRMENT* | | |
| RENAL FAILURE | 0 | 0 |
| ***URINARY TRACT SIGNS AND SYMPTOMS*** | | |
| *BLADDER AND URETHRAL SYMPTOMS* | | |
| BLADDER PAIN | 1 | 1 |
| MICTURITION URGENCY | 0 | 0 |
| POLLAKIURIA | 1 | 1 |
| URINARY INCONTINENCE | 0 | 0 |
| *URINARY ABNORMALITIES* | | |
| HAEMATURIA | 0 | 0 |
| URINE ABNORMALITY | 0 | 0 |
| URINE ODOUR ABNORMAL | 0 | 0 |
| *URINARY TRACT SIGNS AND SYMPTOMS NEC* | | |
| POLYURIA | 0 | 0 |
| RENAL PAIN | 1 | 1 |
| **REPRODUCTIVE SYSTEM AND BREAST DISORDERS** | | |
| ***BREAST DISORDERS*** | | |
| *BREAST DISORDERS NEC* | | |
| BREAST MASS | 1 | 1 |
| NIPPLE ENLARGEMENT | 0 | 0 |
| *BREAST SIGNS AND SYMPTOMS* | | |
| BREAST DISCHARGE | 1 | 1 |
| BREAST PAIN | 2 | 2 |
| BREAST SWELLING | 0 | 0 |
| BREAST TENDERNESS | 0 | 0 |
| NIPPLE PAIN | 0 | 0 |
| ***MENOPAUSE RELATED CONDITIONS*** | | |
| *MENOPAUSAL EFFECTS ON THE GENITOURINARY TRACT* | | |
| POSTMENOPAUSAL HAEMORRHAGE | 0 | 0 |
| ***MENSTRUAL CYCLE AND UTERINE BLEEDING DISORDERS*** | | |
| *MENSTRUATION AND UTERINE BLEEDING NEC* | | |
| DYSMENORRHOEA | 3 | 3 |
| INTERMENSTRUAL BLEEDING | 1 | 1 |
| MENSTRUAL DISORDER | 1 | 1 |
| MENSTRUATION IRREGULAR | 7 | 6 |
| PREMENSTRUAL PAIN | 1 | 1 |
| RETROGRADE MENSTRUATION | 0 | 0 |
| *MENSTRUATION WITH DECREASED BLEEDING* | | |
| AMENORRHOEA | 0 | 0 |
| HYPOMENORRHOEA | 3 | 3 |
| MENSTRUATION DELAYED | 12 | 11 |
| OLIGOMENORRHOEA | 1 | 1 |
| *MENSTRUATION WITH INCREASED BLEEDING* | | |
| HEAVY MENSTRUAL BLEEDING | 9 | 7 |
| POLYMENORRHOEA | 0 | 0 |
| ***PENILE AND SCROTAL DISORDERS (EXCL INFECTIONS AND INFLAMMATIONS)*** | | |
| *SCROTAL DISORDERS NEC* | | |
| SCROTAL SWELLING | 0 | 0 |
| ***REPRODUCTIVE TRACT DISORDERS NEC*** | | |
| *REPRODUCTIVE TRACT DISORDERS NEC (EXCL NEOPLASMS)* | | |
| GENITAL LESION | 0 | 0 |

| | | |
|---|---|---|
| *REPRODUCTIVE TRACT SIGNS AND SYMPTOMS NEC* | | |
| GENITAL DISCOMFORT | 0 | 0 |
| GENITAL PAIN | 0 | 0 |
| ***UTERINE, PELVIC AND BROAD LIGAMENT DISORDERS*** | | |
| *PELVIS AND BROAD LIGAMENT DISORDERS NEC* | | |
| ADNEXA UTERI PAIN | 0 | 0 |
| ***VULVOVAGINAL DISORDERS (EXCL INFECTIONS AND INFLAMMATIONS)*** | | |
| *VULVOVAGINAL DISORDERS NEC* | | |
| VAGINAL HAEMORRHAGE | 0 | 0 |
| *VULVOVAGINAL SIGNS AND SYMPTOMS* | | |
| VAGINAL DISCHARGE | 0 | 0 |
| **RESPIRATORY, THORACIC AND MEDIASTINAL DISORDERS** | | |
| ***BRONCHIAL DISORDERS (EXCL NEOPLASMS)*** | | |
| *BRONCHOSPASM AND OBSTRUCTION* | | |
| ASTHMA | 0 | 0 |
| WHEEZING | 1 | 1 |
| ***LOWER RESPIRATORY TRACT DISORDERS (EXCL OBSTRUCTION AND INFECTION)*** | | |
| *PULMONARY OEDEMAS* | | |
| PULMONARY CONGESTION | 0 | 0 |
| ***PULMONARY VASCULAR DISORDERS*** | | |
| *PULMONARY THROMBOTIC AND EMBOLIC CONDITIONS* | | |
| PULMONARY EMBOLISM | 1 | 1 |
| ***RESPIRATORY DISORDERS NEC*** | | |
| *BREATHING ABNORMALITIES* | | |
| DYSPNOEA | 14 | 13 |
| HYPERVENTILATION | 0 | 0 |
| HYPOPNOEA | 1 | 1 |
| IRREGULAR BREATHING | 0 | 0 |
| MOUTH BREATHING | 0 | 0 |
| RESPIRATION ABNORMAL | 0 | 0 |
| RESPIRATORY ARREST | 0 | 0 |
| RESPIRATORY FATIGUE | 0 | 0 |
| SLEEP APNOEA SYNDROME | 0 | 0 |
| *COUGHING AND ASSOCIATED SYMPTOMS* | | |
| COUGH | 6 | 6 |
| PRODUCTIVE COUGH | 2 | 1 |
| *RESPIRATORY TRACT DISORDERS NEC* | | |
| RESPIRATORY TRACT IRRITATION | 0 | 0 |
| ***RESPIRATORY TRACT SIGNS AND SYMPTOMS*** | | |
| *LOWER RESPIRATORY TRACT SIGNS AND SYMPTOMS* | | |
| PULMONARY PAIN | 0 | 0 |
| *RESPIRATORY SIGNS AND SYMPTOMS NEC* | | |
| RESPIRATORY SYMPTOM | 0 | 0 |
| *UPPER RESPIRATORY TRACT SIGNS AND SYMPTOMS* | | |
| APHONIA | 0 | 0 |
| CATARRH | 1 | 1 |
| DRY THROAT | 1 | 1 |
| DYSPHONIA | 0 | 0 |
| INCREASED UPPER AIRWAY SECRETION | 0 | 0 |
| INCREASED VISCOSITY OF UPPER RESPIRATORY SE( | 0 | 0 |
| NASAL DISCOMFORT | 0 | 0 |
| OROPHARYNGEAL DISCOMFORT | 0 | 0 |
| OROPHARYNGEAL PAIN | 11 | 11 |

| | | |
|---|---|---|
| PARANASAL SINUS DISCOMFORT | 0 | 0 |
| RHINALGIA | 0 | 0 |
| RHINORRHOEA | 8 | 8 |
| SINUS PAIN | 0 | 0 |
| SNEEZING | 2 | 2 |
| THROAT CLEARING | 0 | 0 |
| THROAT IRRITATION | 1 | 1 |
| THROAT TIGHTNESS | 0 | 0 |
| UPPER-AIRWAY COUGH SYNDROME | 0 | 0 |
| YAWNING | 1 | 1 |
| ***UPPER RESPIRATORY TRACT DISORDERS (EXCL INFECTIONS)*** | | |
| *NASAL CONGESTION AND INFLAMMATIONS* | | |
| NASAL CONGESTION | 0 | 0 |
| RHINITIS ALLERGIC | 0 | 0 |
| RHINITIS ATROPHIC | 0 | 0 |
| *NASAL DISORDERS NEC* | | |
| EPISTAXIS | 0 | 0 |
| NASAL DRYNESS | 0 | 0 |
| *PARANASAL SINUS DISORDERS (EXCL INFECTIONS AND NEOPLASMS)* | | |
| SINUS CONGESTION | 0 | 0 |
| *PHARYNGEAL DISORDERS (EXCL INFECTIONS AND NEOPLASMS)* | | |
| PHARYNGEAL SWELLING | 0 | 0 |
| PHARYNGEAL ULCERATION | 0 | 0 |
| TONSILLAR ERYTHEMA | 1 | 1 |
| *TRACHEAL DISORDERS (EXCL INFECTIONS AND NEOPLASMS)* | | |
| TRACHEAL PAIN | 0 | 0 |
| **SKIN AND SUBCUTANEOUS TISSUE DISORDERS** | | |
| ***ANGIOEDEMA AND URTICARIA*** | | |
| *ANGIOEDEMAS* | | |
| ANGIOEDEMA | 0 | 0 |
| *URTICARIAS* | | |
| COLD URTICARIA | 0 | 0 |
| SOLAR URTICARIA | 0 | 0 |
| URTICARIA | 0 | 0 |
| URTICARIA CHRONIC | 1 | 1 |
| ***CORNIFICATION AND DYSTROPHIC SKIN DISORDERS*** | | |
| *SKIN DYSTROPHIES* | | |
| HYPERTROPHIC SCAR | 0 | 0 |
| ***CUTANEOUS NEOPLASMS BENIGN*** | | |
| *SKIN CYSTS AND POLYPS* | | |
| DERMAL CYST | 0 | 0 |
| ***EPIDERMAL AND DERMAL CONDITIONS*** | | |
| *BULLOUS CONDITIONS* | | |
| BLISTER | 1 | 1 |
| BLOOD BLISTER | 1 | 1 |
| PEMPHIGOID | 0 | 0 |
| TOXIC EPIDERMAL NECROLYSIS | 0 | 0 |
| *DERMAL AND EPIDERMAL CONDITIONS NEC* | | |
| DRY SKIN | 4 | 3 |
| PAIN OF SKIN | 3 | 3 |
| SENSITIVE SKIN | 2 | 2 |
| SKIN BURNING SENSATION | 2 | 2 |

| | | |
|---|---|---|
| SKIN DISCOLOURATION | 0 | 0 |
| SKIN FRAGILITY | 0 | 0 |
| SKIN LESION | 1 | 0 |
| SKIN ODOUR ABNORMAL | 0 | 0 |
| SKIN REACTION | 0 | 0 |
| SKIN SENSITISATION | 0 | 0 |
| SKIN WARM | 1 | 1 |
| *DERMATITIS AND ECZEMA* | | |
| DERMATITIS | 0 | 0 |
| DERMATITIS ALLERGIC | 0 | 0 |
| DERMATITIS ATOPIC | 0 | 0 |
| DERMATITIS CONTACT | 0 | 0 |
| ECZEMA | 2 | 1 |
| ECZEMA ASTEATOTIC | 0 | 0 |
| NEURODERMATITIS | 0 | 0 |
| SEBORRHOEIC DERMATITIS | 0 | 0 |
| SKIN IRRITATION | 0 | 0 |
| *ERYTHEMAS* | | |
| ERYTHEMA | 5 | 4 |
| *EXFOLIATIVE CONDITIONS* | | |
| SKIN EXFOLIATION | 0 | 0 |
| *PAPULOSQUAMOUS CONDITIONS* | | |
| LICHEN PLANUS | 0 | 0 |
| PITYRIASIS ROSEA | 0 | 0 |
| *PHOTOSENSITIVITY AND PHOTODERMATOSIS CONDITIONS* | | |
| PHOTOSENSITIVITY REACTION | 0 | 0 |
| *PRURITUS NEC* | | |
| PRURITUS | 13 | 12 |
| *PSORIATIC CONDITIONS* | | |
| PSORIASIS | 1 | 1 |
| *RASHES, ERUPTIONS AND EXANTHEMS NEC* | | |
| RASH | 4 | 4 |
| RASH ERYTHEMATOUS | 4 | 3 |
| RASH MACULAR | 0 | 0 |
| RASH MORBILLIFORM | 0 | 0 |
| RASH PAPULAR | 1 | 1 |
| RASH PRURITIC | 3 | 3 |
| *SKIN INJURIES AND MECHANICAL DERMATOSES* | | |
| DECUBITUS ULCER | 0 | 0 |
| NEEDLE TRACK MARKS | 0 | 0 |
| ***SKIN AND SUBCUTANEOUS TISSUE DISORDERS NEC*** | | |
| *SKIN AND SUBCUTANEOUS TISSUE ULCERATIONS* | | |
| SKIN EROSION | 0 | 0 |
| ***SKIN APPENDAGE CONDITIONS*** | | |
| *ACNES* | | |
| ACNE | 0 | 0 |
| ACNE CYSTIC | 0 | 0 |
| *ALOPECIAS* | | |
| ALOPECIA | 5 | 4 |
| *APOCRINE AND ECCRINE GLAND DISORDERS* | | |
| COLD SWEAT | 0 | 0 |
| HYPERHIDROSIS | 19 | 19 |

| | | |
|---|---|---|
| MILIARIA | 1 | 1 |
| NIGHT SWEATS | 3 | 3 |
| *HYPERTRICHOSES* | | |
| HIRSUTISM | 0 | 0 |
| *NAIL AND NAIL BED CONDITIONS (EXCL INFECTIONS AND INFESTATIONS)* | | |
| NAIL DISCOLOURATION | 1 | 1 |
| ONYCHOCLASIS | 1 | 0 |
| *PILAR DISORDERS NEC* | | |
| PILOERECTION | 0 | 0 |
| ***SKIN VASCULAR ABNORMALITIES*** | | |
| *PURPURA AND RELATED CONDITIONS* | | |
| PETECHIAE | 0 | 0 |
| PURPURA | 0 | 0 |
| *SKIN HAEMORRHAGES* | | |
| SKIN HAEMORRHAGE | 0 | 0 |
| **SOCIAL CIRCUMSTANCES** | | |
| ***ECONOMIC AND HOUSING ISSUES*** | | |
| *EMPLOYMENT ISSUES* | | |
| RETIREMENT | 0 | 0 |
| ***ENVIRONMENTAL ISSUES*** | | |
| *NON-OCCUPATIONAL AND UNSPECIFIED ENVIRONMENTAL PROBLEMS* | | |
| POLLUTION | 0 | 0 |
| **SURGICAL AND MEDICAL PROCEDURES** | | |
| ***BONE AND JOINT THERAPEUTIC PROCEDURES*** | | |
| *JOINT THERAPEUTIC PROCEDURES* | | |
| KNEE OPERATION | 0 | 0 |
| ***BREAST THERAPEUTIC PROCEDURES*** | | |
| *MASTECTOMIES* | | |
| BREAST CONSERVING SURGERY | 0 | 0 |
| ***GASTROINTESTINAL THERAPEUTIC PROCEDURES*** | | |
| *GASTROINTESTINAL THERAPEUTIC PROCEDURES NEC* | | |
| PROPHYLAXIS OF NAUSEA AND VOMITING | 0 | 0 |
| ***HEAD AND NECK THERAPEUTIC PROCEDURES*** | | |
| *DENTAL AND GINGIVAL THERAPEUTIC PROCEDURES* | | |
| DENTAL CARE | 0 | 0 |
| ***NERVOUS SYSTEM, SKULL AND SPINE THERAPEUTIC PROCEDURES*** | | |
| *SKULL AND BRAIN THERAPEUTIC PROCEDURES* | | |
| BRAIN TUMOUR OPERATION | 0 | 0 |
| ***OBSTETRIC AND GYNAECOLOGICAL THERAPEUTIC PROCEDURES*** | | |
| *FERTILITY AND FERTILISATION INTERVENTIONS FEMALE* | | |
| ENDOMETRIAL SCRATCHING | 0 | 0 |
| ***RESPIRATORY TRACT THERAPEUTIC PROCEDURES*** | | |
| *RESPIRATORY TRACT THERAPEUTIC PROCEDURES NEC* | | |
| OXYGEN THERAPY | 1 | 1 |
| ***THERAPEUTIC PROCEDURES AND SUPPORTIVE CARE NEC*** | | |
| *ANAESTHESIA AND ALLIED PROCEDURES* | | |
| NERVE BLOCK | 0 | 0 |
| *DIETARY AND NUTRITIONAL THERAPIES* | | |
| NOTHING BY MOUTH ORDER | 1 | 1 |
| *IMMUNISATIONS* | | |
| COVID-19 IMMUNISATION | 0 | 0 |
| *THERAPEUTIC PROCEDURES NEC* | | |
| INJECTION | 0 | 0 |

| | | |
|---|---|---|
| LOCALISED ALTERNATING HOT AND COLD THERAPY | 0 | 0 |
| MASS EXCISION | 0 | 0 |
| REINFUSION | 0 | 0 |
| **VASCULAR DISORDERS** | | |
| ***ARTERIOSCLEROSIS, STENOSIS, VASCULAR INSUFFICIENCY AND NECROSIS*** | | |
| *NON-SITE SPECIFIC NECROSIS AND VASCULAR INSUFFICIENCY NEC* | | |
| VASOSPASM | 0 | 0 |
| *PERIPHERAL VASOCONSTRICTION, NECROSIS AND VASCULAR INSUFFICIENCY* | | |
| PERIPHERAL COLDNESS | 2 | 2 |
| RAYNAUD'S PHENOMENON | 0 | 0 |
| ***DECREASED AND NONSPECIFIC BLOOD PRESSURE DISORDERS AND SHOCK*** | | |
| *BLOOD PRESSURE DISORDERS NEC* | | |
| BLOOD PRESSURE FLUCTUATION | 0 | 0 |
| *VASCULAR HYPOTENSIVE DISORDERS* | | |
| CAPILLARY LEAK SYNDROME | 0 | 0 |
| HYPOTENSION | 0 | 0 |
| ORTHOSTATIC HYPOTENSION | 0 | 0 |
| ***EMBOLISM AND THROMBOSIS*** | | |
| *NON-SITE SPECIFIC EMBOLISM AND THROMBOSIS* | | |
| EMBOLISM | 0 | 0 |
| THROMBOSIS | 1 | 1 |
| VENOUS THROMBOSIS | 0 | 0 |
| *PERIPHERAL EMBOLISM AND THROMBOSIS* | | |
| BLUE TOE SYNDROME | 0 | 0 |
| DEEP VEIN THROMBOSIS | 1 | 1 |
| SUPERFICIAL VEIN THROMBOSIS | 1 | 1 |
| ***LYMPHATIC VESSEL DISORDERS*** | | |
| *LYMPHOEDEMAS* | | |
| LYMPHOEDEMA | 0 | 0 |
| ***VASCULAR DISORDERS NEC*** | | |
| *NON-SITE SPECIFIC VASCULAR DISORDERS NEC* | | |
| VASCULAR PAIN | 0 | 0 |
| VEIN DISCOLOURATION | 1 | 1 |
| VEIN RUPTURE | 0 | 0 |
| *PERIPHERAL VASCULAR DISORDERS NEC* | | |
| FLUSHING | 1 | 1 |
| HOT FLUSH | 8 | 8 |
| *SITE SPECIFIC VASCULAR DISORDERS NEC* | | |
| PALLOR | 2 | 2 |
| ***VASCULAR HAEMORRHAGIC DISORDERS*** | | |
| *HAEMORRHAGES NEC* | | |
| HAEMATOMA | 0 | 0 |
| HAEMORRHAGE | 1 | 1 |
| ***VASCULAR HYPERTENSIVE DISORDERS*** | | |
| *ACCELERATED AND MALIGNANT HYPERTENSION* | | |
| HYPERTENSIVE URGENCY | 0 | 0 |
| *VASCULAR HYPERTENSIVE DISORDERS NEC* | | |
| HYPERTENSION | 1 | 1 |
| SYSTOLIC HYPERTENSION | 0 | 0 |
| ***VASCULAR INFECTIONS AND INFLAMMATIONS*** | | |
| *ARTERIAL INFECTIONS AND INFLAMMATIONS* | | |
| GIANT CELL ARTERITIS | 0 | 0 |

| | | |
|---|---|---|
| *PHLEBITIS NEC* | | |
| PHLEBITIS | 0 | 0 |
| *VASCULITIDES NEC* | | |
| VASCULITIS | 0 | 0 |
| ***VENOUS VARICES*** | | |
| *VARICOSE VEINS NEC* | | |
| SPIDER VEIN | 0 | 0 |
| VARICOSE VEIN | 0 | 0 |
| **TOTAL ADR EVENTS** | ***2102*** | ***1980*** |
