## Supplementary Material S2. ADR listing in Pregnant and Breastfeeding Cohort for "Implementation and results of active vaccine safety monitoring during the COVID-19 pandemic in the UK"

### Supplementary Material S2. Adverse Events (ADRs) Reported during Pregnancy or Breast-feeding

Total ADR events reported

The tables S2-1 to S2-12 listed below are presented in this file named "Supplementary _S2-ADR_Listing_during_Pregnancy_Breastfeeding". Each of these tables capture the full listing of ADRs reported by MedDRA SOC, HLGT, HLT and PT level terms for each brand and dose within the cohorts of patients indicating they were (i) pregnant or (ii) breastfeeding at the time of a vaccination.

| Table Number | Description |
|---|---|
| | **(i) PREGNANT PATIENTS** |
| **S2-1** | ADR listing for events reported by pregnant patients, in those reporting any vaccination dose. |
| **S2-2** | Pfizer BioNTech COVID-19 vaccine: ADR listing for events reported by pregnant patients, in those reporting any vaccination dose. |
| **S2-3** | AstraZeneca COVID-19 vaccine: ADR listing for events reported by pregnant patients, in those reporting any vaccination dose. |
| **S2-4** | Moderna COVID-19 vaccine: ADR listing for events reported by pregnant patients, in those reporting any vaccination dose. |
| **S2-5** | Others/Unknown COVID-19 vaccine: ADR listings for events reported by pregnant patients, in reporting any vaccination dose. |
| **S2-6** | COVID-19 vaccine ADR listings for events reported by pregnant patients with no dose identification information in those reporting any vaccination dose. |
| | **(ii) BREAST-FEEDING PATIENTS** |
| **S2-7** | ADR listing for events reported by breast-feeding patients, in those reporting any vaccination dose. |
| **S2-8** | Pfizer BioNTech COVID-19 vaccine: ADR listing for events reported by breast-feeding patients, in those reporting any vaccination dose. |
| **S2-9** | AstraZeneca COVID-19 vaccine: ADR listing for events reported by breast-feeding patients, in those reporting any vaccination dose. |
| **S2-10** | Moderna COVID-19 vaccine: ADR listing for events reported by breast-feeding patients, in those reporting any vaccination dose. |
| **S2-11** | Others/Unknown COVID-19 vaccine: ADR listings for events reported by breast-feeding patients, in reporting any vaccination dose. |
| **S2-12** | COVID-19 vaccine ADR listings for events reported by breast-feeding patients with no dose identification information in those reporting any vaccination dose. |

**TABLE S2-1. ADR listing for events reported by pregnant patients in those reporting any vaccination dose.**

| | Patients with any vaccination dose: ADR Counts | | | | | |
|---|---|---|---|---|---|---|
| **REACTION TERM (SOC, *HLGT*, *HLT*, PT)** | All doses | 1st dose | 2nd dose | 3rd dose | Other doses | Unknown |
| ***(freetext)*** | 0 | 0 | 0 | 0 | 0 | 0 |
| **BLOOD AND LYMPHATIC SYSTEM DISORDERS** | | | | | | |
| ***SPLEEN, LYMPHATIC AND RETICULOENDOTHELIAL SYSTEM DISORDERS*** | | | | | | |
| *LYMPHATIC SYSTEM DISORDERS NEC* | | | | | | |
| LYMPH NODE PAIN | 4 | 0 | 2 | 1 | 0 | 1 |
| LYMPHADENITIS | 2 | 1 | 1 | 0 | 0 | 0 |
| LYMPHADENOPATHY | 11 | 7 | 4 | 0 | 0 | 0 |
| **CARDIAC DISORDERS** | | | | | | |
| ***CARDIAC ARRHYTHMIAS*** | | | | | | |
| *RATE AND RHYTHM DISORDERS NEC* | | | | | | |
| TACHYCARDIA | 1 | 0 | 1 | 0 | 0 | 0 |
| ***CARDIAC DISORDERS, SIGNS AND SYMPTOMS NEC*** | | | | | | |
| *CARDIAC SIGNS AND SYMPTOMS NEC* | | | | | | |
| PALPITATIONS | 5 | 2 | 3 | 0 | 0 | 0 |
| **EAR AND LABYRINTH DISORDERS** | | | | | | |
| ***AURAL DISORDERS NEC*** | | | | | | |
| *EAR DISORDERS NEC* | | | | | | |
| EAR PAIN | 1 | 1 | 0 | 0 | 0 | 0 |
| ***INNER EAR AND VIIITH CRANIAL NERVE DISORDERS*** | | | | | | |
| *INNER EAR SIGNS AND SYMPTOMS* | | | | | | |
| TINNITUS | 2 | 0 | 1 | 1 | 0 | 0 |
| VERTIGO | 1 | 1 | 0 | 0 | 0 | 0 |
| **EYE DISORDERS** | | | | | | |
| ***EYE DISORDERS NEC*** | | | | | | |
| *OCULAR DISORDERS NEC* | | | | | | |
| EYE PAIN | 3 | 2 | 1 | 0 | 0 | 0 |
| ***OCULAR SENSORY SYMPTOMS NEC*** | | | | | | |
| *OCULAR SENSATION DISORDERS* | | | | | | |
| ASTHENOPIA | 2 | 1 | 1 | 0 | 0 | 0 |
| PHOTOPHOBIA | 2 | 2 | 0 | 0 | 0 | 0 |
| ***VISION DISORDERS*** | | | | | | |
| *VISUAL DISORDERS NEC* | | | | | | |
| VISION BLURRED | 4 | 2 | 2 | 0 | 0 | 0 |
| **GASTROINTESTINAL DISORDERS** | | | | | | |
| ***DENTAL AND GINGIVAL CONDITIONS*** | | | | | | |
| *DENTAL DISORDERS NEC* | | | | | | |
| TEETHING | 1 | 1 | 0 | 0 | 0 | 0 |
| ***GASTROINTESTINAL INFLAMMATORY CONDITIONS*** | | | | | | |
| *GASTROINTESTINAL INFLAMMATORY DISORDERS NEC* | | | | | | |
| GASTROINTESTINAL TRACT IRRITATION | 1 | 0 | 0 | 1 | 0 | 0 |
| ***GASTROINTESTINAL MOTILITY AND DEFAECATION CONDITIONS*** | | | | | | |
| *DIARRHOEA (EXCL INFECTIVE)* | | | | | | |
| DIARRHOEA | 15 | 9 | 3 | 3 | 0 | 0 |
| *GASTROINTESTINAL ATONIC AND HYPOMOTILITY DISORDERS NEC* | | | | | | |

| | | | | | | |
|---|---|---|---|---|---|---|
| CONSTIPATION | 1 | 0 | 0 | 1 | 0 | 0 |
| ***GASTROINTESTINAL SIGNS AND SYMPTOMS*** | | | | | | |
| *GASTROINTESTINAL AND ABDOMINAL PAINS (EXCL ORAL AND THROAT)* | | | | | | |
| ABDOMINAL PAIN | 1 | 1 | 0 | 0 | 0 | 0 |
| ABDOMINAL PAIN UPPER | 4 | 3 | 1 | 0 | 0 | 0 |
| *GASTROINTESTINAL SIGNS AND SYMPTOMS NEC* | | | | | | |
| ABDOMINAL DISCOMFORT | 2 | 2 | 0 | 0 | 0 | 0 |
| *NAUSEA AND VOMITING SYMPTOMS* | | | | | | |
| NAUSEA | 73 | 43 | 23 | 4 | 0 | 3 |
| VOMITING | 22 | 14 | 7 | 0 | 0 | 1 |
| ***ORAL SOFT TISSUE CONDITIONS*** | | | | | | |
| *ORAL SOFT TISSUE SIGNS AND SYMPTOMS* | | | | | | |
| HYPOAESTHESIA ORAL | 1 | 1 | 0 | 0 | 0 | 0 |
| PARAESTHESIA ORAL | 1 | 1 | 0 | 0 | 0 | 0 |
| ***TONGUE CONDITIONS*** | | | | | | |
| *TONGUE SIGNS AND SYMPTOMS* | | | | | | |
| GLOSSODYNIA | 1 | 1 | 0 | 0 | 0 | 0 |
| **GENERAL DISORDERS AND ADMINISTRATION SITE CONDITIONS** | | | | | | |
| ***ADMINISTRATION SITE REACTIONS*** | | | | | | |
| *ADMINISTRATION SITE REACTIONS NEC* | | | | | | |
| PUNCTURE SITE BRUISE | 2 | 0 | 0 | 1 | 0 | 1 |
| PUNCTURE SITE PAIN | 1 | 1 | 0 | 0 | 0 | 0 |
| *INJECTION SITE REACTIONS* | | | | | | |
| INJECTION SITE BRUISING | 1 | 0 | 1 | 0 | 0 | 0 |
| INJECTION SITE ERYTHEMA | 1 | 1 | 0 | 0 | 0 | 0 |
| INJECTION SITE MASS | 3 | 1 | 1 | 0 | 0 | 1 |
| INJECTION SITE PAIN | 49 | 36 | 9 | 3 | 0 | 1 |
| INJECTION SITE REACTION | 1 | 0 | 1 | 0 | 0 | 0 |
| INJECTION SITE URTICARIA | 1 | 0 | 1 | 0 | 0 | 0 |
| INJECTION SITE WARMTH | 1 | 1 | 0 | 0 | 0 | 0 |
| *VACCINATION SITE REACTIONS* | | | | | | |
| VACCINATION SITE PAIN | 4 | 2 | 1 | 0 | 0 | 1 |
| ***BODY TEMPERATURE CONDITIONS*** | | | | | | |
| *FEBRILE DISORDERS* | | | | | | |
| PYREXIA | 121 | 74 | 33 | 8 | 0 | 6 |
| ***GENERAL SYSTEM DISORDERS NEC*** | | | | | | |
| *ASTHENIC CONDITIONS* | | | | | | |
| ASTHENIA | 6 | 4 | 0 | 1 | 0 | 1 |
| FATIGUE | 276 | 167 | 74 | 17 | 1 | 17 |
| MALAISE | 16 | 8 | 6 | 1 | 1 | 0 |
| *FEELINGS AND SENSATIONS NEC* | | | | | | |
| CHILLS | 59 | 35 | 16 | 7 | 0 | 1 |
| FEELING ABNORMAL | 4 | 2 | 1 | 1 | 0 | 0 |
| FEELING COLD | 3 | 2 | 1 | 0 | 0 | 0 |
| FEELING HOT | 1 | 1 | 0 | 0 | 0 | 0 |
| FEELING OF BODY TEMPERATURE CHANGE | 1 | 1 | 0 | 0 | 0 | 0 |
| HANGOVER | 1 | 0 | 0 | 1 | 0 | 0 |
| HUNGER | 1 | 1 | 0 | 0 | 0 | 0 |
| THIRST | 2 | 2 | 0 | 0 | 0 | 0 |
| *GENERAL SIGNS AND SYMPTOMS NEC* | | | | | | |
| CRYING | 1 | 1 | 0 | 0 | 0 | 0 |
| ILLNESS | 12 | 8 | 3 | 1 | 0 | 0 |

| | | | | | | |
|---|---|---|---|---|---|---|
| INFLUENZA LIKE ILLNESS | 16 | 9 | 5 | 1 | 0 | 1 |
| LOCAL REACTION | 1 | 0 | 1 | 0 | 0 | 0 |
| PERIPHERAL SWELLING | 4 | 3 | 0 | 1 | 0 | 0 |
| SWELLING | 4 | 2 | 1 | 1 | 0 | 0 |
| *PAIN AND DISCOMFORT NEC* | | | | | | |
| AXILLARY PAIN | 10 | 3 | 5 | 2 | 0 | 0 |
| CHEST PAIN | 1 | 1 | 0 | 0 | 0 | 0 |
| PAIN | 67 | 35 | 22 | 5 | 0 | 5 |
| TENDERNESS | 10 | 8 | 2 | 0 | 0 | 0 |
| **INFECTIONS AND INFESTATIONS** | | | | | | |
| ***BACTERIAL INFECTIOUS DISORDERS*** | | | | | | |
| *BACTERIAL INFECTIONS NEC* | | | | | | |
| PERIORBITAL CELLULITIS | 1 | 0 | 1 | 0 | 0 | 0 |
| ***INFECTIONS - PATHOGEN UNSPECIFIED*** | | | | | | |
| *BREAST INFECTIONS* | | | | | | |
| MASTITIS | 1 | 0 | 0 | 1 | 0 | 0 |
| *UPPER RESPIRATORY TRACT INFECTIONS* | | | | | | |
| LARYNGITIS | 2 | 1 | 1 | 0 | 0 | 0 |
| NASOPHARYNGITIS | 12 | 7 | 4 | 1 | 0 | 0 |
| ***VIRAL INFECTIOUS DISORDERS*** | | | | | | |
| *HERPES VIRAL INFECTIONS* | | | | | | |
| GENITAL HERPES | 1 | 1 | 0 | 0 | 0 | 0 |
| HERPES ZOSTER | 1 | 1 | 0 | 0 | 0 | 0 |
| ORAL HERPES | 1 | 0 | 1 | 0 | 0 | 0 |
| *INFLUENZA VIRAL INFECTIONS* | | | | | | |
| INFLUENZA | 24 | 12 | 9 | 3 | 0 | 0 |
| *VIRAL INFECTIONS NEC* | | | | | | |
| VIRAL DIARRHOEA | 1 | 1 | 0 | 0 | 0 | 0 |
| **INJURY, POISONING AND PROCEDURAL COMPLICATIONS** | | | | | | |
| ***INJURIES NEC*** | | | | | | |
| *SKIN INJURIES NEC* | | | | | | |
| CONTUSION | 7 | 7 | 0 | 0 | 0 | 0 |
| ***PROCEDURAL RELATED INJURIES AND COMPLICATIONS NEC*** | | | | | | |
| *NON-SITE SPECIFIC PROCEDURAL COMPLICATIONS* | | | | | | |
| INJECTION RELATED REACTION | 1 | 0 | 0 | 1 | 0 | 0 |
| **INVESTIGATIONS** | | | | | | |
| ***CARDIAC AND VASCULAR INVESTIGATIONS (EXCL ENZYME TESTS)*** | | | | | | |
| *HEART RATE AND PULSE INVESTIGATIONS* | | | | | | |
| HEART RATE | 2 | 2 | 0 | 0 | 0 | 0 |
| HEART RATE INCREASED | 1 | 1 | 0 | 0 | 0 | 0 |
| ***PHYSICAL EXAMINATION AND ORGAN SYSTEM STATUS TOPICS*** | | | | | | |
| *PHYSICAL EXAMINATION PROCEDURES AND ORGAN SYSTEM STATUS* | | | | | | |
| BODY TEMPERATURE INCREASED | 2 | 1 | 0 | 1 | 0 | 0 |
| ***WATER, ELECTROLYTE AND MINERAL INVESTIGATIONS*** | | | | | | |
| *WATER AND ELECTROLYTE ANALYSES NEC* | | | | | | |
| VOLUME BLOOD | 1 | 0 | 1 | 0 | 0 | 0 |
| **METABOLISM AND NUTRITION DISORDERS** | | | | | | |
| ***APPETITE AND GENERAL NUTRITIONAL DISORDERS*** | | | | | | |

| | | | | | | |
|---|---|---|---|---|---|---|
| *APPETITE DISORDERS* | | | | | | |
| DECREASED APPETITE | 3 | 2 | 1 | 0 | 0 | 0 |
| ***ELECTROLYTE AND FLUID BALANCE CONDITIONS*** | | | | | | |
| *TOTAL FLUID VOLUME DECREASED* | | | | | | |
| DEHYDRATION | 1 | 1 | 0 | 0 | 0 | 0 |
| **MUSCULOSKELETAL AND CONNECTIVE TISSUE DISORDERS** | | | | | | |
| ***BONE DISORDERS (EXCL CONGENITAL AND FRACTURES)*** | | | | | | |
| *BONE RELATED SIGNS AND SYMPTOMS* | | | | | | |
| PAIN IN JAW | 3 | 1 | 2 | 0 | 0 | 0 |
| ***JOINT DISORDERS*** | | | | | | |
| *JOINT RELATED SIGNS AND SYMPTOMS* | | | | | | |
| ARTHRALGIA | 37 | 24 | 12 | 0 | 0 | 1 |
| ***MUSCLE DISORDERS*** | | | | | | |
| *MUSCLE PAINS* | | | | | | |
| MYALGIA | 78 | 50 | 21 | 5 | 0 | 2 |
| *MUSCLE RELATED SIGNS AND SYMPTOMS NEC* | | | | | | |
| MUSCLE FATIGUE | 2 | 1 | 0 | 1 | 0 | 0 |
| MUSCLE SPASMS | 1 | 1 | 0 | 0 | 0 | 0 |
| *MUSCLE TONE ABNORMALITIES* | | | | | | |
| TRISMUS | 1 | 0 | 1 | 0 | 0 | 0 |
| ***MUSCULOSKELETAL AND CONNECTIVE TISSUE DISORDERS NEC*** | | | | | | |
| *MUSCULOSKELETAL AND CONNECTIVE TISSUE CONDITIONS NEC* | | | | | | |
| MUSCULOSKELETAL STIFFNESS | 7 | 4 | 3 | 0 | 0 | 0 |
| BACK PAIN | 7 | 4 | 2 | 1 | 0 | 0 |
| LIMB DISCOMFORT | 44 | 34 | 9 | 1 | 0 | 0 |
| NECK PAIN | 3 | 2 | 1 | 0 | 0 | 0 |
| PAIN IN EXTREMITY | 383 | 302 | 54 | 16 | 1 | 10 |
| **NERVOUS SYSTEM DISORDERS** | | | | | | |
| ***CRANIAL NERVE DISORDERS (EXCL NEOPLASMS)*** | | | | | | |
| *FACIAL CRANIAL NERVE DISORDERS* | | | | | | |
| BELL'S PALSY | 1 | 0 | 1 | 0 | 0 | 0 |
| *OLFACTORY NERVE DISORDERS* | | | | | | |
| ANOSMIA | 1 | 1 | 0 | 0 | 0 | 0 |
| PAROSMIA | 2 | 1 | 1 | 0 | 0 | 0 |
| ***HEADACHES*** | | | | | | |
| *HEADACHES NEC* | | | | | | |
| CLUSTER HEADACHE | 1 | 0 | 1 | 0 | 0 | 0 |
| HEADACHE | 220 | 134 | 58 | 14 | 0 | 14 |
| SINUS HEADACHE | 8 | 5 | 3 | 0 | 0 | 0 |
| TENSION HEADACHE | 11 | 8 | 3 | 0 | 0 | 0 |
| *MIGRAINE HEADACHES* | | | | | | |
| MIGRAINE WITH AURA | 1 | 0 | 1 | 0 | 0 | 0 |
| TYPICAL AURA WITHOUT HEADACHE | 1 | 0 | 1 | 0 | 0 | 0 |
| ***MENTAL IMPAIRMENT DISORDERS*** | | | | | | |
| *MENTAL IMPAIRMENT (EXCL DEMENTIA AND MEMORY LOSS)* | | | | | | |
| DISTURBANCE IN ATTENTION | 1 | 0 | 1 | 0 | 0 | 0 |
| ***MOVEMENT DISORDERS (INCL PARKINSONISM)*** | | | | | | |
| *DYSKINESIAS AND MOVEMENT DISORDERS NEC* | | | | | | |
| CLUMSINESS | 1 | 1 | 0 | 0 | 0 | 0 |
| *TREMOR (EXCL CONGENITAL)* | | | | | | |
| TREMOR | 4 | 3 | 1 | 0 | 0 | 0 |
| ***NEUROLOGICAL DISORDERS NEC*** | | | | | | |
| *DISTURBANCES IN CONSCIOUSNESS NEC* | | | | | | |
| LETHARGY | 12 | 8 | 3 | 1 | 0 | 0 |
| SOMNOLENCE | 8 | 5 | 1 | 0 | 0 | 2 |
| SYNCOPE | 2 | 2 | 0 | 0 | 0 | 0 |

| | | | | | | |
|---|---|---|---|---|---|---|
| *NEUROLOGICAL SIGNS AND SYMPTOMS NEC* | | | | | | |
| AGITATION NEONATAL | 1 | 0 | 0 | 0 | 0 | 1 |
| BRAIN FOG | 1 | 1 | 0 | 0 | 0 | 0 |
| DIZZINESS | 37 | 22 | 11 | 3 | 0 | 1 |
| DIZZINESS EXERTIONAL | 1 | 1 | 0 | 0 | 0 | 0 |
| PRESYNCOPE | 1 | 1 | 0 | 0 | 0 | 0 |
| *PARAESTHESIAS AND DYSAESTHESIAS* | | | | | | |
| PARAESTHESIA | 1 | 1 | 0 | 0 | 0 | 0 |
| *SENSORY ABNORMALITIES NEC* | | | | | | |
| DYSGEUSIA | 1 | 1 | 0 | 0 | 0 | 0 |
| NEURALGIA | 1 | 1 | 0 | 0 | 0 | 0 |
| TASTE DISORDER | 1 | 1 | 0 | 0 | 0 | 0 |
| ***SEIZURES (INCL SUBTYPES)*** | | | | | | |
| *SEIZURES AND SEIZURE DISORDERS NEC* | | | | | | |
| SEIZURE | 1 | 1 | 0 | 0 | 0 | 0 |
| **PREGNANCY, PUERPERIUM AND PERINATAL CONDITIONS** | | | | | | |
| ***ABORTIONS AND STILLBIRTH*** | | | | | | |
| *ABORTIONS SPONTANEOUS* | | | | | | |
| ABORTION SPONTANEOUS | 19 | 11 | 8 | 0 | 0 | 0 |
| *STILLBIRTH AND FOETAL DEATH* | | | | | | |
| FOETAL DEATH | 1 | 1 | 0 | 0 | 0 | 0 |
| ***FOETAL COMPLICATIONS*** | | | | | | |
| *FOETAL COMPLICATIONS NEC* | | | | | | |
| FOETAL DISORDER | 1 | 1 | 0 | 0 | 0 | 0 |
| FOETAL HYPOKINESIA | 2 | 1 | 0 | 1 | 0 | 0 |
| *FOETAL GROWTH COMPLICATIONS* | | | | | | |
| FOETAL MACROSOMIA | 1 | 1 | 0 | 0 | 0 | 0 |
| ***MATERNAL COMPLICATIONS OF PREGNANCY*** | | | | | | |
| *MATERNAL COMPLICATIONS OF PREGNANCY NEC* | | | | | | |
| MORNING SICKNESS | 1 | 0 | 0 | 0 | 0 | 1 |
| ***PLACENTAL, AMNIOTIC AND CAVITY DISORDERS (EXCL HAEMORRHAGES)*** | | | | | | |
| *PLACENTAL ABNORMALITIES (EXCL NEOPLASMS)* | | | | | | |
| PLACENTAL INFARCTION | 1 | 1 | 0 | 0 | 0 | 0 |
| ***PREGNANCY, LABOUR, DELIVERY AND POSTPARTUM CONDITIONS*** | | | | | | |
| *NORMAL PREGNANCY, LABOUR AND DELIVERY* | | | | | | |
| PREGNANCY | 1 | 0 | 0 | 0 | 0 | 1 |
| UTERINE CONTRACTIONS DURING PREGNANCY | 1 | 1 | 0 | 0 | 0 | 0 |
| **PSYCHIATRIC DISORDERS** | | | | | | |
| ***ANXIETY DISORDERS AND SYMPTOMS*** | | | | | | |
| *ANXIETY SYMPTOMS* | | | | | | |
| AGITATION | 1 | 0 | 1 | 0 | 0 | 0 |
| ANXIETY | 3 | 1 | 0 | 0 | 0 | 2 |
| ***COGNITIVE AND ATTENTION DISORDERS AND DISTURBANCES*** | | | | | | |
| *COGNITIVE AND ATTENTION DISORDERS AND DISTURBANCES NEC* | | | | | | |
| MENTAL FATIGUE | 1 | 1 | 0 | 0 | 0 | 0 |
| ***DELIRIA (INCL CONFUSION)*** | | | | | | |
| *CONFUSION AND DISORIENTATION* | | | | | | |
| CONFUSIONAL STATE | 1 | 1 | 0 | 0 | 0 | 0 |
| ***DEPRESSED MOOD DISORDERS AND DISTURBANCES*** | | | | | | |
| *MOOD ALTERATIONS WITH DEPRESSIVE SYMPTOMS* | | | | | | |
| DEPRESSED MOOD | 1 | 1 | 0 | 0 | 0 | 0 |
| ***DISTURBANCES IN THINKING AND PERCEPTION*** | | | | | | |
| *HALLUCINATIONS (EXCL SLEEP-RELATED)* | | | | | | |
| HALLUCINATION | 1 | 1 | 0 | 0 | 0 | 0 |
| ***MOOD DISORDERS AND DISTURBANCES NEC*** | | | | | | |
| *EMOTIONAL AND MOOD DISTURBANCES NEC* | | | | | | |

| | | | | | | |
|---|---|---|---|---|---|---|
| EMOTIONAL DISORDER | 1 | 0 | 1 | 0 | 0 | 0 |
| ***SLEEP DISORDERS AND DISTURBANCES*** | | | | | | |
| *DISTURBANCES IN INITIATING AND MAINTAINING SLEEP* | | | | | | |
| INITIAL INSOMNIA | 1 | 1 | 0 | 0 | 0 | 0 |
| INSOMNIA | 7 | 5 | 2 | 0 | 0 | 0 |
| **REPRODUCTIVE SYSTEM AND BREAST DISORDERS** | | | | | | |
| ***MENSTRUAL CYCLE AND UTERINE BLEEDING DISORDERS*** | | | | | | |
| *MENSTRUATION AND UTERINE BLEEDING NEC* | | | | | | |
| MENSTRUAL DISORDER | 1 | 0 | 0 | 1 | 0 | 0 |
| *MENSTRUATION WITH DECREASED BLEEDING* | | | | | | |
| MENSTRUATION DELAYED | 2 | 0 | 1 | 0 | 0 | 1 |
| OLIGOMENORRHOEA | 1 | 0 | 0 | 0 | 0 | 1 |
| *MENSTRUATION WITH INCREASED BLEEDING* | | | | | | |
| HEAVY MENSTRUAL BLEEDING | 2 | 0 | 0 | 1 | 0 | 1 |
| ***VULVOVAGINAL DISORDERS (EXCL INFECTIONS AND INFLAMMATIONS)*** | | | | | | |
| *VULVOVAGINAL DISORDERS NEC* | | | | | | |
| VAGINAL HAEMORRHAGE | 6 | 4 | 2 | 0 | 0 | 0 |
| **RESPIRATORY, THORACIC AND MEDIASTINAL DISORDERS** | | | | | | |
| ***RESPIRATORY DISORDERS NEC*** | | | | | | |
| *BREATHING ABNORMALITIES* | | | | | | |
| DYSPNOEA | 10 | 5 | 4 | 0 | 0 | 1 |
| *COUGHING AND ASSOCIATED SYMPTOMS* | | | | | | |
| COUGH | 5 | 4 | 1 | 0 | 0 | 0 |
| ***RESPIRATORY TRACT SIGNS AND SYMPTOMS*** | | | | | | |
| *UPPER RESPIRATORY TRACT SIGNS AND SYMPTOMS* | | | | | | |
| DRY THROAT | 1 | 0 | 1 | 0 | 0 | 0 |
| OROPHARYNGEAL PAIN | 18 | 14 | 3 | 0 | 0 | 1 |
| PARANASAL SINUS DISCOMFORT | 1 | 1 | 0 | 0 | 0 | 0 |
| RHINORRHOEA | 7 | 4 | 2 | 1 | 0 | 0 |
| SINUS PAIN | 2 | 2 | 0 | 0 | 0 | 0 |
| SNEEZING | 1 | 1 | 0 | 0 | 0 | 0 |
| ***UPPER RESPIRATORY TRACT DISORDERS (EXCL INFECTIONS)*** | | | | | | |
| *NASAL CONGESTION AND INFLAMMATIONS* | | | | | | |
| NASAL CONGESTION | 2 | 0 | 2 | 0 | 0 | 0 |
| **SKIN AND SUBCUTANEOUS TISSUE DISORDERS** | | | | | | |
| ***ANGIOEDEMA AND URTICARIA*** | | | | | | |
| *URTICARIAS* | | | | | | |
| URTICARIA | 1 | 1 | 0 | 0 | 0 | 0 |
| ***CORNIFICATION AND DYSTROPHIC SKIN DISORDERS*** | | | | | | |
| *SKIN DYSTROPHIES* | | | | | | |
| HYPERTROPHIC SCAR | 1 | 1 | 0 | 0 | 0 | 0 |
| ***EPIDERMAL AND DERMAL CONDITIONS*** | | | | | | |
| *DERMAL AND EPIDERMAL CONDITIONS NEC* | | | | | | |
| SKIN WARM | 1 | 1 | 0 | 0 | 0 | 0 |
| *DERMATITIS AND ECZEMA* | | | | | | |
| DERMATITIS ALLERGIC | 1 | 1 | 0 | 0 | 0 | 0 |
| *ERYTHEMAS* | | | | | | |
| ERYTHEMA | 3 | 1 | 1 | 0 | 0 | 1 |
| *PRURITUS NEC* | | | | | | |
| PRURITUS | 5 | 3 | 0 | 1 | 0 | 1 |
| *RASHES, ERUPTIONS AND EXANTHEMS NEC* | | | | | | |
| RASH | 9 | 6 | 1 | 2 | 0 | 0 |
| RASH PRURITIC | 1 | 1 | 0 | 0 | 0 | 0 |
| ***SKIN APPENDAGE CONDITIONS*** | | | | | | |
| *APOCRINE AND ECCRINE GLAND DISORDERS* | | | | | | |
| COLD SWEAT | 2 | 2 | 0 | 0 | 0 | 0 |

| | | | | | | |
|---|---|---|---|---|---|---|
| HYPERHIDROSIS | 6 | 6 | 0 | 0 | 0 | 0 |
| NIGHT SWEATS | 2 | 0 | 2 | 0 | 0 | 0 |
| **VASCULAR DISORDERS** | | | | | | |
| ***DECREASED AND NONSPECIFIC BLOOD PRESSURE DISORDERS AND SHOCK*** | | | | | | |
| *BLOOD PRESSURE DISORDERS NEC* | | | | | | |
| BLOOD PRESSURE FLUCTUATION | 1 | 1 | 0 | 0 | 0 | 0 |
| *VASCULAR HYPOTENSIVE DISORDERS* | | | | | | |
| HYPOTENSION | 2 | 2 | 0 | 0 | 0 | 0 |
| ***EMBOLISM AND THROMBOSIS*** | | | | | | |
| *NON-SITE SPECIFIC EMBOLISM AND THROMBOSIS* | | | | | | |
| EMBOLISM | 1 | 1 | 0 | 0 | 0 | 0 |
| ***VASCULAR DISORDERS NEC*** | | | | | | |
| *NON-SITE SPECIFIC VASCULAR DISORDERS NEC* | | | | | | |
| VASCULAR PAIN | 1 | 0 | 1 | 0 | 0 | 0 |
| *PERIPHERAL VASCULAR DISORDERS NEC* | | | | | | |
| HOT FLUSH | 2 | 2 | 0 | 0 | 0 | 0 |
| ***VASCULAR HAEMORRHAGIC DISORDERS*** | | | | | | |
| *HAEMORRHAGES NEC* | | | | | | |
| HAEMORRHAGE | 1 | 1 | 0 | 0 | 0 | 0 |
| **TOTAL ADR EVENTS** | ***1934*** | ***1255*** | ***476*** | ***118*** | ***3*** | ***82*** |

**TABLE S2-2. Pfizer BioNTech COVID-19 vaccine: ADR listing for events reported by pregnant patients, in those reporting any vaccination dose**

| | Patients with any vaccination dose: ADR Counts | | | | |
|---|---|---|---|---|---|
| **REACTION TERM (SOC, *HLGT*, *HLT*, PT)** | All doses | 1st dose | 2nd dose | 3rd dose | Other doses |
| ***(freetext)*** | | | | | |
| **BLOOD AND LYMPHATIC SYSTEM DISORDERS** | | | | | |
| ***SPLEEN, LYMPHATIC AND RETICULOENDOTHELIAL SYSTEM DISORDERS*** | | | | | |
| *LYMPHATIC SYSTEM DISORDERS NEC* | | | | | |
| LYMPH NODE PAIN | 3 | 0 | 2 | 1 | 0 |
| LYMPHADENITIS | 1 | 0 | 1 | 0 | 0 |
| LYMPHADENOPATHY | 9 | 6 | 3 | 0 | 0 |
| **CARDIAC DISORDERS** | | | | | |
| ***CARDIAC ARRHYTHMIAS*** | | | | | |
| *RATE AND RHYTHM DISORDERS NEC* | | | | | |
| TACHYCARDIA | 0 | 0 | 0 | 0 | 0 |
| ***CARDIAC DISORDERS, SIGNS AND SYMPTOMS NEC*** | | | | | |
| *CARDIAC SIGNS AND SYMPTOMS NEC* | | | | | |
| PALPITATIONS | 3 | 1 | 2 | 0 | 0 |
| **EAR AND LABYRINTH DISORDERS** | | | | | |
| ***AURAL DISORDERS NEC*** | | | | | |
| *EAR DISORDERS NEC* | | | | | |
| EAR PAIN | 1 | 1 | 0 | 0 | 0 |
| ***INNER EAR AND VIIITH CRANIAL NERVE DISORDERS*** | | | | | |
| *INNER EAR SIGNS AND SYMPTOMS* | | | | | |
| TINNITUS | 1 | 0 | 1 | 0 | 0 |
| VERTIGO | 0 | 0 | 0 | 0 | 0 |
| **EYE DISORDERS** | | | | | |
| ***EYE DISORDERS NEC*** | | | | | |
| *OCULAR DISORDERS NEC* | | | | | |
| EYE PAIN | 1 | 1 | 0 | 0 | 0 |
| ***OCULAR SENSORY SYMPTOMS NEC*** | | | | | |
| *OCULAR SENSATION DISORDERS* | | | | | |
| ASTHENOPIA | 2 | 1 | 1 | 0 | 0 |
| PHOTOPHOBIA | 1 | 1 | 0 | 0 | 0 |
| ***VISION DISORDERS*** | | | | | |
| *VISUAL DISORDERS NEC* | | | | | |
| VISION BLURRED | 2 | 1 | 1 | 0 | 0 |
| **GASTROINTESTINAL DISORDERS** | | | | | |
| ***DENTAL AND GINGIVAL CONDITIONS*** | | | | | |
| *DENTAL DISORDERS NEC* | | | | | |
| TEETHING | 1 | 1 | 0 | 0 | 0 |
| ***GASTROINTESTINAL INFLAMMATORY CONDITIONS*** | | | | | |
| *GASTROINTESTINAL INFLAMMATORY DISORDERS NEC* | | | | | |
| GASTROINTESTINAL TRACT IRRITATION | 1 | 0 | 0 | 1 | 0 |
| ***GASTROINTESTINAL MOTILITY AND DEFAECATION CONDITIONS*** | | | | | |
| *DIARRHOEA (EXCL INFECTIVE)* | | | | | |
| DIARRHOEA | 13 | 7 | 3 | 3 | 0 |
| *GASTROINTESTINAL ATONIC AND HYPOMOTILITY DISORDERS NEC* | | | | | |
| CONSTIPATION | 1 | 0 | 0 | 1 | 0 |

| | | | | | |
|---|---|---|---|---|---|
| ***GASTROINTESTINAL SIGNS AND SYMPTOMS*** | | | | | |
| *GASTROINTESTINAL AND ABDOMINAL PAINS (EXCL ORAL AND THROAT)* | | | | | |
| ABDOMINAL PAIN | 0 | 0 | 0 | 0 | 0 |
| ABDOMINAL PAIN UPPER | 4 | 3 | 1 | 0 | 0 |
| *GASTROINTESTINAL SIGNS AND SYMPTOMS NEC* | | | | | |
| ABDOMINAL DISCOMFORT | 0 | 0 | 0 | 0 | 0 |
| *NAUSEA AND VOMITING SYMPTOMS* | | | | | |
| NAUSEA | 45 | 26 | 18 | 1 | 0 |
| VOMITING | 12 | 9 | 3 | 0 | 0 |
| ***ORAL SOFT TISSUE CONDITIONS*** | | | | | |
| *ORAL SOFT TISSUE SIGNS AND SYMPTOMS* | | | | | |
| HYPOAESTHESIA ORAL | 1 | 1 | 0 | 0 | 0 |
| PARAESTHESIA ORAL | 1 | 1 | 0 | 0 | 0 |
| ***TONGUE CONDITIONS*** | | | | | |
| *TONGUE SIGNS AND SYMPTOMS* | | | | | |
| GLOSSODYNIA | 1 | 1 | 0 | 0 | 0 |
| **GENERAL DISORDERS AND ADMINISTRATION SITE CONDITIONS** | | | | | |
| ***ADMINISTRATION SITE REACTIONS*** | | | | | |
| *ADMINISTRATION SITE REACTIONS NEC* | | | | | |
| PUNCTURE SITE BRUISE | 0 | 0 | 0 | 0 | 0 |
| PUNCTURE SITE PAIN | 1 | 1 | 0 | 0 | 0 |
| *INJECTION SITE REACTIONS* | | | | | |
| INJECTION SITE BRUISING | 1 | 0 | 1 | 0 | 0 |
| INJECTION SITE ERYTHEMA | 1 | 1 | 0 | 0 | 0 |
| INJECTION SITE MASS | 1 | 0 | 1 | 0 | 0 |
| INJECTION SITE PAIN | 39 | 31 | 7 | 1 | 0 |
| INJECTION SITE REACTION | 1 | 0 | 1 | 0 | 0 |
| INJECTION SITE URTICARIA | 1 | 0 | 1 | 0 | 0 |
| INJECTION SITE WARMTH | 1 | 1 | 0 | 0 | 0 |
| *VACCINATION SITE REACTIONS* | | | | | |
| VACCINATION SITE PAIN | 1 | 1 | 0 | 0 | 0 |
| ***BODY TEMPERATURE CONDITIONS*** | | | | | |
| *FEBRILE DISORDERS* | | | | | |
| PYREXIA | 40 | 22 | 17 | 1 | 0 |
| ***GENERAL SYSTEM DISORDERS NEC*** | | | | | |
| *ASTHENIC CONDITIONS* | | | | | |
| ASTHENIA | 2 | 2 | 0 | 0 | 0 |
| FATIGUE | 166 | 112 | 48 | 6 | 0 |
| MALAISE | 10 | 4 | 6 | 0 | 0 |
| *FEELINGS AND SENSATIONS NEC* | | | | | |
| CHILLS | 12 | 4 | 7 | 1 | 0 |
| FEELING ABNORMAL | 4 | 2 | 1 | 1 | 0 |
| FEELING COLD | 1 | 0 | 1 | 0 | 0 |
| FEELING HOT | 0 | 0 | 0 | 0 | 0 |
| FEELING OF BODY TEMPERATURE CHANGE | 0 | 0 | 0 | 0 | 0 |
| HANGOVER | 1 | 0 | 0 | 1 | 0 |
| HUNGER | 0 | 0 | 0 | 0 | 0 |
| THIRST | 1 | 1 | 0 | 0 | 0 |
| *GENERAL SIGNS AND SYMPTOMS NEC* | | | | | |
| CRYING | 0 | 0 | 0 | 0 | 0 |
| ILLNESS | 7 | 4 | 3 | 0 | 0 |
| INFLUENZA LIKE ILLNESS | 7 | 3 | 4 | 0 | 0 |

| | | | | | |
|---|---|---|---|---|---|
| LOCAL REACTION | 1 | 0 | 1 | 0 | 0 |
| PERIPHERAL SWELLING | 3 | 2 | 0 | 1 | 0 |
| SWELLING | 2 | 1 | 1 | 0 | 0 |
| *PAIN AND DISCOMFORT NEC* | | | | | |
| AXILLARY PAIN | 9 | 2 | 5 | 2 | 0 |
| CHEST PAIN | 1 | 1 | 0 | 0 | 0 |
| PAIN | 39 | 24 | 12 | 3 | 0 |
| TENDERNESS | 8 | 6 | 2 | 0 | 0 |
| **INFECTIONS AND INFESTATIONS** | | | | | |
| ***BACTERIAL INFECTIOUS DISORDERS*** | | | | | |
| *BACTERIAL INFECTIONS NEC* | | | | | |
| PERIORBITAL CELLULITIS | 1 | 0 | 1 | 0 | 0 |
| ***INFECTIONS - PATHOGEN UNSPECIFIED*** | | | | | |
| *BREAST INFECTIONS* | | | | | |
| MASTITIS | 1 | 0 | 0 | 1 | 0 |
| *UPPER RESPIRATORY TRACT INFECTIONS* | | | | | |
| LARYNGITIS | 1 | 0 | 1 | 0 | 0 |
| NASOPHARYNGITIS | 7 | 4 | 3 | 0 | 0 |
| ***VIRAL INFECTIOUS DISORDERS*** | | | | | |
| *HERPES VIRAL INFECTIONS* | | | | | |
| GENITAL HERPES | 1 | 1 | 0 | 0 | 0 |
| HERPES ZOSTER | 1 | 1 | 0 | 0 | 0 |
| ORAL HERPES | 1 | 0 | 1 | 0 | 0 |
| *INFLUENZA VIRAL INFECTIONS* | | | | | |
| INFLUENZA | 9 | 4 | 4 | 1 | 0 |
| *VIRAL INFECTIONS NEC* | | | | | |
| VIRAL DIARRHOEA | 1 | 1 | 0 | 0 | 0 |
| **INJURY, POISONING AND PROCEDURAL COMPLICATIONS** | | | | | |
| ***INJURIES NEC*** | | | | | |
| *SKIN INJURIES NEC* | | | | | |
| CONTUSION | 6 | 6 | 0 | 0 | 0 |
| ***PROCEDURAL RELATED INJURIES AND COMPLICATIONS NEC*** | | | | | |
| *NON-SITE SPECIFIC PROCEDURAL COMPLICATIONS* | | | | | |
| INJECTION RELATED REACTION | 0 | 0 | 0 | 0 | 0 |
| **INVESTIGATIONS** | | | | | |
| ***CARDIAC AND VASCULAR INVESTIGATIONS (EXCL ENZYME TESTS)*** | | | | | |
| *HEART RATE AND PULSE INVESTIGATIONS* | | | | | |
| HEART RATE | 1 | 1 | 0 | 0 | 0 |
| HEART RATE INCREASED | 0 | 0 | 0 | 0 | 0 |
| ***PHYSICAL EXAMINATION AND ORGAN SYSTEM STATUS TOPICS*** | | | | | |
| *PHYSICAL EXAMINATION PROCEDURES AND ORGAN SYSTEM STATUS* | | | | | |
| BODY TEMPERATURE INCREASED | 0 | 0 | 0 | 0 | 0 |
| ***WATER, ELECTROLYTE AND MINERAL INVESTIGATIONS*** | | | | | |
| *WATER AND ELECTROLYTE ANALYSES NEC* | | | | | |
| VOLUME BLOOD | 1 | 0 | 1 | 0 | 0 |
| **METABOLISM AND NUTRITION DISORDERS** | | | | | |
| ***APPETITE AND GENERAL NUTRITIONAL DISORDERS*** | | | | | |
| *APPETITE DISORDERS* | | | | | |
| DECREASED APPETITE | 0 | 0 | 0 | 0 | 0 |
| ***ELECTROLYTE AND FLUID BALANCE CONDITIONS*** | | | | | |
| *TOTAL FLUID VOLUME DECREASED* | | | | | |
| DEHYDRATION | 0 | 0 | 0 | 0 | 0 |
| **MUSCULOSKELETAL AND CONNECTIVE TISSUE DISORDERS** | | | | | |

| | | | | | |
|---|---|---|---|---|---|
| ***BONE DISORDERS (EXCL CONGENITAL AND FRACTURES)*** | | | | | |
| *BONE RELATED SIGNS AND SYMPTOMS* | | | | | |
| PAIN IN JAW | 1 | 1 | 0 | 0 | 0 |
| ***JOINT DISORDERS*** | | | | | |
| *JOINT RELATED SIGNS AND SYMPTOMS* | | | | | |
| ARTHRALGIA | 26 | 16 | 10 | 0 | 0 |
| ***MUSCLE DISORDERS*** | | | | | |
| *MUSCLE PAINS* | | | | | |
| MYALGIA | 37 | 25 | 11 | 1 | 0 |
| *MUSCLE RELATED SIGNS AND SYMPTOMS NEC* | | | | | |
| MUSCLE FATIGUE | 0 | 0 | 0 | 0 | 0 |
| MUSCLE SPASMS | 1 | 1 | 0 | 0 | 0 |
| *MUSCLE TONE ABNORMALITIES* | | | | | |
| TRISMUS | 0 | 0 | 0 | 0 | 0 |
| ***MUSCULOSKELETAL AND CONNECTIVE TISSUE DISORDERS NEC*** | | | | | |
| *MUSCULOSKELETAL AND CONNECTIVE TISSUE CONDITIONS NEC* | | | | | |
| MUSCULOSKELETAL STIFFNESS | 4 | 3 | 1 | 0 | 0 |
| BACK PAIN | 2 | 1 | 0 | 1 | 0 |
| LIMB DISCOMFORT | 37 | 28 | 8 | 1 | 0 |
| NECK PAIN | 2 | 2 | 0 | 0 | 0 |
| PAIN IN EXTREMITY | 293 | 235 | 45 | 12 | 1 |
| **NERVOUS SYSTEM DISORDERS** | | | | | |
| ***CRANIAL NERVE DISORDERS (EXCL NEOPLASMS)*** | | | | | |
| *FACIAL CRANIAL NERVE DISORDERS* | | | | | |
| BELL'S PALSY | 1 | 0 | 1 | 0 | 0 |
| *OLFACTORY NERVE DISORDERS* | | | | | |
| ANOSMIA | 1 | 1 | 0 | 0 | 0 |
| PAROSMIA | 1 | 1 | 0 | 0 | 0 |
| ***HEADACHES*** | | | | | |
| *HEADACHES NEC* | | | | | |
| CLUSTER HEADACHE | 1 | 0 | 1 | 0 | 0 |
| HEADACHE | 120 | 79 | 33 | 8 | 0 |
| SINUS HEADACHE | 5 | 3 | 2 | 0 | 0 |
| TENSION HEADACHE | 6 | 4 | 2 | 0 | 0 |
| *MIGRAINE HEADACHES* | | | | | |
| MIGRAINE WITH AURA | 0 | 0 | 0 | 0 | 0 |
| TYPICAL AURA WITHOUT HEADACHE | 1 | 0 | 1 | 0 | 0 |
| ***MENTAL IMPAIRMENT DISORDERS*** | | | | | |
| *MENTAL IMPAIRMENT (EXCL DEMENTIA AND MEMORY LOSS)* | | | | | |
| DISTURBANCE IN ATTENTION | 1 | 0 | 1 | 0 | 0 |
| ***MOVEMENT DISORDERS (INCL PARKINSONISM)*** | | | | | |
| *DYSKINESIAS AND MOVEMENT DISORDERS NEC* | | | | | |
| CLUMSINESS | 0 | 0 | 0 | 0 | 0 |
| *TREMOR (EXCL CONGENITAL)* | | | | | |
| TREMOR | 0 | 0 | 0 | 0 | 0 |
| ***NEUROLOGICAL DISORDERS NEC*** | | | | | |
| *DISTURBANCES IN CONSCIOUSNESS NEC* | | | | | |
| LETHARGY | 10 | 6 | 3 | 1 | 0 |
| SOMNOLENCE | 4 | 4 | 0 | 0 | 0 |
| SYNCOPE | 2 | 2 | 0 | 0 | 0 |
| *NEUROLOGICAL SIGNS AND SYMPTOMS NEC* | | | | | |
| AGITATION NEONATAL | 0 | 0 | 0 | 0 | 0 |
| BRAIN FOG | 0 | 0 | 0 | 0 | 0 |

| | | | | | |
|---|---|---|---|---|---|
| DIZZINESS | 19 | 10 | 8 | 1 | 0 |
| DIZZINESS EXERTIONAL | 0 | 0 | 0 | 0 | 0 |
| PRESYNCOPE | 0 | 0 | 0 | 0 | 0 |
| *PARAESTHESIAS AND DYSAESTHESIAS* | | | | | |
| PARAESTHESIA | 0 | 0 | 0 | 0 | 0 |
| *SENSORY ABNORMALITIES NEC* | | | | | |
| DYSGEUSIA | 0 | 0 | 0 | 0 | 0 |
| NEURALGIA | 1 | 1 | 0 | 0 | 0 |
| TASTE DISORDER | 1 | 1 | 0 | 0 | 0 |
| ***SEIZURES (INCL SUBTYPES)*** | | | | | |
| *SEIZURES AND SEIZURE DISORDERS NEC* | | | | | |
| SEIZURE | 1 | 1 | 0 | 0 | 0 |
| **PREGNANCY, PUERPERIUM AND PERINATAL CONDITIONS** | | | | | |
| ***ABORTIONS AND STILLBIRTH*** | | | | | |
| *ABORTIONS SPONTANEOUS* | | | | | |
| ABORTION SPONTANEOUS | 13 | 7 | 6 | 0 | 0 |
| *STILLBIRTH AND FOETAL DEATH* | | | | | |
| FOETAL DEATH | 0 | 0 | 0 | 0 | 0 |
| ***FOETAL COMPLICATIONS*** | | | | | |
| *FOETAL COMPLICATIONS NEC* | | | | | |
| FOETAL DISORDER | 0 | 0 | 0 | 0 | 0 |
| FOETAL HYPOKINESIA | 2 | 1 | 0 | 1 | 0 |
| *FOETAL GROWTH COMPLICATIONS* | | | | | |
| FOETAL MACROSOMIA | 1 | 1 | 0 | 0 | 0 |
| ***MATERNAL COMPLICATIONS OF PREGNANCY*** | | | | | |
| *MATERNAL COMPLICATIONS OF PREGNANCY NEC* | | | | | |
| MORNING SICKNESS | 0 | 0 | 0 | 0 | 0 |
| ***PLACENTAL, AMNIOTIC AND CAVITY DISORDERS (EXCL HAEMORRHAGES)*** | | | | | |
| *PLACENTAL ABNORMALITIES (EXCL NEOPLASMS)* | | | | | |
| PLACENTAL INFARCTION | 0 | 0 | 0 | 0 | 0 |
| ***PREGNANCY, LABOUR, DELIVERY AND POSTPARTUM CONDITIONS*** | | | | | |
| *NORMAL PREGNANCY, LABOUR AND DELIVERY* | | | | | |
| PREGNANCY | 0 | 0 | 0 | 0 | 0 |
| UTERINE CONTRACTIONS DURING PREGNANCY | 1 | 1 | 0 | 0 | 0 |
| **PSYCHIATRIC DISORDERS** | | | | | |
| ***ANXIETY DISORDERS AND SYMPTOMS*** | | | | | |
| *ANXIETY SYMPTOMS* | | | | | |
| AGITATION | 1 | 0 | 1 | 0 | 0 |
| ANXIETY | 1 | 1 | 0 | 0 | 0 |
| ***COGNITIVE AND ATTENTION DISORDERS AND DISTURBANCES*** | | | | | |
| *COGNITIVE AND ATTENTION DISORDERS AND DISTURBANCES NEC* | | | | | |
| MENTAL FATIGUE | 1 | 1 | 0 | 0 | 0 |
| ***DELIRIA (INCL CONFUSION)*** | | | | | |
| *CONFUSION AND DISORIENTATION* | | | | | |
| CONFUSIONAL STATE | 1 | 1 | 0 | 0 | 0 |
| ***DEPRESSED MOOD DISORDERS AND DISTURBANCES*** | | | | | |
| *MOOD ALTERATIONS WITH DEPRESSIVE SYMPTOMS* | | | | | |
| DEPRESSED MOOD | 1 | 1 | 0 | 0 | 0 |
| ***DISTURBANCES IN THINKING AND PERCEPTION*** | | | | | |
| *HALLUCINATIONS (EXCL SLEEP-RELATED)* | | | | | |
| HALLUCINATION | 1 | 1 | 0 | 0 | 0 |
| ***MOOD DISORDERS AND DISTURBANCES NEC*** | | | | | |
| *EMOTIONAL AND MOOD DISTURBANCES NEC* | | | | | |

| | | | | | |
|---|---|---|---|---|---|
| EMOTIONAL DISORDER | 1 | 0 | 1 | 0 | 0 |
| ***SLEEP DISORDERS AND DISTURBANCES*** | | | | | |
| *DISTURBANCES IN INITIATING AND MAINTAINING SLEEP* | | | | | |
| INITIAL INSOMNIA | 1 | 1 | 0 | 0 | 0 |
| INSOMNIA | 3 | 2 | 1 | 0 | 0 |
| **REPRODUCTIVE SYSTEM AND BREAST DISORDERS** | | | | | |
| ***MENSTRUAL CYCLE AND UTERINE BLEEDING DISORDERS*** | | | | | |
| *MENSTRUATION AND UTERINE BLEEDING NEC* | | | | | |
| MENSTRUAL DISORDER | 1 | 0 | 0 | 1 | 0 |
| *MENSTRUATION WITH DECREASED BLEEDING* | | | | | |
| MENSTRUATION DELAYED | 0 | 0 | 0 | 0 | 0 |
| OLIGOMENORRHOEA | 0 | 0 | 0 | 0 | 0 |
| *MENSTRUATION WITH INCREASED BLEEDING* | | | | | |
| HEAVY MENSTRUAL BLEEDING | 1 | 0 | 0 | 1 | 0 |
| ***VULVOVAGINAL DISORDERS (EXCL INFECTIONS AND INFLAMMATIONS)*** | | | | | |
| *VULVOVAGINAL DISORDERS NEC* | | | | | |
| VAGINAL HAEMORRHAGE | 5 | 3 | 2 | 0 | 0 |
| **RESPIRATORY, THORACIC AND MEDIASTINAL DISORDERS** | | | | | |
| ***RESPIRATORY DISORDERS NEC*** | | | | | |
| *BREATHING ABNORMALITIES* | | | | | |
| DYSPNOEA | 5 | 4 | 1 | 0 | 0 |
| *COUGHING AND ASSOCIATED SYMPTOMS* | | | | | |
| COUGH | 4 | 3 | 1 | 0 | 0 |
| ***RESPIRATORY TRACT SIGNS AND SYMPTOMS*** | | | | | |
| *UPPER RESPIRATORY TRACT SIGNS AND SYMPTOMS* | | | | | |
| DRY THROAT | 1 | 0 | 1 | 0 | 0 |
| OROPHARYNGEAL PAIN | 12 | 9 | 3 | 0 | 0 |
| PARANASAL SINUS DISCOMFORT | 1 | 1 | 0 | 0 | 0 |
| RHINORRHOEA | 5 | 3 | 2 | 0 | 0 |
| SINUS PAIN | 1 | 1 | 0 | 0 | 0 |
| SNEEZING | 1 | 1 | 0 | 0 | 0 |
| ***UPPER RESPIRATORY TRACT DISORDERS (EXCL INFECTIONS)*** | | | | | |
| *NASAL CONGESTION AND INFLAMMATIONS* | | | | | |
| NASAL CONGESTION | 1 | 0 | 1 | 0 | 0 |
| **SKIN AND SUBCUTANEOUS TISSUE DISORDERS** | | | | | |
| ***ANGIOEDEMA AND URTICARIA*** | | | | | |
| *URTICARIAS* | | | | | |
| URTICARIA | 1 | 1 | 0 | 0 | 0 |
| ***CORNIFICATION AND DYSTROPHIC SKIN DISORDERS*** | | | | | |
| *SKIN DYSTROPHIES* | | | | | |
| HYPERTROPHIC SCAR | 1 | 1 | 0 | 0 | 0 |
| ***EPIDERMAL AND DERMAL CONDITIONS*** | | | | | |
| *DERMAL AND EPIDERMAL CONDITIONS NEC* | | | | | |
| SKIN WARM | 1 | 1 | 0 | 0 | 0 |
| *DERMATITIS AND ECZEMA* | | | | | |
| DERMATITIS ALLERGIC | 0 | 0 | 0 | 0 | 0 |
| *ERYTHEMAS* | | | | | |
| ERYTHEMA | 2 | 1 | 1 | 0 | 0 |
| *PRURITUS NEC* | | | | | |
| PRURITUS | 1 | 1 | 0 | 0 | 0 |
| *RASHES, ERUPTIONS AND EXANTHEMS NEC* | | | | | |
| RASH | 5 | 4 | 1 | 0 | 0 |
| RASH PRURITIC | 0 | 0 | 0 | 0 | 0 |

| | | | | | |
|---|---|---|---|---|---|
| ***SKIN APPENDAGE CONDITIONS*** | | | | | |
| *APOCRINE AND ECCRINE GLAND DISORDERS* | | | | | |
| COLD SWEAT | 0 | 0 | 0 | 0 | 0 |
| HYPERHIDROSIS | 3 | 3 | 0 | 0 | 0 |
| NIGHT SWEATS | 1 | 0 | 1 | 0 | 0 |
| **VASCULAR DISORDERS** | | | | | |
| ***DECREASED AND NONSPECIFIC BLOOD PRESSURE DISORDERS AND SHOCK*** | | | | | |
| *BLOOD PRESSURE DISORDERS NEC* | | | | | |
| BLOOD PRESSURE FLUCTUATION | 0 | 0 | 0 | 0 | 0 |
| *VASCULAR HYPOTENSIVE DISORDERS* | | | | | |
| HYPOTENSION | 2 | 2 | 0 | 0 | 0 |
| ***EMBOLISM AND THROMBOSIS*** | | | | | |
| *NON-SITE SPECIFIC EMBOLISM AND THROMBOSIS* | | | | | |
| EMBOLISM | 0 | 0 | 0 | 0 | 0 |
| ***VASCULAR DISORDERS NEC*** | | | | | |
| *NON-SITE SPECIFIC VASCULAR DISORDERS NEC* | | | | | |
| VASCULAR PAIN | 1 | 0 | 1 | 0 | 0 |
| *PERIPHERAL VASCULAR DISORDERS NEC* | | | | | |
| HOT FLUSH | 2 | 2 | 0 | 0 | 0 |
| ***VASCULAR HAEMORRHAGIC DISORDERS*** | | | | | |
| *HAEMORRHAGES NEC* | | | | | |
| HAEMORRHAGE | 1 | 1 | 0 | 0 | 0 |
| **TOTAL ADR EVENTS** | ***1161*** | ***789*** | ***317*** | ***54*** | ***1*** |

**TABLE S2-3. AstraZeneca COVID-19 vaccine: ADR listing for events reported by pregnant patients, in those reporting any vaccination dose**

| | Patients with any vaccination dose: ADR Counts | | | | |
|---|---|---|---|---|---|
| **REACTION TERM (SOC, *HLGT*, *HLT*, PT)** | All doses | 1st dose | 2nd dose | 3rd dose | Other doses |
| ***(freetext)*** | 0 | 0 | 0 | 0 | 0 |
| **BLOOD AND LYMPHATIC SYSTEM DISORDERS** | | | | | |
| ***SPLEEN, LYMPHATIC AND RETICULOENDOTHELIAL SYSTEM DISORDERS*** | | | | | |
| *LYMPHATIC SYSTEM DISORDERS NEC* | | | | | |
| LYMPH NODE PAIN | 0 | 0 | 0 | 0 | 0 |
| LYMPHADENITIS | 0 | 0 | 0 | 0 | 0 |
| LYMPHADENOPATHY | 0 | 0 | 0 | 0 | 0 |
| **CARDIAC DISORDERS** | | | | | |
| ***CARDIAC ARRHYTHMIAS*** | | | | | |
| *RATE AND RHYTHM DISORDERS NEC* | | | | | |
| TACHYCARDIA | 1 | 0 | 1 | 0 | 0 |
| ***CARDIAC DISORDERS, SIGNS AND SYMPTOMS NEC*** | | | | | |
| *CARDIAC SIGNS AND SYMPTOMS NEC* | | | | | |
| PALPITATIONS | 0 | 0 | 0 | 0 | 0 |
| **EAR AND LABYRINTH DISORDERS** | | | | | |
| ***AURAL DISORDERS NEC*** | | | | | |
| *EAR DISORDERS NEC* | | | | | |
| EAR PAIN | 0 | 0 | 0 | 0 | 0 |
| ***INNER EAR AND VIIITH CRANIAL NERVE DISORDERS*** | | | | | |
| *INNER EAR SIGNS AND SYMPTOMS* | | | | | |
| TINNITUS | 0 | 0 | 0 | 0 | 0 |
| VERTIGO | 1 | 1 | 0 | 0 | 0 |
| **EYE DISORDERS** | | | | | |
| ***EYE DISORDERS NEC*** | | | | | |
| *OCULAR DISORDERS NEC* | | | | | |
| EYE PAIN | 1 | 0 | 1 | 0 | 0 |
| ***OCULAR SENSORY SYMPTOMS NEC*** | | | | | |
| *OCULAR SENSATION DISORDERS* | | | | | |
| ASTHENOPIA | 0 | 0 | 0 | 0 | 0 |
| PHOTOPHOBIA | 1 | 1 | 0 | 0 | 0 |
| ***VISION DISORDERS*** | | | | | |
| *VISUAL DISORDERS NEC* | | | | | |
| VISION BLURRED | 1 | 0 | 1 | 0 | 0 |
| **GASTROINTESTINAL DISORDERS** | | | | | |
| ***DENTAL AND GINGIVAL CONDITIONS*** | | | | | |
| *DENTAL DISORDERS NEC* | | | | | |
| TEETHING | 0 | 0 | 0 | 0 | 0 |
| ***GASTROINTESTINAL INFLAMMATORY CONDITIONS*** | | | | | |
| *GASTROINTESTINAL INFLAMMATORY DISORDERS NEC* | | | | | |
| GASTROINTESTINAL TRACT IRRITATION | 0 | 0 | 0 | 0 | 0 |
| ***GASTROINTESTINAL MOTILITY AND DEFAECATION CONDITIONS*** | | | | | |
| *DIARRHOEA (EXCL INFECTIVE)* | | | | | |
| DIARRHOEA | 1 | 1 | 0 | 0 | 0 |
| *GASTROINTESTINAL ATONIC AND HYPOMOTILITY DISORDERS NEC* | | | | | |
| CONSTIPATION | 0 | 0 | 0 | 0 | 0 |

| | | | | | |
|---|---|---|---|---|---|
| ***GASTROINTESTINAL SIGNS AND SYMPTOMS*** | | | | | |
| *GASTROINTESTINAL AND ABDOMINAL PAINS (EXCL ORAL AND THROAT)* | | | | | |
| ABDOMINAL PAIN | 1 | 1 | 0 | 0 | 0 |
| ABDOMINAL PAIN UPPER | 0 | 0 | 0 | 0 | 0 |
| *GASTROINTESTINAL SIGNS AND SYMPTOMS NEC* | | | | | |
| ABDOMINAL DISCOMFORT | 2 | 2 | 0 | 0 | 0 |
| *NAUSEA AND VOMITING SYMPTOMS* | | | | | |
| NAUSEA | 13 | 11 | 2 | 0 | 0 |
| VOMITING | 5 | 4 | 1 | 0 | 0 |
| ***ORAL SOFT TISSUE CONDITIONS*** | | | | | |
| *ORAL SOFT TISSUE SIGNS AND SYMPTOMS* | | | | | |
| HYPOAESTHESIA ORAL | 0 | 0 | 0 | 0 | 0 |
| PARAESTHESIA ORAL | 0 | 0 | 0 | 0 | 0 |
| ***TONGUE CONDITIONS*** | | | | | |
| *TONGUE SIGNS AND SYMPTOMS* | | | | | |
| GLOSSODYNIA | 0 | 0 | 0 | 0 | 0 |
| **GENERAL DISORDERS AND ADMINISTRATION SITE CONDITIONS** | | | | | |
| ***ADMINISTRATION SITE REACTIONS*** | | | | | |
| *ADMINISTRATION SITE REACTIONS NEC* | | | | | |
| PUNCTURE SITE BRUISE | 0 | 0 | 0 | 0 | 0 |
| PUNCTURE SITE PAIN | 0 | 0 | 0 | 0 | 0 |
| *INJECTION SITE REACTIONS* | | | | | |
| INJECTION SITE BRUISING | 0 | 0 | 0 | 0 | 0 |
| INJECTION SITE ERYTHEMA | 0 | 0 | 0 | 0 | 0 |
| INJECTION SITE MASS | 0 | 0 | 0 | 0 | 0 |
| INJECTION SITE PAIN | 5 | 4 | 1 | 0 | 0 |
| INJECTION SITE REACTION | 0 | 0 | 0 | 0 | 0 |
| INJECTION SITE URTICARIA | 0 | 0 | 0 | 0 | 0 |
| INJECTION SITE WARMTH | 0 | 0 | 0 | 0 | 0 |
| *VACCINATION SITE REACTIONS* | | | | | |
| VACCINATION SITE PAIN | 2 | 1 | 1 | 0 | 0 |
| ***BODY TEMPERATURE CONDITIONS*** | | | | | |
| *FEBRILE DISORDERS* | | | | | |
| PYREXIA | 51 | 47 | 4 | 0 | 0 |
| ***GENERAL SYSTEM DISORDERS NEC*** | | | | | |
| *ASTHENIC CONDITIONS* | | | | | |
| ASTHENIA | 0 | 0 | 0 | 0 | 0 |
| FATIGUE | 46 | 34 | 12 | 0 | 0 |
| MALAISE | 2 | 2 | 0 | 0 | 0 |
| *FEELINGS AND SENSATIONS NEC* | | | | | |
| CHILLS | 32 | 29 | 3 | 0 | 0 |
| FEELING ABNORMAL | 0 | 0 | 0 | 0 | 0 |
| FEELING COLD | 2 | 2 | 0 | 0 | 0 |
| FEELING HOT | 1 | 1 | 0 | 0 | 0 |
| FEELING OF BODY TEMPERATURE CHANGE | 1 | 1 | 0 | 0 | 0 |
| HANGOVER | 0 | 0 | 0 | 0 | 0 |
| HUNGER | 1 | 1 | 0 | 0 | 0 |
| THIRST | 1 | 1 | 0 | 0 | 0 |
| *GENERAL SIGNS AND SYMPTOMS NEC* | | | | | |
| CRYING | 1 | 1 | 0 | 0 | 0 |
| ILLNESS | 3 | 3 | 0 | 0 | 0 |
| INFLUENZA LIKE ILLNESS | 6 | 6 | 0 | 0 | 0 |

| | | | | | |
|---|---|---|---|---|---|
| LOCAL REACTION | 0 | 0 | 0 | 0 | 0 |
| PERIPHERAL SWELLING | 0 | 0 | 0 | 0 | 0 |
| SWELLING | 0 | 0 | 0 | 0 | 0 |
| *PAIN AND DISCOMFORT NEC* | | | | | |
| AXILLARY PAIN | 0 | 0 | 0 | 0 | 0 |
| CHEST PAIN | 0 | 0 | 0 | 0 | 0 |
| PAIN | 11 | 8 | 3 | 0 | 0 |
| TENDERNESS | 2 | 2 | 0 | 0 | 0 |
| **INFECTIONS AND INFESTATIONS** | | | | | |
| ***BACTERIAL INFECTIOUS DISORDERS*** | | | | | |
| *BACTERIAL INFECTIONS NEC* | | | | | |
| PERIORBITAL CELLULITIS | 0 | 0 | 0 | 0 | 0 |
| ***INFECTIONS - PATHOGEN UNSPECIFIED*** | | | | | |
| *BREAST INFECTIONS* | | | | | |
| MASTITIS | 0 | 0 | 0 | 0 | 0 |
| *UPPER RESPIRATORY TRACT INFECTIONS* | | | | | |
| LARYNGITIS | 0 | 0 | 0 | 0 | 0 |
| NASOPHARYNGITIS | 2 | 2 | 0 | 0 | 0 |
| ***VIRAL INFECTIOUS DISORDERS*** | | | | | |
| *HERPES VIRAL INFECTIONS* | | | | | |
| GENITAL HERPES | 0 | 0 | 0 | 0 | 0 |
| HERPES ZOSTER | 0 | 0 | 0 | 0 | 0 |
| ORAL HERPES | 0 | 0 | 0 | 0 | 0 |
| *INFLUENZA VIRAL INFECTIONS* | | | | | |
| INFLUENZA | 7 | 6 | 1 | 0 | 0 |
| *VIRAL INFECTIONS NEC* | | | | | |
| VIRAL DIARRHOEA | 0 | 0 | 0 | 0 | 0 |
| **INJURY, POISONING AND PROCEDURAL COMPLICATIONS** | | | | | |
| ***INJURIES NEC*** | | | | | |
| *SKIN INJURIES NEC* | | | | | |
| CONTUSION | 1 | 1 | 0 | 0 | 0 |
| ***PROCEDURAL RELATED INJURIES AND COMPLICATIONS NEC*** | | | | | |
| *NON-SITE SPECIFIC PROCEDURAL COMPLICATIONS* | | | | | |
| INJECTION RELATED REACTION | 0 | 0 | 0 | 0 | 0 |
| **INVESTIGATIONS** | | | | | |
| ***CARDIAC AND VASCULAR INVESTIGATIONS (EXCL ENZYME TESTS)*** | | | | | |
| *HEART RATE AND PULSE INVESTIGATIONS* | | | | | |
| HEART RATE | 1 | 1 | 0 | 0 | 0 |
| HEART RATE INCREASED | 1 | 1 | 0 | 0 | 0 |
| ***PHYSICAL EXAMINATION AND ORGAN SYSTEM STATUS TOPICS*** | | | | | |
| *PHYSICAL EXAMINATION PROCEDURES AND ORGAN SYSTEM STATUS* | | | | | |
| BODY TEMPERATURE INCREASED | 1 | 1 | 0 | 0 | 0 |
| ***WATER, ELECTROLYTE AND MINERAL INVESTIGATIONS*** | | | | | |
| *WATER AND ELECTROLYTE ANALYSES NEC* | | | | | |
| VOLUME BLOOD | 0 | 0 | 0 | 0 | 0 |
| **METABOLISM AND NUTRITION DISORDERS** | | | | | |
| ***APPETITE AND GENERAL NUTRITIONAL DISORDERS*** | | | | | |
| *APPETITE DISORDERS* | | | | | |
| DECREASED APPETITE | 3 | 2 | 1 | 0 | 0 |
| ***ELECTROLYTE AND FLUID BALANCE CONDITIONS*** | | | | | |
| *TOTAL FLUID VOLUME DECREASED* | | | | | |
| DEHYDRATION | 1 | 1 | 0 | 0 | 0 |
| **MUSCULOSKELETAL AND CONNECTIVE TISSUE DISORDERS** | | | | | |

| | | | | | |
|---|---|---|---|---|---|
| ***BONE DISORDERS (EXCL CONGENITAL AND FRACTURES)*** | | | | | |
| *BONE RELATED SIGNS AND SYMPTOMS* | | | | | |
| PAIN IN JAW | 1 | 0 | 1 | 0 | 0 |
| ***JOINT DISORDERS*** | | | | | |
| *JOINT RELATED SIGNS AND SYMPTOMS* | | | | | |
| ARTHRALGIA | 5 | 5 | 0 | 0 | 0 |
| ***MUSCLE DISORDERS*** | | | | | |
| *MUSCLE PAINS* | | | | | |
| MYALGIA | 24 | 20 | 4 | 0 | 0 |
| *MUSCLE RELATED SIGNS AND SYMPTOMS NEC* | | | | | |
| MUSCLE FATIGUE | 1 | 1 | 0 | 0 | 0 |
| MUSCLE SPASMS | 0 | 0 | 0 | 0 | 0 |
| *MUSCLE TONE ABNORMALITIES* | | | | | |
| TRISMUS | 0 | 0 | 0 | 0 | 0 |
| ***MUSCULOSKELETAL AND CONNECTIVE TISSUE DISORDERS NEC*** | | | | | |
| *MUSCULOSKELETAL AND CONNECTIVE TISSUE CONDITIONS NEC* | | | | | |
| MUSCULOSKELETAL STIFFNESS | 0 | 0 | 0 | 0 | 0 |
| BACK PAIN | 2 | 2 | 0 | 0 | 0 |
| LIMB DISCOMFORT | 4 | 4 | 0 | 0 | 0 |
| NECK PAIN | 0 | 0 | 0 | 0 | 0 |
| PAIN IN EXTREMITY | 21 | 18 | 3 | 0 | 0 |
| **NERVOUS SYSTEM DISORDERS** | | | | | |
| ***CRANIAL NERVE DISORDERS (EXCL NEOPLASMS)*** | | | | | |
| *FACIAL CRANIAL NERVE DISORDERS* | | | | | |
| BELL'S PALSY | 0 | 0 | 0 | 0 | 0 |
| *OLFACTORY NERVE DISORDERS* | | | | | |
| ANOSMIA | 0 | 0 | 0 | 0 | 0 |
| PAROSMIA | 1 | 0 | 1 | 0 | 0 |
| ***HEADACHES*** | | | | | |
| *HEADACHES NEC* | | | | | |
| CLUSTER HEADACHE | 0 | 0 | 0 | 0 | 0 |
| HEADACHE | 54 | 43 | 11 | 0 | 0 |
| SINUS HEADACHE | 3 | 2 | 1 | 0 | 0 |
| TENSION HEADACHE | 3 | 3 | 0 | 0 | 0 |
| *MIGRAINE HEADACHES* | | | | | |
| MIGRAINE WITH AURA | 0 | 0 | 0 | 0 | 0 |
| TYPICAL AURA WITHOUT HEADACHE | 0 | 0 | 0 | 0 | 0 |
| ***MENTAL IMPAIRMENT DISORDERS*** | | | | | |
| *MENTAL IMPAIRMENT (EXCL DEMENTIA AND MEMORY LOSS)* | | | | | |
| DISTURBANCE IN ATTENTION | 0 | 0 | 0 | 0 | 0 |
| ***MOVEMENT DISORDERS (INCL PARKINSONISM)*** | | | | | |
| *DYSKINESIAS AND MOVEMENT DISORDERS NEC* | | | | | |
| CLUMSINESS | 1 | 1 | 0 | 0 | 0 |
| *TREMOR (EXCL CONGENITAL)* | | | | | |
| TREMOR | 3 | 3 | 0 | 0 | 0 |
| ***NEUROLOGICAL DISORDERS NEC*** | | | | | |
| *DISTURBANCES IN CONSCIOUSNESS NEC* | | | | | |
| LETHARGY | 2 | 2 | 0 | 0 | 0 |
| SOMNOLENCE | 1 | 1 | 0 | 0 | 0 |
| SYNCOPE | 0 | 0 | 0 | 0 | 0 |
| *NEUROLOGICAL SIGNS AND SYMPTOMS NEC* | | | | | |
| AGITATION NEONATAL | 0 | 0 | 0 | 0 | 0 |
| BRAIN FOG | 1 | 1 | 0 | 0 | 0 |

| | | | | | |
|---|---|---|---|---|---|
| DIZZINESS | 9 | 8 | 1 | 0 | 0 |
| DIZZINESS EXERTIONAL | 0 | 0 | 0 | 0 | 0 |
| PRESYNCOPE | 1 | 1 | 0 | 0 | 0 |
| *PARAESTHESIAS AND DYSAESTHESIAS* | | | | | |
| PARAESTHESIA | 1 | 1 | 0 | 0 | 0 |
| *SENSORY ABNORMALITIES NEC* | | | | | |
| DYSGEUSIA | 1 | 1 | 0 | 0 | 0 |
| NEURALGIA | 0 | 0 | 0 | 0 | 0 |
| TASTE DISORDER | 0 | 0 | 0 | 0 | 0 |
| ***SEIZURES (INCL SUBTYPES)*** | | | | | |
| *SEIZURES AND SEIZURE DISORDERS NEC* | | | | | |
| SEIZURE | 0 | 0 | 0 | 0 | 0 |
| **PREGNANCY, PUERPERIUM AND PERINATAL CONDITIONS** | | | | | |
| ***ABORTIONS AND STILLBIRTH*** | | | | | |
| *ABORTIONS SPONTANEOUS* | | | | | |
| ABORTION SPONTANEOUS | 5 | 3 | 2 | 0 | 0 |
| *STILLBIRTH AND FOETAL DEATH* | | | | | |
| FOETAL DEATH | 1 | 1 | 0 | 0 | 0 |
| ***FOETAL COMPLICATIONS*** | | | | | |
| *FOETAL COMPLICATIONS NEC* | | | | | |
| FOETAL DISORDER | 0 | 0 | 0 | 0 | 0 |
| FOETAL HYPOKINESIA | 0 | 0 | 0 | 0 | 0 |
| *FOETAL GROWTH COMPLICATIONS* | | | | | |
| FOETAL MACROSOMIA | 0 | 0 | 0 | 0 | 0 |
| ***MATERNAL COMPLICATIONS OF PREGNANCY*** | | | | | |
| *MATERNAL COMPLICATIONS OF PREGNANCY NEC* | | | | | |
| MORNING SICKNESS | 0 | 0 | 0 | 0 | 0 |
| ***PLACENTAL, AMNIOTIC AND CAVITY DISORDERS (EXCL HAEMORRHAGES)*** | | | | | |
| *PLACENTAL ABNORMALITIES (EXCL NEOPLASMS)* | | | | | |
| PLACENTAL INFARCTION | 1 | 1 | 0 | 0 | 0 |
| ***PREGNANCY, LABOUR, DELIVERY AND POSTPARTUM CONDITIONS*** | | | | | |
| *NORMAL PREGNANCY, LABOUR AND DELIVERY* | | | | | |
| PREGNANCY | 0 | 0 | 0 | 0 | 0 |
| UTERINE CONTRACTIONS DURING PREGNANCY | 0 | 0 | 0 | 0 | 0 |
| **PSYCHIATRIC DISORDERS** | | | | | |
| ***ANXIETY DISORDERS AND SYMPTOMS*** | | | | | |
| *ANXIETY SYMPTOMS* | | | | | |
| AGITATION | 0 | 0 | 0 | 0 | 0 |
| ANXIETY | 0 | 0 | 0 | 0 | 0 |
| ***COGNITIVE AND ATTENTION DISORDERS AND DISTURBANCES*** | | | | | |
| *COGNITIVE AND ATTENTION DISORDERS AND DISTURBANCES NEC* | | | | | |
| MENTAL FATIGUE | 0 | 0 | 0 | 0 | 0 |
| ***DELIRIA (INCL CONFUSION)*** | | | | | |
| *CONFUSION AND DISORIENTATION* | | | | | |
| CONFUSIONAL STATE | 0 | 0 | 0 | 0 | 0 |
| ***DEPRESSED MOOD DISORDERS AND DISTURBANCES*** | | | | | |
| *MOOD ALTERATIONS WITH DEPRESSIVE SYMPTOMS* | | | | | |
| DEPRESSED MOOD | 0 | 0 | 0 | 0 | 0 |
| ***DISTURBANCES IN THINKING AND PERCEPTION*** | | | | | |
| *HALLUCINATIONS (EXCL SLEEP-RELATED)* | | | | | |
| HALLUCINATION | 0 | 0 | 0 | 0 | 0 |
| ***MOOD DISORDERS AND DISTURBANCES NEC*** | | | | | |
| *EMOTIONAL AND MOOD DISTURBANCES NEC* | | | | | |

| | | | | | |
|---|---|---|---|---|---|
| EMOTIONAL DISORDER | 0 | 0 | 0 | 0 | 0 |
| ***SLEEP DISORDERS AND DISTURBANCES*** | | | | | |
| *DISTURBANCES IN INITIATING AND MAINTAINING SLEEP* | | | | | |
| INITIAL INSOMNIA | 0 | 0 | 0 | 0 | 0 |
| INSOMNIA | 3 | 2 | 1 | 0 | 0 |
| **REPRODUCTIVE SYSTEM AND BREAST DISORDERS** | | | | | |
| ***MENSTRUAL CYCLE AND UTERINE BLEEDING DISORDERS*** | | | | | |
| *MENSTRUATION AND UTERINE BLEEDING NEC* | | | | | |
| MENSTRUAL DISORDER | 0 | 0 | 0 | 0 | 0 |
| *MENSTRUATION WITH DECREASED BLEEDING* | | | | | |
| MENSTRUATION DELAYED | 1 | 0 | 1 | 0 | 0 |
| OLIGOMENORRHOEA | 0 | 0 | 0 | 0 | 0 |
| *MENSTRUATION WITH INCREASED BLEEDING* | | | | | |
| HEAVY MENSTRUAL BLEEDING | 0 | 0 | 0 | 0 | 0 |
| ***VULVOVAGINAL DISORDERS (EXCL INFECTIONS AND INFLAMMATIONS)*** | | | | | |
| *VULVOVAGINAL DISORDERS NEC* | | | | | |
| VAGINAL HAEMORRHAGE | 1 | 1 | 0 | 0 | 0 |
| **RESPIRATORY, THORACIC AND MEDIASTINAL DISORDERS** | | | | | |
| ***RESPIRATORY DISORDERS NEC*** | | | | | |
| *BREATHING ABNORMALITIES* | | | | | |
| DYSPNOEA | 1 | 0 | 1 | 0 | 0 |
| *COUGHING AND ASSOCIATED SYMPTOMS* | | | | | |
| COUGH | 1 | 1 | 0 | 0 | 0 |
| ***RESPIRATORY TRACT SIGNS AND SYMPTOMS*** | | | | | |
| *UPPER RESPIRATORY TRACT SIGNS AND SYMPTOMS* | | | | | |
| DRY THROAT | 0 | 0 | 0 | 0 | 0 |
| OROPHARYNGEAL PAIN | 3 | 3 | 0 | 0 | 0 |
| PARANASAL SINUS DISCOMFORT | 0 | 0 | 0 | 0 | 0 |
| RHINORRHOEA | 1 | 1 | 0 | 0 | 0 |
| SINUS PAIN | 0 | 0 | 0 | 0 | 0 |
| SNEEZING | 0 | 0 | 0 | 0 | 0 |
| ***UPPER RESPIRATORY TRACT DISORDERS (EXCL INFECTIONS)*** | | | | | |
| *NASAL CONGESTION AND INFLAMMATIONS* | | | | | |
| NASAL CONGESTION | 0 | 0 | 0 | 0 | 0 |
| **SKIN AND SUBCUTANEOUS TISSUE DISORDERS** | | | | | |
| ***ANGIOEDEMA AND URTICARIA*** | | | | | |
| *URTICARIAS* | | | | | |
| URTICARIA | 0 | 0 | 0 | 0 | 0 |
| ***CORNIFICATION AND DYSTROPHIC SKIN DISORDERS*** | | | | | |
| *SKIN DYSTROPHIES* | | | | | |
| HYPERTROPHIC SCAR | 0 | 0 | 0 | 0 | 0 |
| ***EPIDERMAL AND DERMAL CONDITIONS*** | | | | | |
| *DERMAL AND EPIDERMAL CONDITIONS NEC* | | | | | |
| SKIN WARM | 0 | 0 | 0 | 0 | 0 |
| *DERMATITIS AND ECZEMA* | | | | | |
| DERMATITIS ALLERGIC | 1 | 1 | 0 | 0 | 0 |
| *ERYTHEMAS* | | | | | |
| ERYTHEMA | 0 | 0 | 0 | 0 | 0 |
| *PRURITUS NEC* | | | | | |
| PRURITUS | 1 | 1 | 0 | 0 | 0 |
| *RASHES, ERUPTIONS AND EXANTHEMS NEC* | | | | | |
| RASH | 1 | 1 | 0 | 0 | 0 |
| RASH PRURITIC | 1 | 1 | 0 | 0 | 0 |

| | | | | | |
|---|---|---|---|---|---|
| ***SKIN APPENDAGE CONDITIONS*** | | | | | |
| *APOCRINE AND ECCRINE GLAND DISORDERS* | | | | | |
| COLD SWEAT | 1 | 1 | 0 | 0 | 0 |
| HYPERHIDROSIS | 3 | 3 | 0 | 0 | 0 |
| NIGHT SWEATS | 0 | 0 | 0 | 0 | 0 |
| **VASCULAR DISORDERS** | | | | | |
| ***DECREASED AND NONSPECIFIC BLOOD PRESSURE DISORDERS AND SHOCK*** | | | | | |
| *BLOOD PRESSURE DISORDERS NEC* | | | | | |
| BLOOD PRESSURE FLUCTUATION | 1 | 1 | 0 | 0 | 0 |
| *VASCULAR HYPOTENSIVE DISORDERS* | | | | | |
| HYPOTENSION | 0 | 0 | 0 | 0 | 0 |
| ***EMBOLISM AND THROMBOSIS*** | | | | | |
| *NON-SITE SPECIFIC EMBOLISM AND THROMBOSIS* | | | | | |
| EMBOLISM | 0 | 0 | 0 | 0 | 0 |
| ***VASCULAR DISORDERS NEC*** | | | | | |
| *NON-SITE SPECIFIC VASCULAR DISORDERS NEC* | | | | | |
| VASCULAR PAIN | 0 | 0 | 0 | 0 | 0 |
| *PERIPHERAL VASCULAR DISORDERS NEC* | | | | | |
| HOT FLUSH | 0 | 0 | 0 | 0 | 0 |
| ***VASCULAR HAEMORRHAGIC DISORDERS*** | | | | | |
| *HAEMORRHAGES NEC* | | | | | |
| HAEMORRHAGE | 0 | 0 | 0 | 0 | 0 |
| **TOTAL ADR EVENTS** | ***377*** | ***318*** | ***59*** | ***0*** | ***0*** |

**TABLE S2-4. Moderna COVID-19 vaccine: ADR listing for events reported by pregnant patients, in those reporting any vaccination dose**

| | Patients with any vaccination dose: ADR Counts | | | | |
|---|---|---|---|---|---|
| **REACTION TERM (SOC, *HLGT*, *HLT*, PT)** | All doses | 1st dose | 2nd dose | 3rd dose | Other doses |
| ***(freetext)*** | 0 | 0 | 0 | 0 | 0 |
| **BLOOD AND LYMPHATIC SYSTEM DISORDERS** | | | | | |
| ***SPLEEN, LYMPHATIC AND RETICULOENDOTHELIAL SYSTEM DISORDERS*** | | | | | |
| *LYMPHATIC SYSTEM DISORDERS NEC* | 0 | 0 | 0 | 0 | 0 |
| LYMPH NODE PAIN | 1 | 1 | 0 | 0 | 0 |
| LYMPHADENITIS | 2 | 1 | 1 | 0 | 0 |
| LYMPHADENOPATHY | | | | | |
| **CARDIAC DISORDERS** | | | | | |
| ***CARDIAC ARRHYTHMIAS*** | | | | | |
| *RATE AND RHYTHM DISORDERS NEC* | | | | | |
| TACHYCARDIA | 0 | 0 | 0 | 0 | 0 |
| ***CARDIAC DISORDERS, SIGNS AND SYMPTOMS NEC*** | | | | | |
| *CARDIAC SIGNS AND SYMPTOMS NEC* | | | | | |
| PALPITATIONS | 2 | 1 | 1 | 0 | 0 |
| **EAR AND LABYRINTH DISORDERS** | | | | | |
| ***AURAL DISORDERS NEC*** | | | | | |
| *EAR DISORDERS NEC* | | | | | |
| EAR PAIN | 0 | 0 | 0 | 0 | 0 |
| ***INNER EAR AND VIIITH CRANIAL NERVE DISORDERS*** | | | | | |
| *INNER EAR SIGNS AND SYMPTOMS* | | | | | |
| TINNITUS | 1 | 0 | 0 | 1 | 0 |
| VERTIGO | 0 | 0 | 0 | 0 | 0 |
| **EYE DISORDERS** | | | | | |
| ***EYE DISORDERS NEC*** | | | | | |
| *OCULAR DISORDERS NEC* | | | | | |
| EYE PAIN | 1 | 1 | 0 | 0 | 0 |
| ***OCULAR SENSORY SYMPTOMS NEC*** | | | | | |
| *OCULAR SENSATION DISORDERS* | | | | | |
| ASTHENOPIA | 0 | 0 | 0 | 0 | 0 |
| PHOTOPHOBIA | 0 | 0 | 0 | 0 | 0 |
| ***VISION DISORDERS*** | | | | | |
| *VISUAL DISORDERS NEC* | | | | | |
| VISION BLURRED | 1 | 1 | 0 | 0 | 0 |
| **GASTROINTESTINAL DISORDERS** | | | | | |
| ***DENTAL AND GINGIVAL CONDITIONS*** | | | | | |
| *DENTAL DISORDERS NEC* | | | | | |
| TEETHING | 0 | 0 | 0 | 0 | 0 |
| ***GASTROINTESTINAL INFLAMMATORY CONDITIONS*** | | | | | |
| *GASTROINTESTINAL INFLAMMATORY DISORDERS NEC* | | | | | |
| GASTROINTESTINAL TRACT IRRITATION | 0 | 0 | 0 | 0 | 0 |
| ***GASTROINTESTINAL MOTILITY AND DEFAECATION CONDITIONS*** | | | | | |
| *DIARRHOEA (EXCL INFECTIVE)* | | | | | |
| DIARRHOEA | 1 | 1 | 0 | 0 | 0 |
| *GASTROINTESTINAL ATONIC AND HYPOMOTILITY DISORDERS NEC* | | | | | |
| CONSTIPATION | 0 | 0 | 0 | 0 | 0 |

| | | | | | |
|---|---|---|---|---|---|
| ***GASTROINTESTINAL SIGNS AND SYMPTOMS*** | | | | | |
| *GASTROINTESTINAL AND ABDOMINAL PAINS (EXCL ORAL AND THROAT)* | | | | | |
| ABDOMINAL PAIN | 0 | 0 | 0 | 0 | 0 |
| ABDOMINAL PAIN UPPER | 0 | 0 | 0 | 0 | 0 |
| *GASTROINTESTINAL SIGNS AND SYMPTOMS NEC* | | | | | |
| ABDOMINAL DISCOMFORT | 0 | 0 | 0 | 0 | 0 |
| *NAUSEA AND VOMITING SYMPTOMS* | | | | | |
| NAUSEA | 11 | 6 | 3 | 2 | 0 |
| VOMITING | 4 | 1 | 3 | 0 | 0 |
| ***ORAL SOFT TISSUE CONDITIONS*** | | | | | |
| *ORAL SOFT TISSUE SIGNS AND SYMPTOMS* | | | | | |
| HYPOAESTHESIA ORAL | | | | | |
| PARAESTHESIA ORAL | 0 | 0 | 0 | 0 | 0 |
| ***TONGUE CONDITIONS*** | | | | | |
| *TONGUE SIGNS AND SYMPTOMS* | | | | | |
| GLOSSODYNIA | 0 | 0 | 0 | 0 | 0 |
| **GENERAL DISORDERS AND ADMINISTRATION SITE CONDITIONS** | | | | | |
| ***ADMINISTRATION SITE REACTIONS*** | | | | | |
| *ADMINISTRATION SITE REACTIONS NEC* | | | | | |
| PUNCTURE SITE BRUISE | 1 | 0 | 0 | 1 | 0 |
| PUNCTURE SITE PAIN | 0 | 0 | 0 | 0 | 0 |
| *INJECTION SITE REACTIONS* | | | | | |
| INJECTION SITE BRUISING | 0 | 0 | 0 | 0 | 0 |
| INJECTION SITE ERYTHEMA | 0 | 0 | 0 | 0 | 0 |
| INJECTION SITE MASS | 1 | 1 | 0 | 0 | 0 |
| INJECTION SITE PAIN | 4 | 1 | 1 | 2 | 0 |
| INJECTION SITE REACTION | 0 | 0 | 0 | 0 | 0 |
| INJECTION SITE URTICARIA | 0 | 0 | 0 | 0 | 0 |
| INJECTION SITE WARMTH | 0 | 0 | 0 | 0 | 0 |
| *VACCINATION SITE REACTIONS* | | | | | |
| VACCINATION SITE PAIN | 0 | 0 | 0 | 0 | 0 |
| ***BODY TEMPERATURE CONDITIONS*** | | | | | |
| *FEBRILE DISORDERS* | | | | | |
| PYREXIA | 24 | 5 | 12 | 7 | 0 |
| ***GENERAL SYSTEM DISORDERS NEC*** | | | | | |
| *ASTHENIC CONDITIONS* | | | | | |
| ASTHENIA | 2 | 1 | 0 | 1 | 0 |
| FATIGUE | 47 | 21 | 14 | 11 | 1 |
| MALAISE | 4 | 2 | 0 | 1 | 1 |
| *FEELINGS AND SENSATIONS NEC* | | | | | |
| CHILLS | 14 | 2 | 6 | 6 | 0 |
| FEELING ABNORMAL | 0 | 0 | 0 | 0 | 0 |
| FEELING COLD | 0 | 0 | 0 | 0 | 0 |
| FEELING HOT | 0 | 0 | 0 | 0 | 0 |
| FEELING OF BODY TEMPERATURE CHANGE | 0 | 0 | 0 | 0 | 0 |
| HANGOVER | 0 | 0 | 0 | 0 | 0 |
| HUNGER | 0 | 0 | 0 | 0 | 0 |
| THIRST | 0 | 0 | 0 | 0 | 0 |
| *GENERAL SIGNS AND SYMPTOMS NEC* | | | | | |
| CRYING | 0 | 0 | 0 | 0 | 0 |
| ILLNESS | 2 | 1 | 0 | 1 | 0 |
| INFLUENZA LIKE ILLNESS | 2 | 0 | 1 | 1 | 0 |

| | | | | | |
|---|---|---|---|---|---|
| LOCAL REACTION | 0 | 0 | 0 | 0 | 0 |
| PERIPHERAL SWELLING | 1 | 1 | 0 | 0 | 0 |
| SWELLING | 2 | 1 | 0 | 1 | 0 |
| *PAIN AND DISCOMFORT NEC* | | | | | |
| AXILLARY PAIN | 1 | 1 | 0 | 0 | 0 |
| CHEST PAIN | 0 | 0 | 0 | 0 | 0 |
| PAIN | 11 | 3 | 7 | 1 | 0 |
| TENDERNESS | 0 | 0 | 0 | 0 | 0 |
| **INFECTIONS AND INFESTATIONS** | | | | | |
| ***BACTERIAL INFECTIOUS DISORDERS*** | | | | | |
| *BACTERIAL INFECTIONS NEC* | | | | | |
| PERIORBITAL CELLULITIS | 0 | 0 | 0 | 0 | 0 |
| ***INFECTIONS - PATHOGEN UNSPECIFIED*** | | | | | |
| *BREAST INFECTIONS* | | | | | |
| MASTITIS | 0 | 0 | 0 | 0 | 0 |
| *UPPER RESPIRATORY TRACT INFECTIONS* | | | | | |
| LARYNGITIS | 1 | 1 | 0 | 0 | 0 |
| NASOPHARYNGITIS | 3 | 1 | 1 | 1 | 0 |
| ***VIRAL INFECTIOUS DISORDERS*** | | | | | |
| *HERPES VIRAL INFECTIONS* | | | | | |
| GENITAL HERPES | 0 | 0 | 0 | 0 | 0 |
| HERPES ZOSTER | 0 | 0 | 0 | 0 | 0 |
| ORAL HERPES | 0 | 0 | 0 | 0 | 0 |
| *INFLUENZA VIRAL INFECTIONS* | | | | | |
| INFLUENZA | 8 | 2 | 4 | 2 | 0 |
| *VIRAL INFECTIONS NEC* | | | | | |
| VIRAL DIARRHOEA | 0 | 0 | 0 | 0 | 0 |
| **INJURY, POISONING AND PROCEDURAL COMPLICATIONS** | | | | | |
| ***INJURIES NEC*** | | | | | |
| *SKIN INJURIES NEC* | | | | | |
| CONTUSION | 0 | 0 | 0 | 0 | 0 |
| ***PROCEDURAL RELATED INJURIES AND COMPLICATIONS NEC*** | | | | | |
| *NON-SITE SPECIFIC PROCEDURAL COMPLICATIONS* | | | | | |
| INJECTION RELATED REACTION | 1 | 0 | 0 | 1 | 0 |
| **INVESTIGATIONS** | | | | | |
| ***CARDIAC AND VASCULAR INVESTIGATIONS (EXCL ENZYME TESTS)*** | | | | | |
| *HEART RATE AND PULSE INVESTIGATIONS* | | | | | |
| HEART RATE | 0 | 0 | 0 | 0 | 0 |
| HEART RATE INCREASED | 0 | 0 | 0 | 0 | 0 |
| ***PHYSICAL EXAMINATION AND ORGAN SYSTEM STATUS TOPICS*** | | | | | |
| *PHYSICAL EXAMINATION PROCEDURES AND ORGAN SYSTEM STATUS* | | | | | |
| BODY TEMPERATURE INCREASED | 1 | 0 | 0 | 1 | 0 |
| ***WATER, ELECTROLYTE AND MINERAL INVESTIGATIONS*** | | | | | |
| *WATER AND ELECTROLYTE ANALYSES NEC* | | | | | |
| VOLUME BLOOD | 0 | 0 | 0 | 0 | 0 |
| **METABOLISM AND NUTRITION DISORDERS** | | | | | |
| ***APPETITE AND GENERAL NUTRITIONAL DISORDERS*** | | | | | |
| *APPETITE DISORDERS* | | | | | |
| DECREASED APPETITE | 0 | 0 | 0 | 0 | 0 |
| ***ELECTROLYTE AND FLUID BALANCE CONDITIONS*** | | | | | |
| *TOTAL FLUID VOLUME DECREASED* | | | | | |
| DEHYDRATION | 0 | 0 | 0 | 0 | 0 |
| **MUSCULOSKELETAL AND CONNECTIVE TISSUE DISORDERS** | | | | | |

| | | | | | |
|---|---|---|---|---|---|
| ***BONE DISORDERS (EXCL CONGENITAL AND FRACTURES)*** | | | | | |
| *BONE RELATED SIGNS AND SYMPTOMS* | | | | | |
| PAIN IN JAW | 1 | 0 | 1 | 0 | 0 |
| ***JOINT DISORDERS*** | | | | | |
| *JOINT RELATED SIGNS AND SYMPTOMS* | | | | | |
| ARTHRALGIA | 5 | 3 | 2 | 0 | 0 |
| ***MUSCLE DISORDERS*** | | | | | |
| *MUSCLE PAINS* | | | | | |
| MYALGIA | 15 | 5 | 6 | 4 | 0 |
| *MUSCLE RELATED SIGNS AND SYMPTOMS NEC* | | | | | |
| MUSCLE FATIGUE | 1 | 0 | 0 | 1 | 0 |
| MUSCLE SPASMS | 0 | 0 | 0 | 0 | 0 |
| *MUSCLE TONE ABNORMALITIES* | | | | | |
| TRISMUS | 1 | 0 | 1 | 0 | 0 |
| ***MUSCULOSKELETAL AND CONNECTIVE TISSUE DISORDERS NEC*** | | | | | |
| *MUSCULOSKELETAL AND CONNECTIVE TISSUE CONDITIONS NEC* | | | | | |
| MUSCULOSKELETAL STIFFNESS | 3 | 1 | 2 | 0 | 0 |
| BACK PAIN | 3 | 1 | 2 | 0 | 0 |
| LIMB DISCOMFORT | 3 | 2 | 1 | 0 | 0 |
| NECK PAIN | 1 | 0 | 1 | 0 | 0 |
| PAIN IN EXTREMITY | 59 | 49 | 6 | 4 | 0 |
| **NERVOUS SYSTEM DISORDERS** | | | | | |
| ***CRANIAL NERVE DISORDERS (EXCL NEOPLASMS)*** | | | | | |
| *FACIAL CRANIAL NERVE DISORDERS* | | | | | |
| BELL'S PALSY | 0 | 0 | 0 | 0 | 0 |
| *OLFACTORY NERVE DISORDERS* | | | | | |
| ANOSMIA | 0 | 0 | 0 | 0 | 0 |
| PAROSMIA | 0 | 0 | 0 | 0 | 0 |
| ***HEADACHES*** | | | | | |
| *HEADACHES NEC* | | | | | |
| CLUSTER HEADACHE | 0 | 0 | 0 | 0 | 0 |
| HEADACHE | 31 | 12 | 14 | 5 | 0 |
| SINUS HEADACHE | 0 | 0 | 0 | 0 | 0 |
| TENSION HEADACHE | 2 | 1 | 1 | 0 | 0 |
| *MIGRAINE HEADACHES* | | | | | |
| MIGRAINE WITH AURA | 1 | 0 | 1 | 0 | 0 |
| TYPICAL AURA WITHOUT HEADACHE | 0 | 0 | 0 | 0 | 0 |
| ***MENTAL IMPAIRMENT DISORDERS*** | | | | | |
| *MENTAL IMPAIRMENT (EXCL DEMENTIA AND MEMORY LOSS)* | | | | | |
| DISTURBANCE IN ATTENTION | 0 | 0 | 0 | 0 | 0 |
| ***MOVEMENT DISORDERS (INCL PARKINSONISM)*** | | | | | |
| *DYSKINESIAS AND MOVEMENT DISORDERS NEC* | | | | | |
| CLUMSINESS | 0 | 0 | 0 | 0 | 0 |
| *TREMOR (EXCL CONGENITAL)* | | | | | |
| TREMOR | 1 | 0 | 1 | 0 | 0 |
| ***NEUROLOGICAL DISORDERS NEC*** | | | | | |
| *DISTURBANCES IN CONSCIOUSNESS NEC* | | | | | |
| LETHARGY | 0 | 0 | 0 | 0 | 0 |
| SOMNOLENCE | 1 | 0 | 1 | 0 | 0 |
| SYNCOPE | 0 | 0 | 0 | 0 | 0 |
| *NEUROLOGICAL SIGNS AND SYMPTOMS NEC* | | | | | |
| AGITATION NEONATAL | 0 | 0 | 0 | 0 | 0 |
| BRAIN FOG | 0 | 0 | 0 | 0 | 0 |

| | | | | | |
|---|---|---|---|---|---|
| DIZZINESS | 8 | 4 | 2 | 2 | 0 |
| DIZZINESS EXERTIONAL | 1 | 1 | 0 | 0 | 0 |
| PRESYNCOPE | 0 | 0 | 0 | 0 | 0 |
| *PARAESTHESIAS AND DYSAESTHESIAS* | | | | | |
| PARAESTHESIA | 0 | 0 | 0 | 0 | 0 |
| *SENSORY ABNORMALITIES NEC* | | | | | |
| DYSGEUSIA | 0 | 0 | 0 | 0 | 0 |
| NEURALGIA | 0 | 0 | 0 | 0 | 0 |
| TASTE DISORDER | 0 | 0 | 0 | 0 | 0 |
| ***SEIZURES (INCL SUBTYPES)*** | | | | | |
| *SEIZURES AND SEIZURE DISORDERS NEC* | | | | | |
| SEIZURE | 0 | 0 | 0 | 0 | 0 |
| **PREGNANCY, PUERPERIUM AND PERINATAL CONDITIONS** | | | | | |
| ***ABORTIONS AND STILLBIRTH*** | | | | | |
| *ABORTIONS SPONTANEOUS* | | | | | |
| ABORTION SPONTANEOUS | 1 | 1 | 0 | 0 | 0 |
| *STILLBIRTH AND FOETAL DEATH* | | | | | |
| FOETAL DEATH | 0 | 0 | 0 | 0 | 0 |
| ***FOETAL COMPLICATIONS*** | | | | | |
| *FOETAL COMPLICATIONS NEC* | | | | | |
| FOETAL DISORDER | 0 | 0 | 0 | 0 | 0 |
| FOETAL HYPOKINESIA | 0 | 0 | 0 | 0 | 0 |
| *FOETAL GROWTH COMPLICATIONS* | | | | | |
| FOETAL MACROSOMIA | 0 | 0 | 0 | 0 | 0 |
| ***MATERNAL COMPLICATIONS OF PREGNANCY*** | | | | | |
| *MATERNAL COMPLICATIONS OF PREGNANCY NEC* | | | | | |
| MORNING SICKNESS | 0 | 0 | 0 | 0 | 0 |
| ***PLACENTAL, AMNIOTIC AND CAVITY DISORDERS (EXCL HAEMORRHAGES)*** | | | | | |
| *PLACENTAL ABNORMALITIES (EXCL NEOPLASMS)* | | | | | |
| PLACENTAL INFARCTION | 0 | 0 | 0 | 0 | 0 |
| ***PREGNANCY, LABOUR, DELIVERY AND POSTPARTUM CONDITIONS*** | | | | | |
| *NORMAL PREGNANCY, LABOUR AND DELIVERY* | | | | | |
| PREGNANCY | 0 | 0 | 0 | 0 | 0 |
| UTERINE CONTRACTIONS DURING PREGNANCY | 0 | 0 | 0 | 0 | 0 |
| **PSYCHIATRIC DISORDERS** | | | | | |
| ***ANXIETY DISORDERS AND SYMPTOMS*** | | | | | |
| *ANXIETY SYMPTOMS* | | | | | |
| AGITATION | 0 | 0 | 0 | 0 | 0 |
| ANXIETY | 0 | 0 | 0 | 0 | 0 |
| ***COGNITIVE AND ATTENTION DISORDERS AND DISTURBANCES*** | | | | | |
| *COGNITIVE AND ATTENTION DISORDERS AND DISTURBANCES NEC* | | | | | |
| MENTAL FATIGUE | 0 | 0 | 0 | 0 | 0 |
| ***DELIRIA (INCL CONFUSION)*** | | | | | |
| *CONFUSION AND DISORIENTATION* | | | | | |
| CONFUSIONAL STATE | 0 | 0 | 0 | 0 | 0 |
| ***DEPRESSED MOOD DISORDERS AND DISTURBANCES*** | | | | | |
| *MOOD ALTERATIONS WITH DEPRESSIVE SYMPTOMS* | | | | | |
| DEPRESSED MOOD | 0 | 0 | 0 | 0 | 0 |
| ***DISTURBANCES IN THINKING AND PERCEPTION*** | | | | | |
| *HALLUCINATIONS (EXCL SLEEP-RELATED)* | | | | | |
| HALLUCINATION | 0 | 0 | 0 | 0 | 0 |
| ***MOOD DISORDERS AND DISTURBANCES NEC*** | | | | | |
| *EMOTIONAL AND MOOD DISTURBANCES NEC* | | | | | |

| | | | | | |
|---|---|---|---|---|---|
| EMOTIONAL DISORDER | 0 | 0 | 0 | 0 | 0 |
| ***SLEEP DISORDERS AND DISTURBANCES*** | | | | | |
| *DISTURBANCES IN INITIATING AND MAINTAINING SLEEP* | | | | | |
| INITIAL INSOMNIA | 0 | 0 | 0 | 0 | 0 |
| INSOMNIA | 1 | 1 | 0 | 0 | 0 |
| **REPRODUCTIVE SYSTEM AND BREAST DISORDERS** | | | | | |
| ***MENSTRUAL CYCLE AND UTERINE BLEEDING DISORDERS*** | | | | | |
| *MENSTRUATION AND UTERINE BLEEDING NEC* | | | | | |
| MENSTRUAL DISORDER | 0 | 0 | 0 | 0 | 0 |
| *MENSTRUATION WITH DECREASED BLEEDING* | | | | | |
| MENSTRUATION DELAYED | 0 | 0 | 0 | 0 | 0 |
| OLIGOMENORRHOEA | 0 | 0 | 0 | 0 | 0 |
| *MENSTRUATION WITH INCREASED BLEEDING* | | | | | |
| HEAVY MENSTRUAL BLEEDING | 0 | 0 | 0 | 0 | 0 |
| ***VULVOVAGINAL DISORDERS (EXCL INFECTIONS AND INFLAMMATIONS)*** | | | | | |
| *VULVOVAGINAL DISORDERS NEC* | | | | | |
| VAGINAL HAEMORRHAGE | 0 | 0 | 0 | 0 | 0 |
| **RESPIRATORY, THORACIC AND MEDIASTINAL DISORDERS** | | | | | |
| ***RESPIRATORY DISORDERS NEC*** | | | | | |
| *BREATHING ABNORMALITIES* | | | | | |
| DYSPNOEA | 3 | 1 | 2 | 0 | 0 |
| *COUGHING AND ASSOCIATED SYMPTOMS* | | | | | |
| COUGH | 0 | 0 | 0 | 0 | 0 |
| ***RESPIRATORY TRACT SIGNS AND SYMPTOMS*** | | | | | |
| *UPPER RESPIRATORY TRACT SIGNS AND SYMPTOMS* | | | | | |
| DRY THROAT | 0 | 0 | 0 | 0 | 0 |
| OROPHARYNGEAL PAIN | 2 | 2 | 0 | 0 | 0 |
| PARANASAL SINUS DISCOMFORT | 0 | 0 | 0 | 0 | 0 |
| RHINORRHOEA | 1 | 0 | 0 | 1 | 0 |
| SINUS PAIN | 1 | 1 | 0 | 0 | 0 |
| SNEEZING | 0 | 0 | 0 | 0 | 0 |
| ***UPPER RESPIRATORY TRACT DISORDERS (EXCL INFECTIONS)*** | | | | | |
| *NASAL CONGESTION AND INFLAMMATIONS* | | | | | |
| NASAL CONGESTION | 1 | 0 | 1 | 0 | 0 |
| **SKIN AND SUBCUTANEOUS TISSUE DISORDERS** | | | | | |
| ***ANGIOEDEMA AND URTICARIA*** | | | | | |
| *URTICARIAS* | | | | | |
| URTICARIA | 0 | 0 | 0 | 0 | 0 |
| ***CORNIFICATION AND DYSTROPHIC SKIN DISORDERS*** | | | | | |
| *SKIN DYSTROPHIES* | | | | | |
| HYPERTROPHIC SCAR | 0 | 0 | 0 | 0 | 0 |
| ***EPIDERMAL AND DERMAL CONDITIONS*** | | | | | |
| *DERMAL AND EPIDERMAL CONDITIONS NEC* | | | | | |
| SKIN WARM | 0 | 0 | 0 | 0 | 0 |
| *DERMATITIS AND ECZEMA* | | | | | |
| DERMATITIS ALLERGIC | 0 | 0 | 0 | 0 | 0 |
| *ERYTHEMAS* | | | | | |
| ERYTHEMA | 0 | 0 | 0 | 0 | 0 |
| *PRURITUS NEC* | | | | | |
| PRURITUS | 2 | 1 | 0 | 1 | 0 |
| *RASHES, ERUPTIONS AND EXANTHEMS NEC* | | | | | |
| RASH | 3 | 1 | 0 | 2 | 0 |
| RASH PRURITIC | 0 | 0 | 0 | 0 | 0 |

| | | | | | |
|---|---|---|---|---|---|
| ***SKIN APPENDAGE CONDITIONS*** | | | | | |
| *APOCRINE AND ECCRINE GLAND DISORDERS* | | | | | |
| COLD SWEAT | 1 | 1 | 0 | 0 | 0 |
| HYPERHIDROSIS | 0 | 0 | 0 | 0 | 0 |
| NIGHT SWEATS | 1 | 0 | 1 | 0 | 0 |
| **VASCULAR DISORDERS** | | | | | |
| ***DECREASED AND NONSPECIFIC BLOOD PRESSURE DISORDERS AND SHOCK*** | | | | | |
| *BLOOD PRESSURE DISORDERS NEC* | | | | | |
| BLOOD PRESSURE FLUCTUATION | 0 | 0 | 0 | 0 | 0 |
| *VASCULAR HYPOTENSIVE DISORDERS* | | | | | |
| HYPOTENSION | 0 | 0 | 0 | 0 | 0 |
| ***EMBOLISM AND THROMBOSIS*** | | | | | |
| *NON-SITE SPECIFIC EMBOLISM AND THROMBOSIS* | | | | | |
| EMBOLISM | 1 | 1 | 0 | 0 | 0 |
| ***VASCULAR DISORDERS NEC*** | | | | | |
| *NON-SITE SPECIFIC VASCULAR DISORDERS NEC* | | | | | |
| VASCULAR PAIN | 0 | 0 | 0 | 0 | 0 |
| *PERIPHERAL VASCULAR DISORDERS NEC* | | | | | |
| HOT FLUSH | 0 | 0 | 0 | 0 | 0 |
| ***VASCULAR HAEMORRHAGIC DISORDERS*** | | | | | |
| *HAEMORRHAGES NEC* | | | | | |
| HAEMORRHAGE | 0 | 0 | 0 | 0 | 0 |
| **TOTAL ADR EVENTS** | ***309*** | ***146*** | ***100*** | ***61*** | ***2*** |

**TABLE S2-5. Others/Unknown COVID-19 vaccine: ADR listing for events reported by pregnant patients, in those reporting any vaccination dose**

| | Patients with any vaccination dose: ADR Counts | | | | |
|---|---|---|---|---|---|
| **REACTION TERM (SOC, *HLGT*, *HLT*, PT)** | All doses | 1st dose | 2nd dose | 3rd dose | Other doses |
| ***(freetext)*** | 0 | 0 | 0 | 0 | 0 |
| **BLOOD AND LYMPHATIC SYSTEM DISORDERS** | | | | | |
| ***SPLEEN, LYMPHATIC AND RETICULOENDOTHELIAL SYSTEM DISORDERS*** | | | | | |
| *LYMPHATIC SYSTEM DISORDERS NEC* | | | | | |
| LYMPH NODE PAIN | 0 | 0 | 0 | 0 | 0 |
| LYMPHADENITIS | 0 | 0 | 0 | 0 | 0 |
| LYMPHADENOPATHY | 0 | 0 | 0 | 0 | 0 |
| **CARDIAC DISORDERS** | | | | | |
| ***CARDIAC ARRHYTHMIAS*** | | | | | |
| *RATE AND RHYTHM DISORDERS NEC* | | | | | |
| TACHYCARDIA | 0 | 0 | 0 | 0 | 0 |
| ***CARDIAC DISORDERS, SIGNS AND SYMPTOMS NEC*** | | | | | |
| *CARDIAC SIGNS AND SYMPTOMS NEC* | | | | | |
| PALPITATIONS | 0 | 0 | 0 | 0 | 0 |
| **EAR AND LABYRINTH DISORDERS** | | | | | |
| ***AURAL DISORDERS NEC*** | | | | | |
| *EAR DISORDERS NEC* | | | | | |
| EAR PAIN | 0 | 0 | 0 | 0 | 0 |
| ***INNER EAR AND VIIITH CRANIAL NERVE DISORDERS*** | | | | | |
| *INNER EAR SIGNS AND SYMPTOMS* | | | | | |
| TINNITUS | 0 | 0 | 0 | 0 | 0 |
| VERTIGO | 0 | 0 | 0 | 0 | 0 |
| **EYE DISORDERS** | | | | | |
| ***EYE DISORDERS NEC*** | | | | | |
| *OCULAR DISORDERS NEC* | | | | | |
| EYE PAIN | 0 | 0 | 0 | 0 | 0 |
| ***OCULAR SENSORY SYMPTOMS NEC*** | | | | | |
| *OCULAR SENSATION DISORDERS* | | | | | |
| ASTHENOPIA | 0 | 0 | 0 | 0 | 0 |
| PHOTOPHOBIA | 0 | 0 | 0 | 0 | 0 |
| ***VISION DISORDERS*** | | | | | |
| *VISUAL DISORDERS NEC* | | | | | |
| VISION BLURRED | 0 | 0 | 0 | 0 | 0 |
| **GASTROINTESTINAL DISORDERS** | | | | | |
| ***DENTAL AND GINGIVAL CONDITIONS*** | | | | | |
| *DENTAL DISORDERS NEC* | | | | | |
| TEETHING | 0 | 0 | 0 | 0 | 0 |
| ***GASTROINTESTINAL INFLAMMATORY CONDITIONS*** | | | | | |
| *GASTROINTESTINAL INFLAMMATORY DISORDERS NEC* | | | | | |
| GASTROINTESTINAL TRACT IRRITATION | 0 | 0 | 0 | 0 | 0 |
| ***GASTROINTESTINAL MOTILITY AND DEFAECATION CONDITIONS*** | | | | | |
| *DIARRHOEA (EXCL INFECTIVE)* | | | | | |
| DIARRHOEA | 0 | 0 | 0 | 0 | 0 |
| *GASTROINTESTINAL ATONIC AND HYPOMOTILITY DISORDERS NEC* | | | | | |
| CONSTIPATION | 0 | 0 | 0 | 0 | 0 |

| | | | | | |
|---|---|---|---|---|---|
| ***GASTROINTESTINAL SIGNS AND SYMPTOMS*** | | | | | |
| *GASTROINTESTINAL AND ABDOMINAL PAINS (EXCL ORAL AND THROAT)* | | | | | |
| ABDOMINAL PAIN | 0 | 0 | 0 | 0 | 0 |
| ABDOMINAL PAIN UPPER | 0 | 0 | 0 | 0 | 0 |
| *GASTROINTESTINAL SIGNS AND SYMPTOMS NEC* | | | | | |
| ABDOMINAL DISCOMFORT | 0 | 0 | 0 | 0 | 0 |
| *NAUSEA AND VOMITING SYMPTOMS* | | | | | |
| NAUSEA | 1 | 0 | 0 | 1 | 0 |
| VOMITING | 0 | 0 | 0 | 0 | 0 |
| ***ORAL SOFT TISSUE CONDITIONS*** | | | | | |
| *ORAL SOFT TISSUE SIGNS AND SYMPTOMS* | | | | | |
| HYPOAESTHESIA ORAL | 0 | 0 | 0 | 0 | 0 |
| PARAESTHESIA ORAL | 0 | 0 | 0 | 0 | 0 |
| ***TONGUE CONDITIONS*** | | | | | |
| *TONGUE SIGNS AND SYMPTOMS* | | | | | |
| GLOSSODYNIA | 0 | 0 | 0 | 0 | 0 |
| **GENERAL DISORDERS AND ADMINISTRATION SITE CONDITIONS** | | | | | |
| ***ADMINISTRATION SITE REACTIONS*** | | | | | |
| *ADMINISTRATION SITE REACTIONS NEC* | | | | | |
| PUNCTURE SITE BRUISE | 0 | 0 | 0 | 0 | 0 |
| PUNCTURE SITE PAIN | 0 | 0 | 0 | 0 | 0 |
| *INJECTION SITE REACTIONS* | | | | | |
| INJECTION SITE BRUISING | 0 | 0 | 0 | 0 | 0 |
| INJECTION SITE ERYTHEMA | 0 | 0 | 0 | 0 | 0 |
| INJECTION SITE MASS | 0 | 0 | 0 | 0 | 0 |
| INJECTION SITE PAIN | 0 | 0 | 0 | 0 | 0 |
| INJECTION SITE REACTION | 0 | 0 | 0 | 0 | 0 |
| INJECTION SITE URTICARIA | 0 | 0 | 0 | 0 | 0 |
| INJECTION SITE WARMTH | 0 | 0 | 0 | 0 | 0 |
| *VACCINATION SITE REACTIONS* | | | | | |
| VACCINATION SITE PAIN | 0 | 0 | 0 | 0 | 0 |
| ***BODY TEMPERATURE CONDITIONS*** | | | | | |
| *FEBRILE DISORDERS* | | | | | |
| PYREXIA | 0 | 0 | 0 | 0 | 0 |
| ***GENERAL SYSTEM DISORDERS NEC*** | | | | | |
| *ASTHENIC CONDITIONS* | | | | | |
| ASTHENIA | 1 | 1 | 0 | 0 | 0 |
| FATIGUE | 0 | 0 | 0 | 0 | 0 |
| MALAISE | 0 | 0 | 0 | 0 | 0 |
| *FEELINGS AND SENSATIONS NEC* | | | | | |
| CHILLS | 0 | 0 | 0 | 0 | 0 |
| FEELING ABNORMAL | 0 | 0 | 0 | 0 | 0 |
| FEELING COLD | 0 | 0 | 0 | 0 | 0 |
| FEELING HOT | 0 | 0 | 0 | 0 | 0 |
| FEELING OF BODY TEMPERATURE CHANGE | 0 | 0 | 0 | 0 | 0 |
| HANGOVER | 0 | 0 | 0 | 0 | 0 |
| HUNGER | 0 | 0 | 0 | 0 | 0 |
| THIRST | 0 | 0 | 0 | 0 | 0 |
| *GENERAL SIGNS AND SYMPTOMS NEC* | | | | | |
| CRYING | 0 | 0 | 0 | 0 | 0 |
| ILLNESS | 0 | 0 | 0 | 0 | 0 |

| | | | | | |
|---|---|---|---|---|---|
| INFLUENZA LIKE ILLNESS | 0 | 0 | 0 | 0 | 0 |
| LOCAL REACTION | 0 | 0 | 0 | 0 | 0 |
| PERIPHERAL SWELLING | 0 | 0 | 0 | 0 | 0 |
| SWELLING | 0 | 0 | 0 | 0 | 0 |
| *PAIN AND DISCOMFORT NEC* | | | | | |
| AXILLARY PAIN | 0 | 0 | 0 | 0 | 0 |
| CHEST PAIN | 0 | 0 | 0 | 0 | 0 |
| PAIN | 1 | 0 | 0 | 1 | 0 |
| TENDERNESS | 0 | 0 | 0 | 0 | 0 |
| **INFECTIONS AND INFESTATIONS** | | | | | |
| ***BACTERIAL INFECTIOUS DISORDERS*** | | | | | |
| *BACTERIAL INFECTIONS NEC* | | | | | |
| PERIORBITAL CELLULITIS | 0 | 0 | 0 | 0 | 0 |
| ***INFECTIONS - PATHOGEN UNSPECIFIED*** | | | | | |
| *BREAST INFECTIONS* | | | | | |
| MASTITIS | 0 | 0 | 0 | 0 | 0 |
| *UPPER RESPIRATORY TRACT INFECTIONS* | | | | | |
| LARYNGITIS | 0 | 0 | 0 | 0 | 0 |
| NASOPHARYNGITIS | 0 | 0 | 0 | 0 | 0 |
| ***VIRAL INFECTIOUS DISORDERS*** | | | | | |
| *HERPES VIRAL INFECTIONS* | | | | | |
| GENITAL HERPES | 0 | 0 | 0 | 0 | 0 |
| HERPES ZOSTER | 0 | 0 | 0 | 0 | 0 |
| ORAL HERPES | 0 | 0 | 0 | 0 | 0 |
| *INFLUENZA VIRAL INFECTIONS* | | | | | |
| INFLUENZA | 0 | 0 | 0 | 0 | 0 |
| *VIRAL INFECTIONS NEC* | | | | | |
| VIRAL DIARRHOEA | 0 | 0 | 0 | 0 | 0 |
| **INJURY, POISONING AND PROCEDURAL COMPLICATIONS** | | | | | |
| ***INJURIES NEC*** | | | | | |
| *SKIN INJURIES NEC* | | | | | |
| CONTUSION | 0 | 0 | 0 | 0 | 0 |
| ***PROCEDURAL RELATED INJURIES AND COMPLICATIONS NEC*** | | | | | |
| *NON-SITE SPECIFIC PROCEDURAL COMPLICATIONS* | | | | | |
| INJECTION RELATED REACTION | 0 | 0 | 0 | 0 | 0 |
| **INVESTIGATIONS** | | | | | |
| ***CARDIAC AND VASCULAR INVESTIGATIONS (EXCL ENZYME TESTS)*** | | | | | |
| *HEART RATE AND PULSE INVESTIGATIONS* | | | | | |
| HEART RATE | 0 | 0 | 0 | 0 | 0 |
| HEART RATE INCREASED | 0 | 0 | 0 | 0 | 0 |
| ***PHYSICAL EXAMINATION AND ORGAN SYSTEM STATUS TOPICS*** | | | | | |
| *PHYSICAL EXAMINATION PROCEDURES AND ORGAN SYSTEM STATUS* | | | | | |
| BODY TEMPERATURE INCREASED | 0 | 0 | 0 | 0 | 0 |
| ***WATER, ELECTROLYTE AND MINERAL INVESTIGATIONS*** | | | | | |
| *WATER AND ELECTROLYTE ANALYSES NEC* | | | | | |
| VOLUME BLOOD | 0 | 0 | 0 | 0 | 0 |
| **METABOLISM AND NUTRITION DISORDERS** | | | | | |
| ***APPETITE AND GENERAL NUTRITIONAL DISORDERS*** | | | | | |
| *APPETITE DISORDERS* | | | | | |
| DECREASED APPETITE | 0 | 0 | 0 | 0 | 0 |
| ***ELECTROLYTE AND FLUID BALANCE CONDITIONS*** | | | | | |
| *TOTAL FLUID VOLUME DECREASED* | | | | | |
| DEHYDRATION | 0 | 0 | 0 | 0 | 0 |

| | | | | | |
|---|---|---|---|---|---|
| **MUSCULOSKELETAL AND CONNECTIVE TISSUE DISORDERS** | | | | | |
| ***BONE DISORDERS (EXCL CONGENITAL AND FRACTURES)*** | | | | | |
| *BONE RELATED SIGNS AND SYMPTOMS* | | | | | |
| PAIN IN JAW | 0 | 0 | 0 | 0 | 0 |
| ***JOINT DISORDERS*** | | | | | |
| *JOINT RELATED SIGNS AND SYMPTOMS* | | | | | |
| ARTHRALGIA | 0 | 0 | 0 | 0 | 0 |
| ***MUSCLE DISORDERS*** | | | | | |
| *MUSCLE PAINS* | | | | | |
| MYALGIA | 0 | 0 | 0 | 0 | 0 |
| *MUSCLE RELATED SIGNS AND SYMPTOMS NEC* | | | | | |
| MUSCLE FATIGUE | 0 | 0 | 0 | 0 | 0 |
| MUSCLE SPASMS | 0 | 0 | 0 | 0 | 0 |
| *MUSCLE TONE ABNORMALITIES* | | | | | |
| TRISMUS | 0 | 0 | 0 | 0 | 0 |
| ***MUSCULOSKELETAL AND CONNECTIVE TISSUE DISORDERS NEC*** | | | | | |
| *MUSCULOSKELETAL AND CONNECTIVE TISSUE CONDITIONS NEC* | | | | | |
| MUSCULOSKELETAL STIFFNESS | 0 | 0 | 0 | 0 | 0 |
| BACK PAIN | 0 | 0 | 0 | 0 | 0 |
| LIMB DISCOMFORT | 0 | 0 | 0 | 0 | 0 |
| NECK PAIN | 0 | 0 | 0 | 0 | 0 |
| PAIN IN EXTREMITY | 0 | 0 | 0 | 0 | 0 |
| **NERVOUS SYSTEM DISORDERS** | | | | | |
| ***CRANIAL NERVE DISORDERS (EXCL NEOPLASMS)*** | | | | | |
| *FACIAL CRANIAL NERVE DISORDERS* | | | | | |
| BELL'S PALSY | 0 | 0 | 0 | 0 | 0 |
| *OLFACTORY NERVE DISORDERS* | | | | | |
| ANOSMIA | 0 | 0 | 0 | 0 | 0 |
| PAROSMIA | 0 | 0 | 0 | 0 | 0 |
| ***HEADACHES*** | | | | | |
| *HEADACHES NEC* | | | | | |
| CLUSTER HEADACHE | 0 | 0 | 0 | 0 | 0 |
| HEADACHE | 1 | 0 | 0 | 1 | 0 |
| SINUS HEADACHE | 0 | 0 | 0 | 0 | 0 |
| TENSION HEADACHE | 0 | 0 | 0 | 0 | 0 |
| *MIGRAINE HEADACHES* | | | | | |
| MIGRAINE WITH AURA | 0 | 0 | 0 | 0 | 0 |
| TYPICAL AURA WITHOUT HEADACHE | 0 | 0 | 0 | 0 | 0 |
| ***MENTAL IMPAIRMENT DISORDERS*** | | | | | |
| *MENTAL IMPAIRMENT (EXCL DEMENTIA AND MEMORY LOSS)* | | | | | |
| DISTURBANCE IN ATTENTION | 0 | 0 | 0 | 0 | 0 |
| ***MOVEMENT DISORDERS (INCL PARKINSONISM)*** | | | | | |
| *DYSKINESIAS AND MOVEMENT DISORDERS NEC* | | | | | |
| CLUMSINESS | 0 | 0 | 0 | 0 | 0 |
| *TREMOR (EXCL CONGENITAL)* | | | | | |
| TREMOR | 0 | 0 | 0 | 0 | 0 |
| ***NEUROLOGICAL DISORDERS NEC*** | | | | | |
| *DISTURBANCES IN CONSCIOUSNESS NEC* | | | | | |
| LETHARGY | 0 | 0 | 0 | 0 | 0 |
| SOMNOLENCE | 0 | 0 | 0 | 0 | 0 |
| SYNCOPE | 0 | 0 | 0 | 0 | 0 |
| *NEUROLOGICAL SIGNS AND SYMPTOMS NEC* | | | | | |
| AGITATION NEONATAL | 0 | 0 | 0 | 0 | 0 |

| | | | | | |
|---|---|---|---|---|---|
| BRAIN FOG | 0 | 0 | 0 | 0 | 0 |
| DIZZINESS | 0 | 0 | 0 | 0 | 0 |
| DIZZINESS EXERTIONAL | 0 | 0 | 0 | 0 | 0 |
| PRESYNCOPE | 0 | 0 | 0 | 0 | 0 |
| *PARAESTHESIAS AND DYSAESTHESIAS* | | | | | |
| PARAESTHESIA | 0 | 0 | 0 | 0 | 0 |
| *SENSORY ABNORMALITIES NEC* | | | | | |
| DYSGEUSIA | 0 | 0 | 0 | 0 | 0 |
| NEURALGIA | 0 | 0 | 0 | 0 | 0 |
| TASTE DISORDER | 0 | 0 | 0 | 0 | 0 |
| ***SEIZURES (INCL SUBTYPES)*** | | | | | |
| *SEIZURES AND SEIZURE DISORDERS NEC* | | | | | |
| SEIZURE | 0 | 0 | 0 | 0 | 0 |
| **PREGNANCY, PUERPERIUM AND PERINATAL CONDITIONS** | | | | | |
| ***ABORTIONS AND STILLBIRTH*** | | | | | |
| *ABORTIONS SPONTANEOUS* | | | | | |
| ABORTION SPONTANEOUS | 0 | 0 | 0 | 0 | 0 |
| *STILLBIRTH AND FOETAL DEATH* | | | | | |
| FOETAL DEATH | 0 | 0 | 0 | 0 | 0 |
| ***FOETAL COMPLICATIONS*** | | | | | |
| *FOETAL COMPLICATIONS NEC* | | | | | |
| FOETAL DISORDER | 1 | 1 | 0 | 0 | 0 |
| FOETAL HYPOKINESIA | 0 | 0 | 0 | 0 | 0 |
| *FOETAL GROWTH COMPLICATIONS* | | | | | |
| FOETAL MACROSOMIA | 0 | 0 | 0 | 0 | 0 |
| ***MATERNAL COMPLICATIONS OF PREGNANCY*** | | | | | |
| *MATERNAL COMPLICATIONS OF PREGNANCY NEC* | | | | | |
| MORNING SICKNESS | 0 | 0 | 0 | 0 | 0 |
| ***PLACENTAL, AMNIOTIC AND CAVITY DISORDERS (EXCL HAEMORRHAGES)*** | | | | | |
| *PLACENTAL ABNORMALITIES (EXCL NEOPLASMS)* | | | | | |
| PLACENTAL INFARCTION | 0 | 0 | 0 | 0 | 0 |
| ***PREGNANCY, LABOUR, DELIVERY AND POSTPARTUM CONDITIONS*** | | | | | |
| *NORMAL PREGNANCY, LABOUR AND DELIVERY* | | | | | |
| PREGNANCY | 0 | 0 | 0 | 0 | 0 |
| UTERINE CONTRACTIONS DURING PREGNANCY | 0 | 0 | 0 | 0 | 0 |
| **PSYCHIATRIC DISORDERS** | | | | | |
| ***ANXIETY DISORDERS AND SYMPTOMS*** | | | | | |
| *ANXIETY SYMPTOMS* | | | | | |
| AGITATION | 0 | 0 | 0 | 0 | 0 |
| ANXIETY | 0 | 0 | 0 | 0 | 0 |
| ***COGNITIVE AND ATTENTION DISORDERS AND DISTURBANCES*** | | | | | |
| *COGNITIVE AND ATTENTION DISORDERS AND DISTURBANCES NEC* | | | | | |
| MENTAL FATIGUE | 0 | 0 | 0 | 0 | 0 |
| ***DELIRIA (INCL CONFUSION)*** | | | | | |
| *CONFUSION AND DISORIENTATION* | | | | | |
| CONFUSIONAL STATE | 0 | 0 | 0 | 0 | 0 |
| ***DEPRESSED MOOD DISORDERS AND DISTURBANCES*** | | | | | |
| *MOOD ALTERATIONS WITH DEPRESSIVE SYMPTOMS* | | | | | |
| DEPRESSED MOOD | 0 | 0 | 0 | 0 | 0 |
| ***DISTURBANCES IN THINKING AND PERCEPTION*** | | | | | |
| *HALLUCINATIONS (EXCL SLEEP-RELATED)* | | | | | |
| HALLUCINATION | 0 | 0 | 0 | 0 | 0 |
| ***MOOD DISORDERS AND DISTURBANCES NEC*** | | | | | |

| | | | | | |
|---|---|---|---|---|---|
| *EMOTIONAL AND MOOD DISTURBANCES NEC* | | | | | |
| EMOTIONAL DISORDER | 0 | 0 | 0 | 0 | 0 |
| ***SLEEP DISORDERS AND DISTURBANCES*** | | | | | |
| *DISTURBANCES IN INITIATING AND MAINTAINING SLEEP* | | | | | |
| INITIAL INSOMNIA | 0 | 0 | 0 | 0 | 0 |
| INSOMNIA | 0 | 0 | 0 | 0 | 0 |
| **REPRODUCTIVE SYSTEM AND BREAST DISORDERS** | | | | | |
| ***MENSTRUAL CYCLE AND UTERINE BLEEDING DISORDERS*** | | | | | |
| *MENSTRUATION AND UTERINE BLEEDING NEC* | | | | | |
| MENSTRUAL DISORDER | 0 | 0 | 0 | 0 | 0 |
| *MENSTRUATION WITH DECREASED BLEEDING* | | | | | |
| MENSTRUATION DELAYED | 0 | 0 | 0 | 0 | 0 |
| OLIGOMENORRHOEA | 0 | 0 | 0 | 0 | 0 |
| *MENSTRUATION WITH INCREASED BLEEDING* | | | | | |
| HEAVY MENSTRUAL BLEEDING | 0 | 0 | 0 | 0 | 0 |
| ***VULVOVAGINAL DISORDERS (EXCL INFECTIONS AND INFLAMMATIONS)*** | | | | | |
| *VULVOVAGINAL DISORDERS NEC* | | | | | |
| VAGINAL HAEMORRHAGE | 0 | 0 | 0 | 0 | 0 |
| **RESPIRATORY, THORACIC AND MEDIASTINAL DISORDERS** | | | | | |
| ***RESPIRATORY DISORDERS NEC*** | | | | | |
| *BREATHING ABNORMALITIES* | | | | | |
| DYSPNOEA | 0 | 0 | 0 | 0 | 0 |
| *COUGHING AND ASSOCIATED SYMPTOMS* | | | | | |
| COUGH | 0 | 0 | 0 | 0 | 0 |
| ***RESPIRATORY TRACT SIGNS AND SYMPTOMS*** | | | | | |
| *UPPER RESPIRATORY TRACT SIGNS AND SYMPTOMS* | | | | | |
| DRY THROAT | 0 | 0 | 0 | 0 | 0 |
| OROPHARYNGEAL PAIN | 0 | 0 | 0 | 0 | 0 |
| PARANASAL SINUS DISCOMFORT | 0 | 0 | 0 | 0 | 0 |
| RHINORRHOEA | 0 | 0 | 0 | 0 | 0 |
| SINUS PAIN | 0 | 0 | 0 | 0 | 0 |
| SNEEZING | 0 | 0 | 0 | 0 | 0 |
| ***UPPER RESPIRATORY TRACT DISORDERS (EXCL INFECTIONS)*** | | | | | |
| *NASAL CONGESTION AND INFLAMMATIONS* | | | | | |
| NASAL CONGESTION | 0 | 0 | 0 | 0 | 0 |
| **SKIN AND SUBCUTANEOUS TISSUE DISORDERS** | | | | | |
| ***ANGIOEDEMA AND URTICARIA*** | | | | | |
| *URTICARIAS* | | | | | |
| URTICARIA | 0 | 0 | 0 | 0 | 0 |
| ***CORNIFICATION AND DYSTROPHIC SKIN DISORDERS*** | | | | | |
| *SKIN DYSTROPHIES* | | | | | |
| HYPERTROPHIC SCAR | 0 | 0 | 0 | 0 | 0 |
| ***EPIDERMAL AND DERMAL CONDITIONS*** | | | | | |
| *DERMAL AND EPIDERMAL CONDITIONS NEC* | | | | | |
| SKIN WARM | 0 | 0 | 0 | 0 | 0 |
| *DERMATITIS AND ECZEMA* | | | | | |
| DERMATITIS ALLERGIC | 0 | 0 | 0 | 0 | 0 |
| *ERYTHEMAS* | | | | | |
| ERYTHEMA | 0 | 0 | 0 | 0 | 0 |
| *PRURITUS NEC* | | | | | |
| PRURITUS | 0 | 0 | 0 | 0 | 0 |
| *RASHES, ERUPTIONS AND EXANTHEMS NEC* | | | | | |
| RASH | 0 | 0 | 0 | 0 | 0 |

| | | | | | |
|---|---|---|---|---|---|
| RASH PRURITIC | 0 | 0 | 0 | 0 | 0 |
| ***SKIN APPENDAGE CONDITIONS*** | | | | | |
| *APOCRINE AND ECCRINE GLAND DISORDERS* | | | | | |
| COLD SWEAT | 0 | 0 | 0 | 0 | 0 |
| HYPERHIDROSIS | 0 | 0 | 0 | 0 | 0 |
| NIGHT SWEATS | 0 | 0 | 0 | 0 | 0 |
| **VASCULAR DISORDERS** | | | | | |
| ***DECREASED AND NONSPECIFIC BLOOD PRESSURE DISORDERS AND SHOCK*** | | | | | |
| *BLOOD PRESSURE DISORDERS NEC* | | | | | |
| BLOOD PRESSURE FLUCTUATION | 0 | 0 | 0 | 0 | 0 |
| *VASCULAR HYPOTENSIVE DISORDERS* | | | | | |
| HYPOTENSION | 0 | 0 | 0 | 0 | 0 |
| ***EMBOLISM AND THROMBOSIS*** | | | | | |
| *NON-SITE SPECIFIC EMBOLISM AND THROMBOSIS* | | | | | |
| EMBOLISM | 0 | 0 | 0 | 0 | 0 |
| ***VASCULAR DISORDERS NEC*** | | | | | |
| *NON-SITE SPECIFIC VASCULAR DISORDERS NEC* | | | | | |
| VASCULAR PAIN | 0 | 0 | 0 | 0 | 0 |
| *PERIPHERAL VASCULAR DISORDERS NEC* | | | | | |
| HOT FLUSH | 0 | 0 | 0 | 0 | 0 |
| ***VASCULAR HAEMORRHAGIC DISORDERS*** | | | | | |
| *HAEMORRHAGES NEC* | | | | | |
| HAEMORRHAGE | 0 | 0 | 0 | 0 | 0 |
| **TOTAL ADR EVENTS** | ***5*** | ***2*** | ***0*** | ***3*** | ***0*** |

**TABLE S2-6. COVID-19 vaccine ADR listings for events reported by pregnant patients with no dose identification information in those reporting any vaccination dose**

| | Patients with any vaccination dose: ADR Counts |
|---|---|
| **REACTION TERM (SOC, *HLGT*, *HLT*, PT)** | Unknown dose |
| ***(freetext)*** | 0 |
| **BLOOD AND LYMPHATIC SYSTEM DISORDERS** | |
| ***SPLEEN, LYMPHATIC AND RETICULOENDOTHELIAL SYSTEM DISORDERS*** | |
| *LYMPHATIC SYSTEM DISORDERS NEC* | |
| LYMPH NODE PAIN | 1 |
| LYMPHADENITIS | 0 |
| LYMPHADENOPATHY | 0 |
| **CARDIAC DISORDERS** | |
| ***CARDIAC ARRHYTHMIAS*** | |
| *RATE AND RHYTHM DISORDERS NEC* | |
| TACHYCARDIA | 0 |
| ***CARDIAC DISORDERS, SIGNS AND SYMPTOMS NEC*** | |
| *CARDIAC SIGNS AND SYMPTOMS NEC* | |
| PALPITATIONS | 0 |
| **EAR AND LABYRINTH DISORDERS** | |
| ***AURAL DISORDERS NEC*** | |
| *EAR DISORDERS NEC* | |
| EAR PAIN | 0 |
| ***INNER EAR AND VIIITH CRANIAL NERVE DISORDERS*** | |
| *INNER EAR SIGNS AND SYMPTOMS* | |
| TINNITUS | 0 |
| VERTIGO | 0 |
| **EYE DISORDERS** | |
| ***EYE DISORDERS NEC*** | |
| *OCULAR DISORDERS NEC* | |
| EYE PAIN | 0 |
| ***OCULAR SENSORY SYMPTOMS NEC*** | |
| *OCULAR SENSATION DISORDERS* | |
| ASTHENOPIA | 0 |
| PHOTOPHOBIA | 0 |
| ***VISION DISORDERS*** | |
| *VISUAL DISORDERS NEC* | |
| VISION BLURRED | 0 |
| **GASTROINTESTINAL DISORDERS** | |
| ***DENTAL AND GINGIVAL CONDITIONS*** | |
| *DENTAL DISORDERS NEC* | |
| TEETHING | 0 |
| ***GASTROINTESTINAL INFLAMMATORY CONDITIONS*** | |
| *GASTROINTESTINAL INFLAMMATORY DISORDERS NEC* | |
| GASTROINTESTINAL TRACT IRRITATION | 0 |
| ***GASTROINTESTINAL MOTILITY AND DEFAECATION CONDITIONS*** | |
| *DIARRHOEA (EXCL INFECTIVE)* | |
| DIARRHOEA | 0 |
| *GASTROINTESTINAL ATONIC AND HYPOMOTILITY DISORDERS NEC* | |
| CONSTIPATION | 0 |

| | |
|---|---|
| ***GASTROINTESTINAL SIGNS AND SYMPTOMS*** | |
| *GASTROINTESTINAL AND ABDOMINAL PAINS (EXCL ORAL AND THROAT)* | |
| ABDOMINAL PAIN | 0 |
| ABDOMINAL PAIN UPPER | 0 |
| *GASTROINTESTINAL SIGNS AND SYMPTOMS NEC* | |
| ABDOMINAL DISCOMFORT | 0 |
| *NAUSEA AND VOMITING SYMPTOMS* | |
| NAUSEA | 3 |
| VOMITING | 1 |
| ***ORAL SOFT TISSUE CONDITIONS*** | |
| *ORAL SOFT TISSUE SIGNS AND SYMPTOMS* | |
| HYPOAESTHESIA ORAL | 0 |
| PARAESTHESIA ORAL | 0 |
| ***TONGUE CONDITIONS*** | |
| *TONGUE SIGNS AND SYMPTOMS* | |
| GLOSSODYNIA | 0 |
| **GENERAL DISORDERS AND ADMINISTRATION SITE CONDITIONS** | |
| ***ADMINISTRATION SITE REACTIONS*** | |
| *ADMINISTRATION SITE REACTIONS NEC* | |
| PUNCTURE SITE BRUISE | 1 |
| PUNCTURE SITE PAIN | 0 |
| *INJECTION SITE REACTIONS* | |
| INJECTION SITE BRUISING | 0 |
| INJECTION SITE ERYTHEMA | 0 |
| INJECTION SITE MASS | 1 |
| INJECTION SITE PAIN | 1 |
| INJECTION SITE REACTION | 0 |
| INJECTION SITE URTICARIA | 0 |
| INJECTION SITE WARMTH | 0 |
| *VACCINATION SITE REACTIONS* | |
| VACCINATION SITE PAIN | 1 |
| ***BODY TEMPERATURE CONDITIONS*** | |
| *FEBRILE DISORDERS* | |
| PYREXIA | 6 |
| ***GENERAL SYSTEM DISORDERS NEC*** | |
| *ASTHENIC CONDITIONS* | |
| ASTHENIA | 1 |
| FATIGUE | 17 |
| MALAISE | 0 |
| *FEELINGS AND SENSATIONS NEC* | |
| CHILLS | 1 |
| FEELING ABNORMAL | 0 |
| FEELING COLD | 0 |
| FEELING HOT | 0 |
| FEELING OF BODY TEMPERATURE CHANGE | 0 |
| HANGOVER | 0 |
| HUNGER | 0 |
| THIRST | 0 |
| *GENERAL SIGNS AND SYMPTOMS NEC* | |
| CRYING | 0 |
| ILLNESS | 0 |
| INFLUENZA LIKE ILLNESS | 1 |
| LOCAL REACTION | 0 |
| PERIPHERAL SWELLING | 0 |
| SWELLING | 0 |
| *PAIN AND DISCOMFORT NEC* | |

| | |
|---|---|
| AXILLARY PAIN | 0 |
| CHEST PAIN | 0 |
| PAIN | 5 |
| TENDERNESS | 0 |
| **INFECTIONS AND INFESTATIONS** | |
| ***BACTERIAL INFECTIOUS DISORDERS*** | |
| *BACTERIAL INFECTIONS NEC* | |
| PERIORBITAL CELLULITIS | 0 |
| ***INFECTIONS - PATHOGEN UNSPECIFIED*** | |
| *BREAST INFECTIONS* | |
| MASTITIS | 0 |
| *UPPER RESPIRATORY TRACT INFECTIONS* | |
| LARYNGITIS | 0 |
| NASOPHARYNGITIS | 0 |
| ***VIRAL INFECTIOUS DISORDERS*** | |
| *HERPES VIRAL INFECTIONS* | |
| GENITAL HERPES | 0 |
| HERPES ZOSTER | 0 |
| ORAL HERPES | 0 |
| *INFLUENZA VIRAL INFECTIONS* | |
| INFLUENZA | 0 |
| *VIRAL INFECTIONS NEC* | |
| VIRAL DIARRHOEA | 0 |
| **INJURY, POISONING AND PROCEDURAL COMPLICATIONS** | |
| ***INJURIES NEC*** | |
| *SKIN INJURIES NEC* | |
| CONTUSION | 0 |
| ***PROCEDURAL RELATED INJURIES AND COMPLICATIONS NEC*** | |
| *NON-SITE SPECIFIC PROCEDURAL COMPLICATIONS* | |
| INJECTION RELATED REACTION | 0 |
| **INVESTIGATIONS** | |
| ***CARDIAC AND VASCULAR INVESTIGATIONS (EXCL ENZYME TESTS)*** | |
| *HEART RATE AND PULSE INVESTIGATIONS* | |
| HEART RATE | 0 |
| HEART RATE INCREASED | 0 |
| ***PHYSICAL EXAMINATION AND ORGAN SYSTEM STATUS TOPICS*** | |
| *PHYSICAL EXAMINATION PROCEDURES AND ORGAN SYSTEM STATUS* | |
| BODY TEMPERATURE INCREASED | 0 |
| ***WATER, ELECTROLYTE AND MINERAL INVESTIGATIONS*** | |
| *WATER AND ELECTROLYTE ANALYSES NEC* | |
| VOLUME BLOOD | 0 |
| **METABOLISM AND NUTRITION DISORDERS** | |
| ***APPETITE AND GENERAL NUTRITIONAL DISORDERS*** | |
| *APPETITE DISORDERS* | |
| DECREASED APPETITE | 0 |
| ***ELECTROLYTE AND FLUID BALANCE CONDITIONS*** | |
| *TOTAL FLUID VOLUME DECREASED* | |
| DEHYDRATION | 0 |
| **MUSCULOSKELETAL AND CONNECTIVE TISSUE DISORDERS** | |
| ***BONE DISORDERS (EXCL CONGENITAL AND FRACTURES)*** | |
| *BONE RELATED SIGNS AND SYMPTOMS* | |
| PAIN IN JAW | 0 |
| ***JOINT DISORDERS*** | |
| *JOINT RELATED SIGNS AND SYMPTOMS* | |
| ARTHRALGIA | 1 |
| ***MUSCLE DISORDERS*** | |

| | |
|---|---|
| *MUSCLE PAINS* | |
| MYALGIA | 2 |
| *MUSCLE RELATED SIGNS AND SYMPTOMS NEC* | |
| MUSCLE FATIGUE | 0 |
| MUSCLE SPASMS | 0 |
| *MUSCLE TONE ABNORMALITIES* | |
| TRISMUS | 0 |
| ***MUSCULOSKELETAL AND CONNECTIVE TISSUE DISORDERS NEC*** | |
| *MUSCULOSKELETAL AND CONNECTIVE TISSUE CONDITIONS NEC* | |
| MUSCULOSKELETAL STIFFNESS | 0 |
| BACK PAIN | 0 |
| LIMB DISCOMFORT | 0 |
| NECK PAIN | 0 |
| PAIN IN EXTREMITY | 10 |
| **NERVOUS SYSTEM DISORDERS** | |
| ***CRANIAL NERVE DISORDERS (EXCL NEOPLASMS)*** | |
| *FACIAL CRANIAL NERVE DISORDERS* | |
| BELL'S PALSY | 0 |
| *OLFACTORY NERVE DISORDERS* | |
| ANOSMIA | 0 |
| PAROSMIA | 0 |
| ***HEADACHES*** | |
| *HEADACHES NEC* | |
| CLUSTER HEADACHE | 0 |
| HEADACHE | 14 |
| SINUS HEADACHE | 0 |
| TENSION HEADACHE | 0 |
| *MIGRAINE HEADACHES* | |
| MIGRAINE WITH AURA | 0 |
| TYPICAL AURA WITHOUT HEADACHE | 0 |
| ***MENTAL IMPAIRMENT DISORDERS*** | |
| *MENTAL IMPAIRMENT (EXCL DEMENTIA AND MEMORY LOSS)* | |
| DISTURBANCE IN ATTENTION | 0 |
| ***MOVEMENT DISORDERS (INCL PARKINSONISM)*** | |
| *DYSKINESIAS AND MOVEMENT DISORDERS NEC* | |
| CLUMSINESS | 0 |
| *TREMOR (EXCL CONGENITAL)* | |
| TREMOR | 0 |
| ***NEUROLOGICAL DISORDERS NEC*** | |
| *DISTURBANCES IN CONSCIOUSNESS NEC* | |
| LETHARGY | 0 |
| SOMNOLENCE | 2 |
| SYNCOPE | 0 |
| *NEUROLOGICAL SIGNS AND SYMPTOMS NEC* | |
| AGITATION NEONATAL | 1 |
| BRAIN FOG | 0 |
| DIZZINESS | 1 |
| DIZZINESS EXERTIONAL | 0 |
| PRESYNCOPE | 0 |
| *PARAESTHESIAS AND DYSAESTHESIAS* | |
| PARAESTHESIA | 0 |
| *SENSORY ABNORMALITIES NEC* | |
| DYSGEUSIA | 0 |

| | |
|---|---|
| NEURALGIA | 0 |
| TASTE DISORDER | 0 |
| ***SEIZURES (INCL SUBTYPES)*** | |
| *SEIZURES AND SEIZURE DISORDERS NEC* | |
| SEIZURE | 0 |
| **PREGNANCY, PUERPERIUM AND PERINATAL CONDITIONS** | |
| ***ABORTIONS AND STILLBIRTH*** | |
| *ABORTIONS SPONTANEOUS* | |
| ABORTION SPONTANEOUS | 0 |
| *STILLBIRTH AND FOETAL DEATH* | |
| FOETAL DEATH | 0 |
| ***FOETAL COMPLICATIONS*** | |
| *FOETAL COMPLICATIONS NEC* | |
| FOETAL DISORDER | 0 |
| FOETAL HYPOKINESIA | 0 |
| *FOETAL GROWTH COMPLICATIONS* | |
| FOETAL MACROSOMIA | 0 |
| ***MATERNAL COMPLICATIONS OF PREGNANCY*** | |
| *MATERNAL COMPLICATIONS OF PREGNANCY NEC* | |
| MORNING SICKNESS | 1 |
| ***PLACENTAL, AMNIOTIC AND CAVITY DISORDERS (EXCL HAEMORRHAGES)*** | |
| *PLACENTAL ABNORMALITIES (EXCL NEOPLASMS)* | |
| PLACENTAL INFARCTION | 0 |
| ***PREGNANCY, LABOUR, DELIVERY AND POSTPARTUM CONDITIONS*** | |
| *NORMAL PREGNANCY, LABOUR AND DELIVERY* | |
| PREGNANCY | 1 |
| UTERINE CONTRACTIONS DURING PREGNANCY | 0 |
| **PSYCHIATRIC DISORDERS** | |
| ***ANXIETY DISORDERS AND SYMPTOMS*** | |
| *ANXIETY SYMPTOMS* | |
| AGITATION | 0 |
| ANXIETY | 2 |
| ***COGNITIVE AND ATTENTION DISORDERS AND DISTURBANCES*** | |
| *COGNITIVE AND ATTENTION DISORDERS AND DISTURBANCES NEC* | |
| MENTAL FATIGUE | 0 |
| ***DELIRIA (INCL CONFUSION)*** | |
| *CONFUSION AND DISORIENTATION* | |
| CONFUSIONAL STATE | 0 |
| ***DEPRESSED MOOD DISORDERS AND DISTURBANCES*** | |
| *MOOD ALTERATIONS WITH DEPRESSIVE SYMPTOMS* | |
| DEPRESSED MOOD | 0 |
| ***DISTURBANCES IN THINKING AND PERCEPTION*** | |
| *HALLUCINATIONS (EXCL SLEEP-RELATED)* | |
| HALLUCINATION | 0 |
| ***MOOD DISORDERS AND DISTURBANCES NEC*** | |
| *EMOTIONAL AND MOOD DISTURBANCES NEC* | |
| EMOTIONAL DISORDER | 0 |
| ***SLEEP DISORDERS AND DISTURBANCES*** | |
| *DISTURBANCES IN INITIATING AND MAINTAINING SLEEP* | |
| INITIAL INSOMNIA | 0 |
| INSOMNIA | 0 |
| **REPRODUCTIVE SYSTEM AND BREAST DISORDERS** | |
| ***MENSTRUAL CYCLE AND UTERINE BLEEDING DISORDERS*** | |
| *MENSTRUATION AND UTERINE BLEEDING NEC* | |

| | |
|---|---|
| MENSTRUAL DISORDER | 0 |
| *MENSTRUATION WITH DECREASED BLEEDING* | |
| MENSTRUATION DELAYED | 1 |
| OLIGOMENORRHOEA | 1 |
| *MENSTRUATION WITH INCREASED BLEEDING* | |
| HEAVY MENSTRUAL BLEEDING | 1 |
| ***VULVOVAGINAL DISORDERS (EXCL INFECTIONS AND INFLAMMATIONS)*** | |
| *VULVOVAGINAL DISORDERS NEC* | |
| VAGINAL HAEMORRHAGE | 0 |
| **RESPIRATORY, THORACIC AND MEDIASTINAL DISORDERS** | |
| ***RESPIRATORY DISORDERS NEC*** | |
| *BREATHING ABNORMALITIES* | |
| DYSPNOEA | 1 |
| *COUGHING AND ASSOCIATED SYMPTOMS* | |
| COUGH | 0 |
| ***RESPIRATORY TRACT SIGNS AND SYMPTOMS*** | |
| *UPPER RESPIRATORY TRACT SIGNS AND SYMPTOMS* | |
| DRY THROAT | 0 |
| OROPHARYNGEAL PAIN | 1 |
| PARANASAL SINUS DISCOMFORT | 0 |
| RHINORRHOEA | 0 |
| SINUS PAIN | 0 |
| SNEEZING | 0 |
| ***UPPER RESPIRATORY TRACT DISORDERS (EXCL INFECTIONS)*** | |
| *NASAL CONGESTION AND INFLAMMATIONS* | |
| NASAL CONGESTION | 0 |
| **SKIN AND SUBCUTANEOUS TISSUE DISORDERS** | |
| ***ANGIOEDEMA AND URTICARIA*** | |
| *URTICARIAS* | |
| URTICARIA | 0 |
| ***CORNIFICATION AND DYSTROPHIC SKIN DISORDERS*** | |
| *SKIN DYSTROPHIES* | |
| HYPERTROPHIC SCAR | 0 |
| ***EPIDERMAL AND DERMAL CONDITIONS*** | |
| *DERMAL AND EPIDERMAL CONDITIONS NEC* | |
| SKIN WARM | 0 |
| *DERMATITIS AND ECZEMA* | |
| DERMATITIS ALLERGIC | 0 |
| *ERYTHEMAS* | |
| ERYTHEMA | 1 |
| *PRURITUS NEC* | |
| PRURITUS | 1 |
| *RASHES, ERUPTIONS AND EXANTHEMS NEC* | |
| RASH | 0 |
| RASH PRURITIC | 0 |
| ***SKIN APPENDAGE CONDITIONS*** | |
| *APOCRINE AND ECCRINE GLAND DISORDERS* | |
| COLD SWEAT | |
| HYPERHIDROSIS | 0 |
| NIGHT SWEATS | 0 |
| **VASCULAR DISORDERS** | |
| ***DECREASED AND NONSPECIFIC BLOOD PRESSURE DISORDERS AND SHOCK*** | |
| *BLOOD PRESSURE DISORDERS NEC* | |

| | |
|---|---|
| BLOOD PRESSURE FLUCTUATION | 0 |
| *VASCULAR HYPOTENSIVE DISORDERS* | |
| HYPOTENSION | 0 |
| ***EMBOLISM AND THROMBOSIS*** | |
| *NON-SITE SPECIFIC EMBOLISM AND THROMBOSIS* | |
| EMBOLISM | 0 |
| ***VASCULAR DISORDERS NEC*** | |
| *NON-SITE SPECIFIC VASCULAR DISORDERS NEC* | |
| VASCULAR PAIN | 0 |
| *PERIPHERAL VASCULAR DISORDERS NEC* | |
| HOT FLUSH | 0 |
| ***VASCULAR HAEMORRHAGIC DISORDERS*** | |
| *HAEMORRHAGES NEC* | |
| HAEMORRHAGE | 0 |
| **TOTAL ADR EVENTS** | ***82*** |

**TABLE S2-7. ADR listing for events reported by breast-feeding patients, in those reporting any vaccination dose**

| REACTION TERM (SOC, *HLGT*, *HLT*, PT) | Patients with any vaccination dose: ADR Counts | | | | | |
|---|---|---|---|---|---|---|
| | All doses | 1st dose | 2nd dose | 3rd dose | Other doses | Unknown |
| ***(freetext)*** | 1 | 1 | 0 | 0 | 0 | 0 |
| **BLOOD AND LYMPHATIC SYSTEM DISORDERS** | | | | | | |
| *LYMPHATIC SYSTEM DISORDERS NEC* | | | | | | |
| LYMPH NODE PAIN | 2 | 0 | 1 | 1 | 0 | 0 |
| LYMPHADENOPATHY | 5 | 1 | 3 | 0 | 0 | 1 |
| **CARDIAC DISORDERS** | | | | | | |
| ***CARDIAC DISORDERS, SIGNS AND SYMPTOMS NEC*** | | | | | | |
| *CARDIAC SIGNS AND SYMPTOMS NEC* | | | | | | |
| PALPITATIONS | 6 | 2 | 2 | 0 | 0 | 2 |
| **EAR AND LABYRINTH DISORDERS** | | | | | | |
| ***AURAL DISORDERS NEC*** | | | | | | |
| *EAR DISORDERS NEC* | | | | | | |
| EAR PAIN | 1 | 1 | 0 | 0 | 0 | 0 |
| ***INNER EAR AND VIIITH CRANIAL NERVE DISORDERS*** | | | | | | |
| *INNER EAR SIGNS AND SYMPTOMS* | | | | | | |
| TINNITUS | 2 | 0 | 1 | 1 | 0 | 0 |
| **EYE DISORDERS** | | | | | | |
| ***EYE DISORDERS NEC*** | | | | | | |
| *LACRIMATION DISORDERS* | | | | | | |
| DRY EYE | 1 | 1 | 0 | 0 | 0 | 0 |
| *OCULAR DISORDERS NEC* | | | | | | |
| EYE PAIN | 1 | 1 | 0 | 0 | 0 | 0 |
| ***OCULAR INFECTIONS, IRRITATIONS AND INFLAMMATIONS*** | | | | | | |
| *OCULAR INFECTIONS, INFLAMMATIONS AND ASSOCIATED MANIFESTATIONS* | | | | | | |
| EYE IRRITATION | 1 | 1 | 0 | 0 | 0 | 0 |
| ***OCULAR SENSORY SYMPTOMS NEC*** | | | | | | |
| *OCULAR SENSATION DISORDERS* | | | | | | |
| PHOTOPHOBIA | 2 | 2 | 0 | 0 | 0 | 0 |
| ***VISION DISORDERS*** | | | | | | |
| *VISUAL DISORDERS NEC* | | | | | | |
| VISION BLURRED | 2 | 2 | 0 | 0 | 0 | 0 |
| **GASTROINTESTINAL DISORDERS** | | | | | | |
| ***GASTROINTESTINAL INFLAMMATORY CONDITIONS*** | | | | | | |
| *GASTROINTESTINAL INFLAMMATORY DISORDERS NEC* | | | | | | |
| GASTROINTESTINAL TRACT IRRITATION | 1 | 0 | 0 | 1 | 0 | 0 |
| ***GASTROINTESTINAL MOTILITY AND DEFAECATION CONDITIONS*** | | | | | | |
| *DIARRHOEA (EXCL INFECTIVE)* | | | | | | |
| DIARRHOEA | 11 | 7 | 2 | 2 | 0 | 0 |
| ***GASTROINTESTINAL SIGNS AND SYMPTOMS*** | | | | | | |
| *FLATULENCE, BLOATING AND DISTENSION* | | | | | | |
| ABDOMINAL DISTENSION | 1 | 1 | 0 | 0 | 0 | 0 |
| FLATULENCE | 1 | 1 | 0 | 0 | 0 | 0 |
| *GASTROINTESTINAL AND ABDOMINAL PAINS (EXCL ORAL AND THROAT)* | | | | | | |
| ABDOMINAL PAIN | 5 | 5 | 0 | 0 | 0 | 0 |
| ABDOMINAL PAIN UPPER | 3 | 2 | 1 | 0 | 0 | 0 |

| | | | | | | |
|---|---|---|---|---|---|---|
| *NAUSEA AND VOMITING SYMPTOMS* | | | | | | |
| NAUSEA | 39 | 22 | 8 | 5 | 0 | 4 |
| RETCHING | 1 | 1 | 0 | 0 | 0 | 0 |
| VOMITING | 6 | 4 | 2 | 0 | 0 | 0 |
| ***ORAL SOFT TISSUE CONDITIONS*** | | | | | | |
| *ORAL SOFT TISSUE SIGNS AND SYMPTOMS* | | | | | | |
| PARAESTHESIA ORAL | 1 | 1 | 0 | 0 | 0 | 0 |
| LIP SWELLING | 1 | 0 | 1 | 0 | 0 | 0 |
| **GENERAL DISORDERS AND ADMINISTRATION SITE CONDITIONS** | | | | | | |
| ***ADMINISTRATION SITE REACTIONS*** | | | | | | |
| *ADMINISTRATION SITE REACTIONS NEC* | | | | | | |
| PUNCTURE SITE BRUISE | 1 | 0 | 0 | 1 | 0 | 0 |
| *INJECTION SITE REACTIONS* | | | | | | |
| INJECTION SITE ERYTHEMA | 1 | 1 | 0 | 0 | 0 | 0 |
| INJECTION SITE MASS | 1 | 0 | 0 | 0 | 0 | 1 |
| INJECTION SITE PAIN | 23 | 16 | 3 | 1 | 0 | 3 |
| INJECTION SITE WARMTH | 1 | 1 | 0 | 0 | 0 | 0 |
| *VACCINATION SITE REACTIONS* | | | | | | |
| VACCINATION SITE PAIN | 1 | 0 | 1 | 0 | 0 | 0 |
| ***BODY TEMPERATURE CONDITIONS*** | | | | | | |
| *FEBRILE DISORDERS* | | | | | | |
| PYREXIA | 70 | 44 | 16 | 5 | 0 | 5 |
| ***GENERAL SYSTEM DISORDERS NEC*** | | | | | | |
| *ASTHENIC CONDITIONS* | | | | | | |
| FATIGUE | 113 | 60 | 35 | 10 | 0 | 8 |
| MALAISE | 6 | 3 | 3 | 0 | 0 | 0 |
| *FEELINGS AND SENSATIONS NEC* | | | | | | |
| CHILLS | 42 | 27 | 6 | 6 | 0 | 3 |
| FEELING ABNORMAL | 1 | 0 | 0 | 1 | 0 | 0 |
| FEELING COLD | 9 | 7 | 2 | 0 | 0 | 0 |
| FEELING HOT | 1 | 1 | 0 | 0 | 0 | 0 |
| HANGOVER | 1 | 0 | 0 | 1 | 0 | 0 |
| *GENERAL SIGNS AND SYMPTOMS NEC* | | | | | | |
| INFLUENZA LIKE ILLNESS | 8 | 4 | 1 | 2 | 0 | 1 |
| LOCAL REACTION | 1 | 0 | 1 | 0 | 0 | 0 |
| PERIPHERAL SWELLING | 4 | 3 | 1 | 0 | 0 | 0 |
| SWELLING | 1 | 1 | 0 | 0 | 0 | 0 |
| *PAIN AND DISCOMFORT NEC* | | | | | | |
| AXILLARY PAIN | 4 | 1 | 3 | 0 | 0 | 0 |
| CHEST DISCOMFORT | 1 | 1 | 0 | 0 | 0 | 0 |
| PAIN | 29 | 16 | 6 | 5 | 0 | 2 |
| TENDERNESS | 4 | 2 | 2 | 0 | 0 | 0 |
| **INFECTIONS AND INFESTATIONS** | | | | | | |
| ***INFECTIONS - PATHOGEN UNSPECIFIED*** | | | | | | |
| *BREAST INFECTIONS* | | | | | | |
| MASTITIS | 1 | 0 | 0 | 1 | 0 | 0 |
| *SKIN STRUCTURES AND SOFT TISSUE INFECTIONS* | | | | | | |
| INJECTION SITE PUSTULE | 1 | 0 | 1 | 0 | 0 | 0 |
| *UPPER RESPIRATORY TRACT INFECTIONS* | | | | | | |
| LARYNGITIS | 1 | 0 | 1 | 0 | 0 | 0 |
| NASOPHARYNGITIS | 3 | 2 | 1 | 0 | 0 | 0 |
| ***VIRAL INFECTIOUS DISORDERS*** | | | | | | |
| *HERPES VIRAL INFECTIONS* | | | | | | |

| | | | | | | |
|---|---|---|---|---|---|---|
| ORAL HERPES | 1 | 0 | 1 | 0 | 0 | 0 |
| *INFLUENZA VIRAL INFECTIONS* | | | | | | |
| INFLUENZA | 11 | 6 | 4 | 1 | 0 | 0 |
| **INJURY, POISONING AND PROCEDURAL COMPLICATIONS** | | | | | | |
| ***INJURIES NEC*** | | | | | | |
| *SKIN INJURIES NEC* | | | | | | |
| CONTUSION | 2 | 2 | 0 | 0 | 0 | 0 |
| **INVESTIGATIONS** | | | | | | |
| ***CARDIAC AND VASCULAR INVESTIGATIONS (EXCL ENZYME TESTS)*** | | | | | | |
| *HEART RATE AND PULSE INVESTIGATIONS* | | | | | | |
| HEART RATE | 1 | 1 | 0 | 0 | 0 | 0 |
| ***PHYSICAL EXAMINATION AND ORGAN SYSTEM STATUS TOPICS*** | | | | | | |
| *PHYSICAL EXAMINATION PROCEDURES AND ORGAN SYSTEM STATUS* | | | | | | |
| BODY TEMPERATURE | 3 | 3 | 0 | 0 | 0 | 0 |
| BODY TEMPERATURE INCREASED | 2 | 1 | 0 | 1 | 0 | 0 |
| ***WATER, ELECTROLYTE AND MINERAL INVESTIGATIONS*** | | | | | | |
| *WATER AND ELECTROLYTE ANALYSES NEC* | | | | | | |
| VOLUME BLOOD | 1 | 0 | 1 | 0 | 0 | 0 |
| **METABOLISM AND NUTRITION DISORDERS** | | | | | | |
| ***APPETITE AND GENERAL NUTRITIONAL DISORDERS*** | | | | | | |
| *APPETITE DISORDERS* | | | | | | |
| DECREASED APPETITE | 4 | 3 | 1 | 0 | 0 | 0 |
| **MUSCULOSKELETAL AND CONNECTIVE TISSUE DISORDERS** | | | | | | |
| ***BONE DISORDERS (EXCL CONGENITAL AND FRACTURES)*** | | | | | | |
| *BONE RELATED SIGNS AND SYMPTOMS* | | | | | | |
| BONE PAIN | 1 | 1 | 0 | 0 | 0 | 0 |
| PAIN IN JAW | 1 | 0 | 1 | 0 | 0 | 0 |
| ***JOINT DISORDERS*** | | | | | | |
| *JOINT RELATED SIGNS AND SYMPTOMS* | | | | | | |
| ARTHRALGIA | 17 | 9 | 5 | 1 | 0 | 2 |
| JOINT STIFFNESS | 1 | 1 | 0 | 0 | 0 | 0 |
| ***MUSCLE DISORDERS*** | | | | | | |
| *MUSCLE PAINS* | | | | | | |
| MYALGIA | 51 | 35 | 9 | 5 | 0 | 2 |
| *MUSCLE RELATED SIGNS AND SYMPTOMS NEC* | | | | | | |
| MUSCLE FATIGUE | 1 | 0 | 0 | 1 | 0 | 0 |
| MUSCLE SPASMS | 1 | 1 | 0 | 0 | 0 | 0 |
| ***MUSCULOSKELETAL AND CONNECTIVE TISSUE DISORDERS NEC*** | | | | | | |
| *MUSCULOSKELETAL AND CONNECTIVE TISSUE CONDITIONS NEC* | | | | | | |
| MUSCULOSKELETAL STIFFNESS | 2 | 0 | 0 | 2 | 0 | 0 |
| BACK PAIN | 3 | 2 | 0 | 1 | 0 | 0 |
| LIMB DISCOMFORT | 25 | 19 | 6 | 0 | 0 | 0 |
| PAIN IN EXTREMITY | 134 | 100 | 23 | 8 | 0 | 3 |
| **NERVOUS SYSTEM DISORDERS** | | | | | | |
| ***CRANIAL NERVE DISORDERS (EXCL NEOPLASMS)*** | | | | | | |
| *OLFACTORY NERVE DISORDERS* | | | | | | |
| ANOSMIA | 1 | 1 | 0 | 0 | 0 | 0 |
| PAROSMIA | 1 | 0 | 1 | 0 | 0 | 0 |
| ***HEADACHES*** | | | | | | |
| *HEADACHES NEC* | | | | | | |
| HEADACHE | 101 | 64 | 16 | 13 | 0 | 8 |
| SINUS HEADACHE | 3 | 1 | 2 | 0 | 0 | 0 |
| TENSION HEADACHE | 5 | 5 | 0 | 0 | 0 | 0 |

| | | | | | | |
|---|---|---|---|---|---|---|
| *MIGRAINE HEADACHES* | | | | | | |
| MIGRAINE | 2 | 1 | 0 | 0 | 0 | 1 |
| TYPICAL AURA WITHOUT HEADACHE | 1 | 0 | 1 | 0 | 0 | 0 |
| ***MOVEMENT DISORDERS (INCL PARKINSONISM)*** | | | | | | |
| *TREMOR (EXCL CONGENITAL)* | | | | | | |
| TREMOR | 2 | 1 | 0 | 0 | 0 | 1 |
| ***NEUROLOGICAL DISORDERS NEC*** | | | | | | |
| *DISTURBANCES IN CONSCIOUSNESS NEC* | | | | | | |
| LETHARGY | 5 | 1 | 4 | 0 | 0 | 0 |
| SOMNOLENCE | 5 | 3 | 2 | 0 | 0 | 0 |
| *NEUROLOGICAL SIGNS AND SYMPTOMS NEC* | | | | | | |
| BRAIN FOG | 1 | 1 | 0 | 0 | 0 | 0 |
| DIZZINESS | 18 | 6 | 7 | 2 | 0 | 3 |
| INFANT IRRITABILITY | 1 | 0 | 1 | 0 | 0 | 0 |
| NEUROLOGICAL SYMPTOM | 1 | 1 | 0 | 0 | 0 | 0 |
| PRESYNCOPE | 1 | 1 | 0 | 0 | 0 | 0 |
| *PARAESTHESIAS AND DYSAESTHESIAS* | | | | | | |
| HYPOAESTHESIA | 1 | 1 | 0 | 0 | 0 | 0 |
| PARAESTHESIA | 2 | 2 | 0 | 0 | 0 | 0 |
| *SENSORY ABNORMALITIES NEC* | | | | | | |
| AGEUSIA | 1 | 1 | 0 | 0 | 0 | 0 |
| DYSGEUSIA | 2 | 2 | 0 | 0 | 0 | 0 |
| ***SEIZURES (INCL SUBTYPES)*** | | | | | | |
| *SEIZURES AND SEIZURE DISORDERS NEC* | | | | | | |
| SEIZURE | 2 | 2 | 0 | 0 | 0 | 0 |
| **PREGNANCY, PUERPERIUM AND PERINATAL CONDITIONS** | | | | | | |
| ***PLACENTAL, AMNIOTIC AND CAVITY DISORDERS (EXCL HAEMORRHAGES)*** | | | | | | |
| *PLACENTAL ABNORMALITIES (EXCL NEOPLASMS)* | | | | | | |
| PLACENTAL INFARCTION | 1 | 1 | 0 | 0 | 0 | 0 |
| **PSYCHIATRIC DISORDERS** | | | | | | |
| ***ANXIETY DISORDERS AND SYMPTOMS*** | | | | | | |
| *ANXIETY SYMPTOMS* | | | | | | |
| AGITATION | 1 | 0 | 1 | 0 | 0 | 0 |
| ANXIETY | 2 | 0 | 0 | 0 | 0 | 2 |
| ***DELIRIA (INCL CONFUSION)*** | | | | | | |
| *CONFUSION AND DISORIENTATION* | | | | | | |
| CONFUSIONAL STATE | 2 | 2 | 0 | 0 | 0 | 0 |
| ***DEPRESSED MOOD DISORDERS AND DISTURBANCES*** | | | | | | |
| *MOOD ALTERATIONS WITH DEPRESSIVE SYMPTOMS* | | | | | | |
| DEPRESSED MOOD | 1 | 1 | 0 | 0 | 0 | 0 |
| ***MOOD DISORDERS AND DISTURBANCES NEC*** | | | | | | |
| *EMOTIONAL AND MOOD DISTURBANCES NEC* | | | | | | |
| EMOTIONAL DISORDER | 1 | 0 | 1 | 0 | 0 | 0 |
| IRRITABILITY | 1 | 1 | 0 | 0 | 0 | 0 |
| MOOD ALTERED | 1 | 0 | 0 | 1 | 0 | 0 |
| ***SLEEP DISORDERS AND DISTURBANCES*** | | | | | | |
| *DISTURBANCES IN INITIATING AND MAINTAINING SLEEP* | | | | | | |
| INSOMNIA | 2 | 2 | 0 | 0 | 0 | 0 |
| **REPRODUCTIVE SYSTEM AND BREAST DISORDERS** | | | | | | |
| ***MENSTRUAL CYCLE AND UTERINE BLEEDING DISORDERS*** | | | | | | |
| *MENSTRUATION AND UTERINE BLEEDING NEC* | | | | | | |
| MENSTRUATION IRREGULAR | 1 | 1 | 0 | 0 | 0 | 0 |
| *MENSTRUATION WITH DECREASED BLEEDING* | | | | | | |

| | | | | | | |
|---|---|---|---|---|---|---|
| HYPOMENORRHOEA | 1 | 0 | 1 | 0 | 0 | 0 |
| MENSTRUATION DELAYED | 1 | 0 | 1 | 0 | 0 | 0 |
| OLIGOMENORRHOEA | 1 | 0 | 0 | 0 | 0 | 1 |
| *MENSTRUATION WITH INCREASED BLEEDING* | | | | | | |
| HEAVY MENSTRUAL BLEEDING | 1 | 1 | 0 | 0 | 0 | 0 |
| ***VULVOVAGINAL DISORDERS (EXCL INFECTIONS AND INFLAMMATIONS)*** | | | | | | |
| *VULVOVAGINAL DISORDERS NEC* | | | | | | |
| VAGINAL HAEMORRHAGE | 1 | 1 | 0 | 0 | 0 | 0 |
| **RESPIRATORY, THORACIC AND MEDIASTINAL DISORDERS** | | | | | | |
| ***RESPIRATORY DISORDERS NEC*** | | | | | | |
| *BREATHING ABNORMALITIES* | | | | | | |
| DYSPNOEA | 4 | 3 | 1 | 0 | 0 | 0 |
| *COUGHING AND ASSOCIATED SYMPTOMS* | | | | | | |
| COUGH | 2 | 2 | 0 | 0 | 0 | 0 |
| ***RESPIRATORY TRACT SIGNS AND SYMPTOMS*** | | | | | | |
| *UPPER RESPIRATORY TRACT SIGNS AND SYMPTOMS* | | | | | | |
| OROPHARYNGEAL PAIN | 9 | 7 | 2 | 0 | 0 | 0 |
| PARANASAL SINUS DISCOMFORT | 2 | 2 | 0 | 0 | 0 | 0 |
| RHINORRHOEA | 2 | 1 | 0 | 1 | 0 | 0 |
| **SKIN AND SUBCUTANEOUS TISSUE DISORDERS** | | | | | | |
| ***EPIDERMAL AND DERMAL CONDITIONS*** | | | | | | |
| *DERMAL AND EPIDERMAL CONDITIONS NEC* | | | | | | |
| SENSITIVE SKIN | 1 | 1 | 0 | 0 | 0 | 0 |
| *DERMATITIS AND ECZEMA* | | | | | | |
| DERMATITIS ALLERGIC | 1 | 1 | 0 | 0 | 0 | 0 |
| *ERYTHEMAS* | | | | | | |
| ERYTHEMA | 3 | 1 | 0 | 0 | 0 | 2 |
| *PRURITUS NEC* | | | | | | |
| PRURITUS | 4 | 4 | 0 | 0 | 0 | 0 |
| *RASHES, ERUPTIONS AND EXANTHEMS NEC* | | | | | | |
| RASH | 4 | 2 | 1 | 1 | 0 | 0 |
| ***SKIN APPENDAGE CONDITIONS*** | | | | | | |
| *APOCRINE AND ECCRINE GLAND DISORDERS* | | | | | | |
| COLD SWEAT | 1 | 0 | 1 | 0 | 0 | 0 |
| HYPERHIDROSIS | 4 | 3 | 0 | 0 | 0 | 1 |
| NIGHT SWEATS | 2 | 1 | 1 | 0 | 0 | 0 |
| **VASCULAR DISORDERS** | | | | | | |
| ***EMBOLISM AND THROMBOSIS*** | | | | | | |
| *NON-SITE SPECIFIC EMBOLISM AND THROMBOSIS* | | | | | | |
| EMBOLISM | 1 | 1 | 0 | 0 | 0 | 0 |
| ***VASCULAR DISORDERS NEC*** | | | | | | |
| *PERIPHERAL VASCULAR DISORDERS NEC* | | | | | | |
| HOT FLUSH | 1 | 1 | 0 | 0 | 0 | 0 |
| **TOTAL ADR EVENTS** | ***900*** | ***564*** | ***199*** | ***81*** | ***0*** | ***56*** |

**TABLE S2-8. Pfizer BioNTech COVID-19 vaccine: ADR listing for events reported by breast-feeding patients, in those reporting any vaccination dose**

| | Patients with any vaccination dose: ADR Counts | | | | |
|---|---|---|---|---|---|
| **REACTION TERM (SOC, *HLGT*, *HLT*, PT)** | All doses | 1st dose | 2nd dose | 3rd dose | Other doses |
| ***(freetext)*** | 1 | 1 | 0 | 0 | 0 |
| **BLOOD AND LYMPHATIC SYSTEM DISORDERS** | | | | | |
| *LYMPHATIC SYSTEM DISORDERS NEC* | | | | | |
| LYMPH NODE PAIN | 2 | 0 | 1 | 1 | 0 |
| LYMPHADENOPATHY | 2 | 1 | 1 | 0 | 0 |
| **CARDIAC DISORDERS** | | | | | |
| ***CARDIAC DISORDERS, SIGNS AND SYMPTOMS NEC*** | | | | | |
| *CARDIAC SIGNS AND SYMPTOMS NEC* | | | | | |
| PALPITATIONS | 3 | 1 | 2 | 0 | 0 |
| **EAR AND LABYRINTH DISORDERS** | | | | | |
| ***AURAL DISORDERS NEC*** | | | | | |
| *EAR DISORDERS NEC* | | | | | |
| EAR PAIN | 0 | 0 | 0 | 0 | 0 |
| ***INNER EAR AND VIIITH CRANIAL NERVE DISORDERS*** | | | | | |
| *INNER EAR SIGNS AND SYMPTOMS* | | | | | |
| TINNITUS | 1 | 0 | 1 | 0 | 0 |
| **EYE DISORDERS** | | | | | |
| ***EYE DISORDERS NEC*** | | | | | |
| *LACRIMATION DISORDERS* | | | | | |
| DRY EYE | 0 | 0 | 0 | 0 | 0 |
| *OCULAR DISORDERS NEC* | | | | | |
| EYE PAIN | 0 | 0 | 0 | 0 | 0 |
| ***OCULAR INFECTIONS, IRRITATIONS AND INFLAMMATIONS*** | | | | | |
| *OCULAR INFECTIONS, INFLAMMATIONS AND ASSOCIATED MANIFESTATIONS* | | | | | |
| EYE IRRITATION | 0 | 0 | 0 | 0 | 0 |
| ***OCULAR SENSORY SYMPTOMS NEC*** | | | | | |
| *OCULAR SENSATION DISORDERS* | | | | | |
| PHOTOPHOBIA | 1 | 1 | 0 | 0 | 0 |
| ***VISION DISORDERS*** | | | | | |
| *VISUAL DISORDERS NEC* | | | | | |
| VISION BLURRED | 0 | 0 | 0 | 0 | 0 |
| **GASTROINTESTINAL DISORDERS** | | | | | |
| ***GASTROINTESTINAL INFLAMMATORY CONDITIONS*** | | | | | |
| *GASTROINTESTINAL INFLAMMATORY DISORDERS NEC* | | | | | |
| GASTROINTESTINAL TRACT IRRITATION | 1 | 0 | 0 | 1 | 0 |
| ***GASTROINTESTINAL MOTILITY AND DEFAECATION CONDITIONS*** | | | | | |
| *DIARRHOEA (EXCL INFECTIVE)* | | | | | |
| DIARRHOEA | 8 | 4 | 2 | 2 | 0 |
| ***GASTROINTESTINAL SIGNS AND SYMPTOMS*** | | | | | |
| *FLATULENCE, BLOATING AND DISTENSION* | | | | | |
| ABDOMINAL DISTENSION | 0 | 0 | 0 | 0 | 0 |
| FLATULENCE | 0 | 0 | 0 | 0 | 0 |
| *GASTROINTESTINAL AND ABDOMINAL PAINS (EXCL ORAL AND THROAT)* | | | | | |
| ABDOMINAL PAIN | 0 | 0 | 0 | 0 | 0 |
| ABDOMINAL PAIN UPPER | 1 | 1 | 0 | 0 | 0 |

| | | | | | |
|---|---|---|---|---|---|
| *NAUSEA AND VOMITING SYMPTOMS* | | | | | |
| NAUSEA | 11 | 5 | 6 | 0 | 0 |
| RETCHING | 0 | 0 | 0 | 0 | 0 |
| VOMITING | 2 | 1 | 1 | 0 | 0 |
| ***ORAL SOFT TISSUE CONDITIONS*** | | | | | |
| *ORAL SOFT TISSUE SIGNS AND SYMPTOMS* | | | | | |
| PARAESTHESIA ORAL | 0 | 0 | 0 | 0 | 0 |
| LIP SWELLING | 0 | 0 | 0 | 0 | 0 |
| **GENERAL DISORDERS AND ADMINISTRATION SITE CONDITIONS** | | | | | |
| ***ADMINISTRATION SITE REACTIONS*** | | | | | |
| *ADMINISTRATION SITE REACTIONS NEC* | | | | | |
| PUNCTURE SITE BRUISE | 0 | 0 | 0 | 0 | 0 |
| *INJECTION SITE REACTIONS* | | | | | |
| INJECTION SITE ERYTHEMA | 1 | 1 | 0 | 0 | 0 |
| INJECTION SITE MASS | 0 | 0 | 0 | 0 | 0 |
| INJECTION SITE PAIN | 15 | 12 | 3 | 0 | 0 |
| INJECTION SITE WARMTH | 1 | 1 | 0 | 0 | 0 |
| *VACCINATION SITE REACTIONS* | | | | | |
| VACCINATION SITE PAIN | 0 | 0 | 0 | 0 | 0 |
| ***BODY TEMPERATURE CONDITIONS*** | | | | | |
| *FEBRILE DISORDERS* | | | | | |
| PYREXIA | 14 | 5 | 8 | 1 | 0 |
| ***GENERAL SYSTEM DISORDERS NEC*** | | | | | |
| *ASTHENIC CONDITIONS* | | | | | |
| FATIGUE | 47 | 26 | 18 | 3 | 0 |
| MALAISE | 2 | 0 | 2 | 0 | 0 |
| *FEELINGS AND SENSATIONS NEC* | | | | | |
| CHILLS | 6 | 2 | 3 | 1 | 0 |
| FEELING ABNORMAL | 1 | 0 | 0 | 1 | 0 |
| FEELING COLD | 1 | 0 | 1 | 0 | 0 |
| FEELING HOT | 0 | 0 | 0 | 0 | 0 |
| HANGOVER | 1 | 0 | 0 | 1 | 0 |
| *GENERAL SIGNS AND SYMPTOMS NEC* | | | | | |
| INFLUENZA LIKE ILLNESS | 1 | 0 | 1 | 0 | 0 |
| LOCAL REACTION | 1 | 0 | 1 | 0 | 0 |
| PERIPHERAL SWELLING | 2 | 1 | 1 | 0 | 0 |
| SWELLING | 0 | 0 | 0 | 0 | 0 |
| *PAIN AND DISCOMFORT NEC* | | | | | |
| AXILLARY PAIN | 4 | 1 | 3 | 0 | 0 |
| CHEST DISCOMFORT | 0 | 0 | 0 | 0 | 0 |
| PAIN | 17 | 10 | 4 | 3 | 0 |
| TENDERNESS | 2 | 0 | 2 | 0 | 0 |
| **INFECTIONS AND INFESTATIONS** | | | | | |
| ***INFECTIONS - PATHOGEN UNSPECIFIED*** | | | | | |
| *BREAST INFECTIONS* | | | | | |
| MASTITIS | 0 | 0 | 0 | 0 | 0 |
| *SKIN STRUCTURES AND SOFT TISSUE INFECTIONS* | | | | | |
| INJECTION SITE PUSTULE | 0 | 0 | 0 | 0 | 0 |
| *UPPER RESPIRATORY TRACT INFECTIONS* | | | | | |
| LARYNGITIS | 1 | 0 | 1 | 0 | 0 |
| NASOPHARYNGITIS | 1 | 0 | 1 | 0 | 0 |
| ***VIRAL INFECTIOUS DISORDERS*** | | | | | |
| *HERPES VIRAL INFECTIONS* | | | | | |

| | | | | | |
|---|---|---|---|---|---|
| ORAL HERPES | 1 | 0 | 1 | 0 | 0 |
| *INFLUENZA VIRAL INFECTIONS* | | | | | |
| INFLUENZA | 3 | 0 | 2 | 1 | 0 |
| **INJURY, POISONING AND PROCEDURAL COMPLICATIONS** | | | | | |
| ***INJURIES NEC*** | | | | | |
| *SKIN INJURIES NEC* | | | | | |
| CONTUSION | 1 | 1 | 0 | 0 | 0 |
| **INVESTIGATIONS** | | | | | |
| ***CARDIAC AND VASCULAR INVESTIGATIONS (EXCL ENZYME TESTS)*** | | | | | |
| *HEART RATE AND PULSE INVESTIGATIONS* | | | | | |
| HEART RATE | 0 | 0 | 0 | 0 | 0 |
| ***PHYSICAL EXAMINATION AND ORGAN SYSTEM STATUS TOPICS*** | | | | | |
| *PHYSICAL EXAMINATION PROCEDURES AND ORGAN SYSTEM STATUS* | | | | | |
| BODY TEMPERATURE | 0 | 0 | 0 | 0 | 0 |
| BODY TEMPERATURE INCREASED | 0 | 0 | 0 | 0 | 0 |
| ***WATER, ELECTROLYTE AND MINERAL INVESTIGATIONS*** | | | | | |
| *WATER AND ELECTROLYTE ANALYSES NEC* | | | | | |
| VOLUME BLOOD | 1 | 0 | 1 | 0 | 0 |
| **METABOLISM AND NUTRITION DISORDERS** | | | | | |
| ***APPETITE AND GENERAL NUTRITIONAL DISORDERS*** | | | | | |
| *APPETITE DISORDERS* | | | | | |
| DECREASED APPETITE | 0 | 0 | 0 | 0 | 0 |
| **MUSCULOSKELETAL AND CONNECTIVE TISSUE DISORDERS** | | | | | |
| ***BONE DISORDERS (EXCL CONGENITAL AND FRACTURES)*** | | | | | |
| *BONE RELATED SIGNS AND SYMPTOMS* | | | | | |
| BONE PAIN | 0 | 0 | 0 | 0 | 0 |
| PAIN IN JAW | 0 | 0 | 0 | 0 | 0 |
| ***JOINT DISORDERS*** | | | | | |
| *JOINT RELATED SIGNS AND SYMPTOMS* | | | | | |
| ARTHRALGIA | 6 | 3 | 3 | 0 | 0 |
| JOINT STIFFNESS | 0 | 0 | 0 | 0 | 0 |
| ***MUSCLE DISORDERS*** | | | | | |
| *MUSCLE PAINS* | | | | | |
| MYALGIA | 10 | 5 | 4 | 1 | 0 |
| *MUSCLE RELATED SIGNS AND SYMPTOMS NEC* | | | | | |
| MUSCLE FATIGUE | 0 | 0 | 0 | 0 | 0 |
| MUSCLE SPASMS | 0 | 0 | 0 | 0 | 0 |
| ***MUSCULOSKELETAL AND CONNECTIVE TISSUE DISORDERS NEC*** | | | | | |
| *MUSCULOSKELETAL AND CONNECTIVE TISSUE CONDITIONS NEC* | | | | | |
| MUSCULOSKELETAL STIFFNESS | 2 | 0 | 0 | 2 | 0 |
| BACK PAIN | 1 | 0 | 0 | 1 | 0 |
| LIMB DISCOMFORT | 19 | 14 | 5 | 0 | 0 |
| PAIN IN EXTREMITY | 89 | 69 | 14 | 6 | 0 |
| **NERVOUS SYSTEM DISORDERS** | | | | | |
| ***CRANIAL NERVE DISORDERS (EXCL NEOPLASMS)*** | | | | | |
| *OLFACTORY NERVE DISORDERS* | | | | | |
| ANOSMIA | 1 | 1 | 0 | 0 | 0 |
| PAROSMIA | 0 | 0 | 0 | 0 | 0 |
| ***HEADACHES*** | | | | | |
| *HEADACHES NEC* | | | | | |
| HEADACHE | 40 | 24 | 9 | 7 | 0 |
| SINUS HEADACHE | 2 | 0 | 2 | 0 | 0 |
| TENSION HEADACHE | 1 | 1 | 0 | 0 | 0 |

| | | | | | |
|---|---|---|---|---|---|
| *MIGRAINE HEADACHES* | | | | | |
| MIGRAINE | 0 | 0 | 0 | 0 | 0 |
| TYPICAL AURA WITHOUT HEADACHE | 1 | 0 | 1 | 0 | 0 |
| ***MOVEMENT DISORDERS (INCL PARKINSONISM)*** | | | | | |
| *TREMOR (EXCL CONGENITAL)* | | | | | |
| TREMOR | 0 | 0 | 0 | 0 | 0 |
| ***NEUROLOGICAL DISORDERS NEC*** | | | | | |
| *DISTURBANCES IN CONSCIOUSNESS NEC* | | | | | |
| LETHARGY | 3 | 0 | 3 | 0 | 0 |
| SOMNOLENCE | 2 | 2 | 0 | 0 | 0 |
| *NEUROLOGICAL SIGNS AND SYMPTOMS NEC* | | | | | |
| BRAIN FOG | 0 | 0 | 0 | 0 | 0 |
| DIZZINESS | 8 | 2 | 5 | 1 | 0 |
| INFANT IRRITABILITY | 0 | 0 | 0 | 0 | 0 |
| NEUROLOGICAL SYMPTOM | 0 | 0 | 0 | 0 | 0 |
| PRESYNCOPE | 0 | 0 | 0 | 0 | 0 |
| *PARAESTHESIAS AND DYSAESTHESIAS* | | | | | |
| HYPOAESTHESIA | 1 | 1 | 0 | 0 | 0 |
| PARAESTHESIA | 1 | 1 | 0 | 0 | 0 |
| *SENSORY ABNORMALITIES NEC* | | | | | |
| AGEUSIA | 0 | 0 | 0 | 0 | 0 |
| DYSGEUSIA | 0 | 0 | 0 | 0 | 0 |
| ***SEIZURES (INCL SUBTYPES)*** | | | | | |
| *SEIZURES AND SEIZURE DISORDERS NEC* | | | | | |
| SEIZURE | 0 | 0 | 0 | 0 | 0 |
| **PREGNANCY, PUERPERIUM AND PERINATAL CONDITIONS** | | | | | |
| ***PLACENTAL, AMNIOTIC AND CAVITY DISORDERS (EXCL HAEMORRHAGES)*** | | | | | |
| *PLACENTAL ABNORMALITIES (EXCL NEOPLASMS)* | | | | | |
| PLACENTAL INFARCTION | 0 | 0 | 0 | 0 | 0 |
| **PSYCHIATRIC DISORDERS** | | | | | |
| ***ANXIETY DISORDERS AND SYMPTOMS*** | | | | | |
| *ANXIETY SYMPTOMS* | | | | | |
| AGITATION | 1 | 0 | 1 | 0 | 0 |
| ANXIETY | 0 | 0 | 0 | 0 | 0 |
| ***DELIRIA (INCL CONFUSION)*** | | | | | |
| *CONFUSION AND DISORIENTATION* | | | | | |
| CONFUSIONAL STATE | 1 | 1 | 0 | 0 | 0 |
| ***DEPRESSED MOOD DISORDERS AND DISTURBANCES*** | | | | | |
| *MOOD ALTERATIONS WITH DEPRESSIVE SYMPTOMS* | | | | | |
| DEPRESSED MOOD | 1 | 1 | 0 | 0 | 0 |
| ***MOOD DISORDERS AND DISTURBANCES NEC*** | | | | | |
| *EMOTIONAL AND MOOD DISTURBANCES NEC* | | | | | |
| EMOTIONAL DISORDER | 1 | 0 | 1 | 0 | 0 |
| IRRITABILITY | 0 | 0 | 0 | 0 | 0 |
| MOOD ALTERED | 0 | 0 | 0 | 0 | 0 |
| ***SLEEP DISORDERS AND DISTURBANCES*** | | | | | |
| *DISTURBANCES IN INITIATING AND MAINTAINING SLEEP* | | | | | |
| INSOMNIA | 0 | 0 | 0 | 0 | 0 |
| **REPRODUCTIVE SYSTEM AND BREAST DISORDERS** | | | | | |
| ***MENSTRUAL CYCLE AND UTERINE BLEEDING DISORDERS*** | | | | | |
| *MENSTRUATION AND UTERINE BLEEDING NEC* | | | | | |
| MENSTRUATION IRREGULAR | 1 | 1 | 0 | 0 | 0 |
| *MENSTRUATION WITH DECREASED BLEEDING* | | | | | |

| | | | | | |
|---|---|---|---|---|---|
| HYPOMENORRHOEA | 1 | 0 | 1 | 0 | 0 |
| MENSTRUATION DELAYED | 0 | 0 | 0 | 0 | 0 |
| OLIGOMENORRHOEA | 0 | 0 | 0 | 0 | 0 |
| *MENSTRUATION WITH INCREASED BLEEDING* | | | | | |
| HEAVY MENSTRUAL BLEEDING | 0 | 0 | 0 | 0 | 0 |
| ***VULVOVAGINAL DISORDERS (EXCL INFECTIONS AND INFLAMMATIONS)*** | | | | | |
| *VULVOVAGINAL DISORDERS NEC* | | | | | |
| VAGINAL HAEMORRHAGE | 1 | 1 | 0 | 0 | 0 |
| **RESPIRATORY, THORACIC AND MEDIASTINAL DISORDERS** | | | | | |
| ***RESPIRATORY DISORDERS NEC*** | | | | | |
| *BREATHING ABNORMALITIES* | | | | | |
| DYSPNOEA | 2 | 2 | 0 | 0 | 0 |
| *COUGHING AND ASSOCIATED SYMPTOMS* | | | | | |
| COUGH | 2 | 2 | 0 | 0 | 0 |
| ***RESPIRATORY TRACT SIGNS AND SYMPTOMS*** | | | | | |
| *UPPER RESPIRATORY TRACT SIGNS AND SYMPTOMS* | | | | | |
| OROPHARYNGEAL PAIN | 5 | 3 | 2 | 0 | 0 |
| PARANASAL SINUS DISCOMFORT | 1 | 1 | 0 | 0 | 0 |
| RHINORRHOEA | 1 | 1 | 0 | 0 | 0 |
| **SKIN AND SUBCUTANEOUS TISSUE DISORDERS** | | | | | |
| ***EPIDERMAL AND DERMAL CONDITIONS*** | | | | | |
| *DERMAL AND EPIDERMAL CONDITIONS NEC* | | | | | |
| SENSITIVE SKIN | 0 | 0 | 0 | 0 | 0 |
| *DERMATITIS AND ECZEMA* | | | | | |
| DERMATITIS ALLERGIC | 0 | 0 | 0 | 0 | 0 |
| *ERYTHEMAS* | | | | | |
| ERYTHEMA | 0 | 0 | 0 | 0 | 0 |
| *PRURITUS NEC* | | | | | |
| PRURITUS | 1 | 1 | 0 | 0 | 0 |
| *RASHES, ERUPTIONS AND EXANTHEMS NEC* | | | | | |
| RASH | 2 | 1 | 1 | 0 | 0 |
| ***SKIN APPENDAGE CONDITIONS*** | | | | | |
| *APOCRINE AND ECCRINE GLAND DISORDERS* | | | | | |
| COLD SWEAT | 0 | 0 | 0 | 0 | 0 |
| HYPERHIDROSIS | 0 | 0 | 0 | 0 | 0 |
| NIGHT SWEATS | 1 | 0 | 1 | 0 | 0 |
| **VASCULAR DISORDERS** | | | | | |
| ***EMBOLISM AND THROMBOSIS*** | | | | | |
| *NON-SITE SPECIFIC EMBOLISM AND THROMBOSIS* | | | | | |
| EMBOLISM | 0 | 0 | 0 | 0 | 0 |
| ***VASCULAR DISORDERS NEC*** | | | | | |
| *PERIPHERAL VASCULAR DISORDERS NEC* | | | | | |
| HOT FLUSH | 0 | 0 | 0 | 0 | 0 |
| **TOTAL ADR EVENTS** | ***366*** | ***213*** | ***120*** | ***33*** | ***0*** |

**TABLE S2-9. AstraZeneca COVID-19 vaccine: ADR listing for events reported by breast-feeding patients, in those reporting any vaccination dose**

| REACTION TERM (SOC, ***HLGT***, *HLT*, PT) | Patients with any vaccination dose: ADR Counts | | | | |
|---|---|---|---|---|---|
| | All doses | 1st dose | 2nd dose | 3rd dose | Other doses |
| ***(freetext)*** | 0 | 0 | 0 | 0 | 0 |
| **BLOOD AND LYMPHATIC SYSTEM DISORDERS** | | | | | |
| *LYMPHATIC SYSTEM DISORDERS NEC* | | | | | |
| LYMPH NODE PAIN | 0 | 0 | 0 | 0 | 0 |
| LYMPHADENOPATHY | 1 | 0 | 1 | 0 | 0 |
| **CARDIAC DISORDERS** | | | | | |
| ***CARDIAC DISORDERS, SIGNS AND SYMPTOMS NEC*** | | | | | |
| *CARDIAC SIGNS AND SYMPTOMS NEC* | | | | | |
| PALPITATIONS | 1 | 1 | 0 | 0 | 0 |
| **EAR AND LABYRINTH DISORDERS** | | | | | |
| ***AURAL DISORDERS NEC*** | | | | | |
| *EAR DISORDERS NEC* | | | | | |
| EAR PAIN | 1 | 1 | 0 | 0 | 0 |
| ***INNER EAR AND VIIITH CRANIAL NERVE DISORDERS*** | | | | | |
| *INNER EAR SIGNS AND SYMPTOMS* | | | | | |
| TINNITUS | 0 | 0 | 0 | 0 | 0 |
| **EYE DISORDERS** | | | | | |
| ***EYE DISORDERS NEC*** | | | | | |
| *LACRIMATION DISORDERS* | | | | | |
| DRY EYE | 1 | 1 | 0 | 0 | 0 |
| *OCULAR DISORDERS NEC* | | | | | |
| EYE PAIN | 1 | 1 | 0 | 0 | 0 |
| ***OCULAR INFECTIONS, IRRITATIONS AND INFLAMMATIONS*** | | | | | |
| *OCULAR INFECTIONS, INFLAMMATIONS AND ASSOCIATED MANIFESTATIONS* | | | | | |
| EYE IRRITATION | 0 | 0 | 0 | 0 | 0 |
| ***OCULAR SENSORY SYMPTOMS NEC*** | | | | | |
| *OCULAR SENSATION DISORDERS* | | | | | |
| PHOTOPHOBIA | 1 | 1 | 0 | 0 | 0 |
| ***VISION DISORDERS*** | | | | | |
| *VISUAL DISORDERS NEC* | | | | | |
| VISION BLURRED | 2 | 2 | 0 | 0 | 0 |
| **GASTROINTESTINAL DISORDERS** | | | | | |
| ***GASTROINTESTINAL INFLAMMATORY CONDITIONS*** | | | | | |
| *GASTROINTESTINAL INFLAMMATORY DISORDERS NEC* | | | | | |
| GASTROINTESTINAL TRACT IRRITATION | 0 | 0 | 0 | 0 | 0 |
| ***GASTROINTESTINAL MOTILITY AND DEFAECATION CONDITIONS*** | | | | | |
| *DIARRHOEA (EXCL INFECTIVE)* | | | | | |
| DIARRHOEA | 3 | 3 | 0 | 0 | 0 |
| ***GASTROINTESTINAL SIGNS AND SYMPTOMS*** | | | | | |
| *FLATULENCE, BLOATING AND DISTENSION* | | | | | |
| ABDOMINAL DISTENSION | 1 | 1 | 0 | 0 | 0 |
| FLATULENCE | 1 | 1 | 0 | 0 | 0 |
| *GASTROINTESTINAL AND ABDOMINAL PAINS (EXCL ORAL AND THROAT)* | | | | | |
| ABDOMINAL PAIN | 5 | 5 | 0 | 0 | 0 |
| ABDOMINAL PAIN UPPER | 2 | 1 | 1 | 0 | 0 |

| | | | | | |
|---|---|---|---|---|---|
| *NAUSEA AND VOMITING SYMPTOMS* | | | | | |
| NAUSEA | 17 | 15 | 2 | 0 | 0 |
| RETCHING | 1 | 1 | 0 | 0 | 0 |
| VOMITING | 3 | 3 | 0 | 0 | 0 |
| ***ORAL SOFT TISSUE CONDITIONS*** | | | | | |
| *ORAL SOFT TISSUE SIGNS AND SYMPTOMS* | | | | | |
| PARAESTHESIA ORAL | 1 | 1 | 0 | 0 | 0 |
| LIP SWELLING | 0 | 0 | 0 | 0 | 0 |
| **GENERAL DISORDERS AND ADMINISTRATION SITE CONDITIONS** | | | | | |
| ***ADMINISTRATION SITE REACTIONS*** | | | | | |
| *ADMINISTRATION SITE REACTIONS NEC* | | | | | |
| PUNCTURE SITE BRUISE | 0 | 0 | 0 | 0 | 0 |
| *INJECTION SITE REACTIONS* | | | | | |
| INJECTION SITE ERYTHEMA | 0 | 0 | 0 | 0 | 0 |
| INJECTION SITE MASS | 0 | 0 | 0 | 0 | 0 |
| INJECTION SITE PAIN | 4 | 4 | 0 | 0 | 0 |
| INJECTION SITE WARMTH | 0 | 0 | 0 | 0 | 0 |
| *VACCINATION SITE REACTIONS* | | | | | |
| VACCINATION SITE PAIN | 1 | 0 | 1 | 0 | 0 |
| ***BODY TEMPERATURE CONDITIONS*** | | | | | |
| *FEBRILE DISORDERS* | | | | | |
| PYREXIA | 41 | 38 | 3 | 0 | 0 |
| ***GENERAL SYSTEM DISORDERS NEC*** | | | | | |
| *ASTHENIC CONDITIONS* | | | | | |
| FATIGUE | 38 | 29 | 9 | 0 | 0 |
| MALAISE | 2 | 2 | 0 | 0 | 0 |
| *FEELINGS AND SENSATIONS NEC* | | | | | |
| CHILLS | 25 | 25 | 0 | 0 | 0 |
| FEELING ABNORMAL | 0 | 0 | 0 | 0 | 0 |
| FEELING COLD | 7 | 7 | 0 | 0 | 0 |
| FEELING HOT | 1 | 1 | 0 | 0 | 0 |
| HANGOVER | 0 | 0 | 0 | 0 | 0 |
| *GENERAL SIGNS AND SYMPTOMS NEC* | | | | | |
| INFLUENZA LIKE ILLNESS | 4 | 4 | 0 | 0 | 0 |
| LOCAL REACTION | 0 | 0 | 0 | 0 | 0 |
| PERIPHERAL SWELLING | 1 | 1 | 0 | 0 | 0 |
| SWELLING | 0 | 0 | 0 | 0 | 0 |
| *PAIN AND DISCOMFORT NEC* | | | | | |
| AXILLARY PAIN | 0 | 0 | 0 | 0 | 0 |
| CHEST DISCOMFORT | 1 | 1 | 0 | 0 | 0 |
| PAIN | 7 | 6 | 1 | 0 | 0 |
| TENDERNESS | 2 | 2 | 0 | 0 | 0 |
| **INFECTIONS AND INFESTATIONS** | | | | | |
| ***INFECTIONS - PATHOGEN UNSPECIFIED*** | | | | | |
| *BREAST INFECTIONS* | | | | | |
| MASTITIS | 0 | 0 | 0 | 0 | 0 |
| *SKIN STRUCTURES AND SOFT TISSUE INFECTIONS* | | | | | |
| INJECTION SITE PUSTULE | 1 | 0 | 1 | 0 | 0 |
| *UPPER RESPIRATORY TRACT INFECTIONS* | | | | | |
| LARYNGITIS | 0 | 0 | 0 | 0 | 0 |
| NASOPHARYNGITIS | 2 | 2 | 0 | 0 | 0 |
| ***VIRAL INFECTIOUS DISORDERS*** | | | | | |
| *HERPES VIRAL INFECTIONS* | | | | | |

| | | | | | |
|---|---|---|---|---|---|
| ORAL HERPES | 0 | 0 | 0 | 0 | 0 |
| *INFLUENZA VIRAL INFECTIONS* | | | | | |
| INFLUENZA | 5 | 5 | 0 | 0 | 0 |
| **INJURY, POISONING AND PROCEDURAL COMPLICATIONS** | | | | | |
| ***INJURIES NEC*** | | | | | |
| *SKIN INJURIES NEC* | | | | | |
| CONTUSION | 1 | 1 | 0 | 0 | 0 |
| **INVESTIGATIONS** | | | | | |
| ***CARDIAC AND VASCULAR INVESTIGATIONS (EXCL ENZYME TESTS)*** | | | | | |
| *HEART RATE AND PULSE INVESTIGATIONS* | | | | | |
| HEART RATE | 1 | 1 | 0 | 0 | 0 |
| ***PHYSICAL EXAMINATION AND ORGAN SYSTEM STATUS TOPICS*** | | | | | |
| *PHYSICAL EXAMINATION PROCEDURES AND ORGAN SYSTEM STATUS* | | | | | |
| BODY TEMPERATURE | 3 | 3 | 0 | 0 | 0 |
| BODY TEMPERATURE INCREASED | 1 | 1 | 0 | 0 | 0 |
| ***WATER, ELECTROLYTE AND MINERAL INVESTIGATIONS*** | | | | | |
| *WATER AND ELECTROLYTE ANALYSES NEC* | | | | | |
| VOLUME BLOOD | 0 | 0 | 0 | 0 | 0 |
| **METABOLISM AND NUTRITION DISORDERS** | | | | | |
| ***APPETITE AND GENERAL NUTRITIONAL DISORDERS*** | | | | | |
| *APPETITE DISORDERS* | | | | | |
| DECREASED APPETITE | 4 | 3 | 1 | 0 | 0 |
| **MUSCULOSKELETAL AND CONNECTIVE TISSUE DISORDERS** | | | | | |
| ***BONE DISORDERS (EXCL CONGENITAL AND FRACTURES)*** | | | | | |
| *BONE RELATED SIGNS AND SYMPTOMS* | | | | | |
| BONE PAIN | 1 | 1 | 0 | 0 | 0 |
| PAIN IN JAW | 1 | 0 | 1 | 0 | 0 |
| ***JOINT DISORDERS*** | | | | | |
| *JOINT RELATED SIGNS AND SYMPTOMS* | | | | | |
| ARTHRALGIA | 6 | 6 | 0 | 0 | 0 |
| JOINT STIFFNESS | 1 | 1 | 0 | 0 | 0 |
| ***MUSCLE DISORDERS*** | | | | | |
| *MUSCLE PAINS* | | | | | |
| MYALGIA | 30 | 28 | 2 | 0 | 0 |
| *MUSCLE RELATED SIGNS AND SYMPTOMS NEC* | | | | | |
| MUSCLE FATIGUE | 0 | 0 | 0 | 0 | 0 |
| MUSCLE SPASMS | 1 | 1 | 0 | 0 | 0 |
| ***MUSCULOSKELETAL AND CONNECTIVE TISSUE DISORDERS NEC*** | | | | | |
| *MUSCULOSKELETAL AND CONNECTIVE TISSUE CONDITIONS NEC* | | | | | |
| MUSCULOSKELETAL STIFFNESS | 0 | 0 | 0 | 0 | 0 |
| BACK PAIN | 2 | 2 | 0 | 0 | 0 |
| LIMB DISCOMFORT | 4 | 4 | 0 | 0 | 0 |
| PAIN IN EXTREMITY | 24 | 20 | 4 | 0 | 0 |
| **NERVOUS SYSTEM DISORDERS** | | | | | |
| ***CRANIAL NERVE DISORDERS (EXCL NEOPLASMS)*** | | | | | |
| *OLFACTORY NERVE DISORDERS* | | | | | |
| ANOSMIA | 0 | 0 | 0 | 0 | 0 |
| PAROSMIA | 1 | 0 | 1 | 0 | 0 |
| ***HEADACHES*** | | | | | |
| *HEADACHES NEC* | | | | | |
| HEADACHE | 40 | 37 | 3 | 0 | 0 |
| SINUS HEADACHE | 1 | 1 | 0 | 0 | 0 |
| TENSION HEADACHE | 4 | 4 | 0 | 0 | 0 |

| | | | | | |
|---|---|---|---|---|---|
| *MIGRAINE HEADACHES* | | | | | |
| MIGRAINE | 1 | 1 | 0 | 0 | 0 |
| TYPICAL AURA WITHOUT HEADACHE | 0 | 0 | 0 | 0 | 0 |
| ***MOVEMENT DISORDERS (INCL PARKINSONISM)*** | | | | | |
| *TREMOR (EXCL CONGENITAL)* | | | | | |
| TREMOR | 1 | 1 | 0 | 0 | 0 |
| ***NEUROLOGICAL DISORDERS NEC*** | | | | | |
| *DISTURBANCES IN CONSCIOUSNESS NEC* | | | | | |
| LETHARGY | 2 | 1 | 1 | 0 | 0 |
| SOMNOLENCE | 2 | 1 | 1 | 0 | 0 |
| *NEUROLOGICAL SIGNS AND SYMPTOMS NEC* | | | | | |
| BRAIN FOG | 1 | 1 | 0 | 0 | 0 |
| DIZZINESS | 4 | 3 | 1 | 0 | 0 |
| INFANT IRRITABILITY | 1 | 0 | 1 | 0 | 0 |
| NEUROLOGICAL SYMPTOM | 1 | 1 | 0 | 0 | 0 |
| PRESYNCOPE | 1 | 1 | 0 | 0 | 0 |
| *PARAESTHESIAS AND DYSAESTHESIAS* | | | | | |
| HYPOAESTHESIA | 0 | 0 | 0 | 0 | 0 |
| PARAESTHESIA | 1 | 1 | 0 | 0 | 0 |
| *SENSORY ABNORMALITIES NEC* | | | | | |
| AGEUSIA | 1 | 1 | 0 | 0 | 0 |
| DYSGEUSIA | 2 | 2 | 0 | 0 | 0 |
| ***SEIZURES (INCL SUBTYPES)*** | | | | | |
| *SEIZURES AND SEIZURE DISORDERS NEC* | | | | | |
| SEIZURE | 2 | 2 | 0 | 0 | 0 |
| **PREGNANCY, PUERPERIUM AND PERINATAL CONDITIONS** | | | | | |
| ***PLACENTAL, AMNIOTIC AND CAVITY DISORDERS (EXCL HAEMORRHAGES)*** | | | | | |
| *PLACENTAL ABNORMALITIES (EXCL NEOPLASMS)* | | | | | |
| PLACENTAL INFARCTION | 1 | 1 | 0 | 0 | 0 |
| **PSYCHIATRIC DISORDERS** | | | | | |
| ***ANXIETY DISORDERS AND SYMPTOMS*** | | | | | |
| *ANXIETY SYMPTOMS* | | | | | |
| AGITATION | 0 | 0 | 0 | 0 | 0 |
| ANXIETY | 0 | 0 | 0 | 0 | 0 |
| ***DELIRIA (INCL CONFUSION)*** | | | | | |
| *CONFUSION AND DISORIENTATION* | | | | | |
| CONFUSIONAL STATE | 1 | 1 | 0 | 0 | 0 |
| ***DEPRESSED MOOD DISORDERS AND DISTURBANCES*** | | | | | |
| *MOOD ALTERATIONS WITH DEPRESSIVE SYMPTOMS* | | | | | |
| DEPRESSED MOOD | 0 | 0 | 0 | 0 | 0 |
| ***MOOD DISORDERS AND DISTURBANCES NEC*** | | | | | |
| *EMOTIONAL AND MOOD DISTURBANCES NEC* | | | | | |
| EMOTIONAL DISORDER | 0 | 0 | 0 | 0 | 0 |
| IRRITABILITY | 1 | 1 | 0 | 0 | 0 |
| MOOD ALTERED | 0 | 0 | 0 | 0 | 0 |
| ***SLEEP DISORDERS AND DISTURBANCES*** | | | | | |
| *DISTURBANCES IN INITIATING AND MAINTAINING SLEEP* | | | | | |
| INSOMNIA | 2 | 2 | 0 | 0 | 0 |
| **REPRODUCTIVE SYSTEM AND BREAST DISORDERS** | | | | | |
| ***MENSTRUAL CYCLE AND UTERINE BLEEDING DISORDERS*** | | | | | |
| *MENSTRUATION AND UTERINE BLEEDING NEC* | | | | | |
| MENSTRUATION IRREGULAR | 0 | 0 | 0 | 0 | 0 |
| *MENSTRUATION WITH DECREASED BLEEDING* | | | | | |

| | | | | | |
|---|---|---|---|---|---|
| HYPOMENORRHOEA | 0 | 0 | 0 | 0 | 0 |
| MENSTRUATION DELAYED | 1 | 0 | 1 | 0 | 0 |
| OLIGOMENORRHOEA | 0 | 0 | 0 | 0 | 0 |
| *MENSTRUATION WITH INCREASED BLEEDING* | | | | | |
| HEAVY MENSTRUAL BLEEDING | 1 | 1 | 0 | 0 | 0 |
| ***VULVOVAGINAL DISORDERS (EXCL INFECTIONS AND INFLAMMATIONS)*** | | | | | |
| *VULVOVAGINAL DISORDERS NEC* | | | | | |
| VAGINAL HAEMORRHAGE | 0 | 0 | 0 | 0 | 0 |
| **RESPIRATORY, THORACIC AND MEDIASTINAL DISORDERS** | | | | | |
| ***RESPIRATORY DISORDERS NEC*** | | | | | |
| *BREATHING ABNORMALITIES* | | | | | |
| DYSPNOEA | 1 | 0 | 1 | 0 | 0 |
| *COUGHING AND ASSOCIATED SYMPTOMS* | | | | | |
| COUGH | 0 | 0 | 0 | 0 | 0 |
| ***RESPIRATORY TRACT SIGNS AND SYMPTOMS*** | | | | | |
| *UPPER RESPIRATORY TRACT SIGNS AND SYMPTOMS* | | | | | |
| OROPHARYNGEAL PAIN | 4 | 4 | 0 | 0 | 0 |
| PARANASAL SINUS DISCOMFORT | 1 | 1 | 0 | 0 | 0 |
| RHINORRHOEA | 0 | 0 | 0 | 0 | 0 |
| **SKIN AND SUBCUTANEOUS TISSUE DISORDERS** | | | | | |
| ***EPIDERMAL AND DERMAL CONDITIONS*** | | | | | |
| *DERMAL AND EPIDERMAL CONDITIONS NEC* | | | | | |
| SENSITIVE SKIN | 1 | 1 | 0 | 0 | 0 |
| *DERMATITIS AND ECZEMA* | | | | | |
| DERMATITIS ALLERGIC | 1 | 1 | 0 | 0 | 0 |
| *ERYTHEMAS* | | | | | |
| ERYTHEMA | 1 | 1 | 0 | 0 | 0 |
| *PRURITUS NEC* | | | | | |
| PRURITUS | 1 | 1 | 0 | 0 | 0 |
| *RASHES, ERUPTIONS AND EXANTHEMS NEC* | | | | | |
| RASH | 0 | 0 | 0 | 0 | 0 |
| ***SKIN APPENDAGE CONDITIONS*** | | | | | |
| *APOCRINE AND ECCRINE GLAND DISORDERS* | | | | | |
| COLD SWEAT | 1 | 0 | 1 | 0 | 0 |
| HYPERHIDROSIS | 3 | 3 | 0 | 0 | 0 |
| NIGHT SWEATS | 1 | 1 | 0 | 0 | 0 |
| **VASCULAR DISORDERS** | | | | | |
| ***EMBOLISM AND THROMBOSIS*** | | | | | |
| *NON-SITE SPECIFIC EMBOLISM AND THROMBOSIS* | | | | | |
| EMBOLISM | 0 | 0 | 0 | 0 | 0 |
| ***VASCULAR DISORDERS NEC*** | | | | | |
| *PERIPHERAL VASCULAR DISORDERS NEC* | | | | | |
| HOT FLUSH | 1 | 1 | 0 | 0 | 0 |
| **TOTAL ADR EVENTS** | ***353*** | ***315*** | ***38*** | ***0*** | ***0*** |

**TABLE S2-10. Moderna COVID-19 vaccine: ADR listing for events reported by breast-feeding patients, in those reporting any vaccination dose**

| REACTION TERM (SOC, ***HLGT***, *HLT*, PT) | Patients with any vaccination dose: ADR Counts | | | | |
|---|---|---|---|---|---|
| | All doses | 1st dose | 2nd dose | 3rd dose | Other doses |
| ***(freetext)*** | 0 | 0 | 0 | 0 | 0 |
| **BLOOD AND LYMPHATIC SYSTEM DISORDERS** | | | | | |
| *LYMPHATIC SYSTEM DISORDERS NEC* | | | | | |
| LYMPH NODE PAIN | 0 | 0 | 0 | 0 | 0 |
| LYMPHADENOPATHY | 1 | 0 | 1 | 0 | 0 |
| **CARDIAC DISORDERS** | | | | | |
| ***CARDIAC DISORDERS, SIGNS AND SYMPTOMS NEC*** | | | | | |
| *CARDIAC SIGNS AND SYMPTOMS NEC* | | | | | |
| PALPITATIONS | 0 | 0 | 0 | 0 | 0 |
| **EAR AND LABYRINTH DISORDERS** | | | | | |
| ***AURAL DISORDERS NEC*** | | | | | |
| *EAR DISORDERS NEC* | | | | | |
| EAR PAIN | 0 | 0 | 0 | 0 | 0 |
| ***INNER EAR AND VIIITH CRANIAL NERVE DISORDERS*** | | | | | |
| *INNER EAR SIGNS AND SYMPTOMS* | | | | | |
| TINNITUS | 1 | 0 | 0 | 1 | 0 |
| **EYE DISORDERS** | | | | | |
| ***EYE DISORDERS NEC*** | | | | | |
| *LACRIMATION DISORDERS* | | | | | |
| DRY EYE | 0 | 0 | 0 | 0 | 0 |
| *OCULAR DISORDERS NEC* | | | | | |
| EYE PAIN | 0 | 0 | 0 | 0 | 0 |
| ***OCULAR INFECTIONS, IRRITATIONS AND INFLAMMATIONS*** | | | | | |
| *OCULAR INFECTIONS, INFLAMMATIONS AND ASSOCIATED MANIFESTATIONS* | | | | | |
| EYE IRRITATION | 1 | 1 | 0 | 0 | 0 |
| ***OCULAR SENSORY SYMPTOMS NEC*** | | | | | |
| *OCULAR SENSATION DISORDERS* | | | | | |
| PHOTOPHOBIA | 0 | 0 | 0 | 0 | 0 |
| ***VISION DISORDERS*** | | | | | |
| *VISUAL DISORDERS NEC* | | | | | |
| VISION BLURRED | 0 | 0 | 0 | 0 | 0 |
| **GASTROINTESTINAL DISORDERS** | | | | | |
| ***GASTROINTESTINAL INFLAMMATORY CONDITIONS*** | | | | | |
| *GASTROINTESTINAL INFLAMMATORY DISORDERS NEC* | | | | | |
| GASTROINTESTINAL TRACT IRRITATION | 0 | 0 | 0 | 0 | 0 |
| ***GASTROINTESTINAL MOTILITY AND DEFAECATION CONDITIONS*** | | | | | |
| *DIARRHOEA (EXCL INFECTIVE)* | | | | | |
| DIARRHOEA | 0 | 0 | 0 | 0 | 0 |
| ***GASTROINTESTINAL SIGNS AND SYMPTOMS*** | | | | | |
| *FLATULENCE, BLOATING AND DISTENSION* | | | | | |
| ABDOMINAL DISTENSION | 0 | 0 | 0 | 0 | 0 |
| FLATULENCE | 0 | 0 | 0 | 0 | 0 |
| *GASTROINTESTINAL AND ABDOMINAL PAINS (EXCL ORAL AND THROAT)* | | | | | |
| ABDOMINAL PAIN | 0 | 0 | 0 | 0 | 0 |
| ABDOMINAL PAIN UPPER | 0 | 0 | 0 | 0 | 0 |

| | | | | | |
|---|---|---|---|---|---|
| *NAUSEA AND VOMITING SYMPTOMS* | | | | | |
| NAUSEA | 6 | 2 | 0 | 4 | 0 |
| RETCHING | 0 | 0 | 0 | 0 | 0 |
| VOMITING | 1 | 0 | 1 | 0 | 0 |
| ***ORAL SOFT TISSUE CONDITIONS*** | | | | | |
| *ORAL SOFT TISSUE SIGNS AND SYMPTOMS* | | | | | |
| PARAESTHESIA ORAL | 0 | 0 | 0 | 0 | 0 |
| LIP SWELLING | 1 | 0 | 1 | 0 | 0 |
| **GENERAL DISORDERS AND ADMINISTRATION SITE CONDITIONS** | | | | | |
| ***ADMINISTRATION SITE REACTIONS*** | | | | | |
| *ADMINISTRATION SITE REACTIONS NEC* | | | | | |
| PUNCTURE SITE BRUISE | 1 | 0 | 0 | 1 | 0 |
| *INJECTION SITE REACTIONS* | | | | | |
| INJECTION SITE ERYTHEMA | 0 | 0 | 0 | 0 | 0 |
| INJECTION SITE MASS | 0 | 0 | 0 | 0 | 0 |
| INJECTION SITE PAIN | 1 | 0 | 0 | 1 | 0 |
| INJECTION SITE WARMTH | 0 | 0 | 0 | 0 | 0 |
| *VACCINATION SITE REACTIONS* | | | | | |
| VACCINATION SITE PAIN | 0 | 0 | 0 | 0 | 0 |
| ***BODY TEMPERATURE CONDITIONS*** | | | | | |
| *FEBRILE DISORDERS* | | | | | |
| PYREXIA | 10 | 1 | 5 | 4 | 0 |
| ***GENERAL SYSTEM DISORDERS NEC*** | | | | | |
| *ASTHENIC CONDITIONS* | | | | | |
| FATIGUE | 20 | 5 | 8 | 7 | 0 |
| MALAISE | 2 | 1 | 1 | 0 | 0 |
| *FEELINGS AND SENSATIONS NEC* | | | | | |
| CHILLS | 8 | 0 | 3 | 5 | 0 |
| FEELING ABNORMAL | 0 | 0 | 0 | 0 | 0 |
| FEELING COLD | 1 | 0 | 1 | 0 | 0 |
| FEELING HOT | 0 | 0 | 0 | 0 | 0 |
| HANGOVER | 0 | 0 | 0 | 0 | 0 |
| *GENERAL SIGNS AND SYMPTOMS NEC* | | | | | |
| INFLUENZA LIKE ILLNESS | 2 | 0 | 0 | 2 | 0 |
| LOCAL REACTION | 0 | 0 | 0 | 0 | 0 |
| PERIPHERAL SWELLING | 1 | 1 | 0 | 0 | 0 |
| SWELLING | 1 | 1 | 0 | 0 | 0 |
| *PAIN AND DISCOMFORT NEC* | | | | | |
| AXILLARY PAIN | 0 | 0 | 0 | 0 | 0 |
| CHEST DISCOMFORT | 0 | 0 | 0 | 0 | 0 |
| PAIN | 2 | 0 | 1 | 1 | 0 |
| TENDERNESS | 0 | 0 | 0 | 0 | 0 |
| **INFECTIONS AND INFESTATIONS** | | | | | |
| ***INFECTIONS - PATHOGEN UNSPECIFIED*** | | | | | |
| *BREAST INFECTIONS* | | | | | |
| MASTITIS | 1 | 0 | 0 | 1 | 0 |
| *SKIN STRUCTURES AND SOFT TISSUE INFECTIONS* | | | | | |
| INJECTION SITE PUSTULE | 0 | 0 | 0 | 0 | 0 |
| *UPPER RESPIRATORY TRACT INFECTIONS* | | | | | |
| LARYNGITIS | 0 | 0 | 0 | 0 | 0 |
| NASOPHARYNGITIS | 0 | 0 | 0 | 0 | 0 |
| ***VIRAL INFECTIOUS DISORDERS*** | | | | | |
| *HERPES VIRAL INFECTIONS* | | | | | |

| | | | | | |
|---|---|---|---|---|---|
| ORAL HERPES | 0 | 0 | 0 | 0 | 0 |
| *INFLUENZA VIRAL INFECTIONS* | | | | | |
| INFLUENZA | 3 | 1 | 2 | 0 | 0 |
| **INJURY, POISONING AND PROCEDURAL COMPLICATIONS** | | | | | |
| ***INJURIES NEC*** | | | | | |
| *SKIN INJURIES NEC* | | | | | |
| CONTUSION | 0 | 0 | 0 | 0 | 0 |
| **INVESTIGATIONS** | | | | | |
| ***CARDIAC AND VASCULAR INVESTIGATIONS (EXCL ENZYME TESTS)*** | | | | | |
| *HEART RATE AND PULSE INVESTIGATIONS* | | | | | |
| HEART RATE | 0 | 0 | 0 | 0 | 0 |
| ***PHYSICAL EXAMINATION AND ORGAN SYSTEM STATUS TOPICS*** | | | | | |
| *PHYSICAL EXAMINATION PROCEDURES AND ORGAN SYSTEM STATUS* | | | | | |
| BODY TEMPERATURE | 0 | 0 | 0 | 0 | 0 |
| BODY TEMPERATURE INCREASED | 1 | 0 | 0 | 1 | 0 |
| ***WATER, ELECTROLYTE AND MINERAL INVESTIGATIONS*** | | | | | |
| *WATER AND ELECTROLYTE ANALYSES NEC* | | | | | |
| VOLUME BLOOD | 0 | 0 | 0 | 0 | 0 |
| **METABOLISM AND NUTRITION DISORDERS** | | | | | |
| ***APPETITE AND GENERAL NUTRITIONAL DISORDERS*** | | | | | |
| *APPETITE DISORDERS* | | | | | |
| DECREASED APPETITE | 0 | 0 | 0 | 0 | 0 |
| **MUSCULOSKELETAL AND CONNECTIVE TISSUE DISORDERS** | | | | | |
| ***BONE DISORDERS (EXCL CONGENITAL AND FRACTURES)*** | | | | | |
| *BONE RELATED SIGNS AND SYMPTOMS* | | | | | |
| BONE PAIN | 0 | 0 | 0 | 0 | 0 |
| PAIN IN JAW | 0 | 0 | 0 | 0 | 0 |
| ***JOINT DISORDERS*** | | | | | |
| *JOINT RELATED SIGNS AND SYMPTOMS* | | | | | |
| ARTHRALGIA | 3 | 0 | 2 | 1 | 0 |
| JOINT STIFFNESS | 0 | 0 | 0 | 0 | 0 |
| ***MUSCLE DISORDERS*** | | | | | |
| *MUSCLE PAINS* | | | | | |
| MYALGIA | 9 | 2 | 3 | 4 | 0 |
| *MUSCLE RELATED SIGNS AND SYMPTOMS NEC* | | | | | |
| MUSCLE FATIGUE | 1 | 0 | 0 | 1 | 0 |
| MUSCLE SPASMS | 0 | 0 | 0 | 0 | 0 |
| ***MUSCULOSKELETAL AND CONNECTIVE TISSUE DISORDERS NEC*** | | | | | |
| *MUSCULOSKELETAL AND CONNECTIVE TISSUE CONDITIONS NEC* | | | | | |
| MUSCULOSKELETAL STIFFNESS | 0 | 0 | 0 | 0 | 0 |
| BACK PAIN | 0 | 0 | 0 | 0 | 0 |
| LIMB DISCOMFORT | 2 | 1 | 1 | 0 | 0 |
| PAIN IN EXTREMITY | 18 | 11 | 5 | 2 | 0 |
| **NERVOUS SYSTEM DISORDERS** | | | | | |
| ***CRANIAL NERVE DISORDERS (EXCL NEOPLASMS)*** | | | | | |
| *OLFACTORY NERVE DISORDERS* | | | | | |
| ANOSMIA | 0 | 0 | 0 | 0 | 0 |
| PAROSMIA | 0 | 0 | 0 | 0 | 0 |
| ***HEADACHES*** | | | | | |
| *HEADACHES NEC* | | | | | |
| HEADACHE | 12 | 3 | 4 | 5 | 0 |
| SINUS HEADACHE | 0 | 0 | 0 | 0 | 0 |
| TENSION HEADACHE | 0 | 0 | 0 | 0 | 0 |

| | | | | | |
|---|---|---|---|---|---|
| *MIGRAINE HEADACHES* | | | | | |
| MIGRAINE | 0 | 0 | 0 | 0 | 0 |
| TYPICAL AURA WITHOUT HEADACHE | 0 | 0 | 0 | 0 | 0 |
| ***MOVEMENT DISORDERS (INCL PARKINSONISM)*** | | | | | |
| *TREMOR (EXCL CONGENITAL)* | | | | | |
| TREMOR | 0 | 0 | 0 | 0 | 0 |
| ***NEUROLOGICAL DISORDERS NEC*** | | | | | |
| *DISTURBANCES IN CONSCIOUSNESS NEC* | | | | | |
| LETHARGY | 0 | 0 | 0 | 0 | 0 |
| SOMNOLENCE | 1 | 0 | 1 | 0 | 0 |
| *NEUROLOGICAL SIGNS AND SYMPTOMS NEC* | | | | | |
| BRAIN FOG | 0 | 0 | 0 | 0 | 0 |
| DIZZINESS | 3 | 1 | 1 | 1 | 0 |
| INFANT IRRITABILITY | 0 | 0 | 0 | 0 | 0 |
| NEUROLOGICAL SYMPTOM | 0 | 0 | 0 | 0 | 0 |
| PRESYNCOPE | 0 | 0 | 0 | 0 | 0 |
| *PARAESTHESIAS AND DYSAESTHESIAS* | | | | | |
| HYPOAESTHESIA | 0 | 0 | 0 | 0 | 0 |
| PARAESTHESIA | 0 | 0 | 0 | 0 | 0 |
| *SENSORY ABNORMALITIES NEC* | | | | | |
| AGEUSIA | 0 | 0 | 0 | 0 | 0 |
| DYSGEUSIA | 0 | 0 | 0 | 0 | 0 |
| ***SEIZURES (INCL SUBTYPES)*** | | | | | |
| *SEIZURES AND SEIZURE DISORDERS NEC* | | | | | |
| SEIZURE | 0 | 0 | 0 | 0 | 0 |
| **PREGNANCY, PUERPERIUM AND PERINATAL CONDITIONS** | | | | | |
| ***PLACENTAL, AMNIOTIC AND CAVITY DISORDERS (EXCL HAEMORRHAGES)*** | | | | | |
| *PLACENTAL ABNORMALITIES (EXCL NEOPLASMS)* | | | | | |
| PLACENTAL INFARCTION | 0 | 0 | 0 | 0 | 0 |
| **PSYCHIATRIC DISORDERS** | | | | | |
| ***ANXIETY DISORDERS AND SYMPTOMS*** | | | | | |
| *ANXIETY SYMPTOMS* | | | | | |
| AGITATION | 0 | 0 | 0 | 0 | 0 |
| ANXIETY | 0 | 0 | 0 | 0 | 0 |
| ***DELIRIA (INCL CONFUSION)*** | | | | | |
| *CONFUSION AND DISORIENTATION* | | | | | |
| CONFUSIONAL STATE | 0 | 0 | 0 | 0 | 0 |
| ***DEPRESSED MOOD DISORDERS AND DISTURBANCES*** | | | | | |
| *MOOD ALTERATIONS WITH DEPRESSIVE SYMPTOMS* | | | | | |
| DEPRESSED MOOD | 0 | 0 | 0 | 0 | 0 |
| ***MOOD DISORDERS AND DISTURBANCES NEC*** | | | | | |
| *EMOTIONAL AND MOOD DISTURBANCES NEC* | | | | | |
| EMOTIONAL DISORDER | 0 | 0 | 0 | 0 | 0 |
| IRRITABILITY | 0 | 0 | 0 | 0 | 0 |
| MOOD ALTERED | 1 | 0 | 0 | 1 | 0 |
| ***SLEEP DISORDERS AND DISTURBANCES*** | | | | | |
| *DISTURBANCES IN INITIATING AND MAINTAINING SLEEP* | | | | | |
| INSOMNIA | 0 | 0 | 0 | 0 | 0 |
| **REPRODUCTIVE SYSTEM AND BREAST DISORDERS** | | | | | |
| ***MENSTRUAL CYCLE AND UTERINE BLEEDING DISORDERS*** | | | | | |
| *MENSTRUATION AND UTERINE BLEEDING NEC* | | | | | |
| MENSTRUATION IRREGULAR | 0 | 0 | 0 | 0 | 0 |
| *MENSTRUATION WITH DECREASED BLEEDING* | | | | | |

| | | | | | |
|---|---|---|---|---|---|
| HYPOMENORRHOEA | 0 | 0 | 0 | 0 | 0 |
| MENSTRUATION DELAYED | 0 | 0 | 0 | 0 | 0 |
| OLIGOMENORRHOEA | 0 | 0 | 0 | 0 | 0 |
| *MENSTRUATION WITH INCREASED BLEEDING* | | | | | |
| HEAVY MENSTRUAL BLEEDING | 0 | 0 | 0 | 0 | 0 |
| ***VULVOVAGINAL DISORDERS (EXCL INFECTIONS AND INFLAMMATIONS)*** | | | | | |
| *VULVOVAGINAL DISORDERS NEC* | | | | | |
| VAGINAL HAEMORRHAGE | 0 | 0 | 0 | 0 | 0 |
| **RESPIRATORY, THORACIC AND MEDIASTINAL DISORDERS** | | | | | |
| ***RESPIRATORY DISORDERS NEC*** | | | | | |
| *BREATHING ABNORMALITIES* | | | | | |
| DYSPNOEA | 1 | 1 | 0 | 0 | 0 |
| *COUGHING AND ASSOCIATED SYMPTOMS* | | | | | |
| COUGH | 0 | 0 | 0 | 0 | 0 |
| ***RESPIRATORY TRACT SIGNS AND SYMPTOMS*** | | | | | |
| *UPPER RESPIRATORY TRACT SIGNS AND SYMPTOMS* | | | | | |
| OROPHARYNGEAL PAIN | 0 | 0 | 0 | 0 | 0 |
| PARANASAL SINUS DISCOMFORT | 0 | 0 | 0 | 0 | 0 |
| RHINORRHOEA | 1 | 0 | 0 | 1 | 0 |
| **SKIN AND SUBCUTANEOUS TISSUE DISORDERS** | | | | | |
| ***EPIDERMAL AND DERMAL CONDITIONS*** | | | | | |
| *DERMAL AND EPIDERMAL CONDITIONS NEC* | | | | | |
| SENSITIVE SKIN | 0 | 0 | 0 | 0 | 0 |
| *DERMATITIS AND ECZEMA* | | | | | |
| DERMATITIS ALLERGIC | 0 | 0 | 0 | 0 | 0 |
| *ERYTHEMAS* | | | | | |
| ERYTHEMA | 0 | 0 | 0 | 0 | 0 |
| *PRURITUS NEC* | | | | | |
| PRURITUS | 2 | 2 | 0 | 0 | 0 |
| *RASHES, ERUPTIONS AND EXANTHEMS NEC* | | | | | |
| RASH | 2 | 1 | 0 | 1 | 0 |
| ***SKIN APPENDAGE CONDITIONS*** | | | | | |
| *APOCRINE AND ECCRINE GLAND DISORDERS* | | | | | |
| COLD SWEAT | 0 | 0 | 0 | 0 | 0 |
| HYPERHIDROSIS | 0 | 0 | 0 | 0 | 0 |
| NIGHT SWEATS | 0 | 0 | 0 | 0 | 0 |
| **VASCULAR DISORDERS** | | | | | |
| ***EMBOLISM AND THROMBOSIS*** | | | | | |
| *NON-SITE SPECIFIC EMBOLISM AND THROMBOSIS* | | | | | |
| EMBOLISM | 1 | 1 | 0 | 0 | 0 |
| ***VASCULAR DISORDERS NEC*** | | | | | |
| *PERIPHERAL VASCULAR DISORDERS NEC* | | | | | |
| HOT FLUSH | 0 | 0 | 0 | 0 | 0 |
| **TOTAL ADR EVENTS** | ***122*** | ***36*** | ***41*** | ***45*** | ***0*** |

**TABLE S2-11. Others/Unknown COVID-19 vaccine: ADR listing for events reported by breastfeeding patients, in those reporting any vaccination dose**

| | Patients with any vaccination dose: ADR Counts | | | | |
|---|---|---|---|---|---|
| **REACTION TERM (SOC, *HLGT*, *HLT*, PT)** | All doses | 1st dose | 2nd dose | 3rd dose | Other doses |
| ***(freetext)*** | 0 | 0 | 0 | 0 | 0 |
| **BLOOD AND LYMPHATIC SYSTEM DISORDERS** | | | | | |
| *LYMPHATIC SYSTEM DISORDERS NEC* | | | | | |
| LYMPH NODE PAIN | 0 | 0 | 0 | 0 | 0 |
| LYMPHADENOPATHY | 0 | 0 | 0 | 0 | 0 |
| **CARDIAC DISORDERS** | | | | | |
| ***CARDIAC DISORDERS, SIGNS AND SYMPTOMS NEC*** | | | | | |
| *CARDIAC SIGNS AND SYMPTOMS NEC* | | | | | |
| PALPITATIONS | 0 | 0 | 0 | 0 | 0 |
| **EAR AND LABYRINTH DISORDERS** | | | | | |
| ***AURAL DISORDERS NEC*** | | | | | |
| *EAR DISORDERS NEC* | | | | | |
| EAR PAIN | 0 | 0 | 0 | 0 | 0 |
| ***INNER EAR AND VIIITH CRANIAL NERVE DISORDERS*** | | | | | |
| *INNER EAR SIGNS AND SYMPTOMS* | | | | | |
| TINNITUS | 0 | 0 | 0 | 0 | 0 |
| **EYE DISORDERS** | | | | | |
| ***EYE DISORDERS NEC*** | | | | | |
| *LACRIMATION DISORDERS* | | | | | |
| DRY EYE | 0 | 0 | 0 | 0 | 0 |
| *OCULAR DISORDERS NEC* | | | | | |
| EYE PAIN | 0 | 0 | 0 | 0 | 0 |
| ***OCULAR INFECTIONS, IRRITATIONS AND INFLAMMATIONS*** | | | | | |
| *OCULAR INFECTIONS, INFLAMMATIONS AND ASSOCIATED MANIFESTATIONS* | | | | | |
| EYE IRRITATION | 0 | 0 | 0 | 0 | 0 |
| ***OCULAR SENSORY SYMPTOMS NEC*** | | | | | |
| *OCULAR SENSATION DISORDERS* | | | | | |
| PHOTOPHOBIA | 0 | 0 | 0 | 0 | 0 |
| ***VISION DISORDERS*** | | | | | |
| *VISUAL DISORDERS NEC* | | | | | |
| VISION BLURRED | 0 | 0 | 0 | 0 | 0 |
| **GASTROINTESTINAL DISORDERS** | | | | | |
| ***GASTROINTESTINAL INFLAMMATORY CONDITIONS*** | | | | | |
| *GASTROINTESTINAL INFLAMMATORY DISORDERS NEC* | | | | | |
| GASTROINTESTINAL TRACT IRRITATION | 0 | 0 | 0 | 0 | 0 |
| ***GASTROINTESTINAL MOTILITY AND DEFAECATION CONDITIONS*** | | | | | |
| *DIARRHOEA (EXCL INFECTIVE)* | | | | | |
| DIARRHOEA | 0 | 0 | 0 | 0 | 0 |
| ***GASTROINTESTINAL SIGNS AND SYMPTOMS*** | | | | | |
| *FLATULENCE, BLOATING AND DISTENSION* | | | | | |
| ABDOMINAL DISTENSION | 0 | 0 | 0 | 0 | 0 |
| FLATULENCE | 0 | 0 | 0 | 0 | 0 |
| *GASTROINTESTINAL AND ABDOMINAL PAINS (EXCL ORAL AND THROAT)* | | | | | |
| ABDOMINAL PAIN | 0 | 0 | 0 | 0 | 0 |
| ABDOMINAL PAIN UPPER | 0 | 0 | 0 | 0 | 0 |

| | | | | | |
|---|---|---|---|---|---|
| *NAUSEA AND VOMITING SYMPTOMS* | | | | | |
| NAUSEA | 1 | 0 | 0 | 1 | 0 |
| RETCHING | 0 | 0 | 0 | 0 | 0 |
| VOMITING | 0 | 0 | 0 | 0 | 0 |
| ***ORAL SOFT TISSUE CONDITIONS*** | | | | | |
| *ORAL SOFT TISSUE SIGNS AND SYMPTOMS* | | | | | |
| PARAESTHESIA ORAL | 0 | 0 | 0 | 0 | 0 |
| LIP SWELLING | 0 | 0 | 0 | 0 | 0 |
| **GENERAL DISORDERS AND ADMINISTRATION SITE CONDITIONS** | | | | | |
| ***ADMINISTRATION SITE REACTIONS*** | | | | | |
| *ADMINISTRATION SITE REACTIONS NEC* | | | | | |
| PUNCTURE SITE BRUISE | 0 | 0 | 0 | 0 | 0 |
| *INJECTION SITE REACTIONS* | | | | | |
| INJECTION SITE ERYTHEMA | 0 | 0 | 0 | 0 | 0 |
| INJECTION SITE MASS | 0 | 0 | 0 | 0 | 0 |
| INJECTION SITE PAIN | 0 | 0 | 0 | 0 | 0 |
| INJECTION SITE WARMTH | 0 | 0 | 0 | 0 | 0 |
| *VACCINATION SITE REACTIONS* | | | | | |
| VACCINATION SITE PAIN | 0 | 0 | 0 | 0 | 0 |
| ***BODY TEMPERATURE CONDITIONS*** | | | | | |
| *FEBRILE DISORDERS* | | | | | |
| PYREXIA | 0 | 0 | 0 | 0 | 0 |
| ***GENERAL SYSTEM DISORDERS NEC*** | | | | | |
| *ASTHENIC CONDITIONS* | | | | | |
| FATIGUE | 0 | 0 | 0 | 0 | 0 |
| MALAISE | 0 | 0 | 0 | 0 | 0 |
| *FEELINGS AND SENSATIONS NEC* | | | | | |
| CHILLS | 0 | 0 | 0 | 0 | 0 |
| FEELING ABNORMAL | 0 | 0 | 0 | 0 | 0 |
| FEELING COLD | 0 | 0 | 0 | 0 | 0 |
| FEELING HOT | 0 | 0 | 0 | 0 | 0 |
| HANGOVER | 0 | 0 | 0 | 0 | 0 |
| *GENERAL SIGNS AND SYMPTOMS NEC* | | | | | |
| INFLUENZA LIKE ILLNESS | 0 | 0 | 0 | 0 | 0 |
| LOCAL REACTION | 0 | 0 | 0 | 0 | 0 |
| PERIPHERAL SWELLING | 0 | 0 | 0 | 0 | 0 |
| SWELLING | 0 | 0 | 0 | 0 | 0 |
| *PAIN AND DISCOMFORT NEC* | | | | | |
| AXILLARY PAIN | 0 | 0 | 0 | 0 | 0 |
| CHEST DISCOMFORT | 0 | 0 | 0 | 0 | 0 |
| PAIN | 1 | 0 | 0 | 1 | 0 |
| TENDERNESS | 0 | 0 | 0 | 0 | 0 |
| **INFECTIONS AND INFESTATIONS** | | | | | |
| ***INFECTIONS - PATHOGEN UNSPECIFIED*** | | | | | |
| *BREAST INFECTIONS* | | | | | |
| MASTITIS | 0 | 0 | 0 | 0 | 0 |
| *SKIN STRUCTURES AND SOFT TISSUE INFECTIONS* | | | | | |
| INJECTION SITE PUSTULE | 0 | 0 | 0 | 0 | 0 |
| *UPPER RESPIRATORY TRACT INFECTIONS* | | | | | |
| LARYNGITIS | 0 | 0 | 0 | 0 | 0 |
| NASOPHARYNGITIS | 0 | 0 | 0 | 0 | 0 |
| ***VIRAL INFECTIOUS DISORDERS*** | | | | | |
| *HERPES VIRAL INFECTIONS* | | | | | |

| | | | | | |
|---|---|---|---|---|---|
| ORAL HERPES | 0 | 0 | 0 | 0 | 0 |
| *INFLUENZA VIRAL INFECTIONS* | | | | | |
| INFLUENZA | 0 | 0 | 0 | 0 | 0 |
| **INJURY, POISONING AND PROCEDURAL COMPLICATIONS** | | | | | |
| ***INJURIES NEC*** | | | | | |
| *SKIN INJURIES NEC* | | | | | |
| CONTUSION | 0 | 0 | 0 | 0 | 0 |
| **INVESTIGATIONS** | | | | | |
| ***CARDIAC AND VASCULAR INVESTIGATIONS (EXCL ENZYME TESTS)*** | | | | | |
| *HEART RATE AND PULSE INVESTIGATIONS* | | | | | |
| HEART RATE | 0 | 0 | 0 | 0 | 0 |
| ***PHYSICAL EXAMINATION AND ORGAN SYSTEM STATUS TOPICS*** | | | | | |
| *PHYSICAL EXAMINATION PROCEDURES AND ORGAN SYSTEM STATUS* | | | | | |
| BODY TEMPERATURE | 0 | 0 | 0 | 0 | 0 |
| BODY TEMPERATURE INCREASED | 0 | 0 | 0 | 0 | 0 |
| ***WATER, ELECTROLYTE AND MINERAL INVESTIGATIONS*** | | | | | |
| *WATER AND ELECTROLYTE ANALYSES NEC* | | | | | |
| VOLUME BLOOD | 0 | 0 | 0 | 0 | 0 |
| **METABOLISM AND NUTRITION DISORDERS** | | | | | |
| ***APPETITE AND GENERAL NUTRITIONAL DISORDERS*** | | | | | |
| *APPETITE DISORDERS* | | | | | |
| DECREASED APPETITE | 0 | 0 | 0 | 0 | 0 |
| **MUSCULOSKELETAL AND CONNECTIVE TISSUE DISORDERS** | | | | | |
| ***BONE DISORDERS (EXCL CONGENITAL AND FRACTURES)*** | | | | | |
| *BONE RELATED SIGNS AND SYMPTOMS* | | | | | |
| BONE PAIN | 0 | 0 | 0 | 0 | 0 |
| PAIN IN JAW | 0 | 0 | 0 | 0 | 0 |
| ***JOINT DISORDERS*** | | | | | |
| *JOINT RELATED SIGNS AND SYMPTOMS* | | | | | |
| ARTHRALGIA | 0 | 0 | 0 | 0 | 0 |
| JOINT STIFFNESS | 0 | 0 | 0 | 0 | 0 |
| ***MUSCLE DISORDERS*** | | | | | |
| *MUSCLE PAINS* | | | | | |
| MYALGIA | 0 | 0 | 0 | 0 | 0 |
| *MUSCLE RELATED SIGNS AND SYMPTOMS NEC* | | | | | |
| MUSCLE FATIGUE | 0 | 0 | 0 | 0 | 0 |
| MUSCLE SPASMS | 0 | 0 | 0 | 0 | 0 |
| ***MUSCULOSKELETAL AND CONNECTIVE TISSUE DISORDERS NEC*** | | | | | |
| *MUSCULOSKELETAL AND CONNECTIVE TISSUE CONDITIONS NEC* | | | | | |
| MUSCULOSKELETAL STIFFNESS | 0 | 0 | 0 | 0 | 0 |
| BACK PAIN | 0 | 0 | 0 | 0 | 0 |
| LIMB DISCOMFORT | 0 | 0 | 0 | 0 | 0 |
| PAIN IN EXTREMITY | 0 | 0 | 0 | 0 | 0 |
| **NERVOUS SYSTEM DISORDERS** | | | | | |
| ***CRANIAL NERVE DISORDERS (EXCL NEOPLASMS)*** | | | | | |
| *OLFACTORY NERVE DISORDERS* | | | | | |
| ANOSMIA | 0 | 0 | 0 | 0 | 0 |
| PAROSMIA | 0 | 0 | 0 | 0 | 0 |
| ***HEADACHES*** | | | | | |
| *HEADACHES NEC* | | | | | |
| HEADACHE | 1 | 0 | 0 | 1 | 0 |
| SINUS HEADACHE | 0 | 0 | 0 | 0 | 0 |
| TENSION HEADACHE | 0 | 0 | 0 | 0 | 0 |

| | | | | | |
|---|---|---|---|---|---|
| *MIGRAINE HEADACHES* | | | | | |
| MIGRAINE | 0 | 0 | 0 | 0 | 0 |
| TYPICAL AURA WITHOUT HEADACHE | 0 | 0 | 0 | 0 | 0 |
| ***MOVEMENT DISORDERS (INCL PARKINSONISM)*** | | | | | |
| *TREMOR (EXCL CONGENITAL)* | | | | | |
| TREMOR | 0 | 0 | 0 | 0 | 0 |
| ***NEUROLOGICAL DISORDERS NEC*** | | | | | |
| *DISTURBANCES IN CONSCIOUSNESS NEC* | | | | | |
| LETHARGY | 0 | 0 | 0 | 0 | 0 |
| SOMNOLENCE | 0 | 0 | 0 | 0 | 0 |
| *NEUROLOGICAL SIGNS AND SYMPTOMS NEC* | | | | | |
| BRAIN FOG | 0 | 0 | 0 | 0 | 0 |
| DIZZINESS | 0 | 0 | 0 | 0 | 0 |
| INFANT IRRITABILITY | 0 | 0 | 0 | 0 | 0 |
| NEUROLOGICAL SYMPTOM | 0 | 0 | 0 | 0 | 0 |
| PRESYNCOPE | 0 | 0 | 0 | 0 | 0 |
| *PARAESTHESIAS AND DYSAESTHESIAS* | | | | | |
| HYPOAESTHESIA | 0 | 0 | 0 | 0 | 0 |
| PARAESTHESIA | 0 | 0 | 0 | 0 | 0 |
| *SENSORY ABNORMALITIES NEC* | | | | | |
| AGEUSIA | 0 | 0 | 0 | 0 | 0 |
| DYSGEUSIA | 0 | 0 | 0 | 0 | 0 |
| ***SEIZURES (INCL SUBTYPES)*** | | | | | |
| *SEIZURES AND SEIZURE DISORDERS NEC* | | | | | |
| SEIZURE | 0 | 0 | 0 | 0 | 0 |
| **PREGNANCY, PUERPERIUM AND PERINATAL CONDITIONS** | | | | | |
| ***PLACENTAL, AMNIOTIC AND CAVITY DISORDERS (EXCL HAEMORRHAGES)*** | | | | | |
| *PLACENTAL ABNORMALITIES (EXCL NEOPLASMS)* | | | | | |
| PLACENTAL INFARCTION | 0 | 0 | 0 | 0 | 0 |
| **PSYCHIATRIC DISORDERS** | | | | | |
| ***ANXIETY DISORDERS AND SYMPTOMS*** | | | | | |
| *ANXIETY SYMPTOMS* | | | | | |
| AGITATION | 0 | 0 | 0 | 0 | 0 |
| ANXIETY | 0 | 0 | 0 | 0 | 0 |
| ***DELIRIA (INCL CONFUSION)*** | | | | | |
| *CONFUSION AND DISORIENTATION* | | | | | |
| CONFUSIONAL STATE | 0 | 0 | 0 | 0 | 0 |
| ***DEPRESSED MOOD DISORDERS AND DISTURBANCES*** | | | | | |
| *MOOD ALTERATIONS WITH DEPRESSIVE SYMPTOMS* | | | | | |
| DEPRESSED MOOD | 0 | 0 | 0 | 0 | 0 |
| ***MOOD DISORDERS AND DISTURBANCES NEC*** | | | | | |
| *EMOTIONAL AND MOOD DISTURBANCES NEC* | | | | | |
| EMOTIONAL DISORDER | 0 | 0 | 0 | 0 | 0 |
| IRRITABILITY | 0 | 0 | 0 | 0 | 0 |
| MOOD ALTERED | 0 | 0 | 0 | 0 | 0 |
| ***SLEEP DISORDERS AND DISTURBANCES*** | | | | | |
| *DISTURBANCES IN INITIATING AND MAINTAINING SLEEP* | | | | | |
| INSOMNIA | 0 | 0 | 0 | 0 | 0 |
| **REPRODUCTIVE SYSTEM AND BREAST DISORDERS** | | | | | |
| ***MENSTRUAL CYCLE AND UTERINE BLEEDING DISORDERS*** | | | | | |
| *MENSTRUATION AND UTERINE BLEEDING NEC* | | | | | |
| MENSTRUATION IRREGULAR | 0 | 0 | 0 | 0 | 0 |
| *MENSTRUATION WITH DECREASED BLEEDING* | | | | | |

| | | | | | |
|---|---|---|---|---|---|
| HYPOMENORRHOEA | 0 | 0 | 0 | 0 | 0 |
| MENSTRUATION DELAYED | 0 | 0 | 0 | 0 | 0 |
| OLIGOMENORRHOEA | 0 | 0 | 0 | 0 | 0 |
| *MENSTRUATION WITH INCREASED BLEEDING* | | | | | |
| HEAVY MENSTRUAL BLEEDING | 0 | 0 | 0 | 0 | 0 |
| ***VULVOVAGINAL DISORDERS (EXCL INFECTIONS AND INFLAMMATIONS)*** | | | | | |
| *VULVOVAGINAL DISORDERS NEC* | | | | | |
| VAGINAL HAEMORRHAGE | 0 | 0 | 0 | 0 | 0 |
| **RESPIRATORY, THORACIC AND MEDIASTINAL DISORDERS** | | | | | |
| ***RESPIRATORY DISORDERS NEC*** | | | | | |
| *BREATHING ABNORMALITIES* | | | | | |
| DYSPNOEA | 0 | 0 | 0 | 0 | 0 |
| *COUGHING AND ASSOCIATED SYMPTOMS* | | | | | |
| COUGH | 0 | 0 | 0 | 0 | 0 |
| ***RESPIRATORY TRACT SIGNS AND SYMPTOMS*** | | | | | |
| *UPPER RESPIRATORY TRACT SIGNS AND SYMPTOMS* | | | | | |
| OROPHARYNGEAL PAIN | 0 | 0 | 0 | 0 | 0 |
| PARANASAL SINUS DISCOMFORT | 0 | 0 | 0 | 0 | 0 |
| RHINORRHOEA | 0 | 0 | 0 | 0 | 0 |
| **SKIN AND SUBCUTANEOUS TISSUE DISORDERS** | | | | | |
| ***EPIDERMAL AND DERMAL CONDITIONS*** | | | | | |
| *DERMAL AND EPIDERMAL CONDITIONS NEC* | | | | | |
| SENSITIVE SKIN | 0 | 0 | 0 | 0 | 0 |
| *DERMATITIS AND ECZEMA* | | | | | |
| DERMATITIS ALLERGIC | 0 | 0 | 0 | 0 | 0 |
| *ERYTHEMAS* | | | | | |
| ERYTHEMA | 0 | 0 | 0 | 0 | 0 |
| *PRURITUS NEC* | | | | | |
| PRURITUS | 0 | 0 | 0 | 0 | 0 |
| *RASHES, ERUPTIONS AND EXANTHEMS NEC* | | | | | |
| RASH | 0 | 0 | 0 | 0 | 0 |
| ***SKIN APPENDAGE CONDITIONS*** | | | | | |
| *APOCRINE AND ECCRINE GLAND DISORDERS* | | | | | |
| COLD SWEAT | 0 | 0 | 0 | 0 | 0 |
| HYPERHIDROSIS | 0 | 0 | 0 | 0 | 0 |
| NIGHT SWEATS | 0 | 0 | 0 | 0 | 0 |
| **VASCULAR DISORDERS** | | | | | |
| ***EMBOLISM AND THROMBOSIS*** | | | | | |
| *NON-SITE SPECIFIC EMBOLISM AND THROMBOSIS* | | | | | |
| EMBOLISM | 0 | 0 | 0 | 0 | 0 |
| ***VASCULAR DISORDERS NEC*** | | | | | |
| *PERIPHERAL VASCULAR DISORDERS NEC* | | | | | |
| HOT FLUSH | 0 | 0 | 0 | 0 | 0 |
| **TOTAL ADR EVENTS** | ***3*** | ***0*** | ***0*** | ***3*** | ***0*** |

**TABLE S2-12. COVID-19 vaccine ADR listings for events reported by breast-feeding patients with no dose identification information in those reporting any vaccination dose**

| | Patients with any vaccination dose: ADR Counts |
|---|---|
| **REACTION TERM (SOC, *HLGT*, *HLT*, PT)** | Unknown dose |
| ***(freetext)*** | |
| **BLOOD AND LYMPHATIC SYSTEM DISORDERS** | |
| *LYMPHATIC SYSTEM DISORDERS NEC* | |
| LYMPH NODE PAIN | 0 |
| LYMPHADENOPATHY | 1 |
| **CARDIAC DISORDERS** | |
| ***CARDIAC DISORDERS, SIGNS AND SYMPTOMS NEC*** | |
| *CARDIAC SIGNS AND SYMPTOMS NEC* | |
| PALPITATIONS | 2 |
| **EAR AND LABYRINTH DISORDERS** | |
| ***AURAL DISORDERS NEC*** | |
| *EAR DISORDERS NEC* | |
| EAR PAIN | 0 |
| ***INNER EAR AND VIIITH CRANIAL NERVE DISORDERS*** | |
| *INNER EAR SIGNS AND SYMPTOMS* | |
| TINNITUS | 0 |
| **EYE DISORDERS** | |
| ***EYE DISORDERS NEC*** | |
| *LACRIMATION DISORDERS* | |
| DRY EYE | 0 |
| *OCULAR DISORDERS NEC* | |
| EYE PAIN | 0 |
| ***OCULAR INFECTIONS, IRRITATIONS AND INFLAMMATIONS*** | |
| *OCULAR INFECTIONS, INFLAMMATIONS AND ASSOCIATED MANIFESTATIONS* | |
| EYE IRRITATION | 0 |
| ***OCULAR SENSORY SYMPTOMS NEC*** | |
| *OCULAR SENSATION DISORDERS* | |
| PHOTOPHOBIA | 0 |
| ***VISION DISORDERS*** | |
| *VISUAL DISORDERS NEC* | |
| VISION BLURRED | 0 |
| **GASTROINTESTINAL DISORDERS** | |
| ***GASTROINTESTINAL INFLAMMATORY CONDITIONS*** | |
| *GASTROINTESTINAL INFLAMMATORY DISORDERS NEC* | |
| GASTROINTESTINAL TRACT IRRITATION | 0 |
| ***GASTROINTESTINAL MOTILITY AND DEFAECATION CONDITIONS*** | |
| *DIARRHOEA (EXCL INFECTIVE)* | |
| DIARRHOEA | 0 |
| ***GASTROINTESTINAL SIGNS AND SYMPTOMS*** | |
| *FLATULENCE, BLOATING AND DISTENSION* | |
| ABDOMINAL DISTENSION | 0 |
| FLATULENCE | 0 |
| *GASTROINTESTINAL AND ABDOMINAL PAINS (EXCL ORAL AND THROAT)* | |
| ABDOMINAL PAIN | 0 |

| | |
|---|---|
| ABDOMINAL PAIN UPPER | 0 |
| *NAUSEA AND VOMITING SYMPTOMS* | |
| NAUSEA | 4 |
| RETCHING | 0 |
| VOMITING | 0 |
| ***ORAL SOFT TISSUE CONDITIONS*** | |
| *ORAL SOFT TISSUE SIGNS AND SYMPTOMS* | |
| PARAESTHESIA ORAL | 0 |
| LIP SWELLING | 0 |
| **GENERAL DISORDERS AND ADMINISTRATION SITE CONDITIONS** | |
| ***ADMINISTRATION SITE REACTIONS*** | |
| *ADMINISTRATION SITE REACTIONS NEC* | |
| PUNCTURE SITE BRUISE | 0 |
| *INJECTION SITE REACTIONS* | |
| INJECTION SITE ERYTHEMA | 0 |
| INJECTION SITE MASS | 1 |
| INJECTION SITE PAIN | 3 |
| INJECTION SITE WARMTH | 0 |
| *VACCINATION SITE REACTIONS* | |
| VACCINATION SITE PAIN | 0 |
| ***BODY TEMPERATURE CONDITIONS*** | |
| *FEBRILE DISORDERS* | |
| PYREXIA | 5 |
| ***GENERAL SYSTEM DISORDERS NEC*** | |
| *ASTHENIC CONDITIONS* | |
| FATIGUE | 8 |
| MALAISE | 0 |
| *FEELINGS AND SENSATIONS NEC* | |
| CHILLS | 3 |
| FEELING ABNORMAL | 0 |
| FEELING COLD | 0 |
| FEELING HOT | 0 |
| HANGOVER | 0 |
| *GENERAL SIGNS AND SYMPTOMS NEC* | |
| INFLUENZA LIKE ILLNESS | 1 |
| LOCAL REACTION | 0 |
| PERIPHERAL SWELLING | 0 |
| SWELLING | 0 |
| *PAIN AND DISCOMFORT NEC* | |
| AXILLARY PAIN | 0 |
| CHEST DISCOMFORT | 0 |
| PAIN | 2 |
| TENDERNESS | 0 |
| **INFECTIONS AND INFESTATIONS** | |
| ***INFECTIONS - PATHOGEN UNSPECIFIED*** | |
| *BREAST INFECTIONS* | |
| MASTITIS | 0 |
| *SKIN STRUCTURES AND SOFT TISSUE INFECTIONS* | |
| INJECTION SITE PUSTULE | 0 |
| *UPPER RESPIRATORY TRACT INFECTIONS* | |
| LARYNGITIS | 0 |
| NASOPHARYNGITIS | 0 |
| ***VIRAL INFECTIOUS DISORDERS*** | |
| *HERPES VIRAL INFECTIONS* | |
| ORAL HERPES | 0 |
| *INFLUENZA VIRAL INFECTIONS* | |

| | |
|---|---|
| INFLUENZA | 0 |
| **INJURY, POISONING AND PROCEDURAL COMPLICATIONS** | |
| ***INJURIES NEC*** | |
| *SKIN INJURIES NEC* | |
| CONTUSION | 0 |
| **INVESTIGATIONS** | |
| ***CARDIAC AND VASCULAR INVESTIGATIONS (EXCL ENZYME TESTS)*** | |
| *HEART RATE AND PULSE INVESTIGATIONS* | |
| HEART RATE | 0 |
| ***PHYSICAL EXAMINATION AND ORGAN SYSTEM STATUS TOPICS*** | |
| *PHYSICAL EXAMINATION PROCEDURES AND ORGAN SYSTEM STATUS* | |
| BODY TEMPERATURE | 0 |
| BODY TEMPERATURE INCREASED | 0 |
| ***WATER, ELECTROLYTE AND MINERAL INVESTIGATIONS*** | |
| *WATER AND ELECTROLYTE ANALYSES NEC* | |
| VOLUME BLOOD | 0 |
| **METABOLISM AND NUTRITION DISORDERS** | |
| ***APPETITE AND GENERAL NUTRITIONAL DISORDERS*** | |
| *APPETITE DISORDERS* | |
| DECREASED APPETITE | 0 |
| **MUSCULOSKELETAL AND CONNECTIVE TISSUE DISORDERS** | |
| ***BONE DISORDERS (EXCL CONGENITAL AND FRACTURES)*** | |
| *BONE RELATED SIGNS AND SYMPTOMS* | |
| BONE PAIN | 0 |
| PAIN IN JAW | 0 |
| ***JOINT DISORDERS*** | |
| *JOINT RELATED SIGNS AND SYMPTOMS* | |
| ARTHRALGIA | 2 |
| JOINT STIFFNESS | 0 |
| ***MUSCLE DISORDERS*** | |
| *MUSCLE PAINS* | |
| MYALGIA | 2 |
| *MUSCLE RELATED SIGNS AND SYMPTOMS NEC* | |
| MUSCLE FATIGUE | 0 |
| MUSCLE SPASMS | 0 |
| ***MUSCULOSKELETAL AND CONNECTIVE TISSUE DISORDERS NEC*** | |
| *MUSCULOSKELETAL AND CONNECTIVE TISSUE CONDITIONS NEC* | |
| MUSCULOSKELETAL STIFFNESS | 0 |
| BACK PAIN | 0 |
| LIMB DISCOMFORT | 0 |
| PAIN IN EXTREMITY | 3 |
| **NERVOUS SYSTEM DISORDERS** | |
| ***CRANIAL NERVE DISORDERS (EXCL NEOPLASMS)*** | |
| *OLFACTORY NERVE DISORDERS* | |
| ANOSMIA | 0 |
| PAROSMIA | 0 |
| ***HEADACHES*** | |
| *HEADACHES NEC* | |
| HEADACHE | 8 |
| SINUS HEADACHE | 0 |
| TENSION HEADACHE | 0 |
| *MIGRAINE HEADACHES* | |
| MIGRAINE | 1 |
| TYPICAL AURA WITHOUT HEADACHE | 0 |
| ***MOVEMENT DISORDERS (INCL PARKINSONISM)*** | |
| *TREMOR (EXCL CONGENITAL)* | |

| | |
|---|---|
| TREMOR | 1 |
| ***NEUROLOGICAL DISORDERS NEC*** | |
| *DISTURBANCES IN CONSCIOUSNESS NEC* | |
| LETHARGY | 0 |
| SOMNOLENCE | 0 |
| *NEUROLOGICAL SIGNS AND SYMPTOMS NEC* | |
| BRAIN FOG | 0 |
| DIZZINESS | 3 |
| INFANT IRRITABILITY | 0 |
| NEUROLOGICAL SYMPTOM | 0 |
| PRESYNCOPE | 0 |
| *PARAESTHESIAS AND DYSAESTHESIAS* | |
| HYPOAESTHESIA | 0 |
| PARAESTHESIA | 0 |
| *SENSORY ABNORMALITIES NEC* | |
| AGEUSIA | 0 |
| DYSGEUSIA | 0 |
| ***SEIZURES (INCL SUBTYPES)*** | |
| *SEIZURES AND SEIZURE DISORDERS NEC* | |
| SEIZURE | 0 |
| **PREGNANCY, PUERPERIUM AND PERINATAL CONDITIONS** | |
| ***PLACENTAL, AMNIOTIC AND CAVITY DISORDERS (EXCL HAEMORRHAGES)*** | |
| *PLACENTAL ABNORMALITIES (EXCL NEOPLASMS)* | |
| PLACENTAL INFARCTION | 0 |
| **PSYCHIATRIC DISORDERS** | |
| ***ANXIETY DISORDERS AND SYMPTOMS*** | |
| *ANXIETY SYMPTOMS* | |
| AGITATION | 0 |
| ANXIETY | 2 |
| ***DELIRIA (INCL CONFUSION)*** | |
| *CONFUSION AND DISORIENTATION* | |
| CONFUSIONAL STATE | 0 |
| ***DEPRESSED MOOD DISORDERS AND DISTURBANCES*** | |
| *MOOD ALTERATIONS WITH DEPRESSIVE SYMPTOMS* | |
| DEPRESSED MOOD | 0 |
| ***MOOD DISORDERS AND DISTURBANCES NEC*** | |
| *EMOTIONAL AND MOOD DISTURBANCES NEC* | |
| EMOTIONAL DISORDER | 0 |
| IRRITABILITY | 0 |
| MOOD ALTERED | 0 |
| ***SLEEP DISORDERS AND DISTURBANCES*** | |
| *DISTURBANCES IN INITIATING AND MAINTAINING SLEEP* | |
| INSOMNIA | 0 |
| **REPRODUCTIVE SYSTEM AND BREAST DISORDERS** | |
| ***MENSTRUAL CYCLE AND UTERINE BLEEDING DISORDERS*** | |
| *MENSTRUATION AND UTERINE BLEEDING NEC* | |
| MENSTRUATION IRREGULAR | 0 |
| *MENSTRUATION WITH DECREASED BLEEDING* | |
| HYPOMENORRHOEA | 0 |
| MENSTRUATION DELAYED | 0 |
| OLIGOMENORRHOEA | 1 |
| *MENSTRUATION WITH INCREASED BLEEDING* | |
| HEAVY MENSTRUAL BLEEDING | 0 |
| ***VULVOVAGINAL DISORDERS (EXCL INFECTIONS AND INFLAMMATIONS)*** | |
| *VULVOVAGINAL DISORDERS NEC* | |
| VAGINAL HAEMORRHAGE | 0 |

| | |
|---|---|
| **RESPIRATORY, THORACIC AND MEDIASTINAL DISORDERS** | |
| ***RESPIRATORY DISORDERS NEC*** | |
| *BREATHING ABNORMALITIES* | |
| DYSPNOEA | 0 |
| *COUGHING AND ASSOCIATED SYMPTOMS* | |
| COUGH | 0 |
| ***RESPIRATORY TRACT SIGNS AND SYMPTOMS*** | |
| *UPPER RESPIRATORY TRACT SIGNS AND SYMPTOMS* | |
| OROPHARYNGEAL PAIN | 0 |
| PARANASAL SINUS DISCOMFORT | 0 |
| RHINORRHOEA | 0 |
| **SKIN AND SUBCUTANEOUS TISSUE DISORDERS** | |
| ***EPIDERMAL AND DERMAL CONDITIONS*** | |
| *DERMAL AND EPIDERMAL CONDITIONS NEC* | |
| SENSITIVE SKIN | 0 |
| *DERMATITIS AND ECZEMA* | |
| DERMATITIS ALLERGIC | 0 |
| *ERYTHEMAS* | |
| ERYTHEMA | 2 |
| *PRURITUS NEC* | |
| PRURITUS | 0 |
| *RASHES, ERUPTIONS AND EXANTHEMS NEC* | |
| RASH | 0 |
| ***SKIN APPENDAGE CONDITIONS*** | |
| *APOCRINE AND ECCRINE GLAND DISORDERS* | |
| COLD SWEAT | 0 |
| HYPERHIDROSIS | 1 |
| NIGHT SWEATS | 0 |
| **VASCULAR DISORDERS** | |
| ***EMBOLISM AND THROMBOSIS*** | |
| *NON-SITE SPECIFIC EMBOLISM AND THROMBOSIS* | |
| EMBOLISM | 0 |
| ***VASCULAR DISORDERS NEC*** | |
| *PERIPHERAL VASCULAR DISORDERS NEC* | |
| HOT FLUSH | 0 |
| **TOTAL ADR EVENTS** | ***56*** |
